## Supplementary Data for "Multiplex Target-Redundant RT-LAMP for Robust Detection of SARS-CoV-2 Using Fluorescent Universal Displacement Probes"

### Supplementary Information

**CFX Maestro Software settings configuration note:**

FAM channel baseline was set as 2-35cycles with a manual threshold of 50 RFU. Texas Red channel baseline was set as 20-60 cycles with a manual threshold of 50RFU.

**supp. Fig. S1: Validation of individual targets in the multiplex assay (challenged with fragments corresponding to individual targets)**

**A.**

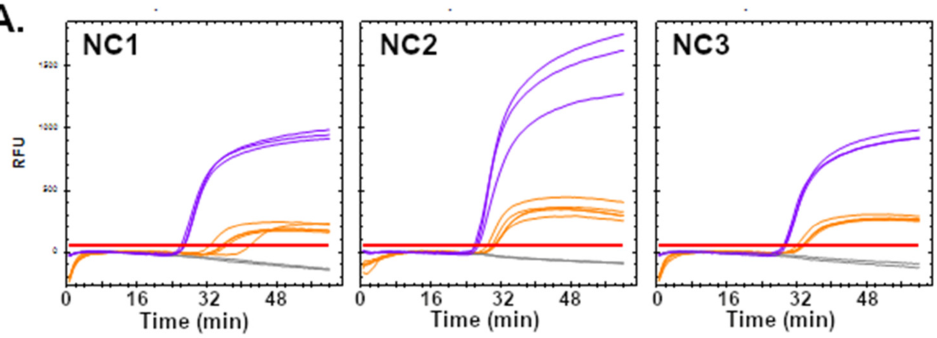


**B.**

| LAMP Target | NC1 | NC2 | NC3 |
|---|---|---|---|
| Target TTD (min) | 26.6, 26.8,25.9 | 25.9, 26.3,26.7 | 28.8, 29.1,29.4 |

**supp. Fig. S1: Validation of individual targets in the multiplex assay (challenged with fragments corresponding to individual targets) A)** The assay was tested with synthetic RNA fragments containing the NC1 sequence, NC2 sequence, or the NC3 sequence as separate fragments. All IAC signals (TEX615) are shown in red, SARS-CoV-2 signals (FAM) from reactions with 0 copies/NTC (n=2) are shown in gray, and SARS-CoV-2 signals (FAM) from reactions with 200 copies (n=3) are shown in blue. **B)** Time for detection of SARS-CoV-2 signals for the assay with individual target inputs from the plots in panel **A**.

**supp. Fig. S2: Effect of commonly used transport medium / contrived specimens on mRT-LAMP**

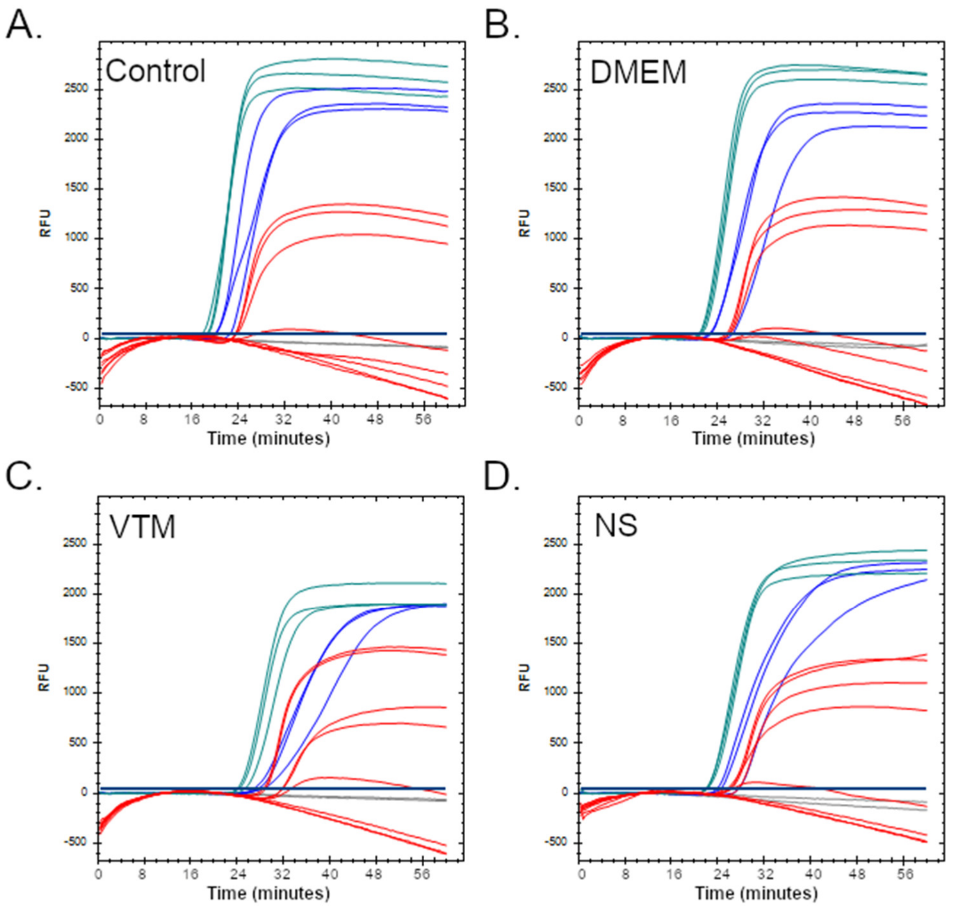


**supp. Fig. S2: Effect of commonly used transport medium / contrived mRT-LAMP** A) Water control (25% final) B) DMEM (25% final). C) VTM (25% final). D) NS (25% final). All IAC signals are shown in red (1e5 copies). SARS-CoV-2 signals for multiplexed reactions are shown as: 0 copies RNA/ NTC in gray (n=2), 20 copies RNA in blue (n=3), 200 copies RNA in teal (n=3). Thresholds are shown as blue and red lines at 50 RFU.

**supp. Table S1:** Clinical specimen panel

| ID | Source | N1 copies | N2 copies | RP Ct | PCR result | COVID LAMP 1 (min) | IAC LAMP 1 (min) | COVID LAMP 2 (min) | IAC LAMP 2 (min) | LAMP result | Other pathogens | Original characterization |
|---|---|---|---|---|---|---|---|---|---|---|---|---|
| 1 | MTB | N/A | 2.3 | 26.1 | INC | N/A | 29.2 | N/A | 29.0 | NEG | No | POS |
| 2 | MTB | 18126.0 | 13828.0 | 29.4 | POS | 17.8 | N/A | 17.7 | N/A | POS | Influenza A | POS |
| 3 | MTB | 455.6 | 731.0 | 32.7 | POS | 20.2 | N/A | 20.2 | N/A | POS | No | POS |
| 4 | MTB | 98480.0 | 104100.0 | 33.0 | POS | 16.1 | N/A | 16.2 | N/A | POS | No | POS |
| 5 | MTB | 939.6 | 831.2 | 31.8 | POS | 19.8 | N/A | 19.9 | N/A | POS | Adenovirus | POS |
| 6 | MTB | N/A | N/A | 28.9 | NEG | N/A | 28.2 | N/A | 28.0 | NEG | Streptococcus pneumoniae | NEG |

| | | | | | | | | | | | | |
|---|---|---|---|---|---|---|---|---|---|---|---|---|
| 7 | MTB | N/A | N/A | 30.8 | NEG | N/A | 28.1 | N/A | 28.2 | NEG | Streptococcus pneumoniae | NEG |
| 8 | MTB | N/A | N/A | 28.7 | NEG | N/A | 28.8 | N/A | 28.3 | NEG | Streptococcus pneumoniae | NEG |
| 9 | MTB | N/A | N/A | N/A | IND | N/A | 28.5 | N/A | 28.0 | NEG | No | NEG |
| 10 | MTB | N/A | N/A | 26.9 | NEG | N/A | 28.2 | N/A | 28.4 | NEG | Human parechovirus | NEG |
| 11 | MTB | N/A | N/A | N/A | IND | N/A | 27.9 | N/A | 28.4 | NEG | Rhinovirus, Influenza B, Adenovirus | NEG |
| 12 | MTB | N/A | N/A | 33.7 | NEG | N/A | 28.4 | N/A | 29.4 | NEG | Rhinovirus, Influenza B | NEG |
| 13 | MTB | 2838.0 | 1972.2 | 24.1 | POS | 19.3 | N/A | 19.2 | N/A | POS | Streptococcus pneumoniae | POS |
| 14 | MTB | N/A | N/A | 30.6 | NEG | N/A | 28.5 | N/A | 28.2 | NEG | No | POS |
| 15 | MTB | 6874.0 | 4214.0 | 27.0 | POS | 18.8 | N/A | 18.7 | N/A | POS | No | POS |
| 17 | MTB | 48.0 | 56.7 | 31.1 | POS | 22.8 | N/A | 23.1 | N/A | POS | No | POS |
| 18 | MTB | N/A | N/A | 30.2 | NEG | N/A | 28.0 | N/A | 28.6 | NEG | Rhinovirus | NEG |
| 19 | MTB | N/A | N/A | N/A | IND | N/A | 28.5 | N/A | 28.9 | NEG | Human coronavirus, Influenza A | NEG |
| 20 | MTB | N/A | N/A | 25.9 | NEG | N/A | 28.6 | N/A | 28.2 | NEG | Streptococcus pneumoniae, Rhinovirus | NEG |
| 21 | MTB | N/A | N/A | 33.6 | NEG | N/A | 28.7 | N/A | 28.5 | NEG | Streptococcus pneumoniae, Adenovirus | NEG |
| 22 | MTB | N/A | N/A | 30.9 | NEG | N/A | 28.9 | N/A | 29.3 | NEG | Human bocavirus | NEG |
| 23 | MTB | N/A | N/A | 34.3 | NEG | N/A | 28.9 | N/A | 28.9 | NEG | Rhinovirus, Influenza B | NEG |
| 24 | MTB | N/A | N/A | 34.7 | NEG | N/A | 29.8 | N/A | 28.5 | NEG | Rhinovirus | NEG |
| 25 | MTB | 11.2 | 5.1 | 27.1 | POS | 28.1 | 30.2 | 27.7 | 30.3 | POS | Adenovirus | POS |
| 26 | MTB | 19.6 | 11.2 | 24.0 | POS | 23.6 | N/A | 22.8 | N/A | POS | No | POS |
| 27 | MTB | 0.1 | 0.7 | 26.3 | POS | N/A | 29.3 | N/A | 28.7 | NEG | No | POS |
| 29 | MTB | 36.8 | 19.6 | 26.6 | POS | N/A | 29.3 | 24.6 | N/A | Discordant | No | POS |
| 30 | MTB | N/A | N/A | 28.8 | NEG | N/A | 28.9 | N/A | 28.4 | NEG | Rhinovirus, Streptococcus pneumoniae | NEG |
| 31 | MTB | N/A | N/A | 27.3 | NEG | N/A | 28.0 | N/A | 28.6 | NEG | Influenza A, Streptococcus pneumoniae | NEG |
| 32 | MTB | N/A | N/A | 23.4 | NEG | N/A | 28.4 | N/A | 28.7 | NEG | Streptococcus pneumoniae | NEG |
| 33 | MTB | N/A | N/A | 37.0 | NEG | N/A | 29.1 | N/A | 28.6 | NEG | Streptococcus pneumoniae | NEG |
| 34 | MTB | N/A | N/A | 31.0 | NEG | N/A | 28.6 | N/A | 29.3 | NEG | Human bocavirus | NEG |
| 35 | MTB | N/A | N/A | 35.2 | NEG | N/A | 28.6 | N/A | 28.3 | NEG | Rhinovirus, Influenza B | NEG |
| 36 | MTB | N/A | N/A | 29.4 | NEG | N/A | 29.5 | N/A | 28.8 | NEG | Streptococcus pneumoniae, Rhinovirus, Influenza B | NEG |
| 37 | MTB | 17.2 | 11.1 | 28.1 | POS | 23.5 | N/A | 23.9 | N/A | POS | Streptococcus pneumoniae | POS |
| 38 | MTB | 219800.0 | 164700.0 | 28.8 | POS | 15.2 | N/A | 15.4 | N/A | POS | Rhinovirus | POS |
| 39 | MTB | 15.2 | 2.7 | 28.6 | POS | 24.5 | N/A | N/A | 28.9 | Discordant | Rhinovirus | POS |
| 41 | MTB | 75100.0 | 72740.0 | 29.8 | POS | 16.5 | N/A | 16.5 | N/A | POS | Streptococcus pneumoniae | NEG |

| | | | | | | | | | | | | |
|---|---|---|---|---|---|---|---|---|---|---|---|---|
| 42 | MTB | N/A | N/A | 26.2 | NEG | N/A | 29.0 | N/A | 28.3 | NEG | Influenza A, Streptococcus pneumoniae | POS |
| 43 | MTB | N/A | N/A | 31.5 | NEG | N/A | 28.1 | N/A | 28.8 | NEG | Rhinovirus, Streptococcus pneumoniae | NEG |
| 44 | MTB | N/A | N/A | 28.8 | NEG | N/A | 29.0 | N/A | 29.0 | NEG | Streptococcus pneumoniae | NEG |
| 45 | MTB | N/A | N/A | 32.1 | NEG | N/A | 29.2 | N/A | 29.4 | NEG | Streptococcus pneumoniae, Rhinovirus, Enterovirus D68 | NEG |
| 46 | MTB | N/A | N/A | N/A | IND | N/A | 28.4 | N/A | 28.5 | NEG | Human parainfluenza | NEG |
| 47 | MTB | N/A | N/A | N/A | IND | N/A | 28.8 | N/A | 27.8 | NEG | Influenza B | NEG |
| 48 | MTB | N/A | N/A | N/A | IND | N/A | 29.0 | N/A | 28.3 | NEG | Influenza B | NEG |
| 49 | MTB | 10718.0 | 6840.0 | 34.7 | POS | 18.7 | N/A | 18.4 | N/A | POS | No | NEG |
| 50 | MTB | 10.7 | 14.7 | 29.2 | POS | 23.9 | N/A | 26.8 | 28.9 | POS | No | NEG |
| 51 | MTB | 1742.6 | 1763.6 | 30.5 | POS | 19.8 | N/A | 19.9 | N/A | POS | No | POS |
| 53 | MTB | 0.2 | N/A | 31.4 | INC | N/A | 29.0 | N/A | 28.6 | NEG | No | POS |
| 54 | MTB | N/A | N/A | 33.4 | NEG | N/A | 28.3 | N/A | 28.4 | NEG | Adenovirus, Human coronavirus, Streptococcus pneumoniae | NEG |
| 55 | MTB | N/A | N/A | 34.0 | NEG | N/A | 28.7 | N/A | 27.9 | NEG | Influenza A, Rhinovirus, Streptococcus pneumoniae | NEG |
| 56 | MTB | N/A | N/A | 23.6 | NEG | N/A | 29.0 | N/A | 28.9 | NEG | Streptococcus pneumoniae | NEG |
| 57 | MTB | N/A | N/A | 38.8 | NEG | N/A | 29.8 | N/A | 28.9 | NEG | Streptococcus pneumoniae | NEG |
| 58 | MTB | N/A | N/A | 29.9 | NEG | N/A | 28.4 | N/A | 28.0 | NEG | Streptococcus pneumoniae | POS |
| 59 | MTB | N/A | N/A | 29.7 | NEG | N/A | 29.0 | N/A | 28.1 | NEG | Rhinovirus, Influenza B | NEG |
| 60 | MTB | N/A | N/A | 31.9 | NEG | N/A | 28.6 | N/A | 28.3 | NEG | No | NEG |
| 61 | MTB | 619.2 | 440.2 | 30.0 | POS | 20.1 | N/A | 20.4 | N/A | POS | No | unk |
| 62 | MTB | 3680.0 | 3550.0 | 30.1 | POS | 18.6 | N/A | 18.5 | N/A | POS | No | POS |
| 63 | NP | 1.1 | 1.2 | 24.5 | POS | N/A | 28.5 | N/A | 28.3 | NEG | No | POS |
| 65 | MTB | 125.8 | 90.0 | 25.5 | POS | 20.8 | N/A | 21.3 | N/A | POS | No | POS |
| 67 | MTB | N/A | N/A | 30.7 | NEG | N/A | 28.0 | N/A | 28.0 | NEG | Human metapneumovirus | NEG |
| 68 | MTB | N/A | N/A | 25.3 | NEG | N/A | 28.2 | N/A | 27.6 | NEG | Rhinovirus, Streptococcus pneumoniae | NEG |
| 69 | MTB | N/A | N/A | 34.0 | NEG | N/A | 29.3 | N/A | 28.3 | NEG | Streptococcus pneumoniae | NEG |
| 70 | MTB | N/A | N/A | 28.2 | NEG | N/A | 28.9 | N/A | 27.8 | NEG | Human parainfluenza | NEG |
| 71 | MTB | N/A | N/A | N/A | IND | N/A | 29.4 | N/A | 28.4 | NEG | Rhinovirus | NEG |
| 72 | MTB | N/A | N/A | 31.3 | NEG | N/A | 29.5 | N/A | 29.6 | NEG | No | NEG |
| 73 | MTB | 2228.0 | 1791.0 | 26.9 | POS | 19.4 | N/A | 19.4 | N/A | POS | No | POS |
| 74 | MTB | 0.2 | N/A | 30.0 | INC | N/A | 29.6 | N/A | 29.8 | NEG | Rhinovirus | POS |
| 75 | MTB | 4.0 | 4.6 | 25.4 | POS | 25.7 | N/A | N/A | 29.1 | Discordant | No | POS |

| | | | | | | | | | | | | |
|---|---|---|---|---|---|---|---|---|---|---|---|---|
| 77 | MTB | 53220.0 | 38920.0 | 23.3 | POS | 16.9 | N/A | 16.8 | N/A | POS | No | POS |
| 78 | MTB | N/A | N/A | 30.6 | NEG | N/A | 28.3 | N/A | 28.1 | NEG | Human coronavius | NEG |
| 79 | MTB | N/A | N/A | 28.8 | NEG | N/A | 28.7 | N/A | 28.1 | NEG | Rhinovirus, Streptococcus pneumoniae | NEG |
| 80 | MTB | N/A | N/A | 26.0 | NEG | N/A | 29.3 | N/A | 28.6 | NEG | Streptococcus pneumoniae, Human coronavirus, Human bocavirus, Adenovirus | NEG |
| 81 | MTB | N/A | N/A | 26.9 | NEG | N/A | 28.7 | N/A | 28.3 | NEG | Human coronavirus | NEG |
| 82 | MTB | N/A | N/A | 29.5 | NEG | N/A | 28.6 | N/A | 28.6 | NEG | Streptococcus pneumoniae | NEG |
| 83 | MTB | N/A | N/A | 39.1 | NEG | N/A | 29.5 | N/A | 29.3 | NEG | Rhinovirus, Influenza B | NEG |
| 84 | MTB | N/A | N/A | 30.1 | NEG | N/A | 29.0 | N/A | 29.4 | NEG | Rhinovirus | NEG |
| 85 | MTB | 733.0 | 407.0 | 27.9 | POS | 20.5 | N/A | 20.8 | N/A | POS | Enterovirus, Rhinovirus | POS |
| 86 | MTB | 60.3 | 37.6 | 33.7 | POS | 21.6 | N/A | 22.8 | N/A | POS | No | POS |
| 87 | NP | 62480.0 | 42300.0 | 28.6 | POS | 16.7 | N/A | 16.8 | N/A | POS | No | POS |
| 88 | MTB | 2178.0 | 1160.4 | 27.4 | POS | 19.9 | N/A | 20.0 | N/A | POS | No | POS |
| 89 | MTB | 364.0 | 220.2 | 27.4 | POS | 21.2 | N/A | 20.9 | N/A | POS | No | POS |
| 90 | MTB | N/A | N/A | 30.1 | NEG | N/A | 28.5 | N/A | 29.1 | NEG | Human coronavirus, Rhinovirus | NEG |
| 91 | MTB | N/A | N/A | 29.1 | NEG | N/A | 29.1 | N/A | 28.6 | NEG | Streptococcu pneumoniae, Rhinovirus | NEG |
| 92 | MTB | N/A | N/A | 27.2 | NEG | N/A | 29.4 | N/A | 29.7 | NEG | No | NEG |
| 93 | MTB | N/A | N/A | N/A | IND | N/A | 29.4 | N/A | 29.3 | NEG | Human bocavirus | NEG |
| 94 | MTB | N/A | N/A | 30.6 | NEG | N/A | 28.8 | N/A | 28.2 | NEG | Rhinovirus | NEG |
| 95 | MTB | N/A | N/A | 32.5 | NEG | N/A | 29.2 | N/A | 29.4 | NEG | Influenza B | NEG |
| 96 | MTB | N/A | N/A | 31.0 | NEG | N/A | 29.1 | N/A | 30.0 | NEG | No | NEG |
| 97 | MTB | N/A | N/A | 40.4 | NEG | N/A | 29.0 | N/A | 29.3 | NEG | No | NEG |
| 98 | MTB | N/A | N/A | 32.8 | NEG | N/A | 29.3 | N/A | 29.6 | NEG | Streptococcus pneumoniae | NEG |
| 99 | MTB | N/A | N/A | N/A | IND | N/A | 27.7 | N/A | 28.7 | NEG | No | NEG |
| 100 | MTB | N/A | N/A | 34.2 | NEG | N/A | 27.5 | N/A | 29.1 | NEG | Rhinovirus | NEG |
| 101 | MTB | N/A | N/A | 36.7 | NEG | N/A | 28.6 | N/A | 28.6 | NEG | No | NEG |
| 102 | MTB | N/A | N/A | 31.0 | NEG | N/A | 29.0 | N/A | 28.9 | NEG | Rhinovirus, Influenza B | NEG |
| 103 | MTB | N/A | N/A | 29.3 | NEG | N/A | 28.8 | N/A | 29.3 | NEG | Streptococcus pneumoniae | NEG |
| 104 | MTB | N/A | N/A | 36.8 | NEG | N/A | 29.2 | N/A | 29.7 | NEG | Rhinovirus | NEG |
| 105 | MTB | N/A | N/A | 33.4 | NEG | N/A | 28.3 | N/A | 28.6 | NEG | No | NEG |
| 106 | MTB | N/A | N/A | 43.3 | NEG | N/A | 29.0 | N/A | 28.7 | NEG | No | NEG |
| 107 | MTB | N/A | N/A | 47.1 | NEG | N/A | 28.8 | N/A | 29.1 | NEG | No | NEG |
| 108 | MTB | N/A | N/A | 32.1 | NEG | N/A | 29.1 | N/A | 28.8 | NEG | No | NEG |
| 109 | MTB | N/A | N/A | 28.4 | NEG | N/A | 29.0 | N/A | 28.8 | NEG | No | NEG |

### supp. Table S2: Alpha

We gratefully acknowledge the following Authors from the Originating laboratories responsible for obtaining the specimens, as well as the Submitting laboratories where the genome data were generated and shared via GISAID, on which this research is based.

All Submitters of data may be contacted directly via www.gisaid.org

Authors are sorted alphabetically.

| Accession ID | Originating Laboratory | Submitting Laboratory | Authors |
|---|---|---|---|
| EPI_ISL_2565995 | Genome Analysis Center, Yamanashi Central Hospital | Genome Analysis Center, Yamanashi Central Hospital | Yosuke Hirotsu |
| EPI_ISL_2597900 | WSSE w Gdasku | 1. Tricity SARS-CoV-2 sequencing consortium: University of Gdansk, Medical University of Gdansk, Vaxican Ltd., Invicta Ltd. 2. National Institute of Public Health - National Institute of Hygiene, Warsaw, Poland | Maciej Kosinski, Celina Cybulska, Krystyna Bienkowska Szewczyk, Maciej Grzybek, Karolina Gackowska, Marcin Lubocki, Katarzyna Groth, Lukasz Rabalski, Katarzyna Zacharczuk, Magdalena Nowakowska, Magorzata Sadkowska-Todys, Tomasz Wokowicz |
| EPI_ISL_2597930 | Nebraska Public Health Laboratory | NPHL COVID-19 Response Team | NPHL COVID-19 Response Team |
| EPI_ISL_2601940, EPI_ISL_2601941, EPI_ISL_2601942, EPI_ISL_2601945, EPI_ISL_2601947, EPI_ISL_2601948, EPI_ISL_2601949 | ZOL | Jessa | Berden et al. on behalf of the Jessa_cmdLab |
| EPI_ISL_2604225, EPI_ISL_2604226, EPI_ISL_2612461, EPI_ISL_2612466 | Montana Public Health Laboratory | Montana Public Health Laboratory | Joy Ritter, Michelle Mozer, Carrie Biskupiak, Deborah Gibson, Michael Dills |
| EPI_ISL_2617988, EPI_ISL_2617993, EPI_ISL_2617996, EPI_ISL_2617999, EPI_ISL_2618000, EPI_ISL_2618006, EPI_ISL_2618008, EPI_ISL_2618014, EPI_ISL_2618015 | UW Virology Lab | UW Virology Lab | Pavitra Roychoudhury, Hong Xie, Lasata Shrestha, Tien V. Nguyen, Shah Mohamed Bakhash, Michelle Lin, Noah R. Baker, Ricardo Perez, Sean Ellis, Nathan Breit, Robert J. Livingston, Meei-Li Huang, Keith R Jerome, Patrick Mathias, Alexander Greninger |
| EPI_ISL_2627922 | Kansas Health and Environmental Lab | Kansas Health and Environmental Lab | Mike Grose, Katherine Wiggins, Jonathan Barnell, Ben Olsen, and Phil Adam |
| EPI_ISL_2628054 | LHUB-ULB | Labo Klinische Biologie, UZA | Marie Le Mercier, Jasmine Coppens, Basil Britto Xavier, Christine Lammens, Veerle Matheeussen, Herman Goossens |
| EPI_ISL_2628076, EPI_ISL_2628077, EPI_ISL_2628078, EPI_ISL_2628079, EPI_ISL_2628080, EPI_ISL_2628081, EPI_ISL_2628082, EPI_ISL_2628083, EPI_ISL_2628084, EPI_ISL_2628085, EPI_ISL_2628087, EPI_ISL_2628089, EPI_ISL_2628090, EPI_ISL_2628091, EPI_ISL_2628092, EPI_ISL_2628093, EPI_ISL_2628094, EPI_ISL_2628095, EPI_ISL_2628096, EPI_ISL_2628098, EPI_ISL_2628101, EPI_ISL_2628102, EPI_ISL_2628104 | | | |
| see above | Platform BIS UZA/UAntwerpen | Labo Klinische Biologie, UZA | Marie Le Mercier, Jasmine Coppens, Basil Britto Xavier, Christine Lammens, Veerle Matheeussen, Herman Goossens |
| EPI_ISL_2635603, EPI_ISL_2635613, EPI_ISL_2635615, EPI_ISL_2635624, EPI_ISL_2635633, EPI_ISL_2635639, EPI_ISL_2635647 | nordlab - Partnerschaftspraxis fur Laboratoriumsmedizin | Robert Koch Institute | unknown |
| EPI_ISL_2635653 | amedes MVZ für Labordiagnostik Rhein-Main | Robert Koch Institute | unknown |
| EPI_ISL_2635663, EPI_ISL_2635672 | Limbach - MVZ Humangenetik Ulm | Robert Koch Institute | unknown |
| EPI_ISL_2635866, EPI_ISL_2635878 | Limbach - MVZ Labor Eveld & Kollegen Essen | Robert Koch Institute | unknown |
| EPI_ISL_2637851 | GH de l'Est Francilien | Department of Virology, Henri Mondor University Hospital, Assistance Publique Hôpitaux de Paris, Université Paris-Est Créteil, INSERM U955 | Christophe Rodriguez, Slim Fourati, Vanessa Demontant, Guillaume Gricourt, Melissa N'Debi, Alexandre Soulier, Elisabeth Trawinski, Jean-Michel Pawlotsky |
| EPI_ISL_2637903, EPI_ISL_2637951 | CH.INTERCOMMUNAL DE CRETEIL | Department of Virology, Henri Mondor University Hospital, Assistance Publique Hôpitaux de Paris, Université Paris-Est Créteil, INSERM U955 | Christophe Rodriguez, Slim Fourati, Vanessa Demontant, Guillaume Gricourt, Melissa N'Debi, Alexandre Soulier, Elisabeth Trawinski, Jean-Michel Pawlotsky |
| EPI_ISL_2637996 | GH de l'Est Francilien | Department of Virology, Henri Mondor University Hospital, Assistance Publique Hôpitaux de Paris, Université Paris-Est Créteil, INSERM U955 | Christophe Rodriguez, Slim Fourati, Vanessa Demontant, Guillaume Gricourt, Melissa N'Debi, Alexandre Soulier, Elisabeth Trawinski, Jean-Michel Pawlotsky |
| EPI_ISL_2638660 | Lighthouse Lab in Milton Keynes | Wellcome Sanger Institute for the COVID-19 Genomics UK (COG-UK) Consortium | The Lighthouse Lab in Milton Keynes and Alex Alderton, Roberto Amato, Jeffrey Barrett, Sonia Goncalves, Ewan Harrison, David K. Jackson, Ian Johnston, Dominic Kwiatkowski, Cordelia Langford, John Sillitoe on behalf of the Wellcome Sanger Institute COVID-19 Surveillance Team |
| EPI_ISL_2638853 | Lighthouse Lab in Glasgow | Wellcome Sanger Institute for the COVID-19 Genomics UK (COG-UK) Consortium | Harper VanSteenhouse, Yumi Kasai, David Gray, Carol Clugston, Anna Dominiczak and Alex Alderton, Roberto Amato, Jeffrey Barrett, Sonia Goncalves, Ewan Harrison, David K. Jackson, Ian Johnston, Dominic Kwiatkowski, Cordelia Langford, John Sillitoe on behalf of the Wellcome Sanger Institute COVID-19 Surveillance Team |
| EPI_ISL_2638871 | Lighthouse Lab in Milton Keynes | Wellcome Sanger Institute for the COVID-19 Genomics UK (COG-UK) Consortium | The Lighthouse Lab in Milton Keynes and Alex Alderton, Roberto Amato, Jeffrey Barrett, Sonia Goncalves, Ewan Harrison, David K. Jackson, Ian Johnston, Dominic Kwiatkowski, Cordelia Langford, John Sillitoe on behalf of the Wellcome Sanger Institute COVID-19 Surveillance Team |
| EPI_ISL_2639029 | Lighthouse Laboratory Plymouth | Wellcome Sanger Institute for the COVID-19 Genomics UK (COG-UK) Consortium | Lighthouse Laboratory Plymouth and Alex Alderton, Roberto Amato, Jeffrey Barrett, Sonia Goncalves, Ewan Harrison, David K. Jackson, Ian Johnston, Dominic Kwiatkowski, Cordelia Langford, John Sillitoe on behalf of the Wellcome Sanger Institute COVID-19 Surveillance Team |
| EPI_ISL_2639092 | Lighthouse Lab in Milton Keynes | Wellcome Sanger Institute for the COVID-19 Genomics UK (COG-UK) Consortium | The Lighthouse Lab in Milton Keynes and Alex Alderton, Roberto Amato, Jeffrey Barrett, Sonia Goncalves, Ewan Harrison, David K. Jackson, Ian Johnston, Dominic Kwiatkowski, Cordelia Langford, John Sillitoe on behalf of the Wellcome Sanger Institute COVID-19 Surveillance Team |
| EPI_ISL_2639330, EPI_ISL_2639334, EPI_ISL_2639348, EPI_ISL_2639367, EPI_ISL_2639584, EPI_ISL_2639650, EPI_ISL_2639709, EPI_ISL_2639865 | Lighthouse Lab in Alderley Park | Wellcome Sanger Institute for the COVID-19 Genomics UK (COG-UK) Consortium | Jacquelyn Wynn, Mairead Hyland, The Lighthouse Lab in Alderley Park and Alex Alderton, Roberto Amato, Jeffrey Barrett, Sonia Goncalves, Ewan Harrison, David K. Jackson, Ian Johnston, Dominic Kwiatkowski, Cordelia Langford, John Sillitoe on behalf of the Wellcome Sanger Institute COVID-19 Surveillance Team |
| EPI_ISL_2641978, EPI_ISL_2641980, EPI_ISL_2641985, EPI_ISL_2641986, EPI_ISL_2641987, EPI_ISL_2641988, EPI_ISL_2641989, EPI_ISL_2642023, EPI_ISL_2642024, EPI_ISL_2642025, EPI_ISL_2642026, EPI_ISL_2642027, EPI_ISL_2642034, EPI_ISL_2642035, EPI_ISL_2642037, EPI_ISL_2642038, EPI_ISL_2642039, EPI_ISL_2642041 | | | |
| see above | Istituto Zooprofilattico Sperimentale del Mezzogiorno | Telethon Institute of Genetics and Medicine (TIGEM) | Antonio Grimaldi Patrizia Annunziata Francesco Panariello Biancamaria Pierri Claudia Tiberio Teresa Giuliano Valentina Bouche Chiara Colantuono Maria Concetta Cuomo Denise Di Concilio Lucio Di Filippo Anna Manfredi Marcello Salvi Antonio Limone Luigi Atripaldi Pellegrino Cerino Andrea Ballabio Davide Cacchiarelli |
| EPI_ISL_2650882, EPI_ISL_2650884, EPI_ISL_2650885, EPI_ISL_2650886 | ASL TO4 | Fondazione del Piemonte per l'Oncologia IRCCS | Antonino Sottile, Silvia Brossa, Paola Marino, Giorgio Giardina |
| EPI_ISL_2650888 | ASL CN1 | Fondazione del Piemonte per l'Oncologia IRCCS | Antonino Sottile, Silvia Brossa, Paola Marino, Giorgio Giardina |
| EPI_ISL_2650890 | ASL TO4 | Fondazione del Piemonte per l'Oncologia IRCCS | Antonino Sottile, Silvia Brossa, Paola Marino, Giorgio Giardina |

| | | | |
|---|---|---|---|
| EPI_ISL_2651107 | Washington State Department of Health Public Health Laboratories | Washington State Department of Health Public Health Laboratories | Drew MacKellar, Philip Dykema, Denny Russell, Joenice Gonzalez, Hannah Gray, Geoff Melly, Vanessa De Los Santos, Darren Lucas, JohnAric Peterson, Avi Singh, Rebecca Cao |
| EPI_ISL_2651786, EPI_ISL_2651896 | Lighthouse Lab in Glasgow | Wellcome Sanger Institute for the COVID-19 Genomics UK (COG-UK) Consortium | Harper VanSteenhouse, Yumi Kasai, David Gray, Carol Clugston, Anna Dominiczak and Alex Alderton, Roberto Amato, Jeffrey Barrett, Sonia Goncalves, Ewan Harrison, David K. Jackson, Ian Johnston, Dominic Kwiatkowski, Cordelia Langford, John Sillitoe on behalf of the Wellcome Sanger Institute COVID-19 Surveillance Team |
| EPI_ISL_2661534 | UHTL, University Hospitals | UHTL, University Hospitals | Sadri,N., Alouani,D., Song,X. |
| EPI_ISL_2663213, EPI_ISL_2663217 | HealthPartners Central Lab | Minnesota Department of Health, Public Health Laboratory | Alexandra Lorentz, Jacob Garfin, Matt Plumb, and Xiong Wang |
| EPI_ISL_2664038 | Lighthouse Lab in Alderley Park | Wellcome Sanger Institute for the COVID-19 Genomics UK (COG-UK) Consortium | Jacquelyn Wynn, Mairead Hyland, The Lighthouse Lab in Alderley Park and Alex Alderton, Roberto Amato, Jeffrey Barrett, Sonia Goncalves, Ewan Harrison, David K. Jackson, Ian Johnston, Dominic Kwiatkowski, Cordelia Langford, John Sillitoe on behalf of the Wellcome Sanger Institute COVID-19 Surveillance Team |
| EPI_ISL_2664937, EPI_ISL_2665215 | Lighthouse Lab in Milton Keynes | Wellcome Sanger Institute for the COVID-19 Genomics UK (COG-UK) Consortium | The Lighthouse Lab in Milton Keynes and Alex Alderton, Roberto Amato, Jeffrey Barrett, Sonia Goncalves, Ewan Harrison, David K. Jackson, Ian Johnston, Dominic Kwiatkowski, Cordelia Langford, John Sillitoe on behalf of the Wellcome Sanger Institute COVID-19 Surveillance Team |
| EPI_ISL_2665417, EPI_ISL_2665432, EPI_ISL_2665492, EPI_ISL_2665506, EPI_ISL_2665508, EPI_ISL_2665510, EPI_ISL_2665511, EPI_ISL_2665521, EPI_ISL_2665522, EPI_ISL_2665523, EPI_ISL_2665532, EPI_ISL_2665535, EPI_ISL_2665547, EPI_ISL_2665573, EPI_ISL_2665575, EPI_ISL_2665583, EPI_ISL_2665638, EPI_ISL_2665643, EPI_ISL_2665644, EPI_ISL_2665660, EPI_ISL_2665669, EPI_ISL_2665673, EPI_ISL_2665695, EPI_ISL_2665696, EPI_ISL_2665697, EPI_ISL_2665741, EPI_ISL_2665746, EPI_ISL_2665748, EPI_ISL_2665757, EPI_ISL_2665768, EPI_ISL_2665770, EPI_ISL_2665774, EPI_ISL_2665776, EPI_ISL_2665784, EPI_ISL_2665804, EPI_ISL_2665819, EPI_ISL_2665831, EPI_ISL_2665841, EPI_ISL_2665852, EPI_ISL_2665874, EPI_ISL_2665896, EPI_ISL_2665898, EPI_ISL_2665900, EPI_ISL_2665909, EPI_ISL_2665912, EPI_ISL_2665941, EPI_ISL_2665947, EPI_ISL_2665954, EPI_ISL_2665955, EPI_ISL_2665968, EPI_ISL_2665980, EPI_ISL_2665985, EPI_ISL_2666007, EPI_ISL_2666013, EPI_ISL_2666045, EPI_ISL_2666046, EPI_ISL_2666047, EPI_ISL_2666059, EPI_ISL_2666067, EPI_ISL_2666068, EPI_ISL_2666084, EPI_ISL_2666090, EPI_ISL_2666113, EPI_ISL_2666127, EPI_ISL_2666134, EPI_ISL_2666135, EPI_ISL_2666140, EPI_ISL_2666158, EPI_ISL_2666171, EPI_ISL_2666191, EPI_ISL_2666199, EPI_ISL_2666204, EPI_ISL_2666217, EPI_ISL_2666227, EPI_ISL_2666240, EPI_ISL_2666248, EPI_ISL_2666250, EPI_ISL_2666252, EPI_ISL_2666271, EPI_ISL_2666276, EPI_ISL_2666289, EPI_ISL_2666296, EPI_ISL_2666306, EPI_ISL_2666327, EPI_ISL_2666351, EPI_ISL_2666353, EPI_ISL_2666361, EPI_ISL_2666363, EPI_ISL_2666364, EPI_ISL_2666370, EPI_ISL_2666377, EPI_ISL_2666379, EPI_ISL_2666389, EPI_ISL_2666423, EPI_ISL_2666429, EPI_ISL_2666514, EPI_ISL_2666526, EPI_ISL_2666527, EPI_ISL_2666541, EPI_ISL_2666543, EPI_ISL_2666546, EPI_ISL_2666549, EPI_ISL_2666565, EPI_ISL_2666566, EPI_ISL_2666583 | | | |
| see above | Department of Bacteria, Parasites and Fungi, Statens Serum Institut, Copenhagen, Denmark | Statens Serum Institut Bioinformatics and Microbial Genomics | Danish Covid-19 Genome Consortium |
| EPI_ISL_2666602 | Molekylær Medicinsk Afdeling, Aarhus University Hospital, Aarhus, Denmark | Statens Serum Institut Bioinformatics and Microbial Genomics | Danish Covid-19 Genome Consortium |
| EPI_ISL_2666607, EPI_ISL_2666621, EPI_ISL_2666622, EPI_ISL_2666671, EPI_ISL_2666703, EPI_ISL_2666757, EPI_ISL_2666766, EPI_ISL_2666776, EPI_ISL_2666793, EPI_ISL_2666801, EPI_ISL_2666806, EPI_ISL_2666851, EPI_ISL_2666857, EPI_ISL_2666862, EPI_ISL_2666878, EPI_ISL_2666904, EPI_ISL_2666923, EPI_ISL_2666929, EPI_ISL_2666932, EPI_ISL_2666933 | | | |
| see above | Department of Bacteria, Parasites and Fungi, Statens Serum Institut, Copenhagen, Denmark | Statens Serum Institut Bioinformatics and Microbial Genomics | Danish Covid-19 Genome Consortium |
| EPI_ISL_2666939 | Molekylær Medicinsk Afdeling, Aarhus University Hospital, Aarhus, Denmark | Statens Serum Institut Bioinformatics and Microbial Genomics | Danish Covid-19 Genome Consortium |
| EPI_ISL_2666960, EPI_ISL_2666990, EPI_ISL_2666998, EPI_ISL_2666999, EPI_ISL_2667002 | Department of Bacteria, Parasites and Fungi, Statens Serum Institut, Copenhagen, Denmark | Statens Serum Institut Bioinformatics and Microbial Genomics | Danish Covid-19 Genome Consortium |
| EPI_ISL_2667025 | Molekylær Medicinsk Afdeling, Aarhus University Hospital, Aarhus, Denmark | Statens Serum Institut Bioinformatics and Microbial Genomics | Danish Covid-19 Genome Consortium |
| EPI_ISL_2667053, EPI_ISL_2667056, EPI_ISL_2667061, EPI_ISL_2667064, EPI_ISL_2667072, EPI_ISL_2667082, EPI_ISL_2667114, EPI_ISL_2667134, EPI_ISL_2667140, EPI_ISL_2667159, EPI_ISL_2667161, EPI_ISL_2667175, EPI_ISL_2667177, EPI_ISL_2667186, EPI_ISL_2667199, EPI_ISL_2667235, EPI_ISL_2667276, EPI_ISL_2667304, EPI_ISL_2667324, EPI_ISL_2667341, EPI_ISL_2667352, EPI_ISL_2667365, EPI_ISL_2667371, EPI_ISL_2667380, EPI_ISL_2667403, EPI_ISL_2667406, EPI_ISL_2667408, EPI_ISL_2667428, EPI_ISL_2667432, EPI_ISL_2667435, EPI_ISL_2667438, EPI_ISL_2667440, EPI_ISL_2667446, EPI_ISL_2667472, EPI_ISL_2667474, EPI_ISL_2667476, EPI_ISL_2667477, EPI_ISL_2667480, EPI_ISL_2667510, EPI_ISL_2667523, EPI_ISL_2667530, EPI_ISL_2667532, EPI_ISL_2667536, EPI_ISL_2667576, EPI_ISL_2667586, EPI_ISL_2667620, EPI_ISL_2667621, EPI_ISL_2667651, EPI_ISL_2667657, EPI_ISL_2667692, EPI_ISL_2667694, EPI_ISL_2667695, EPI_ISL_2667723, EPI_ISL_2667729, EPI_ISL_2667772, EPI_ISL_2667773, EPI_ISL_2667775, EPI_ISL_2667782, EPI_ISL_2667802, EPI_ISL_2667806, EPI_ISL_2667820, EPI_ISL_2667821, EPI_ISL_2667826, EPI_ISL_2667828, EPI_ISL_2667837, EPI_ISL_2667841, EPI_ISL_2667854, EPI_ISL_2667857, EPI_ISL_2667860, EPI_ISL_2667863, EPI_ISL_2667889, EPI_ISL_2667899, EPI_ISL_2667911, EPI_ISL_2667923, EPI_ISL_2667931, EPI_ISL_2667938, EPI_ISL_2667944, EPI_ISL_2667953, EPI_ISL_2667957, EPI_ISL_2667958, EPI_ISL_2668037, EPI_ISL_2668040, EPI_ISL_2668046, EPI_ISL_2668052, EPI_ISL_2668068, EPI_ISL_2668083, EPI_ISL_2668084, EPI_ISL_2668085, EPI_ISL_2668092, EPI_ISL_2668093, EPI_ISL_2668109, EPI_ISL_2668119, EPI_ISL_2668120, EPI_ISL_2668126, EPI_ISL_2668144, EPI_ISL_2668149, EPI_ISL_2668179, EPI_ISL_2668185, EPI_ISL_2668196, EPI_ISL_2668199, EPI_ISL_2668215, EPI_ISL_2668216, EPI_ISL_2668236, EPI_ISL_2668242, EPI_ISL_2668249, EPI_ISL_2668258, EPI_ISL_2668265, EPI_ISL_2668287, EPI_ISL_2668299, EPI_ISL_2668301, EPI_ISL_2668308, EPI_ISL_2668329, EPI_ISL_2668336, EPI_ISL_2668338, EPI_ISL_2668341, EPI_ISL_2668358, EPI_ISL_2668379, EPI_ISL_2668380, EPI_ISL_2668393, EPI_ISL_2668394, EPI_ISL_2668411, EPI_ISL_2668426, EPI_ISL_2668437, EPI_ISL_2668442, EPI_ISL_2668443 | | | |
| see above | Department of Bacteria, Parasites and Fungi, Statens Serum Institut, Copenhagen, Denmark | Statens Serum Institut Bioinformatics and Microbial Genomics | Danish Covid-19 Genome Consortium |
| EPI_ISL_2668447 | Molekylær Medicinsk Afdeling, Aarhus University Hospital, Aarhus, Denmark | Statens Serum Institut Bioinformatics and Microbial Genomics | Danish Covid-19 Genome Consortium |
| EPI_ISL_2668450, EPI_ISL_2668463, EPI_ISL_2668478, EPI_ISL_2668498, EPI_ISL_2668508, EPI_ISL_2668537, EPI_ISL_2668544, EPI_ISL_2668545, EPI_ISL_2668549, EPI_ISL_2668565, EPI_ISL_2668566, EPI_ISL_2668578, EPI_ISL_2668582, EPI_ISL_2668591, EPI_ISL_2668597, EPI_ISL_2668600, EPI_ISL_2668618, EPI_ISL_2668627, EPI_ISL_2668641, EPI_ISL_2668644, EPI_ISL_2668665, EPI_ISL_2668680, EPI_ISL_2668681, EPI_ISL_2668704, EPI_ISL_2668716, EPI_ISL_2668726, EPI_ISL_2668742 | | | |
| see above | Department of Bacteria, Parasites and Fungi, Statens Serum Institut, Copenhagen, Denmark | Statens Serum Institut Bioinformatics and Microbial Genomics | Danish Covid-19 Genome Consortium |
| EPI_ISL_2668775 | Molekylær Medicinsk Afdeling, Aarhus University Hospital, Aarhus, Denmark | Statens Serum Institut Bioinformatics and Microbial Genomics | Danish Covid-19 Genome Consortium |
| EPI_ISL_2668797, EPI_ISL_2668803, EPI_ISL_2668811, EPI_ISL_2668815, EPI_ISL_2668818, EPI_ISL_2668852, EPI_ISL_2668890, EPI_ISL_2668906, EPI_ISL_2668924, EPI_ISL_2668929, EPI_ISL_2668937, EPI_ISL_2668961, EPI_ISL_2668988, EPI_ISL_2669001, EPI_ISL_2669021, EPI_ISL_2669039, EPI_ISL_2669040, EPI_ISL_2669046, EPI_ISL_2669056, EPI_ISL_2669059, EPI_ISL_2669092, EPI_ISL_2669100, EPI_ISL_2669106, EPI_ISL_2669124, EPI_ISL_2669128, EPI_ISL_2669132, EPI_ISL_2669134, EPI_ISL_2669157, EPI_ISL_2669158, EPI_ISL_2669166, EPI_ISL_2669193, EPI_ISL_2669200, EPI_ISL_2669233, EPI_ISL_2669244, EPI_ISL_2669250, EPI_ISL_2669257, EPI_ISL_2669259, EPI_ISL_2669274, EPI_ISL_2669275, EPI_ISL_2669276, EPI_ISL_2669300, EPI_ISL_2669325, EPI_ISL_2669328, EPI_ISL_2669353, EPI_ISL_2669356, EPI_ISL_2669379, EPI_ISL_2669380, EPI_ISL_2669390, EPI_ISL_2669395, EPI_ISL_2669401, EPI_ISL_2669422, EPI_ISL_2669432, EPI_ISL_2669444, EPI_ISL_2669472, EPI_ISL_2669473, EPI_ISL_2669481, EPI_ISL_2669483, EPI_ISL_2669511, EPI_ISL_2669516, EPI_ISL_2669529, EPI_ISL_2669533, EPI_ISL_2669547, EPI_ISL_2669578, EPI_ISL_2669611, EPI_ISL_2669624, EPI_ISL_2669651, EPI_ISL_2669667, EPI_ISL_2669681, EPI_ISL_2669683, EPI_ISL_2669704, EPI_ISL_2669705, EPI_ISL_2669711, EPI_ISL_2669715, EPI_ISL_2669722, EPI_ISL_2669748 | | | |
| see above | Department of Bacteria, Parasites and Fungi, Statens Serum Institut, Copenhagen, Denmark | Statens Serum Institut Bioinformatics and Microbial Genomics | Danish Covid-19 Genome Consortium |
| EPI_ISL_2669755 | Molekylær Medicinsk Afdeling, Aarhus University Hospital, Aarhus, Denmark | Statens Serum Institut Bioinformatics and Microbial Genomics | Danish Covid-19 Genome Consortium |
| EPI_ISL_2669757, EPI_ISL_2669760, EPI_ISL_2669792, EPI_ISL_2669806, EPI_ISL_2669817, EPI_ISL_2669818, EPI_ISL_2669839, EPI_ISL_2669861, EPI_ISL_2669867, EPI_ISL_2669884, EPI_ISL_2669896, EPI_ISL_2669897, EPI_ISL_2669945, EPI_ISL_2669948, EPI_ISL_2669953, EPI_ISL_2669954, EPI_ISL_2669965, EPI_ISL_2669971, EPI_ISL_2669978, EPI_ISL_2669986, EPI_ISL_2670003, EPI_ISL_2670004, EPI_ISL_2670020, EPI_ISL_2670027, EPI_ISL_2670030, EPI_ISL_2670031, EPI_ISL_2670033, EPI_ISL_2670066, EPI_ISL_2670081 | | | |
| see above | Department of Bacteria, Parasites and Fungi, Statens Serum Institut, Copenhagen, Denmark | Statens Serum Institut Bioinformatics and Microbial Genomics | Danish Covid-19 Genome Consortium |
| EPI_ISL_2670089 | Molekylær Medicinsk Afdeling, Aarhus University Hospital, Aarhus, Denmark | Statens Serum Institut Bioinformatics and Microbial Genomics | Danish Covid-19 Genome Consortium |
| EPI_ISL_2670120, EPI_ISL_2670127, EPI_ISL_2670137, EPI_ISL_2670147, EPI_ISL_2670159, EPI_ISL_2670167, EPI_ISL_2670173, EPI_ISL_2670180, EPI_ISL_2670187, EPI_ISL_2670196, EPI_ISL_2670199, EPI_ISL_2670211, EPI_ISL_2670243, EPI_ISL_2670244, EPI_ISL_2670250, EPI_ISL_2670275, EPI_ISL_2670282, EPI_ISL_2670302, EPI_ISL_2670303, EPI_ISL_2670304, EPI_ISL_2670338, EPI_ISL_2670340, EPI_ISL_2670348, EPI_ISL_2670365, EPI_ISL_2670390, EPI_ISL_2670394, EPI_ISL_2670413, EPI_ISL_2670424, EPI_ISL_2670446, EPI_ISL_2670479, EPI_ISL_2670487, EPI_ISL_2670489, EPI_ISL_2670490, EPI_ISL_2670505, EPI_ISL_2670536, EPI_ISL_2670543, EPI_ISL_2670553, EPI_ISL_2670564, EPI_ISL_2670569, EPI_ISL_2670572, EPI_ISL_2670592, EPI_ISL_2670604, EPI_ISL_2670628, EPI_ISL_2670631, EPI_ISL_2670636, EPI_ISL_2670662, EPI_ISL_2670679, EPI_ISL_2670693, EPI_ISL_2670700, EPI_ISL_2670722, EPI_ISL_2670729, EPI_ISL_2670733, EPI_ISL_2670746, EPI_ISL_2670757, EPI_ISL_2670761, EPI_ISL_2670774, EPI_ISL_2670776, EPI_ISL_2670789, EPI_ISL_2670811, EPI_ISL_2670844, EPI_ISL_2670856, EPI_ISL_2670867, EPI_ISL_2670868, EPI_ISL_2670871, EPI_ISL_2670882, EPI_ISL_2670884, EPI_ISL_2670885, EPI_ISL_2670890, EPI_ISL_2670896, EPI_ISL_2670900, EPI_ISL_2670922, EPI_ISL_2670928, EPI_ISL_2670937, EPI_ISL_2670941, EPI_ISL_2670961, EPI_ISL_2670962, EPI_ISL_2670990, EPI_ISL_2671006, EPI_ISL_2671008, EPI_ISL_2671031, EPI_ISL_2671036, EPI_ISL_2671044, EPI_ISL_2671066, EPI_ISL_2671080, EPI_ISL_2671095, EPI_ISL_2671105, EPI_ISL_2671121, EPI_ISL_2671122, EPI_ISL_2671129, EPI_ISL_2671149, EPI_ISL_2671161, EPI_ISL_2671169, EPI_ISL_2671183, EPI_ISL_2671201, EPI_ISL_2671217, EPI_ISL_2671246, EPI_ISL_2671259, EPI_ISL_2671264, EPI_ISL_2671275, EPI_ISL_2671285, EPI_ISL_2671288, EPI_ISL_2671303, EPI_ISL_2671324, EPI_ISL_2671332, EPI_ISL_2671334, EPI_ISL_2671347 | | | |

| | | | |
|---|---|---|---|
| see above | Department of Bacteria, Parasites and Fungi, Statens Serum Institut, Copenhagen, Denmark | Statens Serum Institut Bioinformatics and Microbial Genomics | Danish Covid-19 Genome Consortium |
| EPI_ISL_2671365 | Microbiology Department, Laboratori Clínic Metropolitana Nord. Hospital Universitari Germans Trias i Pujol | Can Ruti SARS-CoV-2 Sequencing Hub (HUGTiP/IrsiCaixa/IGTP) | Marc Noguera-Julian, Pilar Armengol, Ignacio Blanco, Antoni E Bordoy, Francesc Catala-Moll, Pere-Joan Cardona, Maria Casadellà, Cristina Casañ, Gemma Clara, Bonaventura Clotet, Cristina Esteban, Montserrat Giménez, Mercedes Guerrero, Anna Not, Roger Paredes, Mariona Parera, Verónica Saludes, Alba Sánchez, and Elisa Martró on behalf of the Can Ruti SARS-CoV-2 Sequencing Hub |
| EPI_ISL_2671460, EPI_ISL_2671463, EPI_ISL_2671464, EPI_ISL_2671466, EPI_ISL_2671467, EPI_ISL_2671468, EPI_ISL_2671470 | Center for Laboratory Medicine | Center for Laboratory Medicine | Yannick Gerth |
| EPI_ISL_2672103 | IN State Department of Health Laboratory Services | IN State Department of Health Laboratory Services | Cassandra Campion, Jamie Yeadon, Brian Pope, Lixia Liu, Kyle Brownlee, Melissa Hindenlang, Mark Glazier |
| EPI_ISL_2673675 | SIESP CHIETI - DRIVE IN CHIETI | Istituto Zooprofilattico Sperimentale dell'Abruzzo e Molise "G. Caporale" | Lorusso A, Marcacci M, Di Domenico M, Ancora M, Curini V, Di Lollo Valeria, Mangone I, Rinaldi A, Delli Compagni E, Scialabba S, Caporale M, Di Pasquale A, Cammà C, Puglia I, Calistri P, Savini G |
| EPI_ISL_2673679, EPI_ISL_2673680 | PRESIDIO OSPEDALIERO TAGLIACOZZO | Istituto Zooprofilattico Sperimentale dell'Abruzzo e Molise "G. Caporale" | Lorusso A, Marcacci M, Di Domenico M, Ancora M, Curini V, Di Lollo Valeria, Mangone I, Rinaldi A, Delli Compagni E, Scialabba S, Caporale M, Di Pasquale A, Cammà C, Puglia I, Calistri P, Savini G |
| EPI_ISL_2673681, EPI_ISL_2673682 | SIESP CHIETI - DRIVE IN LANCIANO | Istituto Zooprofilattico Sperimentale dell'Abruzzo e Molise "G. Caporale" | Lorusso A, Marcacci M, Di Domenico M, Ancora M, Curini V, Di Lollo Valeria, Mangone I, Rinaldi A, Delli Compagni E, Scialabba S, Caporale M, Di Pasquale A, Cammà C, Puglia I, Calistri P, Savini G |
| EPI_ISL_2673750 | Akershus University Hospital, Department for Microbiology and Infectious Disease Control | Norwegian Institute of Public Health, Department of Virology | Kathrine Stene-Johansen, Kamilla Heddeland Instefjord, Hilde Elshaug, Garcia Llorente Ignacio, Jon Bråte, Engebretsen Serina Beate,Pedersen Benedikte Nevjen, Line Victoria Moen, Debech Nadia, Atiya R Ali,Marie Paulsen Madsen, Rasmus Riis Kopperud, Hilde Vollan, Karoline Bragstad, Olav Hungnes |
| EPI_ISL_2675085, EPI_ISL_2675086, EPI_ISL_2675087, EPI_ISL_2675088, EPI_ISL_2675089, EPI_ISL_2675090, EPI_ISL_2675091, EPI_ISL_2675092 | Vestfold Hospital, Toensberg Department of Microbiology | Norwegian Institute of Public Health, Department of Virology | Kathrine Stene-Johansen, Kamilla Heddeland Instefjord, Hilde Elshaug, Garcia Llorente Ignacio, Jon Bråte, Engebretsen Serina Beate,Pedersen Benedikte Nevjen, Line Victoria Moen, Debech Nadia, Atiya R Ali,Marie Paulsen Madsen, Rasmus Riis Kopperud, Hilde Vollan, Karoline Bragstad, Olav Hungnes |
| EPI_ISL_2675268, EPI_ISL_2675271, EPI_ISL_2675279, EPI_ISL_2675285, EPI_ISL_2675299 | Hospital General Universitario Gregorio Marañón | Hospital General Universitario Gregorio Marañón | Sergio Buenestado Serrano, Pedro Sola Campoy, Laura Pérez-Lago, Cristina Rodriguez-Grande, Marta Herranz Martin, Victor Manuel de la Cueva, Julia Suárez, Pilar Catalán, Patricia Muñoz, Darío García de Viedma |
| EPI_ISL_2675399, EPI_ISL_2675417, EPI_ISL_2675436, EPI_ISL_2675455, EPI_ISL_2675461, EPI_ISL_2675474, EPI_ISL_2675478, EPI_ISL_2675484, EPI_ISL_2675497, EPI_ISL_2675506, EPI_ISL_2675555, EPI_ISL_2675569, EPI_ISL_2675571, EPI_ISL_2675576, EPI_ISL_2675585, EPI_ISL_2675586, EPI_ISL_2675634, EPI_ISL_2675637, EPI_ISL_2675647, EPI_ISL_2675653, EPI_ISL_2675664, EPI_ISL_2675666, EPI_ISL_2675672, EPI_ISL_2675677, EPI_ISL_2675684, EPI_ISL_2675695, EPI_ISL_2675703, EPI_ISL_2675724, EPI_ISL_2675729, EPI_ISL_2675736, EPI_ISL_2675755, EPI_ISL_2675789, EPI_ISL_2675793, EPI_ISL_2675808, EPI_ISL_2675809, EPI_ISL_2675821, EPI_ISL_2675823, EPI_ISL_2675837, EPI_ISL_2675838, EPI_ISL_2675841, EPI_ISL_2675856, EPI_ISL_2675863, EPI_ISL_2675865 | | | |
| see above | Department of Bacteria, Parasites and Fungi, Statens Serum Institut, Copenhagen, Denmark | Statens Serum Institut Bioinformatics and Microbial Genomics | Danish Covid-19 Genome Consortium |
| EPI_ISL_2676577, EPI_ISL_2676627 | CNR Virus des Infections Respiratoires - France SUD | CNR Virus des Infections Respiratoires - France SUD | Antonin Bal, Gregory Destras, Gwendolyne Burfin, Hadrien Regue, Quentin Semanas, Martine Valette, Bruno Lina, Laurence Josset |
| EPI_ISL_2676702, EPI_ISL_2676704 | LABORATOIRE BOUVIER | CNR Virus des Infections Respiratoires - France SUD | Antonin Bal, Gregory Destras, Gwendolyne Burfin, Hadrien Regue, Quentin Semanas, Martine Valette, Bruno Lina, Laurence Josset |
| EPI_ISL_2676753, EPI_ISL_2676758, EPI_ISL_2676759, EPI_ISL_2676760, EPI_ISL_2676762, EPI_ISL_2676763, EPI_ISL_2676764 | MIRIALIS CLUSES BECHET | CNR Virus des Infections Respiratoires - France SUD | Antonin Bal, Gregory Destras, Gwendolyne Burfin, Hadrien Regue, Quentin Semanas, Martine Valette, Bruno Lina, Laurence Josset |
| EPI_ISL_2676793, EPI_ISL_2676797 | CNR Virus des Infections Respiratoires - France SUD | CNR Virus des Infections Respiratoires - France SUD | Antonin Bal, Gregory Destras, Gwendolyne Burfin, Hadrien Regue, Quentin Semanas, Martine Valette, Bruno Lina, Laurence Josset |
| EPI_ISL_2678439, EPI_ISL_2678445 | Dianovis GmbH Greiz | Robert Koch Institute | unknown |
| EPI_ISL_2678446 | Städtisches Klinikum Dresden Institut für Klinische Chemie und Laboratoriumsmedizin | Robert Koch Institute | unknown |
| EPI_ISL_2678447 | LABOR CHEMNITZ - Zentrum für Diagnostik am Klinikum Chemnitz | Robert Koch Institute | unknown |
| EPI_ISL_2678448 | Landesuntersuchungsanstalt für das Gesundheits- und Veterinärwesen (LUA) Sachsen | Robert Koch Institute | unknown |
| EPI_ISL_2678449 | LABOR CHEMNITZ - Zentrum für Diagnostik am Klinikum Chemnitz | Robert Koch Institute | unknown |
| EPI_ISL_2678451 | Uniklinikum Carl Gustav Carus an der TU Dresden; Institut für Virologie | Robert Koch Institute | unknown |
| EPI_ISL_2678454 | Medizinisches Labor Ostsachsen MVZ GbR Nebenbetriebsstätte Dresden | Robert Koch Institute | unknown |
| EPI_ISL_2678455 | Städtisches Klinikum Dresden Institut für Klinische Chemie und Laboratoriumsmedizin | Robert Koch Institute | unknown |
| EPI_ISL_2678457 | Medizinisches Labor Ostsachsen MVZ GbR Nebenbetriebsstätte Dresden | Robert Koch Institute | unknown |
| EPI_ISL_2678458 | LABOR CHEMNITZ - Zentrum für Diagnostik am Klinikum Chemnitz | Robert Koch Institute | unknown |
| EPI_ISL_2678459, EPI_ISL_2678461 | Uniklinikum Carl Gustav Carus an der TU Dresden; Institut für Virologie | Robert Koch Institute | unknown |
| EPI_ISL_2678462, EPI_ISL_2678463 | Medizinisches Labor Ostsachsen MVZ GbR Nebenbetriebsstätte Dresden | Robert Koch Institute | unknown |
| EPI_ISL_2678467 | Städtisches Klinikum Dresden Institut für Klinische Chemie und Laboratoriumsmedizin | Robert Koch Institute | unknown |
| EPI_ISL_2678468 | Landesuntersuchungsanstalt für das Gesundheits- und Veterinärwesen (LUA) Sachsen | Robert Koch Institute | unknown |
| EPI_ISL_2678470 | Uniklinikum Carl Gustav Carus an der TU Dresden; Institut für Virologie | Robert Koch Institute | unknown |
| EPI_ISL_2678471 | Medizinisches Labor Ostsachsen MVZ GbR Nebenbetriebsstätte Dresden | Robert Koch Institute | unknown |
| EPI_ISL_2678646 | MVZ Labor Dr. Quade & Kollegen GmbH | Robert Koch Institute | unknown |
| EPI_ISL_2678741, EPI_ISL_2678742 | Klinikum Ernst von Bergmann gemeinnützige GmbH - | Robert Koch Institute | unknown |

| | | | |
|---|---|---|---|
| | stationärer Bereich | | |
| EPI_ISL_2681444, EPI_ISL_2681449 | Althaia. Xarxa Assistencial Universitària de Manresa | IrsiCaixa | Marc Noguera-Julian, Mariona Parera, Maria Casadellà, Pilar Armengol, Francesc Catala-Moll, Roger Paredes, Bonaventura Clotet Gloria Trujillo, Rafel Perez Vidal, Jaume Trapé, Carolina Gonzalez-Fernandez, Miquel Micó, Antonia Flor, Roger Paredes, Eulalia Grau, Bonaventura Clotet |
| EPI_ISL_2681468, EPI_ISL_2681469 | University Hospitals of Geneva, Laboratory of Virology | HUG, Laboratory of Virology and the Health2030 Genome Center | Samuel Cordey, Ana Rita Goncalves, Laurent Kaiser, Lorenzo Cerutti, Henri Pegeot, Melyssa Elies, Deborah Penet, Keith Harshman, Ioannis Xenarios, Emmanouil Dermitzakis |
| EPI_ISL_2681551, EPI_ISL_2681559, EPI_ISL_2681561, EPI_ISL_2681571, EPI_ISL_2681603, EPI_ISL_2681605, EPI_ISL_2681614 | Microbiology Department, Laboratori Clínic Metropolitana Nord. Hospital Universitari Germans Trias i Pujol | Can Ruti SARS-CoV-2 Sequencing Hub (HUGTiP/IrsiCaixa/IGTP) | Marc Noguera-Julian, Pilar Armengol, Ignacio Blanco, Antoni E Bordoy, Francesc Catala-Moll, Pere-Joan Cardona, Maria Casadellà, Cristina Casañ, Gemma Clara, Bonaventura Clotet, Cristina Esteban, Montserrat Giménez, Mercedes Guerrero, Anna Not, Roger Paredes, Mariona Parera, Verónica Saludes, Alba Sánchez, and Elisa Martró on behalf of the Can Ruti SARS-CoV-2 Sequencing Hub |
| EPI_ISL_2682680, EPI_ISL_2682739 | Lighthouse Lab in Alderley Park | Wellcome Sanger Institute for the COVID-19 Genomics UK (COG-UK) Consortium | Jacquelyn Wynn, Mairead Hyland, The Lighthouse Lab in Alderley Park and Alex Alderton, Roberto Amato, Jeffrey Barrett, Sonia Goncalves, Ewan Harrison, David K. Jackson, Ian Johnston, Dominic Kwiatkowski, Cordelia Langford, John Sillitoe on behalf of the Wellcome Sanger Institute COVID-19 Surveillance Team |
| EPI_ISL_2684242, EPI_ISL_2684243, EPI_ISL_2684253, EPI_ISL_2684259, EPI_ISL_2684268, EPI_ISL_2684279, EPI_ISL_2684334, EPI_ISL_2684353, EPI_ISL_2684357, EPI_ISL_2684359, EPI_ISL_2684364, EPI_ISL_2684395, EPI_ISL_2684401, EPI_ISL_2684432, EPI_ISL_2684436, EPI_ISL_2684439, EPI_ISL_2684465, EPI_ISL_2684500, EPI_ISL_2684501, EPI_ISL_2684527, EPI_ISL_2684536, EPI_ISL_2684557, EPI_ISL_2684575, EPI_ISL_2684621, EPI_ISL_2684628, EPI_ISL_2684642, EPI_ISL_2684676, EPI_ISL_2684678, EPI_ISL_2684681, EPI_ISL_2684683, EPI_ISL_2684691, EPI_ISL_2684694, EPI_ISL_2684726, EPI_ISL_2684738, EPI_ISL_2684759, EPI_ISL_2684764, EPI_ISL_2684766, EPI_ISL_2684767, EPI_ISL_2684769, EPI_ISL_2684778, EPI_ISL_2684785, EPI_ISL_2684790, EPI_ISL_2684799, EPI_ISL_2684810, EPI_ISL_2684850, EPI_ISL_2684851, EPI_ISL_2684869, EPI_ISL_2684876, EPI_ISL_2684904, EPI_ISL_2684944, EPI_ISL_2685027, EPI_ISL_2685039, EPI_ISL_2685044, EPI_ISL_2685067, EPI_ISL_2685073, EPI_ISL_2685076, EPI_ISL_2685091, EPI_ISL_2685095, EPI_ISL_2685112, EPI_ISL_2685192, EPI_ISL_2685225, EPI_ISL_2685233, EPI_ISL_2685241, EPI_ISL_2685242, EPI_ISL_2685323, EPI_ISL_2685328, EPI_ISL_2685335, EPI_ISL_2685349, EPI_ISL_2685353, EPI_ISL_2685470, EPI_ISL_2685472, EPI_ISL_2685480, EPI_ISL_2685498, EPI_ISL_2685505, EPI_ISL_2685508, EPI_ISL_2685559, EPI_ISL_2685560, EPI_ISL_2685567, EPI_ISL_2685583, EPI_ISL_2685589, EPI_ISL_2685590, EPI_ISL_2685598, EPI_ISL_2685604, EPI_ISL_2685613, EPI_ISL_2685618, EPI_ISL_2685637, EPI_ISL_2685666, EPI_ISL_2685669, EPI_ISL_2685671, EPI_ISL_2685676, EPI_ISL_2685683, EPI_ISL_2685696, EPI_ISL_2685715, EPI_ISL_2685721, EPI_ISL_2685723, EPI_ISL_2685740, EPI_ISL_2685755, EPI_ISL_2685767, EPI_ISL_2685808, EPI_ISL_2685833 | | | |
| see above | Statens Serum Institut Bioinformatics and Microbial Genomics | Statens Serum Institut Bioinformatics and Microbial Genomics | Danish Covid-19 Genome Consortium |
| EPI_ISL_2685892, EPI_ISL_2685893 | Department of Clinical Microbiology | GIGA Medical Genomics | Keith Durkin, Maria Artesi, Sébastien Bontems, Raphaël Boreux, Bouchra Boujemla, Nathalie Renotte, Cécile Meex, Pierrette Melin, Marie-Pierre Hayette, Vincent Bours |
| EPI_ISL_2686178 | Amazon Dx, AUV2 Laboratory | Amazon Dx, AUV2 Laboratory | Luke Meredith |
| EPI_ISL_2686618, EPI_ISL_2686619, EPI_ISL_2686656, EPI_ISL_2686657, EPI_ISL_2686694, EPI_ISL_2686739, EPI_ISL_2686740, EPI_ISL_2686751, EPI_ISL_2686786, EPI_ISL_2686789 | Fulgent Genetics | Centers for Disease Control and Prevention Division of Viral Diseases, Pathogen Discovery | Dakota Howard, Dhwani Batra, Peter W. Cook, Kara Moser, Adrian Paskey, Jason Caravas, Benjamin Rambo-Martin, Shatavia Morrison, Christopher Gulvick, Scott Sammons, Yvette Unoarumhi, Darlene Wagner, Matthew Schmerer, Harry Gao, Mickey Li, John Gao, Joseph Fierro, Benafsh Sapra, Becky Tsai, Yan Meng, Doreen Ng, James Xie, Clinton R. Paden, Duncan MacCannell |
| EPI_ISL_2686812, EPI_ISL_2686823, EPI_ISL_2686824, EPI_ISL_2686825, EPI_ISL_2686831, EPI_ISL_2686834, EPI_ISL_2686835, EPI_ISL_2686836 | EHNV | Laboratory of genomics and metagenomics | Trestan Pillonel, Damien Jacot, Sébastien Aeby, Gilbert Greub, Claire Bertelli |
| EPI_ISL_2687999 | Department of Public Health Bucharest | National Institute of Infectious Diseases-Prof. Dr. Matei Bals Molecular Diagnostics Laboratory | Corina Casangiu, Leontina Banica, Marius Surleac, Petre Milu, Robert Hohan, Simona Paraschiv, Dan Otelea |
| EPI_ISL_2688239 | IN State Department of Health Laboratory Services | IN State Department of Health Laboratory Services | Cassandra Campion, Jamie Yeadon, Brian Pope, Lixia Liu, Kyle Brownlee, Melissa Hindenlang, Mark Glazier |
| EPI_ISL_2689088 | MD PHL | MD PHL | Maryland Department of Health Laboratories Administration |
| EPI_ISL_2690195 | Institute of Virology, Biomedical Research Center of the Slovak Academy of Sciences, Bratislava | Faculty of Natural Sciences, Comenius University, Bratislava | Viktoria Cabanova, Kristina Borsova, Brona Brejova, Viktoria Hodorova, Sabina Fumacova Havlikova, Juraj Kopacek, Martina Lickova, Lubomira Lukacikova, Martina Nebohacova, Monika Slavikova, Tomas Vinar, Jozef Nosek, Boris Klempa |
| EPI_ISL_2690210 | Institute of Virology, Biomedical Research Center of the Slovak Academy of Sciences, Bratislava | Faculty of Natural Sciences, Comenius University, Bratislava | Brona Brejova, Viktoria Cabanova, Kristina Borsova, Viktoria Hodorova, Sabina Fumacova Havlikova, Juraj Kopacek, Martina Lickova, Lubomira Lukacikova, Martina Nebohacova, Monika Slavikova, Tomas Vinar, Jozef Nosek, Boris Klempa |
| EPI_ISL_2690355 | Arizona State University | Arizona State University | Peter T. Skidmore, LaRinda A. Holland, Matthew F. Smith, Rabia Maqsood, Nicholas J. Mellor, Joy M. Blain, Valerie Harris, Joshua LaBaer, Vel Murugan, Efrem S. Lim |
| EPI_ISL_2692715 | Salud Digna | Instituto Nacional de Medicina Genomica | Cedro-Tanda A, Hidalgo-Miranda A, Mendoza-Vargas A, Reyes-Grajeda JP, Abraham Campos-Romero, Moreno-Camacho José Luis, Rodriguez-Gallegos Jorge, Luna-Ruiz Marco, Gonzalez-Barrera D, Rangel-DeLeon D, Munguia-Garza P, Ramirez-Vega O, Escobar-Arrazola, M, Herrera-Montalvo LA. |
| EPI_ISL_2693065 | NHLS_VIRO | KRISP, KZN Research Innovation and Sequencing Platform | Giandhari Jennifer, Pillay Sureshnee, Yajna Ramphal, Naidoo Yeshnee, Tshabuila Derek,Tegally Houriiyah, San James, Wilkinson Eduan, de Oliveira Tulio |
| EPI_ISL_2693118, EPI_ISL_2693120, EPI_ISL_2693123, EPI_ISL_2693124, EPI_ISL_2693265, EPI_ISL_2693269, EPI_ISL_2693270, EPI_ISL_2693273, EPI_ISL_2693275, EPI_ISL_2693276, EPI_ISL_2693279, EPI_ISL_2693280, EPI_ISL_2693281 | | | |
| see above | UW Virology Lab | UW Virology Lab | Pavitra Roychoudhury, Hong Xie, Lasata Shrestha, Tien V. Nguyen, Shah Mohamed Bakhash, Michelle Lin, Noah R. Baker, Ricardo Perez, Sean Ellis, Nathan Breit, Robert J. Livingston, Meei-Li Huang, Keith R Jerome, Patrick Mathias, Alexander Greninger |
| EPI_ISL_2693823 | UW Virology Lab | UW Virology Lab | Pavitra Roychoudhury, Hong Xie, Lasata Shrestha, Shah Mohamed Bakhash, Tien V. Nguyen, Noah R. Baker, Sean Ellis, Meei-Li Huang, Keith R Jerome, Alexander Greninger |
| EPI_ISL_2693916, EPI_ISL_2693917, EPI_ISL_2693918 | Virology Unit, Institut Pasteur du Cambodge | Virology Unit, Institut Pasteur du Cambodge | Cecile Troupin, Leakhena Pum, Jurre Y Siegers, Ly Sovann, Kraing Sidonn, Yi Sengdoeurn, Chin Savuth, Chau Darapheak, Veasna Duong, Erik A Karlsson |
| EPI_ISL_2694788, EPI_ISL_2694796 | Diagnostyka. Laboratoria Medyczne. | 1. ViroGenetics - BSL3 Laboratory of Virology, Maopolska Centre of Biotechnology, Jagiellonian University; 2. Diagtron Laboratoria ukasz Rbalski | Rabalski L., Gromowski,T., Rogalska-Kupiec M., Swadzba J., Wydmanski,W., Mazur-Panasiuk,N., Kowalski,M., Maciej Kosinski, Natalia Derewonko, Szulc,P., Sylwia Januszczak, Labaj,P.P., Pyrc,K. |
| EPI_ISL_2695060, EPI_ISL_2695061, EPI_ISL_2695062, EPI_ISL_2695063, EPI_ISL_2695064 | H Divino Espirito Santo - Ponta Delgada | Instituto Nacional de Saude (INSA) | Borges et al |
| EPI_ISL_2695256 | SESARAM | Instituto Nacional de Saude (INSA) | Borges et al |
| EPI_ISL_2695267 | CHU Sao Joao, Porto | Instituto Nacional de Saude (INSA) | Borges et al |
| EPI_ISL_2695382, EPI_ISL_2695386 | Labeto - CAB - Leiria | Instituto Nacional de Saude (INSA) | Borges et al |
| EPI_ISL_2697675 | National Platform bis UMONS/Jolimont | National Platform bis UMONS/Jolimont | François Dufrasne, Guillaume Bayon-Vicente, Florian Juszczak, Gautier Detry, Ruddy Wattiez |
| EPI_ISL_2707632, EPI_ISL_2707634, EPI_ISL_2707691, EPI_ISL_2707701, EPI_ISL_2707702, EPI_ISL_2708085 | Originating lab: Wales Specialist Virology Centre Sequencing lab: Pathogen Genomics Unit | Public Health Wales Microbiology Cardiff Wales Specialist Virology Centre | Catherine Moore, Johnathan Evans, Laura Gifford, Malorie Perry, Simon Cottrell, Angela Marchbank, Alec Birchley, Alexander Adams, Amy Gaskin, Bree Gatica-Wilcox, Jason Coombes, Joel Southgate, Lauren Gilbert, Lee Graham, Nicole Pacchiarini, Sara Kumziene-Summerhayes, Sarah Taylor, Sophie Jones, Sara Rey, Matthew Bull, Joanne Watkins, Sally Corden, Tom Connor |
| EPI_ISL_2709822 | MD PHL | MD PHL | Maryland Department of Health Laboratories Administration |
| EPI_ISL_2710223, EPI_ISL_2710224 | HOSPITAL UNIVERSITARIO SON ESPASES | HOSPITAL UNIVERSITARIO SON ESPASES | Servicio de Microbiología, Hospital Universitario Son Espases |
| EPI_ISL_2712843, EPI_ISL_2712844 | Hospital Universitari Vall d'Hebron - Vall d'Hebron Institut de Recerca | Hospital Universitari Vall d'Hebron - Vall d'Hebron Institut de Recerca | Cristina Andrés, Maria Piñana, Alejandra González-Sánchez, Damir Garcia-Cehic, Ariadna Rando, Juliana Esperalba, Maria Gema Codina, Carla Castillo, Maria Carmen Martin, Tomàs Pumarola, Josep Quer, Andrés Antón |
| EPI_ISL_2712953, EPI_ISL_2712956 | Wyoming Public Health Laboratory | Wyoming Public Health Laboratory | Jim Mildenberger, Taylor Fearing, Channing Weber, Ashley Norberg, Chayse Rowley, Marley Goetz, Brian Dominguez, Elliot Thomasson, Sam Britz, Cari Sloma, Robert Petit, and Rob Christensen |

| | | | |
|---|---|---|---|
| EPI_ISL_2713054 | University of Wisconsin-Madison AIDS Vaccine Research Laboratories | University of Wisconsin-Madison AIDS Vaccine Research Laboratories | Gage Moreno, Katarina Braun, et al. AIDS Vaccine Research Laboratories |
| EPI_ISL_2713063 | Oregon State Public Health Laboratory | Oregon State Public Health Laboratory | Rafia Razzaque, Eugene Yeboah, Vanda Makris, Laura Tsaknaridis, John Fontana and Shane Sevey |
| EPI_ISL_2716209 | The Jackson Laboratory | The Jackson Laboratory | Long J, Renzette N, Adams M, Omerza G, Kelly K, Li L |
| EPI_ISL_2716295 | Kansas Health and Environmental Lab | Kansas Health and Environmental Lab | Katherine Wiggins, Mike Grose, Jonathan Barnell, Ben Olsen, and Phil Adam |
| EPI_ISL_2716934, EPI_ISL_2716936 | Southern Nevada Public Health Laboratory | Southern Nevada Public Health Laboratory | Michael Picker |
| EPI_ISL_2718506, EPI_ISL_2718525 | Lighthouse Lab in Glasgow | Wellcome Sanger Institute for the COVID-19 Genomics UK (COG-UK) Consortium | Harper VanSteenhouse, Yumi Kasai, David Gray, Carol Clugston, Anna Dominiczak and Alex Alderton, Roberto Amato, Jeffrey Barrett, Sonia Goncalves, Ewan Harrison, David K. Jackson, Ian Johnston, Dominic Kwiatkowski, Cordelia Langford, John Sillitoe on behalf of the Wellcome Sanger Institute COVID-19 Surveillance Team |
| EPI_ISL_2724393, EPI_ISL_2724394 | Viollier AG | Department of Biosystems Science and Engineering, ETH Zürich | Christian Beisel, Sarah Nadeau, Chaoran Chen, Ivan Topolsky, Kim Philipp Jablonski, Lara Fuhrmann, Rebecca Denes, Mirjam Feldkamp, Ina Nissen, Natascha Santacroce, Elodie Burcklen, Christiane Beckmann, Maurice Redondo, Olivier Kobel, Christoph Noppen, Niko Beerenwinkel, Tanja Stadler |
| EPI_ISL_2725105 | Innlandet Hospital Trust, Division Lillehammer, Department for Medical Microbiology | Norwegian Institute of Public Health, Department of Virology | Kathrine Stene-Johansen, Kamilla Heddeland Instefjord, Hilde Elshaug, Garcia Llorente Ignacio, Jon Bråte, Engebretsen Serina Beate,Pedersen Benedikte Nevjen, Line Victoria Moen, Debech Nadia, Atiya R Ali,Marie Paulsen Madsen, Rasmus Riis Kopperud, Hilde Vollan, Karoline Bragstad, Olav Hungnes |
| EPI_ISL_2725185, EPI_ISL_2725187, EPI_ISL_2725188, EPI_ISL_2725189, EPI_ISL_2725202, EPI_ISL_2725203 | Lab. Microbiologia e Virologia Cotugno A.O. dei Colli - Istituto Zooprofilattico Sperimentale del Mezzogiorno | Telethon Institute of Genetics and Medicine (TIGEM) | Antonio Grimaldi Patrizia Annunziata Francesco Panariello Biancamaria Pierri Claudia Tiberio Teresa Giuliano Valentina Bouche Chiara Colantuono Maria Concetta Cuomo Denise Di Concilio Lucio Di Filippo Anna Manfredi Marcello Salvi Antonio Limone Luigi Atripaldi Pellegrino Cerino Andrea Ballabio Davide Cacchiarelli |
| EPI_ISL_2725545, EPI_ISL_2725548 | Azienda Sanitaria dell'Alto Adige - Laboratorio Aziendale di Microbiologia e Virologia | Azienda Sanitaria dell'Alto Adige | Irene Bianconi |
| EPI_ISL_2726701 | Eurofins-NMDL | Eurofins-NMDL | Thierry Janssens, Anne Vogel, Lisa Dreesens, Roy Masius, Katja Plathan, Anco Molijn, Maurine Leversteijn-van Hall, Thomas van der Toorn, Cornel Brouwer, Diviana Lakker, Hamza Saleem, Layonel Jacobus, Rosalie Van der Stap, Marisol Williams, Chris van Koppen, Senay Ozturk, Marvin Ruiter, Mark de Jong, Alexandra Hansen, Coen de Jong, Madhavi Kharagjitsing, Ramon Koers, Rens Driel, Thomas van der Toorn, Bram Weijers, Chantal Schenker, Humaira Yousaf, Jelmer Eerland, Melek Aras, Rousjan Amir, Enes Sekerci, Goyaert Roosen, Laura Weijenberg, Mathijs Wielen, Nienke Meijer, Tobias van der Heijde, Chinmoy Saha, Glenn Bohms, Iman El-Idrioui, Jorian Flik, Lida Cen, Simon Lansu |
| EPI_ISL_2727017 | National Institute of Public Health | National Institute of Public Health | Helena Jirincova, Jaromira Vecerova, Timotej Suri, Dusan Trnka, Alexander Nagy |
| EPI_ISL_2727060 | MD PHL | MD PHL | Maryland Department of Health Laboratories Administration |
| EPI_ISL_2727580 | NJDOH, Public Health and Environmental Laboratories | NJ_PHEL | Lindsey Bodnar, Shiv K. Verma, Jacquelyn Deverell, Ryan Pachucki, Dana Woell, Allison Roder, Byeong Jeong |
| EPI_ISL_2727815, EPI_ISL_2727819, EPI_ISL_2727821, EPI_ISL_2727822, EPI_ISL_2727829 | Wisconsin State Laboratory of Hygiene Communicable Disease Division | Wisconsin State Laboratory of Hygiene Communicable Disease Division | Abigail C. Shockey, Alicia J. Mooney, Erika M. Hanson, Tonya Danz, Richard Griesser, Sara Wagner, Kelsey R. Florek |
| EPI_ISL_2728621, EPI_ISL_2728626 | SYNLAB | GIGA Medical Genomics | Keith Durkin, Maria Artesi, Bouchra Boujemla, Nathalie Renotte, Nadine Cambisano, Sébastien Bontems, Cécile Meex, Claire Gourzonès, Olivier Ek, Laurent Gillet, Marie-Pierre Hayette, Vincent Bours |
| EPI_ISL_2731154, EPI_ISL_2731166, EPI_ISL_2731184, EPI_ISL_2731206, EPI_ISL_2731230, EPI_ISL_2731239, EPI_ISL_2731249, EPI_ISL_2731257, EPI_ISL_2731258, EPI_ISL_2731259, EPI_ISL_2731260 | | | |
| see above | Lighthouse Lab in Glasgow | Wellcome Sanger Institute for the COVID-19 Genomics UK (COG-UK) Consortium | Harper VanSteenhouse, Yumi Kasai, David Gray, Carol Clugston, Anna Dominiczak and Alex Alderton, Roberto Amato, Jeffrey Barrett, Sonia Goncalves, Ewan Harrison, David K. Jackson, Ian Johnston, Dominic Kwiatkowski, Cordelia Langford, John Sillitoe on behalf of the Wellcome Sanger Institute COVID-19 Surveillance Team |
| EPI_ISL_2731353, EPI_ISL_2731360 | Microvida | Microvida | Suzan D. Pas, Jaco J. Verweij, Joep J. J. M. Stohr |
| EPI_ISL_2754070 | Wyoming Public Health Laboratory | Wyoming Public Health Laboratory | Jim Mildenberger, Taylor Fearing, Channing Weber, Ashley Norberg, Chayse Rowley, Marley Goetz, Brian Dominguez, Elliot Thomasson, Sam Britz, Cari Sloma, Robert Petit, and Rob Christensen |
| EPI_ISL_2754306 | Arizona State Public Health Laboratory | Arizona State Public Health Laboratory | Trung Huynh, Jessica Escobar, Katherine Fullerton, Nobuko Fukushima, Matthew Contursi, Stacy White, Linda Getsinger, Victor Waddell |
| EPI_ISL_2757689 | Presidio di Brindisi Di Summa - Perrino | Istituto Zooprofilattico Sperimentale della Puglia e della Basilicata | Parisi A., Bianco A., Capozzi L., Del Sambro L., Simone D., Difato L., Santoro A., Ridolfi D., Giannico A. |
| EPI_ISL_2757700 | Ospedale Vito Fazzi | Istituto Zooprofilattico Sperimentale della Puglia e della Basilicata | Parisi A., Bianco A., Capozzi L., Del Sambro L., Simone D., Difato L., Lobreglio G. |
| EPI_ISL_2758028 | Austrian Agency for Health and Food Safety (AGES) | Bergthaler laboratory, CeMM Research Center for Molecular Medicine of the Austrian Academy of Sciences | Lukas Endler, Anna Schedl, Fabian Amman, Petr Triska, Matthew Thornton, Thomas Penz, Benedikt Agerer, Maelle Le Moing, Michael Schuster, Bekir Erguner, Jan Laine, Martin Senekowitsch, Christoph Bock, Andreas Bergthaler |
| EPI_ISL_2758221 | Laboratoire Hôpital Sainte Musse | Laboratoire Hôpital Sainte Musse | Lionel Chollet |
| EPI_ISL_2758371, EPI_ISL_2758376 | Berkeley Medical Center | WVU and Marshall University Combined Genomics Core Facilities | James Denvir, Peter Stoilov, Peter Perrotta, Wesley Kimble, Ryan Percifield |
| EPI_ISL_2758400 | QLabs | WVU and Marshall University Combined Genomics Core Facilities | James Denvir, Peter Stoilov, Peter Perrotta, Wesley Kimble, Ryan Percifield |
| EPI_ISL_2758929 | The Ohio State University Applied Microbiology Services Laboratory | The Ohio State University Applied Microbiology Services Laboratory | Seth A. Faith PhD |
| EPI_ISL_2759726 | SYNLAB MVZ Weiden | Robert Koch Institute | unknown |
| EPI_ISL_2759823 | Uniklinikum Regensburg; Institut für klinische Mikrobiologie und Hygiene | Robert Koch Institute | unknown |
| EPI_ISL_2760230, EPI_ISL_2760251, EPI_ISL_2760254, EPI_ISL_2760260 | Helios Universitätsklinikum Wuppertal: Universität Witten/Herdecke: Institut für Medizinische Labordiagnostik | Robert Koch Institute | unknown |
| EPI_ISL_2760298 | MVZ Labor Krone GbR | Robert Koch Institute | unknown |
| EPI_ISL_2760452 | MVZ Dr. Eberhard & Partner Dortmund | Robert Koch Institute | unknown |
| EPI_ISL_2760460, EPI_ISL_2760466 | amedes MVZ DIAMEDIS Sennestadt | Robert Koch Institute | unknown |
| EPI_ISL_2761153, EPI_ISL_2761156, EPI_ISL_2761164, EPI_ISL_2761254, EPI_ISL_2761316, EPI_ISL_2761453, EPI_ISL_2761489, EPI_ISL_2761533, EPI_ISL_2761556 | Bioscientia Labor Wermsdorf | Robert Koch Institute | unknown |
| EPI_ISL_2761670, EPI_ISL_2761671 | MVZ Labor Dr. Fenner und Kollegen (Standort Hamburg) | Robert Koch Institute | unknown |
| EPI_ISL_2761752, EPI_ISL_2761754, EPI_ISL_2761761, EPI_ISL_2761773, EPI_ISL_2761817 | CENTOGENE Frankfurt Laboratory: Niederlassung Industriepark Höchst | Robert Koch Institute | unknown |

| | | | |
|---|---|---|---|
| EPI_ISL_2761918 | Medizinisch-Diagnostisches Labor Kempten allgäulab | Robert Koch Institute | unknown |
| EPI_ISL_2762045, EPI_ISL_2762073 | Bioscientia Labor Wermsdorf | Robert Koch Institute | unknown |
| EPI_ISL_2762136 | IMD - MVZ Labor Greifswald | Robert Koch Institute | unknown |
| EPI_ISL_2762137, EPI_ISL_2762140 | IMD - MVZ Labor Martinsried | Robert Koch Institute | unknown |
| EPI_ISL_2762228, EPI_ISL_2762272 | Bayerisches Landesamt für Gesundheit und Lebensmittelsicherheit (LGL) | Robert Koch Institute | unknown |
| EPI_ISL_2768099 | Felix-Platter Spital | Clinical Bacteriology | Tim Roloff, Fanny Wegner, Helena MB Seth-Smith, Alfredo Mari, Karoline Leuzinger, Julia Bielicki, Manuel Battegay, Hans Hirsch, Adrian Egli |
| EPI_ISL_2769139 | Spital Männedorf AG | Institute of Medical Virology | Verena Kufner, Gabriela Ziltener, Maryam Zaheri, Stefan Schmutz, Annette Audigé, Maria Grünberg, Kevin Steiner, Jon Huder, Cyril Shah, Riccarda Capaul, Guido Bloemberg, Jürg Böni, Michael Huber, Alexandra Trkola |
| EPI_ISL_2769143 | Swiss Analysis AG | Institute of Medical Virology | Verena Kufner, Gabriela Ziltener, Maryam Zaheri, Stefan Schmutz, Annette Audigé, Maria Grünberg, Kevin Steiner, Jon Huder, Cyril Shah, Riccarda Capaul, Guido Bloemberg, Jürg Böni, Michael Huber, Alexandra Trkola |
| EPI_ISL_2772668 | Max von Pettenkofer Institute, Virology, National Reference Center for Retroviruses, LMU Munich | Laboratory for Functional Genome Analysis; Dept. Genomics; Gene Center of the LMU Munich | Max Muenchhoff; Stefan Krebs; Alexander Graf; Oliver Keppler; Helmut Blum |
| EPI_ISL_2774090 | Multiplex DX | Multiplex DX | Diana Drobna, Veronika Mancikova, Silvia Rybecka, Jakub Kovac, Pavol Cekan |
| EPI_ISL_2774218 | IRCCS San Gallicano Dermatological Institute | IRCCS Regina Elena National Cancer Institute | Ludovica Ciuffreda, Sara Donzelli, Alice Massacci, Valentina Ricca, Fabrizio Ensoli, Fulvia Pimpinelli, Maurizio Fanciulli, Giovanni Blandino, Aldo Morrone, Gennaro Ciliberto |
| EPI_ISL_2774268, EPI_ISL_2774269, EPI_ISL_2774271, EPI_ISL_2774282 | Baylor Scott & White-Temple | Baylor Scott & White-Temple | Ari Rao, Linden Morales, Kimberly Walker, Marcus Volz, Shelby Johnson |
| EPI_ISL_2774440, EPI_ISL_2774450, EPI_ISL_2774451 | Multiplex DX | Multiplex DX | Diana Drobna, Veronika Mancikova, Silvia Rybecka, Jakub Kovac, Pavol Cekan |
| EPI_ISL_2812556 | Communicable Disease Laboratory, Public Health Directorate | Communicable Disease Laboratory, Public Health Directorate | Alwasti,H., AlHujairi,Z., AlAbbas,Z., Marhoon,A., Almoamen,G. |

### supp. Table S3: Beta

We gratefully acknowledge the following Authors from the Originating laboratories responsible for obtaining the specimens, as well as the Submitting laboratories where the genome data were generated and shared via GISAID, on which this research is based.

All Submitters of data may be contacted directly via www.gisaid.org

Authors are sorted alphabetically.

| Accession ID | Originating Laboratory | Submitting Laboratory | Authors |
|---|---|---|---|
| EPI_ISL_2171924 | Lighthouse Lab in Alderley Park | Wellcome Sanger Institute for the COVID-19 Genomics UK (COG-UK) Consortium | Jacquelyn Wynn, Mairead Hyland, The Lighthouse Lab in Alderley Park and Alex Alderton, Roberto Amato, Jeffrey Barrett, Sonia Goncalves, Ewan Harrison, David K. Jackson, Ian Johnston, Dominic Kwiatkowski, Cordelia Langford, John Sillitoe on behalf of the Wellcome Sanger Institute COVID-19 Surveillance Team |
| EPI_ISL_2179665 | National Platform bis UMONS/Jolimont | National Platform bis UMONS/Jolimont | François Dufrasne, Guillaume Bayon-Vicente, Florian Juszczak, Eric Tarantino, Gautier Detry, Ruddy Wattiez |
| EPI_ISL_2179795 | Servicio Microbiología Hospital La Paz | Servicio Microbiología Hospital La Paz | Fernando Lázaro, Rubén Cáceres, Jesús Mingorance Cruz, Elie Dahdouh |
| EPI_ISL_2226240 | Plateforme de testing Namuroise | Plateforme de testing Namuroise | Lesly Nyinkeu Kemamen; Otto Gaetan ; Denis Olivier ; Degosserie Jonathan ; Mullier François |
| EPI_ISL_2228178, EPI_ISL_2228204 | UMC Groningen, Clinical Virology, Department of Medical Microbiology and Infection Prevention | UMC Groningen, Clinical Virology, Department of Medical Microbiology and Infection Prevention | Hubert Niesters, Alexander Friedrich, Erley Lizarazo-Forero, Monika Fliss, Lilli Gard, Sigrid Rosema, Coretta Van Leer-Buter, Xuewei Zhou, Marjolein Knoester |
| EPI_ISL_2228863, EPI_ISL_2228865 | CH.INTERCOMMUNAL DE CRETEIL | Department of Virology, Henri Mondor University Hospital, Assistance Publique Hôpitaux de Paris, Université Paris-Est Créteil, INSERM U955 | Christophe Rodriguez, Slim Fourati, Vanessa Demontant, Guillaume Gricourt, Melissa N'Debi, Alexandre Soulier, Elisabeth Trawinski, Jean-Michel Pawlotsky |
| EPI_ISL_2235024, EPI_ISL_2235031 | Pandemic Response Lab - NYC | Pandemic Response Lab, R&D | Henry Lee, Michael Hammerling, Melissa Hopkins, Cybill del Castillo, Shinyoung Clair Kang, William Ward, Pradeep Bugga, Sol Rey, Dylan Law, Katharine Nelson, Haiping Hao, Jon Laurent |
| EPI_ISL_2245449 | University of Liège COVID-19 testing center | GIGA Medical Genomics | Keith Durkin, Maria Artesi, Bouchra Boujemla, Nathalie Renotte, Cécile Meex, Sébastien Bontems, Fabrice Bureau, Laurent Gillet, Wouter Coppieters, Marie-Pierre Hayette, Vincent Bours |
| EPI_ISL_2253116, EPI_ISL_2253117, EPI_ISL_2253124 | SARS-CoV-2 testing team, National Institute of Infectious Diseases | Pathogen Genomics Center, National Institute of Infectious Diseases | Tsuyoshi Sekizuka, Kentaro Itokawa, Rina Tanaka, Masanori Hashino, Nozomu Hanaoka, Masumichi Saito, Naomi Nojiri, Hazuka Y Furihata, Sana Uchikoba, Tsuguto Fujimoto, Makoto Kuroda |
| EPI_ISL_2262449 | MVZ Labor Dr. Limbach & Kollegen GbR | Robert Koch Institute | unknown |
| EPI_ISL_2263792, EPI_ISL_2263795, EPI_ISL_2263840, EPI_ISL_2263936 | SYNLAB MVZ Leverkusen | Robert Koch Institute | unknown |
| EPI_ISL_2264542 | MDI Limbach Berlin GmbH; MVZ Labor Berlin | Robert Koch Institute | unknown |
| EPI_ISL_2265003 | Medizinisches Labor Wahl Lüdenscheid | Robert Koch Institute | unknown |
| EPI_ISL_2265085 | SYNLAB MVZ Leinfelden-Echterdingen | Robert Koch Institute | unknown |
| EPI_ISL_2266111 | Labor Dr. Heidrich & Kollegen MVZ GmbH Hamburg | Robert Koch Institute | unknown |
| EPI_ISL_2266668 | SYNLAB MVZ Trier | Robert Koch Institute | unknown |
| EPI_ISL_2266982 | SYNLAB MVZ Leverkusen | Robert Koch Institute | unknown |
| EPI_ISL_2267030 | Synlab MVZ Augsburg | Robert Koch Institute | unknown |
| EPI_ISL_2267112, EPI_ISL_2267131, EPI_ISL_2267148, EPI_ISL_2267222 | SYNLAB MVZ Leverkusen | Robert Koch Institute | unknown |
| EPI_ISL_2267408, EPI_ISL_2267536 | SYNLAB MVZ Heidelberg | Robert Koch Institute | unknown |
| EPI_ISL_2267636, EPI_ISL_2267663, EPI_ISL_2267761 | SYNLAB MVZ Leverkusen | Robert Koch Institute | unknown |
| EPI_ISL_2268185 | CENTOGENE Frankfurt Laboratory: Niederlassung Industriepark Höchst | Robert Koch Institute | unknown |
| EPI_ISL_2272557 | Alaska State Virology Laboratory | Alaska State Virology Laboratory | Stephanie DeRonde, Elva House, Jacob Zidek, Lisa Smith, Ph.D., Jack Chen, Ph.D. |
| EPI_ISL_2272765 | Platform BIS UZA/UAntwerpen | Labo Klinische Biologie, UZA | Marie Le Mercier, Jasmine Coppens, Basil Britto Xavier, Christine Lammens, Veerle Matheeussen, Herman Goossens |
| EPI_ISL_2274245 | PHV-FSS | PHV-FSS | Son Nguyen |
| EPI_ISL_2280872 | Aegis Sciences Corporation | Centers for Disease Control and Prevention Division of Viral Diseases, Pathogen Discovery | Dakota Howard, Dhwani Batra, Peter W. Cook, Kara Moser, Adrian Paskey, Jason Caravas, Benjamin Rambo-Martin, Shatavia Morrison, Christopher Gulvick, Scott Sammons, Yvette Unoarumhi, Darlene Wagner, Matthew Schmerer, Cyndi Clark, Patrick Campbell, Rob Case, Vikramsinha Ghorpade, Holly Houdeshell, Ola Kvalvaag, Dillon Nall, Ethan Sanders, Alec Vest, Shaun Westlund, Matthew Hardison, Clinton R. Paden, Duncan MacCannell |
| EPI_ISL_2289468 | LABORATOIRE DE BIOLOGIE MEDICALE | CNR Virus des Infections Respiratoires - France SUD | Antonin Bal, Gregory Destras, Gwendolyne Burfin, Hadrien Regue, Quentin Semanas, Martine Valette, Bruno Lina, Laurence Josset |
| EPI_ISL_2293466 | Hospital | National Reference Center for Viruses of Respiratory Infections, Institut Pasteur, Paris | Marion Barbet, Sylvie Behillil, Méline Bizard, Angela Brisebarre, Camille Capel, Vincent Enouf, Louise Lefrançois, Frédéric Lemoine, Christophe Malabat, Corinne Maufrais, Etienne Simon-Lorière, Maud Vanpeene, Sylvie Van der Werf ,Laurence Louvet |
| EPI_ISL_2303970 | Dutch COVID-19 response team | National Institute for Public Health and the Environment (RIVM) | Adam Meijer, Harry Vennema, Dirk Eggink, Jeroen Cremer, Sharon van den Brink, Bas van der Veer, AnneMarie van den Brandt, Lisa Wijsman, Kim Freriks, Ryanne Jaarsma, Euníce Then, Lynn Aarts, Sanne Bos, Melissa van Tuil, Linda van de Nes, Sjoerd Kuiling, James Groot, Florian Zwagemaker, Dennis Schmitz, Annelies Kroneman, Karim Hajji, Chantal Reusken, on behalf of the national COVID-19 response team |
| EPI_ISL_2305050, EPI_ISL_2306333, EPI_ISL_2306752, EPI_ISL_2306814, EPI_ISL_2306873 | Laboratory Corporation of America | Centers for Disease Control and Prevention Division of Viral Diseases, Pathogen Discovery | Dakota Howard, Dhwani Batra, Peter W. Cook, Kara Moser, Adrian Paskey, Jason Caravas, Benjamin Rambo-Martin, Shatavia Morrison, Christopher Gulvick, Scott Sammons, Yvette Unoarumhi, Darlene Wagner, Matthew Schmerer, Minoo Agarwal, Eyad Almasri, Debbie Boles, Ayla Burns, Nuthawin Charoensri, Oren Cohen, Susan Countryman, Mary Ann Cristobal, Bobbi Croy, Suzanne Dale, Hrushikesh Deshmukh, Amanda Douglas, Vincent Drouillon, Marcia Eisenberg, Howard Engler, Rama Ghatti, Prashant Gupta, Susan Hicks, Jake Humphrey, Lax Iyer, Manoj Jain, Mohan Kolli, Brian Krueger, Tim Kuphal, Stanley Letovsky, Michael Levandoski, Craig Lukasik, Jonathan Meltzer, Brian Norvell, Mindy Nye, Scott Parker, Christos Petropoulos, John Pruitt, Steven Ragan, Scott Ryan, Mike Sapeta, Jana Schroth, Suresh Babu Selvaraju, Goran Stevovic, Amanda Suchanek, Andrea Throop, Lyndon Tilson, Thomas Urban, Joe Voshell, Kimberly Wagner, Jonathan Williams, Mary Williamson, Qian Zeng, Tricia Zwiefelhofer, Clinton R. Paden, Duncan MacCannell |
| EPI_ISL_2308371 | Nebraska Public Health Laboratory | NPHL COVID-19 Response Team | NPHL COVID-19 Response Team |
| EPI_ISL_2313084 | Department of Virology | Department of Virology | Massab Umair, Aamer Ikram, Muhammad Salman, Nazish Badar, Zaira Rehman, Muhammad Ammar, Adnan Haider |
| EPI_ISL_2313118, EPI_ISL_2313181 | Limbach - MVZ Labor Eveld & Kollegen Essen | Robert Koch Institute | unknown |
| EPI_ISL_2313339 | Robert Koch-Institut ZBS1 | Robert Koch Institute | Annika Brinkmann |
| EPI_ISL_2313398 | Bioscientia MVZ Labor Karlsruhe GmbH | Robert Koch Institute | unknown |

| | | | |
|---|---|---|---|
| EPI_ISL_2313849 | Labor 28 MVZ GmbH | Robert Koch Institute | unknown |
| EPI_ISL_2313893 | Laborarztpraxis Osnabrück | Robert Koch Institute | unknown |
| EPI_ISL_2314139 | LabKom - Labor Hannover MVZ GmbH | Robert Koch Institute | unknown |
| EPI_ISL_2314141 | Bioscientia MVZ Labor Karlsruhe GmbH | Robert Koch Institute | unknown |
| EPI_ISL_2314185 | Department of Virology | Department of Virology | Massab Umair, Aamer Ikram, Muhammad Salman, Nazish Badar, Zaira Rehman, Muhammad Ammar, Adnan Haider |
| EPI_ISL_2314239, EPI_ISL_2314251 | Sonic - Labor Dr. von Froreich GmbH | Robert Koch Institute | unknown |
| EPI_ISL_2314738 | MVZ für Laboratoriumsmedizin und Mikrobiologie Koblenz-Mittelrhein (Labor Koblenz) | Robert Koch Institute | unknown |
| EPI_ISL_2314753 | Labor Prof. Dr. G. Enders MVZ GbR | Robert Koch Institute | unknown |
| EPI_ISL_2314773 | Limbach - MVZ Labor Ludwigsburg | Robert Koch Institute | unknown |
| EPI_ISL_2315020 | Diagnosticum - Labor Neukirchen | Robert Koch Institute | unknown |
| EPI_ISL_2315141, EPI_ISL_2315167, EPI_ISL_2315218 | SYNLAB MVZ Leverkusen | Robert Koch Institute | unknown |
| EPI_ISL_2316262 | SYNLAB MVZ Leinfelden-Echterdingen | Robert Koch Institute | unknown |
| EPI_ISL_2316615, EPI_ISL_2316635 | Klinikum Ernst von Bergmann gemeinnützige GmbH - stationärer Bereich | Robert Koch Institute | unknown |
| EPI_ISL_2320451 | Helix/Illumina | Centers for Disease Control and Prevention Division of Viral Diseases, Pathogen Discovery | Dakota Howard, Dhwani Batra, Peter W. Cook, Kara Moser, Adrian Paskey, Jason Caravas, Benjamin Rambo-Martin, Shatavia Morrison, Christopher Gulvick, Scott Sammons, Yvette Unoarumhi, Darlene Wagner, Matthew Schmerer, Eileen de Feo, Jan Antico, Christine Tran, Matthew Tolentino, Shannon Wickline, Kim Gietzen, Brad Sickler, Jingtao Liu, Eric Allen, Phil Febbo, Nicole L. Washington, Simon White, Geraint Levan, Kelly Schiabor Barrett, Elizabeth Cirulli, Alexandre Bolze, Ary Ascencio, Charlotte Rivera-Garcia, Ryan Cho, Jason Nguyen, Sherry Wang, Jimmy Ramirez, Tyler Cassens, Efren Sandoval, Magnus Isaksson, William Lee, David Becker, Marc Laurent, James Lu, Clinton R. Paden, Duncan MacCannell |
| EPI_ISL_2323153 | Nebraska Public Health Laboratory | NPHL COVID-19 Response Team | NPHL COVID-19 Response Team |
| EPI_ISL_2325091 | Mayo Clinic & Mayo Clinic Laboratories | Minnesota Department of Health, Public Health Laboratory | Alexandra Lorentz, Jacob Garfin, Matt Plumb, and Xiong Wang |
| EPI_ISL_2332810 | OLVZ Aalst | OLVZ Aalst | Anne Vankeerberghen |
| EPI_ISL_2332879 | UW Virology Lab | UW Virology Lab | Pavitra Roychoudhury, Hong Xie, Lasata Shrestha, Tien V. Nguyen, Shah Mohamed Bakhash, Michelle Lin, Noah R. Baker, Ricardo Perez, Sean Ellis, Nathan Breit, Robert J. Livingston, Meei-Li Huang, Keith R Jerome, Patrick Mathias, Alexander Greninger |
| EPI_ISL_2333707 | CH.INTERCOMMUNAL DE CRETEIL | Department of Virology, Henri Mondor University Hospital, Assistance Publique Hôpitaux de Paris, Université Paris-Est Créteil, INSERM U955 | Christophe Rodriguez, Slim Fourati, Vanessa Demontant, Guillaume Gricourt, Melissa N'Debi, Alexandre Soulier, Elisabeth Trawinski, Jean-Michel Pawlotsky |
| EPI_ISL_2333760, EPI_ISL_2333761, EPI_ISL_2333762 | Hôpital Avicenne | Department of Virology, Henri Mondor University Hospital, Assistance Publique Hôpitaux de Paris, Université Paris-Est Créteil, INSERM U955 | Christophe Rodriguez, Slim Fourati, Vanessa Demontant, Guillaume Gricourt, Melissa N'Debi, Alexandre Soulier, Elisabeth Trawinski, Jean-Michel Pawlotsky |
| EPI_ISL_2333806 | GH A.CHENEVIER-H.MONDOR | Department of Virology, Henri Mondor University Hospital, Assistance Publique Hôpitaux de Paris, Université Paris-Est Créteil, INSERM U955 | Christophe Rodriguez, Slim Fourati, Vanessa Demontant, Guillaume Gricourt, Melissa N'Debi, Alexandre Soulier, Elisabeth Trawinski, Jean-Michel Pawlotsky |
| EPI_ISL_2339296 | Arizona State University | Arizona State University | Peter T. Skidmore, LaRinda A. Holland, Matthew F. Smith, Rabia Maqsood, Nicholas J. Mellor, Joy M. Blain, Valerie Harris, Joshua LaBaer, Vel Murugan, Efrem S. Lim |
| EPI_ISL_2339612 | UMC Groningen, Clinical Virology, Department of Medical Microbiology and Infection Prevention | UMC Groningen, Clinical Virology, Department of Medical Microbiology and Infection Prevention | Hubert Niesters, Alexander Friedrich, Erley Lizarazo-Forero, Monika Fliss, Lilli Gard, Sigrid Rosema, Coretta Van Leer-Buter, Xuewei Zhou, Marjolein Knoester |
| EPI_ISL_2340666 | University of Wisconsin-Madison AIDS Vaccine Research Laboratories | University of Wisconsin-Madison AIDS Vaccine Research Laboratories | Gage Moreno, Katarina Braun, et al. AIDS Vaccine Research Laboratories |
| EPI_ISL_2347125, EPI_ISL_2347237 | Lighthouse Lab in Milton Keynes | Wellcome Sanger Institute for the COVID-19 Genomics UK (COG-UK) Consortium | The Lighthouse Lab in Milton Keynes and Alex Alderton, Roberto Amato, Jeffrey Barrett, Sonia Goncalves, Ewan Harrison, David K. Jackson, Ian Johnston, Dominic Kwiatkowski, Cordelia Langford, John Sillitoe on behalf of the Wellcome Sanger Institute COVID-19 Surveillance Team |
| EPI_ISL_2348566 | Department of Clinical Microbiology | GIGA Medical Genomics | Keith Durkin, Maria Artesi, Sébastien Bontems, Raphaël Boreux, Bouchra Boujemla, Nathalie Renotte, Cécile Meex, Pierrette Melin, Marie-Pierre Hayette, Vincent Bours |
| EPI_ISL_2348808, EPI_ISL_2348809, EPI_ISL_2348810 | Faculty of Medicine, Prince of Songkla University | COVID-19 Network Investigations (CONI) Alliance | Elizabeth Batty, Wasun Chantratita, Thanat Chookajorn, Stefan Fernandez, Angkana Huang, Anthony R. Jones, Khajohn Joonlasak, Chonticha Klungtong, Theerarat Kochakarn, Namfon Kotanan, Krittikorn Kumpornsin, Duangkamon Loesbanluechai, Wudtichai Manasatienkij, Bhakbhoom Panthan, Ekawat Pasomsub, Kingkan Rakmanee, Insee Sensorn, Janjira Thaipadungpanit, Arporn Wangwiwatsin, Treewat Watthanachockchai, Smonrapat Surasombatpattana |
| EPI_ISL_2348820 | Office of Diseases Prevention and Control Region 12 Songkhla | COVID-19 Network Investigations (CONI) Alliance | Elizabeth Batty, Wasun Chantratita, Thanat Chookajorn, Stefan Fernandez, Angkana Huang, Anthony R. Jones, Khajohn Joonlasak, Chonticha Klungtong, Theerarat Kochakarn, Namfon Kotanan, Krittikorn Kumpornsin, Duangkamon Loesbanluechai, Wudtichai Manasatienkij, Bhakbhoom Panthan, Ekawat Pasomsub, Kingkan Rakmanee, Insee Sensorn, Janjira Thaipadungpanit, Arporn Wangwiwatsin, Treewat Watthanachockchai, Sawanya Chantutanon |
| EPI_ISL_2348930 | Oregon State Public Health Laboratory | Oregon State Public Health Laboratory | Rafia Razzaque, Eugene Yeboah, Vanda Makris, Laura Tsaknaridis, John Fontana and Shane Sevey |
| EPI_ISL_2349834 | National Public Health Laboratory, National Centre for Infectious Diseases | National Public Health Laboratory, National Centre for Infectious Diseases | Tze Minn Mak, Zhenyang Zhou, Royce Ang, Lin Cui, Raymond Tzer Pin Lin |
| EPI_ISL_2350263 | Clinical Microbiology, Infection Prevention and Control | Section for Molecular Diagnostics | Björn Hallström, Jonas Björkman |
| EPI_ISL_2352955 | Lighthouse Lab in Milton Keynes | Wellcome Sanger Institute for the COVID-19 Genomics UK (COG-UK) Consortium | The Lighthouse Lab in Milton Keynes and Alex Alderton, Roberto Amato, Jeffrey Barrett, Sonia Goncalves, Ewan Harrison, David K. Jackson, Ian Johnston, Dominic Kwiatkowski, Cordelia Langford, John Sillitoe on behalf of the Wellcome Sanger Institute COVID-19 Surveillance Team |
| EPI_ISL_2361525 | CHU LILLE | CHU Lille - Laboratoire de Virologie | AIT YAHYA Emilie, ALIDJINOU Enagnon Kazali, BOCKET Laurence, CREPIN Michel, DEMAY Christophe, ENGELMANN Ilka, GEFFROY Sandrine, GUIGON Aurélie, LAMBERT Valérie, LAZREK Mouna, NOBILLIAUX Florian, PREVOST Brigitte, THUILLIER Caroline, TINEZ Claire ,TCHANTCHOU NJOSSE YANICK |
| EPI_ISL_2361527 | CH ROUBAIX | CHU Lille - Laboratoire de Virologie | AIT YAHYA Emilie, ALIDJINOU Enagnon Kazali, BOCKET Laurence, CREPIN Michel, DEMAY Christophe, ENGELMANN Ilka, GEFFROY Sandrine, GUIGON Aurélie, LAMBERT Valérie, LAZREK Mouna, NOBILLIAUX Florian, PREVOST Brigitte, THUILLIER Caroline, TINEZ Claire ,TCHANTCHOU NJOSSE YANICK |
| EPI_ISL_2361628 | Viollier AG | Viollier AG | Andrea Patrizia Salzmann, Henriette Kurth, Christiane Beckmann, Maurice Redondo, Olivier Kobel, Christoph Noppen |
| EPI_ISL_2362786 | Lab voor klinische biologie | Lab voor klinische biologie | Marija Janevska, Hannelore Hamerlinck, Bruno Verhasselt |
| EPI_ISL_2366297 | Lighthouse Lab in Milton Keynes | Wellcome Sanger Institute for the COVID-19 Genomics UK (COG-UK) Consortium | The Lighthouse Lab in Milton Keynes and Alex Alderton, Roberto Amato, Jeffrey Barrett, Sonia Goncalves, Ewan Harrison, David K. Jackson, Ian Johnston, Dominic Kwiatkowski, Cordelia Langford, John Sillitoe on behalf of the Wellcome Sanger Institute COVID-19 Surveillance Team |
| EPI_ISL_2367762 | CH BETHUNE | CHU Lille - Laboratoire de Virologie | AIT YAHYA Emilie, ALIDJINOU Enagnon Kazali, BOCKET Laurence, CREPIN Michel, DEMAY Christophe, ENGELMANN Ilka, GEFFROY Sandrine, GUIGON Aurélie, LAMBERT Valérie, LAZREK Mouna, NOBILLIAUX Florian, PREVOST Brigitte, THUILLIER Caroline, TINEZ Claire ,TCHANTCHOU NJOSSE YANICK |
| EPI_ISL_2368377 | Aegis Sciences Corporation | Centers for Disease Control and Prevention Division of Viral Diseases, Pathogen Discovery | Dakota Howard, Dhwani Batra, Peter W. Cook, Kara Moser, Adrian Paskey, Jason Caravas, Benjamin Rambo-Martin, Shatavia Morrison, Christopher Gulvick, Scott Sammons, Yvette Unoarumhi, Darlene Wagner, Matthew Schmerer, Cyndi Clark, Patrick Campbell, Rob Case, Vikramsinha Ghorpade, Holly Houdeshell, |

| | | | |
|---|---|---|---|
| | | | Ola Kvalvaag, Dillon Nall, Ethan Sanders, Alec Vest, Shaun Westlund, Matthew Hardison, Clinton R. Paden, Duncan MacCannell |
| EPI_ISL_2372701 | Microbiologia CATLAB | Can Ruti SARS-CoV-2 Sequencing Hub (HUGTiP/IrsiCaixa/IGTP) | Marc Noguera-Julian, Pilar Armengol, Ignacio Blanco, Antoni E Bordoy, Francesc Catala-Moll, Pere-Joan Cardona, Maria Casadellà, Cristina Casañ, Gemma Clara, Bonaventura Clotet, Cristina Esteban, Montserrat Giménez, Mercedes Guerrero, Anna Not, Roger Pared |
| EPI_ISL_2373699 | CNR Virus des Infections Respiratoires - France SUD | CNR Virus des Infections Respiratoires - France SUD | Antonin Bal, Gregory Destras, Gwendolyne Burfin, Hadrien Regue, Quentin Semanas, Martine Valette, Bruno Lina, Laurence Josset |
| EPI_ISL_2373713 | LABORATOIRE DE BIOLOGIE MEDICALE | CNR Virus des Infections Respiratoires - France SUD | Antonin Bal, Gregory Destras, Gwendolyne Burfin, Hadrien Regue, Quentin Semanas, Martine Valette, Bruno Lina, Laurence Josset |
| EPI_ISL_2373755, EPI_ISL_2373796 | CNR Virus des Infections Respiratoires - France SUD | CNR Virus des Infections Respiratoires - France SUD | Antonin Bal, Gregory Destras, Gwendolyne Burfin, Hadrien Regue, Quentin Semanas, Martine Valette, Bruno Lina, Laurence Josset |
| EPI_ISL_2373822, EPI_ISL_2373823 | REUNILAB | CNR Virus des Infections Respiratoires - France SUD | Antonin Bal, Gregory Destras, Gwendolyne Burfin, Hadrien Regue, Quentin Semanas, Martine Valette, Bruno Lina, Laurence Josset |
| EPI_ISL_2373858 | LBM UNIBIO VALENTIN | CNR Virus des Infections Respiratoires - France SUD | Antonin Bal, Gregory Destras, Gwendolyne Burfin, Hadrien Regue, Quentin Semanas, Martine Valette, Bruno Lina, Laurence Josset |
| EPI_ISL_2373867 | LBM TRONQUIERES | CNR Virus des Infections Respiratoires - France SUD | Antonin Bal, Gregory Destras, Gwendolyne Burfin, Hadrien Regue, Quentin Semanas, Martine Valette, Bruno Lina, Laurence Josset |
| EPI_ISL_2373877 | LBM UNIBIO VALENTIN | CNR Virus des Infections Respiratoires - France SUD | Antonin Bal, Gregory Destras, Gwendolyne Burfin, Hadrien Regue, Quentin Semanas, Martine Valette, Bruno Lina, Laurence Josset |
| EPI_ISL_2373952 | CNR Virus des Infections Respiratoires - France SUD | CNR Virus des Infections Respiratoires - France SUD | Antonin Bal, Gregory Destras, Gwendolyne Burfin, Hadrien Regue, Quentin Semanas, Martine Valette, Bruno Lina, Laurence Josset |
| EPI_ISL_2374957 | Viollier AG | Department of Biosystems Science and Engineering, ETH Zürich | Christian Beisel, Sarah Nadeau, Chaoran Chen, Ivan Topolsky, Philipp Jablonski, Lara Fuhrmann, David Dreifuss, Katharina Jahn, Rebecca Denes, Mirjam Feldkamp, Ina Nissen, Natascha Santacroce, Elodie Burcklen, Christiane Beckmann, Maurice Redondo, Olivier Kobel, Christoph Noppen, Sophie Seidel, Noemie Santamaria de Souza, Niko Beerenwinkel, Tanja Stadler |
| EPI_ISL_2375390 | Viollier AG | Department of Biosystems Science and Engineering, ETH Zürich | Chaoran Chen, Sarah Nadeau, Catharine Aquino, Ivan Topolsky, Philipp Jablonski, Lara Fuhrmann, David Dreifuss, Katharina Jahn, Daniel Ehrsam, Isabel Stürmer, Andreia Cabral de Gouvea, Maria Domenica Moccia, Simon Grüter, Timothy Sykes, Lennart Opitz, Griffin White, Laura Neff, Doris Popovic, Andrea Patrignani, Jay Tracy, Ralph Schlapbach, Christiane Beckmann, Maurice Redondo, Olivier Kobel, Christoph Noppen, Sophie Seidel, Noemie Santamaria de Souza, Niko Beerenwinkel, Tanja Stadler |
| EPI_ISL_2375591 | Infinity Biologix | Centers for Disease Control and Prevention Division of Viral Diseases, Pathogen Discovery | Dakota Howard, Dhwani Batra, Peter W. Cook, Kara Moser, Adrian Paskey, Jason Caravas, Benjamin Rambo-Martin, Shatavia Morrison, Christopher Gulvick, Scott Sammons, Yvette Unoarumhi, Darlene Wagner, Matthew Schmerer, Christian Bixby, Yihe Wang, Jonathan Schultz, Chirayu Goswami, Russ Hager, Robin Grimwood, Clinton R. Paden, Duncan MacCannell |
| EPI_ISL_2376117 | NJDOH, Public Health and Environmental Laboratories | NJ_PHEL | Lindsey Bodnar, Shiv K. Verma, Jacquelyn Deverell, Dana Woell, Allison Roder, Byeong Jeong |
| EPI_ISL_2376256 | Gundersen Clinical Microbiology Laboratory | Kabara Cancer Research Institute | Craig S. Richmond, Paraic A. Kenny |
| EPI_ISL_2376587 | M Health Fairview | Minnesota Department of Health, Public Health Laboratory | Alexandra Lorentz, Jacob Garfin, Matt Plumb, and Xiong Wang |
| EPI_ISL_2378708 | Department of Health Technology and Informatics, The Hong Kong Polytechnic University | Department of Health Technology and Informatics, The Hong Kong Polytechnic University | Cheng,V.C.-C., Siu,G.K.-H., Wong,S.-C., Chen,H., Lee,L.-K., Leung,J.S.-L., Lu,K.K., Chan,C.T.-M., Lo,H.W.-H., Leung,K.S.-S., Wong,E.Y.-K., Luk,S., Ng,T.T.-L., Jim,H.-C., Lao,H.-Y., Wong,D.S.-H., Tam,K.K.-G., Mok,K.K.-S., Wong,K.N., Yeh,E.Y.-W., Lam,B.P.-H., Lam,J.Y.-W., Wu,A.K.-L., Yau,M.C.-Y., Lai,Y.W.-M., Ho,A.Y.-M., Leung,W.-S., Chan,M.-C., Lam,B.H.-S., To,W.-K., Lee,R.A., Lung,D.C., Tse,H., To,K.K.-W., Yuen,K.-Y. |
| EPI_ISL_2378890 | Washington State Department of Health Public Health Laboratories | Washington State Department of Health Public Health Laboratories | Drew MacKellar, Philip Dykema, Denny Russell, Joenice Gonzalez, Hannah Gray, Geoff Melly, Vanessa De Los Santos, Darren Lucas, JohnAric Peterson, Avi Singh, Rebecca Cao |
| EPI_ISL_2379302, EPI_ISL_2379426 | National Platform bis UMONS/Jolimont | National Platform bis UMONS/Jolimont | François Dufrasne, Guillaume Bayon-Vicente, Florian Juszczak, Gautier Detry, Ruddy Wattiez |
| EPI_ISL_2382840, EPI_ISL_2382855 | University of Liège COVID-19 testing center | GIGA Medical Genomics | Keith Durkin, Maria Artesi, Bouchra Boujemla, Nathalie Renotte, Cécile Meex, Sébastien Bontems, Fabrice Bureau, Laurent Gillet, Wouter Coppieters, Marie-Pierre Hayette, Vincent Bours |
| EPI_ISL_2382899 | Jessa | Jessa | Raymaekers et al. on behalf of Jessa_cmdLab |
| EPI_ISL_2383950 | SC Dept of Health and Env. Control-Bureau of Laboratories | Centers for Disease Control and Prevention Division of Viral Diseases, Pathogen Discovery | Mili Sheth, Sarah Nobles, Jasmine Padilla, Mark Burroughs, Shoshona Le, Katie Dillon, Peter Cook, Clinton R. Paden, Dhwani Batra, Krista Queen, Kristen Knipe, Dakota Howard, Yvette Unoarumhi, Darlene Wagner, Matthew Schmerer, Ben L. Rambo-Martin, Kristine Lacek, Sam Shepard, Alison Laufer Halpin, Dave Wentworth, Vivien Dugan, Suxiang Tong, Justin Lee |
| EPI_ISL_2386188 | MVZ für Laboratoriumsmedizin und Mikrobiologie Koblenz-Mittelrhein (Labor Koblenz) | Robert Koch Institute | unknown |
| EPI_ISL_2386311 | Synlab MVZ Augsburg | Robert Koch Institute | unknown |
| EPI_ISL_2386546 | Labor Dr. Wisplinghoff - Köln | Robert Koch Institute | unknown |
| EPI_ISL_2386796 | Labor ZOTZ\|KLIMAS; MVZ Düsseldorf-Centrum | Robert Koch Institute | unknown |
| EPI_ISL_2388210 | Laborarztpraxis Osnabrück | Robert Koch Institute | unknown |
| EPI_ISL_2388317 | LabKom - Labor Hannover MVZ GmbH | Robert Koch Institute | unknown |
| EPI_ISL_2388328 | LabKom - Labor Augsburg MVZ GmbH | Robert Koch Institute | unknown |
| EPI_ISL_2388569 | Sonic - MVZ Medizinisches Labor Bremen GmbH | Robert Koch Institute | unknown |
| EPI_ISL_2388597, EPI_ISL_2388615 | LabKom - Labor Augsburg MVZ GmbH | Robert Koch Institute | unknown |
| EPI_ISL_2388918 | Bioscientia Labor Wermsdorf | Robert Koch Institute | unknown |
| EPI_ISL_2388957 | Sonic - Bioscientia - MVZ Labor Saar GmbH | Robert Koch Institute | unknown |
| EPI_ISL_2389057 | Bioscientia Labor Wermsdorf | Robert Koch Institute | unknown |
| EPI_ISL_2389351 | Labor Dr. Heidrich & Kollegen MVZ GmbH Hamburg | Robert Koch Institute | unknown |
| EPI_ISL_2389582 | SYNLAB MVZ Berlin | Robert Koch Institute | unknown |
| EPI_ISL_2389619 | SYNLAB MVZ Leverkusen | Robert Koch Institute | unknown |
| EPI_ISL_2389744 | SYNLAB MVZ Trier | Robert Koch Institute | unknown |
| EPI_ISL_2389848 | SYNLAB MVZ Heidelberg | Robert Koch Institute | unknown |
| EPI_ISL_2390144 | MVZ Labor Dr. Fenner und Kollegen (Standort Hamburg) | Robert Koch Institute | unknown |
| EPI_ISL_2392826, EPI_ISL_2395820 | Lighthouse Lab in Milton Keynes | Wellcome Sanger Institute for the COVID-19 Genomics UK (COG-UK) Consortium | The Lighthouse Lab in Milton Keynes and Alex Alderton, Roberto Amato, Jeffrey Barrett, Sonia Goncalves, Ewan Harrison, David K. Jackson, Ian Johnston, Dominic Kwiatkowski, Cordelia Langford, John Sillitoe on behalf of the Wellcome Sanger Institute COVID-19 Surveillance Team |
| EPI_ISL_2398631, EPI_ISL_2398802, EPI_ISL_2398951, EPI_ISL_2398994, EPI_ISL_2399066, EPI_ISL_2399251, EPI_ISL_2399285 | Laboratory Corporation of America | Centers for Disease Control and Prevention Division of Viral Diseases, Pathogen Discovery | Dakota Howard, Dhwani Batra, Peter W. Cook, Kara Moser, Adrian Paskey, Jason Caravas, Benjamin Rambo-Martin, Shatavia Morrison, Christopher Gulvick, Scott Sammons, Yvette Unoarumhi, Darlene Wagner, Matthew Schmerer, Minoo Agarwal, Eyad Almasri, Debbie Boles, Ayla Burns, Nuthawin Charoensri, Oren Cohen, Susan Countryman, Mary Ann Cristobal, Bobbi Croy, Suzanne Dale, Hrushikesh Deshmukh, Amanda Douglas, Vincent Drouillon, Marcia Eisenberg, Howard Engler, Rama Ghatti, Prashant Gupta, Susan Hicks, Jake Humphrey, Lax Iyer, Lisa Pfefferle, Manoj Jain, Matthew Robinson, Mohan Kolli, Brian Krueger, Tim Kuphal, Stanley Letovsky, Michael Levandoski, Craig Lukasik, Jonathan Meltzer, Brian Norvell, Mindy Nye, Scott Parker, Christos Petropoulos, John Pruitt, Steven Ragan, Scott Ryan, Mike Sapeta, Jana Schroth, Suresh Babu Selvaraju, Goran Stevovic, Amanda Suchanek, Andrea Throop, Lyndon Tilson, Thomas Urban, Joe Voshell, Kimberly Wagner, Jonathan Williams, Mary Williamson, Qian Zeng, Tricia Zwiefelhofer, Clinton R. Paden, Duncan MacCannell |
| EPI_ISL_2400368, EPI_ISL_2400369 | University of Michigan Clinical Microbiology Laboratory | Lauring Lab, University of Michigan, Department of Microbiology and Immunology | Gilbert |

| | | | |
|---|---|---|---|
| EPI_ISL_2400808 | Laboratoires d'analyses medicales - Ketterthill | Laboratoire national de sante, Microbiology, Microbial Genomics Platform | Anke Wienecke-Baldacchino, Catherine Ragimbeau,Jessica Tapp, Fatu Djabi, Lise Pignon, Raoul Salmon, Serge Vedy, Caroline Scheiber, Tamir Abdelrahman |
| EPI_ISL_2401594, EPI_ISL_2401595, EPI_ISL_2401596, EPI_ISL_2401670, EPI_ISL_2401706, EPI_ISL_2401733, EPI_ISL_2401740 | BioneXt Lab | Laboratoire national de sante, Microbiology, Microbial Genomics Platform | Anke Wienecke-Baldacchino, Catherine Ragimbeau,Jessica Tapp, Fatu Djabi, Lise Pignon, Raoul Salmon, Thibault Ferrandon, Tamir Abdelrahman |
| EPI_ISL_2401864 | Hospital Center Emile Mayrisch | Laboratoire national de sante, Microbiology, Microbial Genomics Platform | Anke Wienecke-Baldacchino, Catherine Ragimbeau,Jessica Tapp, Fatu Djabi, Lise Pignon, Raoul Salmon, Cynthia Oxacelay, Tamir Abdelrahman |
| EPI_ISL_2402615 | Centracare Laboratory Services | Minnesota Department of Health, Public Health Laboratory | Alexandra Lorentz, Jacob Garfin, Matt Plumb, and Xiong Wang |
| EPI_ISL_2403347, EPI_ISL_2403353, EPI_ISL_2403354 | Ministry of Health Turkey | Ministry of Health Turkey | Fatma Bayrakdar, Yasemin Cosgun, Suleyman Yalcin, Gulay Korukluoglu |
| EPI_ISL_2406058, EPI_ISL_2406090 | Dutch COVID-19 response team | National Institute for Public Health and the Environment (RIVM) | Adam Meijer, Harry Vennema, Dirk Eggink, Jeroen Cremer, Sharon van den Brink, Bas van der Veer, AnneMarie van den Brandt, Lisa Wijsman, Kim Freriks, Ryanne Jaarsma, Euníce Then, Lynn Aarts, Sanne Bos, Melissa van Tuil, Linda van de Nes, Sjoerd Kuiling, James Groot, Florian Zwagemaker, Dennis Schmitz, Annelies Kroneman, Karim Hajji, Chantal Reusken, on behalf of the national COVID-19 response team |
| EPI_ISL_2406492 | Middlemore Hospital | Institute of Environmental Science and Research (ESR) | Rachel Boyle, SallyAnn Harbison, Olivia Stroeven, Xiaoyun Ren, Matt Storey, Nikki Freed, Muhammad Faisal, Jing Wang, Hermes Perez, Anja Werno, Antje van der Linden, Arlo Upton, Chris Mansell, David Hammer, Dragana Drinkovic, Gary McAuliffe, Hana Sofia Andersson, James Ussher, Jill Sherwood, Josh Freeman, Julia Howard, Juliet Elvy, Mary DeAlmeida, Matt Blakiston, Matthew Rogers, Max Bloomfield, Michael Addidle, Michelle Balm, Sally Roberts, Sarah Jefferies, Sharmini Muttaiyah, Susan Morpeth, Susan Taylor, Timothy Blackmore, Vani Sathyendran, Veronica Playle, Virginia Hope, Erasmus Smit, Lauren Jelly, Olin Silander, Joep de Ligt |
| EPI_ISL_2406499, EPI_ISL_2406502 | Laboratoire Virologie Saint Louis APHP | Laboratoire Virologie Saint Louis APHP | Maud Salmona, Marie Laure Chaix, Severine Mercier Delarue, Marie Laure Néré, Linda Feghoul, Jérome Le Goff, Constance Delauguerre, Sophia Achaibou |
| EPI_ISL_2408141, EPI_ISL_2408194 | Swedish national genomic surveillance program of SARS-CoV-2 | The Public Health Agency of Sweden | Maximilian Riess, Maria Lind Karlberg, Alma Brolund, Swedish national genomic surveillance program of SARS-CoV-2 |
| EPI_ISL_2408469 | Labor Team W Ag | Clinical Bacteriology | Tim Roloff, Fanny Wegner, Helena MB Seth-Smith, Alfredo Mari, Karoline Leuzinger, Julia Bielicki, Manuel Battegay, Hans Hirsch, Adrian Egli |
| EPI_ISL_2414909, EPI_ISL_2415495, EPI_ISL_2415508, EPI_ISL_2415699 | Swedish national genomic surveillance program of SARS-CoV-2 | The Public Health Agency of Sweden | Maximilian Riess, Maria Lind Karlberg, Alma Brolund, Swedish national genomic surveillance program of SARS-CoV-2 |
| EPI_ISL_2417501 | Department of Bacteria, Parasites and Fungi, Statens Serum Institut, Copenhagen, Denmark | Statens Serum Institut Bioinformatics and Microbial Genomics | Danish Covid-19 Genome Consortium |
| EPI_ISL_2418035, EPI_ISL_2418047, EPI_ISL_2418218, EPI_ISL_2418225, EPI_ISL_2418291, EPI_ISL_2418292, EPI_ISL_2418301, EPI_ISL_2418309, EPI_ISL_2418324, EPI_ISL_2418576, EPI_ISL_2418792, EPI_ISL_2418816, EPI_ISL_2418824, EPI_ISL_2418865, EPI_ISL_2418875, EPI_ISL_2418881, EPI_ISL_2418887, EPI_ISL_2418888, EPI_ISL_2418897, EPI_ISL_2418903, EPI_ISL_2418905, EPI_ISL_2418916, EPI_ISL_2418935, EPI_ISL_2418936, EPI_ISL_2418968, EPI_ISL_2419549 | | | |
| see above | Swedish national genomic surveillance program of SARS-CoV-2 | The Public Health Agency of Sweden | Maximilian Riess, Maria Lind Karlberg, Alma Brolund, Swedish national genomic surveillance program of SARS-CoV-2 |
| EPI_ISL_2420697, EPI_ISL_2420700, EPI_ISL_2420701 | Viollier AG | Viollier AG | Andrea Patrizia Salzmann, Henriette Kurth, Christiane Beckmann, Maurice Redondo, Olivier Kobel, Christoph Noppen |
| EPI_ISL_2422284, EPI_ISL_2422287, EPI_ISL_2422291, EPI_ISL_2422310 | SUNY UPSTATE MEDICAL UNIVERSITY | Wadsworth Center, New York State Department of Health | Kirsten St. George, Daryl M. Lamson, Alexis Russell, Matthew Shudt, Melissa A Leisner, Jonathan Plitnick, Catharine Prussing, Navjot Singh, John Kelly, Erasmus Schneider, Erica Lasek-Nesselquist |
| EPI_ISL_2422357 | URMC LABS | Wadsworth Center, New York State Department of Health | Kirsten St. George, Daryl M. Lamson, Alexis Russell, Matthew Shudt, Melissa A Leisner, Jonathan Plitnick, Catharine Prussing, Navjot Singh, John Kelly, Erasmus Schneider, Erica Lasek-Nesselquist |
| EPI_ISL_2424246 | Oregon State Public Health Laboratory | Oregon State Public Health Laboratory | Rafia Razzaque, Eugene Yeboah, Vanda Makris, Laura Tsaknaridis, John Fontana and Shane Sevey |
| EPI_ISL_2425456, EPI_ISL_2425469 | Virginia Division of Consolidated Laboratory Services | Virginia Division of Consolidated Laboratory Services | Virginia DCLS |
| EPI_ISL_2426477, EPI_ISL_2426495, EPI_ISL_2426505, EPI_ISL_2426555 | UW Virology Lab | UW Virology Lab | Pavitra Roychoudhury, Hong Xie, Lasata Shrestha, Tien V. Nguyen, Shah Mohamed Bakhash, Michelle Lin, Noah R. Baker, Sean Ellis, Meei-Li Huang, Keith R Jerome, Alexander Greninger |
| EPI_ISL_2426579, EPI_ISL_2426610 | SYNLAB | GIGA Medical Genomics | Keith Durkin, Maria Artesi, Sébastien Bontems, Raphaël Boreux, Bouchra Boujemla, Nathalie Renotte, Cécile Meex, Pierrette Melin, Marie-Pierre Hayette, Vincent Bours |
| EPI_ISL_2426657, EPI_ISL_2426658 | Servicio de Microbiologia Hospital Ramon y Cajal | Servicio de Microbiologia Hospital Ramon y Cajal | Ponce M, Galan JC, Martinez L. Abreu M, y Gonzalez-Alba JM |
| EPI_ISL_2429049 | Clinical Microbiology, Infection Prevention and Control | Section for Molecular Diagnostics | Björn Hallström, Jonas Björkman |
| EPI_ISL_2431800 | Department of Bacteria, Parasites and Fungi, Statens Serum Institut, Copenhagen, Denmark | Statens Serum Institut Bioinformatics and Microbial Genomics | Danish Covid-19 Genome Consortium |
| EPI_ISL_2432662 | UW Virology Lab | UW Virology Lab | Pavitra Roychoudhury, Hong Xie, Lasata Shrestha, Tien V. Nguyen, Shah Mohamed Bakhash, Michelle Lin, Noah R. Baker, Ricardo Perez, Sean Ellis, Nathan Breit, Robert J. Livingston, Meei-Li Huang, Keith R Jerome, Patrick Mathias, Alexander Greninger |
| EPI_ISL_2433565, EPI_ISL_2433566, EPI_ISL_2433567, EPI_ISL_2433568, EPI_ISL_2433569, EPI_ISL_2433570, EPI_ISL_2433571 | Naradhiwas Rajanagarindra Hospital | Division of Genomic Medicine and Innovation support,Department of Medical Sciences, Ministry of Public Health, Thailand | Surakameth Mahasirimongkol,Nuanjun Wichukchinda,Archawin Rojanawiwat,Pilailuk Akkapaiboon Okada ,Waritta Sawaengdee,Penpitcha Thawong,Pundharika Piboonsiri,Jirapha Pakdee,Natthakul Bunneang |
| EPI_ISL_2434247 | Department of Virology | Department of Virology | Massab Umair, Aamer Ikram, Muhammad Salman, Nazish Badar, Zaira Rehman, Muhammad Ammar, Syed Adnan Haider |
| EPI_ISL_2438544 | Lighthouse Lab in Milton Keynes | Wellcome Sanger Institute for the COVID-19 Genomics UK (COG-UK) Consortium | The Lighthouse Lab in Milton Keynes and Alex Alderton, Roberto Amato, Jeffrey Barrett, Sonia Goncalves, Ewan Harrison, David K. Jackson, Ian Johnston, Dominic Kwiatkowski, Cordelia Langford, John Sillitoe on behalf of the Wellcome Sanger Institute COVID-19 Surveillance Team |
| EPI_ISL_2438793 | Dept. of Microbiology and Infection Control, Akershus University Hospital HF | Dept. of Microbiology and Infection Control, Akershus University Hospital HF | Hege Vangstein Aamot, Alexander Hesselberg Løvestad |
| EPI_ISL_2440109 | Helix/Illumina | Centers for Disease Control and Prevention Division of Viral Diseases, Pathogen Discovery | Dakota Howard, Dhwani Batra, Peter W. Cook, Kara Moser, Adrian Paskey, Jason Caravas, Benjamin Rambo-Martin, Shatavia Morrison, Christopher Gulvick, Scott Sammons, Yvette Unoarumhi, Darlene Wagner, Matthew Schmerer, Eileen de Feo, Jan Antico, Christine Tran, Matthew Tolentino, Shannon Wickline, Kim Gietzen, Brad Sickler, Jingtao Liu, Eric Allen, Phil Febbo, Nicole L. Washington, Simon White, Geraint Levan, Kelly Schiabor Barrett, Elizabeth Cirulli, Alexandre Bolze, Ary Ascencio, Charlotte Rivera-Garcia, Ryan Cho, Jason Nguyen, Sherry Wang, Jimmy Ramirez, Tyler Cassens, Efren Sandoval, Magnus Isaksson, William Lee, David Becker, Marc Laurent, James Lu, Clinton R. Paden, Duncan MacCannell |
| EPI_ISL_2442052 | AZDelta | AZ Delta Medical Laboratories in Roeselare, Belgium | Geert Martens, Dieter De Smet, Merijn Vanhee, on behalf of AZ Delta COVID-19 Genomics core (member of Genomic surveillance of SARS-CoV-2 in Belgium network) |
| EPI_ISL_2443143, EPI_ISL_2443194 | Kansas Health and Environmental Lab | Kansas Health and Environmental Lab | Katherine Wiggins, Mike Grose, Jonathan Barnell, Ben Olsen, and Phil Adam |
| EPI_ISL_2444388 | CH TOURCOING | CHU Lille - Laboratoire de Virologie | AIT YAHYA Emilie, ALIDJINOU Enagnon Kazali, BOCKET Laurence, CREPIN Michel, DEMAY Christophe, ENGELMANN Ilka, GEFFROY Sandrine, GUIGON Aurélie, LAMBERT Valérie, LAZREK Mouna, NOBILLIAUX Florian, PREVOST Brigitte, THUILLIER Caroline, TINEZ Claire |
| EPI_ISL_2447604, EPI_ISL_2447608, EPI_ISL_2447610, EPI_ISL_2447639, EPI_ISL_2447662 | SARS-CoV-2 testing team, National Institute of Infectious Diseases | Pathogen Genomics Center, National Institute of Infectious Diseases | Tsuyoshi Sekizuka, Kentaro Itokawa, Rina Tanaka, Masanori Hashino, Nozomu Hanaoka, Masumichi Saito, Naomi Nojiri, Hazuka Y Furihata, Sana Uchikoba, Tsuguto Fujimoto, Makoto Kuroda |

| | | | |
|---|---|---|---|
| EPI_ISL_2448024 | Microbiology Department, Laboratori Clínic Metropolitana Nord. Hospital Universitari Germans Trias i Pujol | Can Ruti SARS-CoV-2 Sequencing Hub (HUGTiP/IrsiCaixa/IGTP) | Marc Noguera-Julian, Pilar Armengol, Ignacio Blanco, Antoni E Bordoy, Francesc Catala-Moll, Pere-Joan Cardona, Maria Casadell, Cristina Casa, Gemma Clara, Bonaventura Clotet, Cristina Esteban, Montserrat Gimnez, Mercedes Guerrero, Anna Not, Roger Paredes, Mariona Parera, Vernica Saludes, Alba S nchez, and Elisa Martron behalf of the Can Ruti SARS-CoV-2 Sequencing Hub. |
| EPI_ISL_2448643 | Medilab | CHU Poitiers | Caroline MICHAUD, Maxime PICHON, Manon PRAT, Valentin BON-BARET, Birama N'DIAYE, Magali GARCIA, Agnes BEBY-DEFAUX, Nicolas LEVEQUE |
| EPI_ISL_2451794, EPI_ISL_2451799 | IL Dept. of Public Health Springfield Laboratory | Centers for Disease Control and Prevention Division of Viral Diseases, Pathogen Discovery | Mili Sheth, Sarah Nobles, Jasmine Padilla, Mark Burroughs, Shoshona Le, Katie Dillon, Peter Cook, Clinton R. Paden, Dhwani Batra, Krista Queen, Kristen Knipe, Dakota Howard, Yvette Unoarumhi, Darlene Wagner, Matthew Schmerer, Ben L. Rambo-Martin, Kristine Lacek, Sam Shepard, Alison Laufer Halpin, Dave Wentworth, Vivien Dugan, Suxiang Tong, Justin Lee |
| EPI_ISL_2455229 | New South Wales Health Pathology Royal Prince Alfred Hospital | Microbiology RPAH | Foster, C.; Au, J.; Ruiz Silva, M.; Deveson, I.; Bull, R.; Van Hal, S.; Rawlinson, W. |
| EPI_ISL_2455266, EPI_ISL_2455275 | Johns Hopkins Hospital Department of Pathology | Johns Hopkins Hospital Department of Pathology | C. Paul Morris, Chun Huai Luo, Adannaya Amadi, Matthew Schwartz, Nicholas Gallagher, Heba H. Mostafa |
| EPI_ISL_2456639 | Lighthouse Lab in Milton Keynes | Wellcome Sanger Institute for the COVID-19 Genomics UK (COG-UK) Consortium | The Lighthouse Lab in Milton Keynes and Alex Alderton, Roberto Amato, Jeffrey Barrett, Sonia Goncalves, Ewan Harrison, David K. Jackson, Ian Johnston, Dominic Kwiatkowski, Cordelia Langford, John Sillitoe on behalf of the Wellcome Sanger Institute COVID-19 Surveillance Team |
| EPI_ISL_2458463 | Utah Public Health Laboratory | Utah Public Health Laboratory | Erin L. Young, Kelly F. Oakeson, Tara Gallagher |
| EPI_ISL_2462870 | AZ Sint-Lucas Gent | AZ Sint-Lucas Gent | Jos Van Acker, Charlotte Verfaillie, Elke Vanlaere, Klara De Rauw |
| EPI_ISL_2462952 | Labo Analyses Med | National Reference Center for Viruses of Respiratory Infections, Institut Pasteur, Paris | Marion Barbet, Sylvie Behillil, Méline Bizard, Angela Brisebarre, Camille Capel, Vincent Enouf, Louise Lefrançois, Frédéric Lemoine, Christophe Malabat, Corinne Maufrais, Etienne Simon-Lorière, Maud Vanpeene, Sylvie Van der Werf ,Patricia Stoessel-Thouvenin |
| EPI_ISL_2462953, EPI_ISL_2462954 | Hospital | National Reference Center for Viruses of Respiratory Infections, Institut Pasteur, Paris | Marion Barbet, Sylvie Behillil, Méline Bizard, Angela Brisebarre, Camille Capel, Vincent Enouf, Louise Lefrançois, Frédéric Lemoine, Christophe Malabat, Corinne Maufrais, Etienne Simon-Lorière, Maud Vanpeene, Sylvie Van der Werf ,Pascale Martres |
| EPI_ISL_2462955 | Labo Analyses Med | National Reference Center for Viruses of Respiratory Infections, Institut Pasteur, Paris | Marion Barbet, Sylvie Behillil, Méline Bizard, Angela Brisebarre, Camille Capel, Vincent Enouf, Louise Lefrançois, Frédéric Lemoine, Christophe Malabat, Corinne Maufrais, Etienne Simon-Lorière, Maud Vanpeene, Sylvie Van der Werf ,Vincent Vieillefond |
| EPI_ISL_2462958 | Hospital | National Reference Center for Viruses of Respiratory Infections, Institut Pasteur, Paris | Marion Barbet, Sylvie Behillil, Méline Bizard, Angela Brisebarre, Camille Capel, Vincent Enouf, Louise Lefrançois, Frédéric Lemoine, Christophe Malabat, Corinne Maufrais, Etienne Simon-Lorière, Maud Vanpeene, Sylvie Van der Werf ,AgnèS Scanvic |
| EPI_ISL_2462960, EPI_ISL_2462962 | Hospital | National Reference Center for Viruses of Respiratory Infections, Institut Pasteur, Paris | Marion Barbet, Sylvie Behillil, Méline Bizard, Angela Brisebarre, Camille Capel, Vincent Enouf, Louise Lefrançois, Frédéric Lemoine, Christophe Malabat, Corinne Maufrais, Etienne Simon-Lorière, Maud Vanpeene, Sylvie Van der Werf ,Axelle Paquin |
| EPI_ISL_2462963 | Hospital | National Reference Center for Viruses of Respiratory Infections, Institut Pasteur, Paris | Marion Barbet, Sylvie Behillil, Méline Bizard, Angela Brisebarre, Camille Capel, Vincent Enouf, Louise Lefrançois, Frédéric Lemoine, Christophe Malabat, Corinne Maufrais, Etienne Simon-Lorière, Maud Vanpeene, Sylvie Van der Werf ,CéLine Ramanantsoa |
| EPI_ISL_2462988 | ZOL | Jessa | Berden et al. on behalf of the Jessa_cmdLab |
| EPI_ISL_2463242 | Quest Diagnostics Incorporated | Centers for Disease Control and Prevention Division of Viral Diseases, Pathogen Discovery | Dakota Howard, Dhwani Batra, Peter W. Cook, Kara Moser, Adrian Paskey, Jason Caravas, Benjamin Rambo-Martin, Shatavia Morrison, Christopher Gulvick, Scott Sammons, Yvette Unoarumhi, Darlene Wagner, Matthew Schmerer, S. H. Rosenthal, A. Gerasimova, R. M. Kagan, B. Anderson, M. Hua, Y. Liu, L.E. Bernstein, K.E. Livingston, A. Perez, I. A. Shlyakhter, R. V. Rolando, R. Owen, P. Tanpaiboon, F. Lacbawan, Clinton R. Paden, Duncan MacCannell |
| EPI_ISL_2463305, EPI_ISL_2463306, EPI_ISL_2463307, EPI_ISL_2463310 | LANCET LABORATORY | National Institute for Communicable Diseases of the National Health Laboratory Service | Sisonke Team, Amoako DG, Everatt J, Viana R, Glass A, Bhiman JN |
| EPI_ISL_2464082 | Hospital | National Reference Center for Viruses of Respiratory Infections, Institut Pasteur, Paris | Marion Barbet, Sylvie Behillil, Méline Bizard, Angela Brisebarre, Camille Capel, Vincent Enouf, Louise Lefrançois, Frédéric Lemoine, Christophe Malabat, Corinne Maufrais, Etienne Simon-Lorière, Maud Vanpeene, Sylvie Van der Werf ,Marianne Burgard |
| EPI_ISL_2464088, EPI_ISL_2464089 | Hospital | National Reference Center for Viruses of Respiratory Infections, Institut Pasteur, Paris | Marion Barbet, Sylvie Behillil, Méline Bizard, Angela Brisebarre, Camille Capel, Vincent Enouf, Louise Lefrançois, Frédéric Lemoine, Christophe Malabat, Corinne Maufrais, Etienne Simon-Lorière, Maud Vanpeene, Sylvie Van der Werf ,AgnèS Scanvic |
| EPI_ISL_2464098 | Armies | National Reference Center for Viruses of Respiratory Infections, Institut Pasteur, Paris | Marion Barbet, Sylvie Behillil, Méline Bizard, Angela Brisebarre, Camille Capel, Vincent Enouf, Louise Lefrançois, Frédéric Lemoine, Christophe Malabat, Corinne Maufrais, Etienne Simon-Lorière, Maud Vanpeene, Sylvie Van der Werf ,Marine Desroches |
| EPI_ISL_2464156 | Labo Analyses Med | National Reference Center for Viruses of Respiratory Infections, Institut Pasteur, Paris | Marion Barbet, Sylvie Behillil, Méline Bizard, Angela Brisebarre, Camille Capel, Vincent Enouf, Louise Lefrançois, Frédéric Lemoine, Christophe Malabat, Corinne Maufrais, Emmanuelle Permal, Etienne Simon-Lorière, Maud Vanpeene, Sylvie Van der Werf ,Arditi Cocco |
| EPI_ISL_2464168, EPI_ISL_2464177, EPI_ISL_2464183 | Labo Analyses Med | National Reference Center for Viruses of Respiratory Infections, Institut Pasteur, Paris | Marion Barbet, Sylvie Behillil, Méline Bizard, Angela Brisebarre, Camille Capel, Vincent Enouf, Louise Lefrançois, Frédéric Lemoine, Christophe Malabat, Corinne Maufrais, Etienne Simon-Lorière, Maud Vanpeene, Sylvie Van der Werf ,Arditi Cocco |
| EPI_ISL_2464242 | Hospital | National Reference Center for Viruses of Respiratory Infections, Institut Pasteur, Paris | Marion Barbet, Sylvie Behillil, Méline Bizard, Angela Brisebarre, Camille Capel, Vincent Enouf, Louise Lefrançois, Frédéric Lemoine, Christophe Malabat, Corinne Maufrais, Etienne Simon-Lorière, Maud Vanpeene, Sylvie Van der Werf ,Marianne Burgard |

| | | | |
|---|---|---|---|
| EPI_ISL_2464248 | Hospital | National Reference Center for Viruses of Respiratory Infections, Institut Pasteur, Paris | Marion Barbet, Sylvie Behillil, Méline Bizard, Angela Brisebarre, Camille Capel, Vincent Enouf, Louise Lefrançois, Frédéric Lemoine, Christophe Malabat, Corinne Maufrais, Etienne Simon-Lorière, Maud Vanpeene, Sylvie Van der Werf ,AgnèS Scanvic |
| EPI_ISL_2464422, EPI_ISL_2464423, EPI_ISL_2464424 | GH de l'Est Francilien | Department of Virology, Henri Mondor University Hospital, Assistance Publique Hôpitaux de Paris, Université Paris-Est Créteil, INSERM U955 | Christophe Rodriguez, Slim Fourati, Vanessa Demontant, Guillaume Gricourt, Melissa N'Debi, Alexandre Soulier, Elisabeth Trawinski, Jean-Michel Pawlotsky |
| EPI_ISL_2464442 | Hôpital Bicêtre | Department of Virology, Henri Mondor University Hospital, Assistance Publique Hôpitaux de Paris, Université Paris-Est Créteil, INSERM U955 | Christophe Rodriguez, Slim Fourati, Vanessa Demontant, Guillaume Gricourt, Melissa N'Debi, Alexandre Soulier, Elisabeth Trawinski, Jean-Michel Pawlotsky |
| EPI_ISL_2464473, EPI_ISL_2464479 | Biogroup Bio Lam-LCD Saint-Denis | Department of Virology, Henri Mondor University Hospital, Assistance Publique Hôpitaux de Paris, Université Paris-Est Créteil, INSERM U955 | Christophe Rodriguez, Slim Fourati, Vanessa Demontant, Guillaume Gricourt, Melissa N'Debi, Alexandre Soulier, Elisabeth Trawinski, Jean-Michel Pawlotsky |
| EPI_ISL_2464484, EPI_ISL_2464493, EPI_ISL_2464495 | Hôpital Avicenne | Department of Virology, Henri Mondor University Hospital, Assistance Publique Hôpitaux de Paris, Université Paris-Est Créteil, INSERM U955 | Christophe Rodriguez, Slim Fourati, Vanessa Demontant, Guillaume Gricourt, Melissa N'Debi, Alexandre Soulier, Elisabeth Trawinski, Jean-Michel Pawlotsky |
| EPI_ISL_2464502, EPI_ISL_2464512, EPI_ISL_2464513 | GH A.CHENEVIER-H.MONDOR | Department of Virology, Henri Mondor University Hospital, Assistance Publique Hôpitaux de Paris, Université Paris-Est Créteil, INSERM U955 | Christophe Rodriguez, Slim Fourati, Vanessa Demontant, Guillaume Gricourt, Melissa N'Debi, Alexandre Soulier, Elisabeth Trawinski, Jean-Michel Pawlotsky |
| EPI_ISL_2464521 | HOPITAL EMILE ROUX | Department of Virology, Henri Mondor University Hospital, Assistance Publique Hôpitaux de Paris, Université Paris-Est Créteil, INSERM U955 | Christophe Rodriguez, Slim Fourati, Vanessa Demontant, Guillaume Gricourt, Melissa N'Debi, Alexandre Soulier, Elisabeth Trawinski, Jean-Michel Pawlotsky |
| EPI_ISL_2464548, EPI_ISL_2464549 | Hôpital Cochin | Department of Virology, Henri Mondor University Hospital, Assistance Publique Hôpitaux de Paris, Université Paris-Est Créteil, INSERM U955 | Christophe Rodriguez, Slim Fourati, Vanessa Demontant, Guillaume Gricourt, Melissa N'Debi, Alexandre Soulier, Elisabeth Trawinski, Jean-Michel Pawlotsky |
| EPI_ISL_2464780 | Laboratoire CBM 25 TERRE ROUGE | Department of Virology, Henri Mondor University Hospital, Assistance Publique Hôpitaux de Paris, Université Paris-Est Créteil, INSERM U955 | Christophe Rodriguez, Slim Fourati, Vanessa Demontant, Guillaume Gricourt, Melissa N'Debi, Alexandre Soulier, Elisabeth Trawinski, Jean-Michel Pawlotsky |
| EPI_ISL_2464788 | Besançon Laboratoire CBM25 Terre Rouge | Department of Virology, Henri Mondor University Hospital, Assistance Publique Hôpitaux de Paris, Université Paris-Est Créteil, INSERM U955 | Christophe Rodriguez, Slim Fourati, Vanessa Demontant, Guillaume Gricourt, Melissa N'Debi, Alexandre Soulier, Elisabeth Trawinski, Jean-Michel Pawlotsky |
| EPI_ISL_2464887 | CH.INTERCOMMUNAL DE CRETEIL | Department of Virology, Henri Mondor University Hospital, Assistance Publique Hôpitaux de Paris, Université Paris-Est Créteil, INSERM U955 | Christophe Rodriguez, Slim Fourati, Vanessa Demontant, Guillaume Gricourt, Melissa N'Debi, Alexandre Soulier, Elisabeth Trawinski, Jean-Michel Pawlotsky |
| EPI_ISL_2464936 | CH. ROBERT BALLANGER | Department of Virology, Henri Mondor University Hospital, Assistance Publique Hôpitaux de Paris, Université Paris-Est Créteil, INSERM U955 | Christophe Rodriguez, Slim Fourati, Vanessa Demontant, Guillaume Gricourt, Melissa N'Debi, Alexandre Soulier, Elisabeth Trawinski, Jean-Michel Pawlotsky |
| EPI_ISL_2464980 | Biogroup Bio Lam-LCD Saint-Denis | Department of Virology, Henri Mondor University Hospital, Assistance | Christophe Rodriguez, Slim Fourati, Vanessa Demontant, Guillaume Gricourt, Melissa N'Debi, Alexandre Soulier, Elisabeth Trawinski, Jean-Michel Pawlotsky |

| | | | |
|---|---|---|---|
| | | Publique Hôpitaux de Paris, Université Paris-Est Créteil, INSERM U955 | |
| EPI_ISL_2465017 | SELAS CERBALLIANCE Normandie ouest | Department of Virology, Henri Mondor University Hospital, Assistance Publique Hôpitaux de Paris, Université Paris-Est Créteil, INSERM U955 | Christophe Rodriguez, Slim Fourati, Vanessa Demontant, Guillaume Gricourt, Melissa N'Debi, Alexandre Soulier, Elisabeth Trawinski, Jean-Michel Pawlotsky |
| EPI_ISL_2466325 | National Platform bis UMONS/Jolimont | National Platform bis UMONS/Jolimont | François Dufrasne, Guillaume Bayon-Vicente, Florian Juszczak, Gautier Detry, Ruddy Wattiez |
| EPI_ISL_2466510 | National Institute of Public Health | State Veterinary Institute Prague | Nagy,A;Jirincova,H;Suri,T;Trnka,D;Vecerova,J |
| EPI_ISL_2466736, EPI_ISL_2466737, EPI_ISL_2466738 | Nebraska Public Health Laboratory | NPHL COVID-19 Response Team | NPHL COVID-19 Response Team |
| EPI_ISL_2467794 | University of Wisconsin-Madison AIDS Vaccine Research Laboratories | University of Wisconsin-Madison AIDS Vaccine Research Laboratories | Gage Moreno, Katarina Braun, et al. AIDS Vaccine Research Laboratories |
| EPI_ISL_2467803 | Alaska State Virology Laboratory | Alaska State Virology Laboratory | Stephanie DeRonde, Elva House, Jacob Zidek, Lisa Smith, Ph.D., Jack Chen, Ph.D. |
| EPI_ISL_2467881 | NC State Laboratory of Public Health | Genomics and Discovery, Respiratory Viruses Branch, Division of Viral Diseases, Centers for Disease Control and Prevention | Ying Tao, Yan Li, Jing Zhang, Anna Kelleher, Brian Lynch, Krista Queen, Anna Uehara, Peter Cook, Han Jia Justin Ng, Rachel Marine, Clinton R. Paden, Dhwani Batra, Haibin Wang, Tara Coalter, Jasmine Padilla, Morgan Davis, Mili Sheth, Sarah Nobles, Mark Burroughs, Justin Lee, Adam Retchless, Suxiang Tong |
| EPI_ISL_2469571 | SYNLAB MVZ Leinfelden-Echterdingen | Robert Koch Institute | unknown |
| EPI_ISL_2470239 | LabKom - Labor Hannover MVZ GmbH | Robert Koch Institute | unknown |
| EPI_ISL_2470249, EPI_ISL_2470309 | Sonic - Bioscientia - MVZ Labor Saar GmbH | Robert Koch Institute | unknown |
| EPI_ISL_2471027 | Labormedizin Darmstadt | Robert Koch Institute | unknown |
| EPI_ISL_2471047, EPI_ISL_2471057 | Labor Dr. Heidrich & Kollegen MVZ GmbH Hamburg | Robert Koch Institute | unknown |
| EPI_ISL_2471241 | CENTOGENE Frankfurt Laboratory: Niederlassung Industriepark Höchst | Robert Koch Institute | unknown |
| EPI_ISL_2471529 | MVZ Medizinisches Labor Hannover GmbH | Robert Koch Institute | unknown |
| EPI_ISL_2471644, EPI_ISL_2471651 | Eurofins LifeCodexx GmbH | Robert Koch Institute | unknown |
| EPI_ISL_2471910 | MVZ Labor Dr. Limbach & Kollegen GbR | Robert Koch Institute | unknown |
| EPI_ISL_2472394 | MDI Limbach Berlin GmbH; MVZ Labor Berlin | Robert Koch Institute | unknown |
| EPI_ISL_2472470, EPI_ISL_2472488, EPI_ISL_2472491, EPI_ISL_2472494, EPI_ISL_2472499 | MVZ Labor Dr. Limbach & Kollegen GbR | Robert Koch Institute | unknown |
| EPI_ISL_2472547 | Labor Dr. Spranger | Robert Koch Institute | unknown |
| EPI_ISL_2472859 | Labor ZOTZ\|KLIMAS; MVZ Düsseldorf-Centrum | Robert Koch Institute | unknown |
| EPI_ISL_2473327, EPI_ISL_2473341, EPI_ISL_2473362, EPI_ISL_2473477, EPI_ISL_2473531 | State Testing Facility | Altius Institute for Biomedical Research | Daniel Bates, Rebecca Bruders, Michael Buckley, Mark Frerker, Amanda Gale, Clem Green, Muhammad Halimun, Kneshay Harper, Matt Hartman, Alex Isner, Audra Johnson, Jessica Kunder, Lauren Mitchell, Jemma Nelson, Alex Nguyen, Sofia Olsson, Sadie Patraw, Tobias Ragoczy, Joshua Richards, Jean Robinston, Jacob Rodriguez, John Stamatoyannopoulos, Eric Thorland, Julia Wald |
| EPI_ISL_2476301, EPI_ISL_2476305, EPI_ISL_2476306 | Dutch COVID-19 response team | National Institute for Public Health and the Environment (RIVM) | Adam Meijer, Harry Vennema, Dirk Eggink, Jeroen Cremer, Sharon van den Brink, Bas van der Veer, AnneMarie van den Brandt, Lisa Wijsman, Kim Freriks, Ryanne Jaarsma, Euníce Then, Lynn Aarts, Sanne Bos, Melissa van Tuil, Linda van de Nes, Florian Zwagemaker, Dennis Schmitz, Annelies Kroneman, Karim Hajji, Chantal Reusken, on behalf of the national COVID-19 response team |
| EPI_ISL_2480245, EPI_ISL_2480912, EPI_ISL_2481076, EPI_ISL_2481340, EPI_ISL_2481776, EPI_ISL_2481896, EPI_ISL_2482231 | Laboratory Corporation of America | Centers for Disease Control and Prevention Division of Viral Diseases, Pathogen Discovery | Dakota Howard, Dhwani Batra, Peter W. Cook, Kara Moser, Adrian Paskey, Jason Caravas, Benjamin Rambo-Martin, Shatavia Morrison, Christopher Gulvick, Scott Sammons, Yvette Unoarumhi, Darlene Wagner, Matthew Schmerer, Minoo Agarwal, Eyad Almasri, Debbie Boles, Ayla Burns, Nuthawin Charoensri, Oren Cohen, Susan Countryman, Mary Ann Cristobal, Bobbi Croy, Suzanne Dale, Hrushikesh Deshmukh, Amanda Douglas, Vincent Drouillon, Marcia Eisenberg, Howard Engler, Rama Ghatti, Prashant Gupta, Susan Hicks, Jake Humphrey, Lax Iyer, Lisa Pfefferle, Manoj Jain, Matthew Robinson, Mohan Kolli, Brian Krueger, Tim Kuphal, Stanley Letovsky, Michael Levandoski, Craig Lukasik, Jonathan Meltzer, Brian Norvell, Mindy Nye, Scott Parker, Christos Petropoulos, John Pruitt, Steven Ragan, Scott Ryan, Mike Sapeta, Jana Schroth, Suresh Babu Selvaraju, |

| | | | |
|---|---|---|---|
| | | | Goran Stevovic, Amanda Suchanek, Andrea Throop, Lyndon Tilson, Thomas Urban, Joe Voshell, Kimberly Wagner, Jonathan Williams, Mary Williamson, Qian Zeng, Tricia Zwiefelhofer, Clinton R. Paden, Duncan MacCannell |
| EPI_ISL_2482521 | LABORATOIRE DE BIOLOGIE MEDICALE | CNR Virus des Infections Respiratoires - France SUD | Antonin Bal, Gregory Destras, Gwendolyne Burfin, Hadrien Regue, Quentin Semanas, Martine Valette, Bruno Lina, Laurence Josset |
| EPI_ISL_2482590 | LABORATOIRE UNILIANS DECINES | CNR Virus des Infections Respiratoires - France SUD | Antonin Bal, Gregory Destras, Gwendolyne Burfin, Hadrien Regue, Quentin Semanas, Martine Valette, Bruno Lina, Laurence Josset |
| EPI_ISL_2482608 | MIRIALIS CLUSES BECHET | CNR Virus des Infections Respiratoires - France SUD | Antonin Bal, Gregory Destras, Gwendolyne Burfin, Hadrien Regue, Quentin Semanas, Martine Valette, Bruno Lina, Laurence Josset |
| EPI_ISL_2482633 | LABORATOIRE UNILIANS DECINES | CNR Virus des Infections Respiratoires - France SUD | Antonin Bal, Gregory Destras, Gwendolyne Burfin, Hadrien Regue, Quentin Semanas, Martine Valette, Bruno Lina, Laurence Josset |
| EPI_ISL_2482669 | LABORATOIRE CREAVALLEE | CNR Virus des Infections Respiratoires - France SUD | Antonin Bal, Gregory Destras, Gwendolyne Burfin, Hadrien Regue, Quentin Semanas, Martine Valette, Bruno Lina, Laurence Josset |
| EPI_ISL_2482787 | LABORATOIRE CERBALLIANCE PLT VILLON | CNR Virus des Infections Respiratoires - France SUD | Antonin Bal, Gregory Destras, Gwendolyne Burfin, Hadrien Regue, Quentin Semanas, Martine Valette, Bruno Lina, Laurence Josset |
| EPI_ISL_2482799 | HOPITAL SAINT ANDRE | CNR Virus des Infections Respiratoires - France SUD | Antonin Bal, Gregory Destras, Gwendolyne Burfin, Hadrien Regue, Quentin Semanas, Martine Valette, Bruno Lina, Laurence Josset |
| EPI_ISL_2482828 | C.H.R.U. MONTPIED | CNR Virus des Infections Respiratoires - France SUD | Antonin Bal, Gregory Destras, Gwendolyne Burfin, Hadrien Regue, Quentin Semanas, Martine Valette, Bruno Lina, Laurence Josset |
| EPI_ISL_2482829 | HOPITAL ALBI | CNR Virus des Infections Respiratoires - France SUD | Antonin Bal, Gregory Destras, Gwendolyne Burfin, Hadrien Regue, Quentin Semanas, Martine Valette, Bruno Lina, Laurence Josset |
| EPI_ISL_2482837, EPI_ISL_2482841, EPI_ISL_2482845 | LBM UNIBIO VALENTIN | CNR Virus des Infections Respiratoires - France SUD | Antonin Bal, Gregory Destras, Gwendolyne Burfin, Hadrien Regue, Quentin Semanas, Martine Valette, Bruno Lina, Laurence Josset |
| EPI_ISL_2482918, EPI_ISL_2482920, EPI_ISL_2482921 | CERBAILLIANCE LA REUNION | CNR Virus des Infections Respiratoires - France SUD | Antonin Bal, Gregory Destras, Gwendolyne Burfin, Hadrien Regue, Quentin Semanas, Martine Valette, Bruno Lina, Laurence Josset |
| EPI_ISL_2482922 | CERBALLIANCE REUNION | CNR Virus des Infections Respiratoires - France SUD | Antonin Bal, Gregory Destras, Gwendolyne Burfin, Hadrien Regue, Quentin Semanas, Martine Valette, Bruno Lina, Laurence Josset |
| EPI_ISL_2482927, EPI_ISL_2482930 | CERBAILLIANCE LA REUNION | CNR Virus des Infections Respiratoires - France SUD | Antonin Bal, Gregory Destras, Gwendolyne Burfin, Hadrien Regue, Quentin Semanas, Martine Valette, Bruno Lina, Laurence Josset |
| EPI_ISL_2482941 | ORIAPOLE - BACTERIOLOGIE | CNR Virus des Infections Respiratoires - France SUD | Antonin Bal, Gregory Destras, Gwendolyne Burfin, Hadrien Regue, Quentin Semanas, Martine Valette, Bruno Lina, Laurence Josset |
| EPI_ISL_2483090 | Furst Medical Laboratory | Norwegian Institute of Public Health, Department of Virology | Kathrine Stene-Johansen, Kamilla Heddeland Instefjord, Hilde Elshaug, Garcia Llorente Ignacio, Jon Bråte, Engebretsen Serina Beate,Pedersen Benedikte Nevjen, Line Victoria Moen, Debech Nadia, Atiya R Ali,Marie Paulsen Madsen, Rasmus Riis Kopperud, Hilde Vollan, Karoline Bragstad, Olav Hungnes |
| EPI_ISL_2483103 | Ostfold Hospital Trust - Kalnes, Centre for Laboratory Medicine, Section for gene technology and infection serology | Norwegian Institute of Public Health, Department of Virology | Kathrine Stene-Johansen, Kamilla Heddeland Instefjord, Hilde Elshaug, Garcia Llorente Ignacio, Jon Bråte, Engebretsen Serina Beate,Pedersen Benedikte Nevjen, Line Victoria Moen, Debech Nadia, Atiya R Ali,Marie Paulsen Madsen, Rasmus Riis Kopperud, Hilde Vollan, Karoline Bragstad, Olav Hungnes |
| EPI_ISL_2483247 | Platform BIS UZA/UAntwerpen | Labo Klinische Biologie, UZA | Jasmine Coppens, Marie Le Mercier, Basil Britto Xavier, Christine Lammens, Veerle Matheeussen, Herman Goossens |
| EPI_ISL_2483708 | National Platform bis UMONS/Jolimont | National Platform bis UMONS/Jolimont | François Dufrasne, Guillaume Bayon-Vicente, Florian Juszczak, Gautier Detry, Ruddy Wattiez |
| EPI_ISL_2483754 | Ospedale Sandro Pertini ASL Roma 2 | INMI Lazzaro Spallanzani IRCCS | G Bonfiglio, O Butera, F Messina, CEM Gruber, G Orlandi, E Sperandio, MC Cava, R Longo, B Bartolini |
| EPI_ISL_2484019 | Hospital General Universitario Gregorio Marañón | Hospital General Universitario Gregorio Marañón | Sergio Buenestado Serrano, Pedro Sola Campoy, Laura Pérez-Lago, Cristina Rodriguez-Grande, Marta Herranz Martin, Victor Manuel de la Cueva, Julia Suárez, Pilar Catalán, Patricia Muñoz, Darío García de Viedma |
| EPI_ISL_2484046 | Arizona State University | Arizona State University | Peter T. Skidmore, LaRinda A. Holland, Matthew F. Smith, Rabia Maqsood, Nicholas J. Mellor, Joy M. Blain, Valerie Harris, Joshua LaBaer, Vel Murugan, Efrem S. Lim |
| EPI_ISL_2484882 | Southern Nevada Public Health Laboratory | Southern Nevada Public Health Laboratory | Michael Picker |
| EPI_ISL_2487022 | Lighthouse Lab in Milton Keynes | Wellcome Sanger Institute for the COVID-19 Genomics UK (COG-UK) Consortium | The Lighthouse Lab in Milton Keynes and Alex Alderton, Roberto Amato, Jeffrey Barrett, Sonia Goncalves, Ewan Harrison, David K. Jackson, Ian Johnston, Dominic Kwiatkowski, Cordelia Langford, John Sillitoe on behalf of the Wellcome Sanger Institute COVID-19 Surveillance Team |
| EPI_ISL_2491660 | Centre Hospitalier Universitaire Clermont-Ferrand | CHU Clermont-Ferrand, service de virologie | Bisseux Maxime, Mirand Audrey, Combes Patricia, Henquell Cécile |
| EPI_ISL_2491664 | Centre hospitalier D'AURILLAC | CHU Clermont-Ferrand, service de virologie | Bisseux Maxime, Mirand Audrey, Combes Patricia, Henquell Cécile |
| EPI_ISL_2493041 | AMPATH | CERI, Centre for Epidemic Response and Innvoation, Stellenbosch University and KRISP, | Giandhari J, Pillay S, Naidoo Y, Ramphal U, Lessells R, Mdlalose K, York D, Khan S, Emmanuel SJ, Tegally H, Wilkinson E, de Oliveira T |

| | | | |
|---|---|---|---|
| | | KZN Research Innovation and Sequencing Platform, UKZN. | |
| EPI_ISL_2493147 | Hospital Universitari Arnau de Vilanova | Hospital Universitari Vall d'Hebron - Vall d'Hebron Institut de Recerca | Cristina Andrés, Maria Piñana, Alejandra González-Sánchez, Damir Garcia-Cehic, Ariadna Rando, Juliana Esperalba, Maria Gema Codina, Carla Castillo, Maria Carmen Martin, Tomàs Pumarola, Josep Quer, Andrés Antón |
| EPI_ISL_2495603 | MB-Cadham Provincial laboratory | National Microbiology Laboratory (NML) | Anna Majer, Shari Tyson, Grace Seo, Philip Mabon, Elsie Grudeski, Rhiannon Huzarewich, Russell Mandes, Anneliese Landgraff, Jennifer Tanner, Natalie Knox, Morag Graham, Gary Van Domselaar, Paul Van Caeseele, Jared Bullard, David Alexander, Kerry Dust, Nathalie Bastien, Yan Li, Timothy Booth, Darian Hole, Madison Chapel, Kirsten Biggar, CanCOGeN's metadata curation team, Public Health Agency of Canada CanCOGeN team |
| EPI_ISL_2497353, EPI_ISL_2497382, EPI_ISL_2497422 | UW Virology Lab | UW Virology Lab | Pavitra Roychoudhury, Hong Xie, Lasata Shrestha, Tien V. Nguyen, Shah Mohamed Bakhash, Michelle Lin, Noah R. Baker, Ricardo Perez, Sean Ellis, Nathan Breit, Robert J. Livingston, Meei-Li Huang, Keith R Jerome, Patrick Mathias, Alexander Greninger |
| EPI_ISL_2499991 | University of Liège COVID-19 testing center | GIGA Medical Genomics | Keith Durkin, Maria Artesi, Bouchra Boujemla, Nathalie Renotte, Cécile Meex, Sébastien Bontems, Fabrice Bureau, Laurent Gillet, Wouter Coppieters, Marie-Pierre Hayette, Vincent Bours |
| EPI_ISL_2500498 | CHUV | Laboratory of genomics and metagenomics | Trestan Pillonel, Damien Jacot, Sébastien Aeby, Gilbert Greub, Claire Bertelli |
| EPI_ISL_2501036 | Virology Unit, Institut Pasteur du Cambodge | Virology Unit, Institut Pasteur du Cambodge | Cecile Troupin, Leakhena Pum, Jurre Y Siegers, Ly Sovann, Kraing Sidonn, Yi Sengdoeurn, Chin Savuth, Chau Darapheak, Veasna Duong, Erik A Karlsson |
| EPI_ISL_2501090 | AMPATH | KRISP, KZN Research Innovation and Sequencing Platform | Sisonke Team, Giandhari J, Pillay S, Naidoo Y, Ramphal U, Lessells R, Mdlalose K, York D, Khan S, Emmanuel SJ, Tegally H, Wilkinson E, de Oliveira T |
| EPI_ISL_2501092, EPI_ISL_2501094, EPI_ISL_2501095 | Netcare St Augustines Hospital | KRISP, KZN Research Innovation and Sequencing Platform | Sisonke Team, Giandhari J, Pillay S, Naidoo Y, Ramphal U, Lessells R, Mdlalose K, York D, Khan S, Emmanuel SJ, Tegally H, Wilkinson E, de Oliveira T |
| EPI_ISL_2501301 | CH LENS | CHU Lille - Laboratoire de Virologie | AIT YAHYA Emilie, ALIDJINOU Enagnon Kazali, BOCKET Laurence, CREPIN Michel, DEMAY Christophe, ENGELMANN Ilka, GEFFROY Sandrine, GUIGON Aurélie, LAMBERT Valérie, LAZREK Mouna, NOBILLIAUX Florian, PREVOST Brigitte, THUILLIER Caroline, TINEZ Claire ,TCHANTCHOU NJOSSE YANICK |
| EPI_ISL_2501719 | Yale Clinical Virology Lab | Grubaugh Lab - Yale School of Public Health | Joseph Fauver, Mallery Breban, Isabel Ott, Tara Alpert, Mary Petrone, Anderson Brito, Chantal Vogels, Annie Watkins, Chaney Kalinich, Jessica Rothman, Marie L. Landry, Nathan Grubaugh |
| EPI_ISL_2501929 | Missouri State Public Health Laboratory | Missouri State Public Health Laboratory | Matthew Sinn, Joshua Barry, Ashley New |
| EPI_ISL_2504068, EPI_ISL_2504071, EPI_ISL_2504072, EPI_ISL_2504073, EPI_ISL_2504088 | Botswana Harvard HIV Reference Laboratory | Botswana Harvard HIV Reference Laboratory | Sikhulile Moyo, Dorcas Maruapula, Wonderful T. Choga, Thongbotho Mphoyakgosi, Boitumelo Zuze, Botshelo Radibe, Legodile Kooepile, Ontlametse T. Bareng, Keoratile Ntshambiwa, Modisa Motswaledi, Madisa Mine, Joseph Makhema, Roger Shapiro, Shahin Lockman, Mosepele Mosepele, Simani Gaseitsiwe |
| EPI_ISL_2504114 | Palapye Primary Hospital Laboratory | Botswana Harvard HIV Reference Laboratory | Sikhulile Moyo, Dorcas Maruapula, Wonderful T. Choga, Thongbotho Mphoyakgosi, Boitumelo Zuze, Botshelo Radibe, Legodile Kooepile, Ontlametse T. Bareng, Letsibogo Gaoraelwe, Thela Tefelo, Keoratile Ntshambiwa, Modisa Motswaledi, Madisa Mine, Joseph Makhema, Roger Shapiro, Shahin Lockman, Mosepele Mosepele, Simani Gaseitsiwe |
| EPI_ISL_2508402, EPI_ISL_2508446, EPI_ISL_2508463 | Microbiology Division, SC DHEC | Microbiology Division, SC DHEC | Freeman,J., Flores,H. |
| EPI_ISL_2508715, EPI_ISL_2508716, EPI_ISL_2508723, EPI_ISL_2508724, EPI_ISL_2508725, EPI_ISL_2508801, EPI_ISL_2508802, EPI_ISL_2508803, EPI_ISL_2508804, EPI_ISL_2508888, EPI_ISL_2509024, EPI_ISL_2509025, EPI_ISL_2509026, EPI_ISL_2509027, EPI_ISL_2509028 | | | |
| see above | National Public Health Laboratory, National Centre for Infectious Diseases | National Public Health Laboratory, National Centre for Infectious Diseases | Tze Minn Mak, Zhenyang Zhou, Royce Ang, Lin Cui, Raymond Tzer Pin Lin |
| EPI_ISL_2509334 | Palapye Primary Hospital Laboratory | Botswana Harvard HIV Reference Laboratory | Sikhulile Moyo, Dorcas Maruapula, Wonderful T. Choga, Thongbotho Mphoyakgosi, Boitumelo Zuze, Botshelo Radibe, Legodile Kooepile, Ontlametse T. Bareng, Letsibogo Gaoraelwe, Thela Tefelo, Keoratile Ntshambiwa, Modisa Motswaledi, Madisa Mine, Joseph Makhema, Roger Shapiro, Shahin Lockman, Mosepele Mosepele, Simani Gaseitsiwe |
| EPI_ISL_2510596 | Servicio de Microbiología Clínica (Complejo Hospitalario de Navarra, Pamplona) | Centro de Secuenciación NASERTIC | Carmen Ezpeleta Baquedano, Ana Navascués, Ana Miqueleiz |
| EPI_ISL_2516556 | Hospital Universitari Bellvitge | Microbiology Department | Sara Marti, Aida Gonzalez-Diaz, Laura Calatayud, Jordi Niubó, Miguel Fernandez-Huerta, Carmen Ardanuy, Jordi Camara, M Angeles Domínguez |
| EPI_ISL_2516672, EPI_ISL_2516682, EPI_ISL_2516690 | Servicio de Microbiología Clínica (Complejo Hospitalario de Navarra, Pamplona) | Centro de Secuenciación NASERTIC | Carmen Ezpeleta Baquedano, Ana Navascués, Ana Miqueleiz |
| EPI_ISL_2521974 | Plateforme de testing Namuroise | Plateforme de testing Namuroise | Nicolas Gilliard; Demars Aurore ; Maschietto Céline ; Nobis Chloé ; Otto Gaetan ; Denis Olivier ; Degosserie Jonathan ; Mullier François |
| EPI_ISL_2521995, EPI_ISL_2521996 | Nucleic Acid Testing, National Reference Laboratory | GIGA Medical Genomics | Yvan Butera, Keith Durkin, Maria Artesi, Bouchra Boujemla, Robert Rutayisire, Patrick Tuyisenge, Esperence Umumararungu, Sébastien Bontems, Marie-Pierre Hayette, Nathalie Renotte, Swaibu Gatare, Jacob Souopgui, Sabin Nsanzimana, Vincent Bours, Léon Mutesa |
| EPI_ISL_2523862 | HELIX LLC | WHO National Influenza Centre Russian Federation | Andrey Komissarov, Artem Fadeev, Kseniya Komissarova, Oula Mansour, Kirill Varchenko, Mikhail Bakaev, Tamila Musaeva, Maria Timofeeva, Veronika Eder, Maria Pisareva, Nikita Yolshin, Daria Danilenko, Ksenia Safina, Elena Nabieva, |

| | | | |
|---|---|---|---|
| | | | Georgii Bazykin, Dmitry Lioznov |
| EPI_ISL_2524774 | Hospital Universitari Arnau de Vilanova | Hospital Universitari Vall d'Hebron - Vall d'Hebron Institut de Recerca | Cristina Andrés, Maria Piñana, Alejandra González-Sánchez, Damir Garcia-Cehic, Ariadna Rando, Juliana Esperalba, Maria Gema Codina, Carla Castillo, Maria Carmen Martin, Tomàs Pumarola, Josep Quer, Andrés Antón |
| EPI_ISL_2531827 | Hospital | National Reference Center for Viruses of Respiratory Infections, Institut Pasteur, Paris | Marion Barbet, Sylvie Behillil, Méline Bizard, Angela Brisebarre, Camille Capel, Vincent Enouf, Louise Lefrançois, Frédéric Lemoine, Christophe Malabat, Corinne Maufrais, Etienne Simon-Lorière, Maud Vanpeene, Sylvie Van der Werf ,Pascale Martres |
| EPI_ISL_2531877 | Hospital | National Reference Center for Viruses of Respiratory Infections, Institut Pasteur, Paris | Marion Barbet, Sylvie Behillil, Méline Bizard, Angela Brisebarre, Camille Capel, Vincent Enouf, Louise Lefrançois, Frédéric Lemoine, Christophe Malabat, Corinne Maufrais, Etienne Simon-Lorière, Maud Vanpeene, Sylvie Van der Werf ,Marianne Burgard |
| EPI_ISL_2531934 | RIIP | National Reference Center for Viruses of Respiratory Infections, Institut Pasteur, Paris | Marion Barbet, Sylvie Behillil, Méline Bizard, Angela Brisebarre, Camille Capel, Vincent Enouf, Louise Lefrançois, Frédéric Lemoine, Christophe Malabat, Corinne Maufrais, Etienne Simon-Lorière, Maud Vanpeene, Sylvie Van der Werf ,StéPhanie Guyomard-Rabenirina |
| EPI_ISL_2531962 | Labo Analyses Med | National Reference Center for Viruses of Respiratory Infections, Institut Pasteur, Paris | Marion Barbet, Sylvie Behillil, Méline Bizard, Angela Brisebarre, Camille Capel, Vincent Enouf, Louise Lefrançois, Frédéric Lemoine, Christophe Malabat, Corinne Maufrais, Etienne Simon-Lorière, Maud Vanpeene, Sylvie Van der Werf ,Vicky Le |
| EPI_ISL_2532279, EPI_ISL_2532280, EPI_ISL_2532281 | Labo Analyses Med | National Reference Center for Viruses of Respiratory Infections, Institut Pasteur, Paris | Marion Barbet, Sylvie Behillil, Méline Bizard, Angela Brisebarre, Camille Capel, Vincent Enouf, Louise Lefrançois, Frédéric Lemoine, Christophe Malabat, Corinne Maufrais, Etienne Simon-Lorière, Maud Vanpeene, Sylvie Van der Werf ,OphéLie Said-Delattre |
| EPI_ISL_2532284, EPI_ISL_2532285, EPI_ISL_2532286 | Labo Analyses Med | National Reference Center for Viruses of Respiratory Infections, Institut Pasteur, Paris | Marion Barbet, Sylvie Behillil, Méline Bizard, Angela Brisebarre, Camille Capel, Vincent Enouf, Louise Lefrançois, Frédéric Lemoine, Christophe Malabat, Corinne Maufrais, Etienne Simon-Lorière, Maud Vanpeene, Sylvie Van der Werf ,Eve Haguenoer |
| EPI_ISL_2532287 | Labo Analyses Med | National Reference Center for Viruses of Respiratory Infections, Institut Pasteur, Paris | Marion Barbet, Sylvie Behillil, Méline Bizard, Angela Brisebarre, Camille Capel, Vincent Enouf, Louise Lefrançois, Frédéric Lemoine, Christophe Malabat, Corinne Maufrais, Etienne Simon-Lorière, Maud Vanpeene, Sylvie Van der Werf ,GréGoire Potiron |
| EPI_ISL_2532311, EPI_ISL_2532312, EPI_ISL_2532313 | Hospital | National Reference Center for Viruses of Respiratory Infections, Institut Pasteur, Paris | Marion Barbet, Sylvie Behillil, Méline Bizard, Angela Brisebarre, Camille Capel, Vincent Enouf, Louise Lefrançois, Frédéric Lemoine, Christophe Malabat, Corinne Maufrais, Etienne Simon-Lorière, Maud Vanpeene, Sylvie Van der Werf ,Pascale Martres |
| EPI_ISL_2532315 | Hospital | National Reference Center for Viruses of Respiratory Infections, Institut Pasteur, Paris | Marion Barbet, Sylvie Behillil, Méline Bizard, Angela Brisebarre, Camille Capel, Vincent Enouf, Louise Lefrançois, Frédéric Lemoine, Christophe Malabat, Corinne Maufrais, Amaury Vaysse, Etienne Simon-Lorière, Maud Vanpeene, Sylvie Van der Werf ,Pascale Martres |
| EPI_ISL_2532388 | Hospital | National Reference Center for Viruses of Respiratory Infections, Institut Pasteur, Paris | Marion Barbet, Sylvie Behillil, Méline Bizard, Angela Brisebarre, Camille Capel, Vincent Enouf, Louise Lefrançois, Frédéric Lemoine, Christophe Malabat, Corinne Maufrais, Etienne Simon-Lorière, Maud Vanpeene, Sylvie Van der Werf ,AgnèS Scanvic |
| EPI_ISL_2532412 | Labo Analyses Med | National Reference Center for Viruses of Respiratory Infections, Institut Pasteur, Paris | Marion Barbet, Sylvie Behillil, Méline Bizard, Angela Brisebarre, Camille Capel, Vincent Enouf, Louise Lefrançois, Frédéric Lemoine, Christophe Malabat, Corinne Maufrais, Etienne Simon-Lorière, Maud Vanpeene, Sylvie Van der Werf ,Pierre-Yves Leonard |
| EPI_ISL_2532465 | Labo Analyses Med | National Reference Center for Viruses of Respiratory Infections, Institut Pasteur, Paris | Marion Barbet, Sylvie Behillil, Méline Bizard, Angela Brisebarre, Camille Capel, Vincent Enouf, Louise Lefrançois, Frédéric Lemoine, Christophe Malabat, Corinne Maufrais, Etienne Simon-Lorière, Maud Vanpeene, Sylvie Van der Werf |
| EPI_ISL_2533826 | Laboratoire Virologie Saint Louis APHP | Laboratoire Virologie Saint Louis APHP | Maud Salmona, Marie Laure Chaix, Severine Mercier Delarue, Marie Laure Néré, Linda Feghoul, Jérome Le Goff, Constance Delauguerre, Sophia Achaibou |
| EPI_ISL_2535237 | Labo Analyses Med | National Reference Center for Viruses of Respiratory Infections, Institut Pasteur, Paris | Marion Barbet, Sylvie Behillil, Méline Bizard, Angela Brisebarre, Camille Capel, Vincent Enouf, Louise Lefrançois, Frédéric Lemoine, Christophe Malabat, Corinne Maufrais, Etienne Simon-Lorière, Maud Vanpeene, Sylvie Van der Werf |
| EPI_ISL_2535271 | Hospital | National Reference Center for Viruses of Respiratory Infections, Institut Pasteur, Paris | Marion Barbet, Sylvie Behillil, Méline Bizard, Angela Brisebarre, Camille Capel, Vincent Enouf, Louise Lefrançois, Frédéric Lemoine, Christophe Malabat, Corinne Maufrais, Damien Mornico, Etienne Simon-Lorière, Maud Vanpeene, Sylvie Van der Werf ,Pascale Martres |
| EPI_ISL_2535274 | Hospital | National Reference Center for Viruses of Respiratory Infections, Institut Pasteur, Paris | Marion Barbet, Sylvie Behillil, Méline Bizard, Angela Brisebarre, Camille Capel, Vincent Enouf, Louise Lefrançois, Frédéric Lemoine, Christophe Malabat, Corinne Maufrais, Etienne Simon-Lorière, Maud Vanpeene, Sylvie Van der Werf ,Pascale Martres |
| EPI_ISL_2535276 | Labo Analyses Med | National Reference Center for Viruses of Respiratory Infections, Institut Pasteur, Paris | Marion Barbet, Sylvie Behillil, Méline Bizard, Angela Brisebarre, Camille Capel, Vincent Enouf, Louise Lefrançois, Frédéric Lemoine, Christophe Malabat, Corinne Maufrais, Victoire Baillet, Etienne Simon-Lorière, Maud Vanpeene, Sylvie Van der Werf ,OphéLie Said-Delattre |
| EPI_ISL_2535281 | Labo Analyses Med | National Reference Center for Viruses of Respiratory Infections, Institut Pasteur, Paris | Marion Barbet, Sylvie Behillil, Méline Bizard, Angela Brisebarre, Camille Capel, Vincent Enouf, Louise Lefrançois, Frédéric Lemoine, Christophe Malabat, Corinne Maufrais, Etienne Simon-Lorière, Maud Vanpeene, Sylvie Van der Werf ,OphéLie Said-Delattre |
| EPI_ISL_2535524 | Labo Analyses Med | National Reference Center for Viruses of Respiratory Infections, Institut Pasteur, Paris | Marion Barbet, Sylvie Behillil, Méline Bizard, Angela Brisebarre, Camille Capel, Vincent Enouf, Louise Lefrançois, Frédéric Lemoine, Christophe Malabat, Corinne Maufrais, Etienne Simon-Lorière, Maud Vanpeene, Sylvie Van der Werf , Hugues Leroy |
| EPI_ISL_2535555 | Hospital | National Reference Center for Viruses of Respiratory Infections, Institut Pasteur, Paris | Marion Barbet, Sylvie Behillil, Méline Bizard, Angela Brisebarre, Camille Capel, Vincent Enouf, Louise Lefrançois, Frédéric Lemoine, Christophe Malabat, Corinne Maufrais, Etienne Simon-Lorière, Maud Vanpeene, Sylvie Van der Werf ,Isabelle Woercel |

| | | | |
|---|---|---|---|
| EPI_ISL_2535559 | Hospital | National Reference Center for Viruses of Respiratory Infections, Institut Pasteur, Paris | Marion Barbet, Sylvie Behillil, Méline Bizard, Angela Brisebarre, Camille Capel, Vincent Enouf, Louise Lefrançois, Frédéric Lemoine, Christophe Malabat, Corinne Maufrais, Etienne Simon-Lorière, Maud Vanpeene, Sylvie Van der Werf ,CéCile Farrugia |
| EPI_ISL_2535560 | Labo Analyses Med | National Reference Center for Viruses of Respiratory Infections, Institut Pasteur, Paris | Marion Barbet, Sylvie Behillil, Méline Bizard, Angela Brisebarre, Camille Capel, Vincent Enouf, Louise Lefrançois, Frédéric Lemoine, Christophe Malabat, Corinne Maufrais, Etienne Simon-Lorière, Maud Vanpeene, Sylvie Van der Werf ,Benedicte Baccouch |
| EPI_ISL_2536001 | Gravity Diagnostics, LLC | Gravity Diagnostics, LLC | Gravity Diagnostics |
| EPI_ISL_2538830 | Lighthouse Lab in Milton Keynes | Wellcome Sanger Institute for the COVID-19 Genomics UK (COG-UK) Consortium | The Lighthouse Lab in Milton Keynes and Alex Alderton, Roberto Amato, Jeffrey Barrett, Sonia Goncalves, Ewan Harrison, David K. Jackson, Ian Johnston, Dominic Kwiatkowski, Cordelia Langford, John Sillitoe on behalf of the Wellcome Sanger Institute COVID-19 Surveillance Team |
| EPI_ISL_2543479 | Genetica Molecular and Subdepartamento de Virologia ISP Chile | Instituto de Salud Publica de Chile | Karen Orostica, Constanza Campano, Barbara Parra, Loredana Arata, Gisselle Barra, Patricia Bustos, Rodrigo Fasce, Javier Tognarelli, Andres Castillo, Soledad Ulloa, Jorge Fernandez |
| EPI_ISL_2544259 | Invenimus AG | Institute of Medical Virology | Verena Kufner, Gabriela Ziltener, Maryam Zaheri, Stefan Schmutz, Annette Audigé, Maria Grünberg, Kevin Steiner, Jon Huder, Cyril Shah, Riccarda Capaul, Guido Bloemberg, Jürg Böni, Michael Huber, Alexandra Trkola |
| EPI_ISL_2544783 | MD PHL | MD PHL | Maryland Department of Health Laboratories Administration |
| EPI_ISL_2544916, EPI_ISL_2544922, EPI_ISL_2544951, EPI_ISL_2544960, EPI_ISL_2544970 | UW Virology Lab | UW Virology Lab | Pavitra Roychoudhury, Hong Xie, Lasata Shrestha, Shah Mohamed Bakhash, Tien V. Nguyen, Noah R. Baker, Sean Ellis, Meei-Li Huang, Keith R Jerome, Alexander Greninger |
| EPI_ISL_2545648 | LHUB-ULB | Labo Klinische Biologie, UZA | Jasmine Coppens, Marie Le Mercier, Basil Britto Xavier, Christine Lammens, Veerle Matheeussen, Herman Goossens |
| EPI_ISL_2547365 | Calvinia Hospital | National Health Laboratory Service/University of Cape Town (NHLS/UCT) | Arash Iranzadeh, Deelan Doolabh, Lynn Tyers, Bruna Galvao, Innocent Mudau, Marvin Hsiao, Gert Marais, Diana Hardie, Stephen Korsman, Carolyn Williamson |
| EPI_ISL_2547367 | Vredendal North Clinic wc VDN | National Health Laboratory Service/University of Cape Town (NHLS/UCT) | Arash Iranzadeh, Deelan Doolabh, Lynn Tyers, Bruna Galvao, Innocent Mudau, Marvin Hsiao, Gert Marais, Diana Hardie, Stephen Korsman, Carolyn Williamson |
| EPI_ISL_2547370 | Kuruman Hospital | National Health Laboratory Service/University of Cape Town (NHLS/UCT) | Arash Iranzadeh, Deelan Doolabh, Lynn Tyers, Bruna Galvao, Innocent Mudau, Marvin Hsiao, Gert Marais, Diana Hardie, Stephen Korsman, Carolyn Williamson |
| EPI_ISL_2547371 | Postdene Clinic | National Health Laboratory Service/University of Cape Town (NHLS/UCT) | Arash Iranzadeh, Deelan Doolabh, Lynn Tyers, Bruna Galvao, Innocent Mudau, Marvin Hsiao, Gert Marais, Diana Hardie, Stephen Korsman, Carolyn Williamson |
| EPI_ISL_2547411, EPI_ISL_2547413 | 2 Military Hospital wc MAA | National Health Laboratory Service/University of Cape Town (NHLS/UCT) | Arash Iranzadeh, Deelan Doolabh, Lynn Tyers, Bruna Galvao, Innocent Mudau, Marvin Hsiao, Gert Marais, Diana Hardie, Stephen Korsman, Carolyn Williamson |
| EPI_ISL_2547415 | Knysna Hospital wc KNY | National Health Laboratory Service/University of Cape Town (NHLS/UCT) | Arash Iranzadeh, Deelan Doolabh, Lynn Tyers, Bruna Galvao, Innocent Mudau, Marvin Hsiao, Gert Marais, Diana Hardie, Stephen Korsman, Carolyn Williamson |
| EPI_ISL_2547418, EPI_ISL_2547419 | Groote Schuur Hospital wc GSH | National Health Laboratory Service/University of Cape Town (NHLS/UCT) | Arash Iranzadeh, Deelan Doolabh, Lynn Tyers, Bruna Galvao, Innocent Mudau, Marvin Hsiao, Gert Marais, Diana Hardie, Stephen Korsman, Carolyn Williamson |
| EPI_ISL_2547420 | Nelspoort Clinic wc NPP | National Health Laboratory Service/University of Cape Town (NHLS/UCT) | Arash Iranzadeh, Deelan Doolabh, Lynn Tyers, Bruna Galvao, Innocent Mudau, Marvin Hsiao, Gert Marais, Diana Hardie, Stephen Korsman, Carolyn Williamson |
| EPI_ISL_2547421, EPI_ISL_2547422 | 2 Military Hospital wc MAA | National Health Laboratory Service/University of Cape Town (NHLS/UCT) | Arash Iranzadeh, Deelan Doolabh, Lynn Tyers, Bruna Galvao, Innocent Mudau, Marvin Hsiao, Gert Marais, Diana Hardie, Stephen Korsman, Carolyn Williamson |
| EPI_ISL_2547424, EPI_ISL_2547425 | Groote Schuur Hospital wc GSH | National Health Laboratory Service/University of Cape Town (NHLS/UCT) | Arash Iranzadeh, Deelan Doolabh, Lynn Tyers, Bruna Galvao, Innocent Mudau, Marvin Hsiao, Gert Marais, Diana Hardie, Stephen Korsman, Carolyn Williamson |
| EPI_ISL_2547426 | Great Brak River Clinic wc GBC | National Health Laboratory Service/University of Cape Town (NHLS/UCT) | Arash Iranzadeh, Deelan Doolabh, Lynn Tyers, Bruna Galvao, Innocent Mudau, Marvin Hsiao, Gert Marais, Diana Hardie, Stephen Korsman, Carolyn Williamson |
| EPI_ISL_2547427 | Hornlee Clinic wc HLC | National Health Laboratory Service/University of Cape Town (NHLS/UCT) | Arash Iranzadeh, Deelan Doolabh, Lynn Tyers, Bruna Galvao, Innocent Mudau, Marvin Hsiao, Gert Marais, Diana Hardie, Stephen Korsman, Carolyn Williamson |
| EPI_ISL_2549077 | Furst Medical Laboratory | Norwegian Institute of Public Health, Department of Virology | Kathrine Stene-Johansen, Kamilla Heddeland Instefjord, Hilde Elshaug, Garcia Llorente Ignacio, Jon Bråte, Engebretsen Serina Beate,Pedersen Benedikte Nevjen, Line Victoria Moen, Debech Nadia, Atiya R Ali,Marie Paulsen Madsen, Rasmus Riis Kopperud, Hilde Vollan, Karoline Bragstad, Olav Hungnes |
| EPI_ISL_2550817 | Emory Molecular Diagnostics Laboratory, Emory Healthcare | Piantadosi Lab, Emory Department of Pathology | Ahmed Babiker, Dara Khosravi, Anne Piantadosi |

| | | | |
|---|---|---|---|
| EPI_ISL_2554885 | Lighthouse Lab in Alderley Park | Wellcome Sanger Institute for the COVID-19 Genomics UK (COG-UK) Consortium | Jacquelyn Wynn, Mairead Hyland, The Lighthouse Lab in Alderley Park and Alex Alderton, Roberto Amato, Jeffrey Barrett, Sonia Goncalves, Ewan Harrison, David K. Jackson, Ian Johnston, Dominic Kwiatkowski, Cordelia Langford, John Sillitoe on behalf of the Wellcome Sanger Institute COVID-19 Surveillance Team |
| EPI_ISL_2557932 | Oregon State Public Health Laboratory | Oregon State Public Health Laboratory | Rafia Razzaque, Eugene Yeboah, Vanda Makris, Laura Tsaknaridis, John Fontana and Shane Sevey |
| EPI_ISL_2558623 | Wisconsin State Laboratory of Hygiene Communicable Disease Division | Wisconsin State Laboratory of Hygiene Communicable Disease Division | Abigail C. Shockey, Alicia J. Mooney, Erika M. Hanson, Tonya Danz, Richard Griesser, Sara Wagner, Kelsey R. Florek |
| EPI_ISL_2562728 | NC State Laboratory of Public Health | Genomics and Discovery, Respiratory Viruses Branch, Division of Viral Diseases, Centers for Disease Control and Prevention | Yan Li, Jing Zhang, Anna Kelleher, Ying Tao, Brian Lynch, Krista Queen, Anna Uehara, Peter Cook, Han Jia Justin Ng, Rachel Marine, Clinton R. Paden, Dhwani Batra, Haibin Wang, Tara Coalter, Jasmine Padilla, Morgan Davis, Mili Sheth, Sarah Nobles, Mark Burroughs, Justin Lee, Adam Retchless, Suxiang Tong |
| EPI_ISL_2566449 | Department of Public Health Bucharest | National Institute of Infectious Diseases-Prof. Dr. Matei Bals Molecular Diagnostics Laboratory | Corina Casangiu, Leontina Banica, Marius Surleac, Petre Milu, Robert Hohan, Simona Paraschiv, Dan Otelea |
| EPI_ISL_2566591 | Hospital Universitari Joan XXIII | Hospital Universitari Joan XXIII | Gemma Recio, Cristina Gutiérrez, Clara Benavent, Carla Martín, Margarida Terrón, Ester Picó, Natalia Bastón |
| EPI_ISL_2567051, EPI_ISL_2567063 | Shamir Medical Center (Asaf Harofe) | Shamir Medical Center (Asaf Harofe) | Nir Rainy,Chen Weiner,Reut Sorek Abramovich,Yevgeni Yegorov,Anna Vishnevsky,Patricia Benveniste-Lekovitz,Abu Hamad Ramzia,Adina Bar Chaim |
| EPI_ISL_2567368 | CENTRE HOSPITALIER SUD GIRONDE | CNR Virus des Infections Respiratoires - France SUD | Antonin Bal, Gregory Destras, Gwendolyne Burfin, Hadrien Regue, Quentin Semanas, Martine Valette, Bruno Lina, Laurence Josset |
| EPI_ISL_2567373 | CENTRE HOSPITALIER POITIERS | CNR Virus des Infections Respiratoires - France SUD | Antonin Bal, Gregory Destras, Gwendolyne Burfin, Hadrien Regue, Quentin Semanas, Martine Valette, Bruno Lina, Laurence Josset |
| EPI_ISL_2567384, EPI_ISL_2567391 | LBM UNIBIO VALENTIN | CNR Virus des Infections Respiratoires - France SUD | Antonin Bal, Gregory Destras, Gwendolyne Burfin, Hadrien Regue, Quentin Semanas, Martine Valette, Bruno Lina, Laurence Josset |
| EPI_ISL_2567437, EPI_ISL_2567460 | MIRIALIS CLUSES BECHET | CNR Virus des Infections Respiratoires - France SUD | Antonin Bal, Gregory Destras, Gwendolyne Burfin, Hadrien Regue, Quentin Semanas, Martine Valette, Bruno Lina, Laurence Josset |
| EPI_ISL_2567463, EPI_ISL_2567464, EPI_ISL_2567465 | GROUPE LCD - UNIBIO LA PLAINE | CNR Virus des Infections Respiratoires - France SUD | Antonin Bal, Gregory Destras, Gwendolyne Burfin, Hadrien Regue, Quentin Semanas, Martine Valette, Bruno Lina, Laurence Josset |
| EPI_ISL_2567469 | BIOGROUP LABORATOIRE ST DIE | CNR Virus des Infections Respiratoires - France SUD | Antonin Bal, Gregory Destras, Gwendolyne Burfin, Hadrien Regue, Quentin Semanas, Martine Valette, Bruno Lina, Laurence Josset |
| EPI_ISL_2567508 | LBM UNIBIO VALENTIN | CNR Virus des Infections Respiratoires - France SUD | Antonin Bal, Gregory Destras, Gwendolyne Burfin, Hadrien Regue, Quentin Semanas, Martine Valette, Bruno Lina, Laurence Josset |
| EPI_ISL_2568858, EPI_ISL_2570464 | Lighthouse Lab in Milton Keynes | Wellcome Sanger Institute for the COVID-19 Genomics UK (COG-UK) Consortium | The Lighthouse Lab in Milton Keynes and Alex Alderton, Roberto Amato, Jeffrey Barrett, Sonia Goncalves, Ewan Harrison, David K. Jackson, Ian Johnston, Dominic Kwiatkowski, Cordelia Langford, John Sillitoe on behalf of the Wellcome Sanger Institute COVID-19 Surveillance Team |
| EPI_ISL_2574071 | Center for Laboratory Medicine | Center for Laboratory Medicine | Yannick Gerth |
| EPI_ISL_2598363 | Hospital General Universitario de Alicante - Instituto de Investigación Sanitaria y Biomédica de Alicante | SeqCOVID-SPAIN consortium/IBV(CSIC) | Maripaz Ventero Martín, Carmen Molina Pardines and SeqCOVID-SPAIN consortium |
| EPI_ISL_2599733 | Quest Diagnostics Incorporated | Centers for Disease Control and Prevention Division of Viral Diseases, Pathogen Discovery | Dakota Howard, Dhwani Batra, Peter W. Cook, Kara Moser, Adrian Paskey, Jason Caravas, Benjamin Rambo-Martin, Shatavia Morrison, Christopher Gulvick, Scott Sammons, Yvette Unoarumhi, Darlene Wagner, Matthew Schmerer, S. H. Rosenthal, A. Gerasimova, R. M. Kagan, B. Anderson, M. Hua, Y. Liu, L.E. Bernstein, K.E. Livingston, A. Perez, I. A. Shlyakhter, R. V. Rolando, R. Owen, P. Tanpaiboon, F. Lacbawan, Clinton R. Paden, Duncan MacCannell |
| EPI_ISL_2599792 | Dianalabs SA | Genesupport | Tanguy ARAUD, Geraldine Jost, Katia Jaton, Nadia Liassine |
| EPI_ISL_2599879 | Quest Diagnostics Incorporated | Centers for Disease Control and Prevention Division of Viral Diseases, Pathogen Discovery | Dakota Howard, Dhwani Batra, Peter W. Cook, Kara Moser, Adrian Paskey, Jason Caravas, Benjamin Rambo-Martin, Shatavia Morrison, Christopher Gulvick, Scott Sammons, Yvette Unoarumhi, Darlene Wagner, Matthew Schmerer, S. H. Rosenthal, A. Gerasimova, R. M. Kagan, B. Anderson, M. Hua, Y. Liu, L.E. Bernstein, K.E. Livingston, A. Perez, I. A. Shlyakhter, R. V. Rolando, R. Owen, P. Tanpaiboon, F. Lacbawan, Clinton R. Paden, Duncan MacCannell |
| EPI_ISL_2600368 | Institute of Epidemiology, Disease Control and Research (IEDCR) | IEDCR-ideSHi-icddr,b | Hassan Afrad, Sadia Rahman, Manjur Hossain Khan, Firdausi Qadri, Tahmina Shirin |
| EPI_ISL_2602434 | Laboratorio di Genetica Medica Ospedale Belcolle | INMI Lazzaro Spallanzani IRCCS | F Santini, F Messina, O Butera, M Rueca, S Donzelli, M Pallocca, F Natoni, G Pessina, E Giombini |
| EPI_ISL_2602697, EPI_ISL_2602707 | KEMRI-Wellcome Trust Research Programme,Kilifi | KEMRI-Wellcome Trust Research Programme,Kilifi | Githinji G.,Mohamed K.S.,de Laurent Z.,Mburu M.W.,Matoke D.,Onyango C., |
| EPI_ISL_2606787 | Lighthouse Lab in Milton Keynes | Wellcome Sanger Institute for the COVID-19 Genomics UK (COG-UK) Consortium | The Lighthouse Lab in Milton Keynes and Alex Alderton, Roberto Amato, Jeffrey Barrett, Sonia Goncalves, Ewan Harrison, David K. Jackson, Ian Johnston, Dominic Kwiatkowski, Cordelia Langford, John Sillitoe on behalf of the Wellcome Sanger Institute COVID-19 Surveillance Team |

| | | | |
|---|---|---|---|
| EPI_ISL_2612528 | Centracare Laboratory Services | Minnesota Department of Health, Public Health Laboratory | Alexandra Lorentz, Jacob Garfin, Matt Plumb, and Xiong Wang |
| EPI_ISL_2612834, EPI_ISL_2612835 | SYNLAB | GIGA Medical Genomics | Keith Durkin, Maria Artesi, Sébastien Bontems, Raphaël Boreux, Bouchra Boujemla, Nathalie Renotte, Cécile Meex, Pierrette Melin, Marie-Pierre Hayette, Vincent Bours |
| EPI_ISL_2615283, EPI_ISL_2615285, EPI_ISL_2615335, EPI_ISL_2615385 | Swedish national genomic surveillance program of SARS-CoV-2 | The Public Health Agency of Sweden | Maximilian Riess, Maria Lind Karlberg, Alma Brolund, Swedish national genomic surveillance program of SARS-CoV-2 |
| EPI_ISL_2615889 | Berkeley Medical Center | WVU and Marshall University Combined Genomics Core Facilities | James Denvir, Peter Stoilov, Peter Perrotta, Wesley Kimble, Ryan Percifield |
| EPI_ISL_2616364 | Austrian Agency for Health and Food Safety (AGES) | Bergthaler laboratory, CeMM Research Center for Molecular Medicine of the Austrian Academy of Sciences | Lukas Endler, Anna Schedl, Fabian Amman, Petr Triska, Matthew Thornton, Thomas Penz, Benedikt Agerer, Maelle Le Moing, Michael Schuster, Bekir Erguner, Jan Laine, Martin Senekowitsch, Christoph Bock, Andreas Bergthaler |
| EPI_ISL_2618719 | UW Virology Lab | UW Virology Lab | Pavitra Roychoudhury, Hong Xie, Lasata Shrestha, Shah Mohamed Bakhash, Tien V. Nguyen, Noah R. Baker, Sean Ellis, Meei-Li Huang, Keith R Jerome, Alexander Greninger |
| EPI_ISL_2618857, EPI_ISL_2618993, EPI_ISL_2619469, EPI_ISL_2619470, EPI_ISL_2619551, EPI_ISL_2619560, EPI_ISL_2619730, EPI_ISL_2619773, EPI_ISL_2619778, EPI_ISL_2619968, EPI_ISL_2619981, EPI_ISL_2620413 | | | |
| see above | Swedish national genomic surveillance program of SARS-CoV-2 | The Public Health Agency of Sweden | Maximilian Riess, Maria Lind Karlberg, Alma Brolund, Swedish national genomic surveillance program of SARS-CoV-2 |
| EPI_ISL_2621132, EPI_ISL_2621133 | Groote Schuur Hospital wc GSH | NHLS/UCT | Arash Iranzadeh, Deelan Doolabh, Lynn Tyers, Bruna Galvao, Innocent Mudau, Marvin Hsiao, Gert Marais, Diana Hardie, Stephen Korsman, Carolyn Williamson |
| EPI_ISL_2621136 | 2 Military Hospital wc MAA | NHLS/UCT | Arash Iranzadeh, Deelan Doolabh, Lynn Tyers, Bruna Galvao, Innocent Mudau, Marvin Hsiao, Gert Marais, Diana Hardie, Stephen Korsman, Carolyn Williamson |
| EPI_ISL_2621137 | George Hospital wc GRH | NHLS/UCT | Arash Iranzadeh, Deelan Doolabh, Lynn Tyers, Bruna Galvao, Innocent Mudau, Marvin Hsiao, Gert Marais, Diana Hardie, Stephen Korsman, Carolyn Williamson |
| EPI_ISL_2621143 | D'Almeida Clinic wc DAL | NHLS/UCT | Arash Iranzadeh, Deelan Doolabh, Lynn Tyers, Bruna Galvao, Innocent Mudau, Marvin Hsiao, Gert Marais, Diana Hardie, Stephen Korsman, Carolyn Williamson |
| EPI_ISL_2621144 | 2 Military Hospital wc MAA | NHLS/UCT | Arash Iranzadeh, Deelan Doolabh, Lynn Tyers, Bruna Galvao, Innocent Mudau, Marvin Hsiao, Gert Marais, Diana Hardie, Stephen Korsman, Carolyn Williamson |
| EPI_ISL_2621145 | Knysna CDC wc WLC | NHLS/UCT | Arash Iranzadeh, Deelan Doolabh, Lynn Tyers, Bruna Galvao, Innocent Mudau, Marvin Hsiao, Gert Marais, Diana Hardie, Stephen Korsman, Carolyn Williamson |
| EPI_ISL_2621146 | Dysselsdorp Clinic wc DDC | NHLS/UCT | Arash Iranzadeh, Deelan Doolabh, Lynn Tyers, Bruna Galvao, Innocent Mudau, Marvin Hsiao, Gert Marais, Diana Hardie, Stephen Korsman, Carolyn Williamson |
| EPI_ISL_2621738, EPI_ISL_2621752, EPI_ISL_2621763, EPI_ISL_2621765, EPI_ISL_2621769, EPI_ISL_2621773 | SARS-CoV-2 testing team, National Institute of Infectious Diseases | Pathogen Genomics Center, National Institute of Infectious Diseases | Tsuyoshi Sekizuka, Kentaro Itokawa, Rina Tanaka, Masanori Hashino, Nozomu Hanaoka, Masumichi Saito, Naomi Nojiri, Hazuka Y Furihata, Sana Uchikoba, Hiromizu Takahashi, Tsuguto Fujimoto, Makoto Kuroda |
| EPI_ISL_2621921 | National Public Health Laboratory, National Centre for Infectious Diseases | National Public Health Laboratory, National Centre for Infectious Diseases | Tze Minn Mak, Zhenyang Zhou, Royce Ang, Lin Cui, Raymond Tzer Pin Lin |
| EPI_ISL_2622100 | CHUV | Laboratory of genomics and metagenomics | Trestan Pillonel, Damien Jacot, Sébastien Aeby, Gilbert Greub, Claire Bertelli |
| EPI_ISL_2623677 | Quadram Institute Bioscience | COVID-19 Genomics UK (COG-UK) Consortium | Dave J. Baker, Gemma L. Kay, Alp Aydin, Thanh Le-Viet, Steven Rudder, Ana P. Tedim, Anastasia Kolyva, Maria Diaz, Leonardo de Oliveira Martins, Nabil-Fareed Alikhan, Lizzie Meadows, Rachael Stanley, Ngozi Elumogo, Muhammed Yasir, Nicholas M. Thomson, Alexander J Trotter, Rachel Gilroy, Samuel Bloomfield, Claire Stuart, Andrew Bell, Reenesh Prakash, Samir Dervisevic, Alison E. Mather, John Wain, Mark Webber, Andrew J. Page, Justin O'Grady |
| EPI_ISL_2624573 | Oxford Viromics, NDM, University of Oxford; Oxford University Hospitals; Basingstoke and North Hampshire Hospital | COVID-19 Genomics UK (COG-UK) Consortium | Tanya Golubchik, David Bonsall, George Macintyre, Amy Trebes, Mariateresa de Cesare, Catrin Moore, Alex Mobbs, Anita Justice, Robert Shaw, Monique Andersson, Timothy Peto, Emma Wise, Nathan Moore, Jessica Lynch, Nick Cortes, Matilde Mori, Stephen Kidd, David Buck, John Todd, Christophe Fraser |
| EPI_ISL_2626081 | Clinique Saint-Pierre Ottignies | UCLouvain/IREC/MBLG | Jean Ruelle, Bertrand Bearzatto, Benoit Kabamba Mukadi |
| EPI_ISL_2626175 | HOSPITAL UNIVERSITARIO SON ESPASES | HOSPITAL UNIVERSITARIO SON ESPASES | Servicio de Microbiología, Hospital Universitario Son Espases |
| EPI_ISL_2626181 | Usansolo-Galdakao University Hospital | Cruces University Hospital | Ana Gual-de-Torrella, Izaskun Alejo-Cancho, Mikel Gallego, Ana Belén de la Hoz |
| EPI_ISL_2626872 | CENTRUM VOOR MEDISCHE ANALYSE | Labo Klinische Biologie, UZA | Marie Le Mercier, Jasmine Coppens, Basil Britto Xavier, Christine Lammens, Veerle Matheeussen, Herman Goossens |
| EPI_ISL_2626938 | TEMPUS LABS INC | Wadsworth Center, New York State Department of Health | Kirsten St. George, Daryl M. Lamson, Alexis Russell, Matthew Shudt, Melissa A Leisner, Jonathan Plitnick, Catharine Prussing, Navjot Singh, John Kelly, Erasmus Schneider, Erica Lasek-Nesselquist |
| EPI_ISL_2627001 | ALBANY MEDICAL CENTER | Wadsworth Center, New York State Department of Health | Kirsten St. George, Daryl M. Lamson, Alexis Russell, Matthew Shudt, Melissa A Leisner, Jonathan Plitnick, Catharine Prussing, Navjot Singh, John Kelly, Erasmus Schneider, Erica Lasek-Nesselquist |
| EPI_ISL_2627062 | URMC LABS | Wadsworth Center, New York State Department of Health | Kirsten St. George, Daryl M. Lamson, Alexis Russell, Matthew Shudt, Melissa A Leisner, Jonathan Plitnick, Catharine Prussing, Navjot Singh, John Kelly, Erasmus Schneider, Erica Lasek-Nesselquist |

| | | | |
|---|---|---|---|
| EPI_ISL_2627948 | Hospital | National Reference Center for Viruses of Respiratory Infections, Institut Pasteur, Paris | Marion Barbet, Sylvie Behillil, Méline Bizard, Angela Brisebarre, Camille Capel, Vincent Enouf, Louise Lefrançois, Frédéric Lemoine, Christophe Malabat, Corinne Maufrais, Etienne Simon-Lorière, Maud Vanpeene, Sylvie Van der Werf ,ValéRie Serazin |
| EPI_ISL_2627953, EPI_ISL_2627954, EPI_ISL_2627955 | Hospital | National Reference Center for Viruses of Respiratory Infections, Institut Pasteur, Paris | Marion Barbet, Sylvie Behillil, Méline Bizard, Angela Brisebarre, Camille Capel, Vincent Enouf, Louise Lefrançois, Frédéric Lemoine, Christophe Malabat, Corinne Maufrais, Etienne Simon-Lorière, Maud Vanpeene, Sylvie Van der Werf ,Pascale Martres |
| EPI_ISL_2627961, EPI_ISL_2627963 | Hospital | National Reference Center for Viruses of Respiratory Infections, Institut Pasteur, Paris | Marion Barbet, Sylvie Behillil, Méline Bizard, Angela Brisebarre, Camille Capel, Vincent Enouf, Louise Lefrançois, Frédéric Lemoine, Christophe Malabat, Corinne Maufrais, Etienne Simon-Lorière, Maud Vanpeene, Sylvie Van der Werf ,JéRome Guinard |
| EPI_ISL_2627975, EPI_ISL_2627976, EPI_ISL_2627978 | Hospital | National Reference Center for Viruses of Respiratory Infections, Institut Pasteur, Paris | Marion Barbet, Sylvie Behillil, Méline Bizard, Angela Brisebarre, Camille Capel, Vincent Enouf, Louise Lefrançois, Frédéric Lemoine, Christophe Malabat, Corinne Maufrais, Etienne Simon-Lorière, Maud Vanpeene, Sylvie Van der Werf ,Pascale Martres |
| EPI_ISL_2627993, EPI_ISL_2627994 | Hospital | National Reference Center for Viruses of Respiratory Infections, Institut Pasteur, Paris | Marion Barbet, Sylvie Behillil, Méline Bizard, Angela Brisebarre, Camille Capel, Vincent Enouf, Louise Lefrançois, Frédéric Lemoine, Christophe Malabat, Corinne Maufrais, Etienne Simon-Lorière, Maud Vanpeene, Sylvie Van der Werf ,Nabil Gastli |
| EPI_ISL_2628151 | Hospital Universitari Vall d'Hebron - Vall d'Hebron Institut de Recerca | Hospital Universitari Vall d'Hebron - Vall d'Hebron Institut de Recerca | Cristina Andrés, Maria Piñana, Alejandra González-Sánchez, Damir Garcia-Cehic, Ariadna Rando, Juliana Esperalba, Maria Gema Codina, Carla Castillo, Maria Carmen Martin, Tomàs Pumarola, Josep Quer, Andrés Antón |
| EPI_ISL_2629257, EPI_ISL_2629313 | LAM GEN-BIO GRAVANCHES | CNR Virus des Infections Respiratoires - France SUD | Antonin Bal, Gregory Destras, Gwendolyne Burfin, Hadrien Regue, Quentin Semanas, Martine Valette, Bruno Lina, Laurence Josset |
| EPI_ISL_2629520 | PLATEAU TECHNIQUE L'ABO+ | CNR Virus des Infections Respiratoires - France SUD | Antonin Bal, Gregory Destras, Gwendolyne Burfin, Hadrien Regue, Quentin Semanas, Martine Valette, Bruno Lina, Laurence Josset |
| EPI_ISL_2629523, EPI_ISL_2629525, EPI_ISL_2629528, EPI_ISL_2629529, EPI_ISL_2629532, EPI_ISL_2629533 | CERBAILLIANCE LA REUNION | CNR Virus des Infections Respiratoires - France SUD | Antonin Bal, Gregory Destras, Gwendolyne Burfin, Hadrien Regue, Quentin Semanas, Martine Valette, Bruno Lina, Laurence Josset |
| EPI_ISL_2629564 | LABM DU PRE - LABOMAINE | CNR Virus des Infections Respiratoires - France SUD | Antonin Bal, Gregory Destras, Gwendolyne Burfin, Hadrien Regue, Quentin Semanas, Martine Valette, Bruno Lina, Laurence Josset |
| EPI_ISL_2631153 | Oregon State Public Health Laboratory | Oregon State Public Health Laboratory | Rafia Razzaque, Eugene Yeboah, Vanda Makris, Laura Tsaknaridis, John Fontana and Shane Sevey |
| EPI_ISL_2631566, EPI_ISL_2631582 | SYNLAB MVZ Leinfelden-Echterdingen | Robert Koch Institute | unknown |
| EPI_ISL_2631601 | SYNLAB MVZ Leverkusen | Robert Koch Institute | unknown |
| EPI_ISL_2631616, EPI_ISL_2631694 | SYNLAB MVZ Leinfelden-Echterdingen | Robert Koch Institute | unknown |
| EPI_ISL_2631828 | Labor Dr. Wisplinghoff - Köln | Robert Koch Institute | unknown |
| EPI_ISL_2631923, EPI_ISL_2631924, EPI_ISL_2631930, EPI_ISL_2631931 | Eurofins LifeCodexx GmbH | Robert Koch Institute | unknown |
| EPI_ISL_2632328 | Niedersächsisches Landesgesundheitsamt (NLGA) | Robert Koch Institute | unknown |
| EPI_ISL_2632382 | Sonic - Bioscientia - MVZ Labor Saar GmbH | Robert Koch Institute | unknown |
| EPI_ISL_2632814, EPI_ISL_2632815, EPI_ISL_2632822 | Niedersächsisches Landesgesundheitsamt (NLGA) | Robert Koch Institute | unknown |
| EPI_ISL_2633408, EPI_ISL_2633420, EPI_ISL_2633452, EPI_ISL_2633457 | Laborarztpraxis Osnabrück | Robert Koch Institute | unknown |
| EPI_ISL_2634518, EPI_ISL_2634543 | Eurofins LifeCodexx GmbH | Robert Koch Institute | unknown |
| EPI_ISL_2635142 | Missouri State Public Health Laboratory | Missouri State Public Health Laboratory | Matthew Sinn, Joshua Barry, Ashley New |
| EPI_ISL_2635450 | SYNLAB MVZ Weiden | Robert Koch Institute | unknown |
| EPI_ISL_2635538 | Labor Becker & Kollegen (Standort München) | Robert Koch Institute | unknown |
| EPI_ISL_2635612 | amedes MVZ für Labordiagnostik Rhein-Main | Robert Koch Institute | unknown |
| EPI_ISL_2635908, EPI_ISL_2635920 | Labor Becker & Kollegen (Standort München) | Robert Koch Institute | unknown |
| EPI_ISL_2636152 | Israel Central Virology laboratory | Israel National Consortium for SARS-CoV-2 sequencing | Neta Zuckerman, Efrat Dahan Bucris, Michal Mandelboim, Dana Bar-Ilan, Miranda Geva, Oran Erster, Efrat Glick-Saar, Omri Nayshool, Gideon Rechavi, Ella Mendelson, Orna Mor |
| EPI_ISL_2636232, EPI_ISL_2636245, EPI_ISL_2636303 | Diagnosticum - Labor Neukirchen | Robert Koch Institute | unknown |

| | | | |
|---|---|---|---|
| EPI_ISL_2636305 | Israel Central Virology laboratory | Israel National Consortium for SARS-CoV-2 sequencing | Neta Zuckerman, Efrat Dahan Bucris, Michal Mandelboim, Dana Bar-Ilan, Miranda Geva, Oran Erster, Efrat Glick-Saar, Omri Nayshool, Gideon Rechavi, Ella Mendelson, Orna Mor |
| EPI_ISL_2636320, EPI_ISL_2636327 | Diagnosticum - Labor Neukirchen | Robert Koch Institute | unknown |
| EPI_ISL_2636778 | MVZ Labor Krone GbR | Robert Koch Institute | unknown |
| EPI_ISL_2636891 | Laborarztpraxis Osnabrück | Robert Koch Institute | unknown |
| EPI_ISL_2637364 | CLILAB | Microbiology Department | Sara Marti, Aida Gonzalez-Diaz, Laura Calatayud, Jordi Niubó, Miguel Fernandez-Huerta, Carmen Ardanuy, Jordi Camara, M Angeles Domínguez |
| EPI_ISL_2637398 | Hôpital Bichat-Claude Bernard | Department of Virology, Henri Mondor University Hospital, Assistance Publique Hôpitaux de Paris, Université Paris-Est Créteil, INSERM U955 | Christophe Rodriguez, Slim Fourati, Vanessa Demontant, Guillaume Gricourt, Melissa N'Debi, Alexandre Soulier, Elisabeth Trawinski, Jean-Michel Pawlotsky |
| EPI_ISL_2637399 | Hôpital Avicenne | Department of Virology, Henri Mondor University Hospital, Assistance Publique Hôpitaux de Paris, Université Paris-Est Créteil, INSERM U955 | Christophe Rodriguez, Slim Fourati, Vanessa Demontant, Guillaume Gricourt, Melissa N'Debi, Alexandre Soulier, Elisabeth Trawinski, Jean-Michel Pawlotsky |
| EPI_ISL_2637437 | laboratoire Belle Epine | Department of Virology, Henri Mondor University Hospital, Assistance Publique Hôpitaux de Paris, Université Paris-Est Créteil, INSERM U955 | Christophe Rodriguez, Slim Fourati, Vanessa Demontant, Guillaume Gricourt, Melissa N'Debi, Alexandre Soulier, Elisabeth Trawinski, Jean-Michel Pawlotsky |
| EPI_ISL_2637449 | Hôpital Ambroise-Paré | Department of Virology, Henri Mondor University Hospital, Assistance Publique Hôpitaux de Paris, Université Paris-Est Créteil, INSERM U955 | Christophe Rodriguez, Slim Fourati, Vanessa Demontant, Guillaume Gricourt, Melissa N'Debi, Alexandre Soulier, Elisabeth Trawinski, Jean-Michel Pawlotsky |
| EPI_ISL_2637466 | laboratoire Belle Epine | Department of Virology, Henri Mondor University Hospital, Assistance Publique Hôpitaux de Paris, Université Paris-Est Créteil, INSERM U955 | Christophe Rodriguez, Slim Fourati, Vanessa Demontant, Guillaume Gricourt, Melissa N'Debi, Alexandre Soulier, Elisabeth Trawinski, Jean-Michel Pawlotsky |
| EPI_ISL_2637467, EPI_ISL_2637468 | GROUPE HOSPITALIER SUD ILE DE FRANCE | Department of Virology, Henri Mondor University Hospital, Assistance Publique Hôpitaux de Paris, Université Paris-Est Créteil, INSERM U955 | Christophe Rodriguez, Slim Fourati, Vanessa Demontant, Guillaume Gricourt, Melissa N'Debi, Alexandre Soulier, Elisabeth Trawinski, Jean-Michel Pawlotsky |
| EPI_ISL_2637473, EPI_ISL_2637476, EPI_ISL_2637481 | laboratoire Belle Epine | Department of Virology, Henri Mondor University Hospital, Assistance Publique Hôpitaux de Paris, Université Paris-Est Créteil, INSERM U955 | Christophe Rodriguez, Slim Fourati, Vanessa Demontant, Guillaume Gricourt, Melissa N'Debi, Alexandre Soulier, Elisabeth Trawinski, Jean-Michel Pawlotsky |
| EPI_ISL_2637494 | Hôpital Cochin | Department of Virology, Henri Mondor University Hospital, Assistance Publique Hôpitaux de Paris, Université Paris-Est Créteil, INSERM U955 | Christophe Rodriguez, Slim Fourati, Vanessa Demontant, Guillaume Gricourt, Melissa N'Debi, Alexandre Soulier, Elisabeth Trawinski, Jean-Michel Pawlotsky |
| EPI_ISL_2637524 | laboratoire Belle Epine | Department of Virology, Henri Mondor University Hospital, Assistance Publique Hôpitaux de Paris, Université Paris-Est Créteil, INSERM U955 | Christophe Rodriguez, Slim Fourati, Vanessa Demontant, Guillaume Gricourt, Melissa N'Debi, Alexandre Soulier, Elisabeth Trawinski, Jean-Michel Pawlotsky |
| EPI_ISL_2637532, EPI_ISL_2637567 | BIO ARD'AISNE | Department of Virology, Henri Mondor University Hospital, Assistance Publique Hôpitaux de Paris, Université Paris-Est Créteil, INSERM U955 | Christophe Rodriguez, Slim Fourati, Vanessa Demontant, Guillaume Gricourt, Melissa N'Debi, Alexandre Soulier, Elisabeth Trawinski, Jean-Michel Pawlotsky |
| EPI_ISL_2637615 | Hôpital Cochin | Department of Virology, Henri Mondor University Hospital, Assistance Publique Hôpitaux de | Christophe Rodriguez, Slim Fourati, Vanessa Demontant, Guillaume Gricourt, Melissa N'Debi, Alexandre Soulier, Elisabeth Trawinski, Jean-Michel Pawlotsky |

| | | | |
|---|---|---|---|
| | | Paris, Université Paris-Est Créteil, INSERM U955 | |
| EPI_ISL_2637638 | laboratoire Belle Epine | Department of Virology, Henri Mondor University Hospital, Assistance Publique Hôpitaux de Paris, Université Paris-Est Créteil, INSERM U955 | Christophe Rodriguez, Slim Fourati, Vanessa Demontant, Guillaume Gricourt, Melissa N'Debi, Alexandre Soulier, Elisabeth Trawinski, Jean-Michel Pawlotsky |
| EPI_ISL_2637653 | BROUSSAIS | Department of Virology, Henri Mondor University Hospital, Assistance Publique Hôpitaux de Paris, Université Paris-Est Créteil, INSERM U955 | Christophe Rodriguez, Slim Fourati, Vanessa Demontant, Guillaume Gricourt, Melissa N'Debi, Alexandre Soulier, Elisabeth Trawinski, Jean-Michel Pawlotsky |
| EPI_ISL_2637668, EPI_ISL_2637683 | laboratoire Belle Epine | Department of Virology, Henri Mondor University Hospital, Assistance Publique Hôpitaux de Paris, Université Paris-Est Créteil, INSERM U955 | Christophe Rodriguez, Slim Fourati, Vanessa Demontant, Guillaume Gricourt, Melissa N'Debi, Alexandre Soulier, Elisabeth Trawinski, Jean-Michel Pawlotsky |
| EPI_ISL_2637816 | BIOMNIS EUROFINS IVRY | Department of Virology, Henri Mondor University Hospital, Assistance Publique Hôpitaux de Paris, Université Paris-Est Créteil, INSERM U955 | Christophe Rodriguez, Slim Fourati, Vanessa Demontant, Guillaume Gricourt, Melissa N'Debi, Alexandre Soulier, Elisabeth Trawinski, Jean-Michel Pawlotsky |
| EPI_ISL_2637828 | CHU Rouen Normandie | Department of Virology, Henri Mondor University Hospital, Assistance Publique Hôpitaux de Paris, Université Paris-Est Créteil, INSERM U955 | Christophe Rodriguez, Slim Fourati, Vanessa Demontant, Guillaume Gricourt, Melissa N'Debi, Alexandre Soulier, Elisabeth Trawinski, Jean-Michel Pawlotsky |
| EPI_ISL_2637858 | BIO ARD'AISNE | Department of Virology, Henri Mondor University Hospital, Assistance Publique Hôpitaux de Paris, Université Paris-Est Créteil, INSERM U955 | Christophe Rodriguez, Slim Fourati, Vanessa Demontant, Guillaume Gricourt, Melissa N'Debi, Alexandre Soulier, Elisabeth Trawinski, Jean-Michel Pawlotsky |
| EPI_ISL_2637880 | GROUPE HOSPITALIER SUD ILE DE FRANCE | Department of Virology, Henri Mondor University Hospital, Assistance Publique Hôpitaux de Paris, Université Paris-Est Créteil, INSERM U955 | Christophe Rodriguez, Slim Fourati, Vanessa Demontant, Guillaume Gricourt, Melissa N'Debi, Alexandre Soulier, Elisabeth Trawinski, Jean-Michel Pawlotsky |
| EPI_ISL_2637890 | BIOMNIS EUROFINS IVRY | Department of Virology, Henri Mondor University Hospital, Assistance Publique Hôpitaux de Paris, Université Paris-Est Créteil, INSERM U955 | Christophe Rodriguez, Slim Fourati, Vanessa Demontant, Guillaume Gricourt, Melissa N'Debi, Alexandre Soulier, Elisabeth Trawinski, Jean-Michel Pawlotsky |
| EPI_ISL_2637926 | laboratoire Belle Epine | Department of Virology, Henri Mondor University Hospital, Assistance Publique Hôpitaux de Paris, Université Paris-Est Créteil, INSERM U955 | Christophe Rodriguez, Slim Fourati, Vanessa Demontant, Guillaume Gricourt, Melissa N'Debi, Alexandre Soulier, Elisabeth Trawinski, Jean-Michel Pawlotsky |
| EPI_ISL_2637932 | BIO ARD'AISNE | Department of Virology, Henri Mondor University Hospital, Assistance Publique Hôpitaux de Paris, Université Paris-Est Créteil, INSERM U955 | Christophe Rodriguez, Slim Fourati, Vanessa Demontant, Guillaume Gricourt, Melissa N'Debi, Alexandre Soulier, Elisabeth Trawinski, Jean-Michel Pawlotsky |
| EPI_ISL_2637944 | BIOMNIS EUROFINS IVRY | Department of Virology, Henri Mondor University Hospital, Assistance Publique Hôpitaux de Paris, Université Paris-Est Créteil, INSERM U955 | Christophe Rodriguez, Slim Fourati, Vanessa Demontant, Guillaume Gricourt, Melissa N'Debi, Alexandre Soulier, Elisabeth Trawinski, Jean-Michel Pawlotsky |
| EPI_ISL_2638017 | GROUPE HOSPITALIER SUD ILE DE FRANCE | Department of Virology, Henri Mondor University Hospital, Assistance Publique Hôpitaux de | Christophe Rodriguez, Slim Fourati, Vanessa Demontant, Guillaume Gricourt, Melissa N'Debi, Alexandre Soulier, Elisabeth Trawinski, Jean-Michel Pawlotsky |

| | | | |
|---|---|---|---|
| | | Paris, Université Paris-Est Créteil, INSERM U955 | |
| EPI_ISL_2638109 | AULSS 2 Marca Trevigiana | Istituto Zooprofilattico Sperimentale delle Venezie | Adelaide Milani, Alessia Schivo, Annalisa Salviato, Elisa Palumbo, Erika Giorgia Quaranta, Luca Tassoni, Ambra Pastori, Edoardo Giussani, Alice Fusaro, Isabella Monne, Calogero Terregino, Antonia Ricci |
| EPI_ISL_2645092 | Fimlab Laboratoriot Oy Tampere | Expert Microbiology, National Institute for Health and Welfare | Soile Blomqvist, Jani Halkilahti, Kirsi Liitsola, Haider al-Hello, Päivi Laurila, Erika Lindh, Teemu Smura, Ravi Kant, Phuoc Truong, Olli Vapalahti, Sari Hannula, Pekka Ellonen, Niina Ikonen, Carita Savolainen-Kopra |
| EPI_ISL_2645124 | Islab, Pohjois-Savon aluelaboratorio | Expert Microbiology, National Institute for Health and Welfare | Soile Blomqvist, Jani Halkilahti, Kirsi Liitsola, Haider al-Hello, Päivi Laurila, Erika Lindh, Teemu Smura, Ravi Kant, Phuoc Truong, Olli Vapalahti, Sari Hannula, Pekka Ellonen, Niina Ikonen, Carita Savolainen-Kopra |
| EPI_ISL_2645201 | Fimlab Laboratoriot Oy Tampere | Expert Microbiology, National Institute for Health and Welfare | Soile Blomqvist, Jani Halkilahti, Kirsi Liitsola, Haider al-Hello, Päivi Laurila, Erika Lindh, Teemu Smura, Ravi Kant, Phuoc Truong, Olli Vapalahti, Sari Hannula, Pekka Ellonen, Niina Ikonen, Carita Savolainen-Kopra |
| EPI_ISL_2646145 | CH CALAIS | CHU Lille - Laboratoire de Virologie | AIT YAHYA Emilie, ALIDJINOU Enagnon Kazali, BOCKET Laurence, CREPIN Michel, DEMAY Christophe, ENGELMANN Ilka, GEFFROY Sandrine, GUIGON Aurélie, LAMBERT Valérie, LAZREK Mouna, NOBILLIAUX Florian, PREVOST Brigitte, THUILLIER Caroline, TINEZ Claire ,TCHANTCHOU NJOSSE YANICK |
| EPI_ISL_2646175 | CH BETHUNE | CHU Lille - Laboratoire de Virologie | AIT YAHYA Emilie, ALIDJINOU Enagnon Kazali, BOCKET Laurence, CREPIN Michel, DEMAY Christophe, ENGELMANN Ilka, GEFFROY Sandrine, GUIGON Aurélie, LAMBERT Valérie, LAZREK Mouna, NOBILLIAUX Florian, PREVOST Brigitte, THUILLIER Caroline, TINEZ Claire ,TCHANTCHOU NJOSSE YANICK |
| EPI_ISL_2646195 | CH ROUBAIX | CHU Lille - Laboratoire de Virologie | AIT YAHYA Emilie, ALIDJINOU Enagnon Kazali, BOCKET Laurence, CREPIN Michel, DEMAY Christophe, ENGELMANN Ilka, GEFFROY Sandrine, GUIGON Aurélie, LAMBERT Valérie, LAZREK Mouna, NOBILLIAUX Florian, PREVOST Brigitte, THUILLIER Caroline, TINEZ Claire ,TCHANTCHOU NJOSSE YANICK |
| EPI_ISL_2646285 | CHU de la Réunion - Sud | Laboratoire de virologie, CNR arbovirus Associé, Chu de la Réunion | Etienne Frumence, Marie-Christine Jaffar Bandjee, Nicolas Traversier, Sabrina Petit Genet |
| EPI_ISL_2646288 | CHU de la Réunion - Nord | Laboratoire de virologie, CNR arbovirus Associé, Chu de la Réunion | Etienne Frumence, Marie-Christine Jaffar Bandjee, Nicolas Traversier, Sabrina Petit Genet |
| EPI_ISL_2646289, EPI_ISL_2646292, EPI_ISL_2646293, EPI_ISL_2646297 | CHU de la Réunion - Sud | Laboratoire de virologie, CNR arbovirus Associé, Chu de la Réunion | Etienne Frumence, Marie-Christine Jaffar Bandjee, Nicolas Traversier, Sabrina Petit Genet |
| EPI_ISL_2646298 | CHU de la Réunion - Nord | Laboratoire de virologie, CNR arbovirus Associé, Chu de la Réunion | Etienne Frumence, Marie-Christine Jaffar Bandjee, Nicolas Traversier, Sabrina Petit Genet |
| EPI_ISL_2647311 | Mako Medical | Centers for Disease Control and Prevention Division of Viral Diseases, Pathogen Discovery | Dakota Howard, Dhwani Batra, Peter W. Cook, Kara Moser, Adrian Paskey, Jason Caravas, Benjamin Rambo-Martin, Shatavia Morrison, Christopher Gulvick, Scott Sammons, Yvette Unoarumhi, Darlene Wagner, Matthew Schmerer,Matthew Tugwell, Lauren Moon, Clinton R. Paden, Duncan MacCannell |
| EPI_ISL_2648206 | Debswana Orapa Mine Hospital Laboratory | Botswana Harvard HIV Reference Laboratory | Sikhulile Moyo, Wonderful T. Choga, Dorcas Maruapula, Keoratile Ntshambiwa, Lesedi Magama, Mpo Molapisi, Koketso Maotwe, Thongbotho Mphoyakgosi, Boitumelo Zuze, Botshelo Radibe, Legodile Kooepile, Ontlametse T. Bareng, Roger Shapiro, Shahin Lockman, Joseph Makhema, Madisa Mine, Mosepele Mosepele, Simani Gaseitsiwe |
| EPI_ISL_2648239 | Kasane Primary Hospital Laboratory | Botswana Harvard HIV Reference Laboratory | Sikhulile Moyo, Wonderful T. Choga, Dorcas Maruapula, Agnes Karutwaeng, Rose Munyere, Keoratile Ntshambiwa, Thongbotho Mphoyakgosi, Boitumelo Zuze, Botshelo Radibe, Legodile Kooepile, Ontlametse T. Bareng, Roger Shapiro, Shahin Lockman, Joseph Makhema, Madisa Mine, Mosepele Mosepele, Simani Gaseitsiwe |
| EPI_ISL_2649991 | Kudat Hospital | Institute for Medical Research, Infectious Disease Research Centre, National Institutes of Health, Ministry of Health Malaysia | Suppiah J, Kamel K, Mohd Zawawi Z, Azizan MA, Ramly N, Robert F, Thayan R |
| EPI_ISL_2649994 | Tuanku Fauziah Hopsital | Institute for Medical Research, Infectious Disease Research Centre, National Institutes of Health, Ministry of Health Malaysia | Suppiah J, Kamel K, Mohd Zawawi Z, Azizan MA, Ramly N, Robert F, Thayan R |
| EPI_ISL_2649998, EPI_ISL_2649999 | Penang General Hospital | Institute for Medical Research, Infectious Disease Research Centre, National Institutes of Health, Ministry of Health Malaysia | Suppiah J, Kamel K, Mohd Zawawi Z, Azizan MA, Ramly N, Robert F, Thayan R |
| EPI_ISL_2650000 | Sultanah Nur Zahirah Hospital | Institute for Medical Research, Infectious Disease Research Centre, National Institutes of Health, Ministry of Health Malaysia | Suppiah J, Kamel K, Mohd Zawawi Z, Azizan MA, Ramly N, Robert F, Thayan R |

| | | | |
|---|---|---|---|
| EPI_ISL_2650001 | Hospital Sultanah Nur Zahirah Kuala Terengganu | Institute for Medical Research, Infectious Disease Research Centre, National Institutes of Health, Ministry of Health Malaysia | Suppiah J, Kamel K, Mohd Zawawi Z, Azizan MA, Ramly N, Robert F, Thayan R |
| EPI_ISL_2650004, EPI_ISL_2650005 | Sungai Buloh Hospital | Institute for Medical Research, Infectious Disease Research Centre, National Institutes of Health, Ministry of Health Malaysia | Suppiah J, Kamel K, Mohd Zawawi Z, Azizan MA, Ramly N, Robert F, Thayan R |
| EPI_ISL_2650006 | Tumpat Hospital | Institute for Medical Research, Infectious Disease Research Centre, National Institutes of Health, Ministry of Health Malaysia | Suppiah J, Kamel K, Mohd Zawawi Z, Azizan MA, Ramly N, Robert F, Thayan R |
| EPI_ISL_2650007 | Ipoh Public Health Laboratory (MKAI), Ministry of Health Malaysia | Institute for Medical Research, Infectious Disease Research Centre, National Institutes of Health, Ministry of Health Malaysia | Suppiah J, Kamel K, Mohd Zawawi Z, Azizan MA, Ramly N, Robert F, Thayan R |
| EPI_ISL_2650010 | Sungai Buloh Hospital | Institute for Medical Research, Infectious Disease Research Centre, National Institutes of Health, Ministry of Health Malaysia | Suppiah J, Kamel K, Mohd Zawawi Z, Azizan MA, Ramly N, Robert F, Thayan R |
| EPI_ISL_2650028 | Sultanah Aminah Hospital | Institute for Medical Research, Infectious Disease Research Centre, National Institutes of Health, Ministry of Health Malaysia | Suppiah J, Kamel K, Mohd Zawawi Z, Azizan MA, Ramly N, Robert F, Thayan R |
| EPI_ISL_2650250 | Microbiology Department, Laboratori Clínic Metropolitana Nord. Hospital Universitari Germans Trias i Pujol | Can Ruti SARS-CoV-2 Sequencing Hub (HUGTiP/IrsiCaixa/IGTP) | Marc Noguera-Julian, Pilar Armengol, Ignacio Blanco, Antoni E Bordoy, Francesc Catala-Moll, Pere-Joan Cardona, Maria Casadellà, Cristina Casañ, Gemma Clara, Bonaventura Clotet, Cristina Esteban, Montserrat Giménez, Mercedes Guerrero, Anna Not, Roger Paredes, Mariona Parera, Verónica Saludes, Alba Sánchez, and Elisa Martró on behalf of the Can Ruti SARS-CoV-2 Sequencing Hub |
| EPI_ISL_2650259 | Microbiologia CATLAB | Can Ruti SARS-CoV-2 Sequencing Hub (HUGTiP/IrsiCaixa/IGTP) | Marc Noguera-Julian, Pilar Armengol, Ignacio Blanco, Antoni E Bordoy, Francesc Catala-Moll, Pere-Joan Cardona, Maria Casadellà, Cristina Casañ, Gemma Clara, Bonaventura Clotet, Cristina Esteban, Montserrat Giménez, Mercedes Guerrero, Anna Not, Roger Paredes, Mariona Parera, Verónica Saludes, Alba Sánchez, and Elisa Martró on behalf of the Can Ruti SARS-CoV-2 Sequencing Hub |
| EPI_ISL_2650430 | Queen Elizabeth Hospital | Institute for Medical Research, Infectious Disease Research Centre, National Institutes of Health, Ministry of Health Malaysia | Suppiah J, Kamel K, Mohd Zawawi Z, Azizan MA, Ramly N, Robert F, Thayan R |
| EPI_ISL_2650468 | Sydney South West Pathology Service (SSWPS) - Royal Prince Alfred Hospital - NSW Health Pathology | NSW Health Pathology - Institute of Clinical Pathology and Medical Research; Westmead Hospital; University of Sydney | CIDM-PH et al. |
| EPI_ISL_2650499, EPI_ISL_2650505 | Queen Elizabeth Hospital | Institute for Medical Research, Infectious Disease Research Centre, National Institutes of Health, Ministry of Health Malaysia | Suppiah J, Kamel K, Mohd Zawawi Z, Azizan MA, Ramly N, Robert F, Thayan R |
| EPI_ISL_2651123 | Temporary Specimen Collection Centre at the AsiaWorld-Expo | Hong Kong Department of Health | Alan K.L. Tsang, Peter C.W. Yip, Ken H.L. Ng, Edman T.K. Lam, Rickjason C.W. Chan, Dominic N.C. Tsang |
| EPI_ISL_2651266, EPI_ISL_2651267 | CHU de la Réunion - Nord | Laboratoire de virologie, CNR arbovirus Associé, Chu de la Réunion | Etienne Frumence, Marie-Christine Jaffar Bandjee, Nicolas Traversier, Sabrina Petit Genet |
| EPI_ISL_2651270, EPI_ISL_2651274, EPI_ISL_2651275, EPI_ISL_2651276 | CHU de la Réunion - Sud | Laboratoire de virologie, CNR arbovirus Associé, Chu de la Réunion | Etienne Frumence, Marie-Christine Jaffar Bandjee, Nicolas Traversier, Sabrina Petit Genet |
| EPI_ISL_2651279 | CHU de la Réunion - Nord | Laboratoire de virologie, CNR arbovirus Associé, | Etienne Frumence, Marie-Christine Jaffar Bandjee, Nicolas Traversier, Sabrina Petit Genet |

| | | | |
|---|---|---|---|
| | | Chu de la Réunion | |
| EPI_ISL_2651365 | CHU de la Réunion - Sud | Laboratoire de virologie, CNR arbovirus Associé, Chu de la Réunion | Etienne Frumence, Marie-Christine Jaffar Bandjee, Nicolas Traversier, Sabrina Petit Genet |
| EPI_ISL_2651367, EPI_ISL_2651368 | CHU de la Réunion - Nord | Laboratoire de virologie, CNR arbovirus Associé, Chu de la Réunion | Etienne Frumence, Marie-Christine Jaffar Bandjee, Nicolas Traversier, Sabrina Petit Genet |
| EPI_ISL_2651370, EPI_ISL_2651372, EPI_ISL_2651373, EPI_ISL_2651374, EPI_ISL_2651378, EPI_ISL_2651380, EPI_ISL_2651381, EPI_ISL_2651382 | CHU de la Réunion - Sud | Laboratoire de virologie, CNR arbovirus Associé, Chu de la Réunion | Etienne Frumence, Marie-Christine Jaffar Bandjee, Nicolas Traversier, Sabrina Petit Genet |
| EPI_ISL_2651386, EPI_ISL_2651388 | CHU de la Réunion - Nord | Laboratoire de virologie, CNR arbovirus Associé, Chu de la Réunion | Etienne Frumence, Marie-Christine Jaffar Bandjee, Nicolas Traversier, Sabrina Petit Genet |
| EPI_ISL_2651391 | CHU de la Réunion - Sud | Laboratoire de virologie, CNR arbovirus Associé, Chu de la Réunion | Etienne Frumence, Marie-Christine Jaffar Bandjee, Nicolas Traversier, Sabrina Petit Genet |
| EPI_ISL_2651395, EPI_ISL_2651396 | CHU de la Réunion - Nord | Laboratoire de virologie, CNR arbovirus Associé, Chu de la Réunion | Etienne Frumence, Marie-Christine Jaffar Bandjee, Nicolas Traversier, Sabrina Petit Genet |
| EPI_ISL_2651404, EPI_ISL_2651405 | CHU de la Réunion - Sud | Laboratoire de virologie, CNR arbovirus Associé, Chu de la Réunion | Etienne Frumence, Marie-Christine Jaffar Bandjee, Nicolas Traversier, Sabrina Petit Genet |
| EPI_ISL_2652536, EPI_ISL_2652577, EPI_ISL_2652723, EPI_ISL_2652753, EPI_ISL_2652755, EPI_ISL_2652798, EPI_ISL_2652920 | Infinity Biologix | Centers for Disease Control and Prevention Division of Viral Diseases, Pathogen Discovery | Dakota Howard, Dhwani Batra, Peter W. Cook, Kara Moser, Adrian Paskey, Jason Caravas, Benjamin Rambo-Martin, Shatavia Morrison, Christopher Gulvick, Scott Sammons, Yvette Unoarumhi, Darlene Wagner, Matthew Schmerer, Christian Bixby, Yihe Wang, Jonathan Schultz, Chirayu Goswami, Russ Hager, Robin Grimwood, Clinton R. Paden, Duncan MacCannell |
| EPI_ISL_2657919 | Public Health Authority of the Slovak Republic | Laboratory of Genomics and Bioinformatics, Comenius University Science Park | Tatiana Sedláková, Diana Rusáková, Miroslav Böhmer, Anna Giová, Jaroslav Budiš, Tomáš Szemes, Jakub Styk |
| EPI_ISL_2658517 | Minnesota Department of Health, Public Health Laboratory | Minnesota Department of Health, Public Health Laboratory | Alexandra Lorentz, Jacob Garfin, Matt Plumb, and Xiong Wang |
| EPI_ISL_2662306, EPI_ISL_2662429, EPI_ISL_2662437 | Viollier AG | Department of Biosystems Science and Engineering, ETH Zürich | Andrea Patrizia Salzmann, Henriette Kurth, Christiane Beckmann, Ivan Topolsky, Chaoran Chen, Sarah Nadeau, Kim Philipp Jablonski, Lara Fuhrmann, Niko Beerenwinkel, Christoph Noppen, Olivier Kobel, Maurice Redondo, Tanja Stadler |
| EPI_ISL_2662570, EPI_ISL_2662572, EPI_ISL_2662575, EPI_ISL_2662577, EPI_ISL_2662579, EPI_ISL_2662580, EPI_ISL_2662584, EPI_ISL_2662587, EPI_ISL_2662601, EPI_ISL_2662602, EPI_ISL_2662612 | | | |
| see above | TAMBO MEMORIAL LABORATORY | National Institute for Communicable Diseases of the National Health Laboratory Service | Amoako DG, Everatt J, Scheepers C, Mohale T, Ntuli N, Mahlangu B, Mnguni A, Ismail A, Bhiman JN |
| EPI_ISL_2662620, EPI_ISL_2662638, EPI_ISL_2662644, EPI_ISL_2662645, EPI_ISL_2662656 | CHRIS HANI BARAGWANATH LABORATORY | National Institute for Communicable Diseases of the National Health Laboratory Service | Amoako DG, Everatt J, Scheepers C, Mohale T, Ntuli N, Mahlangu B, Mnguni A, Ismail A, Bhiman JN |
| EPI_ISL_2662664 | KOPANONG LABORATORY | National Institute for Communicable Diseases of the National Health Laboratory Service | Amoako DG, Everatt J, Scheepers C, Mohale T, Ntuli N, Mahlangu B, Mnguni A, Ismail A, Bhiman JN |
| EPI_ISL_2662710, EPI_ISL_2662712, EPI_ISL_2662714, EPI_ISL_2662716, EPI_ISL_2662723, EPI_ISL_2662725 | National Institute for Communicable Diseases of the National Health Laboratory Service | National Institute for Communicable Diseases of the National Health Laboratory Service | Amoako DG, Everatt J, Scheepers C, Mohale T, Ntuli N, Mahlangu B, Mnguni A, Ismail A, Bhiman JN |
| EPI_ISL_2665942, EPI_ISL_2670086 | Department of Bacteria, Parasites and Fungi, Statens Serum Institut, Copenhagen, Denmark | Statens Serum Institut Bioinformatics and Microbial Genomics | Danish Covid-19 Genome Consortium |
| EPI_ISL_2674251 | National Institute of Public Health | State Veterinary Institute Prague | Nagy,A;Cernikov,L;Stara,M;Suri,T;Vecerova,J;Jirincova,H |
| EPI_ISL_2674806 | SARS-CoV-2 testing team, National Institute of Infectious Diseases | Pathogen Genomics Center, National Institute of Infectious Diseases | Tsuyoshi Sekizuka, Kentaro Itokawa, Rina Tanaka, Masanori Hashino, Nozomu Hanaoka, Masumichi Saito, Naomi Nojiri, Hazuka Y Furihata, Hiromizu Takahashi, Tsuguto Fujimoto, Makoto Kuroda |
| EPI_ISL_2676579 | LABORATOIRE DE BIOLOGIE MEDICALE | CNR Virus des Infections Respiratoires - France SUD | Antonin Bal, Gregory Destras, Gwendolyne Burfin, Hadrien Regue, Quentin Semanas, Martine Valette, Bruno Lina, Laurence Josset |
| EPI_ISL_2676654, EPI_ISL_2676657, EPI_ISL_2676667, EPI_ISL_2676668, EPI_ISL_2676671 | REUNILAB | CNR Virus des Infections Respiratoires - France SUD | Antonin Bal, Gregory Destras, Gwendolyne Burfin, Hadrien Regue, Quentin Semanas, Martine Valette, Bruno Lina, Laurence Josset |

| | | | |
|---|---|---|---|
| EPI_ISL_2676706, EPI_ISL_2676708 | ASTRALAB TASSIGNY | CNR Virus des Infections Respiratoires - France SUD | Antonin Bal, Gregory Destras, Gwendolyne Burfin, Hadrien Regue, Quentin Semanas, Martine Valette, Bruno Lina, Laurence Josset |
| EPI_ISL_2676713 | CHR LA REUNION FELIX GUYON | CNR Virus des Infections Respiratoires - France SUD | Antonin Bal, Gregory Destras, Gwendolyne Burfin, Hadrien Regue, Quentin Semanas, Martine Valette, Bruno Lina, Laurence Josset |
| EPI_ISL_2676721, EPI_ISL_2676725 | LAM DE SAINT BENOIT | CNR Virus des Infections Respiratoires - France SUD | Antonin Bal, Gregory Destras, Gwendolyne Burfin, Hadrien Regue, Quentin Semanas, Martine Valette, Bruno Lina, Laurence Josset |
| EPI_ISL_2676741 | LBM UNIBIO VALENTIN | CNR Virus des Infections Respiratoires - France SUD | Antonin Bal, Gregory Destras, Gwendolyne Burfin, Hadrien Regue, Quentin Semanas, Martine Valette, Bruno Lina, Laurence Josset |
| EPI_ISL_2678111, EPI_ISL_2678112 | Office of Diseases Prevention and Control Region 12 Songkhla | COVID-19 Network Investigations (CONI) Alliance | Elizabeth Batty, Wasun Chantratita, Thanat Chookajorn, Stefan Fernandez, Angkana Huang, Anthony R. Jones, Khajohn Joonlasak, Chonticha Klungtong, Theerarat Kochakarn, Namfon Kotanan, Krittikorn Kumpornsin, Duangkamon Loesbanluechai, Wudtichai Manasatienkij, Bhakbhoom Panthan, Ekawat Pasomsub, Kingkan Rakmanee, Insee Sensorn, Janjira Thaipadungpanit, Arporn Wangwiwatsin, Treewat Watthanachockchai, Wiphaporn Thabthimthong, Sawanya Chantutanon |
| EPI_ISL_2678988, EPI_ISL_2679374, EPI_ISL_2679376, EPI_ISL_2679441, EPI_ISL_2679473, EPI_ISL_2679475 | Utah Public Health Laboratory | Utah Public Health Laboratory | Erin L. Young, Kelly F. Oakeson, Tara Gallagher |
| EPI_ISL_2684231, EPI_ISL_2684232, EPI_ISL_2684233 | Institute for Medical Research, Infectious Disease Research Centre, National Institutes of Health, Ministry of Health Malaysia | Institute for Medical Research, Infectious Disease Research Centre, National Institutes of Health, Ministry of Health Malaysia | Suppiah J, Kamel K, Mohd Zawawi Z, Azizan MA, Ramly N, Robert F, Thayan R |
| EPI_ISL_2684234, EPI_ISL_2684235 | Melaka Hospital | Institute for Medical Research, Infectious Disease Research Centre, National Institutes of Health, Ministry of Health Malaysia | Suppiah J, Kamel K, Mohd Zawawi Z, Azizan MA, Ramly N, Robert F, Thayan R |
| EPI_ISL_2684236 | Sultanah Bahiyah Hospital, Alor Setar | Institute for Medical Research, Infectious Disease Research Centre, National Institutes of Health, Ministry of Health Malaysia | Suppiah J, Kamel K, Mohd Zawawi Z, Azizan MA, Ramly N, Robert F, Thayan R |
| EPI_ISL_2684545 | Institute for Medical Research, Infectious Disease Research Centre, National Institutes of Health, Ministry of Health Malaysia | Institute for Medical Research, Infectious Disease Research Centre, National Institutes of Health, Ministry of Health Malaysia | Suppiah J, Kamel K, Mohd Zawawi Z, Azizan MA, Ramly N, Robert F, Thayan R |
| EPI_ISL_2684686 | Statens Serum Institut Bioinformatics and Microbial Genomics | Statens Serum Institut Bioinformatics and Microbial Genomics | Danish Covid-19 Genome Consortium |
| EPI_ISL_2685359, EPI_ISL_2685766, EPI_ISL_2685835, EPI_ISL_2685836, EPI_ISL_2685841, EPI_ISL_2685843, EPI_ISL_2685844, EPI_ISL_2685845 | Institute for Medical Research, Infectious Disease Research Centre, National Institutes of Health, Ministry of Health Malaysia | Institute for Medical Research, Infectious Disease Research Centre, National Institutes of Health, Ministry of Health Malaysia | Suppiah J, Kamel K, Mohd Zawawi Z, Azizan MA, Ramly N, Robert F, Thayan R |
| EPI_ISL_2686278, EPI_ISL_2686279 | National Institute of Public Health | National Institute of Public Health | Helena Jirincova, Jaromira Vecerova, Timotej Suri, Dusan Trnka, Alexander Nagy |
| EPI_ISL_2687328 | Aegis Sciences Corporation | Centers for Disease Control and Prevention Division of Viral Diseases, Pathogen Discovery | Dakota Howard, Dhwani Batra, Peter W. Cook, Kara Moser, Adrian Paskey, Jason Caravas, Benjamin Rambo-Martin, Shatavia Morrison, Christopher Gulvick, Scott Sammons, Yvette Unoarumhi, Darlene Wagner, Matthew Schmerer, Cyndi Clark, Patrick Campbell, Rob Case, Vikramsinha Ghorpade, Holly Houdeshell, Ola Kvalvaag, Dillon Nall, Ethan Sanders, Alec Vest, Shaun Westlund, Matthew Hardison, Clinton R. Paden, Duncan MacCannell |
| EPI_ISL_2687946 | National Platform bis UMONS/Jolimont | National Platform bis UMONS/Jolimont | François Dufrasne, Guillaume Bayon-Vicente, Florian Juszczak, Gautier Detry, Ruddy Wattiez |
| EPI_ISL_2688532, EPI_ISL_2688534, EPI_ISL_2688537 | National Health Laboratory Service, South Africa | KRISP, KZn Research Innovation and Sequencing Platform | Giandhari Jennifer, Pillay Sureshnee, Yajna Ramphal, Naidoo Yeshnee, Tshabuila Derek,Tegally Houriiyah, San James, Wilkinson Eduan, de Oliveira Tulio |
| EPI_ISL_2688547, EPI_ISL_2688548 | KRISP, KZn Research Innovation and Sequencing Platform | KRISP, KZn Research Innovation and Sequencing Platform | Giandhari Jennifer, Pillay Sureshnee, Yajna Ramphal, Naidoo Yeshnee, Tshabuila Derek,Tegally Houriiyah, San James, Wilkinson Eduan, de Oliveira Tulio |
| EPI_ISL_2688549, EPI_ISL_2688557, EPI_ISL_2688560, | National Health Laboratory Service, South Africa | KRISP, KZn Research Innovation and Sequencing Platform | Giandhari Jennifer, Pillay Sureshnee, Yajna Ramphal, Naidoo Yeshnee, Tshabuila Derek,Tegally Houriiyah, San James, Wilkinson Eduan, de Oliveira Tulio |

| | | | |
|---|---|---|---|
| EPI_ISL_2688561, EPI_ISL_2688562, EPI_ISL_2688568, EPI_ISL_2688570, EPI_ISL_2688571, EPI_ISL_2688572, EPI_ISL_2688574 | | | |
| EPI_ISL_2689595, EPI_ISL_2689597 | GA Department of Public Health Laboratory | Genomics and Discovery, Respiratory Viruses Branch, Division of Viral Diseases, Centers for Disease Control and Prevention | Jing Zhang, Anna Kelleher, Ying Tao, Yan Li, Brian Lynch, Krista Queen, Anna Uehara, Peter Cook, Han Jia Justin Ng, Rachel Marine, Clinton R. Paden, Dhwani Batra, Haibin Wang, Tara Coalter, Jasmine Padilla, Morgan Davis, Mili Sheth, Sarah Nobles, Mark Burroughs, Justin Lee, Adam Retchless, Suxiang Tong |
| EPI_ISL_2689731 | Servicio de Microbiologia Hospital Ramon y Cajal | Servicio de Microbiologia Hospital Ramon y Cajal | Ponce M, Galan JC, Martinez L. Abreu M, y Gonzalez-Alba JM |
| EPI_ISL_2690056 | KS Health and Environmental Laboratories | Centers for Disease Control and Prevention Division of Viral Diseases, Pathogen Discovery | Mili Sheth, Sarah Nobles, Jasmine Padilla, Mark Burroughs, Shoshona Le, Katie Dillon, Peter Cook, Clinton R. Paden, Dhwani Batra, Krista Queen, Kristen Knipe, Dakota Howard, Yvette Unoarumhi, Darlene Wagner, Matthew Schmerer, Ben L. Rambo-Martin, Kristine Lacek, Sam Shepard, Alison Laufer Halpin, Dave Wentworth, Vivien Dugan, Suxiang Tong, Justin Lee |
| EPI_ISL_2691928 | Rush University Medical Center | RIPHL at Rush University Medical Center | Stefan Green, Kevin Kunstman, Max Kolton, Marieta Hyde, Laura Furtado, Felix Araujo Perez, Mary Hayden, Joyce Houlihan, Diane Springer |
| EPI_ISL_2693028 | Oregon State Public Health Laboratory | Oregon State Public Health Laboratory | Rafia Razzaque, Eugene Yeboah, Vanda Makris, Laura Tsaknaridis, John Fontana and Shane Sevey |
| EPI_ISL_2693075 | NHLS_VIRO | KRISP, KZN Research Innovation and Sequencing Platform | Giandhari Jennifer, Pillay Sureshnee, Yajna Ramphal, Naidoo Yeshnee, Tshabuila Derek,Tegally Houriiyah, San James, Wilkinson Eduan, de Oliveira Tulio |
| EPI_ISL_2693321 | Swellendam PHC Clinic wc SHC | NHLS/UCT | Arash Iranzadeh, Deelan Doolabh, Lynn Tyers, Bruna Galvao, Innocent Mudau, Marvin Hsiao, Gert Marais, Diana Hardie, Stephen Korsman, Carolyn Williamson |
| EPI_ISL_2693324, EPI_ISL_2693325, EPI_ISL_2693327 | Groote Schuur Hospital wc GSH | NHLS/UCT | Arash Iranzadeh, Deelan Doolabh, Lynn Tyers, Bruna Galvao, Innocent Mudau, Marvin Hsiao, Gert Marais, Diana Hardie, Stephen Korsman, Carolyn Williamson |
| EPI_ISL_2693330, EPI_ISL_2693332 | NHLS/UCT | NHLS/UCT | Arash Iranzadeh, Deelan Doolabh, Lynn Tyers, Bruna Galvao, Innocent Mudau, Marvin Hsiao, Gert Marais, Diana Hardie, Stephen Korsman, Carolyn Williamson |
| EPI_ISL_2693736 | UW Virology Lab | UW Virology Lab | Pavitra Roychoudhury, Hong Xie, Lasata Shrestha, Shah Mohamed Bakhash, Tien V. Nguyen, Noah R. Baker, Sean Ellis, Meei-Li Huang, Keith R Jerome, Alexander Greninger |
| EPI_ISL_2693857 | NHLS_VIRO Eastern Cape | KRISP, KZN Research Innovation and Sequencing Platform | Giandhari Jennifer, Pillay Sureshnee, Yajna Ramphal, Naidoo Yeshnee, Tshabuila Derek,Tegally Houriiyah, San James, Wilkinson Eduan, de Oliveira Tulio |
| EPI_ISL_2694612 | TAMBO MEMORIAL | National Institute for Communicable Diseases of the National Health Laboratory Service | Amoako DG, Everatt J, Scheepers C, Mohale T, Ntuli N, Mahlangu B, Mnguni A, Ismail A, Bhiman JN |
| EPI_ISL_2694614, EPI_ISL_2694615 | KOPANONG | National Institute for Communicable Diseases of the National Health Laboratory Service | Amoako DG, Everatt J, Scheepers C, Mohale T, Ntuli N, Mahlangu B, Mnguni A, Ismail A, Bhiman JN |
| EPI_ISL_2694618, EPI_ISL_2694620, EPI_ISL_2694625 | TAMBO MEMORIAL | National Institute for Communicable Diseases of the National Health Laboratory Service | Amoako DG, Everatt J, Scheepers C, Mohale T, Ntuli N, Mahlangu B, Mnguni A, Ismail A, Bhiman JN |
| EPI_ISL_2694629, EPI_ISL_2694630, EPI_ISL_2694631, EPI_ISL_2694633, EPI_ISL_2694635, EPI_ISL_2694647, EPI_ISL_2694656, EPI_ISL_2694660, EPI_ISL_2694673, EPI_ISL_2694674, EPI_ISL_2694675, EPI_ISL_2694676 | | | |
| see above | CHRIS HANI BARAGWANATH | National Institute for Communicable Diseases of the National Health Laboratory Service | Amoako DG, Everatt J, Scheepers C, Mohale T, Ntuli N, Mahlangu B, Mnguni A, Ismail A, Bhiman JN |
| EPI_ISL_2694682, EPI_ISL_2694695, EPI_ISL_2694703, EPI_ISL_2694707, EPI_ISL_2694712, EPI_ISL_2694717, EPI_ISL_2694720, EPI_ISL_2694721, EPI_ISL_2694722, EPI_ISL_2695504 | TAMBO MEMORIAL | National Institute for Communicable Diseases of the National Health Laboratory Service | Amoako DG, Everatt J, Scheepers C, Mohale T, Ntuli N, Mahlangu B, Mnguni A, Ismail A, Bhiman JN |
| EPI_ISL_2695506 | Bertha gxowa hospital | National Institute for Communicable Diseases of the National Health Laboratory Service | Amoako DG, Everatt J, Scheepers C, Mohale T, Ntuli N, Mahlangu B, Mnguni A, Ismail A, Bhiman JN |
| EPI_ISL_2695509, EPI_ISL_2695511, EPI_ISL_2695528 | TAMBO MEMORIAL | National Institute for Communicable Diseases of the National Health Laboratory Service | Amoako DG, Everatt J, Scheepers C, Mohale T, Ntuli N, Mahlangu B, Mnguni A, Ismail A, Bhiman JN |
| EPI_ISL_2695541, EPI_ISL_2695542, EPI_ISL_2695544, EPI_ISL_2695545, EPI_ISL_2695546, EPI_ISL_2695547, EPI_ISL_2695552, EPI_ISL_2695553, EPI_ISL_2695554, EPI_ISL_2695558, EPI_ISL_2695559, EPI_ISL_2695560, EPI_ISL_2695563, EPI_ISL_2695567, EPI_ISL_2695568, EPI_ISL_2695571, EPI_ISL_2695575, EPI_ISL_2695577, EPI_ISL_2695582 | | | |
| see above | POLOKWANE MANKWENG HOSPITAL | National Institute for Communicable Diseases | Amoako DG, Everatt J, Scheepers C, Mohale T, Ntuli N, Mahlangu B, Mnguni A, Ismail A, Bhiman JN |

| | | | |
|---|---|---|---|
| | | of the National Health Laboratory Service | |
| EPI_ISL_2695584, EPI_ISL_2695585 | PATHCARE LABORATORY | National Institute for Communicable Diseases of the National Health Laboratory Service | Amoako DG, Everatt J, Scheepers C, Mohale T, Ntuli N, Mahlangu B, Mnguni A, Ismail A, Bhiman JN |
| EPI_ISL_2695590, EPI_ISL_2695606 | HELEN JOSEPH | National Institute for Communicable Diseases of the National Health Laboratory Service | Amoako DG, Everatt J, Scheepers C, Mohale T, Ntuli N, Mahlangu B, Mnguni A, Ismail A, Bhiman JN |
| EPI_ISL_2695608, EPI_ISL_2695609 | ROB FERREIRA LABORATORY | National Institute for Communicable Diseases of the National Health Laboratory Service | Amoako DG, Everatt J, Scheepers C, Mohale T, Ntuli N, Mahlangu B, Mnguni A, Ismail A, Bhiman JN |
| EPI_ISL_2695732 | HELEN JOSEPH | National Institute for Communicable Diseases of the National Health Laboratory Service | Amoako DG, Everatt J, Scheepers C, Mohale T, Ntuli N, Mahlangu B, Mnguni A, Ismail A, Bhiman JN |
| EPI_ISL_2695760, EPI_ISL_2695761 | KOPANONG LABORATORY | National Institute for Communicable Diseases of the National Health Laboratory Service | Amoako DG, Everatt J, Scheepers C, Mohale T, Ntuli N, Mahlangu B, Mnguni A, Ismail A, Bhiman JN |
| EPI_ISL_2695811, EPI_ISL_2695812, EPI_ISL_2695813, EPI_ISL_2695814, EPI_ISL_2695816, EPI_ISL_2695824, EPI_ISL_2695826, EPI_ISL_2695827, EPI_ISL_2695830, EPI_ISL_2695831, EPI_ISL_2695832, EPI_ISL_2695833, EPI_ISL_2695838, EPI_ISL_2695839, EPI_ISL_2695840, EPI_ISL_2695848 | | | |
| see above | PathCare, Cape Town | Division of Medical Virology, National Health Laboratory Service (NHLS), Tygerberg Hospital / Stellenbosch University | Susan Engelbrecht, Tongai Maponga, Bronwyn Kleinhans, Tania Stander, Jean Maritz, Petra Raimond, San Emmanual James, Tulio de Oliveira, Gert van Zyl, Wolfgang Preiser |
| EPI_ISL_2695850, EPI_ISL_2695858, EPI_ISL_2695863, EPI_ISL_2695867 | Division of Medical Virology, National Health Laboratory Service (NHLS), Tygerberg Hospital / Stellenbosch University | Division of Medical Virology, National Health Laboratory Service (NHLS), Tygerberg Hospital / Stellenbosch University | Susan Engelbrecht, Tongai Maponga, Bronwyn Kleinhans, Tania Stander, Gert van Zyl, San Emmanual James, Tulio de Oliveira, Wolfgang Preiser |
| EPI_ISL_2709261 | Labo Analyses Med | National Reference Center for Viruses of Respiratory Infections, Institut Pasteur, Paris | Marion Barbet, Sylvie Behillil, Méline Bizard, Angela Brisebarre, Camille Capel, Vincent Enouf, Louise Lefrançois, Frédéric Lemoine, Christophe Malabat, Corinne Maufrais, Etienne Simon-Lorière, Maud Vanpeene, Sylvie Van der Werf ,Patricia Tamby |
| EPI_ISL_2709270 | Labo Analyses Med | National Reference Center for Viruses of Respiratory Infections, Institut Pasteur, Paris | Marion Barbet, Sylvie Behillil, Méline Bizard, Angela Brisebarre, Camille Capel, Vincent Enouf, Louise Lefrançois, Frédéric Lemoine, Christophe Malabat, Corinne Maufrais, Etienne Simon-Lorière, Maud Vanpeene, Sylvie Van der Werf ,Anne-Laure Garand |
| EPI_ISL_2709294 | Labo Analyses med | National Reference Center for Viruses of Respiratory Infections, Institut Pasteur, Paris | Marion Barbet, Sylvie Behillil, Méline Bizard, Angela Brisebarre, Camille Capel, Vincent Enouf, Louise Lefrançois, Frédéric Lemoine, Christophe Malabat, Corinne Maufrais, Damien Mornico, Etienne Simon-Lorière, Maud Vanpeene, Sylvie Van der Werf ,OphéLie Said-Delattre |
| EPI_ISL_2709304 | Labo Analyses med | National Reference Center for Viruses of Respiratory Infections, Institut Pasteur, Paris | Marion Barbet, Sylvie Behillil, Méline Bizard, Angela Brisebarre, Camille Capel, Vincent Enouf, Louise Lefrançois, Frédéric Lemoine, Christophe Malabat, Corinne Maufrais, Etienne Simon-Lorière, Maud Vanpeene, Sylvie Van der Werf ,OphéLie Said-Delattre |
| EPI_ISL_2709851, EPI_ISL_2709862 | POLOKWANE MANKWENG HOSPITAL | National Institute for Communicable Diseases of the National Health Laboratory Service | Amoako DG, Everatt J, Scheepers C, Mohale T, Ntuli N, Mahlangu B, Mnguni A, Ismail A, Bhiman JN |
| EPI_ISL_2709863, EPI_ISL_2709872, EPI_ISL_2709875, EPI_ISL_2709878 | HELEN JOSEPH | National Institute for Communicable Diseases of the National Health Laboratory Service | Amoako DG, Everatt J, Scheepers C, Mohale T, Ntuli N, Mahlangu B, Mnguni A, Ismail A, Bhiman JN |
| EPI_ISL_2709880, EPI_ISL_2709883, EPI_ISL_2709884 | CHRIS HANI BARAGWANATH | National Institute for Communicable Diseases of the National Health Laboratory Service | Amoako DG, Everatt J, Scheepers C, Mohale T, Ntuli N, Mahlangu B, Mnguni A, Ismail A, Bhiman JN |
| EPI_ISL_2709885 | HELEN JOSEPH | National Institute for Communicable Diseases of the National Health Laboratory Service | Amoako DG, Everatt J, Scheepers C, Mohale T, Ntuli N, Mahlangu B, Mnguni A, Ismail A, Bhiman JN |
| EPI_ISL_2709897, EPI_ISL_2709900, EPI_ISL_2709901, EPI_ISL_2709906, EPI_ISL_2709908, EPI_ISL_2709909, EPI_ISL_2709911, EPI_ISL_2709917, EPI_ISL_2709919, EPI_ISL_2709924, EPI_ISL_2709926, EPI_ISL_2709930, EPI_ISL_2709931, EPI_ISL_2709933, EPI_ISL_2709940 | | | |
| see above | CHRIS HANI BARAGWANATH | National Institute for Communicable Diseases of the National Health Laboratory Service | Amoako DG, Everatt J, Scheepers C, Mohale T, Ntuli N, Mahlangu B, Mnguni A, Ismail A, Bhiman JN |
| EPI_ISL_2709953, EPI_ISL_2709957, EPI_ISL_2709960, EPI_ISL_2709979, EPI_ISL_2709982, EPI_ISL_2709987, EPI_ISL_2709988 | TAMBO MEMORIAL LABORATORY | National Institute for Communicable Diseases of the National Health Laboratory Service | Amoako DG, Everatt J, Scheepers C, Mohale T, Ntuli N, Mahlangu B, Mnguni A, Ismail A, Bhiman JN |
| EPI_ISL_2710263, | NHLS Charlotte Maxeke | KRISP, KZN Research | Florette Treurnicht, Bulelani Manene, Kathleen Subramoney, Giandhari Jennifer, |

| | | | |
|---|---|---|---|
| EPI_ISL_2710289, EPI_ISL_2710291 | Johannesburg Academic Hospital and the University of the Witwatersrand | Innovation and Sequencing Platform | Pillay Sureshnee, Yajna Ramphal, Naidoo Yeshnee, Tshabuila Derek,Tegally Houriiyah, San James, Wilkinson Eduan, de Oliveira Tulio |
| EPI_ISL_2710309, EPI_ISL_2710310, EPI_ISL_2710311, EPI_ISL_2710317, EPI_ISL_2710321 | BARC / Lancet | KRISP, KZN Research Innovation and Sequencing Platform | Sisonke, Giandhari Jennifer, Pillay Sureshnee, Yajna Ramphal, Naidoo Yeshnee, Tshabuila Derek,Tegally Houriiyah, San James, Wilkinson Eduan, de Oliveira Tulio |
| EPI_ISL_2710799 | Orebro University Hospital, Dept Laboratory Medicine, Clinical Microbiology | Orebro University Hospital | Sundqvist M et al |
| EPI_ISL_2713000 | TXDSHS | TXDSHS | Rashmi Tuladhar, Bonnie Oh, Jenny Zhang, Maliha Rahman, Mayela Pedrueza, Anita Pokharel, Karen Bobier, Lorraine Rodriguez, Myong Koag, Chun Wang, Rachel Lee, Grace Kubin |
| EPI_ISL_2715018 | Atlas Genomics | Seattle Flu Study | Deborah A. Nickerson, Chris D. Frazar, Jover Lee, Benjamin Pelle, Erica Ryke, Matthew Richardson, Amanda Adler, Elisabeth Brandstetter, Peter D. Han, Kairsten Fay, Misja Ilcisin, Kirsten Lacombe, Thomas R. Sibley, Melissa Truong, Caitlin R. Wolf, Romesh Gautom, Geoff Melly, Brian Hiatt, Philip Dykema, Scott Lindquist, Michael Boeckh, Janet A. Englund, Michael Famulare, Barry R. Lutz, Mark J. Rieder, Lea M. Starita, Matthew Thompson, Helen Y. Chu, Jay Shendure, Trevor Bedford |
| EPI_ISL_2715174 | Northwest Laboratory | Seattle Flu Study | Deborah A. Nickerson, Chris D. Frazar, Jover Lee, Benjamin Pelle, Erica Ryke, Matthew Richardson, Amanda Adler, Elisabeth Brandstetter, Peter D. Han, Kairsten Fay, Misja Ilcisin, Kirsten Lacombe, Thomas R. Sibley, Melissa Truong, Caitlin R. Wolf, Romesh Gautom, Geoff Melly, Brian Hiatt, Philip Dykema, Scott Lindquist, Michael Boeckh, Janet A. Englund, Michael Famulare, Barry R. Lutz, Mark J. Rieder, Lea M. Starita, Matthew Thompson, Helen Y. Chu, Jay Shendure, Trevor Bedford |
| EPI_ISL_2726322, EPI_ISL_2726331, EPI_ISL_2726335, EPI_ISL_2726336, EPI_ISL_2726337, EPI_ISL_2726338, EPI_ISL_2726339, EPI_ISL_2726340, EPI_ISL_2726341, EPI_ISL_2726342, EPI_ISL_2726512, EPI_ISL_2726514 | | | |
| see above | KU Leuven, Rega Institute, Clinical and Epidemiological Virology | KU Leuven, Rega Institute, Clinical and Epidemiological Virology | Tony Wawina-Bokalanga, Bert Vanmechelen, Joan Marti-Carerras, Piet Maes |
| EPI_ISL_2727331, EPI_ISL_2727333, EPI_ISL_2727336, EPI_ISL_2727337, EPI_ISL_2727339, EPI_ISL_2727340, EPI_ISL_2727341, EPI_ISL_2727342, EPI_ISL_2727344, EPI_ISL_2727346, EPI_ISL_2727348 | | | |
| see above | ZARV/NHLS, Department Medical Virology, University of Pretoria | KRISP, KZn Research Innovation and Sequencing Platform | Amy Strydom, Adriano Mendes, Micheala Davids, Sim Mayaphi and Marietjie Venter, Giandhari J, Pillay S, Naidoo Y, Ramphal U, Lessells R, Emmanuel SJ, Tegally H, Wilkinson E, de Oliveira T |
| EPI_ISL_2736453 | Seattle Flu Study | Seattle Flu Study | Deborah A. Nickerson, Chris D. Frazar, Jover Lee, Benjamin Pelle, Erica Ryke, Matthew Richardson, Amanda Adler, Elisabeth Brandstetter, Peter D. Han, Kairsten Fay, Misja Ilcisin, Kirsten Lacombe, Thomas R. Sibley, Melissa Truong, Caitlin R. Wolf, Michael Boeckh, Janet A. Englund, Michael Famulare, Barry R. Lutz, Mark J. Rieder, Lea M. Starita, Matthew Thompson, Jay Shendure, Trevor Bedford, Helen Y. Chu |
| EPI_ISL_2757735, EPI_ISL_2757740, EPI_ISL_2757745, EPI_ISL_2757746, EPI_ISL_2757747, EPI_ISL_2757748, EPI_ISL_2757750, EPI_ISL_2757751, EPI_ISL_2757753, EPI_ISL_2757754 | Department of Virology | Department of Virology | Massab Umair, Aamer Ikram, Muhammad Salman, Nazish Badar, Zaira Rehman, Muhammad Ammar, Syed Adnan Haider |
| EPI_ISL_2758864, EPI_ISL_2758869 | The Ohio State University Applied Microbiology Services Laboratory | The Ohio State University Applied Microbiology Services Laboratory | Seth A. Faith PhD |
| EPI_ISL_2760111 | Bayerisches Landesamt für Gesundheit und Lebensmittelsicherheit (LGL) | Robert Koch Institute | unknown |
| EPI_ISL_2761339 | Sonic - Bioscientia - MVZ Labor Saar GmbH | Robert Koch Institute | unknown |
| EPI_ISL_2761420 | LabKom - MVZ Labor Bochum MLB GmbH | Robert Koch Institute | unknown |
| EPI_ISL_2761798 | CENTOGENE Frankfurt Laboratory: Niederlassung Industriepark Höchst | Robert Koch Institute | unknown |
| EPI_ISL_2762020, EPI_ISL_2762074 | Sonic - Bioscientia - MVZ Labor Saar GmbH | Robert Koch Institute | unknown |
| EPI_ISL_2762313 | Diagnosticum - Labor Neukirchen | Robert Koch Institute | unknown |
| EPI_ISL_2768014 | National Institute of Laboratory Medicine and Referral Center | Genomic Research Lab, BCSIR | Md. Murshed Hasan Sarkar, Mohammad Samir Uzzaman, Eshrar Osman, Md. Ahasan Habib, Shahina Akter, Tanjina Akhter Banu, Barna Goswami, Iffat Jahan, Mohammad Mohi Uddin, Md. Kamrul Islam, Tasnim Nafisa, Md. Maruf Ahmed Molla, Mahmuda Yeasmin, Asish Kumar Ghosh, Arifa Akram, Md. Salim Khan |
| EPI_ISL_2769141, EPI_ISL_2769142 | UniversitätsSpital Zürich | Institute of Medical Virology | Verena Kufner, Gabriela Ziltener, Maryam Zaheri, Stefan Schmutz, Annette Audigé, Maria Grünberg, Kevin Steiner, Jon Huder, Cyril Shah, Riccarda Capaul, Guido Bloemberg, Jürg Böni, Michael Huber, Alexandra Trkola |
| EPI_ISL_2770441, EPI_ISL_2770481 | NHLS Charlotte Maxeke Johannesburg Academic | KRISP, KZN Research Innovation and | Florette Treurnicht, Bulelani Manene, Kathleen Subramoney, Giandhari Jennifer, Pillay Sureshnee, Yajna Ramphal, Naidoo Yeshnee, Tshabuila Derek,Tegally |

| | | | |
|---|---|---|---|
| | Hospital and the University of the Witwatersrand | Sequencing Platform | Houriiyah, San James, Wilkinson Eduan, de Oliveira Tulio |
| EPI_ISL_2772528 | National Center of Infectious and Parasitic Diseases | National Center of Infectious and Parasitic Diseases | Alexiev et al |

### supp. Table S4: Delta

We gratefully acknowledge the following Authors from the Originating laboratories responsible for obtaining the specimens, as well as the Submitting laboratories where the genome data were generated and shared via GISAID, on which this research is based.

All Submitters of data may be contacted directly via www.gisaid.org

Authors are sorted alphabetically.

| Accession ID | Originating Laboratory | Submitting Laboratory | Authors |
|---|---|---|---|
| EPI_ISL_2597929, EPI_ISL_2597932 | Nebraska Public Health Laboratory | NPHL COVID-19 Response Team | NPHL COVID-19 Response Team |
| EPI_ISL_2617990, EPI_ISL_2617991, EPI_ISL_2618009, EPI_ISL_2618010, EPI_ISL_2618013, EPI_ISL_2618016, EPI_ISL_2618018, EPI_ISL_2618019 | UW Virology Lab | UW Virology Lab | Pavitra Roychoudhury, Hong Xie, Lasata Shrestha, Tien V. Nguyen, Shah Mohamed Bakhash, Michelle Lin, Noah R. Baker, Ricardo Perez, Sean Ellis, Nathan Breit, Robert J. Livingston, Meei-Li Huang, Keith R Jerome, Patrick Mathias, Alexander Greninger |
| EPI_ISL_2620723 | Genomik Solidaritas Indonesia Laboratorium | Genomik Solidaritas Indonesia Laboratorium | Dhahlia Agustina Cahyono, Alfin Mohammad Abdillah, Normastuti Adhini Tantyo, Carissa Sintca Wijaya, Vania Gavrila Wikasa, Ahmad Zoebad Foeady, Annisa Muthiah Sukirman, Gracia Felias Enos Korompis, Anuraj Shankar, Meutia Ayuputeri Kumaheri, Ariel Pradipta |
| EPI_ISL_2623094 | University College London, Great Ormond Street Hospital for Children NHS Foundation Trust, Imperial College Healthcare NHS Trust | COVID-19 Genomics UK (COG-UK) Consortium | Sergi Castellano, Rachel Williams, Mark Kristiansen, Paola Resende Silva, Sunando Roy, Tony Brooks, Helena Tutill, Paola Niola, Patricia Dyal, Charlotte Williams, Leysa Forrest, Yasmin Panchbhaya, Jacqueline Findlay, Samuel Weeks, Julianne Brown, Kathryn Harris, Paul Randell, James Price, Alison Holmes, Judith Breuer |
| EPI_ISL_2625108, EPI_ISL_2625119, EPI_ISL_2625142, EPI_ISL_2625143, EPI_ISL_2625144, EPI_ISL_2625145, EPI_ISL_2625146, EPI_ISL_2625149, EPI_ISL_2625151, EPI_ISL_2625153, EPI_ISL_2625157, EPI_ISL_2625159, EPI_ISL_2625160, EPI_ISL_2625165, EPI_ISL_2625166, EPI_ISL_2625168, EPI_ISL_2625169, EPI_ISL_2625171, EPI_ISL_2625172, EPI_ISL_2625173, EPI_ISL_2625177, EPI_ISL_2625178, EPI_ISL_2625180, EPI_ISL_2625181, EPI_ISL_2625182, EPI_ISL_2625184, EPI_ISL_2625185, EPI_ISL_2625186, EPI_ISL_2625187, EPI_ISL_2625188, EPI_ISL_2625189, EPI_ISL_2625190, EPI_ISL_2625191, EPI_ISL_2625192, EPI_ISL_2625195, EPI_ISL_2625196, EPI_ISL_2625197, EPI_ISL_2625199 | | | |
| see above | Centre for Enzyme Innovation, University of Portsmouth / Translational Research Laboratory, Portsmouth Hospitals NHS Trust / Hampshire Hospitals NHS Foundation Trust / Bournemouth University / University Hospitals Dorset NHS Foundation Trust / Brighton and Sussex University Hospitals NHS Trust / Dartford and Gravesham NHS Trust / Isle of Wight NHS Trust / Maidstone and Tunbridge Wells NHS Trust / University Hospital Southampton NHS Foundation Trust | COVID-19 Genomics UK (COG-UK) Consortium | Samuel Robson, Angela Beckett, Salman Goudarzi, Christopher Fearn, Kate Cook, Katie Loveson, Sharon Glaysher, Scott Elliott, Kelly Bicknell, Sarah Wyllie, Allyson Lloyd, Robert Impey, Anoop Chauhan, Stephen Kidd, Nathan Moore, Nick Cortes, Claire Thomas, Anna Mantzaouratou, Sarah Buchan, Magdalena Barrow, Andrew Butt, Liz Sheridan, Jonnie Seymour, Dorian Crudgington, Ben Macklin, Mohammed Hassan-Ibrahim, Cassandra Malone, Benjamin Cogger, Kevin Tucker, Samirakhon Raupova, Rachael Jeremiah, Anibolina Castigador, Emily Macnaughton, Karen Withell, Kordo Saeed, Jacqui Prieto, Adhyana Mahanama, Buddhini Samaraweera, Siona Silvieri, Emanuela Pelosi, Eleri Wilson-Davies, Sarah Jeremiah, Helen Wheeler, Matthew Harvey, Thea Sass, Helen Umpleby, Stephen Aplin |
| EPI_ISL_2627547, EPI_ISL_2627555, EPI_ISL_2627562, EPI_ISL_2627566, EPI_ISL_2627567, EPI_ISL_2627571, EPI_ISL_2627573, EPI_ISL_2627575, EPI_ISL_2627578 | National Public Health Laboratory, National Centre for Infectious Diseases | National Public Health Laboratory, National Centre for Infectious Diseases | Tze Minn Mak, Katherine Ching, Zhenyang Zhou, Royce Ang, Lin Cui, Raymond Tzer Pin Lin |
| EPI_ISL_2628025, EPI_ISL_2628026 | Montana Public Health Laboratory | Montana Public Health Laboratory | Joy Ritter, Michelle Mozer, Carrie Biskupiak, Deborah Gibson, Michael Dills |
| EPI_ISL_2628086, EPI_ISL_2628100, EPI_ISL_2628103 | Platform BIS UZA/UAntwerpen | Labo Klinische Biologie, UZA | Marie Le Mercier, Jasmine Coppens, Basil Britto Xavier, Christine Lammens, Veerle Matheeussen, Herman Goossens |
| EPI_ISL_2635462 | Synlab MVZ Augsburg | Robert Koch Institute | unknown |
| EPI_ISL_2635591 | Medizinische Laboratorien Düsseldorf | Robert Koch Institute | unknown |
| EPI_ISL_2635602, EPI_ISL_2635611, EPI_ISL_2635629, EPI_ISL_2635648, EPI_ISL_2635657, EPI_ISL_2635660 | nordlab - Partnerschaftspraxis fur Laboratoriumsmedizin | Robert Koch Institute | unknown |
| EPI_ISL_2635903, EPI_ISL_2635905 | Limbach - MVZ Labor Ludwigsburg | Robert Koch Institute | unknown |
| EPI_ISL_2638165 | Lighthouse Lab in Glasgow | Wellcome Sanger Institute for the COVID-19 Genomics UK (COG-UK) Consortium | Harper VanSteenhouse, Yumi Kasai, David Gray, Carol Clugston, Anna Dominiczak and Alex Alderton, Roberto Amato, Jeffrey Barrett, Sonia Goncalves, Ewan Harrison, David K. Jackson, Ian Johnston, Dominic Kwiatkowski, Cordelia Langford, John Sillitoe on behalf of the Wellcome Sanger Institute COVID-19 Surveillance Team |
| EPI_ISL_2638169, EPI_ISL_2638171, EPI_ISL_2638203, EPI_ISL_2638205 | Lighthouse Lab in Alderley Park | Wellcome Sanger Institute for the COVID-19 Genomics UK (COG-UK) Consortium | Jacquelyn Wynn, Mairead Hyland, The Lighthouse Lab in Alderley Park and Alex Alderton, Roberto Amato, Jeffrey Barrett, Sonia Goncalves, Ewan Harrison, David K. Jackson, Ian Johnston, Dominic Kwiatkowski, Cordelia Langford, John Sillitoe on behalf of the Wellcome Sanger Institute COVID-19 Surveillance Team |
| EPI_ISL_2638223 | Lighthouse Lab in Glasgow | Wellcome Sanger Institute for the COVID-19 Genomics UK (COG-UK) Consortium | Harper VanSteenhouse, Yumi Kasai, David Gray, Carol Clugston, Anna Dominiczak and Alex Alderton, Roberto Amato, Jeffrey Barrett, Sonia Goncalves, Ewan Harrison, David K. Jackson, Ian Johnston, Dominic Kwiatkowski, Cordelia Langford, John Sillitoe on behalf of the Wellcome Sanger Institute COVID-19 Surveillance Team |
| EPI_ISL_2638224, EPI_ISL_2638228, EPI_ISL_2638231, EPI_ISL_2638248 | Lighthouse Lab in Alderley Park | Wellcome Sanger Institute for the COVID-19 Genomics UK (COG-UK) Consortium | Jacquelyn Wynn, Mairead Hyland, The Lighthouse Lab in Alderley Park and Alex Alderton, Roberto Amato, Jeffrey Barrett, Sonia Goncalves, Ewan Harrison, David K. Jackson, Ian Johnston, Dominic Kwiatkowski, Cordelia Langford, John Sillitoe on behalf of the Wellcome Sanger Institute COVID-19 Surveillance Team |
| EPI_ISL_2638265 | Lighthouse Lab in Milton Keynes | Wellcome Sanger Institute for the COVID-19 Genomics UK (COG-UK) Consortium | The Lighthouse Lab in Milton Keynes and Alex Alderton, Roberto Amato, Jeffrey Barrett, Sonia Goncalves, Ewan Harrison, David K. Jackson, Ian Johnston, Dominic Kwiatkowski, Cordelia Langford, John Sillitoe on behalf of the Wellcome Sanger Institute COVID-19 Surveillance Team |
| EPI_ISL_2638269, EPI_ISL_2638275, EPI_ISL_2638278, EPI_ISL_2638283, EPI_ISL_2638290 | Lighthouse Lab in Alderley Park | Wellcome Sanger Institute for the COVID-19 Genomics UK (COG-UK) Consortium | Jacquelyn Wynn, Mairead Hyland, The Lighthouse Lab in Alderley Park and Alex Alderton, Roberto Amato, Jeffrey Barrett, Sonia Goncalves, Ewan Harrison, David K. Jackson, Ian Johnston, Dominic Kwiatkowski, Cordelia Langford, John Sillitoe on behalf of the Wellcome Sanger Institute COVID-19 Surveillance Team |
| EPI_ISL_2638377 | Lighthouse Lab in Glasgow | Wellcome Sanger Institute for the COVID-19 Genomics UK (COG-UK) Consortium | Harper VanSteenhouse, Yumi Kasai, David Gray, Carol Clugston, Anna Dominiczak and Alex Alderton, Roberto Amato, Jeffrey Barrett, Sonia Goncalves, Ewan Harrison, David K. Jackson, Ian Johnston, Dominic Kwiatkowski, Cordelia Langford, John Sillitoe on behalf of the Wellcome Sanger Institute COVID-19 Surveillance Team |
| EPI_ISL_2638384, EPI_ISL_2638388 | Lighthouse Lab in Alderley Park | Wellcome Sanger Institute for the COVID-19 Genomics UK (COG-UK) Consortium | Jacquelyn Wynn, Mairead Hyland, The Lighthouse Lab in Alderley Park and Alex Alderton, Roberto Amato, Jeffrey Barrett, Sonia Goncalves, Ewan Harrison, David K. Jackson, Ian Johnston, Dominic Kwiatkowski, Cordelia Langford, John Sillitoe on behalf of the Wellcome Sanger Institute COVID-19 Surveillance Team |
| EPI_ISL_2638404, EPI_ISL_2638406, EPI_ISL_2638409 | Lighthouse Lab in Milton Keynes | Wellcome Sanger Institute for the COVID-19 Genomics UK (COG-UK) Consortium | The Lighthouse Lab in Milton Keynes and Alex Alderton, Roberto Amato, Jeffrey Barrett, Sonia Goncalves, Ewan Harrison, David K. Jackson, Ian Johnston, Dominic Kwiatkowski, Cordelia Langford, John Sillitoe on behalf of the Wellcome Sanger Institute COVID-19 Surveillance Team |
| EPI_ISL_2638419 | Lighthouse Lab in Glasgow | Wellcome Sanger Institute for the COVID-19 Genomics UK (COG-UK) Consortium | Harper VanSteenhouse, Yumi Kasai, David Gray, Carol Clugston, Anna Dominiczak and Alex Alderton, Roberto Amato, Jeffrey Barrett, Sonia Goncalves, Ewan Harrison, David K. Jackson, Ian Johnston, Dominic Kwiatkowski, Cordelia Langford, John Sillitoe on behalf of the Wellcome Sanger Institute COVID-19 Surveillance Team |
| EPI_ISL_2638428, EPI_ISL_2638430 | Lighthouse Lab in Milton Keynes | Wellcome Sanger Institute for the COVID-19 Genomics UK | The Lighthouse Lab in Milton Keynes and Alex Alderton, Roberto Amato, Jeffrey Barrett, Sonia Goncalves, Ewan Harrison, David K. Jackson, Ian Johnston, |

| | | | |
|---|---|---|---|
| | | (COG-UK) Consortium | Dominic Kwiatkowski, Cordelia Langford, John Sillitoe on behalf of the Wellcome Sanger Institute COVID-19 Surveillance Team |
| EPI_ISL_2638435 | Lighthouse Lab in Glasgow | Wellcome Sanger Institute for the COVID-19 Genomics UK (COG-UK) Consortium | Harper VanSteenhouse, Yumi Kasai, David Gray, Carol Clugston, Anna Dominiczak and Alex Alderton, Roberto Amato, Jeffrey Barrett, Sonia Goncalves, Ewan Harrison, David K. Jackson, Ian Johnston, Dominic Kwiatkowski, Cordelia Langford, John Sillitoe on behalf of the Wellcome Sanger Institute COVID-19 Surveillance Team |
| EPI_ISL_2638437, EPI_ISL_2638441 | Lighthouse Lab in Milton Keynes | Wellcome Sanger Institute for the COVID-19 Genomics UK (COG-UK) Consortium | The Lighthouse Lab in Milton Keynes and Alex Alderton, Roberto Amato, Jeffrey Barrett, Sonia Goncalves, Ewan Harrison, David K. Jackson, Ian Johnston, Dominic Kwiatkowski, Cordelia Langford, John Sillitoe on behalf of the Wellcome Sanger Institute COVID-19 Surveillance Team |
| EPI_ISL_2638453 | Lighthouse Lab in Glasgow | Wellcome Sanger Institute for the COVID-19 Genomics UK (COG-UK) Consortium | Harper VanSteenhouse, Yumi Kasai, David Gray, Carol Clugston, Anna Dominiczak and Alex Alderton, Roberto Amato, Jeffrey Barrett, Sonia Goncalves, Ewan Harrison, David K. Jackson, Ian Johnston, Dominic Kwiatkowski, Cordelia Langford, John Sillitoe on behalf of the Wellcome Sanger Institute COVID-19 Surveillance Team |
| EPI_ISL_2638478, EPI_ISL_2638487, EPI_ISL_2638554, EPI_ISL_2638588 | Lighthouse Lab in Milton Keynes | Wellcome Sanger Institute for the COVID-19 Genomics UK (COG-UK) Consortium | The Lighthouse Lab in Milton Keynes and Alex Alderton, Roberto Amato, Jeffrey Barrett, Sonia Goncalves, Ewan Harrison, David K. Jackson, Ian Johnston, Dominic Kwiatkowski, Cordelia Langford, John Sillitoe on behalf of the Wellcome Sanger Institute COVID-19 Surveillance Team |
| EPI_ISL_2638595 | Lighthouse Lab in Glasgow | Wellcome Sanger Institute for the COVID-19 Genomics UK (COG-UK) Consortium | Harper VanSteenhouse, Yumi Kasai, David Gray, Carol Clugston, Anna Dominiczak and Alex Alderton, Roberto Amato, Jeffrey Barrett, Sonia Goncalves, Ewan Harrison, David K. Jackson, Ian Johnston, Dominic Kwiatkowski, Cordelia Langford, John Sillitoe on behalf of the Wellcome Sanger Institute COVID-19 Surveillance Team |
| EPI_ISL_2638597, EPI_ISL_2638621, EPI_ISL_2638624 | Lighthouse Lab in Milton Keynes | Wellcome Sanger Institute for the COVID-19 Genomics UK (COG-UK) Consortium | The Lighthouse Lab in Milton Keynes and Alex Alderton, Roberto Amato, Jeffrey Barrett, Sonia Goncalves, Ewan Harrison, David K. Jackson, Ian Johnston, Dominic Kwiatkowski, Cordelia Langford, John Sillitoe on behalf of the Wellcome Sanger Institute COVID-19 Surveillance Team |
| EPI_ISL_2638625 | Lighthouse Lab in Glasgow | Wellcome Sanger Institute for the COVID-19 Genomics UK (COG-UK) Consortium | Harper VanSteenhouse, Yumi Kasai, David Gray, Carol Clugston, Anna Dominiczak and Alex Alderton, Roberto Amato, Jeffrey Barrett, Sonia Goncalves, Ewan Harrison, David K. Jackson, Ian Johnston, Dominic Kwiatkowski, Cordelia Langford, John Sillitoe on behalf of the Wellcome Sanger Institute COVID-19 Surveillance Team |
| EPI_ISL_2638631, EPI_ISL_2638639, EPI_ISL_2638640, EPI_ISL_2638645 | Lighthouse Lab in Milton Keynes | Wellcome Sanger Institute for the COVID-19 Genomics UK (COG-UK) Consortium | The Lighthouse Lab in Milton Keynes and Alex Alderton, Roberto Amato, Jeffrey Barrett, Sonia Goncalves, Ewan Harrison, David K. Jackson, Ian Johnston, Dominic Kwiatkowski, Cordelia Langford, John Sillitoe on behalf of the Wellcome Sanger Institute COVID-19 Surveillance Team |
| EPI_ISL_2638651, EPI_ISL_2638665 | Lighthouse Laboratory Plymouth | Wellcome Sanger Institute for the COVID-19 Genomics UK (COG-UK) Consortium | Lighthouse Laboratory Plymouth and Alex Alderton, Roberto Amato, Jeffrey Barrett, Sonia Goncalves, Ewan Harrison, David K. Jackson, Ian Johnston, Dominic Kwiatkowski, Cordelia Langford, John Sillitoe on behalf of the Wellcome Sanger Institute COVID-19 Surveillance Team |
| EPI_ISL_2638667 | Lighthouse Lab in Milton Keynes | Wellcome Sanger Institute for the COVID-19 Genomics UK (COG-UK) Consortium | The Lighthouse Lab in Milton Keynes and Alex Alderton, Roberto Amato, Jeffrey Barrett, Sonia Goncalves, Ewan Harrison, David K. Jackson, Ian Johnston, Dominic Kwiatkowski, Cordelia Langford, John Sillitoe on behalf of the Wellcome Sanger Institute COVID-19 Surveillance Team |
| EPI_ISL_2638674, EPI_ISL_2638675, EPI_ISL_2638676 | Lighthouse Lab in Glasgow | Wellcome Sanger Institute for the COVID-19 Genomics UK (COG-UK) Consortium | Harper VanSteenhouse, Yumi Kasai, David Gray, Carol Clugston, Anna Dominiczak and Alex Alderton, Roberto Amato, Jeffrey Barrett, Sonia Goncalves, Ewan Harrison, David K. Jackson, Ian Johnston, Dominic Kwiatkowski, Cordelia Langford, John Sillitoe on behalf of the Wellcome Sanger Institute COVID-19 Surveillance Team |
| EPI_ISL_2638678 | Lighthouse Lab in Milton Keynes | Wellcome Sanger Institute for the COVID-19 Genomics UK (COG-UK) Consortium | The Lighthouse Lab in Milton Keynes and Alex Alderton, Roberto Amato, Jeffrey Barrett, Sonia Goncalves, Ewan Harrison, David K. Jackson, Ian Johnston, Dominic Kwiatkowski, Cordelia Langford, John Sillitoe on behalf of the Wellcome Sanger Institute COVID-19 Surveillance Team |
| EPI_ISL_2638681 | Lighthouse Lab in Glasgow | Wellcome Sanger Institute for the COVID-19 Genomics UK (COG-UK) Consortium | Harper VanSteenhouse, Yumi Kasai, David Gray, Carol Clugston, Anna Dominiczak and Alex Alderton, Roberto Amato, Jeffrey Barrett, Sonia Goncalves, Ewan Harrison, David K. Jackson, Ian Johnston, Dominic Kwiatkowski, Cordelia Langford, John Sillitoe on behalf of the Wellcome Sanger Institute COVID-19 Surveillance Team |
| EPI_ISL_2638682 | Lighthouse Lab in Milton Keynes | Wellcome Sanger Institute for the COVID-19 Genomics UK (COG-UK) Consortium | The Lighthouse Lab in Milton Keynes and Alex Alderton, Roberto Amato, Jeffrey Barrett, Sonia Goncalves, Ewan Harrison, David K. Jackson, Ian Johnston, Dominic Kwiatkowski, Cordelia Langford, John Sillitoe on behalf of the Wellcome Sanger Institute COVID-19 Surveillance Team |
| EPI_ISL_2638683 | Lighthouse Lab in Glasgow | Wellcome Sanger Institute for the COVID-19 Genomics UK (COG-UK) Consortium | Harper VanSteenhouse, Yumi Kasai, David Gray, Carol Clugston, Anna Dominiczak and Alex Alderton, Roberto Amato, Jeffrey Barrett, Sonia Goncalves, Ewan Harrison, David K. Jackson, Ian Johnston, Dominic Kwiatkowski, Cordelia Langford, John Sillitoe on behalf of the Wellcome Sanger Institute COVID-19 Surveillance Team |
| EPI_ISL_2638687 | Lighthouse Lab in Milton Keynes | Wellcome Sanger Institute for the COVID-19 Genomics UK (COG-UK) Consortium | The Lighthouse Lab in Milton Keynes and Alex Alderton, Roberto Amato, Jeffrey Barrett, Sonia Goncalves, Ewan Harrison, David K. Jackson, Ian Johnston, Dominic Kwiatkowski, Cordelia Langford, John Sillitoe on behalf of the Wellcome Sanger Institute COVID-19 Surveillance Team |
| EPI_ISL_2638691, EPI_ISL_2638692, EPI_ISL_2638696 | Lighthouse Lab in Glasgow | Wellcome Sanger Institute for the COVID-19 Genomics UK (COG-UK) Consortium | Harper VanSteenhouse, Yumi Kasai, David Gray, Carol Clugston, Anna Dominiczak and Alex Alderton, Roberto Amato, Jeffrey Barrett, Sonia Goncalves, Ewan Harrison, David K. Jackson, Ian Johnston, Dominic Kwiatkowski, Cordelia Langford, John Sillitoe on behalf of the Wellcome Sanger Institute COVID-19 Surveillance Team |
| EPI_ISL_2638705, EPI_ISL_2638713, EPI_ISL_2638714, EPI_ISL_2638723 | Lighthouse Lab in Milton Keynes | Wellcome Sanger Institute for the COVID-19 Genomics UK (COG-UK) Consortium | The Lighthouse Lab in Milton Keynes and Alex Alderton, Roberto Amato, Jeffrey Barrett, Sonia Goncalves, Ewan Harrison, David K. Jackson, Ian Johnston, Dominic Kwiatkowski, Cordelia Langford, John Sillitoe on behalf of the Wellcome Sanger Institute COVID-19 Surveillance Team |
| EPI_ISL_2638729, EPI_ISL_2638731 | Lighthouse Lab in Glasgow | Wellcome Sanger Institute for the COVID-19 Genomics UK (COG-UK) Consortium | Harper VanSteenhouse, Yumi Kasai, David Gray, Carol Clugston, Anna Dominiczak and Alex Alderton, Roberto Amato, Jeffrey Barrett, Sonia Goncalves, Ewan Harrison, David K. Jackson, Ian Johnston, Dominic Kwiatkowski, Cordelia Langford, John Sillitoe on behalf of the Wellcome Sanger Institute COVID-19 Surveillance Team |
| EPI_ISL_2638732, EPI_ISL_2638737, EPI_ISL_2638794, EPI_ISL_2638795, EPI_ISL_2638862 | Lighthouse Lab in Milton Keynes | Wellcome Sanger Institute for the COVID-19 Genomics UK (COG-UK) Consortium | The Lighthouse Lab in Milton Keynes and Alex Alderton, Roberto Amato, Jeffrey Barrett, Sonia Goncalves, Ewan Harrison, David K. Jackson, Ian Johnston, Dominic Kwiatkowski, Cordelia Langford, John Sillitoe on behalf of the Wellcome Sanger Institute COVID-19 Surveillance Team |
| EPI_ISL_2638865 | Lighthouse Lab in Glasgow | Wellcome Sanger Institute for the COVID-19 Genomics UK (COG-UK) Consortium | Harper VanSteenhouse, Yumi Kasai, David Gray, Carol Clugston, Anna Dominiczak and Alex Alderton, Roberto Amato, Jeffrey Barrett, Sonia Goncalves, Ewan Harrison, David K. Jackson, Ian Johnston, Dominic Kwiatkowski, Cordelia Langford, John Sillitoe on behalf of the Wellcome Sanger Institute COVID-19 Surveillance Team |
| EPI_ISL_2638870 | Lighthouse Lab in Milton Keynes | Wellcome Sanger Institute for the COVID-19 Genomics UK (COG-UK) Consortium | The Lighthouse Lab in Milton Keynes and Alex Alderton, Roberto Amato, Jeffrey Barrett, Sonia Goncalves, Ewan Harrison, David K. Jackson, Ian Johnston, Dominic Kwiatkowski, Cordelia Langford, John Sillitoe on behalf of the Wellcome Sanger Institute COVID-19 Surveillance Team |
| EPI_ISL_2638873 | Lighthouse Laboratory Plymouth | Wellcome Sanger Institute for the COVID-19 Genomics UK (COG-UK) Consortium | Lighthouse Laboratory Plymouth and Alex Alderton, Roberto Amato, Jeffrey Barrett, Sonia Goncalves, Ewan Harrison, David K. Jackson, Ian Johnston, Dominic Kwiatkowski, Cordelia Langford, John Sillitoe on behalf of the Wellcome Sanger Institute COVID-19 Surveillance Team |
| EPI_ISL_2638876, EPI_ISL_2638880, EPI_ISL_2638882, EPI_ISL_2638886 | Lighthouse Lab in Milton Keynes | Wellcome Sanger Institute for the COVID-19 Genomics UK (COG-UK) Consortium | The Lighthouse Lab in Milton Keynes and Alex Alderton, Roberto Amato, Jeffrey Barrett, Sonia Goncalves, Ewan Harrison, David K. Jackson, Ian Johnston, Dominic Kwiatkowski, Cordelia Langford, John Sillitoe on behalf of the Wellcome Sanger Institute COVID-19 Surveillance Team |
| EPI_ISL_2638887 | Lighthouse Lab in Glasgow | Wellcome Sanger Institute for the COVID-19 Genomics UK (COG-UK) Consortium | Harper VanSteenhouse, Yumi Kasai, David Gray, Carol Clugston, Anna Dominiczak and Alex Alderton, Roberto Amato, Jeffrey Barrett, Sonia Goncalves, Ewan Harrison, David K. Jackson, Ian Johnston, Dominic Kwiatkowski, Cordelia Langford, John Sillitoe on behalf of the Wellcome Sanger Institute COVID-19 Surveillance Team |
| EPI_ISL_2638888, EPI_ISL_2638892 | Lighthouse Lab in Milton Keynes | Wellcome Sanger Institute for the COVID-19 Genomics UK (COG-UK) Consortium | The Lighthouse Lab in Milton Keynes and Alex Alderton, Roberto Amato, Jeffrey Barrett, Sonia Goncalves, Ewan Harrison, David K. Jackson, Ian Johnston, Dominic Kwiatkowski, Cordelia Langford, John Sillitoe on behalf of the Wellcome Sanger Institute COVID-19 Surveillance Team |
| EPI_ISL_2638894, EPI_ISL_2638901 | Lighthouse Laboratory Plymouth | Wellcome Sanger Institute for the COVID-19 Genomics UK (COG-UK) Consortium | Lighthouse Laboratory Plymouth and Alex Alderton, Roberto Amato, Jeffrey Barrett, Sonia Goncalves, Ewan Harrison, David K. Jackson, Ian Johnston, Dominic Kwiatkowski, Cordelia Langford, John Sillitoe on behalf of the Wellcome Sanger Institute COVID-19 Surveillance Team |
| EPI_ISL_2638902, EPI_ISL_2638904 | Lighthouse Lab in Glasgow | Wellcome Sanger Institute for the COVID-19 Genomics UK (COG-UK) Consortium | Harper VanSteenhouse, Yumi Kasai, David Gray, Carol Clugston, Anna Dominiczak and Alex Alderton, Roberto Amato, Jeffrey Barrett, Sonia Goncalves, Ewan Harrison, David K. Jackson, Ian Johnston, Dominic Kwiatkowski, Cordelia Langford, John Sillitoe on behalf of the Wellcome Sanger Institute COVID-19 Surveillance Team |
| EPI_ISL_2638907 | Lighthouse Lab in Milton Keynes | Wellcome Sanger Institute for the COVID-19 Genomics UK (COG-UK) Consortium | The Lighthouse Lab in Milton Keynes and Alex Alderton, Roberto Amato, Jeffrey Barrett, Sonia Goncalves, Ewan Harrison, David K. Jackson, Ian Johnston, Dominic Kwiatkowski, Cordelia Langford, John Sillitoe on behalf of the Wellcome Sanger Institute COVID-19 Surveillance Team |
| EPI_ISL_2638923 | Lighthouse Laboratory Plymouth | Wellcome Sanger Institute for the COVID-19 Genomics UK | Lighthouse Laboratory Plymouth and Alex Alderton, Roberto Amato, Jeffrey Barrett, Sonia Goncalves, Ewan Harrison, David K. Jackson, Ian Johnston, |

| | | | |
|---|---|---|---|
| | | (COG-UK) Consortium | Dominic Kwiatkowski, Cordelia Langford, John Sillitoe on behalf of the Wellcome Sanger Institute COVID-19 Surveillance Team |
| EPI_ISL_2638961 | Health Services Laboratories | Wellcome Sanger Institute for the COVID-19 Genomics UK (COG-UK) Consortium | Health Services Laboratories and Alex Alderton, Roberto Amato, Jeffrey Barrett, Sonia Goncalves, Ewan Harrison, David K. Jackson, Ian Johnston, Dominic Kwiatkowski, Cordelia Langford, John Sillitoe on behalf of the Wellcome Sanger Institute COVID-19 Surveillance Team |
| EPI_ISL_2638975 | Lighthouse Laboratory Plymouth | Wellcome Sanger Institute for the COVID-19 Genomics UK (COG-UK) Consortium | Lighthouse Laboratory Plymouth and Alex Alderton, Roberto Amato, Jeffrey Barrett, Sonia Goncalves, Ewan Harrison, David K. Jackson, Ian Johnston, Dominic Kwiatkowski, Cordelia Langford, John Sillitoe on behalf of the Wellcome Sanger Institute COVID-19 Surveillance Team |
| EPI_ISL_2639001 | Health Services Laboratories | Wellcome Sanger Institute for the COVID-19 Genomics UK (COG-UK) Consortium | Health Services Laboratories and Alex Alderton, Roberto Amato, Jeffrey Barrett, Sonia Goncalves, Ewan Harrison, David K. Jackson, Ian Johnston, Dominic Kwiatkowski, Cordelia Langford, John Sillitoe on behalf of the Wellcome Sanger Institute COVID-19 Surveillance Team |
| EPI_ISL_2639002, EPI_ISL_2639004, EPI_ISL_2639018 | Lighthouse Lab in Milton Keynes | Wellcome Sanger Institute for the COVID-19 Genomics UK (COG-UK) Consortium | The Lighthouse Lab in Milton Keynes and Alex Alderton, Roberto Amato, Jeffrey Barrett, Sonia Goncalves, Ewan Harrison, David K. Jackson, Ian Johnston, Dominic Kwiatkowski, Cordelia Langford, John Sillitoe on behalf of the Wellcome Sanger Institute COVID-19 Surveillance Team |
| EPI_ISL_2639023 | Lighthouse Laboratory Plymouth | Wellcome Sanger Institute for the COVID-19 Genomics UK (COG-UK) Consortium | Lighthouse Laboratory Plymouth and Alex Alderton, Roberto Amato, Jeffrey Barrett, Sonia Goncalves, Ewan Harrison, David K. Jackson, Ian Johnston, Dominic Kwiatkowski, Cordelia Langford, John Sillitoe on behalf of the Wellcome Sanger Institute COVID-19 Surveillance Team |
| EPI_ISL_2639028 | Lighthouse Lab in Milton Keynes | Wellcome Sanger Institute for the COVID-19 Genomics UK (COG-UK) Consortium | The Lighthouse Lab in Milton Keynes and Alex Alderton, Roberto Amato, Jeffrey Barrett, Sonia Goncalves, Ewan Harrison, David K. Jackson, Ian Johnston, Dominic Kwiatkowski, Cordelia Langford, John Sillitoe on behalf of the Wellcome Sanger Institute COVID-19 Surveillance Team |
| EPI_ISL_2639104 | Lighthouse Laboratory Plymouth | Wellcome Sanger Institute for the COVID-19 Genomics UK (COG-UK) Consortium | Lighthouse Laboratory Plymouth and Alex Alderton, Roberto Amato, Jeffrey Barrett, Sonia Goncalves, Ewan Harrison, David K. Jackson, Ian Johnston, Dominic Kwiatkowski, Cordelia Langford, John Sillitoe on behalf of the Wellcome Sanger Institute COVID-19 Surveillance Team |
| EPI_ISL_2639110 | Lighthouse Lab in Milton Keynes | Wellcome Sanger Institute for the COVID-19 Genomics UK (COG-UK) Consortium | The Lighthouse Lab in Milton Keynes and Alex Alderton, Roberto Amato, Jeffrey Barrett, Sonia Goncalves, Ewan Harrison, David K. Jackson, Ian Johnston, Dominic Kwiatkowski, Cordelia Langford, John Sillitoe on behalf of the Wellcome Sanger Institute COVID-19 Surveillance Team |
| EPI_ISL_2639113 | Health Services Laboratories | Wellcome Sanger Institute for the COVID-19 Genomics UK (COG-UK) Consortium | Health Services Laboratories and Alex Alderton, Roberto Amato, Jeffrey Barrett, Sonia Goncalves, Ewan Harrison, David K. Jackson, Ian Johnston, Dominic Kwiatkowski, Cordelia Langford, John Sillitoe on behalf of the Wellcome Sanger Institute COVID-19 Surveillance Team |
| EPI_ISL_2639119 | Lighthouse Laboratory Plymouth | Wellcome Sanger Institute for the COVID-19 Genomics UK (COG-UK) Consortium | Lighthouse Laboratory Plymouth and Alex Alderton, Roberto Amato, Jeffrey Barrett, Sonia Goncalves, Ewan Harrison, David K. Jackson, Ian Johnston, Dominic Kwiatkowski, Cordelia Langford, John Sillitoe on behalf of the Wellcome Sanger Institute COVID-19 Surveillance Team |
| EPI_ISL_2639145 | Lighthouse Lab in Milton Keynes | Wellcome Sanger Institute for the COVID-19 Genomics UK (COG-UK) Consortium | The Lighthouse Lab in Milton Keynes and Alex Alderton, Roberto Amato, Jeffrey Barrett, Sonia Goncalves, Ewan Harrison, David K. Jackson, Ian Johnston, Dominic Kwiatkowski, Cordelia Langford, John Sillitoe on behalf of the Wellcome Sanger Institute COVID-19 Surveillance Team |
| EPI_ISL_2639168 | Health Services Laboratories | Wellcome Sanger Institute for the COVID-19 Genomics UK (COG-UK) Consortium | Health Services Laboratories and Alex Alderton, Roberto Amato, Jeffrey Barrett, Sonia Goncalves, Ewan Harrison, David K. Jackson, Ian Johnston, Dominic Kwiatkowski, Cordelia Langford, John Sillitoe on behalf of the Wellcome Sanger Institute COVID-19 Surveillance Team |
| EPI_ISL_2639169 | Lighthouse Laboratory Plymouth | Wellcome Sanger Institute for the COVID-19 Genomics UK (COG-UK) Consortium | Lighthouse Laboratory Plymouth and Alex Alderton, Roberto Amato, Jeffrey Barrett, Sonia Goncalves, Ewan Harrison, David K. Jackson, Ian Johnston, Dominic Kwiatkowski, Cordelia Langford, John Sillitoe on behalf of the Wellcome Sanger Institute COVID-19 Surveillance Team |
| EPI_ISL_2639170, EPI_ISL_2639195, EPI_ISL_2639197, EPI_ISL_2639198 | Lighthouse Lab in Milton Keynes | Wellcome Sanger Institute for the COVID-19 Genomics UK (COG-UK) Consortium | The Lighthouse Lab in Milton Keynes and Alex Alderton, Roberto Amato, Jeffrey Barrett, Sonia Goncalves, Ewan Harrison, David K. Jackson, Ian Johnston, Dominic Kwiatkowski, Cordelia Langford, John Sillitoe on behalf of the Wellcome Sanger Institute COVID-19 Surveillance Team |
| EPI_ISL_2639200 | Lighthouse Laboratory Plymouth | Wellcome Sanger Institute for the COVID-19 Genomics UK (COG-UK) Consortium | Lighthouse Laboratory Plymouth and Alex Alderton, Roberto Amato, Jeffrey Barrett, Sonia Goncalves, Ewan Harrison, David K. Jackson, Ian Johnston, Dominic Kwiatkowski, Cordelia Langford, John Sillitoe on behalf of the Wellcome Sanger Institute COVID-19 Surveillance Team |
| EPI_ISL_2639202 | Lighthouse Lab in Milton Keynes | Wellcome Sanger Institute for the COVID-19 Genomics UK (COG-UK) Consortium | The Lighthouse Lab in Milton Keynes and Alex Alderton, Roberto Amato, Jeffrey Barrett, Sonia Goncalves, Ewan Harrison, David K. Jackson, Ian Johnston, Dominic Kwiatkowski, Cordelia Langford, John Sillitoe on behalf of the Wellcome Sanger Institute COVID-19 Surveillance Team |
| EPI_ISL_2639204 | Health Services Laboratories | Wellcome Sanger Institute for the COVID-19 Genomics UK (COG-UK) Consortium | Health Services Laboratories and Alex Alderton, Roberto Amato, Jeffrey Barrett, Sonia Goncalves, Ewan Harrison, David K. Jackson, Ian Johnston, Dominic Kwiatkowski, Cordelia Langford, John Sillitoe on behalf of the Wellcome Sanger Institute COVID-19 Surveillance Team |
| EPI_ISL_2639223, EPI_ISL_2639252 | Lighthouse Lab in Milton Keynes | Wellcome Sanger Institute for the COVID-19 Genomics UK (COG-UK) Consortium | The Lighthouse Lab in Milton Keynes and Alex Alderton, Roberto Amato, Jeffrey Barrett, Sonia Goncalves, Ewan Harrison, David K. Jackson, Ian Johnston, Dominic Kwiatkowski, Cordelia Langford, John Sillitoe on behalf of the Wellcome Sanger Institute COVID-19 Surveillance Team |
| EPI_ISL_2639257, EPI_ISL_2639258, EPI_ISL_2639260, EPI_ISL_2639261, EPI_ISL_2639262, EPI_ISL_2639263, EPI_ISL_2639264, EPI_ISL_2639267, EPI_ISL_2639268, EPI_ISL_2639269, EPI_ISL_2639300, EPI_ISL_2639302, EPI_ISL_2639303, EPI_ISL_2639306, EPI_ISL_2639307, EPI_ISL_2639308, EPI_ISL_2639309, EPI_ISL_2639310, EPI_ISL_2639311, EPI_ISL_2639316, EPI_ISL_2639317, EPI_ISL_2639319, EPI_ISL_2639320, EPI_ISL_2639321, EPI_ISL_2639322, EPI_ISL_2639323, EPI_ISL_2639324, EPI_ISL_2639325, EPI_ISL_2639326, EPI_ISL_2639327, EPI_ISL_2639328, EPI_ISL_2639329, EPI_ISL_2639331, EPI_ISL_2639332, EPI_ISL_2639333, EPI_ISL_2639335, EPI_ISL_2639336, EPI_ISL_2639337, EPI_ISL_2639338, EPI_ISL_2639339, EPI_ISL_2639341, EPI_ISL_2639343, EPI_ISL_2639344, EPI_ISL_2639345, EPI_ISL_2639347, EPI_ISL_2639352, EPI_ISL_2639354, EPI_ISL_2639355, EPI_ISL_2639356, EPI_ISL_2639357, EPI_ISL_2639358, EPI_ISL_2639359, EPI_ISL_2639361, EPI_ISL_2639362, EPI_ISL_2639363, EPI_ISL_2639364, EPI_ISL_2639365, EPI_ISL_2639366, EPI_ISL_2639368, EPI_ISL_2639369, EPI_ISL_2639370, EPI_ISL_2639371, EPI_ISL_2639372, EPI_ISL_2639373, EPI_ISL_2639374, EPI_ISL_2639375, EPI_ISL_2639376, EPI_ISL_2639378, EPI_ISL_2639379, EPI_ISL_2639380, EPI_ISL_2639381, EPI_ISL_2639383, EPI_ISL_2639384, EPI_ISL_2639385, EPI_ISL_2639387, EPI_ISL_2639388, EPI_ISL_2639389, EPI_ISL_2639390, EPI_ISL_2639391, EPI_ISL_2639392, EPI_ISL_2639394, EPI_ISL_2639395, EPI_ISL_2639396, EPI_ISL_2639397, EPI_ISL_2639398, EPI_ISL_2639400, EPI_ISL_2639401, EPI_ISL_2639403, EPI_ISL_2639430, EPI_ISL_2639431, EPI_ISL_2639433, EPI_ISL_2639436, EPI_ISL_2639438, EPI_ISL_2639439, EPI_ISL_2639441, EPI_ISL_2639442, EPI_ISL_2639443, EPI_ISL_2639446, EPI_ISL_2639448, EPI_ISL_2639449, EPI_ISL_2639451, EPI_ISL_2639453, EPI_ISL_2639456, EPI_ISL_2639457, EPI_ISL_2639459, EPI_ISL_2639460, EPI_ISL_2639461, EPI_ISL_2639462, EPI_ISL_2639463, EPI_ISL_2639465, EPI_ISL_2639466, EPI_ISL_2639469, EPI_ISL_2639470, EPI_ISL_2639473, EPI_ISL_2639474, EPI_ISL_2639477, EPI_ISL_2639478, EPI_ISL_2639479, EPI_ISL_2639480, EPI_ISL_2639481, EPI_ISL_2639482, EPI_ISL_2639483, EPI_ISL_2639485, EPI_ISL_2639486, EPI_ISL_2639487, EPI_ISL_2639488, EPI_ISL_2639489, EPI_ISL_2639490, EPI_ISL_2639491, EPI_ISL_2639492, EPI_ISL_2639494, EPI_ISL_2639495, EPI_ISL_2639497, EPI_ISL_2639503, EPI_ISL_2639505, EPI_ISL_2639506, EPI_ISL_2639507, EPI_ISL_2639508, EPI_ISL_2639510, EPI_ISL_2639513, EPI_ISL_2639514, EPI_ISL_2639515, EPI_ISL_2639517, EPI_ISL_2639519, EPI_ISL_2639520, EPI_ISL_2639521, EPI_ISL_2639522, EPI_ISL_2639524, EPI_ISL_2639525, EPI_ISL_2639527, EPI_ISL_2639528, EPI_ISL_2639529, EPI_ISL_2639530, EPI_ISL_2639532, EPI_ISL_2639533, EPI_ISL_2639534, EPI_ISL_2639536, EPI_ISL_2639537, EPI_ISL_2639538, EPI_ISL_2639540, EPI_ISL_2639541, EPI_ISL_2639542, EPI_ISL_2639545, EPI_ISL_2639546, EPI_ISL_2639549, EPI_ISL_2639550, EPI_ISL_2639551, EPI_ISL_2639554, EPI_ISL_2639555, EPI_ISL_2639556, EPI_ISL_2639557, EPI_ISL_2639558, EPI_ISL_2639559, EPI_ISL_2639560, EPI_ISL_2639561, EPI_ISL_2639562, EPI_ISL_2639563, EPI_ISL_2639564, EPI_ISL_2639565, EPI_ISL_2639567, EPI_ISL_2639569, EPI_ISL_2639572, EPI_ISL_2639573, EPI_ISL_2639574, EPI_ISL_2639577, EPI_ISL_2639581, EPI_ISL_2639582, EPI_ISL_2639591, EPI_ISL_2639594, EPI_ISL_2639595, EPI_ISL_2639597, EPI_ISL_2639670 | | | |
| see above | Lighthouse Lab in Alderley Park | Wellcome Sanger Institute for the COVID-19 Genomics UK (COG-UK) Consortium | Jacquelyn Wynn, Mairead Hyland, The Lighthouse Lab in Alderley Park and Alex Alderton, Roberto Amato, Jeffrey Barrett, Sonia Goncalves, Ewan Harrison, David K. Jackson, Ian Johnston, Dominic Kwiatkowski, Cordelia Langford, John Sillitoe on behalf of the Wellcome Sanger Institute COVID-19 Surveillance Team |
| EPI_ISL_2639677 | Lighthouse Lab in Glasgow | Wellcome Sanger Institute for the COVID-19 Genomics UK (COG-UK) Consortium | Harper VanSteenhouse, Yumi Kasai, David Gray, Carol Clugston, Anna Dominiczak and Alex Alderton, Roberto Amato, Jeffrey Barrett, Sonia Goncalves, Ewan Harrison, David K. Jackson, Ian Johnston, Dominic Kwiatkowski, Cordelia Langford, John Sillitoe on behalf of the Wellcome Sanger Institute COVID-19 Surveillance Team |
| EPI_ISL_2639678, EPI_ISL_2639680, EPI_ISL_2639681, EPI_ISL_2639687, EPI_ISL_2639692, EPI_ISL_2639711, EPI_ISL_2639714, EPI_ISL_2639715, EPI_ISL_2639719 | Lighthouse Lab in Alderley Park | Wellcome Sanger Institute for the COVID-19 Genomics UK (COG-UK) Consortium | Jacquelyn Wynn, Mairead Hyland, The Lighthouse Lab in Alderley Park and Alex Alderton, Roberto Amato, Jeffrey Barrett, Sonia Goncalves, Ewan Harrison, David K. Jackson, Ian Johnston, Dominic Kwiatkowski, Cordelia Langford, John Sillitoe on behalf of the Wellcome Sanger Institute COVID-19 Surveillance Team |
| EPI_ISL_2639733 | Lighthouse Lab in Glasgow | Wellcome Sanger Institute for the COVID-19 Genomics UK (COG-UK) Consortium | Harper VanSteenhouse, Yumi Kasai, David Gray, Carol Clugston, Anna Dominiczak and Alex Alderton, Roberto Amato, Jeffrey Barrett, Sonia Goncalves, Ewan Harrison, David K. Jackson, Ian Johnston, Dominic Kwiatkowski, Cordelia Langford, John Sillitoe on behalf of the Wellcome Sanger Institute COVID-19 Surveillance Team |
| EPI_ISL_2639734, EPI_ISL_2639735, EPI_ISL_2639739, EPI_ISL_2639744, EPI_ISL_2639745, EPI_ISL_2639749, EPI_ISL_2639754, EPI_ISL_2639761, EPI_ISL_2639763, EPI_ISL_2639766, EPI_ISL_2639767, EPI_ISL_2639768, EPI_ISL_2639771, EPI_ISL_2639772, EPI_ISL_2639776, EPI_ISL_2639785, EPI_ISL_2639786, EPI_ISL_2639787, EPI_ISL_2639795, EPI_ISL_2639800, EPI_ISL_2639801, EPI_ISL_2639803, EPI_ISL_2639808, EPI_ISL_2639809, EPI_ISL_2639810, EPI_ISL_2639817, EPI_ISL_2639819, EPI_ISL_2639821, EPI_ISL_2639826, EPI_ISL_2639881, EPI_ISL_2639882, EPI_ISL_2639883 | | | |
| see above | Lighthouse Lab in Alderley Park | Wellcome Sanger Institute for the COVID-19 Genomics UK (COG-UK) Consortium | Jacquelyn Wynn, Mairead Hyland, The Lighthouse Lab in Alderley Park and Alex Alderton, Roberto Amato, Jeffrey Barrett, Sonia Goncalves, Ewan Harrison, David K. Jackson, Ian Johnston, Dominic Kwiatkowski, Cordelia Langford, John Sillitoe on behalf of the Wellcome Sanger Institute COVID-19 Surveillance Team |
| EPI_ISL_2639887 | Lighthouse Laboratory Plymouth | Wellcome Sanger Institute for the COVID-19 Genomics UK (COG-UK) Consortium | Lighthouse Laboratory Plymouth and Alex Alderton, Roberto Amato, Jeffrey Barrett, Sonia Goncalves, Ewan Harrison, David K. Jackson, Ian Johnston, Dominic Kwiatkowski, Cordelia Langford, John Sillitoe on behalf of the Wellcome Sanger Institute COVID-19 Surveillance Team |
| EPI_ISL_2651414, EPI_ISL_2651425, EPI_ISL_2651449, EPI_ISL_2651461, EPI_ISL_2651478 | Lighthouse Lab in Milton Keynes | Wellcome Sanger Institute for the COVID-19 Genomics UK (COG-UK) Consortium | The Lighthouse Lab in Milton Keynes and Alex Alderton, Roberto Amato, Jeffrey Barrett, Sonia Goncalves, Ewan Harrison, David K. Jackson, Ian Johnston, Dominic Kwiatkowski, Cordelia Langford, John Sillitoe on behalf of the Wellcome Sanger Institute COVID-19 Surveillance Team |

| | | | |
|---|---|---|---|
| EPI_ISL_2651661, EPI_ISL_2651697, EPI_ISL_2651847, EPI_ISL_2651867, EPI_ISL_2651892, EPI_ISL_2651893, EPI_ISL_2651894, EPI_ISL_2651895, EPI_ISL_2651897, EPI_ISL_2651898, EPI_ISL_2651900, EPI_ISL_2651901, EPI_ISL_2651902, EPI_ISL_2651903, EPI_ISL_2651904, EPI_ISL_2651905, EPI_ISL_2651907, EPI_ISL_2651908, EPI_ISL_2651909, EPI_ISL_2651910, EPI_ISL_2651911, EPI_ISL_2651912, EPI_ISL_2651913, EPI_ISL_2651915, EPI_ISL_2651916, EPI_ISL_2651917, EPI_ISL_2651918, EPI_ISL_2651920, EPI_ISL_2651921, EPI_ISL_2651922, EPI_ISL_2651923, EPI_ISL_2651924, EPI_ISL_2651925, EPI_ISL_2651926, EPI_ISL_2651927, EPI_ISL_2651928, EPI_ISL_2651931, EPI_ISL_2651932, EPI_ISL_2651933, EPI_ISL_2651934, EPI_ISL_2651935, EPI_ISL_2651936, EPI_ISL_2651937, EPI_ISL_2651939, EPI_ISL_2651940, EPI_ISL_2651941 | | | |
| see above | Lighthouse Lab in Glasgow | Wellcome Sanger Institute for the COVID-19 Genomics UK (COG-UK) Consortium | Harper VanSteenhouse, Yumi Kasai, David Gray, Carol Clugston, Anna Dominiczak and Alex Alderton, Roberto Amato, Jeffrey Barrett, Sonia Goncalves, Ewan Harrison, David K. Jackson, Ian Johnston, Dominic Kwiatkowski, Cordelia Langford, John Sillitoe on behalf of the Wellcome Sanger Institute COVID-19 Surveillance Team |
| EPI_ISL_2651943 | Lighthouse Laboratory Plymouth | Wellcome Sanger Institute for the COVID-19 Genomics UK (COG-UK) Consortium | Lighthouse Laboratory Plymouth and Alex Alderton, Roberto Amato, Jeffrey Barrett, Sonia Goncalves, Ewan Harrison, David K. Jackson, Ian Johnston, Dominic Kwiatkowski, Cordelia Langford, John Sillitoe on behalf of the Wellcome Sanger Institute COVID-19 Surveillance Team |
| EPI_ISL_2651947 | Lighthouse Lab in Alderley Park | Wellcome Sanger Institute for the COVID-19 Genomics UK (COG-UK) Consortium | Jacquelyn Wynn, Mairead Hyland, The Lighthouse Lab in Alderley Park and Alex Alderton, Roberto Amato, Jeffrey Barrett, Sonia Goncalves, Ewan Harrison, David K. Jackson, Ian Johnston, Dominic Kwiatkowski, Cordelia Langford, John Sillitoe on behalf of the Wellcome Sanger Institute COVID-19 Surveillance Team |
| EPI_ISL_2663219 | HealthPartners Central Lab | Minnesota Department of Health, Public Health Laboratory | Alexandra Lorentz, Jacob Garfin, Matt Plumb, and Xiong Wang |
| EPI_ISL_2663553 | NYU Langone Health | Departments of Pathology and Medicine, New York University School of Medicine | Adriana Heguy, Dacia Dimartino, Emily Guzman, Christian Marier, Peter Meyn, Sitharam Ramaswami, Gael Westby, Paul Zappile, Yutong Zhang, Paolo Cotzia, Guiqing Wang |
| EPI_ISL_2663880 | Lighthouse Lab in Milton Keynes | Wellcome Sanger Institute for the COVID-19 Genomics UK (COG-UK) Consortium | The Lighthouse Lab in Milton Keynes and Alex Alderton, Roberto Amato, Jeffrey Barrett, Sonia Goncalves, Ewan Harrison, David K. Jackson, Ian Johnston, Dominic Kwiatkowski, Cordelia Langford, John Sillitoe on behalf of the Wellcome Sanger Institute COVID-19 Surveillance Team |
| EPI_ISL_2664104, EPI_ISL_2664123, EPI_ISL_2664125, EPI_ISL_2664140, EPI_ISL_2664166, EPI_ISL_2664168, EPI_ISL_2664170, EPI_ISL_2664188, EPI_ISL_2664194, EPI_ISL_2664198, EPI_ISL_2664201, EPI_ISL_2664205, EPI_ISL_2664209, EPI_ISL_2664212, EPI_ISL_2664301, EPI_ISL_2664304, EPI_ISL_2664319, EPI_ISL_2664346, EPI_ISL_2664366, EPI_ISL_2664369, EPI_ISL_2664373, EPI_ISL_2664389, EPI_ISL_2664390, EPI_ISL_2664392, EPI_ISL_2664397, EPI_ISL_2664401, EPI_ISL_2664408, EPI_ISL_2664422 | | | |
| see above | Lighthouse Lab in Alderley Park | Wellcome Sanger Institute for the COVID-19 Genomics UK (COG-UK) Consortium | Jacquelyn Wynn, Mairead Hyland, The Lighthouse Lab in Alderley Park and Alex Alderton, Roberto Amato, Jeffrey Barrett, Sonia Goncalves, Ewan Harrison, David K. Jackson, Ian Johnston, Dominic Kwiatkowski, Cordelia Langford, John Sillitoe on behalf of the Wellcome Sanger Institute COVID-19 Surveillance Team |
| EPI_ISL_2664423 | Lighthouse Lab in Milton Keynes | Wellcome Sanger Institute for the COVID-19 Genomics UK (COG-UK) Consortium | The Lighthouse Lab in Milton Keynes and Alex Alderton, Roberto Amato, Jeffrey Barrett, Sonia Goncalves, Ewan Harrison, David K. Jackson, Ian Johnston, Dominic Kwiatkowski, Cordelia Langford, John Sillitoe on behalf of the Wellcome Sanger Institute COVID-19 Surveillance Team |
| EPI_ISL_2664440, EPI_ISL_2664549, EPI_ISL_2664566, EPI_ISL_2664584, EPI_ISL_2664680, EPI_ISL_2664706, EPI_ISL_2664709, EPI_ISL_2665042, EPI_ISL_2665225, EPI_ISL_2665260, EPI_ISL_2665296 | | | |
| see above | Lighthouse Lab in Alderley Park | Wellcome Sanger Institute for the COVID-19 Genomics UK (COG-UK) Consortium | Jacquelyn Wynn, Mairead Hyland, The Lighthouse Lab in Alderley Park and Alex Alderton, Roberto Amato, Jeffrey Barrett, Sonia Goncalves, Ewan Harrison, David K. Jackson, Ian Johnston, Dominic Kwiatkowski, Cordelia Langford, John Sillitoe on behalf of the Wellcome Sanger Institute COVID-19 Surveillance Team |
| EPI_ISL_2665447, EPI_ISL_2665873, EPI_ISL_2666131, EPI_ISL_2666384, EPI_ISL_2666515, EPI_ISL_2666744, EPI_ISL_2666809, EPI_ISL_2666828, EPI_ISL_2666913, EPI_ISL_2667010, EPI_ISL_2667233, EPI_ISL_2667607, EPI_ISL_2667871, EPI_ISL_2668328, EPI_ISL_2668510, EPI_ISL_2668554, EPI_ISL_2668747, EPI_ISL_2668987, EPI_ISL_2669191, EPI_ISL_2669196, EPI_ISL_2669198, EPI_ISL_2669409, EPI_ISL_2669604, EPI_ISL_2669906, EPI_ISL_2670378, EPI_ISL_2670389, EPI_ISL_2670699, EPI_ISL_2670831, EPI_ISL_2671046 | | | |
| see above | Department of Bacteria, Parasites and Fungi, Statens Serum Institut, Copenhagen, Denmark | Statens Serum Institut Bioinformatics and Microbial Genomics | Danish Covid-19 Genome Consortium |
| EPI_ISL_2671462 | Center for Laboratory Medicine | Center for Laboratory Medicine | Yannick Gerth |
| EPI_ISL_2673381, EPI_ISL_2673385 | Dutch COVID-19 response team | National Institute for Public Health and the Environment (RIVM) | Adam Meijer, Harry Vennema, Dirk Eggink, Jeroen Cremer, Sharon van den Brink, Bas van der Veer, AnneMarie van den Brandt, Lisa Wijsman, Kim Freriks, Ryanne Jaarsma, Euníce Then, Lynn Aarts, Sanne Bos, Melissa van Tuil, Linda van de Nes, Florian Zwagemaker, Dennis Schmitz, Annelies Kroneman, Karim Hajji, Chantal Reusken, on behalf of the national COVID-19 response team |
| EPI_ISL_2673676, EPI_ISL_2673677, EPI_ISL_2673678 | OSPEDALE CIVILE TERAMO - CENTRO TRASFUSIONALE | Istituto Zooprofilattico Sperimentale dell'Abruzzo e Molise "G. Caporale" | Lorusso A, Marcacci M, Di Domenico M, Ancora M, Curini V, Di Lollo Valeria, Mangone I, Rinaldi A, Delli Compagni E, Scialabba S, Caporale M, Di Pasquale A, Cammà C, Puglia I, Calistri P, Savini G |
| EPI_ISL_2674808 | SARS-CoV-2 testing team, National Institute of Infectious Diseases | Pathogen Genomics Center, National Institute of Infectious Diseases | Tsuyoshi Sekizuka, Kentaro Itokawa, Rina Tanaka, Masanori Hashino, Nozomu Hanaoka, Masumichi Saito, Naomi Nojiri, Hazuka Y Furihata, Hiromizu Takahashi, Tsuguto Fujimoto, Makoto Kuroda |
| EPI_ISL_2675078 | Vestfold Hospital, Toensberg Department of Microbiology | Norwegian Institute of Public Health, Department of Virology | Kathrine Stene-Johansen, Kamilla Heddeland Instefjord, Hilde Elshaug, Garcia Llorente Ignacio, Jon Bråte, Engebretsen Serina Beate,Pedersen Benedikte Nevjen, Line Victoria Moen, Debech Nadia, Atiya R Ali,Marie Paulsen Madsen, Rasmus Riis Kopperud, Hilde Vollan, Karoline Bragstad, Olav Hungnes |
| EPI_ISL_2675096 | Furst Medical Laboratory | Norwegian Institute of Public Health, Department of Virology | Kathrine Stene-Johansen, Kamilla Heddeland Instefjord, Hilde Elshaug, Garcia Llorente Ignacio, Jon Bråte, Engebretsen Serina Beate,Pedersen Benedikte Nevjen, Line Victoria Moen, Debech Nadia, Atiya R Ali,Marie Paulsen Madsen, Rasmus Riis Kopperud, Hilde Vollan, Karoline Bragstad, Olav Hungnes |
| EPI_ISL_2675204 | Houston Health Dept. | Houston Health Dept. | Ryker Penn, Pamela Brown, Adolpho Lara, Yanlai Lai |
| EPI_ISL_2675462 | Department of Bacteria, Parasites and Fungi, Statens Serum Institut, Copenhagen, Denmark | Statens Serum Institut Bioinformatics and Microbial Genomics | Danish Covid-19 Genome Consortium |
| EPI_ISL_2678743 | Klinikum Ernst von Bergmann gemeinnützige GmbH - stationärer Bereich | Robert Koch Institute | unknown |
| EPI_ISL_2681473, EPI_ISL_2681474 | University Hospitals of Geneva, Laboratory of Virology | HUG, Laboratory of Virology and the Health2030 Genome Center | Samuel Cordey, Ana Rita Goncalves, Laurent Kaiser, Lorenzo Cerutti, Henri Pegeot, Melyssa Elies, Deborah Penet, Keith Harshman, Ioannis Xenarios, Emmanouil Dermitzakis |
| EPI_ISL_2681759, EPI_ISL_2681767 | Lighthouse Lab in Alderley Park | Wellcome Sanger Institute for the COVID-19 Genomics UK (COG-UK) Consortium | Jacquelyn Wynn, Mairead Hyland, The Lighthouse Lab in Alderley Park and Alex Alderton, Roberto Amato, Jeffrey Barrett, Sonia Goncalves, Ewan Harrison, David K. Jackson, Ian Johnston, Dominic Kwiatkowski, Cordelia Langford, John Sillitoe on behalf of the Wellcome Sanger Institute COVID-19 Surveillance Team |
| EPI_ISL_2682063, EPI_ISL_2682198 | Lighthouse Lab in Glasgow | Wellcome Sanger Institute for the COVID-19 Genomics UK (COG-UK) Consortium | Harper VanSteenhouse, Yumi Kasai, David Gray, Carol Clugston, Anna Dominiczak and Alex Alderton, Roberto Amato, Jeffrey Barrett, Sonia Goncalves, Ewan Harrison, David K. Jackson, Ian Johnston, Dominic Kwiatkowski, Cordelia Langford, John Sillitoe on behalf of the Wellcome Sanger Institute COVID-19 Surveillance Team |
| EPI_ISL_2682388, EPI_ISL_2682402, EPI_ISL_2682433, EPI_ISL_2682562, EPI_ISL_2682662, EPI_ISL_2682666, EPI_ISL_2682802, EPI_ISL_2682804, EPI_ISL_2682812, EPI_ISL_2682816, EPI_ISL_2682818, EPI_ISL_2682821, EPI_ISL_2682822, EPI_ISL_2682823, EPI_ISL_2682825, EPI_ISL_2682830, EPI_ISL_2682832, EPI_ISL_2682838, EPI_ISL_2682843, EPI_ISL_2682844, EPI_ISL_2682854, EPI_ISL_2682856, EPI_ISL_2682857, EPI_ISL_2682862, EPI_ISL_2682864, EPI_ISL_2682865, EPI_ISL_2682866, EPI_ISL_2682876, EPI_ISL_2682882, EPI_ISL_2682884, EPI_ISL_2682890, EPI_ISL_2682893, EPI_ISL_2682897, EPI_ISL_2682898 | | | |
| see above | Lighthouse Lab in Alderley Park | Wellcome Sanger Institute for the COVID-19 Genomics UK (COG-UK) Consortium | Jacquelyn Wynn, Mairead Hyland, The Lighthouse Lab in Alderley Park and Alex Alderton, Roberto Amato, Jeffrey Barrett, Sonia Goncalves, Ewan Harrison, David K. Jackson, Ian Johnston, Dominic Kwiatkowski, Cordelia Langford, John Sillitoe on behalf of the Wellcome Sanger Institute COVID-19 Surveillance Team |
| EPI_ISL_2683070, EPI_ISL_2683435, EPI_ISL_2683468, EPI_ISL_2683483 | Lighthouse Lab in Glasgow | Wellcome Sanger Institute for the COVID-19 Genomics UK (COG-UK) Consortium | Harper VanSteenhouse, Yumi Kasai, David Gray, Carol Clugston, Anna Dominiczak and Alex Alderton, Roberto Amato, Jeffrey Barrett, Sonia Goncalves, Ewan Harrison, David K. Jackson, Ian Johnston, Dominic Kwiatkowski, Cordelia Langford, John Sillitoe on behalf of the Wellcome Sanger Institute COVID-19 Surveillance Team |
| EPI_ISL_2686643, EPI_ISL_2686652, EPI_ISL_2686653, EPI_ISL_2686660, EPI_ISL_2686666, EPI_ISL_2686670, EPI_ISL_2686673, EPI_ISL_2686674, EPI_ISL_2686676, EPI_ISL_2686679, EPI_ISL_2686685, EPI_ISL_2686686, EPI_ISL_2686687, EPI_ISL_2686688, EPI_ISL_2686692, EPI_ISL_2686693 | | | |
| see above | Fulgent Genetics | Centers for Disease Control and Prevention Division of Viral Diseases, Pathogen Discovery | Dakota Howard, Dhwani Batra, Peter W. Cook, Kara Moser, Adrian Paskey, Jason Caravas, Benjamin Rambo-Martin, Shatavia Morrison, Christopher Gulvick, Scott Sammons, Yvette Unoarumhi, Darlene Wagner, Matthew Schmerer, Harry Gao, Mickey Li, John Gao, Joseph Fierro, Benafsh Sapra, Becky Tsai, Yan Meng, Doreen Ng, James Xie, Clinton R. Paden, Duncan MacCannell |
| EPI_ISL_2687996 | National Institute of Infectious Diseases-Prof. Dr. Matei Bals Molecular Diagnostics Laboratory | National Institute of Infectious Diseases-Prof. Dr. Matei Bals Molecular Diagnostics Laboratory | Corina Casangiu, Leontina Banica, Marius Surleac, Petre Milu, Robert Hohan, Simona Paraschiv, Dan Otelea |
| EPI_ISL_2693033 | Oregon State Public Health Laboratory | Oregon State Public Health Laboratory | Rafia Razzaque, Eugene Yeboah, Vanda Makris, Laura Tsaknaridis, John Fontana and Shane Sevey |

| | | | |
|---|---|---|---|
| EPI_ISL_2693086 | NHLS_VIRO | KRISP, KZN Research Innovation and Sequencing Platform | Giandhari Jennifer, Pillay Sureshnee, Yajna Ramphal, Naidoo Yeshnee, Tshabuila Derek,Tegally Houriiyah, San James, Wilkinson Eduan, de Oliveira Tulio |
| EPI_ISL_2693271, EPI_ISL_2693272, EPI_ISL_2693274 | UW Virology Lab | UW Virology Lab | Pavitra Roychoudhury, Hong Xie, Lasata Shrestha, Tien V. Nguyen, Shah Mohamed Bakhash, Michelle Lin, Noah R. Baker, Ricardo Perez, Sean Ellis, Nathan Breit, Robert J. Livingston, Meei-Li Huang, Keith R Jerome, Patrick Mathias, Alexander Greninger |
| EPI_ISL_2694808, EPI_ISL_2694821, EPI_ISL_2694832, EPI_ISL_2694847, EPI_ISL_2694859 | Labor Berlin Charite Vivantes GmbH / Institut fur Virologie | Charite Universitatsmedizin Berlin, Institut fur Virologie/Labor Berlin | Peter Menzel, Christine Stephan, Rolf Schwarzer, Victor M Corman, Barbara Muhlemann, Terry Jones, Christian Drosten |
| EPI_ISL_2694898, EPI_ISL_2694899 | H Santarem | Instituto Nacional de Saude (INSA) | Borges et al |
| EPI_ISL_2695088 | H Vila Franca Xira | Instituto Nacional de Saude (INSA) | Borges et al |
| EPI_ISL_2695162 | CH Setubal | Instituto Nacional de Saude (INSA) | Borges et al |
| EPI_ISL_2695206, EPI_ISL_2695207, EPI_ISL_2695208, EPI_ISL_2695209, EPI_ISL_2695210, EPI_ISL_2695211, EPI_ISL_2695212, EPI_ISL_2695214, EPI_ISL_2695215 | ULS Litoral Alentejano | Instituto Nacional de Saude (INSA) and Institute of Biomedicine (iBiMed), Universidade de Aveiro | Borges et al |
| EPI_ISL_2695257 | SESARAM | Instituto Nacional de Saude (INSA) | Borges et al |
| EPI_ISL_2695268 | CHU Sao Joao, Porto | Instituto Nacional de Saude (INSA) | Borges et al |
| EPI_ISL_2695287 | H Braga | Instituto Nacional de Saude (INSA) | Borges et al |
| EPI_ISL_2695295, EPI_ISL_2695296, EPI_ISL_2695297 | CHTMAD | Instituto Nacional de Saude (INSA) | Borges et al |
| EPI_ISL_2695358, EPI_ISL_2695359 | Lab Dra Celeste Formosinho | Instituto Nacional de Saude (INSA) | Borges et al |
| EPI_ISL_2695381, EPI_ISL_2695383, EPI_ISL_2695384, EPI_ISL_2695385, EPI_ISL_2695387 | Labeto - CAB - Leiria | Instituto Nacional de Saude (INSA) | Borges et al |
| EPI_ISL_2695403 | Lab La Salete Robles - VN Famalicao | Instituto Nacional de Saude (INSA) | Borges et al |
| EPI_ISL_2695416 | H Guimaraes | Instituto Nacional de Saude (INSA) | Borges et al |
| EPI_ISL_2695466, EPI_ISL_2695467, EPI_ISL_2695468, EPI_ISL_2695498, EPI_ISL_2695499, EPI_ISL_2695500 | ARS Algarve - Laboratorio Laura Ayres | Instituto Nacional de Saude (INSA) | Borges et al |
| EPI_ISL_2698595 | WHO National Influenza Centre Russian Federation | WHO National Influenza Centre Russian Federation | Andrey Komissarov, Artem Fadeev, Kseniya Komissarova, Oula Mansour, Kirill Varchenko, Mikhail Bakaev, Tamila Musaeva, Veronika Eder, Maria Pisareva, Nikita Yolshin, Daria Danilenko, Ksenia Safina, Elena Nabieva, Georgii Bazykin, Dmitry Lioznov |
| EPI_ISL_2700097, EPI_ISL_2700999, EPI_ISL_2701131 | Lighthouse Lab in Alderley Park | Wellcome Sanger Institute for the COVID-19 Genomics UK (COG-UK) Consortium | Jacquelyn Wynn, Mairead Hyland, The Lighthouse Lab in Alderley Park and Alex Alderton, Roberto Amato, Jeffrey Barrett, Sonia Goncalves, Ewan Harrison, David K. Jackson, Ian Johnston, Dominic Kwiatkowski, Cordelia Langford, John Sillitoe on behalf of the Wellcome Sanger Institute COVID-19 Surveillance Team |
| EPI_ISL_2701140 | Lighthouse Lab in Glasgow | Wellcome Sanger Institute for the COVID-19 Genomics UK (COG-UK) Consortium | Harper VanSteenhouse, Yumi Kasai, David Gray, Carol Clugston, Anna Dominiczak and Alex Alderton, Roberto Amato, Jeffrey Barrett, Sonia Goncalves, Ewan Harrison, David K. Jackson, Ian Johnston, Dominic Kwiatkowski, Cordelia Langford, John Sillitoe on behalf of the Wellcome Sanger Institute COVID-19 Surveillance Team |
| EPI_ISL_2701344, EPI_ISL_2701352, EPI_ISL_2701377, EPI_ISL_2701392, EPI_ISL_2701412, EPI_ISL_2701419, EPI_ISL_2701503, EPI_ISL_2701510, EPI_ISL_2701524, EPI_ISL_2701566, EPI_ISL_2701573, EPI_ISL_2701598, EPI_ISL_2701603, EPI_ISL_2701607, EPI_ISL_2701611, EPI_ISL_2701717, EPI_ISL_2701720, EPI_ISL_2701749, EPI_ISL_2701777, EPI_ISL_2701782, EPI_ISL_2701795, EPI_ISL_2701808, EPI_ISL_2701809, EPI_ISL_2701815, EPI_ISL_2701820, EPI_ISL_2701821, EPI_ISL_2701853, EPI_ISL_2701854, EPI_ISL_2701883, EPI_ISL_2701888, EPI_ISL_2701941, EPI_ISL_2701973, EPI_ISL_2701979, EPI_ISL_2701987, EPI_ISL_2701993, EPI_ISL_2701996, EPI_ISL_2702022, EPI_ISL_2702024, EPI_ISL_2702035, EPI_ISL_2702482, EPI_ISL_2702487, EPI_ISL_2702607, EPI_ISL_2702610, EPI_ISL_2702611, EPI_ISL_2702630, EPI_ISL_2702631, EPI_ISL_2702661, EPI_ISL_2702665, EPI_ISL_2702689, EPI_ISL_2702706, EPI_ISL_2702708, EPI_ISL_2702713, EPI_ISL_2702717, EPI_ISL_2702722, EPI_ISL_2702728, EPI_ISL_2702738, EPI_ISL_2702812, EPI_ISL_2703186, EPI_ISL_2703199, EPI_ISL_2703300, EPI_ISL_2703353, EPI_ISL_2703387, EPI_ISL_2703388, EPI_ISL_2703390, EPI_ISL_2703785, EPI_ISL_2703793 | | | |
| see above | Lighthouse Lab in Alderley Park | Wellcome Sanger Institute for the COVID-19 Genomics UK (COG-UK) Consortium | Jacquelyn Wynn, Mairead Hyland, The Lighthouse Lab in Alderley Park and Alex Alderton, Roberto Amato, Jeffrey Barrett, Sonia Goncalves, Ewan Harrison, David K. Jackson, Ian Johnston, Dominic Kwiatkowski, Cordelia Langford, John Sillitoe on behalf of the Wellcome Sanger Institute COVID-19 Surveillance Team |
| EPI_ISL_2703804 | Lighthouse Lab in Milton Keynes | Wellcome Sanger Institute for the COVID-19 Genomics UK (COG-UK) Consortium | The Lighthouse Lab in Milton Keynes and Alex Alderton, Roberto Amato, Jeffrey Barrett, Sonia Goncalves, Ewan Harrison, David K. Jackson, Ian Johnston, Dominic Kwiatkowski, Cordelia Langford, John Sillitoe on behalf of the Wellcome Sanger Institute COVID-19 Surveillance Team |
| EPI_ISL_2704283, EPI_ISL_2704287, EPI_ISL_2704424, EPI_ISL_2704491, EPI_ISL_2704668 | Lighthouse Lab in Alderley Park | Wellcome Sanger Institute for the COVID-19 Genomics UK (COG-UK) Consortium | Jacquelyn Wynn, Mairead Hyland, The Lighthouse Lab in Alderley Park and Alex Alderton, Roberto Amato, Jeffrey Barrett, Sonia Goncalves, Ewan Harrison, David K. Jackson, Ian Johnston, Dominic Kwiatkowski, Cordelia Langford, John Sillitoe on behalf of the Wellcome Sanger Institute COVID-19 Surveillance Team |
| EPI_ISL_2705571 | University College London, Great Ormond Street Hospital for Children NHS Foundation Trust, Imperial College Healthcare NHS Trust | COVID-19 Genomics UK (COG-UK) Consortium | Sergi Castellano, Rachel Williams, Mark Kristiansen, Paola Resende Silva, Sunando Roy, Tony Brooks, Helena Tutill, Paola Niola, Patricia Dyal, Charlotte Williams, Leysa Forrest, Yasmin Panchbhaya, Jacqueline Findlay, Samuel Weeks, Julianne Brown, Kathryn Harris, Paul Randell, James Price, Alison Holmes, Judith Breuer |
| EPI_ISL_2705575 | Centers for Disease Control, R.O.C. (Taiwan) | Centers for Disease Control, R.O.C. (Taiwan) | Ji-Rong Yang, Yu-Chi-Lin, Jung-Jung Mu, Ming-Tsan-Liu |
| EPI_ISL_2705590, EPI_ISL_2705700, EPI_ISL_2705702, EPI_ISL_2705705, EPI_ISL_2705706, EPI_ISL_2705785 | University College London, Great Ormond Street Hospital for Children NHS Foundation Trust, Imperial College Healthcare NHS Trust | COVID-19 Genomics UK (COG-UK) Consortium | Sergi Castellano, Rachel Williams, Mark Kristiansen, Paola Resende Silva, Sunando Roy, Tony Brooks, Helena Tutill, Paola Niola, Patricia Dyal, Charlotte Williams, Leysa Forrest, Yasmin Panchbhaya, Jacqueline Findlay, Samuel Weeks, Julianne Brown, Kathryn Harris, Paul Randell, James Price, Alison Holmes, Judith Breuer |
| EPI_ISL_2706170, EPI_ISL_2706177, EPI_ISL_2706187, EPI_ISL_2706263, EPI_ISL_2706285, EPI_ISL_2706418, EPI_ISL_2706444, EPI_ISL_2706475, EPI_ISL_2706494 | Northumbria University / South Tees Hospitals NHS Foundation Trust / North Cumbria Integrated Care NHS Foundation Trust / North Tees and Hartlepool NHS Foundation Trust / Newcastle Hospitals NHS Foundation Trust | COVID-19 Genomics UK (COG-UK) Consortium | Darren L Smith,Andrew Nelson,Matthew Bashton,Greg R Young,Joshua Loh,John Allan,Mohammad A Tariq,Giles S Holt,Gary Black,Wen C Yew,Lynn Dover,Paul Baker,Steve Liggett,Sarah Essex,Jane Greenaway,Debra Padgett,Clive Graham,Garren Scott,Edward Barton,Emma Swindells,Brendan Payne,Jennifer Collins,Yusri Taha,Gary Eltringham |
| EPI_ISL_2707798, EPI_ISL_2707908, EPI_ISL_2707910, EPI_ISL_2707911, EPI_ISL_2707912, EPI_ISL_2707913, EPI_ISL_2707915, EPI_ISL_2707918, EPI_ISL_2707919, EPI_ISL_2707920, EPI_ISL_2707923, EPI_ISL_2707925, EPI_ISL_2707927, EPI_ISL_2707933, EPI_ISL_2707937, EPI_ISL_2707940, EPI_ISL_2707943, EPI_ISL_2707946, EPI_ISL_2707952, EPI_ISL_2707953, EPI_ISL_2707954, EPI_ISL_2707957, EPI_ISL_2707959, EPI_ISL_2707960, EPI_ISL_2707964, EPI_ISL_2707970, EPI_ISL_2707981, EPI_ISL_2707984, EPI_ISL_2707986, EPI_ISL_2708008, EPI_ISL_2708010, EPI_ISL_2708022, EPI_ISL_2708023, EPI_ISL_2708024, EPI_ISL_2708026, EPI_ISL_2708032, EPI_ISL_2708044, EPI_ISL_2708046, EPI_ISL_2708047, EPI_ISL_2708048, EPI_ISL_2708104, EPI_ISL_2708105, EPI_ISL_2708113, EPI_ISL_2708114, EPI_ISL_2708116, EPI_ISL_2708120, EPI_ISL_2708130, EPI_ISL_2708136, EPI_ISL_2708137 | | | |
| see above | Originating lab: Wales Specialist Virology Centre Sequencing lab: Pathogen Genomics Unit | Public Health Wales Microbiology Cardiff Wales Specialist Virology Centre | Catherine Moore, Johnathan Evans, Laura Gifford, Malorie Perry, Simon Cottrell, Angela Marchbank, Alec Birchley, Alexander Adams, Amy Gaskin, Bree Gatica-Wilcox, Jason Coombes, Joel Southgate, Lauren Gilbert, Lee Graham, Nicole Pacchiarini, Sara Kumziene-Summerhayes, Sarah Taylor, Sophie Jones, Sara Rey, Matthew Bull, Joanne Watkins, Sally Corden, Tom Connor |
| EPI_ISL_2709441 | West Java Health Laboratory | Eijkman Institute for Molecular Biology, National Research and Innovation Agency; West Java Health Laboratory | Lydia V. Panggalo, Iskandar Adnan, Sukma Oktavianthi, Lidwina Priliani, Edison Johar, Frilasita A Yudhaputri, Muhammad Rezki Rasyak, Willy Agustine, Hidayat Trimarsanto, Azzania Fibriani, Ema Rahmawati, Ryan Bayusantika Ristandi, Rifky Waluyajati Rachman, Cut Nur Cinthia Alamanda, Safarina G Malik, Khin Saw Myint, Amin Soebandrio |
| EPI_ISL_2709442 | West Java Health Laboratory | Eijkman Institute for Molecular Biology, National Research and Innovation Agency; West Java Health Laboratory | Iskandar Adnan, Sukma Oktavianthi, Lidwina Priliani, Edison Johar, Frilasita A Yudhaputri, Muhammad Rezki Rasyak, Willy Agustine, Hidayat Trimarsanto, Lydia V. Panggalo, Azzania Fibriani, Ema Rahmawati, Ryan Bayusantika Ristandi, Rifky Waluyajati Rachman, Cut Nur Cinthia Alamanda, Safarina G Malik, Khin Saw Myint, Amin Soebandrio |

| | | | |
|---|---|---|---|
| EPI_ISL_2709736, EPI_ISL_2709737 | National Institute of Public Health | State Veterinary Institute Prague | Nagy,A;Cernikov,L;Stara,M;Suri,T;Vecerova,J;Jirincova,H |
| EPI_ISL_2710258 | NHLS Charlotte Maxeke Johannesburg Academic Hospital and the University of the Witwatersrand | KRISP, KZN Research Innovation and Sequencing Platform | Florette Treurnicht, Bulelani Manene, Kathleen Subramoney, Giandhari Jennifer, Pillay Sureshnee, Yajna Ramphal, Naidoo Yeshnee, Tshabuila Derek,Tegally Houriiyah, San James, Wilkinson Eduan, de Oliveira Tulio |
| EPI_ISL_2712849, EPI_ISL_2712854, EPI_ISL_2712856 | Hospital Universitari Vall d'Hebron - Vall d'Hebron Institut de Recerca | Hospital Universitari Vall d'Hebron - Vall d'Hebron Institut de Recerca | Cristina Andrés, Maria Piñana, Alejandra González-Sánchez, Damir Garcia-Cehic, Ariadna Rando, Juliana Esperalba, Maria Gema Codina, Carla Castillo, Maria Carmen Martin, Tomàs Pumarola, Josep Quer, Andrés Antón |
| EPI_ISL_2712939 | Wyoming Public Health Laboratory | Wyoming Public Health Laboratory | Jim Mildenberger, Taylor Fearing, Channing Weber, Ashley Norberg, Chayse Rowley, Marley Goetz, Brian Dominguez, Elliot Thomasson, Sam Britz, Cari Sloma, Robert Petit, and Rob Christensen |
| EPI_ISL_2716210 | The Jackson Laboratory | The Jackson Laboratory | Long J, Renzette N, Adams M, Omerza G, Kelly K, Li L |
| EPI_ISL_2716404 | Arizona State Public Health Laboratory | Arizona State Public Health Laboratory | Trung Huynh, Jessica Escobar, Katherine Fullerton, Nobuko Fukushima, Matthew Contursi, Stacy White, Linda Getsinger, Victor Waddell |
| EPI_ISL_2716468 | Washington State Department of Health Public Health Laboratories | Washington State Department of Health Public Health Laboratories | Drew MacKellar, Philip Dykema, Denny Russell, Joenice Gonzalez, Hannah Gray, Geoff Melly, Vanessa De Los Santos, Darren Lucas, JohnAric Peterson, Avi Singh, Rebecca Cao |
| EPI_ISL_2717791, EPI_ISL_2718283 | Lighthouse Lab in Alderley Park | Wellcome Sanger Institute for the COVID-19 Genomics UK (COG-UK) Consortium | Jacquelyn Wynn, Mairead Hyland, The Lighthouse Lab in Alderley Park and Alex Alderton, Roberto Amato, Jeffrey Barrett, Sonia Goncalves, Ewan Harrison, David K. Jackson, Ian Johnston, Dominic Kwiatkowski, Cordelia Langford, John Sillitoe on behalf of the Wellcome Sanger Institute COVID-19 Surveillance Team |
| EPI_ISL_2718295, EPI_ISL_2718297, EPI_ISL_2718376, EPI_ISL_2718377, EPI_ISL_2718389, EPI_ISL_2718394, EPI_ISL_2718395, EPI_ISL_2718397, EPI_ISL_2718398, EPI_ISL_2718410, EPI_ISL_2718416, EPI_ISL_2718421, EPI_ISL_2718425, EPI_ISL_2718429, EPI_ISL_2718435, EPI_ISL_2718438, EPI_ISL_2718439, EPI_ISL_2718440, EPI_ISL_2718445, EPI_ISL_2718446, EPI_ISL_2718451, EPI_ISL_2718453, EPI_ISL_2718458, EPI_ISL_2718462, EPI_ISL_2718466, EPI_ISL_2718467, EPI_ISL_2718470, EPI_ISL_2718477, EPI_ISL_2718478, EPI_ISL_2718480, EPI_ISL_2718486, EPI_ISL_2718488 | | | |
| see above | Lighthouse Lab in Glasgow | Wellcome Sanger Institute for the COVID-19 Genomics UK (COG-UK) Consortium | Harper VanSteenhouse, Yumi Kasai, David Gray, Carol Clugston, Anna Dominiczak and Alex Alderton, Roberto Amato, Jeffrey Barrett, Sonia Goncalves, Ewan Harrison, David K. Jackson, Ian Johnston, Dominic Kwiatkowski, Cordelia Langford, John Sillitoe on behalf of the Wellcome Sanger Institute COVID-19 Surveillance Team |
| EPI_ISL_2725550 | Azienda Sanitaria dell'Alto Adige - Laboratorio Aziendale di Microbiologia e Virologia | Azienda Sanitaria dell'Alto Adige | Irene Bianconi |
| EPI_ISL_2728627 | SYNLAB | GIGA Medical Genomics | Keith Durkin, Maria Artesi, Bouchra Boujemla, Nathalie Renotte, Nadine Cambisano, Sébastien Bontems, Cécile Meex, Claire Gourzonès, Olivier Ek, Laurent Gillet, Marie-Pierre Hayette, Vincent Bours |
| EPI_ISL_2731148, EPI_ISL_2731149, EPI_ISL_2731150, EPI_ISL_2731151, EPI_ISL_2731152, EPI_ISL_2731153, EPI_ISL_2731155, EPI_ISL_2731156, EPI_ISL_2731157, EPI_ISL_2731158, EPI_ISL_2731159, EPI_ISL_2731160, EPI_ISL_2731161, EPI_ISL_2731162, EPI_ISL_2731163, EPI_ISL_2731164, EPI_ISL_2731165, EPI_ISL_2731168, EPI_ISL_2731170, EPI_ISL_2731171, EPI_ISL_2731172, EPI_ISL_2731173, EPI_ISL_2731174, EPI_ISL_2731175, EPI_ISL_2731176, EPI_ISL_2731177, EPI_ISL_2731179, EPI_ISL_2731180, EPI_ISL_2731181, EPI_ISL_2731182, EPI_ISL_2731183, EPI_ISL_2731185, EPI_ISL_2731186, EPI_ISL_2731189, EPI_ISL_2731190, EPI_ISL_2731191, EPI_ISL_2731192, EPI_ISL_2731194, EPI_ISL_2731195, EPI_ISL_2731196, EPI_ISL_2731197, EPI_ISL_2731198, EPI_ISL_2731199, EPI_ISL_2731200, EPI_ISL_2731201, EPI_ISL_2731202, EPI_ISL_2731203, EPI_ISL_2731205, EPI_ISL_2731207, EPI_ISL_2731208, EPI_ISL_2731209, EPI_ISL_2731210, EPI_ISL_2731211, EPI_ISL_2731212, EPI_ISL_2731213, EPI_ISL_2731214, EPI_ISL_2731215, EPI_ISL_2731216, EPI_ISL_2731217, EPI_ISL_2731219, EPI_ISL_2731220, EPI_ISL_2731221, EPI_ISL_2731222, EPI_ISL_2731224, EPI_ISL_2731225, EPI_ISL_2731226, EPI_ISL_2731227, EPI_ISL_2731228, EPI_ISL_2731229, EPI_ISL_2731231, EPI_ISL_2731232, EPI_ISL_2731233, EPI_ISL_2731235, EPI_ISL_2731236, EPI_ISL_2731237, EPI_ISL_2731238, EPI_ISL_2731240, EPI_ISL_2731241, EPI_ISL_2731242, EPI_ISL_2731243, EPI_ISL_2731245, EPI_ISL_2731246, EPI_ISL_2731247, EPI_ISL_2731248, EPI_ISL_2731250, EPI_ISL_2731251, EPI_ISL_2731252, EPI_ISL_2731254, EPI_ISL_2731255, EPI_ISL_2731256, EPI_ISL_2731261, EPI_ISL_2731262, EPI_ISL_2731263, EPI_ISL_2731264, EPI_ISL_2731265, EPI_ISL_2731266, EPI_ISL_2731267, EPI_ISL_2731268, EPI_ISL_2731269 | | | |
| see above | Lighthouse Lab in Glasgow | Wellcome Sanger Institute for the COVID-19 Genomics UK (COG-UK) Consortium | Harper VanSteenhouse, Yumi Kasai, David Gray, Carol Clugston, Anna Dominiczak and Alex Alderton, Roberto Amato, Jeffrey Barrett, Sonia Goncalves, Ewan Harrison, David K. Jackson, Ian Johnston, Dominic Kwiatkowski, Cordelia Langford, John Sillitoe on behalf of the Wellcome Sanger Institute COVID-19 Surveillance Team |
| EPI_ISL_2735701, EPI_ISL_2735706 | Oxford Viromics, NDM, University of Oxford; Oxford University Hospitals; Basingstoke and North Hampshire Hospital | COVID-19 Genomics UK (COG-UK) Consortium | Tanya Golubchik, David Bonsall, George Macintyre, Amy Trebes, Mariateresa de Cesare, Catrin Moore, Alex Mobbs, Anita Justice, Robert Shaw, Monique Andersson, Timothy Peto, Emma Wise, Nathan Moore, Jessica Lynch, Nick Cortes, Matilde Mori, Stephen Kidd, David Buck, John Todd, Christophe Fraser |
| EPI_ISL_2745817, EPI_ISL_2745851, EPI_ISL_2746004, EPI_ISL_2746007, EPI_ISL_2746023, EPI_ISL_2746026, EPI_ISL_2746045, EPI_ISL_2746046, EPI_ISL_2746060, EPI_ISL_2746066, EPI_ISL_2746070, EPI_ISL_2746080, EPI_ISL_2746099, EPI_ISL_2746406, EPI_ISL_2746532, EPI_ISL_2746543, EPI_ISL_2746616 | | | |
| see above | Respiratory Virus Unit, Microbiology Services Colindale, Public Health England | COVID-19 Genomics UK (COG-UK) Consortium | PHE Covid Sequencing Team |
| EPI_ISL_2750042, EPI_ISL_2750043 | Originating lab: Wales Specialist Virology Centre Sequencing lab: Pathogen Genomics Unit | Public Health Wales Microbiology Cardiff Wales Specialist Virology Centre | Catherine Moore, Johnathan Evans, Laura Gifford, Malorie Perry, Simon Cottrell, Angela Marchbank, Alec Birchley, Alexander Adams, Amy Gaskin, Bree Gatica-Wilcox, Jason Coombes, Joel Southgate, Lauren Gilbert, Lee Graham, Nicole Pacchiarini, Sara Kumziene-Summerhayes, Sarah Taylor, Sophie Jones, Sara Rey, Matthew Bull, Joanne Watkins, Sally Corden, Tom Connor |
| EPI_ISL_2754296 | Arizona State Public Health Laboratory | Arizona State Public Health Laboratory | Trung Huynh, Jessica Escobar, Katherine Fullerton, Nobuko Fukushima, Matthew Contursi, Stacy White, Linda Getsinger, Victor Waddell |
| EPI_ISL_2757667, EPI_ISL_2757669, EPI_ISL_2757670 | Hospital | National Reference Center for Viruses of Respiratory Infections, Institut Pasteur, Paris | Marion Barbet, Sylvie Behillil, Méline Bizard, Angela Brisebarre, Camille Capel, Vincent Enouf, Louise Lefrançois, Frédéric Lemoine, Christophe Malabat, Corinne Maufrais, Etienne Simon-Lorière, Maud Vanpeene, Sylvie Van der Werf ,JéRôMe Durivault |
| EPI_ISL_2757817 | OSPEDALE CIVILE TERAMO - CENTRO TRASFUSIONALE | Istituto Zooprofilattico Sperimentale dell'Abruzzo e Molise "G. Caporale" | Lorusso A, Marcacci M, Di Domenico M, Ancora M, Curini V, Di Lollo Valeria, Mangone I, Rinaldi A, Delli Compagni E, Scialabba S, Caporale M, Di Pasquale A, Cammà C, Puglia I, Calistri P, Savini G |
| EPI_ISL_2758029, EPI_ISL_2758032, EPI_ISL_2758034, EPI_ISL_2758035, EPI_ISL_2758036 | Austrian Agency for Health and Food Safety (AGES) | Bergthaler laboratory, CeMM Research Center for Molecular Medicine of the Austrian Academy of Sciences | Lukas Endler, Anna Schedl, Fabian Amman, Petr Triska, Matthew Thornton, Thomas Penz, Benedikt Agerer, Maelle Le Moing, Michael Schuster, Bekir Erguner, Jan Laine, Martin Senekowitsch, Christoph Bock, Andreas Bergthaler |
| EPI_ISL_2759829 | Bioscientia Labor Wermsdorf | Robert Koch Institute | unknown |
| EPI_ISL_2759999 | SYNLAB MVZ Weiden | Robert Koch Institute | unknown |
| EPI_ISL_2761070, EPI_ISL_2761093, EPI_ISL_2761292, EPI_ISL_2761400, EPI_ISL_2761413, EPI_ISL_2761460, EPI_ISL_2761477 | Bioscientia Labor Wermsdorf | Robert Koch Institute | unknown |
| EPI_ISL_2761654 | Landesgesundheitsamt Baden-Württemberg | Robert Koch Institute | unknown |
| EPI_ISL_2761673, EPI_ISL_2761682 | MVZ Labor Dr. Fenner und Kollegen (Standort Hamburg) | Robert Koch Institute | unknown |
| EPI_ISL_2761692 | Labor Dr. Heidrich & Kollegen MVZ GmbH Hamburg | Robert Koch Institute | unknown |
| EPI_ISL_2761846 | CENTOGENE Frankfurt Laboratory: Niederlassung Industriepark Höchst | Robert Koch Institute | unknown |
| EPI_ISL_2761909 | Medizinisch-Diagnostisches Labor Kempten allgäulab | Robert Koch Institute | unknown |
| EPI_ISL_2762082 | Bioscientia Labor Wermsdorf | Robert Koch Institute | unknown |
| EPI_ISL_2768037 | Clinical Virology | Clinical Bacteriology | Tim Roloff, Fanny Wegner, Helena MB Seth-Smith, Alfredo Mari, Karoline Leuzinger, Julia Bielicki, Manuel Battegay, Hans Hirsch, Adrian Egli |
| EPI_ISL_2774386, EPI_ISL_2774388, EPI_ISL_2774390 | Illinois Department of Public Health - Springfield Lab | Illinois Department of Public Health - Springfield Lab | Bryan Sim, Gordon McCall |

### supp. Table S5: Epsilon

We gratefully acknowledge the following Authors from the Originating laboratories responsible for obtaining the specimens, as well as the Submitting laboratories where the genome data were generated and shared via GISAID, on which this research is based.

All Submitters of data may be contacted directly via www.gisaid.org

Authors are sorted alphabetically.

| Accession ID | Originating Laboratory | Submitting Laboratory | Authors |
|---|---|---|---|
| EPI_ISL_1993258 | Fulgent Genetics | Centers for Disease Control and Prevention Division of Viral Diseases, Pathogen Discovery | Dakota Howard, Dhwani Batra, Peter W. Cook, Kara Moser, Adrian Paskey, Jason Caravas, Benjamin Rambo-Martin, Shatavia Morrison, Christopher Gulvick, Scott Sammons, Yvette Unoarumhi, Darlene Wagner, Matthew Schmerer, Harry Gao, Mickey Li, John Gao, Joseph Fierro, Benafsh Sapra, Becky Tsai, Yan Meng, Doreen Ng, James Xie, Clinton R. Paden, Duncan MacCannell |
| EPI_ISL_2018101 | Wyoming Public Health Laboratory | Wyoming Public Health Laboratory | Jim Mildenberger, Taylor Fearing, Channing Weber, Ashley Norberg, Chayse Rowley, Marley Goetz, Brian Dominguez, Elliot Thomasson, Brittany Oher, Sam Britz, Cari Sloma, and Rob Christensen |
| EPI_ISL_2037398 | Kansas Health and Environmental Lab | Kansas Health and Environmental Lab | Mike Grose, Jonathan Barnell, Ben Olsen, and Phil Adam |
| EPI_ISL_2043725 | Fulgent Genetics | Centers for Disease Control and Prevention Division of Viral Diseases, Pathogen Discovery | Dakota Howard, Dhwani Batra, Peter W. Cook, Kara Moser, Adrian Paskey, Jason Caravas, Benjamin Rambo-Martin, Shatavia Morrison, Christopher Gulvick, Scott Sammons, Yvette Unoarumhi, Darlene Wagner, Matthew Schmerer, Harry Gao, Mickey Li, John Gao, Joseph Fierro, Benafsh Sapra, Becky Tsai, Yan Meng, Doreen Ng, James Xie, Clinton R. Paden, Duncan MacCannell |
| EPI_ISL_2046753, EPI_ISL_2046773 | Laboratory Corporation of America | Centers for Disease Control and Prevention Division of Viral Diseases, Pathogen Discovery | Dakota Howard, Dhwani Batra, Peter W. Cook, Kara Moser, Adrian Paskey, Jason Caravas, Benjamin Rambo-Martin, Shatavia Morrison, Christopher Gulvick, Scott Sammons, Yvette Unoarumhi, Darlene Wagner, Matthew Schmerer, Minoo Agarwal, Eyad Almasri, Debbie Boles, Ayla Burns, Nuthawin Charoensri, Oren Cohen, Susan Countryman, Mary Ann Cristobal, Bobbi Croy, Suzanne Dale, Hrushikesh Deshmukh, Amanda Douglas, Vincent Drouillon, Marcia Eisenberg, Howard Engler, Rama Ghatti, Prashant Gupta, Susan Hicks, Jake Humphrey, Lax Iyer, Manoj Jain, Mohan Kolli, Brian Krueger, Tim Kuphal, Stanley Letovsky, Michael Levandoski, Craig Lukasik, Jonathan Meltzer, Brian Norvell, Mindy Nye, Scott Parker, Christos Petropoulos, John Pruitt, Steven Ragan, Scott Ryan, Mike Sapeta, Jana Schroth, Suresh Babu Selvaraju, Goran Stevovic, Amanda Suchanek, Andrea Throop, Lyndon Tilson, Thomas Urban, Joe Voshell, Kimberly Wagner, Jonathan Williams, Mary Williamson, Qian Zeng, Tricia Zwiefelhofer, Clinton R. Paden, Duncan MacCannell |
| EPI_ISL_2090645 | Idaho Bureau of Laboratories | Idaho Bureau of Laboratories | R. Beukelman, Matthew Charles Burns, Aimee Ceniseros, Robert L. Voermans, Christopher Ball |
| EPI_ISL_2090896 | Nebraska Public Health Laboratory | NPHL COVID-19 Response Team | NPHL COVID-19 Response Team |
| EPI_ISL_2096131 | Washington State Department of Health Public Health Laboratories | Washington State Department of Health Public Health Laboratories | Drew MacKellar, Philip Dykema, Denny Russell, Joenice Gonzalez, Hannah Gray, Geoff Melly, Vanessa De Los Santos, Darren Lucas, JohnAric Peterson, Avi Singh, Rebecca Cao |
| EPI_ISL_2103887, EPI_ISL_2103888, EPI_ISL_2103893, EPI_ISL_2104402 | Broad Institute Clinical Research Sequencing Platform | Infectious Disease Program, Broad Institute of Harvard and MIT | Siddle,K.J., Adams,G., Pearlman,L., Gladden-Young,A., Vicente,G., Blumenstiel,B., DeFelice,M., Lee,M., McGovern,S., Lagerborg,K., Rudy,M., DeRuff,K., Carter,A., Normandin,E., Bauer,M., Reilly,S., Tomkins-Tinch,C., Loreth,C., Chaluvadi,S., Meldrim,J., Granger,B., Lemieux,J.E., Birren,B.W., Sabeti,P.C., Larkin,K., Dodge,S., Lennon,N., Madoff,L., Brown,C., Gallagher,G., Smole,S., Park,D.J., Gabriel,S., and MacInnis,B.L. |
| EPI_ISL_2104891, EPI_ISL_2104953, EPI_ISL_2104994, EPI_ISL_2105012, EPI_ISL_2105016, EPI_ISL_2105042, EPI_ISL_2105093 | UW Virology Lab | UW Virology Lab | Pavitra Roychoudhury, Hong Xie, Lasata Shrestha, Tien V. Nguyen, Shah Mohamed Bakhash, Michelle Lin, Noah R. Baker, Ricardo Perez, Sean Ellis, Nathan Breit, Robert J. Livingston, Meei-Li Huang, Keith R Jerome, Patrick Mathias, Alexander Greninger |
| EPI_ISL_2106441 | Pandemic Response Lab - NYC | Pandemic Response Lab, R&D | Henry Lee, Michael Hammerling, Melissa Hopkins, Cybill del Castillo, Shinyoung Clair Kang, William Ward, Pradeep Bugga, Sol Rey, Dylan Law, Katharine Nelson, Haiping Hao, Jon Laurent |
| EPI_ISL_2134018, EPI_ISL_2134111, EPI_ISL_2134183, EPI_ISL_2134327, EPI_ISL_2134372, EPI_ISL_2134432 | Fulgent Genetics | Centers for Disease Control and Prevention Division of Viral Diseases, Pathogen Discovery | Dakota Howard, Dhwani Batra, Peter W. Cook, Kara Moser, Adrian Paskey, Jason Caravas, Benjamin Rambo-Martin, Shatavia Morrison, Christopher Gulvick, Scott Sammons, Yvette Unoarumhi, Darlene Wagner, Matthew Schmerer, Harry Gao, Mickey Li, John Gao, Joseph Fierro, Benafsh Sapra, Becky Tsai, Yan Meng, Doreen Ng, James Xie, Clinton R. Paden, Duncan MacCannell |
| EPI_ISL_2135835 | Arizona State Public Health Laboratory | Arizona State Public Health Laboratory | Trung Huynh, Jessica Escobar, Katherine Fullerton, Nobuko Fukushima, Stacy White, Linda Getsinger, Victor Waddell |
| EPI_ISL_2136449 | Alaska State Virology Laboratory | Alaska State Virology Laboratory | Stephanie DeRonde, Elva House, Jacob Zidek, Lisa Smith, Ph.D., Jack Chen, Ph.D. |
| EPI_ISL_2138873, EPI_ISL_2138907, EPI_ISL_2139642 | Oregon State Public Health Laboratory | Oregon State Public Health Laboratory | Rafia Razzaque, Eugene Yeboah, Vanda Makris, Laura Tsaknaridis, John Fontana and Shane Sevey |
| EPI_ISL_2140161 | Idaho Bureau of Laboratories | Idaho Bureau of Laboratories | R. Beukelman, Matthew Charles Burns, Aimee Ceniseros, Robert L. Voermans, Christopher Ball |
| EPI_ISL_2146105, EPI_ISL_2146116, EPI_ISL_2146207, EPI_ISL_2146226, EPI_ISL_2146291, EPI_ISL_2146338, EPI_ISL_2146410, EPI_ISL_2146501, EPI_ISL_2146558, EPI_ISL_2146572, EPI_ISL_2146578, EPI_ISL_2146658, EPI_ISL_2146799, EPI_ISL_2146857, EPI_ISL_2146956, EPI_ISL_2147097, EPI_ISL_2147215, EPI_ISL_2147413, EPI_ISL_2147414, EPI_ISL_2147415, EPI_ISL_2147416, EPI_ISL_2147518, EPI_ISL_2147628, EPI_ISL_2147934, EPI_ISL_2147958, EPI_ISL_2147986, EPI_ISL_2148114, EPI_ISL_2148241, EPI_ISL_2148927, EPI_ISL_2148941, EPI_ISL_2148964, EPI_ISL_2148965, EPI_ISL_2148969, EPI_ISL_2149161, EPI_ISL_2149176, EPI_ISL_2149179 | | | |
| see above | Aegis Sciences Corporation | Centers for Disease Control and Prevention Division of Viral Diseases, Pathogen Discovery | Dakota Howard, Dhwani Batra, Peter W. Cook, Kara Moser, Adrian Paskey, Jason Caravas, Benjamin Rambo-Martin, Shatavia Morrison, Christopher Gulvick, Scott Sammons, Yvette Unoarumhi, Darlene Wagner, Matthew Schmerer, Cyndi Clark, Patrick Campbell, Rob Case, Vikramsinha Ghorpade, Holly Houdeshell, Ola Kvalvaag, Dillon Nall, Ethan Sanders, Alec Vest, Shaun Westlund, Matthew Hardison, Clinton R. Paden, Duncan MacCannell |
| EPI_ISL_2152865 | Lighthouse Lab in Milton Keynes | Wellcome Sanger Institute for the COVID-19 Genomics UK (COG-UK) Consortium | The Lighthouse Lab in Milton Keynes and Alex Alderton, Roberto Amato, Jeffrey Barrett, Sonia Goncalves, Ewan Harrison, David K. Jackson, Ian Johnston, Dominic Kwiatkowski, Cordelia Langford, John Sillitoe on behalf of the Wellcome Sanger Institute COVID-19 Surveillance Team |
| EPI_ISL_2153101 | Wyoming Public Health Laboratory | Wyoming Public Health Laboratory | Jim Mildenberger, Taylor Fearing, Channing Weber, Ashley Norberg, Chayse Rowley, Marley Goetz, Brian Dominguez, Elliot Thomasson, Brittany Oher, Sam Britz, Cari Sloma, and Rob Christensen |
| EPI_ISL_2158836 | Servicio Virosis Respiratorias-Departamento Virología-INEI | Instituto Nacional Enfermedades Infecciosas C.G.Malbran | Baumeister E., Avaro M., Benedetti E., Russo M., Dattero ME, Pontoriero A., Cisterna D., Molina V., Perandones C., Tuduri E., Lorenzo F., Poklepovich T., Campos J. |
| EPI_ISL_2158869, EPI_ISL_2158901, EPI_ISL_2158953, EPI_ISL_2158954, EPI_ISL_2158956, EPI_ISL_2158960, EPI_ISL_2159062, EPI_ISL_2159131, EPI_ISL_2159155, EPI_ISL_2159193, EPI_ISL_2159360, EPI_ISL_2159371 | | | |
| see above | Aegis Sciences Corporation | Centers for Disease Control and Prevention Division of Viral Diseases, Pathogen Discovery | Dakota Howard, Dhwani Batra, Peter W. Cook, Kara Moser, Adrian Paskey, Jason Caravas, Benjamin Rambo-Martin, Shatavia Morrison, Christopher Gulvick, Scott Sammons, Yvette Unoarumhi, Darlene Wagner, Matthew Schmerer, Cyndi Clark, Patrick Campbell, Rob Case, Vikramsinha Ghorpade, Holly Houdeshell, Ola Kvalvaag, Dillon Nall, Ethan Sanders, Alec Vest, Shaun Westlund, Matthew Hardison, Clinton R. Paden, Duncan MacCannell |
| EPI_ISL_2160981, EPI_ISL_2161009 | Fulgent Genetics | Centers for Disease Control and Prevention Division of Viral Diseases, Pathogen Discovery | Dakota Howard, Dhwani Batra, Peter W. Cook, Kara Moser, Adrian Paskey, Jason Caravas, Benjamin Rambo-Martin, Shatavia Morrison, Christopher Gulvick, Scott Sammons, Yvette Unoarumhi, Darlene Wagner, Matthew Schmerer, Harry Gao, Mickey Li, John Gao, Joseph Fierro, Benafsh Sapra, Becky Tsai, Yan Meng, Doreen Ng, James Xie, Clinton R. Paden, Duncan MacCannell |
| EPI_ISL_2161821, EPI_ISL_2161824, EPI_ISL_2161825, EPI_ISL_2161828, EPI_ISL_2161830, EPI_ISL_2161864, EPI_ISL_2161868, EPI_ISL_2161870, EPI_ISL_2161877, EPI_ISL_2161884, EPI_ISL_2161889, EPI_ISL_2161890, EPI_ISL_2161898, EPI_ISL_2161899, EPI_ISL_2161933, EPI_ISL_2161946, EPI_ISL_2161948, EPI_ISL_2161987 | | | |
| see above | UW Virology Lab | UW Virology Lab | Pavitra Roychoudhury, Hong Xie, Lasata Shrestha, Tien V. Nguyen, Shah Mohamed Bakhash, Michelle Lin, Noah R. Baker, Ricardo Perez, Sean Ellis, Nathan Breit, Robert J. Livingston, Meei-Li Huang, Keith R Jerome, Patrick Mathias, Alexander Greninger |
| EPI_ISL_2180605, EPI_ISL_2180615, EPI_ISL_2180625, EPI_ISL_2180687, EPI_ISL_2180777, EPI_ISL_2180903, EPI_ISL_2180904, EPI_ISL_2180936, EPI_ISL_2181140, EPI_ISL_2181355, EPI_ISL_2181369, EPI_ISL_2181427, EPI_ISL_2181454, EPI_ISL_2181468, EPI_ISL_2181484, EPI_ISL_2181546, EPI_ISL_2181585, | | | |

| | | | |
|---|---|---|---|
| EPI_ISL_2181587 | | | |
| see above | Aegis Sciences Corporation | Centers for Disease Control and Prevention Division of Viral Diseases, Pathogen Discovery | Dakota Howard, Dhwani Batra, Peter W. Cook, Kara Moser, Adrian Paskey, Jason Caravas, Benjamin Rambo-Martin, Shatavia Morrison, Christopher Gulvick, Scott Sammons, Yvette Unoarumhi, Darlene Wagner, Matthew Schmerer, Cyndi Clark, Patrick Campbell, Rob Case, Vikramsinha Ghorpade, Holly Houdeshell, Ola Kvalvaag, Dillon Nall, Ethan Sanders, Alec Vest, Shaun Westlund, Matthew Hardison, Clinton R. Paden, Duncan MacCannell |
| EPI_ISL_2182205, EPI_ISL_2182468, EPI_ISL_2182815, EPI_ISL_2182825, EPI_ISL_2182861, EPI_ISL_2182899, EPI_ISL_2183979, EPI_ISL_2184230, EPI_ISL_2184253, EPI_ISL_2184943, EPI_ISL_2185081, EPI_ISL_2185202, EPI_ISL_2185268, EPI_ISL_2185465, EPI_ISL_2185516 | | | |
| see above | Laboratory Corporation of America | Centers for Disease Control and Prevention Division of Viral Diseases, Pathogen Discovery | Dakota Howard, Dhwani Batra, Peter W. Cook, Kara Moser, Adrian Paskey, Jason Caravas, Benjamin Rambo-Martin, Shatavia Morrison, Christopher Gulvick, Scott Sammons, Yvette Unoarumhi, Darlene Wagner, Matthew Schmerer, Minoo Agarwal, Eyad Almasri, Debbie Boles, Ayla Burns, Nuthawin Charoensri, Oren Cohen, Susan Countryman, Mary Ann Cristobal, Bobbi Croy, Suzanne Dale, Hrushikesh Deshmukh, Amanda Douglas, Vincent Drouillon, Marcia Eisenberg, Howard Engler, Rama Ghatti, Prashant Gupta, Susan Hicks, Jake Humphrey, Lax Iyer, Manoj Jain, Mohan Kolli, Brian Krueger, Tim Kuphal, Stanley Letovsky, Michael Levandoski, Craig Lukasik, Jonathan Meltzer, Brian Norvell, Mindy Nye, Scott Parker, Christos Petropoulos, John Pruitt, Steven Ragan, Scott Ryan, Mike Sapeta, Jana Schroth, Suresh Babu Selvaraju, Goran Stevovic, Amanda Suchanek, Andrea Throop, Lyndon Tilson, Thomas Urban, Joe Voshell, Kimberly Wagner, Jonathan Williams, Mary Williamson, Qian Zeng, Tricia Zwiefelhofer, Clinton R. Paden, Duncan MacCannell |
| EPI_ISL_2186559, EPI_ISL_2186706 | Infinity Biologix | Centers for Disease Control and Prevention Division of Viral Diseases, Pathogen Discovery | Dakota Howard, Dhwani Batra, Peter W. Cook, Kara Moser, Adrian Paskey, Jason Caravas, Benjamin Rambo-Martin, Shatavia Morrison, Christopher Gulvick, Scott Sammons, Yvette Unoarumhi, Darlene Wagner, Matthew Schmerer, Christian Bixby, Yihe Wang, Jonathan Schultz, Chirayu Goswami, Russ Hager, Robin Grimwood, Clinton R. Paden, Duncan MacCannell |
| EPI_ISL_2187526, EPI_ISL_2187595 | Aegis Sciences Corporation | Centers for Disease Control and Prevention Division of Viral Diseases, Pathogen Discovery | Dakota Howard, Dhwani Batra, Peter W. Cook, Kara Moser, Adrian Paskey, Jason Caravas, Benjamin Rambo-Martin, Shatavia Morrison, Christopher Gulvick, Scott Sammons, Yvette Unoarumhi, Darlene Wagner, Matthew Schmerer, Cyndi Clark, Patrick Campbell, Rob Case, Vikramsinha Ghorpade, Holly Houdeshell, Ola Kvalvaag, Dillon Nall, Ethan Sanders, Alec Vest, Shaun Westlund, Matthew Hardison, Clinton R. Paden, Duncan MacCannell |
| EPI_ISL_2187634 | Helix/Illumina | Centers for Disease Control and Prevention Division of Viral Diseases, Pathogen Discovery | Dakota Howard, Dhwani Batra, Peter W. Cook, Kara Moser, Adrian Paskey, Jason Caravas, Benjamin Rambo-Martin, Shatavia Morrison, Christopher Gulvick, Scott Sammons, Yvette Unoarumhi, Darlene Wagner, Matthew Schmerer, Eileen de Feo, Jan Antico, Christine Tran, Matthew Tolentino, Shannon Wickline, Kim Gietzen, Brad Sickler, Jingtao Liu, Eric Allen, Phil Febbo, Nicole L. Washington, Simon White, Geraint Levan, Kelly Schiabor Barrett, Elizabeth Cirulli, Alexandre Bolze, Ary Ascencio, Charlotte Rivera-Garcia, Ryan Cho, Jason Nguyen, Sherry Wang, Jimmy Ramirez, Tyler Cassens, Efren Sandoval, Magnus Isaksson, William Lee, David Becker, Marc Laurent, James Lu, Clinton R. Paden, Duncan MacCannell |
| EPI_ISL_2192845 | Broad Institute Clinical Research Sequencing Platform | Infectious Disease Program, Broad Institute of Harvard and MIT | Siddle,K.J., Adams,G., Pearlman,L., Gladden-Young,A., Vicente,G., Blumenstiel,B., DeFelice,M., Lee,M., McGovern,S., Lagerborg,K., Rudy,M., DeRuff,K., Carter,A., Normandin,E., Bauer,M., Reilly,S., Tomkins-Tinch,C., Loreth,C., Chaluvadi,S., Meldrim,J., Granger,B., Lemieux,J.E., Birren,B.W., Sabeti,P.C., Larkin,K., Dodge,S., Lennon,N., Madoff,L., Brown,C., Gallagher,G., Smole,S., Park,D.J., Gabriel,S., and MacInnis,B.L. |
| EPI_ISL_2194040, EPI_ISL_2194043, EPI_ISL_2194064, EPI_ISL_2194066, EPI_ISL_2194073, EPI_ISL_2194077, EPI_ISL_2194096, EPI_ISL_2194107, EPI_ISL_2194124, EPI_ISL_2194145, EPI_ISL_2194155, EPI_ISL_2194164, EPI_ISL_2194166, EPI_ISL_2194168, EPI_ISL_2194169, EPI_ISL_2194181 | | | |
| see above | UW Virology Lab | UW Virology Lab | Pavitra Roychoudhury, Hong Xie, Lasata Shrestha, Tien V. Nguyen, Shah Mohamed Bakhash, Michelle Lin, Noah R. Baker, Ricardo Perez, Sean Ellis, Nathan Breit, Robert J. Livingston, Meei-Li Huang, Keith R Jerome, Patrick Mathias, Alexander Greninger |
| EPI_ISL_2203056, EPI_ISL_2203080, EPI_ISL_2203085 | Oregon State Public Health Laboratory | Oregon State Public Health Laboratory | Rafia Razzaque, Eugene Yeboah, Vanda Makris, Laura Tsaknaridis, John Fontana and Shane Sevey |
| EPI_ISL_2203812, EPI_ISL_2203833 | Quest Diagnostics Incorporated | Centers for Disease Control and Prevention Division of Viral Diseases, Pathogen Discovery | Dakota Howard, Dhwani Batra, Peter W. Cook, Kara Moser, Adrian Paskey, Jason Caravas, Benjamin Rambo-Martin, Shatavia Morrison, Christopher Gulvick, Scott Sammons, Yvette Unoarumhi, Darlene Wagner, Matthew Schmerer, S. H. Rosenthal, A. Gerasimova, R. M. Kagan, B. Anderson, M. Hua, Y. Liu, L.E. Bernstein, K.E. Livingston, A. Perez, I. A. Shlyakhter, R. V. Rolando, R. Owen, P. Tanpaiboon, F. Lacbawan, Clinton R. Paden, Duncan MacCannell |
| EPI_ISL_2204946 | WVU Rapid Development Lab | WVU and Marshall University Combined Genomics Core Facilities | James Denvir, Peter Stoilov, Peter Perrotta, Wesley Kimble, Ryan Percifield |
| EPI_ISL_2211077 | Nevada State Public Health Laboratory | Nevada State Public Health Laboratory | Andrew Gorzalski, Mark Pandori |
| EPI_ISL_2225571 | University of Wisconsin-Madison AIDS Vaccine Research Laboratories | University of Wisconsin-Madison AIDS Vaccine Research Laboratories | Gage Moreno, Katarina Braun, et al. AIDS Vaccine Research Laboratories |
| EPI_ISL_2226828 | Wyoming Public Health Laboratory | Wyoming Public Health Laboratory | Jim Mildenberger, Wanda Manley, Noah Hull, Taylor Fearing, Lynette Gumbleton, Channing Weber, Ashley Norberg, Chayse Rowley, Marley Goetz, Brian Dominguez, Elliot Thomasson, Sam Britz, Cari Sloma, and Rob Christensen |
| EPI_ISL_2229285 | "NM Dept. Health, Scientific Laboratory Division " | Centers for Disease Control and Prevention Division of Viral Diseases, Pathogen Discovery | Mili Sheth, Sarah Nobles, Jasmine Padilla, Mark Burroughs, Shoshona Le, Katie Dillon, Peter Cook, Clinton R. Paden, Dhwani Batra, Krista Queen, Kristen Knipe, Dakota Howard, Yvette Unoarumhi, Darlene Wagner, Matthew Schmerer, Ben L. Rambo-Martin, Kristine Lacek, Sam Shepard, Alison Laufer Halpin, Dave Wentworth, Vivien Dugan, Suxiang Tong, Justin Lee |
| EPI_ISL_2229291, EPI_ISL_2229292 | MT Public Health Laboratory | Centers for Disease Control and Prevention Division of Viral Diseases, Pathogen Discovery | Mili Sheth, Sarah Nobles, Jasmine Padilla, Mark Burroughs, Shoshona Le, Katie Dillon, Peter Cook, Clinton R. Paden, Dhwani Batra, Krista Queen, Kristen Knipe, Dakota Howard, Yvette Unoarumhi, Darlene Wagner, Matthew Schmerer, Ben L. Rambo-Martin, Kristine Lacek, Sam Shepard, Alison Laufer Halpin, Dave Wentworth, Vivien Dugan, Suxiang Tong, Justin Lee |
| EPI_ISL_2232562 | Minnesota Department of Health, Public Health Laboratory | Minnesota Department of Health, Public Health Laboratory | Alexandra Lorentz, Jacob Garfin, Matt Plumb, and Xiong Wang |
| EPI_ISL_2233255, EPI_ISL_2233260, EPI_ISL_2233294, EPI_ISL_2233299, EPI_ISL_2233304, EPI_ISL_2233306, EPI_ISL_2233312, EPI_ISL_2233314, EPI_ISL_2233327 | UW Virology Lab | UW Virology Lab | Pavitra Roychoudhury, Hong Xie, Lasata Shrestha, Tien V. Nguyen, Shah Mohamed Bakhash, Michelle Lin, Noah R. Baker, Sean Ellis, Meei-Li Huang, Keith R Jerome, Alexander Greninger |
| EPI_ISL_2234923, EPI_ISL_2235044 | Pandemic Response Lab - NYC | Pandemic Response Lab, R&D | Henry Lee, Michael Hammerling, Melissa Hopkins, Cybill del Castillo, Shinyoung Clair Kang, William Ward, Pradeep Bugga, Sol Rey, Dylan Law, Katharine Nelson, Haiping Hao, Jon Laurent |
| EPI_ISL_2242509 | IN State Department of Health Laboratory Services | IN State Department of Health Laboratory Services | Cassandra Campion, Jamie Yeadon, Brian Pope, Lixia Liu, Kyle Brownlee, Melissa Hindenlang, Mark Glazier |
| EPI_ISL_2244949 | M Health Fairview | Minnesota Department of Health, Public Health Laboratory | Alexandra Lorentz, Jacob Garfin, Matt Plumb, and Xiong Wang |
| EPI_ISL_2245492, EPI_ISL_2245591, EPI_ISL_2245592 | Maine Health and Environmental Testing Laboratory | Tewhey Lab, The Jackson Laboratory | Matluk,N., Dewey,H., Iosue,F., Barter,M., Lynch,R., Munger,H. and Tewhey,R. |
| EPI_ISL_2247067, EPI_ISL_2247157, EPI_ISL_2247179, EPI_ISL_2247191 | Quest Diagnostics Incorporated | Centers for Disease Control and Prevention Division of Viral Diseases, Pathogen Discovery | Dakota Howard, Dhwani Batra, Peter W. Cook, Kara Moser, Adrian Paskey, Jason Caravas, Benjamin Rambo-Martin, Shatavia Morrison, Christopher Gulvick, Scott Sammons, Yvette Unoarumhi, Darlene Wagner, Matthew Schmerer, S. H. Rosenthal, A. Gerasimova, R. M. Kagan, B. Anderson, M. Hua, Y. Liu, L.E. Bernstein, K.E. Livingston, A. Perez, I. A. Shlyakhter, R. V. Rolando, R. Owen, P. Tanpaiboon, F. Lacbawan, Clinton R. Paden, Duncan MacCannell |
| EPI_ISL_2247419, EPI_ISL_2247488, EPI_ISL_2247514, EPI_ISL_2247607, EPI_ISL_2248020, EPI_ISL_2248451 | Helix/Illumina | Centers for Disease Control and Prevention Division of Viral Diseases, Pathogen Discovery | Dakota Howard, Dhwani Batra, Peter W. Cook, Kara Moser, Adrian Paskey, Jason Caravas, Benjamin Rambo-Martin, Shatavia Morrison, Christopher Gulvick, Scott Sammons, Yvette Unoarumhi, Darlene Wagner, Matthew Schmerer, Eileen de Feo, Jan Antico, Christine Tran, Matthew Tolentino, Shannon Wickline, Kim Gietzen, Brad Sickler, Jingtao Liu, Eric Allen, Phil Febbo, Nicole L. Washington, Simon White, Geraint Levan, Kelly Schiabor Barrett, Elizabeth Cirulli, Alexandre Bolze, Ary Ascencio, Charlotte Rivera-Garcia, Ryan Cho, Jason Nguyen, Sherry Wang, Jimmy Ramirez, Tyler Cassens, Efren Sandoval, Magnus Isaksson, William Lee, David Becker, Marc Laurent, James Lu, Clinton R. Paden, Duncan MacCannell |
| EPI_ISL_2248553, EPI_ISL_2248555, EPI_ISL_2248572, EPI_ISL_2248607, EPI_ISL_2248637, EPI_ISL_2248672 | Quest Diagnostics Incorporated | Centers for Disease Control and Prevention Division of Viral Diseases, Pathogen Discovery | Dakota Howard, Dhwani Batra, Peter W. Cook, Kara Moser, Adrian Paskey, Jason Caravas, Benjamin Rambo-Martin, Shatavia Morrison, Christopher Gulvick, Scott Sammons, Yvette Unoarumhi, Darlene Wagner, Matthew Schmerer, S. H. Rosenthal, A. Gerasimova, R. M. Kagan, B. Anderson, M. Hua, Y. Liu, L.E. Bernstein, K.E. Livingston, A. Perez, I. A. Shlyakhter, R. V. Rolando, R. Owen, P. Tanpaiboon, F. Lacbawan, Clinton R. Paden, Duncan MacCannell |

| | | | |
|---|---|---|---|
| EPI_ISL_2249244, EPI_ISL_2249245 | Hospital Margarita Maza de Juárez | Microbial Genomics Laboratory | Bruno Gomez-Gil, Julissa Enciso-Ibarra, Alejandra Garcia-Gasca, Daniel Fregoso-Rueda |
| EPI_ISL_2249569 | M Health Fairview | Minnesota Department of Health, Public Health Laboratory | Alexandra Lorentz, Jacob Garfin, Matt Plumb, and Xiong Wang |
| EPI_ISL_2250089, EPI_ISL_2250091 | Arizona State University | Arizona State University | Peter T. Skidmore, LaRinda A. Holland, Matthew F. Smith, Rabia Maqsood, Nicholas J. Mellor, Joy M. Blain, Valerie Harris, Joshua LaBaer, Vel Murugan, Efrem S. Lim |
| EPI_ISL_2250158, EPI_ISL_2250176 | Oregon State Public Health Laboratory | Oregon State Public Health Laboratory | Rafia Razzaque, Eugene Yeboah, Vanda Makris, Laura Tsaknaridis, John Fontana and Shane Sevey |
| EPI_ISL_2263490 | Arizona State Public Health Laboratory | Arizona State Public Health Laboratory | Trung Huynh, Jessica Escobar, Katherine Fullerton, Nobuko Fukushima, Stacy White, Linda Getsinger, Victor Waddell |
| EPI_ISL_2268345 | Helix/Illumina | Centers for Disease Control and Prevention Division of Viral Diseases, Pathogen Discovery | Dakota Howard, Dhwani Batra, Peter W. Cook, Kara Moser, Adrian Paskey, Jason Caravas, Benjamin Rambo-Martin, Shatavia Morrison, Christopher Gulvick, Scott Sammons, Yvette Unoarumhi, Darlene Wagner, Matthew Schmerer, Eileen de Feo, Jan Antico, Christine Tran, Matthew Tolentino, Shannon Wickline, Kim Gietzen, Brad Sickler, Jingtao Liu, Eric Allen, Phil Febbo, Nicole L. Washington, Simon White, Geraint Levan, Kelly Schiabor Barrett, Elizabeth Cirulli, Alexandre Bolze, Ary Ascencio, Charlotte Rivera-Garcia, Ryan Cho, Jason Nguyen, Sherry Wang, Jimmy Ramirez, Tyler Cassens, Efren Sandoval, Magnus Isaksson, William Lee, David Becker, Marc Laurent, James Lu, Clinton R. Paden, Duncan MacCannell |
| EPI_ISL_2268397, EPI_ISL_2268485, EPI_ISL_2268690, EPI_ISL_2268751, EPI_ISL_2268958, EPI_ISL_2269031, EPI_ISL_2269179, EPI_ISL_2269201 | Quest Diagnostics Incorporated | Centers for Disease Control and Prevention Division of Viral Diseases, Pathogen Discovery | Dakota Howard, Dhwani Batra, Peter W. Cook, Kara Moser, Adrian Paskey, Jason Caravas, Benjamin Rambo-Martin, Shatavia Morrison, Christopher Gulvick, Scott Sammons, Yvette Unoarumhi, Darlene Wagner, Matthew Schmerer, S. H. Rosenthal, A. Gerasimova, R. M. Kagan, B. Anderson, M. Hua, Y. Liu, L.E. Bernstein, K.E. Livingston, A. Perez, I. A. Shlyakhter, R. V. Rolando, R. Owen, P. Tanpaiboon, F. Lacbawan, Clinton R. Paden, Duncan MacCannell |
| EPI_ISL_2269269, EPI_ISL_2269316, EPI_ISL_2269518, EPI_ISL_2269561, EPI_ISL_2269610 | Fulgent Genetics | Centers for Disease Control and Prevention Division of Viral Diseases, Pathogen Discovery | Dakota Howard, Dhwani Batra, Peter W. Cook, Kara Moser, Adrian Paskey, Jason Caravas, Benjamin Rambo-Martin, Shatavia Morrison, Christopher Gulvick, Scott Sammons, Yvette Unoarumhi, Darlene Wagner, Matthew Schmerer, Harry Gao, Mickey Li, John Gao, Joseph Fierro, Benafsh Sapra, Becky Tsai, Yan Meng, Doreen Ng, James Xie, Clinton R. Paden, Duncan MacCannell |
| EPI_ISL_2269886, EPI_ISL_2269916, EPI_ISL_2269987, EPI_ISL_2270044, EPI_ISL_2270164, EPI_ISL_2270183, EPI_ISL_2270233, EPI_ISL_2270275, EPI_ISL_2270298, EPI_ISL_2270319, EPI_ISL_2270337, EPI_ISL_2270390, EPI_ISL_2270432 | | | |
| see above | Helix/Illumina | Centers for Disease Control and Prevention Division of Viral Diseases, Pathogen Discovery | Dakota Howard, Dhwani Batra, Peter W. Cook, Kara Moser, Adrian Paskey, Jason Caravas, Benjamin Rambo-Martin, Shatavia Morrison, Christopher Gulvick, Scott Sammons, Yvette Unoarumhi, Darlene Wagner, Matthew Schmerer, Eileen de Feo, Jan Antico, Christine Tran, Matthew Tolentino, Shannon Wickline, Kim Gietzen, Brad Sickler, Jingtao Liu, Eric Allen, Phil Febbo, Nicole L. Washington, Simon White, Geraint Levan, Kelly Schiabor Barrett, Elizabeth Cirulli, Alexandre Bolze, Ary Ascencio, Charlotte Rivera-Garcia, Ryan Cho, Jason Nguyen, Sherry Wang, Jimmy Ramirez, Tyler Cassens, Efren Sandoval, Magnus Isaksson, William Lee, David Becker, Marc Laurent, James Lu, Clinton R. Paden, Duncan MacCannell |
| EPI_ISL_2271693, EPI_ISL_2271700, EPI_ISL_2271702 | Laboratorio Central, Ministerio de Salud Cordoba | Instituto de Patologia Vegetal (CIAP-INTA) on behalf of 'Proyecto Argentino Interinstitucional de genomica de SARS-CoV-2' (PAIS Consortium) | Fernandez, FD; Marquez, N.; Debat, HJ.; M; Re, V.; Pisano, M.B.; Castro, G.; Barbas, G. |
| EPI_ISL_2272499, EPI_ISL_2272500 | LANIIA | LANIIA | Cristobal Chaidez, Bruno Gomez-Gil, Jean Pierre González-Gómez, Juan Daniel Lira-Morales, Irvin González-López, Julissa Enciso-Ibarra, Célida Isabel Martínez-Rodríguez |
| EPI_ISL_2272521 | Servicio Diagnóstico Clínico | LANIIA | Cristobal Chaidez, Bruno Gomez-Gil, Jean Pierre González-Gómez, Juan Daniel Lira-Morales, Irvin González-López, Julissa Enciso-Ibarra, Célida Isabel Martínez-Rodríguez |
| EPI_ISL_2272545, EPI_ISL_2272546 | Alaska State Virology Laboratory | Alaska State Virology Laboratory | Stephanie DeRonde, Elva House, Jacob Zidek, Lisa Smith, Ph.D., Jack Chen, Ph.D. |
| EPI_ISL_2272610, EPI_ISL_2272619, EPI_ISL_2272645, EPI_ISL_2272646, EPI_ISL_2273780, EPI_ISL_2273791, EPI_ISL_2273793, EPI_ISL_2273824, EPI_ISL_2273828 | UW Virology Lab | UW Virology Lab | Pavitra Roychoudhury, Hong Xie, Lasata Shrestha, Tien V. Nguyen, Shah Mohamed Bakhash, Michelle Lin, Noah R. Baker, Ricardo Perez, Sean Ellis, Nathan Breit, Robert J. Livingston, Meei-Li Huang, Keith R Jerome, Patrick Mathias, Alexander Greninger |
| EPI_ISL_2280335, EPI_ISL_2280384, EPI_ISL_2280429, EPI_ISL_2280596, EPI_ISL_2280792, EPI_ISL_2280824, EPI_ISL_2280915, EPI_ISL_2281096, EPI_ISL_2281118, EPI_ISL_2281120, EPI_ISL_2281129, EPI_ISL_2281131, EPI_ISL_2281172 | | | |
| see above | Aegis Sciences Corporation | Centers for Disease Control and Prevention Division of Viral Diseases, Pathogen Discovery | Dakota Howard, Dhwani Batra, Peter W. Cook, Kara Moser, Adrian Paskey, Jason Caravas, Benjamin Rambo-Martin, Shatavia Morrison, Christopher Gulvick, Scott Sammons, Yvette Unoarumhi, Darlene Wagner, Matthew Schmerer, Cyndi Clark, Patrick Campbell, Rob Case, Vikramsinha Ghorpade, Holly Houdeshell, Ola Kvalvaag, Dillon Nall, Ethan Sanders, Alec Vest, Shaun Westlund, Matthew Hardison, Clinton R. Paden, Duncan MacCannell |
| EPI_ISL_2282336, EPI_ISL_2282431, EPI_ISL_2282476, EPI_ISL_2282487, EPI_ISL_2282539, EPI_ISL_2282546, EPI_ISL_2282598, EPI_ISL_2282642, EPI_ISL_2282681, EPI_ISL_2282821, EPI_ISL_2282858, EPI_ISL_2282864 | | | |
| see above | Infinity Biologix | Centers for Disease Control and Prevention Division of Viral Diseases, Pathogen Discovery | Dakota Howard, Dhwani Batra, Peter W. Cook, Kara Moser, Adrian Paskey, Jason Caravas, Benjamin Rambo-Martin, Shatavia Morrison, Christopher Gulvick, Scott Sammons, Yvette Unoarumhi, Darlene Wagner, Matthew Schmerer, Christian Bixby, Yihe Wang, Jonathan Schultz, Chirayu Goswami, Russ Hager, Robin Grimwood, Clinton R. Paden, Duncan MacCannell |
| EPI_ISL_2285099 | Mako Medical | Centers for Disease Control and Prevention Division of Viral Diseases, Pathogen Discovery | Dakota Howard, Dhwani Batra, Peter W. Cook, Kara Moser, Adrian Paskey, Jason Caravas, Benjamin Rambo-Martin, Shatavia Morrison, Christopher Gulvick, Scott Sammons, Yvette Unoarumhi, Darlene Wagner, Matthew Schmerer,Matthew Tugwell, Lauren Moon, Clinton R. Paden, Duncan MacCannell |
| EPI_ISL_2287853 | New Mexico Department of Health Scientific Laboratory | New Mexico Department of Health Scientific Laboratory | Ellie Johnson, Anastacia Griego-Fisher, D'eldra Malone, Jennifer Benoit |
| EPI_ISL_2291653, EPI_ISL_2292777, EPI_ISL_2292834, EPI_ISL_2292846, EPI_ISL_2292878, EPI_ISL_2292917 | Utah Public Health Laboratory | Utah Public Health Laboratory | Erin L. Young, Kelly F. Oakeson, Tara Gallagher |
| EPI_ISL_2293023, EPI_ISL_2293024 | LANIIA | LANIIA | Cristobal Chaidez, Bruno Gomez-Gil, Jean Pierre González-Gómez, Juan Daniel Lira-Morales, Irvin González-López, Julissa Enciso-Ibarra, Célida Isabel Martínez-Rodríguez |
| EPI_ISL_2296103 | LESP Sinaloa | Instituto de Diagnostico y Referencia Epidemiologicos (INDRE) | Claudia Wong-Arambula, Abril Rodriguez-Maldonado, Vanessa Rivero-Arredondo, Ariadna Medina-Benitez, Joaquin Quiroz-Mercado, Sergio Rangel-Guerrero, Natividad Cruz-Ortiz, Tatiana Nunez-Garcia, Gisela Barrera-Badillo, Lucia Hernandez-Rivas, Irma Lopez-Martinez, Ernesto Ramirez-Gonzalez. |
| EPI_ISL_2296104 | LESP Baja California | Instituto de Diagnostico y Referencia Epidemiologicos (INDRE) | Claudia Wong-Arambula, Abril Rodriguez-Maldonado, Vanessa Rivero-Arredondo, Ariadna Medina-Benitez, Joaquin Quiroz-Mercado, Sergio Rangel-Guerrero, Natividad Cruz-Ortiz, Tatiana Nunez-Garcia, Gisela Barrera-Badillo, Lucia Hernandez-Rivas, Irma Lopez-Martinez, Ernesto Ramirez-Gonzalez. |
| EPI_ISL_2296512 | Centracare Laboratory Services | Minnesota Department of Health, Public Health Laboratory | Alexandra Lorentz, Jacob Garfin, Matt Plumb, and Xiong Wang |
| EPI_ISL_2299306, EPI_ISL_2299368, EPI_ISL_2299402, EPI_ISL_2299495, EPI_ISL_2300671, EPI_ISL_2300679, EPI_ISL_2300692, EPI_ISL_2300693, EPI_ISL_2300694, EPI_ISL_2300701 | UW Virology Lab | UW Virology Lab | Pavitra Roychoudhury, Hong Xie, Lasata Shrestha, Tien V. Nguyen, Shah Mohamed Bakhash, Michelle Lin, Noah R. Baker, Sean Ellis, Meei-Li Huang, Keith R Jerome, Alexander Greninger |
| EPI_ISL_2304505, EPI_ISL_2304518, EPI_ISL_2304565, EPI_ISL_2304618, EPI_ISL_2304623 | Quest Diagnostics Incorporated | Centers for Disease Control and Prevention Division of Viral Diseases, Pathogen Discovery | Dakota Howard, Dhwani Batra, Peter W. Cook, Kara Moser, Adrian Paskey, Jason Caravas, Benjamin Rambo-Martin, Shatavia Morrison, Christopher Gulvick, Scott Sammons, Yvette Unoarumhi, Darlene Wagner, Matthew Schmerer, S. H. Rosenthal, A. Gerasimova, R. M. Kagan, B. Anderson, M. Hua, Y. Liu, L.E. Bernstein, K.E. Livingston, A. Perez, I. A. Shlyakhter, R. V. Rolando, R. Owen, P. Tanpaiboon, F. Lacbawan, Clinton R. Paden, Duncan MacCannell |
| EPI_ISL_2304961, EPI_ISL_2305141, EPI_ISL_2305384, EPI_ISL_2306197, EPI_ISL_2306351, EPI_ISL_2306352, EPI_ISL_2306353, EPI_ISL_2306537, EPI_ISL_2306549, EPI_ISL_2306665, EPI_ISL_2306672 | | | |
| see above | Laboratory Corporation of America | Centers for Disease Control and Prevention Division of Viral Diseases, Pathogen Discovery | Dakota Howard, Dhwani Batra, Peter W. Cook, Kara Moser, Adrian Paskey, Jason Caravas, Benjamin Rambo-Martin, Shatavia Morrison, Christopher Gulvick, Scott Sammons, Yvette Unoarumhi, Darlene Wagner, Matthew Schmerer, Minoo Agarwal, Eyad Almasri, Debbie Boles, Ayla Burns, Nuthawin Charoensri, Oren Cohen, Susan Countryman, Mary Ann Cristobal, Bobbi Croy, Suzanne Dale, Hrushikesh Deshmukh, Amanda Douglas, Vincent Drouillon, |

| | | | |
|---|---|---|---|
| | | | Marcia Eisenberg, Howard Engler, Rama Ghatti, Prashant Gupta, Susan Hicks, Jake Humphrey, Lax Iyer, Manoj Jain, Mohan Kolli, Brian Krueger, Tim Kuphal, Stanley Letovsky, Michael Levandoski, Craig Lukasik, Jonathan Meltzer, Brian Norvell, Mindy Nye, Scott Parker, Christos Petropoulos, John Pruitt, Steven Ragan, Scott Ryan, Mike Sapeta, Jana Schroth, Suresh Babu Selvaraju, Goran Stevovic, Amanda Suchanek, Andrea Throop, Lyndon Tilson, Thomas Urban, Joe Voshell, Kimberly Wagner, Jonathan Williams, Mary Williamson, Qian Zeng, Tricia Zwiefelhofer, Clinton R. Paden, Duncan MacCannell |
| EPI_ISL_2307199, EPI_ISL_2307243, EPI_ISL_2307286, EPI_ISL_2307304 | Mako Medical | Centers for Disease Control and Prevention Division of Viral Diseases, Pathogen Discovery | Dakota Howard, Dhwani Batra, Peter W. Cook, Kara Moser, Adrian Paskey, Jason Caravas, Benjamin Rambo-Martin, Shatavia Morrison, Christopher Gulvick, Scott Sammons, Yvette Unoarumhi, Darlene Wagner, Matthew Schmerer,Matthew Tugwell, Lauren Moon, Clinton R. Paden, Duncan MacCannell |
| EPI_ISL_2308343 | Montana Public Health Laboratory | Montana Public Health Laboratory | Joy Ritter, Michelle Mozer, Carrie Biskupiak, Deborah Gibson, Michael Dills |
| EPI_ISL_2308503 | Kansas Health and Environmental Lab | Kansas Health and Environmental Lab | Mike Grose, Katherine Wiggins, Jonathan Barnell, Ben Olsen, and Phil Adam |
| EPI_ISL_2308682 | Clinical Reference Laboratory | Kansas Health and Environmental Lab | Mike Grose, Katherine Wiggins, Jonathan Barnell, Ben Olsen, and Phil Adam |
| EPI_ISL_2309988, EPI_ISL_2309989, EPI_ISL_2310134, EPI_ISL_2310168 | Colorado Department of Public Health and Environment | Colorado Department of Public Health and Environment | Laura Bankers, Molly C. Hetherington-Rauth, Diana Ir, Alexandria Rossheim, Shannon R. Matzinger, Sarah Elizabeth Totten, Emily A. Travanty |
| EPI_ISL_2319353 | INDRE | Instituto Nacional de Medicina Genomica | Hidalgo-Miranda A, Mendoza-Vargas A, Reyes-Grajeda JP, Cedro-Tanda A, Gisela Barrera-Badillo, Irma Lopez-Martinez, Jose Ernesto Ramirez González, Gonzalez-Barrera D, Rangel-DeLeon D, Munguia-Garza P, Garcia-Cardenas FJ, Gonzalez-Woge MA, Herrera-Montalvo LA. |
| EPI_ISL_2320134, EPI_ISL_2320220, EPI_ISL_2320226, EPI_ISL_2320314, EPI_ISL_2320748, EPI_ISL_2320774, EPI_ISL_2320779, EPI_ISL_2320864, EPI_ISL_2320978, EPI_ISL_2321019, EPI_ISL_2321112 | | | |
| see above | Helix/Illumina | Centers for Disease Control and Prevention Division of Viral Diseases, Pathogen Discovery | Dakota Howard, Dhwani Batra, Peter W. Cook, Kara Moser, Adrian Paskey, Jason Caravas, Benjamin Rambo-Martin, Shatavia Morrison, Christopher Gulvick, Scott Sammons, Yvette Unoarumhi, Darlene Wagner, Matthew Schmerer, Eileen de Feo, Jan Antico, Christine Tran, Matthew Tolentino, Shannon Wickline, Kim Gietzen, Brad Sickler, Jingtao Liu, Eric Allen, Phil Febbo, Nicole L. Washington, Simon White, Geraint Levan, Kelly Schiabor Barrett, Elizabeth Cirulli, Alexandre Bolze, Ary Ascencio, Charlotte Rivera-Garcia, Ryan Cho, Jason Nguyen, Sherry Wang, Jimmy Ramirez, Tyler Cassens, Efren Sandoval, Magnus Isaksson, William Lee, David Becker, Marc Laurent, James Lu, Clinton R. Paden, Duncan MacCannell |
| EPI_ISL_2321319, EPI_ISL_2321366, EPI_ISL_2321379 | Aegis Sciences Corporation | Centers for Disease Control and Prevention Division of Viral Diseases, Pathogen Discovery | Dakota Howard, Dhwani Batra, Peter W. Cook, Kara Moser, Adrian Paskey, Jason Caravas, Benjamin Rambo-Martin, Shatavia Morrison, Christopher Gulvick, Scott Sammons, Yvette Unoarumhi, Darlene Wagner, Matthew Schmerer, Cyndi Clark, Patrick Campbell, Rob Case, Vikramsinha Ghorpade, Holly Houdeshell, Ola Kvalvaag, Dillon Nall, Ethan Sanders, Alec Vest, Shaun Westlund, Matthew Hardison, Clinton R. Paden, Duncan MacCannell |
| EPI_ISL_2322813, EPI_ISL_2322814, EPI_ISL_2322815, EPI_ISL_2322816, EPI_ISL_2322817, EPI_ISL_2322818 | New Mexico Department of Health Scientific Laboratory | New Mexico Department of Health Scientific Laboratory | Ellie Johnson, Anastacia Griego-Fisher, D'eldra Malone, Jennifer Benoit |
| EPI_ISL_2323735, EPI_ISL_2323755, EPI_ISL_2323910, EPI_ISL_2323939, EPI_ISL_2324053, EPI_ISL_2324081, EPI_ISL_2324082 | Infinity Biologix | Centers for Disease Control and Prevention Division of Viral Diseases, Pathogen Discovery | Dakota Howard, Dhwani Batra, Peter W. Cook, Kara Moser, Adrian Paskey, Jason Caravas, Benjamin Rambo-Martin, Shatavia Morrison, Christopher Gulvick, Scott Sammons, Yvette Unoarumhi, Darlene Wagner, Matthew Schmerer, Christian Bixby, Yihe Wang, Jonathan Schultz, Chirayu Goswami, Russ Hager, Robin Grimwood, Clinton R. Paden, Duncan MacCannell |
| EPI_ISL_2324480, EPI_ISL_2324527, EPI_ISL_2324550 | Laboratory Corporation of America | Centers for Disease Control and Prevention Division of Viral Diseases, Pathogen Discovery | Dakota Howard, Dhwani Batra, Peter W. Cook, Kara Moser, Adrian Paskey, Jason Caravas, Benjamin Rambo-Martin, Shatavia Morrison, Christopher Gulvick, Scott Sammons, Yvette Unoarumhi, Darlene Wagner, Matthew Schmerer, Minoo Agarwal, Eyad Almasri, Debbie Boles, Ayla Burns, Nuthawin Charoensri, Oren Cohen, Susan Countryman, Mary Ann Cristobal, Bobbi Croy, Suzanne Dale, Hrushikesh Deshmukh, Amanda Douglas, Vincent Drouillon, Marcia Eisenberg, Howard Engler, Rama Ghatti, Prashant Gupta, Susan Hicks, Jake Humphrey, Lax Iyer, Lisa Pfefferle, Manoj Jain, Matthew Robinson, Mohan Kolli, Brian Krueger, Tim Kuphal, Stanley Letovsky, Michael Levandoski, Craig Lukasik, Jonathan Meltzer, Brian Norvell, Mindy Nye, Scott Parker, Christos Petropoulos, John Pruitt, Steven Ragan, Scott Ryan, Mike Sapeta, Jana Schroth, Suresh Babu Selvaraju, Goran Stevovic, Amanda Suchanek, Andrea Throop, Lyndon Tilson, Thomas Urban, Joe Voshell, Kimberly Wagner, Jonathan Williams, Mary Williamson, Qian Zeng, Tricia Zwiefelhofer, Clinton R. Paden, Duncan MacCannell |
| EPI_ISL_2325081 | Minnesota Department of Health, Public Health Laboratory | Minnesota Department of Health, Public Health Laboratory | Alexandra Lorentz, Jacob Garfin, Matt Plumb, and Xiong Wang |
| EPI_ISL_2329444 | Sharp HealthCare Laboratory | Andersen lab at Scripps Research | SEARCH Alliance San Diego with Aaron Harding, Jacquelyn Berumen, Cathy Woerle, Liam McGinnis, Art Mendoza, Omid Bakhtar |
| EPI_ISL_2332703 | Centracare Laboratory Services | Minnesota Department of Health, Public Health Laboratory | Alexandra Lorentz, Jacob Garfin, Matt Plumb, and Xiong Wang |
| EPI_ISL_2332863, EPI_ISL_2332882, EPI_ISL_2332892, EPI_ISL_2332895, EPI_ISL_2332910, EPI_ISL_2332912, EPI_ISL_2332927, EPI_ISL_2332930, EPI_ISL_2332935, EPI_ISL_2332957, EPI_ISL_2332958, EPI_ISL_2332996, EPI_ISL_2332999, EPI_ISL_2333015, EPI_ISL_2333058 | | | |
| see above | UW Virology Lab | UW Virology Lab | Pavitra Roychoudhury, Hong Xie, Lasata Shrestha, Tien V. Nguyen, Shah Mohamed Bakhash, Michelle Lin, Noah R. Baker, Ricardo Perez, Sean Ellis, Nathan Breit, Robert J. Livingston, Meei-Li Huang, Keith R Jerome, Patrick Mathias, Alexander Greninger |
| EPI_ISL_2335092, EPI_ISL_2335094 | University of Michigan Clinical Microbiology Laboratory | Lauring Lab, University of Michigan, Department of Microbiology and Immunology | Gilbert |
| EPI_ISL_2336195, EPI_ISL_2336198, EPI_ISL_2336219, EPI_ISL_2336221 | Washington State Department of Health Public Health Laboratories | Washington State Department of Health Public Health Laboratories | Drew MacKellar, Philip Dykema, Denny Russell, Joenice Gonzalez, Hannah Gray, Geoff Melly, Vanessa De Los Santos, Darren Lucas, JohnAric Peterson, Avi Singh, Rebecca Cao |
| EPI_ISL_2339333, EPI_ISL_2339341 | Oregon State Public Health Laboratory | Oregon State Public Health Laboratory | Rafia Razzaque, Eugene Yeboah, Vanda Makris, Laura Tsaknaridis, John Fontana and Shane Sevey |
| EPI_ISL_2339395 | Illinois Department of Public Health - Springfield Lab | Illinois Department of Public Health - Springfield Lab | Bryan Sim, Gordon McCall |
| EPI_ISL_2339997, EPI_ISL_2340022 | North Dakota Department of Health, Public Health Laboratory | North Dakota Department of Health, Public Health Laboratory | Lisa Wingerter |
| EPI_ISL_2340691, EPI_ISL_2340704, EPI_ISL_2340714, EPI_ISL_2340724, EPI_ISL_2340732, EPI_ISL_2340734, EPI_ISL_2340740, EPI_ISL_2340742, EPI_ISL_2340749, EPI_ISL_2340752, EPI_ISL_2340783, EPI_ISL_2340784, EPI_ISL_2340791, EPI_ISL_2340795, EPI_ISL_2340796 | | | |
| see above | UW Virology Lab | UW Virology Lab | Pavitra Roychoudhury, Hong Xie, Lasata Shrestha, Tien V. Nguyen, Shah Mohamed Bakhash, Michelle Lin, Noah R. Baker, Sean Ellis, Meei-Li Huang, Keith R Jerome, Alexander Greninger |
| EPI_ISL_2341035 | Salud Digna | Instituto Nacional de Medicina Genomica | Hidalgo-Miranda A, Cedro-Tanda A, Cisneros-Villanueva M, Orjuela-Rodríguez M, Mendoza-Vargas A, Reyes-Grajeda JP, Abraham Campos-Romero, Moreno-Camacho José Luis, Rodriguez-Gallegos Jorge, Luna-Ruiz Marco, Gonzalez-Barrera D, Rangel-DeLeon D, Munguia-Garza P, Ramirez-Vega O, Escobar-Arrazola, M, Herrera-Montalvo LA. |
| EPI_ISL_2344511 | Virginia Division of Consolidated Laboratory Services | Virginia Division of Consolidated Laboratory Services | Virginia DCLS |
| EPI_ISL_2348932, EPI_ISL_2348934, EPI_ISL_2348935 | Oregon State Public Health Laboratory | Oregon State Public Health Laboratory | Rafia Razzaque, Eugene Yeboah, Vanda Makris, Laura Tsaknaridis, John Fontana and Shane Sevey |
| EPI_ISL_2349901, EPI_ISL_2349923, EPI_ISL_2349993 | Salud Digna | Instituto Nacional de Medicina Genomica | Hidalgo-Miranda A, Cedro-Tanda A, Mendoza-Vargas A, Reyes-Grajeda JP, Abraham Campos-Romero, Moreno-Camacho José Luis, Rodriguez-Gallegos Jorge, Luna-Ruiz Marco, Gonzalez-Barrera D, Rangel-DeLeon D, Munguia-Garza P, Ramirez-Vega O, Escobar-Arrazola, M, Herrera-Montalvo LA. |
| EPI_ISL_2367807, EPI_ISL_2367809, EPI_ISL_2367903, EPI_ISL_2367909, EPI_ISL_2367934 | Quest Diagnostics Incorporated | Centers for Disease Control and Prevention Division of Viral Diseases, Pathogen Discovery | Dakota Howard, Dhwani Batra, Peter W. Cook, Kara Moser, Adrian Paskey, Jason Caravas, Benjamin Rambo-Martin, Shatavia Morrison, Christopher Gulvick, Scott Sammons, Yvette Unoarumhi, Darlene Wagner, Matthew Schmerer, S. H. Rosenthal, A. Gerasimova, R. M. Kagan, B. Anderson, M. Hua, Y. Liu, L.E. Bernstein, K.E. Livingston, A. Perez, I. A. Shlyakhter, R. V. Rolando, R. Owen, P. Tanpaiboon, F. Lacbawan, Clinton R. Paden, Duncan MacCannell |
| EPI_ISL_2368136, EPI_ISL_2368200, EPI_ISL_2368201 | Fulgent Genetics | Centers for Disease Control and Prevention Division of Viral Diseases, Pathogen Discovery | Dakota Howard, Dhwani Batra, Peter W. Cook, Kara Moser, Adrian Paskey, Jason Caravas, Benjamin Rambo-Martin, Shatavia Morrison, Christopher Gulvick, Scott Sammons, Yvette Unoarumhi, Darlene Wagner, Matthew Schmerer, Harry Gao, Mickey Li, John Gao, Joseph Fierro, Benafsh Sapra, Becky Tsai, Yan Meng, Doreen Ng, James Xie, Clinton R. Paden, Duncan MacCannell |
| EPI_ISL_2368275, EPI_ISL_2368400, EPI_ISL_2368409, EPI_ISL_2368699 | Aegis Sciences Corporation | Centers for Disease Control and Prevention Division of Viral Diseases, Pathogen Discovery | Dakota Howard, Dhwani Batra, Peter W. Cook, Kara Moser, Adrian Paskey, Jason Caravas, Benjamin Rambo-Martin, Shatavia Morrison, Christopher Gulvick, Scott Sammons, Yvette Unoarumhi, Darlene Wagner, Matthew Schmerer, Cyndi Clark, Patrick Campbell, Rob Case, Vikramsinha Ghorpade, Holly Houdeshell, Ola Kvalvaag, Dillon Nall, Ethan Sanders, Alec Vest, Shaun Westlund, Matthew Hardison, Clinton R. Paden, Duncan MacCannell |

| | | | |
|---|---|---|---|
| EPI_ISL_2369028, EPI_ISL_2369192, EPI_ISL_2369416 | Quest Diagnostics Incorporated | Centers for Disease Control and Prevention Division of Viral Diseases, Pathogen Discovery | Dakota Howard, Dhwani Batra, Peter W. Cook, Kara Moser, Adrian Paskey, Jason Caravas, Benjamin Rambo-Martin, Shatavia Morrison, Christopher Gulvick, Scott Sammons, Yvette Unoarumhi, Darlene Wagner, Matthew Schmerer, S. H. Rosenthal, A. Gerasimova, R. M. Kagan, B. Anderson, M. Hua, Y. Liu, L.E. Bernstein, K.E. Livingston, A. Perez, I. A. Shlyakhter, R. V. Rolando, R. Owen, P. Tanpaiboon, F. Lacbawan, Clinton R. Paden, Duncan MacCannell |
| EPI_ISL_2370054, EPI_ISL_2370117, EPI_ISL_2370530, EPI_ISL_2370536 | Aegis Sciences Corporation | Centers for Disease Control and Prevention Division of Viral Diseases, Pathogen Discovery | Dakota Howard, Dhwani Batra, Peter W. Cook, Kara Moser, Adrian Paskey, Jason Caravas, Benjamin Rambo-Martin, Shatavia Morrison, Christopher Gulvick, Scott Sammons, Yvette Unoarumhi, Darlene Wagner, Matthew Schmerer, Cyndi Clark, Patrick Campbell, Rob Case, Vikramsinha Ghorpade, Holly Houdeshell, Ola Kvalvaag, Dillon Nall, Ethan Sanders, Alec Vest, Shaun Westlund, Matthew Hardison, Clinton R. Paden, Duncan MacCannell |
| EPI_ISL_2371560, EPI_ISL_2371568, EPI_ISL_2371572, EPI_ISL_2371583, EPI_ISL_2371671 | Fulgent Genetics | Centers for Disease Control and Prevention Division of Viral Diseases, Pathogen Discovery | Dakota Howard, Dhwani Batra, Peter W. Cook, Kara Moser, Adrian Paskey, Jason Caravas, Benjamin Rambo-Martin, Shatavia Morrison, Christopher Gulvick, Scott Sammons, Yvette Unoarumhi, Darlene Wagner, Matthew Schmerer, Harry Gao, Mickey Li, John Gao, Joseph Fierro, Benafsh Sapra, Becky Tsai, Yan Meng, Doreen Ng, James Xie, Clinton R. Paden, Duncan MacCannell |
| EPI_ISL_2371748, EPI_ISL_2372141, EPI_ISL_2372158, EPI_ISL_2372182, EPI_ISL_2372228 | Quest Diagnostics Incorporated | Centers for Disease Control and Prevention Division of Viral Diseases, Pathogen Discovery | Dakota Howard, Dhwani Batra, Peter W. Cook, Kara Moser, Adrian Paskey, Jason Caravas, Benjamin Rambo-Martin, Shatavia Morrison, Christopher Gulvick, Scott Sammons, Yvette Unoarumhi, Darlene Wagner, Matthew Schmerer, S. H. Rosenthal, A. Gerasimova, R. M. Kagan, B. Anderson, M. Hua, Y. Liu, L.E. Bernstein, K.E. Livingston, A. Perez, I. A. Shlyakhter, R. V. Rolando, R. Owen, P. Tanpaiboon, F. Lacbawan, Clinton R. Paden, Duncan MacCannell |
| EPI_ISL_2375569, EPI_ISL_2375583, EPI_ISL_2375651, EPI_ISL_2375775 | Infinity Biologix | Centers for Disease Control and Prevention Division of Viral Diseases, Pathogen Discovery | Dakota Howard, Dhwani Batra, Peter W. Cook, Kara Moser, Adrian Paskey, Jason Caravas, Benjamin Rambo-Martin, Shatavia Morrison, Christopher Gulvick, Scott Sammons, Yvette Unoarumhi, Darlene Wagner, Matthew Schmerer, Christian Bixby, Yihe Wang, Jonathan Schultz, Chirayu Goswami, Russ Hager, Robin Grimwood, Clinton R. Paden, Duncan MacCannell |
| EPI_ISL_2376012, EPI_ISL_2376013, EPI_ISL_2376014, EPI_ISL_2376015, EPI_ISL_2376016 | Alaska State Virology Laboratory | Alaska State Virology Laboratory | Stephanie DeRonde, Elva House, Jacob Zidek, Lisa Smith, Ph.D., Jack Chen, Ph.D. |
| EPI_ISL_2376248, EPI_ISL_2376258, EPI_ISL_2376259 | Gundersen Clinical Microbiology Laboratory | Kabara Cancer Research Institute | Craig S. Richmond, Paraic A. Kenny |
| EPI_ISL_2378556, EPI_ISL_2378568, EPI_ISL_2378583, EPI_ISL_2378587, EPI_ISL_2378590, EPI_ISL_2378593 | State Testing Facility | Altius Institute for Biomedical Research | Daniel Bates, Rebecca Bruders, Michael Buckley, Mark Frerker, Amanda Gale, Clem Green, Muhammad Halimun, Kneshay Harper, Matt Hartman, Alex Isner, Audra Johnson, Jessica Kunder, Lauren Mitchell, Jemma Nelson, Alex Nguyen, Sofia Olsson, Sadie Patraw, Tobias Ragoczy, Joshua Richards, Jean Robinston, Jacob Rodriguez, John Stamatoyannopoulos, Eric Thorland, Julia Wald |
| EPI_ISL_2378837, EPI_ISL_2378878, EPI_ISL_2378888 | Washington State Department of Health Public Health Laboratories | Washington State Department of Health Public Health Laboratories | Drew MacKellar, Philip Dykema, Denny Russell, Joenice Gonzalez, Hannah Gray, Geoff Melly, Vanessa De Los Santos, Darren Lucas, JohnAric Peterson, Avi Singh, Rebecca Cao |
| EPI_ISL_2381933, EPI_ISL_2381934, EPI_ISL_2381940 | New Mexico Department of Health Scientific Laboratory | New Mexico Department of Health Scientific Laboratory | Ellie Johnson,D'eldra Malone, Jennifer Benoit, Ratheesh Rajan, Linda Salazar, Keila Gutierrez, Anastacia Griego-Fisher |
| EPI_ISL_2384011 | MT Public Health Laboratory | Centers for Disease Control and Prevention Division of Viral Diseases, Pathogen Discovery | Mili Sheth, Sarah Nobles, Jasmine Padilla, Mark Burroughs, Shoshona Le, Katie Dillon, Peter Cook, Clinton R. Paden, Dhwani Batra, Krista Queen, Kristen Knipe, Dakota Howard, Yvette Unoarumhi, Darlene Wagner, Matthew Schmerer, Ben L. Rambo-Martin, Kristine Lacek, Sam Shepard, Alison Laufer Halpin, Dave Wentworth, Vivien Dugan, Suxiang Tong, Justin Lee |
| EPI_ISL_2388567 | LabKom - Labor Augsburg MVZ GmbH | Robert Koch Institute | unknown |
| EPI_ISL_2391329 | Genetica Molecular and Subdepartamento de Virologia ISP Chile | Instituto de Salud Publica de Chile | Karen Orostica, Constanza Campano, Barbara Parra, Loredana Arata, Gisselle Barra, Patricia Bustos, Rodrigo Fasce, Javier Tognarelli, Andres Castillo, Soledad Ulloa, Jorge Fernandez |
| EPI_ISL_2397427, EPI_ISL_2397608, EPI_ISL_2397718 | Quest Diagnostics Incorporated | Centers for Disease Control and Prevention Division of Viral Diseases, Pathogen Discovery | Dakota Howard, Dhwani Batra, Peter W. Cook, Kara Moser, Adrian Paskey, Jason Caravas, Benjamin Rambo-Martin, Shatavia Morrison, Christopher Gulvick, Scott Sammons, Yvette Unoarumhi, Darlene Wagner, Matthew Schmerer, S. H. Rosenthal, A. Gerasimova, R. M. Kagan, B. Anderson, M. Hua, Y. Liu, L.E. Bernstein, K.E. Livingston, A. Perez, I. A. Shlyakhter, R. V. Rolando, R. Owen, P. Tanpaiboon, F. Lacbawan, Clinton R. Paden, Duncan MacCannell |
| EPI_ISL_2398161 | Laboratory Corporation of America | Centers for Disease Control and Prevention Division of Viral Diseases, Pathogen Discovery | Dakota Howard, Dhwani Batra, Peter W. Cook, Kara Moser, Adrian Paskey, Jason Caravas, Benjamin Rambo-Martin, Shatavia Morrison, Christopher Gulvick, Scott Sammons, Yvette Unoarumhi, Darlene Wagner, Matthew Schmerer, Minoo Agarwal, Eyad Almasri, Debbie Boles, Ayla Burns, Nuthawin Charoensri, Oren Cohen, Susan Countryman, Mary Ann Cristobal, Bobbi Croy, Suzanne Dale, Hrushikesh Deshmukh, Amanda Douglas, Vincent Drouillon, Marcia Eisenberg, Howard Engler, Rama Ghatti, Prashant Gupta, Susan Hicks, Jake Humphrey, Lax Iyer, Lisa Pfefferle, Manoj Jain, Matthew Robinson, Mohan Kolli, Brian Krueger, Tim Kuphal, Stanley Letovsky, Michael Levandoski, Craig Lukasik, Jonathan Meltzer, Brian Norvell, Mindy Nye, Scott Parker, Christos Petropoulos, John Pruitt, Steven Ragan, Scott Ryan, Mike Sapeta, Jana Schroth, Suresh Babu Selvaraju, Goran Stevovic, Amanda Suchanek, Andrea Throop, Lyndon Tilson, Thomas Urban, Joe Voshell, Kimberly Wagner, Jonathan Williams, Mary Williamson, Qian Zeng, Tricia Zwiefelhofer, Clinton R. Paden, Duncan MacCannell |
| EPI_ISL_2403718 | Minnesota Department of Health, Public Health Laboratory | Minnesota Department of Health, Public Health Laboratory | Alexandra Lorentz, Jacob Garfin, Matt Plumb, and Xiong Wang |
| EPI_ISL_2404749 | Oregon State Public Health Laboratory | Oregon State Public Health Laboratory | Rafia Razzaque, Eugene Yeboah, Vanda Makris, Laura Tsaknaridis, John Fontana and Shane Sevey |
| EPI_ISL_2405202 | Pomona Valley Hospital Medical Center | Los Angeles County PHL | P. Hemarajata et al. |
| EPI_ISL_2421151 | URMC LABS | Wadsworth Center, New York State Department of Health | Kirsten St. George, Daryl M. Lamson, Alexis Russell, Matthew Shudt, Melissa A Leisner, Jonathan Plitnick, Catharine Prussing, Navjot Singh, John Kelly, Erasmus Schneider, Erica Lasek-Nesselquist |
| EPI_ISL_2422114 | Aegis Sciences Corporation | Centers for Disease Control and Prevention Division of Viral Diseases, Pathogen Discovery | Dakota Howard, Dhwani Batra, Peter W. Cook, Kara Moser, Adrian Paskey, Jason Caravas, Benjamin Rambo-Martin, Shatavia Morrison, Christopher Gulvick, Scott Sammons, Yvette Unoarumhi, Darlene Wagner, Matthew Schmerer, Cyndi Clark, Patrick Campbell, Rob Case, Vikramsinha Ghorpade, Holly Houdeshell, Ola Kvalvaag, Dillon Nall, Ethan Sanders, Alec Vest, Shaun Westlund, Matthew Hardison, Clinton R. Paden, Duncan MacCannell |
| EPI_ISL_2423015, EPI_ISL_2423016 | San Diego County Public Health Laboratory | Andersen lab at Scripps Research | SEARCH Alliance San Diego with Ashleigh Murphy, Jovan Shephard, Brett Austin |
| EPI_ISL_2423551 | TXDSHS | TXDSHS | Rashmi Tuladhar, Bonnie Oh, Jenny Zhang, Maliha Rahman, Mayela Pedrueza, Anita Pokharel, Karen Bobier, Lorraine Rodriguez, Myong Koag, Chun Wang, Rachel Lee, Grace Kubin |
| EPI_ISL_2424189, EPI_ISL_2424190 | UCLA Clinical Micro Lab | Los Angeles County PHL | P. Hemarajata et al. |
| EPI_ISL_2424239, EPI_ISL_2424240 | Oregon State Public Health Laboratory | Oregon State Public Health Laboratory | Rafia Razzaque, Eugene Yeboah, Vanda Makris, Laura Tsaknaridis, John Fontana and Shane Sevey |
| EPI_ISL_2425402, EPI_ISL_2425403 | M Health Fairview | Minnesota Department of Health, Public Health Laboratory | Alexandra Lorentz, Jacob Garfin, Matt Plumb, and Xiong Wang |
| EPI_ISL_2426294, EPI_ISL_2426305, EPI_ISL_2426308, EPI_ISL_2426380 | UW Virology Lab | UW Virology Lab | Pavitra Roychoudhury, Hong Xie, Lasata Shrestha, Tien V. Nguyen, Shah Mohamed Bakhash, Michelle Lin, Noah R. Baker, Ricardo Perez, Sean Ellis, Nathan Breit, Robert J. Livingston, Meei-Li Huang, Keith R Jerome, Patrick Mathias, Alexander Greninger |
| EPI_ISL_2426416, EPI_ISL_2426455, EPI_ISL_2426516 | UW Virology Lab | UW Virology Lab | Pavitra Roychoudhury, Hong Xie, Lasata Shrestha, Tien V. Nguyen, Shah Mohamed Bakhash, Michelle Lin, Noah R. Baker, Sean Ellis, Meei-Li Huang, Keith R Jerome, Alexander Greninger |
| EPI_ISL_2432618, EPI_ISL_2432695, EPI_ISL_2432734, EPI_ISL_2432763, EPI_ISL_2432790, EPI_ISL_2432819, EPI_ISL_2432824, EPI_ISL_2432832, EPI_ISL_2432833 | UW Virology Lab | UW Virology Lab | Pavitra Roychoudhury, Hong Xie, Lasata Shrestha, Tien V. Nguyen, Shah Mohamed Bakhash, Michelle Lin, Noah R. Baker, Ricardo Perez, Sean Ellis, Nathan Breit, Robert J. Livingston, Meei-Li Huang, Keith R Jerome, Patrick Mathias, Alexander Greninger |
| EPI_ISL_2432845, EPI_ISL_2432861, EPI_ISL_2432865, EPI_ISL_2432871, EPI_ISL_2432895, EPI_ISL_2432905 | UW Virology Lab | UW Virology Lab | Pavitra Roychoudhury, Hong Xie, Lasata Shrestha, Tien V. Nguyen, Shah Mohamed Bakhash, Michelle Lin, Noah R. Baker, Sean Ellis, Meei-Li Huang, Keith R Jerome, Alexander Greninger |

| | | | |
|---|---|---|---|
| EPI_ISL_2439165 | IN State Department of Health Laboratory Services | IN State Department of Health Laboratory Services | Cassandra Campion, Jamie Yeadon, Brian Pope, Lixia Liu, Kyle Brownlee, Melissa Hindenlang, Mark Glazier |
| EPI_ISL_2439415, EPI_ISL_2439559, EPI_ISL_2439616, EPI_ISL_2439773, EPI_ISL_2439793 | Quest Diagnostics Incorporated | Centers for Disease Control and Prevention Division of Viral Diseases, Pathogen Discovery | Dakota Howard, Dhwani Batra, Peter W. Cook, Kara Moser, Adrian Paskey, Jason Caravas, Benjamin Rambo-Martin, Shatavia Morrison, Christopher Gulvick, Scott Sammons, Yvette Unoarumhi, Darlene Wagner, Matthew Schmerer, S. H. Rosenthal, A. Gerasimova, R. M. Kagan, B. Anderson, M. Hua, Y. Liu, L.E. Bernstein, K.E. Livingston, A. Perez, I. A. Shlyakhter, R. V. Rolando, R. Owen, P. Tanpaiboon, F. Lacbawan, Clinton R. Paden, Duncan MacCannell |
| EPI_ISL_2440207 | Helix/Illumina | Centers for Disease Control and Prevention Division of Viral Diseases, Pathogen Discovery | Dakota Howard, Dhwani Batra, Peter W. Cook, Kara Moser, Adrian Paskey, Jason Caravas, Benjamin Rambo-Martin, Shatavia Morrison, Christopher Gulvick, Scott Sammons, Yvette Unoarumhi, Darlene Wagner, Matthew Schmerer, Eileen de Feo, Jan Antico, Christine Tran, Matthew Tolentino, Shannon Wickline, Kim Gietzen, Brad Sickler, Jingtao Liu, Eric Allen, Phil Febbo, Nicole L. Washington, Simon White, Geraint Levan, Kelly Schiabor Barrett, Elizabeth Cirulli, Alexandre Bolze, Ary Ascencio, Charlotte Rivera-Garcia, Ryan Cho, Jason Nguyen, Sherry Wang, Jimmy Ramirez, Tyler Cassens, Efren Sandoval, Magnus Isaksson, William Lee, David Becker, Marc Laurent, James Lu, Clinton R. Paden, Duncan MacCannell |
| EPI_ISL_2440622 | Quest Diagnostics Incorporated | Centers for Disease Control and Prevention Division of Viral Diseases, Pathogen Discovery | Dakota Howard, Dhwani Batra, Peter W. Cook, Kara Moser, Adrian Paskey, Jason Caravas, Benjamin Rambo-Martin, Shatavia Morrison, Christopher Gulvick, Scott Sammons, Yvette Unoarumhi, Darlene Wagner, Matthew Schmerer, S. H. Rosenthal, A. Gerasimova, R. M. Kagan, B. Anderson, M. Hua, Y. Liu, L.E. Bernstein, K.E. Livingston, A. Perez, I. A. Shlyakhter, R. V. Rolando, R. Owen, P. Tanpaiboon, F. Lacbawan, Clinton R. Paden, Duncan MacCannell |
| EPI_ISL_2440901 | Mako Medical | Centers for Disease Control and Prevention Division of Viral Diseases, Pathogen Discovery | Dakota Howard, Dhwani Batra, Peter W. Cook, Kara Moser, Adrian Paskey, Jason Caravas, Benjamin Rambo-Martin, Shatavia Morrison, Christopher Gulvick, Scott Sammons, Yvette Unoarumhi, Darlene Wagner, Matthew Schmerer,Matthew Tugwell, Lauren Moon, Clinton R. Paden, Duncan MacCannell |
| EPI_ISL_2441902, EPI_ISL_2441903 | Fulgent Genetics | Centers for Disease Control and Prevention Division of Viral Diseases, Pathogen Discovery | Dakota Howard, Dhwani Batra, Peter W. Cook, Kara Moser, Adrian Paskey, Jason Caravas, Benjamin Rambo-Martin, Shatavia Morrison, Christopher Gulvick, Scott Sammons, Yvette Unoarumhi, Darlene Wagner, Matthew Schmerer, Harry Gao, Mickey Li, John Gao, Joseph Fierro, Benafsh Sapra, Becky Tsai, Yan Meng, Doreen Ng, James Xie, Clinton R. Paden, Duncan MacCannell |
| EPI_ISL_2443165, EPI_ISL_2443174 | IN State Department of Health Laboratory Services | IN State Department of Health Laboratory Services | Cassandra Campion, Jamie Yeadon, Brian Pope, Lixia Liu, Kyle Brownlee, Melissa Hindenlang, Mark Glazier |
| EPI_ISL_2443292 | TXDSHS | TXDSHS | Rashmi Tuladhar, Bonnie Oh, Jenny Zhang, Maliha Rahman, Mayela Pedrueza, Anita Pokharel, Karen Bobier, Lorraine Rodriguez, Myong Koag, Chun Wang, Rachel Lee, Grace Kubin |
| EPI_ISL_2443854 | Minnesota Department of Health, Public Health Laboratory | Minnesota Department of Health, Public Health Laboratory | Alexandra Lorentz, Jacob Garfin, Matt Plumb, and Xiong Wang |
| EPI_ISL_2445619, EPI_ISL_2445632 | Humboldt County Public Health Laboratory | Humboldt County Public Health Laboratory | Jeremy Corrigan |
| EPI_ISL_2451230 | Mako Medical | Centers for Disease Control and Prevention Division of Viral Diseases, Pathogen Discovery | Dakota Howard, Dhwani Batra, Peter W. Cook, Kara Moser, Adrian Paskey, Jason Caravas, Benjamin Rambo-Martin, Shatavia Morrison, Christopher Gulvick, Scott Sammons, Yvette Unoarumhi, Darlene Wagner, Matthew Schmerer,Matthew Tugwell, Lauren Moon, Clinton R. Paden, Duncan MacCannell |
| EPI_ISL_2451300, EPI_ISL_2451330, EPI_ISL_2451376 | Quest Diagnostics Incorporated | Centers for Disease Control and Prevention Division of Viral Diseases, Pathogen Discovery | Dakota Howard, Dhwani Batra, Peter W. Cook, Kara Moser, Adrian Paskey, Jason Caravas, Benjamin Rambo-Martin, Shatavia Morrison, Christopher Gulvick, Scott Sammons, Yvette Unoarumhi, Darlene Wagner, Matthew Schmerer, S. H. Rosenthal, A. Gerasimova, R. M. Kagan, B. Anderson, M. Hua, Y. Liu, L.E. Bernstein, K.E. Livingston, A. Perez, I. A. Shlyakhter, R. V. Rolando, R. Owen, P. Tanpaiboon, F. Lacbawan, Clinton R. Paden, Duncan MacCannell |
| EPI_ISL_2451592 | NV-Southern Nevada Public Health Laboratory | Centers for Disease Control and Prevention Division of Viral Diseases, Pathogen Discovery | Mili Sheth, Sarah Nobles, Jasmine Padilla, Mark Burroughs, Shoshona Le, Katie Dillon, Peter Cook, Clinton R. Paden, Dhwani Batra, Krista Queen, Kristen Knipe, Dakota Howard, Yvette Unoarumhi, Darlene Wagner, Matthew Schmerer, Ben L. Rambo-Martin, Kristine Lacek, Sam Shepard, Alison Laufer Halpin, Dave Wentworth, Vivien Dugan, Suxiang Tong, Justin Lee |
| EPI_ISL_2451806 | IL Dept. of Public Health Springfield Laboratory | Centers for Disease Control and Prevention Division of Viral Diseases, Pathogen Discovery | Mili Sheth, Sarah Nobles, Jasmine Padilla, Mark Burroughs, Shoshona Le, Katie Dillon, Peter Cook, Clinton R. Paden, Dhwani Batra, Krista Queen, Kristen Knipe, Dakota Howard, Yvette Unoarumhi, Darlene Wagner, Matthew Schmerer, Ben L. Rambo-Martin, Kristine Lacek, Sam Shepard, Alison Laufer Halpin, Dave Wentworth, Vivien Dugan, Suxiang Tong, Justin Lee |
| EPI_ISL_2451903 | ND Dept. of Health Laboratory Services-Microbiology | Centers for Disease Control and Prevention Division of Viral Diseases, Pathogen Discovery | Mili Sheth, Sarah Nobles, Jasmine Padilla, Mark Burroughs, Shoshona Le, Katie Dillon, Peter Cook, Clinton R. Paden, Dhwani Batra, Krista Queen, Kristen Knipe, Dakota Howard, Yvette Unoarumhi, Darlene Wagner, Matthew Schmerer, Ben L. Rambo-Martin, Kristine Lacek, Sam Shepard, Alison Laufer Halpin, Dave Wentworth, Vivien Dugan, Suxiang Tong, Justin Lee |
| EPI_ISL_2451937, EPI_ISL_2451943 | AK State Public Health Lab, State Health Department | Centers for Disease Control and Prevention Division of Viral Diseases, Pathogen Discovery | Mili Sheth, Sarah Nobles, Jasmine Padilla, Mark Burroughs, Shoshona Le, Katie Dillon, Peter Cook, Clinton R. Paden, Dhwani Batra, Krista Queen, Kristen Knipe, Dakota Howard, Yvette Unoarumhi, Darlene Wagner, Matthew Schmerer, Ben L. Rambo-Martin, Kristine Lacek, Sam Shepard, Alison Laufer Halpin, Dave Wentworth, Vivien Dugan, Suxiang Tong, Justin Lee |
| EPI_ISL_2451984, EPI_ISL_2451985 | OR State PHL-Virology/Immunology Section | Centers for Disease Control and Prevention Division of Viral Diseases, Pathogen Discovery | Mili Sheth, Sarah Nobles, Jasmine Padilla, Mark Burroughs, Shoshona Le, Katie Dillon, Peter Cook, Clinton R. Paden, Dhwani Batra, Krista Queen, Kristen Knipe, Dakota Howard, Yvette Unoarumhi, Darlene Wagner, Matthew Schmerer, Ben L. Rambo-Martin, Kristine Lacek, Sam Shepard, Alison Laufer Halpin, Dave Wentworth, Vivien Dugan, Suxiang Tong, Justin Lee |
| EPI_ISL_2452125, EPI_ISL_2452134 | HI Dept. of Health, State Laboratories Division | Centers for Disease Control and Prevention Division of Viral Diseases, Pathogen Discovery | Mili Sheth, Sarah Nobles, Jasmine Padilla, Mark Burroughs, Shoshona Le, Katie Dillon, Peter Cook, Clinton R. Paden, Dhwani Batra, Krista Queen, Kristen Knipe, Dakota Howard, Yvette Unoarumhi, Darlene Wagner, Matthew Schmerer, Ben L. Rambo-Martin, Kristine Lacek, Sam Shepard, Alison Laufer Halpin, Dave Wentworth, Vivien Dugan, Suxiang Tong, Justin Lee |
| EPI_ISL_2454335 | TriCore Reference Laboratories | Center for Global Health, University of New Mexico Health Sciences Center | Daryl Domman, Kurt Schwalm, Valerie Morley, Cecilia Thompson, Kendra Pesko, Karissa Culbreath, Darrell Dinwiddie |
| EPI_ISL_2455437 | Alaska State Virology Laboratory | Alaska State Virology Laboratory | Stephanie DeRonde, Elva House, Jacob Zidek, Lisa Smith, Ph.D., Jack Chen, Ph.D. |
| EPI_ISL_2456111, EPI_ISL_2456114, EPI_ISL_2456149, EPI_ISL_2456592, EPI_ISL_2458494, EPI_ISL_2458530, EPI_ISL_2458542, EPI_ISL_2458551, EPI_ISL_2458555, EPI_ISL_2458589, EPI_ISL_2458596, EPI_ISL_2458598, EPI_ISL_2458599, EPI_ISL_2458617, EPI_ISL_2458620 | | | |
| see above | Utah Public Health Laboratory | Utah Public Health Laboratory | Erin L. Young, Kelly F. Oakeson, Tara Gallagher |
| EPI_ISL_2463209, EPI_ISL_2463213, EPI_ISL_2463229 | Quest Diagnostics Incorporated | Centers for Disease Control and Prevention Division of Viral Diseases, Pathogen Discovery | Dakota Howard, Dhwani Batra, Peter W. Cook, Kara Moser, Adrian Paskey, Jason Caravas, Benjamin Rambo-Martin, Shatavia Morrison, Christopher Gulvick, Scott Sammons, Yvette Unoarumhi, Darlene Wagner, Matthew Schmerer, S. H. Rosenthal, A. Gerasimova, R. M. Kagan, B. Anderson, M. Hua, Y. Liu, L.E. Bernstein, K.E. Livingston, A. Perez, I. A. Shlyakhter, R. V. Rolando, R. Owen, P. Tanpaiboon, F. Lacbawan, Clinton R. Paden, Duncan MacCannell |
| EPI_ISL_2466048, EPI_ISL_2466049 | Alaska State Virology Lab, Alaska State Public Health Labs | Alaska State Virology Lab, Alaska State Public Health Labs | DeRonde,S., House,E., Zidek,J., Redlinger,M., Smith,L., Chen,J. |
| EPI_ISL_2467527 | GA Department of Public Health | GA Department of Public Health | Stacy Reeves, Jonathan Edwards, Cynthia Dixey, Tonia Parrott, Aliyah Fields, Taylor Smith |
| EPI_ISL_2472968, EPI_ISL_2472969, EPI_ISL_2472978, EPI_ISL_2472979, EPI_ISL_2473002, EPI_ISL_2473004, EPI_ISL_2473048, EPI_ISL_2473050, EPI_ISL_2473095, EPI_ISL_2473337, EPI_ISL_2473348, EPI_ISL_2473360, EPI_ISL_2473361, EPI_ISL_2473369, EPI_ISL_2473372, EPI_ISL_2473376, EPI_ISL_2473418, EPI_ISL_2473425, EPI_ISL_2473430, EPI_ISL_2473435, EPI_ISL_2473438, EPI_ISL_2473456, EPI_ISL_2473492, EPI_ISL_2473501, EPI_ISL_2473504, EPI_ISL_2473519, EPI_ISL_2473544, EPI_ISL_2473556, EPI_ISL_2473593, EPI_ISL_2473594, EPI_ISL_2473595, EPI_ISL_2473600, EPI_ISL_2473616, EPI_ISL_2473629, EPI_ISL_2473636, EPI_ISL_2473640, EPI_ISL_2473641, EPI_ISL_2473651, EPI_ISL_2473652 | | | |
| see above | State Testing Facility | Altius Institute for Biomedical Research | Daniel Bates, Rebecca Bruders, Michael Buckley, Mark Frerker, Amanda Gale, Clem Green, Muhammad Halimun, Kneshay Harper, Matt Hartman, Alex Isner, Audra Johnson, Jessica Kunder, Lauren Mitchell, Jemma Nelson, Alex Nguyen, Sofia Olsson, Sadie Patraw, Tobias Ragoczy, Joshua Richards, Jean Robinson, Jacob Rodriguez, John Stamatoyannopoulos, Eric Thorland, Julia Wald |
| EPI_ISL_2473676 | Oregon State Public Health Laboratory | Oregon State Public Health Laboratory | Rafia Razzaque, Eugene Yeboah, Vanda Makris, Laura Tsaknaridis, John Fontana and Shane Sevey |
| EPI_ISL_2476441 | LESP Chihuahua | Instituto de Diagnostico y Referencia Epidemiologicos (INDRE) | Claudia Wong-Arambula, Abril Rodriguez-Maldonado, Vanessa Rivero-Arredondo, Ariadna Medina-Benitez, Joaquin Quiroz-Mercado, Sergio Rangel-Guerrero, Natividad Cruz-Ortiz, Tatiana Nunez-Garcia, Gisela Barrera-Badillo, Lucia Hernandez-Rivas, Irma Lopez-Martinez, Ernesto Ramirez-Gonzalez. |
| EPI_ISL_2476442 | LESP Campeche | Instituto de Diagnostico y Referencia Epidemiologicos (INDRE) | Claudia Wong-Arambula, Abril Rodriguez-Maldonado, Vanessa Rivero-Arredondo, Ariadna Medina-Benitez, Joaquin Quiroz-Mercado, Sergio Rangel-Guerrero, Natividad Cruz-Ortiz, Tatiana Nunez-Garcia, Gisela Barrera-Badillo, Lucia Hernandez-Rivas, Irma Lopez-Martinez, Ernesto Ramirez-Gonzalez. |
| EPI_ISL_2476443 | LESP Baja California | Instituto de Diagnostico y Referencia Epidemiologicos | Claudia Wong-Arambula, Abril Rodriguez-Maldonado, Vanessa Rivero-Arredondo, Ariadna Medina-Benitez, Joaquin Quiroz-Mercado, Sergio |

| | | | |
|---|---|---|---|
| | | (INDRE) | Rangel-Guerrero, Natividad Cruz-Ortiz, Tatiana Nunez-Garcia, Gisela Barrera-Badillo, Lucia Hernandez-Rivas, Irma Lopez-Martinez, Ernesto Ramirez-Gonzalez. |
| EPI_ISL_2476444 | LESP Sinaloa | Instituto de Diagnostico y Referencia Epidemiologicos (INDRE) | Claudia Wong-Arambula, Abril Rodriguez-Maldonado, Vanessa Rivero-Arredondo, Ariadna Medina-Benitez, Joaquin Quiroz-Mercado, Sergio Rangel-Guerrero, Natividad Cruz-Ortiz, Tatiana Nunez-Garcia, Gisela Barrera-Badillo, Lucia Hernandez-Rivas, Irma Lopez-Martinez, Ernesto Ramirez-Gonzalez. |
| EPI_ISL_2479894, EPI_ISL_2479898 | LESP Nuevo Leon | Instituto de Diagnostico y Referencia Epidemiologicos (INDRE) | Claudia Wong-Arambula, Abril Rodriguez-Maldonado, Vanessa Rivero-Arredondo, Ariadna Medina-Benitez, Joaquin Quiroz-Mercado, Sergio Rangel-Guerrero, Natividad Cruz-Ortiz, Tatiana Nunez-Garcia, Gisela Barrera-Badillo, Lucia Hernandez-Rivas, Irma Lopez-Martinez, Ernesto Ramirez-Gonzalez. |
| EPI_ISL_2479906 | LESP Tamaulipas | Instituto de Diagnostico y Referencia Epidemiologicos (INDRE) | Claudia Wong-Arambula, Abril Rodriguez-Maldonado, Vanessa Rivero-Arredondo, Ariadna Medina-Benitez, Joaquin Quiroz-Mercado, Sergio Rangel-Guerrero, Natividad Cruz-Ortiz, Tatiana Nunez-Garcia, Gisela Barrera-Badillo, Lucia Hernandez-Rivas, Irma Lopez-Martinez, Ernesto Ramirez-Gonzalez. |
| EPI_ISL_2480128 | Fulgent Genetics | Centers for Disease Control and Prevention Division of Viral Diseases, Pathogen Discovery | Dakota Howard, Dhwani Batra, Peter W. Cook, Kara Moser, Adrian Paskey, Jason Caravas, Benjamin Rambo-Martin, Shatavia Morrison, Christopher Gulvick, Scott Sammons, Yvette Unoarumhi, Darlene Wagner, Matthew Schmerer, Harry Gao, Mickey Li, John Gao, Joseph Fierro, Benafsh Sapra, Becky Tsai, Yan Meng, Doreen Ng, James Xie, Clinton R. Paden, Duncan MacCannell |
| EPI_ISL_2480209, EPI_ISL_2480293, EPI_ISL_2481627, EPI_ISL_2481707, EPI_ISL_2481730, EPI_ISL_2482011, EPI_ISL_2482056, EPI_ISL_2482167, EPI_ISL_2482257 | Laboratory Corporation of America | Centers for Disease Control and Prevention Division of Viral Diseases, Pathogen Discovery | Dakota Howard, Dhwani Batra, Peter W. Cook, Kara Moser, Adrian Paskey, Jason Caravas, Benjamin Rambo-Martin, Shatavia Morrison, Christopher Gulvick, Scott Sammons, Yvette Unoarumhi, Darlene Wagner, Matthew Schmerer, Minoo Agarwal, Eyad Almasri, Debbie Boles, Ayla Burns, Nuthawin Charoensri, Oren Cohen, Susan Countryman, Mary Ann Cristobal, Bobbi Croy, Suzanne Dale, Hrushikesh Deshmukh, Amanda Douglas, Vincent Drouillon, Marcia Eisenberg, Howard Engler, Rama Ghatti, Prashant Gupta, Susan Hicks, Jake Humphrey, Lax Iyer, Lisa Pfefferle, Manoj Jain, Matthew Robinson, Mohan Kolli, Brian Krueger, Tim Kuphal, Stanley Letovsky, Michael Levandoski, Craig Lukasik, Jonathan Meltzer, Brian Norvell, Mindy Nye, Scott Parker, Christos Petropoulos, John Pruitt, Steven Ragan, Scott Ryan, Mike Sapeta, Jana Schroth, Suresh Babu Selvaraju, Goran Stevovic, Amanda Suchanek, Andrea Throop, Lyndon Tilson, Thomas Urban, Joe Voshell, Kimberly Wagner, Jonathan Williams, Mary Williamson, Qian Zeng, Tricia Zwiefelhofer, Clinton R. Paden, Duncan MacCannell |
| EPI_ISL_2488881, EPI_ISL_2488892, EPI_ISL_2488929, EPI_ISL_2488968, EPI_ISL_2489018, EPI_ISL_2489031 | Mako Medical | Centers for Disease Control and Prevention Division of Viral Diseases, Pathogen Discovery | Dakota Howard, Dhwani Batra, Peter W. Cook, Kara Moser, Adrian Paskey, Jason Caravas, Benjamin Rambo-Martin, Shatavia Morrison, Christopher Gulvick, Scott Sammons, Yvette Unoarumhi, Darlene Wagner, Matthew Schmerer,Matthew Tugwell, Lauren Moon, Clinton R. Paden, Duncan MacCannell |
| EPI_ISL_2489072, EPI_ISL_2489133, EPI_ISL_2489160, EPI_ISL_2489349, EPI_ISL_2489450, EPI_ISL_2489458, EPI_ISL_2489492 | Aegis Sciences Corporation | Centers for Disease Control and Prevention Division of Viral Diseases, Pathogen Discovery | Dakota Howard, Dhwani Batra, Peter W. Cook, Kara Moser, Adrian Paskey, Jason Caravas, Benjamin Rambo-Martin, Shatavia Morrison, Christopher Gulvick, Scott Sammons, Yvette Unoarumhi, Darlene Wagner, Matthew Schmerer, Cyndi Clark, Patrick Campbell, Rob Case, Vikramsinha Ghorpade, Holly Houdeshell, Ola Kvalvaag, Dillon Nall, Ethan Sanders, Alec Vest, Shaun Westlund, Matthew Hardison, Clinton R. Paden, Duncan MacCannell |
| EPI_ISL_2489790, EPI_ISL_2489792 | State Testing Facility | Altius Institute for Biomedical Research | Daniel Bates, Rebecca Bruders, Michael Buckley, Mark Frerker, Amanda Gale, Clem Green, Muhammad Halimun, Kneshay Harper, Matt Hartman, Alex Isner, Audra Johnson, Jessica Kunder, Lauren Mitchell, Jemma Nelson, Alex Nguyen, Sofia Olsson, Sadie Patraw, Tobias Ragoczy, Joshua Richards, Jean Robinston, Jacob Rodriguez, John Stamatoyannopoulos, Eric Thorland, Julia Wald |
| EPI_ISL_2490046, EPI_ISL_2490077 | Seattle Flu Study | Seattle Flu Study | Deborah A. Nickerson, Chris D. Frazar, Jover Lee, Benjamin Pelle, Erica Ryke, Matthew Richardson, Amanda Adler, Elisabeth Brandstetter, Peter D. Han, Kairsten Fay, Misja Ilcisin, Kirsten Lacombe, Thomas R. Sibley, Melissa Truong, Caitlin R. Wolf, Karen Cowgill, Stephanie Schrag, Jeff Duchin, Michael Boeckh, Janet A. Englund, Michael Famulare, Barry R. Lutz, Mark J. Rieder, Lea M. Starita, Matthew Thompson, Helen Y. Chu, Trevor Bedford, Jay Shendure |
| EPI_ISL_2490181, EPI_ISL_2490198 | Northwest Laboratory | Seattle Flu Study | Deborah A. Nickerson, Chris D. Frazar, Jover Lee, Benjamin Pelle, Erica Ryke, Matthew Richardson, Amanda Adler, Elisabeth Brandstetter, Peter D. Han, Kairsten Fay, Misja Ilcisin, Kirsten Lacombe, Thomas R. Sibley, Melissa Truong, Caitlin R. Wolf, Romesh Gautom, Geoff Melly, Brian Hiatt, Philip Dykema, Scott Lindquist, Michael Boeckh, Janet A. Englund, Michael Famulare, Barry R. Lutz, Mark J. Rieder, Lea M. Starita, Matthew Thompson, Helen Y. Chu, Jay Shendure, Trevor Bedford |
| EPI_ISL_2490266, EPI_ISL_2490289 | Incyte Diagnostics Spokane | Seattle Flu Study | Deborah A. Nickerson, Chris D. Frazar, Jover Lee, Benjamin Pelle, Erica Ryke, Matthew Richardson, Amanda Adler, Elisabeth Brandstetter, Peter D. Han, Kairsten Fay, Misja Ilcisin, Kirsten Lacombe, Thomas R. Sibley, Melissa Truong, Caitlin R. Wolf, Romesh Gautom, Geoff Melly, Brian Hiatt, Philip Dykema, Scott Lindquist, Michael Boeckh, Janet A. Englund, Michael Famulare, Barry R. Lutz, Mark J. Rieder, Lea M. Starita, Matthew Thompson, Helen Y. Chu, Jay Shendure, Trevor Bedford |
| EPI_ISL_2490394, EPI_ISL_2490409, EPI_ISL_2490411, EPI_ISL_2490445, EPI_ISL_2490457, EPI_ISL_2490462 | Centro de Investigación Biomédica del Noreste (CIBIN) | Centro de Investigación en Enfermedades Infecciosas (CIENI), Instituto Nacional de Enfermedades Respiratorias (INER) | Consorcio Mexicano de Vigilancia Genómica (CoViGen-Mex). Authors (in alphabetical order): Julio Elias Alvarado-Yaah, Carlos F. Arias, Santiago Ávila-Ríos, Eduardo Becerril-Vargas, Víctor Hugo Borja-Aburto, Celia Boukadida, Cristóbal Cháidez-Quiróz, Juan Bautista Chale-Dzul, Ricardo Ciria Merce, Andreu Comas-García, Célida Duque Molina, Julissa Enciso-Ibarra, José Antonio Enciso-Moreno, Gloria Elena Espinosa-Ayala, Fernando Fontove-Herrera, Daniel Fregoso-Rueda, Victor Eduardo Garcia-Arias, Alejandra Garcia-Gasca, Bruno Gomez-Gil, Jean Pierre González, Irvin González-López, Concepción Grajales-Muñiz, Ricardo Grande, Rosa María Gutierrez Rios, Alejandra Hernández-Terán, Alfredo Herrera-Estrella, Carla Ivón Herrera-Najera, Pavel Isa, Daniel Lira Morales, Susana Lopez, Antonio Loza Román, Brenda Irasema Maldonado-Meza, Bernardo Martínez-Miguel, José Arturo Martínez-Orozco, Célida Martínez- Rodríguez, Margarita Matías-Florentino, Fidencio Mejía-Nepomuceno, María Guadalupe de Jesús Mireles-Rivera, Gloria María Molina-Salinas, Hector Montoya-Fuentes, , Mario Mújica-Sánchez, José Esteban Muñoz-Medina, José de Jesús Nuñez-Contreras, Alicia Ocaña-Mondragón, Luis Alberto Ochoa-Carrera, Hector Esteban Paz-Juárez, Marissa Perez-Garcia, Francisco Pulido, Helen Haydee Fernanda Ramírez-Plascencia, Jorge Salas-Hernández, Angel Gustavo Salas-Lais, Alejandro Sanchez-Flores, Clara Esperanza Santacruz-Tinoco, María Guadalupe Santiago-Mauricio, Selene Zárate, Nelly Sélem-Mojica, Blanca Taboada, Gloria Vazquez, Joel Armando Vázquez-Pérez. |
| EPI_ISL_2490501, EPI_ISL_2490504, EPI_ISL_2490506, EPI_ISL_2490514, EPI_ISL_2490515, EPI_ISL_2490524, EPI_ISL_2490531, EPI_ISL_2490533, EPI_ISL_2490535 | Unidad de Investigación Biomédica de Zacatecas (UIBZ) | Centro de Investigación en Enfermedades Infecciosas (CIENI), Instituto Nacional de Enfermedades Respiratorias (INER) | Consorcio Mexicano de Vigilancia Genómica (CoViGen-Mex). Authors (in alphabetical order): Julio Elias Alvarado-Yaah, Carlos F. Arias, Santiago Ávila-Ríos, Eduardo Becerril-Vargas, Víctor Hugo Borja-Aburto, Celia Boukadida, Cristóbal Cháidez-Quiróz, Juan Bautista Chale-Dzul, Ricardo Ciria Merce, Andreu Comas-García, Célida Duque Molina, Julissa Enciso-Ibarra, José Antonio Enciso-Moreno, Gloria Elena Espinosa-Ayala, Fernando Fontove-Herrera, Daniel Fregoso-Rueda, Victor Eduardo Garcia-Arias, Alejandra Garcia-Gasca, Bruno Gomez-Gil, Jean Pierre González, Irvin González-López, Concepción Grajales-Muñiz, Ricardo Grande, Rosa María Gutierrez Rios, Alejandra Hernández-Terán, Alfredo Herrera-Estrella, Carla Ivón Herrera-Najera, Pavel Isa, Daniel Lira Morales, Susana Lopez, Antonio Loza Román, Brenda Irasema Maldonado-Meza, Bernardo Martínez-Miguel, José Arturo Martínez-Orozco, Célida Martínez- Rodríguez, Margarita Matías-Florentino, Fidencio Mejía-Nepomuceno, María Guadalupe de Jesús Mireles-Rivera, Gloria María Molina-Salinas, Hector Montoya-Fuentes, , Mario Mújica-Sánchez, José Esteban Muñoz-Medina, José de Jesús Nuñez-Contreras, Alicia Ocaña-Mondragón, Luis Alberto Ochoa-Carrera, Hector Esteban Paz-Juárez, Marissa Perez-Garcia, Francisco Pulido, Helen Haydee Fernanda Ramírez-Plascencia, Jorge Salas-Hernández, Angel Gustavo Salas-Lais, Alejandro Sanchez-Flores, Clara Esperanza Santacruz-Tinoco, María Guadalupe Santiago-Mauricio, Selene Zárate, Nelly Sélem-Mojica, Blanca Taboada, Gloria Vazquez, Joel Armando Vázquez-Pérez. |
| EPI_ISL_2490567 | Laboratorio Central de Epidemiología (LCE) | Centro de Investigación en Enfermedades Infecciosas (CIENI), Instituto Nacional de Enfermedades Respiratorias (INER) | Consorcio Mexicano de Vigilancia Genómica (CoViGen-Mex). Authors (in alphabetical order): Julio Elias Alvarado-Yaah, Carlos F. Arias, Santiago Ávila-Ríos, Eduardo Becerril-Vargas, Víctor Hugo Borja-Aburto, Celia Boukadida, Cristóbal Cháidez-Quiróz, Juan Bautista Chale-Dzul, Ricardo Ciria Merce, Andreu Comas-García, Célida Duque Molina, Julissa Enciso-Ibarra, José Antonio Enciso-Moreno, Gloria Elena Espinosa-Ayala, Fernando Fontove-Herrera, Daniel Fregoso-Rueda, Victor Eduardo Garcia-Arias, Alejandra Garcia-Gasca, Bruno Gomez-Gil, Jean Pierre González, Irvin González-López, Concepción Grajales-Muñiz, Ricardo Grande, Rosa María Gutierrez Rios, Alejandra Hernández-Terán, Alfredo Herrera-Estrella, Carla Ivón Herrera-Najera, Pavel Isa, Daniel Lira Morales, Susana Lopez, Antonio Loza Román, Brenda Irasema Maldonado-Meza, Bernardo Martínez-Miguel, José Arturo Martínez-Orozco, Célida Martínez- Rodríguez, Margarita Matías-Florentino, Fidencio Mejía-Nepomuceno, María Guadalupe de Jesús Mireles-Rivera, Gloria María Molina-Salinas, Hector Montoya-Fuentes, , Mario Mújica-Sánchez, José Esteban Muñoz-Medina, José de Jesús Nuñez-Contreras, Alicia Ocaña-Mondragón, Luis Alberto Ochoa-Carrera, Hector Esteban Paz-Juárez, Marissa Perez-Garcia, Francisco Pulido, Helen Haydee Fernanda Ramírez-Plascencia, Jorge Salas-Hernández, Angel Gustavo Salas-Lais, Alejandro Sanchez-Flores, Clara Esperanza Santacruz-Tinoco, María Guadalupe Santiago-Mauricio, Selene Zárate, Nelly Sélem-Mojica, Blanca Taboada, Gloria Vazquez, Joel Armando Vázquez-Pérez. |

| | | | |
|---|---|---|---|
| EPI_ISL_2494634 | UCSC Genomics Institute | UCSC Genomics Institute | Mark Akeson, Ikenna Anigbogu, Eric Beraut, Isabel Bjork, Molly Cassatt-Johnstone, Terren Chang, Russell Corbett-Detig, Namrita Dhillon, Maximilian Haeussler, David Haussler, Angie Hinrichs, Miten Jain, Joshua Kapp, A. Marm Kilpatrick, Jakob McBroome, Hugh Olsen, Jeremy Sanford, Beth Shapiro, Michael Stone, Bryan Thornlow, Yatish Turakhia, Ciara Wanket |
| EPI_ISL_2496915, EPI_ISL_2496963, EPI_ISL_2496970, EPI_ISL_2496974 | CA DPH Viral and Rickettsial Disease Laboratory | Chan-Zuckerberg Biohub | CZB Cliahub Consortium |
| EPI_ISL_2496976 | Santa Clara County Public Health Laboratory | Chan-Zuckerberg Biohub | CZB Cliahub Consortium |
| EPI_ISL_2496984, EPI_ISL_2496985, EPI_ISL_2496991, EPI_ISL_2496993, EPI_ISL_2496994, EPI_ISL_2496997 | CA DPH Viral and Rickettsial Disease Laboratory | Chan-Zuckerberg Biohub | CZB Cliahub Consortium |
| EPI_ISL_2497254, EPI_ISL_2497306, EPI_ISL_2497326, EPI_ISL_2497335, EPI_ISL_2497360, EPI_ISL_2497369, EPI_ISL_2497373, EPI_ISL_2497404, EPI_ISL_2497411 | UW Virology Lab | UW Virology Lab | Pavitra Roychoudhury, Hong Xie, Lasata Shrestha, Tien V. Nguyen, Shah Mohamed Bakhash, Michelle Lin, Noah R. Baker, Ricardo Perez, Sean Ellis, Nathan Breit, Robert J. Livingston, Meei-Li Huang, Keith R Jerome, Patrick Mathias, Alexander Greninger |
| EPI_ISL_2497697 | Humboldt County Public Health Laboratory | Chan-Zuckerberg Biohub | CZB Cliahub Consortium |
| EPI_ISL_2497710, EPI_ISL_2497768 | CA DPH Viral and Rickettsial Disease Laboratory | Chan-Zuckerberg Biohub | CZB Cliahub Consortium |
| EPI_ISL_2497931 | UW Virology Lab | UW Virology Lab | Pavitra Roychoudhury, Hong Xie, Lasata Shrestha, Tien V. Nguyen, Shah Mohamed Bakhash, Michelle Lin, Noah R. Baker, Ricardo Perez, Sean Ellis, Nathan Breit, Robert J. Livingston, Meei-Li Huang, Keith R Jerome, Patrick Mathias, Alexander Greninger |
| EPI_ISL_2499966 | LESP Sinaloa | Instituto de Diagnostico y Referencia Epidemiologicos (INDRE) | Claudia Wong-Arambula, Abril Rodriguez-Maldonado, Vanessa Rivero-Arredondo, Ariadna Medina-Benitez, Joaquin Quiroz-Mercado, Sergio Rangel-Guerrero, Natividad Cruz-Ortiz, Tatiana Nunez-Garcia, Gisela Barrera-Badillo, Lucia Hernandez-Rivas, Irma Lopez-Martinez, Ernesto Ramirez-Gonzalez. |
| EPI_ISL_2499967 | LESP Michoacan | Instituto de Diagnostico y Referencia Epidemiologicos (INDRE) | Claudia Wong-Arambula, Abril Rodriguez-Maldonado, Vanessa Rivero-Arredondo, Ariadna Medina-Benitez, Joaquin Quiroz-Mercado, Sergio Rangel-Guerrero, Natividad Cruz-Ortiz, Tatiana Nunez-Garcia, Gisela Barrera-Badillo, Lucia Hernandez-Rivas, Irma Lopez-Martinez, Ernesto Ramirez-Gonzalez. |
| EPI_ISL_2500078, EPI_ISL_2500202 | Humboldt County Public Health Laboratory | Chan-Zuckerberg Biohub | CZB Cliahub Consortium |
| EPI_ISL_2501000 | Gundersen Clinical Microbiology Laboratory | Kabara Cancer Research Institute | Craig S. Richmond, Paraic A. Kenny |
| EPI_ISL_2501523, EPI_ISL_2501524 | UW Virology Lab | UW Virology Lab | Pavitra Roychoudhury, Hong Xie, Lasata Shrestha, Tien V. Nguyen, Shah Mohamed Bakhash, Michelle Lin, Noah R. Baker, Ricardo Perez, Sean Ellis, Nathan Breit, Robert J. Livingston, Meei-Li Huang, Keith R Jerome, Patrick Mathias, Alexander Greninger |
| EPI_ISL_2502729 | HOSPITAL LA ANEXION | Inciensa, Instituto Costarricense de InvestigaciOn y Enseñanza en NutriciOn y Salud | Francisco Duarte, Hebleen Porras, Claudio Soto-Garita, Estela Cordero, Adriana Godínez, Melany CalderOn, Jose Luis Vargas, Mariela Gutierrez, Joselyn Prado & Ivanna Krize-Morún |
| EPI_ISL_2504066 | A. Krumbholz, Labor Dr. Krause und Kollegen MVZ GmbH, Kiel | Charite Universitatsmedizin Berlin, Institut fur Virologie/Labor Berlin | Victor M Corman, Andi Krumbholz, Joern Beheim-Schwarzbach, Barbara Müehlemann, Julia Schneider, Talitha Veith, Tobias Bleicker, Julia Tesch, Peter Menzel, Christine Stephan, Rolf Schwarzer, Terry Jones, Christian Drosten |
| EPI_ISL_2508590 | Genetica Molecular and Subdepartamento de Virologia ISP Chile | Instituto de Salud Publica de Chile | Karen Orostica, Constanza Campano, Barbara Parra, Loredana Arata, Gisselle Barra, Patricia Bustos, Rodrigo Fasce, Javier Tognarelli, Andres Castillo, Soledad Ulloa, Jorge Fernandez |
| EPI_ISL_2525466, EPI_ISL_2525484, EPI_ISL_2525533, EPI_ISL_2525654, EPI_ISL_2525752, EPI_ISL_2527584 | Fulgent Genetics | Centers for Disease Control and Prevention Division of Viral Diseases, Pathogen Discovery | Dakota Howard, Dhwani Batra, Peter W. Cook, Kara Moser, Adrian Paskey, Jason Caravas, Benjamin Rambo-Martin, Shatavia Morrison, Christopher Gulvick, Scott Sammons, Yvette Unoarumhi, Darlene Wagner, Matthew Schmerer, Harry Gao, Mickey Li, John Gao, Joseph Fierro, Benafsh Sapra, Becky Tsai, Yan Meng, Doreen Ng, James Xie, Clinton R. Paden, Duncan MacCannell |
| EPI_ISL_2528580, EPI_ISL_2528933 | Aegis Sciences Corporation | Centers for Disease Control and Prevention Division of Viral Diseases, Pathogen Discovery | Dakota Howard, Dhwani Batra, Peter W. Cook, Kara Moser, Adrian Paskey, Jason Caravas, Benjamin Rambo-Martin, Shatavia Morrison, Christopher Gulvick, Scott Sammons, Yvette Unoarumhi, Darlene Wagner, Matthew Schmerer, Cyndi Clark, Patrick Campbell, Rob Case, Vikramsinha Ghorpade, Holly Houdeshell, Ola Kvalvaag, Dillon Nall, Ethan Sanders, Alec Vest, Shaun Westlund, Matthew Hardison, Clinton R. Paden, Duncan MacCannell |
| EPI_ISL_2529223 | Quest Diagnostics Incorporated | Centers for Disease Control and Prevention Division of Viral Diseases, Pathogen Discovery | Dakota Howard, Dhwani Batra, Peter W. Cook, Kara Moser, Adrian Paskey, Jason Caravas, Benjamin Rambo-Martin, Shatavia Morrison, Christopher Gulvick, Scott Sammons, Yvette Unoarumhi, Darlene Wagner, Matthew Schmerer, S. H. Rosenthal, A. Gerasimova, R. M. Kagan, B. Anderson, M. Hua, Y. Liu, L.E. Bernstein, K.E. Livingston, A. Perez, I. A. Shlyakhter, R. V. Rolando, R. Owen, P. Tanpaiboon, F. Lacbawan, Clinton R. Paden, Duncan MacCannell |
| EPI_ISL_2533921 | Hospital Sharp | Microbial Genomics Laboratory | Consorcio Mexicano de Vigilancia Genómica (CoViGen-Mex). Authors (in alphabetical order): Julio Elias Alvarado-Yaah, Carlos F. Arias, Santiago Ávila-Ríos, Eduardo Becerril-Vargas, Víctor Hugo Borja-Aburto, Celia Boukadida, Cristóbal Cháidez-Quiróz, Juan Bautista Chale-Dzul, Ricardo Ciria Merce, Andreu Comas-García, Célida Duque Molina, Julissa Enciso-Ibarra, José Antonio Enciso-Moreno, Gloria Elena Espinosa-Ayala, Fernando Fontove-Herrera, Daniel Fregoso-Rueda, Victor Eduardo García-Arias, Alejandra García-Gasca, Bruno Gómez-Gil, Jean Pierre González, Irvin González-López, Concepción Grajales-Muñiz, Ricardo Grande, Rosa María Gutiérrez Rios, Jesús Hernández, Alejandra Hernández-Terán, Alfredo Herrera-Estrella, Carla Ivón Herrera-Najera, Pavel Isa, Verónica Mata-Haro, Daniel Lira Morales, Susana Lopez, Antonio Loza Román, Brenda Irasema Maldonado-Meza, Bernardo Martínez-Miguel, José Arturo Martínez-Orozco, Célida Martínez- Rodríguez, Margarita Matías-Florentino, Fidencio Mejía-Nepomuceno, María Guadalupe de Jesús Mireles-Rivera, Gloria María Molina-Salinas, Hector Montoya-Fuentes, , Mario Mújica-Sánchez, José Esteban Muñoz-Medina, José de Jesús Nuñez-Contreras, Alicia Ocaña-Mondragón, Luis Alberto Ochoa-Carrera, Hector Esteban Paz-Juárez, Marissa Perez-Garcia, Francisco Pulido, Helen Haydee Fernanda Ramírez-Plascencia, Jorge Salas-Hernández, Angel Gustavo Salas-Lais, Alejandro Sánchez-Flores, Clara Esperanza Santacruz-Tinoco, María Guadalupe Santiago-Mauricio, Selene Zárate, Nelly Sélem-Mojica, Blanca Taboada, Gloria Vazquez, Joel Armando Vázquez-Pérez. |
| EPI_ISL_2536580 | Northwest Laboratory | Seattle Flu Study | Deborah A. Nickerson, Chris D. Frazar, Jover Lee, Benjamin Pelle, Erica Ryke, Matthew Richardson, Amanda Adler, Elisabeth Brandstetter, Peter D. Han, Kairsten Fay, Misja Ilcisin, Kirsten Lacombe, Thomas R. Sibley, Melissa Truong, Caitlin R. Wolf, Romesh Gautom, Geoff Melly, Brian Hiatt, Philip Dykema, Scott Lindquist, Michael Boeckh, Janet A. Englund, Michael Famulare, Barry R. Lutz, Mark J. Rieder, Lea M. Starita, Matthew Thompson, Helen Y. Chu, Jay Shendure, Trevor Bedford |
| EPI_ISL_2536939 | Centracare Laboratory Services | Minnesota Department of Health, Public Health Laboratory | Alexandra Lorentz, Jacob Garfin, Matt Plumb, and Xiong Wang |
| EPI_ISL_2543569 | Genetica Molecular and Subdepartamento de Virologia ISP Chile | Instituto de Salud Publica de Chile | Karen Orostica, Constanza Campano, Barbara Parra, Loredana Arata, Gisselle Barra, Patricia Bustos, Rodrigo Fasce, Javier Tognarelli, Andres Castillo, Soledad Ulloa, Jorge Fernandez |
| EPI_ISL_2544528 | Basurto University Hospital: Clinical Microbiology Laboratory | Biocruces Bizkaia | Mikel Urrutikoetxea-Gutiérrez, Mª Carmen Nieto Toboso, Estibaliz Ugalde Zarraga, Mikel Gallego Rodrigo, Ana de la Hoz, José Luis Díaz de Tuesta del Arco |
| EPI_ISL_2544984, EPI_ISL_2545001, EPI_ISL_2545002, EPI_ISL_2545060, EPI_ISL_2545128, EPI_ISL_2545178 | UW Virology Lab | UW Virology Lab | Pavitra Roychoudhury, Hong Xie, Lasata Shrestha, Shah Mohamed Bakhash, Tien V. Nguyen, Noah R. Baker, Sean Ellis, Meei-Li Huang, Keith R Jerome, Alexander Greninger |
| EPI_ISL_2545700, EPI_ISL_2545701 | LESP Queretaro | Instituto de Diagnostico y Referencia Epidemiologicos (INDRE) | Claudia Wong-Arambula, Abril Rodriguez-Maldonado, Vanessa Rivero-Arredondo, Ariadna Medina-Benitez, Joaquin Quiroz-Mercado, Sergio Rangel-Guerrero, Natividad Cruz-Ortiz, Tatiana Nunez-Garcia, Gisela Barrera-Badillo, Lucia Hernandez-Rivas, Irma Lopez-Martinez, Ernesto Ramirez-Gonzalez. |
| EPI_ISL_2545702 | LESP Michoacan | Instituto de Diagnostico y Referencia Epidemiologicos (INDRE) | Claudia Wong-Arambula, Abril Rodriguez-Maldonado, Vanessa Rivero-Arredondo, Ariadna Medina-Benitez, Joaquin Quiroz-Mercado, Sergio Rangel-Guerrero, Natividad Cruz-Ortiz, Tatiana Nunez-Garcia, Gisela Barrera-Badillo, Lucia Hernandez-Rivas, Irma Lopez-Martinez, Ernesto Ramirez-Gonzalez. |

| | | | |
|---|---|---|---|
| EPI_ISL_2545703, EPI_ISL_2545704 | LESP Sinaloa | Instituto de Diagnostico y Referencia Epidemiologicos (INDRE) | Claudia Wong-Arambula, Abril Rodriguez-Maldonado, Vanessa Rivero-Arredondo, Ariadna Medina-Benitez, Joaquin Quiroz-Mercado, Sergio Rangel-Guerrero, Natividad Cruz-Ortiz, Tatiana Nunez-Garcia, Gisela Barrera-Badillo, Lucia Hernandez-Rivas, Irma Lopez-Martinez, Ernesto Ramirez-Gonzalez. |
| EPI_ISL_2549333, EPI_ISL_2549339, EPI_ISL_2549415, EPI_ISL_2549456, EPI_ISL_2549508, EPI_ISL_2549547, EPI_ISL_2549550, EPI_ISL_2549554 | UW Virology Lab | UW Virology Lab | Pavitra Roychoudhury, Hong Xie, Lasata Shrestha, Tien V. Nguyen, Shah Mohamed Bakhash, Michelle Lin, Noah R. Baker, Ricardo Perez, Sean Ellis, Nathan Breit, Robert J. Livingston, Meei-Li Huang, Keith R Jerome, Patrick Mathias, Alexander Greninger |
| EPI_ISL_2550795 | Maryland Public Health Laboratory (MD PHL) | Maryland Public Health Laboratory (MD PHL) | Maryland Department of Health Laboratories Administration |
| EPI_ISL_2558098, EPI_ISL_2558191, EPI_ISL_2558212, EPI_ISL_2558257 | Wisconsin State Laboratory of Hygiene Communicable Disease Division | Wisconsin State Laboratory of Hygiene Communicable Disease Division | Abigail C. Shockey, Alicia J. Mooney, Erika M. Hanson, Tonya Danz, Richard Griesser, Sara Wagner, Kelsey R. Florek |
| EPI_ISL_2566354, EPI_ISL_2566358 | Operation Infectious Diseases, Naval Health Research Center | Operation Infectious Diseases, Naval Health Research Center | Pan,R.W., Underwood,R.S., Balansay-Ames,M.S., Myers,C.A., Fierro,M., Kriner,P., Iniguez-Stevens,E., Moser,K., Phippard,A.E. |
| EPI_ISL_2567110 | Florida Bureau of Public Health Laboratories | Florida Bureau of Public Health Laboratories | Sarah Schmedes, Jason Blanton |
| EPI_ISL_2587237, EPI_ISL_2587416, EPI_ISL_2587751 | SK-Roy Romanow Provincial Laboratory | National Microbiology Laboratory (NML) | Anna Majer, Shari Tyson, Grace Seo, Philip Mabon, Elsie Grudeski, Rhiannon Huzarewich, Russell Mandes, Anneliese Landgraff, Jennifer Tanner, Natalie Knox, Morag Graham, Gary Van Domselaar, Ryan McDonald, Amanda Lang, Rachel DePaulo, Jessica Minion, Nathalie Bastien, Yan Li, Timothy Booth, Darian Hole, Madison Chapel, Kirsten Biggar, CanCOGeN's metadata curation team, Public Health Agency of Canada CanCOGeN team |
| EPI_ISL_2598722 | Fulgent Genetics | Centers for Disease Control and Prevention Division of Viral Diseases, Pathogen Discovery | Dakota Howard, Dhwani Batra, Peter W. Cook, Kara Moser, Adrian Paskey, Jason Caravas, Benjamin Rambo-Martin, Shatavia Morrison, Christopher Gulvick, Scott Sammons, Yvette Unoarumhi, Darlene Wagner, Matthew Schmerer, Harry Gao, Mickey Li, John Gao, Joseph Fierro, Benafsh Sapra, Becky Tsai, Yan Meng, Doreen Ng, James Xie, Clinton R. Paden, Duncan MacCannell |
| EPI_ISL_2599608 | Helix/Illumina | Centers for Disease Control and Prevention Division of Viral Diseases, Pathogen Discovery | Dakota Howard, Dhwani Batra, Peter W. Cook, Kara Moser, Adrian Paskey, Jason Caravas, Benjamin Rambo-Martin, Shatavia Morrison, Christopher Gulvick, Scott Sammons, Yvette Unoarumhi, Darlene Wagner, Matthew Schmerer, Eileen de Feo, Jan Antico, Christine Tran, Matthew Tolentino, Shannon Wickline, Kim Gietzen, Brad Sickler, Jingtao Liu, Eric Allen, Phil Febbo, Nicole L. Washington, Simon White, Geraint Levan, Kelly Schiabor Barrett, Elizabeth Cirulli, Alexandre Bolze, Ary Ascencio, Charlotte Rivera-Garcia, Ryan Cho, Jason Nguyen, Sherry Wang, Jimmy Ramirez, Tyler Cassens, Efren Sandoval, Magnus Isaksson, William Lee, David Becker, Marc Laurent, James Lu, Clinton R. Paden, Duncan MacCannell |
| EPI_ISL_2602373 | Gravity Diagnostics, LLC | Gravity Diagnostics, LLC | Gravity Diagnostics |
| EPI_ISL_2611257, EPI_ISL_2611278, EPI_ISL_2611348 | Laboratory Corporation of America | Centers for Disease Control and Prevention Division of Viral Diseases, Pathogen Discovery | Dakota Howard, Dhwani Batra, Peter W. Cook, Kara Moser, Adrian Paskey, Jason Caravas, Benjamin Rambo-Martin, Shatavia Morrison, Christopher Gulvick, Scott Sammons, Yvette Unoarumhi, Darlene Wagner, Matthew Schmerer, Minoo Agarwal, Eyad Almasri, Debbie Boles, Ayla Burns, Nuthawin Charoensri, Oren Cohen, Susan Countryman, Mary Ann Cristobal, Bobbi Croy, Suzanne Dale, Hrushikesh Deshmukh, Amanda Douglas, Vincent Drouillon, Marcia Eisenberg, Howard Engler, Rama Ghatti, Prashant Gupta, Susan Hicks, Jake Humphrey, Lax Iyer, Lisa Pfefferle, Manoj Jain, Matthew Robinson, Mohan Kolli, Brian Krueger, Tim Kuphal, Stanley Letovsky, Michael Levandoski, Craig Lukasik, Jonathan Meltzer, Brian Norvell, Mindy Nye, Scott Parker, Christos Petropoulos, John Pruitt, Steven Ragan, Scott Ryan, Mike Sapeta, Jana Schroth, Suresh Babu Selvaraju, Goran Stevovic, Amanda Suchanek, Andrea Throop, Lyndon Tilson, Thomas Urban, Joe Voshell, Kimberly Wagner, Jonathan Williams, Mary Williamson, Qian Zeng, Tricia Zwiefelhofer, Clinton R. Paden, Duncan MacCannell |
| EPI_ISL_2612520 | Centracare Laboratory Services | Minnesota Department of Health, Public Health Laboratory | Alexandra Lorentz, Jacob Garfin, Matt Plumb, and Xiong Wang |
| EPI_ISL_2612548 | Oregon State Public Health Laboratory | Oregon State Public Health Laboratory | Rafia Razzaque, Eugene Yeboah, Vanda Makris, Laura Tsaknaridis, John Fontana and Shane Sevey |
| EPI_ISL_2613845 | Gravity Diagnostics, LLC | Gravity Diagnostics, LLC | Gravity Diagnostics |
| EPI_ISL_2617995, EPI_ISL_2618032, EPI_ISL_2618034, EPI_ISL_2618082 | UW Virology Lab | UW Virology Lab | Pavitra Roychoudhury, Hong Xie, Lasata Shrestha, Tien V. Nguyen, Shah Mohamed Bakhash, Michelle Lin, Noah R. Baker, Ricardo Perez, Sean Ellis, Nathan Breit, Robert J. Livingston, Meei-Li Huang, Keith R Jerome, Patrick Mathias, Alexander Greninger |
| EPI_ISL_2618141, EPI_ISL_2618150, EPI_ISL_2618183, EPI_ISL_2618229, EPI_ISL_2618235, EPI_ISL_2618344, EPI_ISL_2618348, EPI_ISL_2618670, EPI_ISL_2618673, EPI_ISL_2618683, EPI_ISL_2618703, EPI_ISL_2618707, EPI_ISL_2618725 | | | |
| see above | UW Virology Lab | UW Virology Lab | Pavitra Roychoudhury, Hong Xie, Lasata Shrestha, Shah Mohamed Bakhash, Tien V. Nguyen, Noah R. Baker, Sean Ellis, Meei-Li Huang, Keith R Jerome, Alexander Greninger |
| EPI_ISL_2627045, EPI_ISL_2627046 | URMC LABS | Wadsworth Center, New York State Department of Health | Kirsten St. George, Daryl M. Lamson, Alexis Russell, Matthew Shudt, Melissa A Leisner, Jonathan Plitnick, Catharine Prussing, Navjot Singh, John Kelly, Erasmus Schneider, Erica Lasek-Nesselquist |
| EPI_ISL_2628812, EPI_ISL_2628819 | San Diego County Public Health Laboratory | Andersen lab at Scripps Research | SEARCH Alliance San Diego with Ashleigh Murphy, Jovan Shephard, Brett Austin |
| EPI_ISL_2632714 | Limbach - MVZ Humangenetik Ulm | Robert Koch Institute | unknown |
| EPI_ISL_2645346 | MI - Michigan Department of Health and Human Services - Bureau of Laboratories | Centers for Disease Control and Prevention Division of Viral Diseases, Pathogen Discovery | Mili Sheth, Sarah Nobles, Jasmine Padilla, Mark Burroughs, Shoshona Le, Katie Dillon, Peter Cook, Clinton R. Paden, Dhwani Batra, Krista Queen, Kristen Knipe, Dakota Howard, Yvette Unoarumhi, Darlene Wagner, Matthew Schmerer, Ben L. Rambo-Martin, Kristine Lacek, Sam Shepard, Alison Laufer Halpin, Dave Wentworth, Vivien Dugan, Suxiang Tong, Justin Lee |
| EPI_ISL_2646370, EPI_ISL_2646424, EPI_ISL_2646438, EPI_ISL_2646471, EPI_ISL_2646522, EPI_ISL_2646586, EPI_ISL_2646594, EPI_ISL_2646649, EPI_ISL_2646693, EPI_ISL_2646701, EPI_ISL_2647239, EPI_ISL_2647259, EPI_ISL_2647395, EPI_ISL_2647496, EPI_ISL_2647615, EPI_ISL_2647685, EPI_ISL_2647797, EPI_ISL_2647875 | | | |
| see above | Mako Medical | Centers for Disease Control and Prevention Division of Viral Diseases, Pathogen Discovery | Dakota Howard, Dhwani Batra, Peter W. Cook, Kara Moser, Adrian Paskey, Jason Caravas, Benjamin Rambo-Martin, Shatavia Morrison, Christopher Gulvick, Scott Sammons, Yvette Unoarumhi, Darlene Wagner, Matthew Schmerer,Matthew Tugwell, Lauren Moon, Clinton R. Paden, Duncan MacCannell |
| EPI_ISL_2650519 | University of Oregon COVID-19 MAP Laboratory | University of Oregon Genomics and Cell Charactericzation Core Facility (GC3F) | Douglas Turnbull, Ariana White, Jeff Bishop, Jason Carriere, Jason Sydes, Peter Batzel |
| EPI_ISL_2651069, EPI_ISL_2651072, EPI_ISL_2651078 | Washington State Department of Health Public Health Laboratories | Washington State Department of Health Public Health Laboratories | Drew MacKellar, Philip Dykema, Denny Russell, Joenice Gonzalez, Hannah Gray, Geoff Melly, Vanessa De Los Santos, Darren Lucas, JohnAric Peterson, Avi Singh, Rebecca Cao |
| EPI_ISL_2652303, EPI_ISL_2652377, EPI_ISL_2652384 | Quest Diagnostics Incorporated | Centers for Disease Control and Prevention Division of Viral Diseases, Pathogen Discovery | Dakota Howard, Dhwani Batra, Peter W. Cook, Kara Moser, Adrian Paskey, Jason Caravas, Benjamin Rambo-Martin, Shatavia Morrison, Christopher Gulvick, Scott Sammons, Yvette Unoarumhi, Darlene Wagner, Matthew Schmerer, S. H. Rosenthal, A. Gerasimova, R. M. Kagan, B. Anderson, M. Hua, Y. Liu, L.E. Bernstein, K.E. Livingston, A. Perez, I. A. Shlyakhter, R. V. Rolando, R. Owen, P. Tanpaiboon, F. Lacbawan, Clinton R. Paden, Duncan MacCannell |
| EPI_ISL_2652628, EPI_ISL_2652681 | Infinity Biologix | Centers for Disease Control and Prevention Division of Viral Diseases, Pathogen Discovery | Dakota Howard, Dhwani Batra, Peter W. Cook, Kara Moser, Adrian Paskey, Jason Caravas, Benjamin Rambo-Martin, Shatavia Morrison, Christopher Gulvick, Scott Sammons, Yvette Unoarumhi, Darlene Wagner, Matthew Schmerer, Christian Bixby, Yihe Wang, Jonathan Schultz, Chirayu Goswami, Russ Hager, Robin Grimwood, Clinton R. Paden, Duncan MacCannell |
| EPI_ISL_2656549, EPI_ISL_2656553, EPI_ISL_2656664, EPI_ISL_2656820 | BCCDC Public Health Laboratory | BCCDC Public Health Laboratory | Prystajecky Natalie, Linda Hoang, Dan Fornika, John Tyson, Shannon Russell, Kim Macdonald, Kimia Kamelian, Ana Pacagnella, Corrinne Ng, Loretta Janz, Robert Azana, Mel Krajden |
| EPI_ISL_2658683, EPI_ISL_2658685 | CA DPH Viral and Rickettsial Disease Laboratory | Chan-Zuckerberg Biohub | CZB Cliahub Consortium |
| EPI_ISL_2658951, EPI_ISL_2658952 | LESP San Luis Potosi | Instituto de Diagnostico y Referencia Epidemiologicos (INDRE) | Claudia Wong-Arambula, Abril Rodriguez-Maldonado, Vanessa Rivero-Arredondo, Ariadna Medina-Benitez, Joaquin Quiroz-Mercado, Sergio Rangel-Guerrero, Natividad Cruz-Ortiz, Tatiana Nunez-Garcia, Gisela Barrera-Badillo, Lucia Hernandez-Rivas, Irma Lopez-Martinez, Ernesto Ramirez-Gonzalez. |
| EPI_ISL_2658953 | LESP Nuevo Leon | Instituto de Diagnostico y Referencia Epidemiologicos (INDRE) | Claudia Wong-Arambula, Abril Rodriguez-Maldonado, Vanessa Rivero-Arredondo, Ariadna Medina-Benitez, Joaquin Quiroz-Mercado, Sergio Rangel-Guerrero, Natividad Cruz-Ortiz, Tatiana Nunez-Garcia, Gisela Barrera-Badillo, Lucia Hernandez-Rivas, Irma Lopez-Martinez, Ernesto Ramirez-Gonzalez. |
| EPI_ISL_2659153, EPI_ISL_2663428, | Genetica Molecular and Subdepartamento de Virologia ISP | Instituto de Salud Publica de Chile | Karen Orostica, Constanza Campano, Barbara Parra, Loredana Arata, Gisselle Barra, Patricia Bustos, Rodrigo Fasce, Javier Tognarelli, Andres Castillo, |

| | | | |
|---|---|---|---|
| EPI_ISL_2663589, EPI_ISL_2663590, EPI_ISL_2663591 | Chile | | Soledad Ulloa, Jorge Fernandez |
| EPI_ISL_2663663, EPI_ISL_2663664, EPI_ISL_2663681, EPI_ISL_2663689, EPI_ISL_2663709, EPI_ISL_2663731, EPI_ISL_2663737, EPI_ISL_2663778 | OHSU Lab Services Molecular Microbiology Lab | Oregon SARS-CoV-2 Genome Sequencing Center | Brendan L. O'Connell, Sonia Acharya, Cierra LaBlanc, Ruth V. Nichols, Alec J. Hirsch, Donna Hansel, Guang Fan, Xuan Qin, Daniel N. Streblow, William B. Messer, Andrew C. Adey, Benjamin N. Bimber, Brian J. O'Roak |
| EPI_ISL_2671571 | Centro de Investigacion Biomedica de Occidente (CIBO) | Unidad de Genomica Avanzada | Consorcio Mexicano de Vigilancia Genomica (CoViGen-Mex). Authors (in alphabetical order): Julio Elias Alvarado-Yaah, Carlos F. Arias, Santiago avila-Rios, Eduardo Becerril-Vargas, Victor Hugo Borja-Aburto, Celia Boukadida, Cristobal Chaidez-Quiroz, Juan Bautista Chale-Dzul, Ricardo Ciria Merce, Andreu Comas-Garcia, Celida Duque Molina, Julissa Enciso-Ibarra, Jose Antonio Enciso-Moreno, Gloria Elena Espinosa-Ayala, Fernando Fontove-Herrera, Daniel Fregoso-Rueda, Victor Eduardo Garcia-Arias, Alejandra Garcia-Gasca, Bruno Gomez-Gil, Jean Pierre Gonzalez, Irvin Gonzalez-Lopez, Concepcion Grajales-Muñiz, Ricardo Grande, Rosa Maria Gutierrez Rios, Jesus Hernandez, Alejandra Hernandez-Teran, Alfredo Herrera-Estrella, Carla Ivon Herrera-Najera, Pavel Isa, Veronica Mata-Haro, Daniel Lira Morales, Susana Lopez, Antonio Loza Roman, Brenda Irasema Maldonado-Meza, Bernardo Martinez-Miguel, Jose Arturo Martinez-Orozco, Celida Martinez- Rodriguez, Margarita Matias-Florentino, Fidencio Mejia-Nepomuceno, Maria Guadalupe de Jesus Mireles-Rivera, Gloria Maria Molina-Salinas, Hector Montoya-Fuentes, , Mario Mujica-Sanchez, Jose Esteban Muñoz-Medina, Jose de Jesus Nuñez-Contreras, Alicia Ocaña-Mondragon, Luis Alberto Ochoa-Carrera, Hector Esteban Paz-Juarez, Marissa Perez-Garcia, Francisco Pulido, Helen Haydee Fernanda Ramirez-Plascencia, Jorge Salas-Hernandez, Angel Gustavo Salas-Lais, Alejandro Sanchez-Flores, Clara Esperanza Santacruz-Tinoco, Maria Guadalupe Santiago-Mauricio, Selene Zarate, Nelly Selem-Mojica, Blanca Taboada, Gloria Vazquez, Joel Armando Vazquez-Perez. |
| EPI_ISL_2671620, EPI_ISL_2671630 | Centro de Investigacion Biomedica del Noreste (CIBIN) | Unidad de Genomica Avanzada | Consorcio Mexicano de Vigilancia Genomica (CoViGen-Mex). Authors (in alphabetical order): Julio Elias Alvarado-Yaah, Carlos F. Arias, Santiago avila-Rios, Eduardo Becerril-Vargas, Victor Hugo Borja-Aburto, Celia Boukadida, Cristobal Chaidez-Quiroz, Juan Bautista Chale-Dzul, Ricardo Ciria Merce, Andreu Comas-Garcia, Celida Duque Molina, Julissa Enciso-Ibarra, Jose Antonio Enciso-Moreno, Gloria Elena Espinosa-Ayala, Fernando Fontove-Herrera, Daniel Fregoso-Rueda, Victor Eduardo Garcia-Arias, Alejandra Garcia-Gasca, Bruno Gomez-Gil, Jean Pierre Gonzalez, Irvin Gonzalez-Lopez, Concepcion Grajales-Muñiz, Ricardo Grande, Rosa Maria Gutierrez Rios, Jesus Hernandez, Alejandra Hernandez-Teran, Alfredo Herrera-Estrella, Carla Ivon Herrera-Najera, Pavel Isa, Veronica Mata-Haro, Daniel Lira Morales, Susana Lopez, Antonio Loza Roman, Brenda Irasema Maldonado-Meza, Bernardo Martinez-Miguel, Jose Arturo Martinez-Orozco, Celida Martinez- Rodriguez, Margarita Matias-Florentino, Fidencio Mejia-Nepomuceno, Maria Guadalupe de Jesus Mireles-Rivera, Gloria Maria Molina-Salinas, Hector Montoya-Fuentes, , Mario Mujica-Sanchez, Jose Esteban Muñoz-Medina, Jose de Jesus Nuñez-Contreras, Alicia Ocaña-Mondragon, Luis Alberto Ochoa-Carrera, Hector Esteban Paz-Juarez, Marissa Perez-Garcia, Francisco Pulido, Helen Haydee Fernanda Ramirez-Plascencia, Jorge Salas-Hernandez, Angel Gustavo Salas-Lais, Alejandro Sanchez-Flores, Clara Esperanza Santacruz-Tinoco, Maria Guadalupe Santiago-Mauricio, Selene Zarate, Nelly Selem-Mojica, Blanca Taboada, Gloria Vazquez, Joel Armando Vazquez-Perez. |
| EPI_ISL_2671701 | Unidad de Investigacion Biomedica de Zacatecas (UIBZ) | Unidad de Genomica Avanzada | Consorcio Mexicano de Vigilancia Genomica (CoViGen-Mex). Authors (in alphabetical order): Julio Elias Alvarado-Yaah, Carlos F. Arias, Santiago avila-Rios, Eduardo Becerril-Vargas, Victor Hugo Borja-Aburto, Celia Boukadida, Cristobal Chaidez-Quiroz, Juan Bautista Chale-Dzul, Ricardo Ciria Merce, Andreu Comas-Garcia, Celida Duque Molina, Julissa Enciso-Ibarra, Jose Antonio Enciso-Moreno, Gloria Elena Espinosa-Ayala, Fernando Fontove-Herrera, Daniel Fregoso-Rueda, Victor Eduardo Garcia-Arias, Alejandra Garcia-Gasca, Bruno Gomez-Gil, Jean Pierre Gonzalez, Irvin Gonzalez-Lopez, Concepcion Grajales-Muñiz, Ricardo Grande, Rosa Maria Gutierrez Rios, Jesus Hernandez, Alejandra Hernandez-Teran, Alfredo Herrera-Estrella, Carla Ivon Herrera-Najera, Pavel Isa, Veronica Mata-Haro, Daniel Lira Morales, Susana Lopez, Antonio Loza Roman, Brenda Irasema Maldonado-Meza, Bernardo Martinez-Miguel, Jose Arturo Martinez-Orozco, Celida Martinez- Rodriguez, Margarita Matias-Florentino, Fidencio Mejia-Nepomuceno, Maria Guadalupe de Jesus Mireles-Rivera, Gloria Maria Molina-Salinas, Hector Montoya-Fuentes, , Mario Mujica-Sanchez, Jose Esteban Muñoz-Medina, Jose de Jesus Nuñez-Contreras, Alicia Ocaña-Mondragon, Luis Alberto Ochoa-Carrera, Hector Esteban Paz-Juarez, Marissa Perez-Garcia, Francisco Pulido, Helen Haydee Fernanda Ramirez-Plascencia, Jorge Salas-Hernandez, Angel Gustavo Salas-Lais, Alejandro Sanchez-Flores, Clara Esperanza Santacruz-Tinoco, Maria Guadalupe Santiago-Mauricio, Selene Zarate, Nelly Selem-Mojica, Blanca Taboada, Gloria Vazquez, Joel Armando Vazquez-Perez. |
| EPI_ISL_2671747 | Unidad de Investigacion Medica de Yucatan (UIMY) | Unidad de Genomica Avanzada | Consorcio Mexicano de Vigilancia Genomica (CoViGen-Mex). Authors (in alphabetical order): Julio Elias Alvarado-Yaah, Carlos F. Arias, Santiago avila-Rios, Eduardo Becerril-Vargas, Victor Hugo Borja-Aburto, Celia Boukadida, Cristobal Chaidez-Quiroz, Juan Bautista Chale-Dzul, Ricardo Ciria Merce, Andreu Comas-Garcia, Celida Duque Molina, Julissa Enciso-Ibarra, Jose Antonio Enciso-Moreno, Gloria Elena Espinosa-Ayala, Fernando Fontove-Herrera, Daniel Fregoso-Rueda, Victor Eduardo Garcia-Arias, Alejandra Garcia-Gasca, Bruno Gomez-Gil, Jean Pierre Gonzalez, Irvin Gonzalez-Lopez, Concepcion Grajales-Muñiz, Ricardo Grande, Rosa Maria Gutierrez Rios, Jesus Hernandez, Alejandra Hernandez-Teran, Alfredo Herrera-Estrella, Carla Ivon Herrera-Najera, Pavel Isa, Veronica Mata-Haro, Daniel Lira Morales, Susana Lopez, Antonio Loza Roman, Brenda Irasema Maldonado-Meza, Bernardo Martinez-Miguel, Jose Arturo Martinez-Orozco, Celida Martinez- Rodriguez, Margarita Matias-Florentino, Fidencio Mejia-Nepomuceno, Maria Guadalupe de Jesus Mireles-Rivera, Gloria Maria Molina-Salinas, Hector Montoya-Fuentes, , Mario Mujica-Sanchez, Jose Esteban Muñoz-Medina, Jose de Jesus Nuñez-Contreras, Alicia Ocaña-Mondragon, Luis Alberto Ochoa-Carrera, Hector Esteban Paz-Juarez, Marissa Perez-Garcia, Francisco Pulido, Helen Haydee Fernanda Ramirez-Plascencia, Jorge Salas-Hernandez, Angel Gustavo Salas-Lais, Alejandro Sanchez-Flores, Clara Esperanza Santacruz-Tinoco, Maria Guadalupe Santiago-Mauricio, Selene Zarate, Nelly Selem-Mojica, Blanca Taboada, Gloria Vazquez, Joel Armando Vazquez-Perez. |
| EPI_ISL_2678983, EPI_ISL_2679276 | Utah Public Health Laboratory | Utah Public Health Laboratory | Erin L. Young, Kelly F. Oakeson, Tara Gallagher |
| EPI_ISL_2680485 | Humboldt County Public Health Laboratory | Humboldt County Public Health Laboratory | Jeremy Corrigan |
| EPI_ISL_2680922 | Laboratorios Delia Barraza | Laboratorio nacional-LANIIA-CIAD | Consorcio Mexicano de Vigilancia Genómica (CoViGen-Mex). Authors (in alphabetical order): Julio Elias Alvarado-Yaah, Carlos F. Arias, Santiago Ávila-Ríos, Eduardo Becerril-Vargas, Víctor Hugo Borja-Aburto, Celia Boukadida, Cristóbal Cháidez-Quiróz, Juan Bautista Chale-Dzul, Ricardo Ciria Merce, Andreu Comas-García, Célida Duque Molina, Julissa Enciso-Ibarra, José Antonio Enciso-Moreno, Gloria Elena Espinosa-Ayala, Fernando Fontove-Herrera, Daniel Fregoso-Rueda, Victor Eduardo García-Arias, Alejandra García-Gasca, Bruno Gómez-Gil, Jean Pierre González, Irvin González-López, Concepción Grajales-Muñiz, Ricardo Grande, Rosa María Gutiérrez Rios, Jesús Hernández, Alejandra Hernández-Terán, Alfredo Herrera-Estrella, Carla Ivón Herrera-Najera, Pavel Isa, Verónica Mata-Haro, Daniel Lira Morales, Susana Lopez, Antonio Loza Román, Brenda Irasema Maldonado-Meza, Bernardo Martínez-Miguel, José Arturo Martínez-Orozco, Célida Martínez- Rodríguez, Margarita Matías-Florentino, Fidencio Mejía-Nepomuceno, María Guadalupe de Jesús Mireles-Rivera, Gloria María Molina-Salinas, Hector Montoya-Fuentes, , Mario Mújica-Sánchez, José Esteban Muñoz-Medina, José de Jesús Nuñez-Contreras, Alicia Ocaña-Mondragón, Luis Alberto Ochoa-Carrera, Hector Esteban Paz-Juárez, Marissa Perez-Garcia, Francisco Pulido, Helen Haydee Fernanda Ramírez-Plascencia, Jorge Salas-Hernández, Angel Gustavo Salas-Lais, Alejandro Sánchez-Flores, Clara Esperanza Santacruz-Tinoco, María Guadalupe Santiago-Mauricio, Selene Zárate, Nelly Sélem-Mojica, Blanca Taboada, Gloria Vazquez, Joel Armando Vázquez-Pérez. Laboratorios Delia Barraza: María Delia Barraza Sámano, Claudia Soto Félix, Melissa García Angulo. |
| EPI_ISL_2681236 | Unidad de Investigación Médica de Yucatán (UIMY) | Instituto de Biotecnología de la UNAM | Consorcio Mexicano de Vigilancia Genómica (CoViGen-Mex). Authors (in alphabetical order): Julio Elias Alvarado-Yaah, Carlos F. Arias, Santiago Ávila-Ríos, Eduardo Becerril-Vargas, Víctor Hugo Borja-Aburto, Celia Boukadida, Cristóbal Cháidez-Quiróz, Juan Bautista Chale-Dzul, Ricardo Ciria Merce, Andreu Comas-García, Célida Duque Molina, Julissa Enciso-Ibarra, José Antonio Enciso-Moreno, Gloria Elena Espinosa-Ayala, Fernando Fontove-Herrera, Daniel Fregoso-Rueda, Victor Eduardo García-Arias, Alejandra García-Gasca, Bruno Gómez-Gil, Jean Pierre González, Irvin González-López, Concepción Grajales-Muñiz, Ricardo Grande, Rosa María Gutiérrez Rios, Jesús Hernández, Alejandra Hernández-Terán, Alfredo Herrera-Estrella, Carla Ivón Herrera-Najera, Pavel Isa, Verónica Mata-Haro, Daniel Lira Morales, Susana Lopez, Antonio Loza Román, Brenda Irasema Maldonado-Meza, Bernardo Martínez-Miguel, José Arturo Martínez-Orozco, Célida Martínez- Rodríguez, Margarita Matías-Florentino, Fidencio Mejía-Nepomuceno, María Guadalupe de Jesús Mireles-Rivera, Gloria María Molina-Salinas, Hector Montoya-Fuentes, , Mario Mújica-Sánchez, José Esteban Muñoz-Medina, José de Jesús Nuñez-Contreras, Alicia Ocaña-Mondragón, Luis Alberto Ochoa-Carrera, Hector Esteban Paz-Juárez, Marissa Perez-Garcia, Francisco Pulido, Helen Haydee Fernanda Ramírez-Plascencia, Jorge Salas-Hernández, Angel Gustavo Salas-Lais, Alejandro Sánchez-Flores, Clara Esperanza Santacruz-Tinoco, María Guadalupe Santiago-Mauricio, Selene Zárate, Nelly Sélem-Mojica, Blanca Taboada, Gloria Vazquez, Joel Armando Vázquez-Pérez. |
| EPI_ISL_2681252 | Laboratorio Central de Epidemiología (LCE) | Instituto de Biotecnología de la UNAM | Consorcio Mexicano de Vigilancia Genómica (CoViGen-Mex). Authors (in alphabetical order): Julio Elias Alvarado-Yaah, Carlos F. Arias, Santiago Ávila-Ríos, Eduardo Becerril-Vargas, Víctor Hugo Borja-Aburto, Celia Boukadida, Cristóbal Cháidez-Quiróz, Juan Bautista Chale-Dzul, Ricardo Ciria |

| | | | |
|---|---|---|---|
| | | | Merce, Andreu Comas-García, Célida Duque Molina, Julissa Enciso-Ibarra, José Antonio Enciso-Moreno, Gloria Elena Espinosa-Ayala, Fernando Fontove-Herrera, Daniel Fregoso-Rueda, Victor Eduardo García-Arias, Alejandra García-Gasca, Bruno Gómez-Gil, Jean Pierre González, Irvin González-López, Concepción Grajales-Muñiz, Ricardo Grande, Rosa María Gutiérrez Rios, Jesús Hernández, Alejandra Hernández-Terán, Alfredo Herrera-Estrella, Carla Ivón Herrera-Najera, Pavel Isa, Verónica Mata-Haro, Daniel Lira Morales, Susana Lopez, Antonio Loza Román, Brenda Irasema Maldonado-Meza, Bernardo Martínez-Miguel, José Arturo Martínez-Orozco, Célida Martínez- Rodríguez, Margarita Matías-Florentino, Fidencio Mejía-Nepomuceno, María Guadalupe de Jesús Mireles-Rivera, Gloria María Molina-Salinas, Hector Montoya-Fuentes, , Mario Mújica-Sánchez, José Esteban Muñoz-Medina, José de Jesús Nuñez-Contreras, Alicia Ocaña-Mondragón, Luis Alberto Ochoa-Carrera, Hector Esteban Paz-Juárez, Marissa Perez-Garcia, Francisco Pulido, Helen Haydee Fernanda Ramírez-Plascencia, Jorge Salas-Hernández, Angel Gustavo Salas-Lais, Alejandro Sánchez-Flores, Clara Esperanza Santacruz-Tinoco, María Guadalupe Santiago-Mauricio, Selene Zárate, Nelly Sélem-Mojica, Blanca Taboada, Gloria Vazquez, Joel Armando Vázquez-Pérez. |
| EPI_ISL_2681271 | Centro de Investigación Biomédica del Noreste (CIBIN) | Instituto de Biotecnología de la UNAM | Consorcio Mexicano de Vigilancia Genómica (CoViGen-Mex). Authors (in alphabetical order): Julio Elias Alvarado-Yaah, Carlos F. Arias, Santiago Ávila-Ríos, Eduardo Becerril-Vargas, Víctor Hugo Borja-Aburto, Celia Boukadida, Cristóbal Cháidez-Quiróz, Juan Bautista Chale-Dzul, Ricardo Ciria Merce, Andreu Comas-García, Célida Duque Molina, Julissa Enciso-Ibarra, José Antonio Enciso-Moreno, Gloria Elena Espinosa-Ayala, Fernando Fontove-Herrera, Daniel Fregoso-Rueda, Victor Eduardo García-Arias, Alejandra García-Gasca, Bruno Gómez-Gil, Jean Pierre González, Irvin González-López, Concepción Grajales-Muñiz, Ricardo Grande, Rosa María Gutiérrez Rios, Jesús Hernández, Alejandra Hernández-Terán, Alfredo Herrera-Estrella, Carla Ivón Herrera-Najera, Pavel Isa, Verónica Mata-Haro, Daniel Lira Morales, Susana Lopez, Antonio Loza Román, Brenda Irasema Maldonado-Meza, Bernardo Martínez-Miguel, José Arturo Martínez-Orozco, Célida Martínez- Rodríguez, Margarita Matías-Florentino, Fidencio Mejía-Nepomuceno, María Guadalupe de Jesús Mireles-Rivera, Gloria María Molina-Salinas, Hector Montoya-Fuentes, , Mario Mújica-Sánchez, José Esteban Muñoz-Medina, José de Jesús Nuñez-Contreras, Alicia Ocaña-Mondragón, Luis Alberto Ochoa-Carrera, Hector Esteban Paz-Juárez, Marissa Perez-Garcia, Francisco Pulido, Helen Haydee Fernanda Ramírez-Plascencia, Jorge Salas-Hernández, Angel Gustavo Salas-Lais, Alejandro Sánchez-Flores, Clara Esperanza Santacruz-Tinoco, María Guadalupe Santiago-Mauricio, Selene Zárate, Nelly Sélem-Mojica, Blanca Taboada, Gloria Vazquez, Joel Armando Vázquez-Pérez. |
| EPI_ISL_2686407, EPI_ISL_2686419, EPI_ISL_2686426, EPI_ISL_2686597 | Fulgent Genetics | Centers for Disease Control and Prevention Division of Viral Diseases, Pathogen Discovery | Dakota Howard, Dhwani Batra, Peter W. Cook, Kara Moser, Adrian Paskey, Jason Caravas, Benjamin Rambo-Martin, Shatavia Morrison, Christopher Gulvick, Scott Sammons, Yvette Unoarumhi, Darlene Wagner, Matthew Schmerer, Harry Gao, Mickey Li, John Gao, Joseph Fierro, Benafsh Sapra, Becky Tsai, Yan Meng, Doreen Ng, James Xie, Clinton R. Paden, Duncan MacCannell |
| EPI_ISL_2686938, EPI_ISL_2687004, EPI_ISL_2687041, EPI_ISL_2687042 | Aegis Sciences Corporation | Centers for Disease Control and Prevention Division of Viral Diseases, Pathogen Discovery | Dakota Howard, Dhwani Batra, Peter W. Cook, Kara Moser, Adrian Paskey, Jason Caravas, Benjamin Rambo-Martin, Shatavia Morrison, Christopher Gulvick, Scott Sammons, Yvette Unoarumhi, Darlene Wagner, Matthew Schmerer, Cyndi Clark, Patrick Campbell, Rob Case, Vikramsinha Ghorpade, Holly Houdeshell, Ola Kvalvaag, Dillon Nall, Ethan Sanders, Alec Vest, Shaun Westlund, Matthew Hardison, Clinton R. Paden, Duncan MacCannell |
| EPI_ISL_2689673 | Nebraska Public Health Laboratory | NPHL COVID-19 Response Team | NPHL COVID-19 Response Team |
| EPI_ISL_2689985 | OR State PHL-Virology/Immunology Section | Centers for Disease Control and Prevention Division of Viral Diseases, Pathogen Discovery | Mili Sheth, Sarah Nobles, Jasmine Padilla, Mark Burroughs, Shoshona Le, Katie Dillon, Peter Cook, Clinton R. Paden, Dhwani Batra, Krista Queen, Kristen Knipe, Dakota Howard, Yvette Unoarumhi, Darlene Wagner, Matthew Schmerer, Ben L. Rambo-Martin, Kristine Lacek, Sam Shepard, Alison Laufer Halpin, Dave Wentworth, Vivien Dugan, Suxiang Tong, Justin Lee |
| EPI_ISL_2690075 | ID Bureau of Laboratories | Centers for Disease Control and Prevention Division of Viral Diseases, Pathogen Discovery | Mili Sheth, Sarah Nobles, Jasmine Padilla, Mark Burroughs, Shoshona Le, Katie Dillon, Peter Cook, Clinton R. Paden, Dhwani Batra, Krista Queen, Kristen Knipe, Dakota Howard, Yvette Unoarumhi, Darlene Wagner, Matthew Schmerer, Ben L. Rambo-Martin, Kristine Lacek, Sam Shepard, Alison Laufer Halpin, Dave Wentworth, Vivien Dugan, Suxiang Tong, Justin Lee |
| EPI_ISL_2690111 | WA State Department of Health | Centers for Disease Control and Prevention Division of Viral Diseases, Pathogen Discovery | Mili Sheth, Sarah Nobles, Jasmine Padilla, Mark Burroughs, Shoshona Le, Katie Dillon, Peter Cook, Clinton R. Paden, Dhwani Batra, Krista Queen, Kristen Knipe, Dakota Howard, Yvette Unoarumhi, Darlene Wagner, Matthew Schmerer, Ben L. Rambo-Martin, Kristine Lacek, Sam Shepard, Alison Laufer Halpin, Dave Wentworth, Vivien Dugan, Suxiang Tong, Justin Lee |
| EPI_ISL_2692208, EPI_ISL_2692220, EPI_ISL_2692221, EPI_ISL_2692222, EPI_ISL_2692223, EPI_ISL_2692224, EPI_ISL_2692225 | Seattle Flu Study | Seattle Flu Study | Deborah A. Nickerson, Chris D. Frazar, Jover Lee, Benjamin Pelle, Erica Ryke, Matthew Richardson, Amanda Adler, Elisabeth Brandstetter, Peter D. Han, Kairsten Fay, Misja Ilcisin, Kirsten Lacombe, Thomas R. Sibley, Melissa Truong, Caitlin R. Wolf, Karen Cowgill, Stephanie Schrag, Jeff Duchin, Michael Boeckh, Janet A. Englund, Michael Famulare, Barry R. Lutz, Mark J. Rieder, Lea M. Starita, Matthew Thompson, Helen Y. Chu, Trevor Bedford, Jay Shendure |
| EPI_ISL_2692243, EPI_ISL_2692266, EPI_ISL_2692284 | Atlas Genomics | Seattle Flu Study | Deborah A. Nickerson, Chris D. Frazar, Jover Lee, Benjamin Pelle, Erica Ryke, Matthew Richardson, Amanda Adler, Elisabeth Brandstetter, Peter D. Han, Kairsten Fay, Misja Ilcisin, Kirsten Lacombe, Thomas R. Sibley, Melissa Truong, Caitlin R. Wolf, Romesh Gautom, Geoff Melly, Brian Hiatt, Philip Dykema, Scott Lindquist, Michael Boeckh, Janet A. Englund, Michael Famulare, Barry R. Lutz, Mark J. Rieder, Lea M. Starita, Matthew Thompson, Helen Y. Chu, Jay Shendure, Trevor Bedford |
| EPI_ISL_2692309 | Seattle Flu Study | Seattle Flu Study | Deborah A. Nickerson, Chris D. Frazar, Jover Lee, Benjamin Pelle, Erica Ryke, Matthew Richardson, Amanda Adler, Elisabeth Brandstetter, Peter D. Han, Kairsten Fay, Misja Ilcisin, Kirsten Lacombe, Thomas R. Sibley, Melissa Truong, Caitlin R. Wolf, Karen Cowgill, Stephanie Schrag, Jeff Duchin, Michael Boeckh, Janet A. Englund, Michael Famulare, Barry R. Lutz, Mark J. Rieder, Lea M. Starita, Matthew Thompson, Helen Y. Chu, Trevor Bedford, Jay Shendure |
| EPI_ISL_2692395, EPI_ISL_2692398 | Northwest Laboratory | Seattle Flu Study | Deborah A. Nickerson, Chris D. Frazar, Jover Lee, Benjamin Pelle, Erica Ryke, Matthew Richardson, Amanda Adler, Elisabeth Brandstetter, Peter D. Han, Kairsten Fay, Misja Ilcisin, Kirsten Lacombe, Thomas R. Sibley, Melissa Truong, Caitlin R. Wolf, Romesh Gautom, Geoff Melly, Brian Hiatt, Philip Dykema, Scott Lindquist, Michael Boeckh, Janet A. Englund, Michael Famulare, Barry R. Lutz, Mark J. Rieder, Lea M. Starita, Matthew Thompson, Helen Y. Chu, Jay Shendure, Trevor Bedford |
| EPI_ISL_2692424, EPI_ISL_2692432 | Incyte Diagnostics Spokane | Seattle Flu Study | Deborah A. Nickerson, Chris D. Frazar, Jover Lee, Benjamin Pelle, Erica Ryke, Matthew Richardson, Amanda Adler, Elisabeth Brandstetter, Peter D. Han, Kairsten Fay, Misja Ilcisin, Kirsten Lacombe, Thomas R. Sibley, Melissa Truong, Caitlin R. Wolf, Romesh Gautom, Geoff Melly, Brian Hiatt, Philip Dykema, Scott Lindquist, Michael Boeckh, Janet A. Englund, Michael Famulare, Barry R. Lutz, Mark J. Rieder, Lea M. Starita, Matthew Thompson, Helen Y. Chu, Jay Shendure, Trevor Bedford |
| EPI_ISL_2692462, EPI_ISL_2692464, EPI_ISL_2692466, EPI_ISL_2692473, EPI_ISL_2692480, EPI_ISL_2692485 | Atlas Genomics | Seattle Flu Study | Deborah A. Nickerson, Chris D. Frazar, Jover Lee, Benjamin Pelle, Erica Ryke, Matthew Richardson, Amanda Adler, Elisabeth Brandstetter, Peter D. Han, Kairsten Fay, Misja Ilcisin, Kirsten Lacombe, Thomas R. Sibley, Melissa Truong, Caitlin R. Wolf, Romesh Gautom, Geoff Melly, Brian Hiatt, Philip Dykema, Scott Lindquist, Michael Boeckh, Janet A. Englund, Michael Famulare, Barry R. Lutz, Mark J. Rieder, Lea M. Starita, Matthew Thompson, Helen Y. Chu, Jay Shendure, Trevor Bedford |
| EPI_ISL_2692515 | Overlake Hospital | Seattle Flu Study | Deborah A. Nickerson, Chris D. Frazar, Jover Lee, Benjamin Pelle, Erica Ryke, Matthew Richardson, Amanda Adler, Elisabeth Brandstetter, Peter D. Han, Kairsten Fay, Misja Ilcisin, Kirsten Lacombe, Thomas R. Sibley, Melissa Truong, Caitlin R. Wolf, Romesh Gautom, Geoff Melly, Brian Hiatt, Philip Dykema, Scott Lindquist, Michael Boeckh, Janet A. Englund, Michael Famulare, Barry R. Lutz, Mark J. Rieder, Lea M. Starita, Matthew Thompson, Helen Y. Chu, Jay Shendure, Trevor Bedford |
| EPI_ISL_2693294, EPI_ISL_2693298, EPI_ISL_2693305 | UW Virology Lab | UW Virology Lab | Pavitra Roychoudhury, Hong Xie, Lasata Shrestha, Tien V. Nguyen, Shah Mohamed Bakhash, Michelle Lin, Noah R. Baker, Ricardo Perez, Sean Ellis, Nathan Breit, Robert J. Livingston, Meei-Li Huang, Keith R Jerome, Patrick Mathias, Alexander Greninger |
| EPI_ISL_2693356, EPI_ISL_2693357 | ISDH | Quantigen Biosciences | Paul J. Childress |
| EPI_ISL_2693699, EPI_ISL_2693700, EPI_ISL_2693701, EPI_ISL_2693727, EPI_ISL_2693733, EPI_ISL_2693750, EPI_ISL_2693785, EPI_ISL_2693800 | UW Virology Lab | UW Virology Lab | Pavitra Roychoudhury, Hong Xie, Lasata Shrestha, Shah Mohamed Bakhash, Tien V. Nguyen, Noah R. Baker, Sean Ellis, Meei-Li Huang, Keith R Jerome, Alexander Greninger |
| EPI_ISL_2696869 | Laboratory, Gravity Diagnostics | Laboratory, Gravity Diagnostics | Diagnostics,G. |

EPI_ISL_2698120, EPI_ISL_2698128, EPI_ISL_2698143, EPI_ISL_2698145, EPI_ISL_2698195, EPI_ISL_2698206, EPI_ISL_2698217, EPI_ISL_2698244, EPI_ISL_2698255, EPI_ISL_2698263, EPI_ISL_2698311, EPI_ISL_2698315, EPI_ISL_2698454, EPI_ISL_2698456, EPI_ISL_2698459, EPI_ISL_2698485, EPI_ISL_2698497, EPI_ISL_2698508, EPI_ISL_2698569, EPI_ISL_2698573, EPI_ISL_2699233, EPI_ISL_2699252, EPI_ISL_2699254, EPI_ISL_2699264, EPI_ISL_2699270, EPI_ISL_2699280, EPI_ISL_2699309, EPI_ISL_2699323, EPI_ISL_2699343, EPI_ISL_2699349, EPI_ISL_2699360, EPI_ISL_2699373, EPI_ISL_2699382, EPI_ISL_2709029, EPI_ISL_2709040, EPI_ISL_2709071, EPI_ISL_2709090, EPI_ISL_2709118, EPI_ISL_2709134, EPI_ISL_2709148, EPI_ISL_2709156, EPI_ISL_2709185

| | | | |
|---|---|---|---|
| see above | Public Health Laboratory, Minnesota Department of Health | University of Minnesota Genomics Center | Daryl M. Gohl, John Garbe, Jaquelyn Kuriger-Laber, Corbin Dirkx, and Sean Wang |
| EPI_ISL_2712539 | Sharp HealthCare Laboratory | Andersen lab at Scripps Research | SEARCH Alliance San Diego with Aaron Harding, Jacquelyn Berumen, Cathy Woerle, Liam McGinnis, Art Mendoza, Omid Bakhtar |
| EPI_ISL_2712831 | San Diego County Public Health Laboratory | Andersen lab at Scripps Research | SEARCH Alliance San Diego with Ashleigh Murphy, Jovan Shephard, Brett Austin |
| EPI_ISL_2714923, EPI_ISL_2714936 | Incyte Diagnostics Spokane | Seattle Flu Study | Deborah A. Nickerson, Chris D. Frazar, Jover Lee, Benjamin Pelle, Erica Ryke, Matthew Richardson, Amanda Adler, Elisabeth Brandstetter, Peter D. Han, Kairsten Fay, Misja Ilcisin, Kirsten Lacombe, Thomas R. Sibley, Melissa Truong, Caitlin R. Wolf, Romesh Gautom, Geoff Melly, Brian Hiatt, Philip Dykema, Scott Lindquist, Michael Boeckh, Janet A. Englund, Michael Famulare, Barry R. Lutz, Mark J. Rieder, Lea M. Starita, Matthew Thompson, Helen Y. Chu, Jay Shendure, Trevor Bedford |
| EPI_ISL_2714947, EPI_ISL_2714950, EPI_ISL_2714961, EPI_ISL_2714979, EPI_ISL_2714984, EPI_ISL_2714985, EPI_ISL_2715000, EPI_ISL_2715001, EPI_ISL_2715006 | Interpath Laboratory | Seattle Flu Study | Deborah A. Nickerson, Chris D. Frazar, Jover Lee, Benjamin Pelle, Erica Ryke, Matthew Richardson, Amanda Adler, Elisabeth Brandstetter, Peter D. Han, Kairsten Fay, Misja Ilcisin, Kirsten Lacombe, Thomas R. Sibley, Melissa Truong, Caitlin R. Wolf, Romesh Gautom, Geoff Melly, Brian Hiatt, Philip Dykema, Scott Lindquist, Michael Boeckh, Janet A. Englund, Michael Famulare, Barry R. Lutz, Mark J. Rieder, Lea M. Starita, Matthew Thompson, Helen Y. Chu, Jay Shendure, Trevor Bedford |
| EPI_ISL_2715089 | Incyte Diagnostics Spokane | Seattle Flu Study | Deborah A. Nickerson, Chris D. Frazar, Jover Lee, Benjamin Pelle, Erica Ryke, Matthew Richardson, Amanda Adler, Elisabeth Brandstetter, Peter D. Han, Kairsten Fay, Misja Ilcisin, Kirsten Lacombe, Thomas R. Sibley, Melissa Truong, Caitlin R. Wolf, Romesh Gautom, Geoff Melly, Brian Hiatt, Philip Dykema, Scott Lindquist, Michael Boeckh, Janet A. Englund, Michael Famulare, Barry R. Lutz, Mark J. Rieder, Lea M. Starita, Matthew Thompson, Helen Y. Chu, Jay Shendure, Trevor Bedford |
| EPI_ISL_2715203 | Atlas Genomics | Seattle Flu Study | Deborah A. Nickerson, Chris D. Frazar, Jover Lee, Benjamin Pelle, Erica Ryke, Matthew Richardson, Amanda Adler, Elisabeth Brandstetter, Peter D. Han, Kairsten Fay, Misja Ilcisin, Kirsten Lacombe, Thomas R. Sibley, Melissa Truong, Caitlin R. Wolf, Romesh Gautom, Geoff Melly, Brian Hiatt, Philip Dykema, Scott Lindquist, Michael Boeckh, Janet A. Englund, Michael Famulare, Barry R. Lutz, Mark J. Rieder, Lea M. Starita, Matthew Thompson, Helen Y. Chu, Jay Shendure, Trevor Bedford |
| EPI_ISL_2720367, EPI_ISL_2720671, EPI_ISL_2720750, EPI_ISL_2721512 | BCCDC Public Health Laboratory | BCCDC Public Health Laboratory | Prystajecky Natalie, Linda Hoang, Dan Fornika, John Tyson, Shannon Russell, Kim Macdonald, Kimia Kamelian, Ana Pacagnella, Corrinne Ng, Loretta Janz, Robert Azana, Mel Krajden |
| EPI_ISL_2724528, EPI_ISL_2724541, EPI_ISL_2724566, EPI_ISL_2724591, EPI_ISL_2724594, EPI_ISL_2724595, EPI_ISL_2724596, EPI_ISL_2724604, EPI_ISL_2724616, EPI_ISL_2724623, EPI_ISL_2724625, EPI_ISL_2724626, EPI_ISL_2724632, EPI_ISL_2724634, EPI_ISL_2724638, EPI_ISL_2724640, EPI_ISL_2724648, EPI_ISL_2724653, EPI_ISL_2724654, EPI_ISL_2724662, EPI_ISL_2724663, EPI_ISL_2724668, EPI_ISL_2724674, EPI_ISL_2724675, EPI_ISL_2724682, EPI_ISL_2724688, EPI_ISL_2724702, EPI_ISL_2724706, EPI_ISL_2724714, EPI_ISL_2724716, EPI_ISL_2724726, EPI_ISL_2724736, EPI_ISL_2724747, EPI_ISL_2724758, EPI_ISL_2724765, EPI_ISL_2724802, EPI_ISL_2724804, EPI_ISL_2724808, EPI_ISL_2724809, EPI_ISL_2724810, EPI_ISL_2724817, EPI_ISL_2724821, EPI_ISL_2724827, EPI_ISL_2724838, EPI_ISL_2724840, EPI_ISL_2724875, EPI_ISL_2724895, EPI_ISL_2724907, EPI_ISL_2724912 | | | |
| see above | State Laboratories Division, Hawaii State Department of Health | State Laboratories Division, Hawaii State Department of Health | Pamela O'Brien, Drew Kuwazaki, Ayana Garnet, Razvan Sultana, Edward Desmond |
| EPI_ISL_2735972, EPI_ISL_2736001 | Incyte Diagnostics Spokane | Seattle Flu Study | Deborah A. Nickerson, Chris D. Frazar, Jover Lee, Benjamin Pelle, Erica Ryke, Matthew Richardson, Amanda Adler, Elisabeth Brandstetter, Peter D. Han, Kairsten Fay, Misja Ilcisin, Kirsten Lacombe, Thomas R. Sibley, Melissa Truong, Caitlin R. Wolf, Romesh Gautom, Geoff Melly, Brian Hiatt, Philip Dykema, Scott Lindquist, Michael Boeckh, Janet A. Englund, Michael Famulare, Barry R. Lutz, Mark J. Rieder, Lea M. Starita, Matthew Thompson, Helen Y. Chu, Jay Shendure, Trevor Bedford |
| EPI_ISL_2736825 | LESP Michoacan | Instituto de Diagnostico y Referencia Epidemiologicos (INDRE) | Claudia Wong-Arambula, Abril Rodriguez-Maldonado, Vanessa Rivero-Arredondo, Ariadna Medina-Benitez, Joaquin Quiroz-Mercado, Sergio Rangel-Guerrero, Natividad Cruz-Ortiz, Tatiana Nunez-Garcia, Gisela Barrera-Badillo, Lucia Hernandez-Rivas, Irma Lopez-Martinez, Ernesto Ramirez-Gonzalez. |
| EPI_ISL_2736827 | LESP Guanajuato | Instituto de Diagnostico y Referencia Epidemiologicos (INDRE) | Claudia Wong-Arambula, Abril Rodriguez-Maldonado, Vanessa Rivero-Arredondo, Ariadna Medina-Benitez, Joaquin Quiroz-Mercado, Sergio Rangel-Guerrero, Natividad Cruz-Ortiz, Tatiana Nunez-Garcia, Gisela Barrera-Badillo, Lucia Hernandez-Rivas, Irma Lopez-Martinez, Ernesto Ramirez-Gonzalez. |
| EPI_ISL_2736829 | LESP Coahuila | Instituto de Diagnostico y Referencia Epidemiologicos (INDRE) | Claudia Wong-Arambula, Abril Rodriguez-Maldonado, Vanessa Rivero-Arredondo, Ariadna Medina-Benitez, Joaquin Quiroz-Mercado, Sergio Rangel-Guerrero, Natividad Cruz-Ortiz, Tatiana Nunez-Garcia, Gisela Barrera-Badillo, Lucia Hernandez-Rivas, Irma Lopez-Martinez, Ernesto Ramirez-Gonzalez. |
| EPI_ISL_2736832 | LESP Baja California | Instituto de Diagnostico y Referencia Epidemiologicos (INDRE) | Claudia Wong-Arambula, Abril Rodriguez-Maldonado, Vanessa Rivero-Arredondo, Ariadna Medina-Benitez, Joaquin Quiroz-Mercado, Sergio Rangel-Guerrero, Natividad Cruz-Ortiz, Tatiana Nunez-Garcia, Gisela Barrera-Badillo, Lucia Hernandez-Rivas, Irma Lopez-Martinez, Ernesto Ramirez-Gonzalez. |
| EPI_ISL_2736834 | LESP Michoacan | Instituto de Diagnostico y Referencia Epidemiologicos (INDRE) | Claudia Wong-Arambula, Abril Rodriguez-Maldonado, Vanessa Rivero-Arredondo, Ariadna Medina-Benitez, Joaquin Quiroz-Mercado, Sergio Rangel-Guerrero, Natividad Cruz-Ortiz, Tatiana Nunez-Garcia, Gisela Barrera-Badillo, Lucia Hernandez-Rivas, Irma Lopez-Martinez, Ernesto Ramirez-Gonzalez. |
| EPI_ISL_2756322 | Genetica Molecular and Subdepartamento de Virologia ISP Chile | Instituto de Salud Publica de Chile | Karen Orostica, Constanza Campano, Barbara Parra, Loredana Arata, Gisselle Barra, Patricia Bustos, Rodrigo Fasce, Javier Tognarelli, Andres Castillo, Soledad Ulloa, Jorge Fernandez |
| EPI_ISL_2759228, EPI_ISL_2759231, EPI_ISL_2759243, EPI_ISL_2759331, EPI_ISL_2759334 | Population Medicine and Diagnostic Sciences, Cornell University | Population Medicine and Diagnostic Sciences, Cornell University | Mitchell,P.K., Caserta,L.C., Cronk,B.D., Anderson,R.R., Laverack,M., Venugopalan,R., Plocharczyk,E., Diel,D.G. |

### supp. Table S6: Gamma

We gratefully acknowledge the following Authors from the Originating laboratories responsible for obtaining the specimens, as well as the Submitting laboratories where the genome data were generated and shared via GISAID, on which this research is based.

All Submitters of data may be contacted directly via www.gisaid.org

Authors are sorted alphabetically.

| Accession ID | Originating Laboratory | Submitting Laboratory | Authors |
|---|---|---|---|
| EPI_ISL_2466750 | Nebraska Public Health Laboratory | NPHL COVID-19 Response Team | NPHL COVID-19 Response Team |
| EPI_ISL_2466754, EPI_ISL_2466757 | Wichita State University - Molecular Diagnostics Lab | Kansas Health and Environmental Lab | Mike Grose, Katherine Wiggins, Jonathan Barnell, Ben Olsen, and Phil Adam |
| EPI_ISL_2473665 | Oregon State Public Health Laboratory | Oregon State Public Health Laboratory | Rafia Razzaque, Eugene Yeboah, Vanda Makris, Laura Tsaknaridis, John Fontana and Shane Sevey |
| EPI_ISL_2479923 | UPA UNIDADE DE PRONTO ATENDIMENTO 24 HORAS BOM JESUS | Instituto Butantan | Dimas Tadeu Covas, Antonio Jorge Martins, Claudia Renata dos Santos Barros, David Schlesinger, Debora Botequio Moretti, Elaine Cristina Marqueze, Elaine Vieira Santos, Evandra Strazza Rodrigues, Heidge Fukumasu, Jayme Augusto de Souza-Neto, José Salvatore Leister Patané, Luiz Alcantara, Luiz Lehmann Coutinho, Maria Carolina Elias, Maurício Lacerda Nogueira, Rafael dos Santos Bezerra, Raul Machado Neto, Rejane Maria Tommasini Grotto, Ricardo Haddad, Sandra Coccuzzo Sampaio Vessoni, Simone Kashima, Svetoslav Nanev Slavov, Vincent Louis Viala |
| EPI_ISL_2483256, EPI_ISL_2483279, EPI_ISL_2483291, EPI_ISL_2483294, EPI_ISL_2483300 | Platform BIS UZA/UAntwerpen | Labo Klinische Biologie, UZA | Jasmine Coppens, Marie Le Mercier, Basil Britto Xavier, Christine Lammens, Veerle Matheeussen, Herman Goossens |
| EPI_ISL_2484017, EPI_ISL_2484018 | Hospital General Universitario Gregorio Marañón | Hospital General Universitario Gregorio Marañón | Sergio Buenestado Serrano, Pedro Sola Campoy, Laura Pérez-Lago, Cristina Rodriguez-Grande, Marta Herranz Martin, Victor Manuel de la Cueva, Julia Suárez, Pilar Catalán, Patricia Muñoz, Darío García de Viedma |
| EPI_ISL_2492423, EPI_ISL_2492425 | Cliniques universitaires Saint-Luc | UCLouvain/IREC/MBLG | Jean Ruelle, Benoit Kabamba Mukadi |
| EPI_ISL_2499942, EPI_ISL_2499954 | Wichita State University - Molecular Diagnostics Lab | Kansas Health and Environmental Lab | Mike Grose, Katherine Wiggins, Jonathan Barnell, Ben Olsen, and Phil Adam |
| EPI_ISL_2499975, EPI_ISL_2499977, EPI_ISL_2499978, EPI_ISL_2499979, EPI_ISL_2499981, EPI_ISL_2499983 | SYNLAB | GIGA Medical Genomics | Keith Durkin, Maria Artesi, Sébastien Bontems, Raphaël Boreux, Bouchra Boujemla, Nathalie Renotte, Cécile Meex, Pierrette Melin, Marie-Pierre Hayette, Vincent Bours |
| EPI_ISL_2500004, EPI_ISL_2500005 | University of Liège COVID-19 testing center | GIGA Medical Genomics | Keith Durkin, Maria Artesi, Bouchra Boujemla, Nathalie Renotte, Cécile Meex, Sébastien Bontems, Fabrice Bureau, Laurent Gillet, Wouter Coppieters, Marie-Pierre Hayette, Vincent Bours |
| EPI_ISL_2501152 | Minnesota Department of Health, Public Health Laboratory | Minnesota Department of Health, Public Health Laboratory | Alexandra Lorentz, Jacob Garfin, Matt Plumb, and Xiong Wang |
| EPI_ISL_2504004 | Alaska State Virology Laboratory | Alaska State Virology Laboratory | Stephanie DeRonde, Elva House, Jacob Zidek, Lisa Smith, Ph.D., Jack Chen, Ph.D. |
| EPI_ISL_2516752, EPI_ISL_2516755, EPI_ISL_2516859 | Servicio de Microbiología Clínica (Complejo Hospitalario de Navarra, Pamplona) | Centro de Secuenciación NASERTIC | Carmen Ezpeleta Baquedano, Ana Navascués, Ana Miqueleiz |
| EPI_ISL_2521960, EPI_ISL_2521969 | Plateforme de testing Namuroise | Plateforme de testing Namuroise | Nicolas Gilliard; Demars Aurore ; Maschietto Céline ; Nobis Chloé ; Otto Gaetan ; Denis Olivier ; Degosserie Jonathan ; Mullier François |
| EPI_ISL_2524776 | Hospital Universitari Arnau de Vilanova | Hospital Universitari Vall d'Hebron - Vall d'Hebron Institut de Recerca | Cristina Andrés, Maria Piñana, Alejandra González-Sánchez, Damir Garcia-Cehic, Ariadna Rando, Juliana Esperalba, Maria Gema Codina, Carla Castillo, Maria Carmen Martin, Tomàs Pumarola, Josep Quer, Andrés Antón |
| EPI_ISL_2527565, EPI_ISL_2527592, EPI_ISL_2527650, EPI_ISL_2527735, EPI_ISL_2527739, EPI_ISL_2527741, EPI_ISL_2527753, EPI_ISL_2527757, EPI_ISL_2527760, EPI_ISL_2527801, EPI_ISL_2527811, EPI_ISL_2527816 | | | |
| see above | Fulgent Genetics | Centers for Disease Control and Prevention Division of Viral Diseases, Pathogen Discovery | Dakota Howard, Dhwani Batra, Peter W. Cook, Kara Moser, Adrian Paskey, Jason Caravas, Benjamin Rambo-Martin, Shatavia Morrison, Christopher Gulvick, Scott Sammons, Yvette Unoarumhi, Darlene Wagner, Matthew Schmerer, Harry Gao, Mickey Li, John Gao, Joseph Fierro, Benafsh Sapra, Becky Tsai, Yan Meng, Doreen Ng, James Xie, Clinton R. Paden, Duncan MacCannell |
| EPI_ISL_2527882, EPI_ISL_2527899, EPI_ISL_2527909, EPI_ISL_2527937, EPI_ISL_2527954, EPI_ISL_2527961, EPI_ISL_2527974, EPI_ISL_2527995, EPI_ISL_2528163, EPI_ISL_2528165, EPI_ISL_2528218, EPI_ISL_2528226, EPI_ISL_2528233, EPI_ISL_2528235, EPI_ISL_2528237, EPI_ISL_2528283, EPI_ISL_2528299, EPI_ISL_2528310, EPI_ISL_2528318, EPI_ISL_2528372, EPI_ISL_2528380, EPI_ISL_2528394, EPI_ISL_2528419, EPI_ISL_2528425, EPI_ISL_2528428, EPI_ISL_2528862, EPI_ISL_2529005 | | | |
| see above | Aegis Sciences Corporation | Centers for Disease Control and Prevention Division of Viral Diseases, Pathogen Discovery | Dakota Howard, Dhwani Batra, Peter W. Cook, Kara Moser, Adrian Paskey, Jason Caravas, Benjamin Rambo-Martin, Shatavia Morrison, Christopher Gulvick, Scott Sammons, Yvette Unoarumhi, Darlene Wagner, Matthew Schmerer, Cyndi Clark, Patrick Campbell, Rob Case, Vikramsinha Ghorpade, Holly Houdeshell, Ola Kvalvaag, Dillon Nall, Ethan Sanders, Alec Vest, Shaun Westlund, Matthew Hardison, Clinton R. Paden, Duncan MacCannell |
| EPI_ISL_2534256, EPI_ISL_2534262 | Wichita State University - Molecular Diagnostics Lab | Kansas Health and Environmental Lab | Mike Grose, Katherine Wiggins, Jonathan Barnell, Ben Olsen, and Phil Adam |
| EPI_ISL_2534552 | Aegis Sciences Corporation | Centers for Disease Control and Prevention Division of Viral Diseases, Pathogen Discovery | Dakota Howard, Dhwani Batra, Peter W. Cook, Kara Moser, Adrian Paskey, Jason Caravas, Benjamin Rambo-Martin, Shatavia Morrison, Christopher Gulvick, Scott Sammons, Yvette Unoarumhi, Darlene Wagner, Matthew Schmerer, Cyndi Clark, Patrick Campbell, Rob Case, Vikramsinha Ghorpade, Holly Houdeshell, Ola Kvalvaag, Dillon Nall, Ethan Sanders, Alec Vest, Shaun Westlund, Matthew Hardison, Clinton R. Paden, Duncan MacCannell |
| EPI_ISL_2536905 | Centracare Laboratory Services | Minnesota Department of Health, Public Health Laboratory | Alexandra Lorentz, Jacob Garfin, Matt Plumb, and Xiong Wang |
| EPI_ISL_2544257 | Medica | Institute of Medical Virology | Verena Kufner, Gabriela Ziltener, Maryam Zaheri, Stefan Schmutz, Annette Audigé, Maria Grünberg, Kevin Steiner, Jon Huder, Cyril Shah, Riccarda Capaul, Guido Bloemberg, Jürg Böni, Michael Huber, Alexandra Trkola |
| EPI_ISL_2544269 | UniversitätsSpital Zürich 009 | Institute of Medical Virology | Verena Kufner, Gabriela Ziltener, Maryam Zaheri, Stefan Schmutz, Annette Audigé, Maria Grünberg, Kevin Steiner, Jon Huder, Cyril Shah, Riccarda Capaul, Guido Bloemberg, Jürg Böni, Michael Huber, Alexandra Trkola |
| EPI_ISL_2544982 | UW Virology Lab | UW Virology Lab | Pavitra Roychoudhury, Hong Xie, Lasata Shrestha, Shah Mohamed Bakhash, Tien V. Nguyen, Noah R. Baker, Sean Ellis, Meei-Li Huang, Keith R Jerome, Alexander Greninger |
| EPI_ISL_2545446, EPI_ISL_2545468 | National Platform bis UMONS/Jolimont | National Platform bis UMONS/Jolimont | François Dufrasne, Guillaume Bayon-Vicente, Florian Juszczak, Gautier Detry, Ruddy Wattiez |
| EPI_ISL_2545596, EPI_ISL_2545598, EPI_ISL_2545599, EPI_ISL_2545600, EPI_ISL_2545610, EPI_ISL_2545636 | Platform BIS UZA/UAntwerpen | Labo Klinische Biologie, UZA | Jasmine Coppens, Marie Le Mercier, Basil Britto Xavier, Christine Lammens, Veerle Matheeussen, Herman Goossens |
| EPI_ISL_2545657, EPI_ISL_2545658 | LHUB-ULB | Labo Klinische Biologie, UZA | Jasmine Coppens, Marie Le Mercier, Basil Britto Xavier, Christine Lammens, Veerle Matheeussen, Herman Goossens |
| EPI_ISL_2549337, EPI_ISL_2549343, EPI_ISL_2549346, EPI_ISL_2549354, EPI_ISL_2549359, EPI_ISL_2549371, EPI_ISL_2549378, EPI_ISL_2549382, EPI_ISL_2549385, EPI_ISL_2549399, EPI_ISL_2549409, EPI_ISL_2549432, EPI_ISL_2549442, EPI_ISL_2549447, EPI_ISL_2549450, EPI_ISL_2549451, EPI_ISL_2549458, EPI_ISL_2549471, EPI_ISL_2549476, EPI_ISL_2549478, EPI_ISL_2549479, EPI_ISL_2549481, EPI_ISL_2549493, EPI_ISL_2549495, EPI_ISL_2549496, EPI_ISL_2549504, EPI_ISL_2549507, EPI_ISL_2549519 | | | |
| see above | UW Virology Lab | UW Virology Lab | Pavitra Roychoudhury, Hong Xie, Lasata Shrestha, Tien V. Nguyen, Shah Mohamed Bakhash, Michelle Lin, Noah R. Baker, Ricardo Perez, Sean Ellis, Nathan Breit, Robert J. Livingston, Meei-Li Huang, Keith R Jerome, Patrick Mathias, Alexander Greninger |
| EPI_ISL_2550983, EPI_ISL_2551117, EPI_ISL_2551127 | Istituto Zooprofilattico Sperimentale del Mezzogiorno | TIGEM | Antonio Grimaldi Patrizia Annunziata Francesco Panariello Biancamaria Pierri Claudia Tiberio Teresa Giuliano Valentina Bouche Chiara Colantuono Maria Concetta Cuomo Denise Di Concilio Lucio Di Filippo Anna Manfredi Marcello Salvi Antonio Limone Luigi Atripaldi Pellegrino Cerino Andrea Ballabio Davide Cacchiarelli |
| EPI_ISL_2557148, EPI_ISL_2557157 | NJDOH, Public Health and Environmental Laboratories | NJ_PHEL | Lindsey Bodnar, Shiv K. Verma, Ryan Pachucki, Chelsea San Filippo, Dana Woell, Allison Roder, Byeong Jeong |

| | | | |
|---|---|---|---|
| EPI_ISL_2557242 | IN State Department of Health Laboratory Services | IN State Department of Health Laboratory Services | Cassandra Campion, Jamie Yeadon, Brian Pope, Lixia Liu, Kyle Brownlee, Melissa Hindenlang, Mark Glazier |
| EPI_ISL_2557409, EPI_ISL_2557410 | Laboratory of Respiratory Viruses and Measles, Oswaldo Cruz Institute, FIOCRUZ | Laboratory of Respiratory Viruses and Measles, Oswaldo Cruz Institute, FIOCRUZ | Paola Resende, Luciana Appolinario, Fernando Motta, Ana Carolina Paixao, Ana Carolina Mendonca, Alice Sampaio Rocha, Taina Venas, Elisa Cavalcante Pereira, Renata Serrano Lopes, Marilda Siqueira on behalf of the Fiocruz COVID-19 Genomic Surveillance Network |
| EPI_ISL_2558702 | Kansas Health and Environmental Lab | Kansas Health and Environmental Lab | Katherine Wiggins, Mike Grose, Jonathan Barnell, Ben Olsen, and Phil Adam |
| EPI_ISL_2574072, EPI_ISL_2574075 | Center for Laboratory Medicine | Center for Laboratory Medicine | Yannick Gerth |
| EPI_ISL_2597899 | WSSE w Gdasku | 1. Tricity SARS-CoV-2 sequencing consortium: University of Gdansk, Medical University of Gdansk, Vaxican Ltd., Invicta Ltd. 2. National Institute of Public Health - National Institute of Hygiene, Warsaw, Poland | Maciej Kosinski, Celina Cybulska, Krystyna Bienkowska Szewczyk, Maciej Grzybek, Karolina Gackowska, Marcin Lubocki, Katarzyna Groth, Lukasz Rabalski, Katarzyna Zacharczuk, Magdalena Nowakowska, Magorzata Sadkowska-Todys, Tomasz Wokowicz |
| EPI_ISL_2597931 | Nebraska Public Health Laboratory | NPHL COVID-19 Response Team | NPHL COVID-19 Response Team |
| EPI_ISL_2598160 | Minnesota Department of Health, Public Health Laboratory | Minnesota Department of Health, Public Health Laboratory | Alexandra Lorentz, Jacob Garfin, Matt Plumb, and Xiong Wang |
| EPI_ISL_2598704, EPI_ISL_2598710, EPI_ISL_2598724, EPI_ISL_2598726, EPI_ISL_2598733, EPI_ISL_2598764, EPI_ISL_2598778, EPI_ISL_2598791, EPI_ISL_2598822, EPI_ISL_2598828, EPI_ISL_2599922, EPI_ISL_2599923 | | | |
| see above | Fulgent Genetics | Centers for Disease Control and Prevention Division of Viral Diseases, Pathogen Discovery | Dakota Howard, Dhwani Batra, Peter W. Cook, Kara Moser, Adrian Paskey, Jason Caravas, Benjamin Rambo-Martin, Shatavia Morrison, Christopher Gulvick, Scott Sammons, Yvette Unoarumhi, Darlene Wagner, Matthew Schmerer, Harry Gao, Mickey Li, John Gao, Joseph Fierro, Benafsh Sapra, Becky Tsai, Yan Meng, Doreen Ng, James Xie, Clinton R. Paden, Duncan MacCannell |
| EPI_ISL_2600917 | Servicio de Microbiología. Hospital Clínico Universitario de Valencia | SeqCOVID-SPAIN consortium/IBV(CSIC) | David Navarro Ortega, Eliseo Albert Vicent, Ignacio Torres and SeqCOVID-SPAIN consortium |
| EPI_ISL_2600943, EPI_ISL_2600944, EPI_ISL_2600948 | Plateforme de testing Namuroise | Plateforme de testing Namuroise | Nicolas GILLIARD, Lesly NYINKEU KEMAMEN; Demars Aurore ; Maschietto Céline ; Nobis Chloé ; Otto Gaetan ; Denis Olivier ; Degosserie Jonathan ; Mullier François |
| EPI_ISL_2601701 | Hospital General Universitario Gregorio Marañón | Hospital General Universitario Gregorio Marañón | Sergio Buenestado Serrano, Pedro Sola Campoy, Laura Pérez-Lago, Cristina Rodriguez-Grande, Marta Herranz Martin, Victor Manuel de la Cueva, Julia Suárez, Pilar Catalán, Patricia Muñoz, Darío García de Viedma |
| EPI_ISL_2601973, EPI_ISL_2601975, EPI_ISL_2601979 | Jessa | Jessa | Berden et al. on behalf of the Jessa_cmdLab |
| EPI_ISL_2604198, EPI_ISL_2604209 | Oregon State Public Health Laboratory | Oregon State Public Health Laboratory | Rafia Razzaque, Eugene Yeboah, Vanda Makris, Laura Tsaknaridis, John Fontana and Shane Sevey |
| EPI_ISL_2604871 | Omics Sciences Laboratory | Omics Sciences Laboratory | Derly Andrade Molina, Gabriel Morey León, Darlyn Amaya, Rubén Armas González, Juan Carlos Fernández Cadena |
| EPI_ISL_2604875, EPI_ISL_2604876, EPI_ISL_2604877 | INTERLAB | Omics Sciences Laboratory | Derly Andrade Molina, Gabriel Morey León, Darlyn Amaya, Rubén Armas González, Juan Carlos Fernández Cadena |
| EPI_ISL_2610665, EPI_ISL_2610672, EPI_ISL_2610673, EPI_ISL_2610674, EPI_ISL_2610691, EPI_ISL_2610697, EPI_ISL_2610700, EPI_ISL_2610733, EPI_ISL_2610743, EPI_ISL_2610753, EPI_ISL_2610754, EPI_ISL_2610765, EPI_ISL_2610766, EPI_ISL_2610776 | | | |
| see above | Dutch COVID-19 response team | National Institute for Public Health and the Environment (RIVM) | Adam Meijer, Harry Vennema, Dirk Eggink, Jeroen Cremer, Sharon van den Brink, Bas van der Veer, AnneMarie van den Brandt, Lisa Wijsman, Kim Freriks, Ryanne Jaarsma, Euníce Then, Lynn Aarts, Sanne Bos, Melissa van Tuil, Linda van de Nes, Florian Zwagemaker, Dennis Schmitz, Annelies Kroneman, Karim Hajji, Chantal Reusken, on behalf of the national COVID-19 response team |
| EPI_ISL_2611074 | Viollier AG | Department of Biosystems Science and Engineering, ETH Zürich | Christian Beisel, Sarah Nadeau, Chaoran Chen, Ivan Topolsky, Kim Philipp Jablonski, Lara Fuhrmann, Rebecca Denes, Mirjam Feldkamp, Ina Nissen, Natascha Santacroce, Elodie Burcklen, Christiane Beckmann, Maurice Redondo, Olivier Kobel, Christoph Noppen, Niko Beerenwinkel, Tanja Stadler |
| EPI_ISL_2611209, EPI_ISL_2611229, EPI_ISL_2611230, EPI_ISL_2611232, EPI_ISL_2611233, EPI_ISL_2611238, EPI_ISL_2611239, EPI_ISL_2611240, EPI_ISL_2611242, EPI_ISL_2611304, EPI_ISL_2611313, EPI_ISL_2611314, EPI_ISL_2611321, EPI_ISL_2611323, EPI_ISL_2611324, EPI_ISL_2611332, EPI_ISL_2611334, EPI_ISL_2611338, EPI_ISL_2611344, EPI_ISL_2611349, EPI_ISL_2611364, EPI_ISL_2611372, EPI_ISL_2611380, EPI_ISL_2611384, EPI_ISL_2611392, EPI_ISL_2611397, EPI_ISL_2611398, EPI_ISL_2611413, EPI_ISL_2611418, EPI_ISL_2611420, EPI_ISL_2611428, EPI_ISL_2611607, EPI_ISL_2611626, EPI_ISL_2611627, EPI_ISL_2611631, EPI_ISL_2611645, EPI_ISL_2611646, EPI_ISL_2611656, EPI_ISL_2611657, EPI_ISL_2611692, EPI_ISL_2611767, EPI_ISL_2611772, EPI_ISL_2611812, EPI_ISL_2611823 | | | |
| see above | Laboratory Corporation of America | Centers for Disease Control and Prevention Division of Viral Diseases, Pathogen Discovery | Dakota Howard, Dhwani Batra, Peter W. Cook, Kara Moser, Adrian Paskey, Jason Caravas, Benjamin Rambo-Martin, Shatavia Morrison, Christopher Gulvick, Scott Sammons, Yvette Unoarumhi, Darlene Wagner, Matthew Schmerer, Minoo Agarwal, Eyad Almasri, Debbie Boles, Ayla Burns, Nuthawin Charoensri, Oren Cohen, Susan Countryman, Mary Ann Cristobal, Bobbi Croy, Suzanne Dale, Hrushikesh Deshmukh, Amanda Douglas, Vincent Drouillon, Marcia Eisenberg, Howard Engler, Rama Ghatti, Prashant Gupta, Susan Hicks, Jake Humphrey, Lax Iyer, Lisa Pfefferle, Manoj Jain, Matthew Robinson, Mohan Kolli, Brian Krueger, Tim Kuphal, Stanley Letovsky, Michael Levandoski, Craig Lukasik, Jonathan Meltzer, Brian Norvell, Mindy Nye, Scott Parker, Christos Petropoulos, John Pruitt, Steven Ragan, Scott Ryan, Mike Sapeta, Jana Schroth, Suresh Babu Selvaraju, Goran Stevovic, Amanda Suchanek, Andrea Throop, Lyndon Tilson, Thomas Urban, Joe Voshell, Kimberly Wagner, Jonathan Williams, Mary Williamson, Qian Zeng, Tricia Zwiefelhofer, Clinton R. Paden, Duncan MacCannell |
| EPI_ISL_2612191, EPI_ISL_2612195 | Wichita State University - Molecular Diagnostics Lab | Kansas Health and Environmental Lab | Mike Grose, Katherine Wiggins, Jonathan Barnell, Ben Olsen, and Phil Adam |
| EPI_ISL_2612507 | Nevada State Public Health Laboratory | Nevada State Public Health Laboratory | Andrew Gorzalski, Mark Pandori |
| EPI_ISL_2612540, EPI_ISL_2612541, EPI_ISL_2612545 | Oregon State Public Health Laboratory | Oregon State Public Health Laboratory | Rafia Razzaque, Eugene Yeboah, Vanda Makris, Laura Tsaknaridis, John Fontana and Shane Sevey |
| EPI_ISL_2612553, EPI_ISL_2612554, EPI_ISL_2612555 | DOHMH Corona | New York City Public Health Laboratory | Jade Wang, et al. |
| EPI_ISL_2616771 | Houston Health Dept. | Houston Health Dept. | Ryker Penn, Pamela Brown, Adolpho Lara, Yanlai Lai |
| EPI_ISL_2617009, EPI_ISL_2617010, EPI_ISL_2617011, EPI_ISL_2617012, EPI_ISL_2617013, EPI_ISL_2617015, EPI_ISL_2617016, EPI_ISL_2617017 | Salud Digna | Instituto Nacional de Medicina Genomica | Hidalgo-Miranda A, Cedro-Tanda A, Mendoza-Vargas A, Reyes-Grajeda JP, Abraham Campos-Romero, Moreno-Camacho José Luis, Rodriguez-Gallegos Jorge, Luna-Ruiz Marco, Gonzalez-Barrera D, Rangel-DeLeon D, Munguia-Garza P, Ramirez-Vega O, Escobar-Arrazola, M, Herrera-Montalvo LA. |
| EPI_ISL_2617603, EPI_ISL_2617605, EPI_ISL_2617606, EPI_ISL_2617622, EPI_ISL_2617623, EPI_ISL_2617624, EPI_ISL_2617625 | HLAGYN - Laboratorio de Imunologia de Transplantes de Goias | HLAGYN - Laboratorio de Imunologia de Transplantes de Goias | Fernando Antonio Vinhal dos Santos, Erika Lopes Rocha Batista, Alessandro Leonardo Alvares Magalhaes, Frederico Rodrigues Vinhal, Sabrina Sara Moreira Duarte, Lucas Carlos Gomes Pereira, Daniel Ferreira de Sousa |
| EPI_ISL_2617628, EPI_ISL_2617970, EPI_ISL_2617975, EPI_ISL_2617987, EPI_ISL_2617992, EPI_ISL_2617997, EPI_ISL_2617998, EPI_ISL_2618002, EPI_ISL_2618003, EPI_ISL_2618004, EPI_ISL_2618005, EPI_ISL_2618012, EPI_ISL_2618023, EPI_ISL_2618038, EPI_ISL_2618049, EPI_ISL_2618053, EPI_ISL_2618056, EPI_ISL_2618070, EPI_ISL_2618085, EPI_ISL_2618088, EPI_ISL_2618098 | | | |
| see above | UW Virology Lab | UW Virology Lab | Pavitra Roychoudhury, Hong Xie, Lasata Shrestha, Tien V. Nguyen, Shah Mohamed Bakhash, Michelle Lin, Noah R. Baker, Ricardo Perez, Sean Ellis, Nathan Breit, Robert J. Livingston, Meei-Li Huang, Keith R Jerome, Patrick Mathias, Alexander Greninger |
| EPI_ISL_2620963, EPI_ISL_2620966 | Hospital Universitario Virgen de las Nieves de Granada-SAS | SeqCOVID-SPAIN consortium/IBV(CSIC) | Sara Sanbonmatsu Gámez, Irene Pedrosa Corral, José M. Navarro-Marí and SeqCOVID-SPAIN consortium |
| EPI_ISL_2621554 | Genomics, Poplar Healthcare | Genomics, Poplar Healthcare | Truong,M. |
| EPI_ISL_2621962, EPI_ISL_2621963 | National Public Health Laboratory, National Centre for Infectious Diseases | National Public Health Laboratory, National Centre for Infectious Diseases | Tze Minn Mak, Zhenyang Zhou, Royce Ang, Lin Cui, Raymond Tzer Pin Lin |
| EPI_ISL_2625749, EPI_ISL_2625753 | AZDelta | AZ Delta Medical Laboratories in Roeselare, Belgium | Geert Martens, Dieter De Smet, Merijn Vanhee, on behalf of AZ Delta COVID-19 Genomics core (member of Genomic surveillance of SARS-CoV-2 in Belgium network) |
| EPI_ISL_2626011, EPI_ISL_2626014, EPI_ISL_2626019, EPI_ISL_2626029, | Cliniques universitaires Saint-Luc | UCLouvain/IREC/MBLG | Jean Ruelle, Bertrand Bearzatto, Benoit Kabamba Mukadi |

| | | | |
|---|---|---|---|
| EPI_ISL_2626043 | | | |
| EPI_ISL_2626066 | Clinique Saint-Pierre Ottignies | UCLouvain/IREC/MBLG | Jean Ruelle, Bertrand Bearzatto, Benoit Kabamba Mukadi |
| EPI_ISL_2626082 | Cliniques universitaires Saint-Luc | UCLouvain/IREC/MBLG | Jean Ruelle, Bertrand Bearzatto, Benoit Kabamba Mukadi |
| EPI_ISL_2626156 | HOSPITAL MANACOR | HOSPITAL UNIVERSITARIO SON ESPASES | Servicio de Microbiología, Hospital Universitario Son Espases |
| EPI_ISL_2626160 | HOSPITAL UNIVERSITARIO SON ESPASES | HOSPITAL UNIVERSITARIO SON ESPASES | Servicio de Microbiología, Hospital Universitario Son Espases |
| EPI_ISL_2626722, EPI_ISL_2626723, EPI_ISL_2626727, EPI_ISL_2626735, EPI_ISL_2626737, EPI_ISL_2626738, EPI_ISL_2626743, EPI_ISL_2626745 | Alaska State Virology Laboratory | Alaska State Virology Laboratory | Stephanie DeRonde, Elva House, Jacob Zidek, Lisa Smith, Ph.D., Jack Chen, Ph.D. |
| EPI_ISL_2626831 | Baylor Scott & White-Temple | Baylor Scott & White-Temple | Ari Rao, Linden Morales, Kimberly Walker, Marcus Volz, Shelby Johnson |
| EPI_ISL_2626853, EPI_ISL_2626882, EPI_ISL_2626883, EPI_ISL_2626884, EPI_ISL_2626903 | Platform BIS UZA/UAntwerpen | Labo Klinische Biologie, UZA | Marie Le Mercier, Jasmine Coppens, Basil Britto Xavier, Christine Lammens, Veerle Matheeussen, Herman Goossens |
| EPI_ISL_2627274 | Platform BIS UZA/UAntwerpen | Labo Klinische Biologie, UZA | Jasmine Coppens, Marie Le Mercier, Basil Britto Xavier, Christine Lammens, Veerle Matheeussen, Herman Goossens |
| EPI_ISL_2627864, EPI_ISL_2627865, EPI_ISL_2627866 | CNR Institut Pasteur de la Guyane | Institut Pasteur de la Guyane | Anne Lavergne, Dominique Rousset, Antoine Enfissi, Arielle Salmier |
| EPI_ISL_2627930, EPI_ISL_2627938, EPI_ISL_2627939 | Wichita State University - Molecular Diagnostics Lab | Kansas Health and Environmental Lab | Mike Grose, Katherine Wiggins, Jonathan Barnell, Ben Olsen, and Phil Adam |
| EPI_ISL_2628061, EPI_ISL_2628062, EPI_ISL_2628063, EPI_ISL_2628064, EPI_ISL_2628065, EPI_ISL_2628069 | Platform BIS UZA/UAntwerpen | Labo Klinische Biologie, UZA | Marie Le Mercier, Jasmine Coppens, Basil Britto Xavier, Christine Lammens, Veerle Matheeussen, Herman Goossens |
| EPI_ISL_2628717, EPI_ISL_2628825, EPI_ISL_2628962 | UNILABS | Instituto Nacional de Saude (INSA) | Borges et al |
| EPI_ISL_2629224, EPI_ISL_2629232, EPI_ISL_2629233, EPI_ISL_2629234 | CHC Andrée Rosemon | Institut Pasteur de la Guyane | Anne Lavergne, Dominique Rousset, Antoine Enfissi, Arielle Salmier |
| EPI_ISL_2629235, EPI_ISL_2629236, EPI_ISL_2629237 | CNR Institut Pasteur de la Guyane | Institut Pasteur de la Guyane | Anne Lavergne, Dominique Rousset, Antoine Enfissi, Arielle Salmier |
| EPI_ISL_2631155 | Oregon State Public Health Laboratory | Oregon State Public Health Laboratory | Rafia Razzaque, Eugene Yeboah, Vanda Makris, Laura Tsaknaridis, John Fontana and Shane Sevey |
| EPI_ISL_2631350, EPI_ISL_2631354, EPI_ISL_2631359, EPI_ISL_2631360, EPI_ISL_2631386, EPI_ISL_2631407 | Pandemic Response Lab - NYC | Pandemic Response Lab, R&D | Henry Lee, Michael Hammerling, Melissa Hopkins, Cybill del Castillo, Shinyoung Clair Kang, William Ward, Pradeep Bugga, Sol Rey, Dylan Law, Katharine Nelson, Haiping Hao, Jon Laurent |
| EPI_ISL_2632831 | MVZ Labor Dr. Limbach & Kollegen GbR | Robert Koch Institute | unknown |
| EPI_ISL_2633310 | SYNLAB MVZ Leverkusen | Robert Koch Institute | unknown |
| EPI_ISL_2633321 | SYNLAB MVZ Leinfelden-Echterdingen | Robert Koch Institute | unknown |
| EPI_ISL_2633914 | Bioscientia MVZ Labor Karlsruhe GmbH | Robert Koch Institute | unknown |
| EPI_ISL_2634916, EPI_ISL_2634943, EPI_ISL_2634965 | Pandemic Response Lab - NYC | Pandemic Response Lab, R&D | Henry Lee, Michael Hammerling, Melissa Hopkins, Cybill del Castillo, Shinyoung Clair Kang, William Ward, Pradeep Bugga, Sol Rey, Dylan Law, Katharine Nelson, Haiping Hao, Jon Laurent |
| EPI_ISL_2635200, EPI_ISL_2635291 | SYNLAB MVZ Leverkusen | Robert Koch Institute | unknown |
| EPI_ISL_2635480 | MVZ für Laboratoriumsmedizin und Mikrobiologie Würzburg | Robert Koch Institute | unknown |
| EPI_ISL_2635865, EPI_ISL_2635874, EPI_ISL_2635888 | MVZ Labor Dr. Limbach & Kollegen GbR | Robert Koch Institute | unknown |
| EPI_ISL_2636711 | Diagnosticum - Labor Neukirchen | Robert Koch Institute | unknown |
| EPI_ISL_2636815 | Medizinisch-Diagnostisches Labor Kempten allgäulab | Robert Koch Institute | unknown |
| EPI_ISL_2637060, EPI_ISL_2637156, EPI_ISL_2637157 | Bioscientia Labor Wermsdorf | Robert Koch Institute | unknown |
| EPI_ISL_2637186 | Labor Dr. Schumacher MVZ | Robert Koch Institute | unknown |
| EPI_ISL_2637260 | CLILAB | Microbiology Department | Sara Marti, Aida Gonzalez-Diaz, Laura Calatayud, Jordi Niubó, Miguel Fernandez-Huerta, Carmen Ardanuy, Jordi Camara, M Angeles Domínguez |
| EPI_ISL_2637324 | Hospital Universitari Bellvitge | Microbiology Department | Sara Marti, Aida Gonzalez-Diaz, Laura Calatayud, Jordi Niubó, Miguel Fernandez-Huerta, Carmen Ardanuy, Jordi Camara, M Angeles Domínguez |
| EPI_ISL_2637629, EPI_ISL_2637761 | Laboratoire LENYS | Department of Virology, Henri Mondor University Hospital, Assistance Publique Hôpitaux de Paris, Université Paris-Est Créteil, INSERM U955 | Christophe Rodriguez, Slim Fourati, Vanessa Demontant, Guillaume Gricourt, Melissa N'Debi, Alexandre Soulier, Elisabeth Trawinski, Jean-Michel Pawlotsky |
| EPI_ISL_2638007 | Biogroup Bio Lam-LCD Saint-Denis | Department of Virology, Henri Mondor University Hospital, Assistance Publique Hôpitaux de Paris, Université Paris-Est Créteil, INSERM U955 | Christophe Rodriguez, Slim Fourati, Vanessa Demontant, Guillaume Gricourt, Melissa N'Debi, Alexandre Soulier, Elisabeth Trawinski, Jean-Michel Pawlotsky |
| EPI_ISL_2641746, EPI_ISL_2641751, EPI_ISL_2641754, EPI_ISL_2641771, EPI_ISL_2641775, EPI_ISL_2641782, EPI_ISL_2641789, EPI_ISL_2641793, EPI_ISL_2641794, EPI_ISL_2641800, EPI_ISL_2641816, EPI_ISL_2641817, EPI_ISL_2641818, EPI_ISL_2641820, EPI_ISL_2641824, EPI_ISL_2641829, EPI_ISL_2641835, EPI_ISL_2641862, EPI_ISL_2641865, EPI_ISL_2641868, EPI_ISL_2641884, EPI_ISL_2641889, EPI_ISL_2641906, EPI_ISL_2641907, EPI_ISL_2641926, EPI_ISL_2641938, EPI_ISL_2641973, EPI_ISL_2641984, EPI_ISL_2642007, EPI_ISL_2642011, EPI_ISL_2642029, EPI_ISL_2642040 | | | |
| see above | Istituto Zooprofilattico Sperimentale del Mezzogiorno | Telethon Institute of Genetics and Medicine (TIGEM) | Antonio Grimaldi Patrizia Annunziata Francesco Panariello Biancamaria Pierri Claudia Tiberio Teresa Giuliano Valentina Bouche Chiara Colantuono Maria Concetta Cuomo Denise Di Concilio Lucio Di Filippo Anna Manfredi Marcello Salvi Antonio Limone Luigi Atripaldi Pellegrino Cerino Andrea Ballabio Davide Cacchiarelli |
| EPI_ISL_2645408, EPI_ISL_2645409, EPI_ISL_2645410 | Laboratorio Central de Saude Publica do Estado da Bahia (LACEN/BA) | Laboratory of Respiratory Viruses and Measles, Oswaldo Cruz Institute, FIOCRUZ | Paola Resende, Luciana Appolinario, Fernando Motta, Anna Carolina Paixao, Ana Carolina Mendonca, Alice Sampaio Rocha, Taina Venas, Elisa Cavalcante Pereira, Renata Serrano Lopes,Felicidade Pereira, Marilda Siqueira on behalf of the Fiocruz COVID-19 Genomic Surveillance Network |
| EPI_ISL_2645995, EPI_ISL_2646099 | UNILABS | Instituto Nacional de Saude (INSA) | Borges et al |
| EPI_ISL_2646210, EPI_ISL_2646212 | CH Valenciennes | CHU Lille - Laboratoire de Virologie | AIT YAHYA Emilie, ALIDJINOU Enagnon Kazali, BOCKET Laurence, CREPIN Michel, DEMAY Christophe, ENGELMANN Ilka, GEFFROY Sandrine, GUIGON Aurélie, LAMBERT Valérie, LAZREK Mouna, NOBILLIAUX Florian, PREVOST Brigitte, THUILLIER Caroline, TINEZ Claire ,TCHANTCHOU NJOSSE YANICK |
| EPI_ISL_2646337, EPI_ISL_2646340 | MS Public Health Laboratory | University of Mississippi Medical Center, Molecular and Genomics Core Facility | Ashley C. Johnson, Wenjie Wu, Ithiel J. Frame, Krishna K. Ayyalasomayajula, Michael R. Garrett, D. Ashley Robinson |
| EPI_ISL_2649831 | NJDOH, Public Health and Environmental Laboratories | NJ_PHEL | Lindsey Bodnar, Shiv K. Verma, Ryan Pachucki, Chelsea San Filippo, Jacquelyn Deverell, Dana Woell, Allison Roder, Byeong Jeong |

| | | | |
|---|---|---|---|
| EPI_ISL_2649965, EPI_ISL_2649966, EPI_ISL_2649967, EPI_ISL_2649968, EPI_ISL_2649969, EPI_ISL_2649976, EPI_ISL_2649978, EPI_ISL_2649979, EPI_ISL_2649980, EPI_ISL_2649981, EPI_ISL_2649982, EPI_ISL_2649984, EPI_ISL_2649986, EPI_ISL_2649987, EPI_ISL_2649988, EPI_ISL_2649989, EPI_ISL_2649990 | | | |
| see above | Quest Diagnostics Incorporated | Centers for Disease Control and Prevention Division of Viral Diseases, Pathogen Discovery | Dakota Howard, Dhwani Batra, Peter W. Cook, Kara Moser, Adrian Paskey, Jason Caravas, Benjamin Rambo-Martin, Shatavia Morrison, Christopher Gulvick, Scott Sammons, Yvette Unoarumhi, Darlene Wagner, Matthew Schmerer, S. H. Rosenthal, A. Gerasimova, R. M. Kagan, B. Anderson, M. Hua, Y. Liu, L.E. Bernstein, K.E. Livingston, A. Perez, I. A. Shlyakhter, R. V. Rolando, R. Owen, P. Tanpaiboon, F. Lacbawan, Clinton R. Paden, Duncan MacCannell |
| EPI_ISL_2650136, EPI_ISL_2650148 | Arizona State Public Health Laboratory | Arizona State Public Health Laboratory | Trung Huynh, Jessica Escobar, Katherine Fullerton, Nobuko Fukushima, Matthew Contursi, Stacy White, Linda Getsinger, Victor Waddell |
| EPI_ISL_2651062, EPI_ISL_2651066, EPI_ISL_2651067, EPI_ISL_2651083, EPI_ISL_2651084, EPI_ISL_2651086, EPI_ISL_2651097, EPI_ISL_2651105 | Washington State Department of Health Public Health Laboratories | Washington State Department of Health Public Health Laboratories | Drew MacKellar, Philip Dykema, Denny Russell, Joenice Gonzalez, Hannah Gray, Geoff Melly, Vanessa De Los Santos, Darren Lucas, JohnAric Peterson, Avi Singh, Rebecca Cao |
| EPI_ISL_2652076 | New Mexico Department of Health Scientific Laboratory | New Mexico Department of Health Scientific Laboratory | Ellie Johnson, D'eldra Malone, Jennifer Benoit, Ratheesh Rajan, Linda Salazar, Keila Gutierrez, Anastacia Griego-Fisher |
| EPI_ISL_2652157 | AZ Klina | AZ Klina | Carl Vael - Lynsey Berckmans |
| EPI_ISL_2653015, EPI_ISL_2653035, EPI_ISL_2653056, EPI_ISL_2653059 | Infinity Biologix | Centers for Disease Control and Prevention Division of Viral Diseases, Pathogen Discovery | Dakota Howard, Dhwani Batra, Peter W. Cook, Kara Moser, Adrian Paskey, Jason Caravas, Benjamin Rambo-Martin, Shatavia Morrison, Christopher Gulvick, Scott Sammons, Yvette Unoarumhi, Darlene Wagner, Matthew Schmerer, Christian Bixby, Yihe Wang, Jonathan Schultz, Chirayu Goswami, Russ Hager, Robin Grimwood, Clinton R. Paden, Duncan MacCannell |
| EPI_ISL_2658950 | Basurto University Hospital: Clinical Microbiology Laboratory | Biocruces Bizkaia | Mikel Urrutikoetxea-Gutiérrez, Mª Carmen Nieto Toboso, Estibaliz Ugalde Zarraga, Mikel Gallego Rodrigo, Ana de la Hoz, José Luis Díaz de Tuesta del Arco |
| EPI_ISL_2658982 | County of San Luis Obispo Public Health Laboratory | County of San Luis Obispo Public Health Laboratory | Katrina Erwin, Frances Sidhu |
| EPI_ISL_2659142, EPI_ISL_2659174, EPI_ISL_2659193 | Genetica Molecular and Subdepartamento de Virologia ISP Chile | Instituto de Salud Publica de Chile | Karen Orostica, Constanza Campano, Barbara Parra, Loredana Arata, Gisselle Barra, Patricia Bustos, Rodrigo Fasce, Javier Tognarelli, Andres Castillo, Soledad Ulloa, Jorge Fernandez |
| EPI_ISL_2661538, EPI_ISL_2661546, EPI_ISL_2661547, EPI_ISL_2661548, EPI_ISL_2661549 | UHTL, University Hospitals | UHTL, University Hospitals | Sadri,N., Alouani,D., Song,X. |
| EPI_ISL_2662403 | Viollier AG | Department of Biosystems Science and Engineering, ETH Zürich | Andrea Patrizia Salzmann, Henriette Kurth, Christiane Beckmann, Ivan Topolsky, Chaoran Chen, Sarah Nadeau, Kim Philipp Jablonski, Lara Fuhrmann, Niko Beerenwinkel, Christoph Noppen, Olivier Kobel, Maurice Redondo, Tanja Stadler |
| EPI_ISL_2662776 | DPHL | Delaware Public Health Lab | Rebecca Savage |
| EPI_ISL_2663225 | HealthPartners Central Lab | Minnesota Department of Health, Public Health Laboratory | Alexandra Lorentz, Jacob Garfin, Matt Plumb, and Xiong Wang |
| EPI_ISL_2663232, EPI_ISL_2663238 | Mayo Clinic & Mayo Clinic Laboratories | Minnesota Department of Health, Public Health Laboratory | Alexandra Lorentz, Jacob Garfin, Matt Plumb, and Xiong Wang |
| EPI_ISL_2665775, EPI_ISL_2667603, EPI_ISL_2669458, EPI_ISL_2670239, EPI_ISL_2671258 | Department of Bacteria, Parasites and Fungi, Statens Serum Institut, Copenhagen, Denmark | Statens Serum Institut Bioinformatics and Microbial Genomics | Danish Covid-19 Genome Consortium |
| EPI_ISL_2671459, EPI_ISL_2671469 | Center for Laboratory Medicine | Center for Laboratory Medicine | Yannick Gerth |
| EPI_ISL_2672303, EPI_ISL_2672626, EPI_ISL_2672638, EPI_ISL_2672650, EPI_ISL_2672760, EPI_ISL_2672934, EPI_ISL_2672978, EPI_ISL_2673074, EPI_ISL_2673075, EPI_ISL_2673093, EPI_ISL_2673192, EPI_ISL_2673214, EPI_ISL_2673225, EPI_ISL_2673243, EPI_ISL_2673244, EPI_ISL_2673275 | | | |
| see above | Dutch COVID-19 response team | National Institute for Public Health and the Environment (RIVM) | Adam Meijer, Harry Vennema, Dirk Eggink, Jeroen Cremer, Sharon van den Brink, Bas van der Veer, AnneMarie van den Brandt, Lisa Wijsman, Kim Freriks, Ryanne Jaarsma, Euníce Then, Lynn Aarts, Sanne Bos, Melissa van Tuil, Linda van de Nes, Florian Zwagemaker, Dennis Schmitz, Annelies Kroneman, Karim Hajji, Chantal Reusken, on behalf of the national COVID-19 response team |
| EPI_ISL_2674636, EPI_ISL_2674637, EPI_ISL_2674638, EPI_ISL_2674639, EPI_ISL_2674640 | LESP Tabasco | Instituto de Diagnostico y Referencia Epidemiologicos (INDRE) | Claudia Wong-Arambula, Abril Rodriguez-Maldonado, Vanessa Rivero-Arredondo, Ariadna Medina-Benitez, Joaquin Quiroz-Mercado, Sergio Rangel-Guerrero, Natividad Cruz-Ortiz, Tatiana Nunez-Garcia, Gisela Barrera-Badillo, Lucia Hernandez-Rivas, Irma Lopez-Martinez, Ernesto Ramirez-Gonzalez. |
| EPI_ISL_2674644, EPI_ISL_2674645, EPI_ISL_2674646, EPI_ISL_2674647, EPI_ISL_2674648, EPI_ISL_2674649 | LESP Sonora | Instituto de Diagnostico y Referencia Epidemiologicos (INDRE) | Claudia Wong-Arambula, Abril Rodriguez-Maldonado, Vanessa Rivero-Arredondo, Ariadna Medina-Benitez, Joaquin Quiroz-Mercado, Sergio Rangel-Guerrero, Natividad Cruz-Ortiz, Tatiana Nunez-Garcia, Gisela Barrera-Badillo, Lucia Hernandez-Rivas, Irma Lopez-Martinez, Ernesto Ramirez-Gonzalez. |
| EPI_ISL_2674650 | LESP Jalisco | Instituto de Diagnostico y Referencia Epidemiologicos (INDRE) | Claudia Wong-Arambula, Abril Rodriguez-Maldonado, Vanessa Rivero-Arredondo, Ariadna Medina-Benitez, Joaquin Quiroz-Mercado, Sergio Rangel-Guerrero, Natividad Cruz-Ortiz, Tatiana Nunez-Garcia, Gisela Barrera-Badillo, Lucia Hernandez-Rivas, Irma Lopez-Martinez, Ernesto Ramirez-Gonzalez. |
| EPI_ISL_2674674, EPI_ISL_2674675, EPI_ISL_2674676, EPI_ISL_2674677, EPI_ISL_2674678, EPI_ISL_2674679, EPI_ISL_2674680, EPI_ISL_2674681, EPI_ISL_2674682 | LESP Hidalgo | Instituto de Diagnostico y Referencia Epidemiologicos (INDRE) | Claudia Wong-Arambula, Abril Rodriguez-Maldonado, Vanessa Rivero-Arredondo, Ariadna Medina-Benitez, Joaquin Quiroz-Mercado, Sergio Rangel-Guerrero, Natividad Cruz-Ortiz, Tatiana Nunez-Garcia, Gisela Barrera-Badillo, Lucia Hernandez-Rivas, Irma Lopez-Martinez, Ernesto Ramirez-Gonzalez. |
| EPI_ISL_2674683 | LESP Queretaro | Instituto de Diagnostico y Referencia Epidemiologicos (INDRE) | Claudia Wong-Arambula, Abril Rodriguez-Maldonado, Vanessa Rivero-Arredondo, Ariadna Medina-Benitez, Joaquin Quiroz-Mercado, Sergio Rangel-Guerrero, Natividad Cruz-Ortiz, Tatiana Nunez-Garcia, Gisela Barrera-Badillo, Lucia Hernandez-Rivas, Irma Lopez-Martinez, Ernesto Ramirez-Gonzalez. |
| EPI_ISL_2674689 | LESP Coahuila | Instituto de Diagnostico y Referencia Epidemiologicos (INDRE) | Claudia Wong-Arambula, Abril Rodriguez-Maldonado, Vanessa Rivero-Arredondo, Ariadna Medina-Benitez, Joaquin Quiroz-Mercado, Sergio Rangel-Guerrero, Natividad Cruz-Ortiz, Tatiana Nunez-Garcia, Gisela Barrera-Badillo, Lucia Hernandez-Rivas, Irma Lopez-Martinez, Ernesto Ramirez-Gonzalez. |
| EPI_ISL_2674690 | LESP Hidalgo | Instituto de Diagnostico y Referencia Epidemiologicos (INDRE) | Claudia Wong-Arambula, Abril Rodriguez-Maldonado, Vanessa Rivero-Arredondo, Ariadna Medina-Benitez, Joaquin Quiroz-Mercado, Sergio Rangel-Guerrero, Natividad Cruz-Ortiz, Tatiana Nunez-Garcia, Gisela Barrera-Badillo, Lucia Hernandez-Rivas, Irma Lopez-Martinez, Ernesto Ramirez-Gonzalez. |
| EPI_ISL_2674695, EPI_ISL_2674696 | LESP Guanajuato | Instituto de Diagnostico y Referencia Epidemiologicos (INDRE) | Claudia Wong-Arambula, Abril Rodriguez-Maldonado, Vanessa Rivero-Arredondo, Ariadna Medina-Benitez, Joaquin Quiroz-Mercado, Sergio Rangel-Guerrero, Natividad Cruz-Ortiz, Tatiana Nunez-Garcia, Gisela Barrera-Badillo, Lucia Hernandez-Rivas, Irma Lopez-Martinez, Ernesto Ramirez-Gonzalez. |
| EPI_ISL_2674698 | LESP Ciudad de Mexico | Instituto de Diagnostico y Referencia Epidemiologicos (INDRE) | Claudia Wong-Arambula, Abril Rodriguez-Maldonado, Vanessa Rivero-Arredondo, Ariadna Medina-Benitez, Joaquin Quiroz-Mercado, Sergio Rangel-Guerrero, Natividad Cruz-Ortiz, Tatiana Nunez-Garcia, Gisela Barrera-Badillo, Lucia Hernandez-Rivas, Irma Lopez-Martinez, Ernesto Ramirez-Gonzalez. |
| EPI_ISL_2674700 | LESP Puebla | Instituto de Diagnostico y Referencia Epidemiologicos (INDRE) | Claudia Wong-Arambula, Abril Rodriguez-Maldonado, Vanessa Rivero-Arredondo, Ariadna Medina-Benitez, Joaquin Quiroz-Mercado, Sergio Rangel-Guerrero, Natividad Cruz-Ortiz, Tatiana Nunez-Garcia, Gisela Barrera-Badillo, Lucia Hernandez-Rivas, Irma Lopez-Martinez, Ernesto Ramirez-Gonzalez. |
| EPI_ISL_2674703, EPI_ISL_2674704, EPI_ISL_2674705, EPI_ISL_2674706, EPI_ISL_2674707, EPI_ISL_2674708, EPI_ISL_2674709, EPI_ISL_2674710, EPI_ISL_2674711, EPI_ISL_2674712, EPI_ISL_2674713 | | | |
| see above | LESP Yucatan | Instituto de Diagnostico y Referencia Epidemiologicos (INDRE) | Claudia Wong-Arambula, Abril Rodriguez-Maldonado, Vanessa Rivero-Arredondo, Ariadna Medina-Benitez, Joaquin Quiroz-Mercado, Sergio Rangel-Guerrero, Natividad Cruz-Ortiz, Tatiana Nunez-Garcia, Gisela Barrera-Badillo, Lucia Hernandez-Rivas, Irma Lopez-Martinez, Ernesto Ramirez-Gonzalez. |

| | | | |
|---|---|---|---|
| EPI_ISL_2674714 | LESP Quintana Roo | Instituto de Diagnostico y Referencia Epidemiologicos (INDRE) | Claudia Wong-Arambula, Abril Rodriguez-Maldonado, Vanessa Rivero-Arredondo, Ariadna Medina-Benitez, Joaquin Quiroz-Mercado, Sergio Rangel-Guerrero, Natividad Cruz-Ortiz, Tatiana Nunez-Garcia, Gisela Barrera-Badillo, Lucia Hernandez-Rivas, Irma Lopez-Martinez, Ernesto Ramirez-Gonzalez. |
| EPI_ISL_2674715 | LESP Yucatan | Instituto de Diagnostico y Referencia Epidemiologicos (INDRE) | Claudia Wong-Arambula, Abril Rodriguez-Maldonado, Vanessa Rivero-Arredondo, Ariadna Medina-Benitez, Joaquin Quiroz-Mercado, Sergio Rangel-Guerrero, Natividad Cruz-Ortiz, Tatiana Nunez-Garcia, Gisela Barrera-Badillo, Lucia Hernandez-Rivas, Irma Lopez-Martinez, Ernesto Ramirez-Gonzalez. |
| EPI_ISL_2674718, EPI_ISL_2674719 | LESP Sonora | Instituto de Diagnostico y Referencia Epidemiologicos (INDRE) | Claudia Wong-Arambula, Abril Rodriguez-Maldonado, Vanessa Rivero-Arredondo, Ariadna Medina-Benitez, Joaquin Quiroz-Mercado, Sergio Rangel-Guerrero, Natividad Cruz-Ortiz, Tatiana Nunez-Garcia, Gisela Barrera-Badillo, Lucia Hernandez-Rivas, Irma Lopez-Martinez, Ernesto Ramirez-Gonzalez. |
| EPI_ISL_2676629 | DYOMEDEA-LABORATOIRE DE LA SAUVEGARDE | CNR Virus des Infections Respiratoires - France SUD | Antonin Bal, Gregory Destras, Gwendolyne Burfin, Hadrien Regue, Quentin Semanas, Martine Valette, Bruno Lina, Laurence Josset |
| EPI_ISL_2678714 | MVZ Labor Dr. Limbach & Kollegen GbR | Robert Koch Institute | unknown |
| EPI_ISL_2680902, EPI_ISL_2680903, EPI_ISL_2680904, EPI_ISL_2680905, EPI_ISL_2680906, EPI_ISL_2680907, EPI_ISL_2680908, EPI_ISL_2680909, EPI_ISL_2680910, EPI_ISL_2680911, EPI_ISL_2680912, EPI_ISL_2680914, EPI_ISL_2680915 | | | |
| see above | HLAGYN - Laboratorio de Imunologia de Transplantes de Goias | HLAGYN - Laboratorio de Imunologia de Transplantes de Goias | Fernando Antonio Vinhal dos Santos, Erika Lopes Rocha Batista, Alessandro Leonardo Alvares Magalhaes, Frederico Rodrigues Vinhal, Sabrina Sara Moreira Duarte, Lucas Carlos Gomes Pereira, Daniel Ferreira de Sousa, Danielle de Paiva Rezende |
| EPI_ISL_2681091, EPI_ISL_2681109 | Unidad de Investigación Médica de Yucatán (UIMY) | Instituto de Biotecnología de la UNAM | Consorcio Mexicano de Vigilancia Genómica (CoViGen-Mex). Authors (in alphabetical order): Julio Elias Alvarado-Yaah, Carlos F. Arias, Santiago Ávila-Ríos, Eduardo Becerril-Vargas, Víctor Hugo Borja-Aburto, Celia Boukadida, Cristóbal Cháidez-Quiróz, Juan Bautista Chale-Dzul, Ricardo Ciria Merce, Andreu Comas-García, Célida Duque Molina, Julissa Enciso-Ibarra, José Antonio Enciso-Moreno, Gloria Elena Espinosa-Ayala, Fernando Fontove-Herrera, Daniel Fregoso-Rueda, Victor Eduardo García-Arias, Alejandra García-Gasca, Bruno Gómez-Gil, Jean Pierre González, Irvin González-López, Concepción Grajales-Muñiz, Ricardo Grande, Rosa María Gutiérrez Rios, Jesús Hernández, Alejandra Hernández-Terán, Alfredo Herrera-Estrella, Carla Ivón Herrera-Najera, Pavel Isa, Verónica Mata-Haro, Daniel Lira Morales, Susana Lopez, Antonio Loza Román, Brenda Irasema Maldonado-Meza, Bernardo Martínez-Miguel, José Arturo Martínez-Orozco, Célida Martínez- Rodríguez, Margarita Matías-Florentino, Fidencio Mejía-Nepomuceno, María Guadalupe de Jesús Mireles-Rivera, Gloria María Molina-Salinas, Hector Montoya-Fuentes, , Mario Mújica-Sánchez, José Esteban Muñoz-Medina, José de Jesús Nuñez-Contreras, Alicia Ocaña-Mondragón, Luis Alberto Ochoa-Carrera, Hector Esteban Paz-Juárez, Marissa Perez-Garcia, Francisco Pulido, Helen Haydee Fernanda Ramírez-Plascencia, Jorge Salas-Hernández, Angel Gustavo Salas-Lais, Alejandro Sánchez-Flores, Clara Esperanza Santacruz-Tinoco, María Guadalupe Santiago-Mauricio, Selene Zárate, Nelly Sélem-Mojica, Blanca Taboada, Gloria Vazquez, Joel Armando Vázquez-Pérez. |
| EPI_ISL_2681457 | University Hospitals of Geneva, Laboratory of Virology | HUG, Laboratory of Virology and the Health2030 Genome Center | Samuel Cordey, Ana Rita Goncalves, Laurent Kaiser, Lorenzo Cerutti, Henri Pegeot, Melyssa Elies, Deborah Penet, Keith Harshman, Ioannis Xenarios, Emmanouil Dermitzakis |
| EPI_ISL_2685850, EPI_ISL_2685878 | SYNLAB | GIGA Medical Genomics | Keith Durkin, Maria Artesi, Sébastien Bontems, Raphaël Boreux, Bouchra Boujemla, Nathalie Renotte, Cécile Meex, Pierrette Melin, Marie-Pierre Hayette, Vincent Bours |
| EPI_ISL_2685890, EPI_ISL_2685891 | Department of Clinical Microbiology | GIGA Medical Genomics | Keith Durkin, Maria Artesi, Sébastien Bontems, Raphaël Boreux, Bouchra Boujemla, Nathalie Renotte, Cécile Meex, Pierrette Melin, Marie-Pierre Hayette, Vincent Bours |
| EPI_ISL_2686065 | Azienda USL Umbria 2 | Istituto Zooprofilattico Sperimentale dell'Abruzzo e Molise "G. Caporale" | Proietti A, Pistoni E, Lorusso A, Marcacci M, Di Domenico M, Ancora M, Curini V, Mangone I, Rinaldi A, Delli Compagni E, Scialabba S, Di Pasquale A, Cammà C, Puglia I, Calistri P, Savini G |
| EPI_ISL_2686359, EPI_ISL_2686360, EPI_ISL_2686361, EPI_ISL_2686362, EPI_ISL_2686363, EPI_ISL_2686364, EPI_ISL_2686365, EPI_ISL_2686366, EPI_ISL_2686367, EPI_ISL_2686368, EPI_ISL_2686369, EPI_ISL_2686370, EPI_ISL_2686371, EPI_ISL_2686372, EPI_ISL_2686373, EPI_ISL_2686374, EPI_ISL_2686375, EPI_ISL_2686376, EPI_ISL_2686377, EPI_ISL_2686378, EPI_ISL_2686379, EPI_ISL_2686380, EPI_ISL_2686381, EPI_ISL_2686382, EPI_ISL_2686383, EPI_ISL_2686384, EPI_ISL_2686386, EPI_ISL_2686387, EPI_ISL_2686388 | | | |
| see above | Quest Diagnostics Incorporated | Centers for Disease Control and Prevention Division of Viral Diseases, Pathogen Discovery | Dakota Howard, Dhwani Batra, Peter W. Cook, Kara Moser, Adrian Paskey, Jason Caravas, Benjamin Rambo-Martin, Shatavia Morrison, Christopher Gulvick, Scott Sammons, Yvette Unoarumhi, Darlene Wagner, Matthew Schmerer, S. H. Rosenthal, A. Gerasimova, R. M. Kagan, B. Anderson, M. Hua, Y. Liu, L.E. Bernstein, K.E. Livingston, A. Perez, I. A. Shlyakhter, R. V. Rolando, R. Owen, P. Tanpaiboon, F. Lacbawan, Clinton R. Paden, Duncan MacCannell |
| EPI_ISL_2686408, EPI_ISL_2686431, EPI_ISL_2686436, EPI_ISL_2686449, EPI_ISL_2686452, EPI_ISL_2686552, EPI_ISL_2686554, EPI_ISL_2686555, EPI_ISL_2686561, EPI_ISL_2686573, EPI_ISL_2686609, EPI_ISL_2686623, EPI_ISL_2686632, EPI_ISL_2686651, EPI_ISL_2686717, EPI_ISL_2686720, EPI_ISL_2686732, EPI_ISL_2686733, EPI_ISL_2686762, EPI_ISL_2686764, EPI_ISL_2686777, EPI_ISL_2686790, EPI_ISL_2686793, EPI_ISL_2686795, EPI_ISL_2686797, EPI_ISL_2686799 | | | |
| see above | Fulgent Genetics | Centers for Disease Control and Prevention Division of Viral Diseases, Pathogen Discovery | Dakota Howard, Dhwani Batra, Peter W. Cook, Kara Moser, Adrian Paskey, Jason Caravas, Benjamin Rambo-Martin, Shatavia Morrison, Christopher Gulvick, Scott Sammons, Yvette Unoarumhi, Darlene Wagner, Matthew Schmerer, Harry Gao, Mickey Li, John Gao, Joseph Fierro, Benafsh Sapra, Becky Tsai, Yan Meng, Doreen Ng, James Xie, Clinton R. Paden, Duncan MacCannell |
| EPI_ISL_2686856, EPI_ISL_2686857, EPI_ISL_2686880, EPI_ISL_2686883, EPI_ISL_2686889 | Infinity Biologix | Centers for Disease Control and Prevention Division of Viral Diseases, Pathogen Discovery | Dakota Howard, Dhwani Batra, Peter W. Cook, Kara Moser, Adrian Paskey, Jason Caravas, Benjamin Rambo-Martin, Shatavia Morrison, Christopher Gulvick, Scott Sammons, Yvette Unoarumhi, Darlene Wagner, Matthew Schmerer, Christian Bixby, Yihe Wang, Jonathan Schultz, Chirayu Goswami, Russ Hager, Robin Grimwood, Clinton R. Paden, Duncan MacCannell |
| EPI_ISL_2686906, EPI_ISL_2686908, EPI_ISL_2686911, EPI_ISL_2686917, EPI_ISL_2686919, EPI_ISL_2686920, EPI_ISL_2686921, EPI_ISL_2686950, EPI_ISL_2686962, EPI_ISL_2686974, EPI_ISL_2686981, EPI_ISL_2686982, EPI_ISL_2686984, EPI_ISL_2686985, EPI_ISL_2686989, EPI_ISL_2686997, EPI_ISL_2687015, EPI_ISL_2687021, EPI_ISL_2687034, EPI_ISL_2687035, EPI_ISL_2687046, EPI_ISL_2687047, EPI_ISL_2687057, EPI_ISL_2687058, EPI_ISL_2687063, EPI_ISL_2687094, EPI_ISL_2687095, EPI_ISL_2687096, EPI_ISL_2687098, EPI_ISL_2687120, EPI_ISL_2687121, EPI_ISL_2687136, EPI_ISL_2687162, EPI_ISL_2687165, EPI_ISL_2687179, EPI_ISL_2687186, EPI_ISL_2687196, EPI_ISL_2687202, EPI_ISL_2687205, EPI_ISL_2687213, EPI_ISL_2687242, EPI_ISL_2687243, EPI_ISL_2687248, EPI_ISL_2687253, EPI_ISL_2687265, EPI_ISL_2687276, EPI_ISL_2687287, EPI_ISL_2687292, EPI_ISL_2687301, EPI_ISL_2687303, EPI_ISL_2687308, EPI_ISL_2687321, EPI_ISL_2687326, EPI_ISL_2687378, EPI_ISL_2687380, EPI_ISL_2687381, EPI_ISL_2687388, EPI_ISL_2687403, EPI_ISL_2687407, EPI_ISL_2687408, EPI_ISL_2687413, EPI_ISL_2687416, EPI_ISL_2687419, EPI_ISL_2687444, EPI_ISL_2687455, EPI_ISL_2687488, EPI_ISL_2687489, EPI_ISL_2687499, EPI_ISL_2687503, EPI_ISL_2687551, EPI_ISL_2687554, EPI_ISL_2687558, EPI_ISL_2687559 | | | |
| see above | Aegis Sciences Corporation | Centers for Disease Control and Prevention Division of Viral Diseases, Pathogen Discovery | Dakota Howard, Dhwani Batra, Peter W. Cook, Kara Moser, Adrian Paskey, Jason Caravas, Benjamin Rambo-Martin, Shatavia Morrison, Christopher Gulvick, Scott Sammons, Yvette Unoarumhi, Darlene Wagner, Matthew Schmerer, Cyndi Clark, Patrick Campbell, Rob Case, Vikramsinha Ghorpade, Holly Houdeshell, Ola Kvalvaag, Dillon Nall, Ethan Sanders, Alec Vest, Shaun Westlund, Matthew Hardison, Clinton R. Paden, Duncan MacCannell |
| EPI_ISL_2688819 | Massachusetts State Public Health Laboratory | Massachusetts State Public Health Laboratory | Andrew Lang, Timelia Fink, Glen Gallagher, Sandra Smole |
| EPI_ISL_2689682 | Nebraska Public Health Laboratory | NPHL COVID-19 Response Team | NPHL COVID-19 Response Team |
| EPI_ISL_2690127 | MT Public Health Laboratory | Centers for Disease Control and Prevention Division of Viral Diseases, Pathogen Discovery | Mili Sheth, Sarah Nobles, Jasmine Padilla, Mark Burroughs, Shoshona Le, Katie Dillon, Peter Cook, Clinton R. Paden, Dhwani Batra, Krista Queen, Kristen Knipe, Dakota Howard, Yvette Unoarumhi, Darlene Wagner, Matthew Schmerer, Ben L. Rambo-Martin, Kristine Lacek, Sam Shepard, Alison Laufer Halpin, Dave Wentworth, Vivien Dugan, Suxiang Tong, Justin Lee |
| EPI_ISL_2690251 | University of Mississippi Medical Center, Department of Pathology | University of Mississippi Medical Center, Molecular and Genomics Core Facility | Ashley C. Johnson, Wenjie Wu, Ithiel J. Frame, Krishna K. Ayyalasomayajula, Michael R. Garrett, D. Ashley Robinson |
| EPI_ISL_2690272 | Nevada State Public Health Laboratory | Nevada State Public Health Laboratory | Andrew Gorzalski, Mark Pandori |
| EPI_ISL_2691501, EPI_ISL_2691502, EPI_ISL_2691503, EPI_ISL_2691504, EPI_ISL_2691511, EPI_ISL_2691512, EPI_ISL_2691517, EPI_ISL_2691518, EPI_ISL_2691520, EPI_ISL_2691521, EPI_ISL_2691522, EPI_ISL_2691525, EPI_ISL_2691533, EPI_ISL_2691547, EPI_ISL_2691548, EPI_ISL_2691549, EPI_ISL_2691550, EPI_ISL_2691551, EPI_ISL_2691552, EPI_ISL_2691568 | | | |
| see above | Laboratorio Central Noel Nutels | Bioinformatics Laboratory / LNCC | Luiz G P de Almeida, Alessandra P Lamarca, Ronaldo da Silva F Jr, Liliane Cavalcante, Alexandra L Gerber, Ana Paula de C Guimaraes, Douglas Terra Machado, Cassia Alves, Diana Mariani, Cintia Policarpo, Gleidson da Silva de Oliveira, Mario Sergio Ribeiro, Silvia Carvalho, Flavio Dias da Silva, Marcio Henrique de Oliveira Garcia, Leandro Magalhaes de Souza, Cristiane Gomes da Silva, Caio Luiz Pereira Ribeiro, Andrea Cony Cavalcanti, Claudia Maria Braga de Mello, Amilcar Tanuri, Ana Tereza R Vasconcelos |
| EPI_ISL_2691573 | Unidade de apoio ao diagnostico da COVID - UNADIG | Bioinformatics Laboratory / LNCC | Luiz G P de Almeida, Alessandra P Lamarca, Ronaldo da Silva F Jr, Liliane Cavalcante, Alexandra L Gerber, Ana Paula de C Guimaraes, Douglas Terra Machado, Cassia Alves, Diana Mariani, Cintia Policarpo, Gleidson da Silva de Oliveira, Mario Sergio Ribeiro, Silvia Carvalho, Flavio Dias da Silva, Marcio Henrique de Oliveira Garcia, Leandro Magalhaes de Souza, Cristiane Gomes da Silva, Caio Luiz Pereira Ribeiro, Andrea Cony Cavalcanti, Claudia Maria Braga de Mello, Amilcar Tanuri, Ana Tereza R Vasconcelos |

| | | | |
|---|---|---|---|
| EPI_ISL_2692541, EPI_ISL_2692544, EPI_ISL_2692549, EPI_ISL_2692552, EPI_ISL_2692554, EPI_ISL_2692556, EPI_ISL_2692560, EPI_ISL_2692562, EPI_ISL_2692564, EPI_ISL_2692565, EPI_ISL_2692566, EPI_ISL_2692572, EPI_ISL_2692580, EPI_ISL_2692584, EPI_ISL_2692587, EPI_ISL_2692590, EPI_ISL_2692593, EPI_ISL_2692614, EPI_ISL_2692619, EPI_ISL_2692622, EPI_ISL_2692626, EPI_ISL_2692629, EPI_ISL_2692632, EPI_ISL_2692634, EPI_ISL_2692638, EPI_ISL_2692643, EPI_ISL_2692644, EPI_ISL_2692648, EPI_ISL_2692653, EPI_ISL_2692654, EPI_ISL_2692658, EPI_ISL_2692659, EPI_ISL_2692661, EPI_ISL_2692667, EPI_ISL_2692669, EPI_ISL_2692670, EPI_ISL_2692673, EPI_ISL_2692677, EPI_ISL_2692678, EPI_ISL_2692680, EPI_ISL_2692685, EPI_ISL_2692690, EPI_ISL_2692692, EPI_ISL_2692699 | | | |
| see above | Salud Digna | Instituto Nacional de Medicina Genomica | Cedro-Tanda A, Hidalgo-Miranda A, Mendoza-Vargas A, Reyes-Grajeda JP, Abraham Campos-Romero, Moreno-Camacho José Luis, Rodriguez-Gallegos Jorge, Luna-Ruiz Marco, Gonzalez-Barrera D, Rangel-DeLeon D, Munguia-Garza P, Ramirez-Vega O, Escobar-Arrazola, M, Herrera-Montalvo LA. |
| EPI_ISL_2693129, EPI_ISL_2693139, EPI_ISL_2693277, EPI_ISL_2693278, EPI_ISL_2693299, EPI_ISL_2693301, EPI_ISL_2693307, EPI_ISL_2693310, EPI_ISL_2693312 | UW Virology Lab | UW Virology Lab | Pavitra Roychoudhury, Hong Xie, Lasata Shrestha, Tien V. Nguyen, Shah Mohamed Bakhash, Michelle Lin, Noah R. Baker, Ricardo Perez, Sean Ellis, Nathan Breit, Robert J. Livingston, Meei-Li Huang, Keith R Jerome, Patrick Mathias, Alexander Greninger |
| EPI_ISL_2693684, EPI_ISL_2693739, EPI_ISL_2693746, EPI_ISL_2693753, EPI_ISL_2693763, EPI_ISL_2693801, EPI_ISL_2693813 | UW Virology Lab | UW Virology Lab | Pavitra Roychoudhury, Hong Xie, Lasata Shrestha, Shah Mohamed Bakhash, Tien V. Nguyen, Noah R. Baker, Sean Ellis, Meei-Li Huang, Keith R Jerome, Alexander Greninger |
| EPI_ISL_2694805, EPI_ISL_2694858 | Labor Berlin Charite Vivantes GmbH / Institut fur Virologie | Charite Universitatsmedizin Berlin, Institut fur Virologie/Labor Berlin | Peter Menzel, Christine Stephan, Rolf Schwarzer, Victor M Corman, Barbara Muhlemann, Terry Jones, Christian Drosten |
| EPI_ISL_2695098 | CHULN - H Santa Maria | Instituto Nacional de Saude (INSA) | Borges et al |
| EPI_ISL_2695275 | H Braga | Instituto Nacional de Saude (INSA) | Borges et al |
| EPI_ISL_2695325, EPI_ISL_2695326 | ULSM - Matosinhos | Instituto Nacional de Saude (INSA) | Borges et al |
| EPI_ISL_2695345 | CHEDV | Instituto Nacional de Saude (INSA) | Borges et al |
| EPI_ISL_2695390, EPI_ISL_2695391 | CH Tondela Viseu | Instituto Nacional de Saude (INSA) | Borges et al |
| EPI_ISL_2695394 | Lab La Salete Robles - VN Famalicao | Instituto Nacional de Saude (INSA) | Borges et al |
| EPI_ISL_2695447 | ARS Algarve - Laboratorio Laura Ayres | Instituto Nacional de Saude (INSA) | Borges et al |
| EPI_ISL_2709012 | UMC Groningen, Clinical Virology, Department of Medical Microbiology and Infection Prevention | UMC Groningen, Clinical Virology, Department of Medical Microbiology and Infection Prevention | Hubert Niesters, Alexander Friedrich, Erley Lizarazo-Forero, Monika Fliss, Lilli Gard, Sigrid Rosema, Coretta Van Leer-Buter, Xuewei Zhou, Marjolein Knoester |
| EPI_ISL_2709231 | Public Health Laboratory, Minnesota Department of Health | University of Minnesota Genomics Center | Daryl M. Gohl, John Garbe, Jaquelyn Kuriger-Laber, Corbin Dirkx, and Sean Wang |
| EPI_ISL_2709498, EPI_ISL_2709517, EPI_ISL_2709541, EPI_ISL_2709543 | Aegis Sciences Corporation | Centers for Disease Control and Prevention Division of Viral Diseases, Pathogen Discovery | Dakota Howard, Dhwani Batra, Peter W. Cook, Kara Moser, Adrian Paskey, Jason Caravas, Benjamin Rambo-Martin, Shatavia Morrison, Christopher Gulvick, Scott Sammons, Yvette Unoarumhi, Darlene Wagner, Matthew Schmerer, Cyndi Clark, Patrick Campbell, Rob Case, Vikramsinha Ghorpade, Holly Houdeshell, Ola Kvalvaag, Dillon Nall, Ethan Sanders, Alec Vest, Shaun Westlund, Matthew Hardison, Clinton R. Paden, Duncan MacCannell |
| EPI_ISL_2709816 | MD PHL | MD PHL | Maryland Department of Health Laboratories Administration |
| EPI_ISL_2712536, EPI_ISL_2712537 | Sharp HealthCare Laboratory | Andersen lab at Scripps Research | SEARCH Alliance San Diego with Aaron Harding, Jacquelyn Berumen, Cathy Woerle, Liam McGinnis, Art Mendoza, Omid Bakhtar |
| EPI_ISL_2712921 | Wyoming Public Health Laboratory | Wyoming Public Health Laboratory | Jim Mildenberger, Taylor Fearing, Channing Weber, Ashley Norberg, Chayse Rowley, Marley Goetz, Brian Dominguez, Elliot Thomasson, Sam Britz, Cari Sloma, Robert Petit, and Rob Christensen |
| EPI_ISL_2716293 | Kansas Health and Environmental Lab | Kansas Health and Environmental Lab | Katherine Wiggins, Mike Grose, Jonathan Barnell, Ben Olsen, and Phil Adam |
| EPI_ISL_2716405, EPI_ISL_2716406, EPI_ISL_2716407, EPI_ISL_2716411 | Arizona State Public Health Laboratory | Arizona State Public Health Laboratory | Trung Huynh, Jessica Escobar, Katherine Fullerton, Nobuko Fukushima, Matthew Contursi, Stacy White, Linda Getsinger, Victor Waddell |
| EPI_ISL_2716463, EPI_ISL_2716464, EPI_ISL_2716466 | Washington State Department of Health Public Health Laboratories | Washington State Department of Health Public Health Laboratories | Drew MacKellar, Philip Dykema, Denny Russell, Joenice Gonzalez, Hannah Gray, Geoff Melly, Vanessa De Los Santos, Darren Lucas, JohnAric Peterson, Avi Singh, Rebecca Cao |
| EPI_ISL_2716479, EPI_ISL_2716488, EPI_ISL_2716734 | UW Virology Lab | UW Virology Lab | Pavitra Roychoudhury, Hong Xie, Lasata Shrestha, Shah Mohamed Bakhash, Tien V. Nguyen, Noah R. Baker, Sean Ellis, Meei-Li Huang, Keith R Jerome, Alexander Greninger |
| EPI_ISL_2716912, EPI_ISL_2716924, EPI_ISL_2716935 | Southern Nevada Public Health Laboratory | Southern Nevada Public Health Laboratory | Michael Picker |
| EPI_ISL_2724792, EPI_ISL_2724855, EPI_ISL_2724856, EPI_ISL_2724857, EPI_ISL_2724858, EPI_ISL_2724862, EPI_ISL_2724869, EPI_ISL_2724873, EPI_ISL_2724876, EPI_ISL_2724880, EPI_ISL_2724885, EPI_ISL_2724889, EPI_ISL_2724891, EPI_ISL_2724899, EPI_ISL_2724900, EPI_ISL_2724901, EPI_ISL_2724902 | | | |
| see above | State Laboratories Division, Hawaii State Department of Health | State Laboratories Division, Hawaii State Department of Health | Pamela O'Brien, Drew Kuwazaki, Ayana Garnet, Razvan Sultana, Edward Desmond |
| EPI_ISL_2725184, EPI_ISL_2725186, EPI_ISL_2725190, EPI_ISL_2725204, EPI_ISL_2725206, EPI_ISL_2725209, EPI_ISL_2725210, EPI_ISL_2725212, EPI_ISL_2725213, EPI_ISL_2725223, EPI_ISL_2725229, EPI_ISL_2725238, EPI_ISL_2725239, EPI_ISL_2725245, EPI_ISL_2725248, EPI_ISL_2725249, EPI_ISL_2725250, EPI_ISL_2725252, EPI_ISL_2725256, EPI_ISL_2725257, EPI_ISL_2725263, EPI_ISL_2725265, EPI_ISL_2725266, EPI_ISL_2725268, EPI_ISL_2725271, EPI_ISL_2725275, EPI_ISL_2725276, EPI_ISL_2725281, EPI_ISL_2725282, EPI_ISL_2725294 | | | |
| see above | Lab. Microbiologia e Virologia Cotugno A.O. dei Colli - Istituto Zooprofilattico Sperimentale del Mezzogiorno | Telethon Institute of Genetics and Medicine (TIGEM) | Antonio Grimaldi Patrizia Annunziata Francesco Panariello Biancamaria Pierri Claudia Tiberio Teresa Giuliano Valentina Bouche Chiara Colantuono Maria Concetta Cuomo Denise Di Concilio Lucio Di Filippo Anna Manfredi Marcello Salvi Antonio Limone Luigi Atripaldi Pellegrino Cerino Andrea Ballabio Davide Cacchiarelli |
| EPI_ISL_2726819 | Wyoming Public Health Laboratory | Wyoming Public Health Laboratory | Jim Mildenberger, Taylor Fearing, Channing Weber, Ashley Norberg, Chayse Rowley, Marley Goetz, Brian Dominguez, Elliot Thomasson, Sam Britz, Cari Sloma, Robert Petit, and Rob Christensen |
| EPI_ISL_2726937, EPI_ISL_2726940 | ISDH | Quantigen Biosciences | Paul J. Childress |
| EPI_ISL_2727766, EPI_ISL_2727767, EPI_ISL_2727775, EPI_ISL_2727782, EPI_ISL_2727792, EPI_ISL_2727817, EPI_ISL_2727826 | Wisconsin State Laboratory of Hygiene Communicable Disease Division | Wisconsin State Laboratory of Hygiene Communicable Disease Division | Abigail C. Shockey, Alicia J. Mooney, Erika M. Hanson, Tonya Danz, Richard Griesser, Sara Wagner, Kelsey R. Florek |
| EPI_ISL_2727868 | HealthPartners Central Lab | Minnesota Department of Health, Public Health Laboratory | Alexandra Lorentz, Jacob Garfin, Matt Plumb, and Xiong Wang |
| EPI_ISL_2731453, EPI_ISL_2731454, EPI_ISL_2731455, EPI_ISL_2731456 | Laboratorio Central de Saude Publica do Estado do Para (LACEN/PA) | Laboratory of Respiratory Viruses and Measles, Oswaldo Cruz Institute, FIOCRUZ | Paola Resende, Luciana Appolinario, Fernando Motta, Anna Carolina Paixao, Ana Carolina Mendonca, Alice Sampaio Rocha, Taina Venas, Elisa Cavalcante Pereira, Renata Serrano Lopes, Valnete Andrade, Marilda Siqueira on behalf of the Fiocruz COVID-19 Genomic Surveillance Network |
| EPI_ISL_2736280 | Atlas Genomics | Seattle Flu Study | Deborah A. Nickerson, Chris D. Frazar, Jover Lee, Benjamin Pelle, Erica Ryke, Matthew Richardson, Amanda Adler, Elisabeth Brandstetter, Peter D. Han, Kairsten Fay, Misja Ilcisin, Kirsten Lacombe, Thomas R. Sibley, Melissa Truong, Caitlin R. Wolf, Romesh Gautom, Geoff Melly, Brian Hiatt, Philip Dykema, Scott Lindquist, Michael Boeckh, Janet A. Englund, Michael Famulare, Barry R. Lutz, Mark J. Rieder, Lea M. Starita, Matthew Thompson, Helen Y. Chu, Jay Shendure, Trevor Bedford |
| EPI_ISL_2753648 | Missouri State Public Health Laboratory | Missouri State Public Health Laboratory | Matthew Sinn, Joshua Barry, Ashley New |
| EPI_ISL_2753941 | NYU Langone Health | Departments of Pathology and Medicine, New York University School of Medicine | Adriana Heguy, Dacia Dimartino, Emily Guzman, Christian Marier, Peter Meyn, Sitharam Ramaswami, Gael Westby, Paul Zappile, Yutong Zhang, Paolo Cotzia, Guiqing Wang |
| EPI_ISL_2754064 | Wyoming Public Health Laboratory | Wyoming Public Health Laboratory | Jim Mildenberger, Taylor Fearing, Channing Weber, Ashley Norberg, Chayse Rowley, Marley Goetz, Brian Dominguez, Elliot Thomasson, Sam Britz, Cari Sloma, Robert Petit, and Rob Christensen |
| EPI_ISL_2754305 | Arizona State Public Health Laboratory | Arizona State Public Health Laboratory | Trung Huynh, Jessica Escobar, Katherine Fullerton, Nobuko Fukushima, Matthew Contursi, Stacy White, Linda Getsinger, Victor Waddell |

| | | | |
|---|---|---|---|
| EPI_ISL_2756057, EPI_ISL_2756058, EPI_ISL_2756060, EPI_ISL_2756061, EPI_ISL_2756062, EPI_ISL_2756063, EPI_ISL_2756064, EPI_ISL_2756066, EPI_ISL_2756067, EPI_ISL_2756068, EPI_ISL_2756069, EPI_ISL_2756070, EPI_ISL_2756071, EPI_ISL_2756072, EPI_ISL_2756073, EPI_ISL_2756077, EPI_ISL_2756078, EPI_ISL_2756079, EPI_ISL_2756080, EPI_ISL_2756081, EPI_ISL_2756082, EPI_ISL_2756085, EPI_ISL_2756088, EPI_ISL_2756092, EPI_ISL_2756094, EPI_ISL_2756095, EPI_ISL_2756097, EPI_ISL_2756100, EPI_ISL_2756101, EPI_ISL_2756104, EPI_ISL_2756105, EPI_ISL_2756106, EPI_ISL_2756108, EPI_ISL_2756109, EPI_ISL_2756112, EPI_ISL_2756114, EPI_ISL_2756115, EPI_ISL_2756116, EPI_ISL_2756119, EPI_ISL_2756121, EPI_ISL_2756122, EPI_ISL_2756124, EPI_ISL_2756125, EPI_ISL_2756126, EPI_ISL_2756128, EPI_ISL_2756129, EPI_ISL_2756130, EPI_ISL_2756134, EPI_ISL_2756138, EPI_ISL_2756139, EPI_ISL_2756140, EPI_ISL_2756141, EPI_ISL_2756142, EPI_ISL_2756143, EPI_ISL_2756144, EPI_ISL_2756147, EPI_ISL_2756148, EPI_ISL_2756149, EPI_ISL_2756172, EPI_ISL_2756173, EPI_ISL_2756175, EPI_ISL_2756180, EPI_ISL_2756181, EPI_ISL_2756182, EPI_ISL_2756183, EPI_ISL_2756184, EPI_ISL_2756185, EPI_ISL_2756186, EPI_ISL_2756187, EPI_ISL_2756188, EPI_ISL_2756189, EPI_ISL_2756190, EPI_ISL_2756191, EPI_ISL_2756192, EPI_ISL_2756193, EPI_ISL_2756194, EPI_ISL_2756195 | | | |
| see above | Genetica Molecular and Subdepartamento de Virologia ISP Chile | Instituto de Salud Publica de Chile | Karen Orostica, Constanza Campano, Barbara Parra, Loredana Arata, Gisselle Barra, Patricia Bustos, Rodrigo Fasce, Javier Tognarelli, Andres Castillo, Soledad Ulloa, Jorge Fernandez |
| EPI_ISL_2756212 | National Platform bis UMONS/Jolimont | National Platform bis UMONS/Jolimont | François Dufrasne, Guillaume Bayon-Vicente, Florian Juszczak, Eric Tarantino, Gautier Detry, Ruddy Wattiez |
| EPI_ISL_2756222, EPI_ISL_2756223, EPI_ISL_2756226, EPI_ISL_2756228, EPI_ISL_2756230, EPI_ISL_2756233, EPI_ISL_2756234, EPI_ISL_2756236, EPI_ISL_2756237, EPI_ISL_2756240, EPI_ISL_2756243, EPI_ISL_2756244, EPI_ISL_2756245, EPI_ISL_2756246, EPI_ISL_2756247, EPI_ISL_2756249, EPI_ISL_2756251, EPI_ISL_2756252, EPI_ISL_2756253, EPI_ISL_2756256, EPI_ISL_2756257, EPI_ISL_2756260, EPI_ISL_2756264, EPI_ISL_2756269, EPI_ISL_2756272, EPI_ISL_2756273, EPI_ISL_2756274, EPI_ISL_2756286, EPI_ISL_2756287, EPI_ISL_2756289, EPI_ISL_2756291, EPI_ISL_2756293, EPI_ISL_2756294, EPI_ISL_2756295, EPI_ISL_2756296, EPI_ISL_2756297, EPI_ISL_2756299, EPI_ISL_2756302, EPI_ISL_2756303, EPI_ISL_2756304, EPI_ISL_2756306, EPI_ISL_2756309, EPI_ISL_2756310, EPI_ISL_2756311, EPI_ISL_2756313, EPI_ISL_2756314, EPI_ISL_2756315, EPI_ISL_2756317, EPI_ISL_2756325, EPI_ISL_2756327, EPI_ISL_2756331, EPI_ISL_2756335, EPI_ISL_2756338 | | | |
| see above | Genetica Molecular and Subdepartamento de Virologia ISP Chile | Instituto de Salud Publica de Chile | Karen Orostica, Constanza Campano, Barbara Parra, Loredana Arata, Gisselle Barra, Patricia Bustos, Rodrigo Fasce, Javier Tognarelli, Andres Castillo, Soledad Ulloa, Jorge Fernandez |
| EPI_ISL_2758010, EPI_ISL_2758012 | Austrian Agency for Health and Food Safety (AGES) | Bergthaler laboratory, CeMM Research Center for Molecular Medicine of the Austrian Academy of Sciences | Lukas Endler, Anna Schedl, Fabian Amman, Petr Triska, Matthew Thornton, Thomas Penz, Benedikt Agerer, Maelle Le Moing, Michael Schuster, Bekir Erguner, Jan Laine, Martin Senekowitsch, Christoph Bock, Andreas Bergthaler |
| EPI_ISL_2758372, EPI_ISL_2758373 | Berkeley Medical Center | WVU and Marshall University Combined Genomics Core Facilities | James Denvir, Peter Stoilov, Peter Perrotta, Wesley Kimble, Ryan Percifield |
| EPI_ISL_2758565, EPI_ISL_2758594 | Servicio de Microbiologia Hospital Ramon y Cajal | Servicio de Microbiologia Hospital Ramon y Cajal | Ponce M, Galan JC, Martinez L. Abreu M, y Gonzalez-Alba JM |
| EPI_ISL_2758925 | The Ohio State University Applied Microbiology Services Laboratory | The Ohio State University Applied Microbiology Services Laboratory | Seth A. Faith PhD |
| EPI_ISL_2759880 | Washington State Department of Health Public Health Laboratories | Washington State Department of Health Public Health Laboratories | Drew MacKellar, Philip Dykema, Denny Russell, Joenice Gonzalez, Hannah Gray, Geoff Melly, Vanessa De Los Santos, Darren Lucas, JohnAric Peterson, Avi Singh, Rebecca Cao |
| EPI_ISL_2761224 | Sonic - Bioscientia - MVZ Labor Saar GmbH | Robert Koch Institute | unknown |
| EPI_ISL_2761541 | LabKom - Labor Hannover MVZ GmbH | Robert Koch Institute | unknown |
| EPI_ISL_2762312, EPI_ISL_2762347, EPI_ISL_2762356, EPI_ISL_2762365, EPI_ISL_2762395, EPI_ISL_2762431 | Diagnosticum - Labor Neukirchen | Robert Koch Institute | unknown |
| EPI_ISL_2769118 | UniversitätsSpital Zürich | Institute of Medical Virology | Verena Kufner, Gabriela Ziltener, Maryam Zaheri, Stefan Schmutz, Annette Audigé, Maria Grünberg, Kevin Steiner, Jon Huder, Cyril Shah, Riccarda Capaul, Guido Bloemberg, Jürg Böni, Michael Huber, Alexandra Trkola |
| EPI_ISL_2769191 | TGen North | TGen North | "Jolene Bowers, Heather Centner, Brett Van Tassel, Chris French, Hayley Yaglom, Darrin Lemmer, Dave Engelthaler" |
| EPI_ISL_2773856 | Hospital Universitario Virgen de las Nieves de Granada-SAS | SeqCOVID-SPAIN consortium/IBV(CSIC) | Sara Sanbonmatsu Gámez, Irene Pedrosa Corral, José M. Navarro-Marí and SeqCOVID-SPAIN consortium |
| EPI_ISL_2774104 | Microbiology Department, University Hospital Donostia | Microbiology Department, University Hospital Donostia | Marimon JM, Montes M, Piñeiro L, Martin-Peñaranda T, Gomez M, Sorarrain A, Cilla G. |
| EPI_ISL_2774367, EPI_ISL_2774391 | Illinois Department of Public Health - Springfield Lab | Illinois Department of Public Health - Springfield Lab | Bryan Sim, Gordon McCall |
| EPI_ISL_2774572 | Torrance Memorial Medical Center | Los Angeles County PHL | P. Hemarajata et al. |
| EPI_ISL_2811913 | Laboratory Medicine, University of Washington | Laboratory Medicine, University of Washington | Castor,J.L. |

### supp. Table S7: Eta

We gratefully acknowledge the following Authors from the Originating laboratories responsible for obtaining the specimens, as well as the Submitting laboratories where the genome data were generated and shared via GISAID, on which this research is based.

All Submitters of data may be contacted directly via www.gisaid.org

Authors are sorted alphabetically.

| Accession ID | Originating Laboratory | Submitting Laboratory | Authors |
|---|---|---|---|
| EPI_ISL_1471863, EPI_ISL_1471954 | Pandemic Response Lab - NYC | Pandemic Response Lab, R&D | Henry Lee, Michael Hammerling, Melissa Hopkins, Cybill del Castillo, Shinyoung Clair Kang, William Ward, Pradeep Bugga, Sol Rey, Dylan Law, Haiping Hao, Jon Laurent |
| EPI_ISL_1477036, EPI_ISL_1477037 | National Public Health Laboratory, National Centre for Infectious Diseases | National Public Health Laboratory, National Centre for Infectious Diseases | Tze Minn Mak, Zhenyang Zhou, Grace Jie Yin Ngan, Royce Ang, Lin Cui, Raymond Tzer Pin Lin |
| EPI_ISL_1529030, EPI_ISL_1529047, EPI_ISL_1529049, EPI_ISL_1529060 | Berkeley Medical Center | WVU and Marshall University Combined Genomics Core Facilities | "James Denvir, Peter Stoilov, Peter Perrotta, Wesley Kimble, Ryan Percifield" |
| EPI_ISL_1531739 | School of Pharmacy, Shenandoah University | School of Pharmacy, Shenandoah University | Adams,S.M., Harralson,A.F., Kidd,R.S., Sawyer,G.W. |
| EPI_ISL_1534465, EPI_ISL_1534477, EPI_ISL_1534478 | Ministry of Health Turkey | Ministry of Health Turkey | Fatma Bayrakdar, Yasemin Cosgun, Suleyman Yalcin, Gulay Korukluoglu |
| EPI_ISL_1534725, EPI_ISL_1534731 | AZDelta | AZDelta | Geert Martens; Dieter De Smet |
| EPI_ISL_1536186 | Lighthouse Lab in Alderley Park | Wellcome Sanger Institute for the COVID-19 Genomics UK (COG-UK) Consortium | Jacquelyn Wynn, Mairead Hyland, The Lighthouse Lab in Alderley Park and Alex Alderton, Roberto Amato, Jeffrey Barrett, Sonia Goncalves, Ewan Harrison, David K. Jackson, Ian Johnston, Dominic Kwiatkowski, Cordelia Langford, John Sillitoe on behalf of the Wellcome Sanger Institute COVID-19 Surveillance Team |
| EPI_ISL_1542264, EPI_ISL_1543103 | Pandemic Response Lab - NYC | Pandemic Response Lab, R&D | Henry Lee, Michael Hammerling, Melissa Hopkins, Cybill del Castillo, Shinyoung Clair Kang, William Ward, Pradeep Bugga, Sol Rey, Dylan Law, Katharine Nelson, Haiping Hao, Jon Laurent |
| EPI_ISL_1547506 | Cerballiance Pont Audemer | Cerba Lab | Haïm-Boukobza S, Prigent A, Leriche A, Verdurme L, Trombert-Paolantoni S, Lecorche E, Roquebert B |
| EPI_ISL_1567223 | Limbach - MVZ Humangenetik Ulm | Robert Koch Institute | unknown |
| EPI_ISL_1571169, EPI_ISL_1571188 | Limbach - MVZ Labor Eveld & Kollegen Essen | Robert Koch Institute | unknown |
| EPI_ISL_1573183 | MVZ Labor Dr. Limbach & Kollegen GbR | Robert Koch Institute | unknown |
| EPI_ISL_1593082 | Helix/Illumina | Centers for Disease Control and Prevention Division of Viral Diseases, Pathogen Discovery | Dakota Howard, Dhwani Batra, Peter W. Cook, Kara Moser, Adrian Paskey, Jason Caravas, Benjamin Rambo-Martin, Shatavia Morrison, Christopher Gulvick, Scott Sammons, Yvette Unoarumhi, Darlene Wagner, Matthew Schmerer, Eileen de Feo, Jan Antico, Christine Tran, Matthew Tolentino, Shannon Wickline, Kim Gietzen, Brad Sickler, Jingtao Liu, Eric Allen, Phil Febbo, Nicole L. Washington, Simon White, Geraint Levan, Kelly Schiabor Barrett, Elizabeth Cirulli, Alexandre Bolze, Ary Ascencio, Charlotte Rivera-Garcia, Ryan Cho, Jason Nguyen, Sherry Wang, Jimmy Ramirez, Tyler Cassens, Efren Sandoval, Magnus Isaksson, William Lee, David Becker, Marc Laurent, James Lu, Clinton R. Paden, Duncan MacCannell |
| EPI_ISL_1595840 | Armed Forces Institute of Pathology (AFIP), Dhaka Cantonment | Genomic Research Lab, BCSIR | Md. Saddam Hossain, Md. Murshed Hasan Sarkar, Mohammad Samir Uzzaman, Eshrar Osman, Md. Ahasan Habib, Shahina Akter, Tanjina Akhter Banu, Abu Sayeed Mohammad Mahmud, Barna Goswami, Iffat Jahan, Mohammad Mohi Uddin, Md. Kamrul Islam, Mohammad Mizanur Rahman, Susane Giti, Md. Salim Khan |
| EPI_ISL_1601510 | UW Virology Lab | UW Virology Lab | Pavitra Roychoudhury, Hong Xie, Lasata Shrestha, Shah Mohamed Bakhash, Michelle Lin, Noah R. Baker, Sean Ellis, Saraswathi Sathees, Meei-Li Huang, Keith R Jerome, Alexander Greninger |
| EPI_ISL_1623609 | CH VALENCE | CNR Virus des Infections Respiratoires - France SUD | Antonin Bal, Gregory Destras, Gwendolyne Burfin, Hadrien Regue, Quentin Semanas, Martine Valette, Bruno Lina, Laurence Josset |
| EPI_ISL_1624779, EPI_ISL_1624804, EPI_ISL_1624823, EPI_ISL_1624837, EPI_ISL_1624845, EPI_ISL_1624878, EPI_ISL_1624884 | Berkeley Medical Center | WVU and Marshall University Combined Genomics Core Facilities | James Denvir, Peter Stoilov, Peter Perrotta, Wesley Kimble, Ryan Percifield |
| EPI_ISL_1624920 | QLabs | WVU and Marshall University Combined Genomics Core Facilities | James Denvir, Peter Stoilov, Peter Perrotta, Wesley Kimble, Ryan Percifield |
| EPI_ISL_1624963, EPI_ISL_1624964, EPI_ISL_1624966, EPI_ISL_1624968, EPI_ISL_1624971, EPI_ISL_1624981 | Berkeley Medical Center | WVU and Marshall University Combined Genomics Core Facilities | James Denvir, Peter Stoilov, Peter Perrotta, Wesley Kimble, Ryan Percifield |
| EPI_ISL_1628193 | UW Virology Lab | UW Virology Lab | Pavitra Roychoudhury, Hong Xie, Lasata Shrestha, Shah Mohamed Bakhash, Michelle Lin, Noah R. Baker, Sean Ellis, Meei-Li Huang, Keith R Jerome, Alexander Greninger |
| EPI_ISL_1632926, EPI_ISL_1632929, EPI_ISL_1632930, EPI_ISL_1632933 | Istituto Zooprofilattico Sperimentale del Mezzogiorno | Telethon Institute of Genetics and Medicine (TIGEM) | Antonio Grimaldi Patrizia Annunziata Francesco Panariello Biancamaria Pierri Claudia Tiberio Teresa Giuliano Valentina Bouche Chiara Colantuono Maria Concetta Cuomo Denise Di Concilio Lucio Di Filippo Anna Manfredi Marcello Salvi Antonio Limone Luigi Atripaldi Pellegrino Cerino Andrea Ballabio Davide Cacchiarelli |
| EPI_ISL_1633953, EPI_ISL_1634233, EPI_ISL_1636298 | Pandemic Response Lab - NYC | Pandemic Response Lab, R&D | Henry Lee, Michael Hammerling, Melissa Hopkins, Cybill del Castillo, Shinyoung Clair Kang, William Ward, Pradeep Bugga, Sol Rey, Dylan Law, Katharine Nelson, Haiping Hao, Jon Laurent |
| EPI_ISL_1636430 | Microbiology Department, Laboratori Clínic Metropolitana Nord. Hospital Universitari Germans Trias i Pujol. | Can Ruti SARS-CoV-2 Sequencing Hub (HUGTiP/IrsiCaixa/IGTP) | Marc Noguera-Julian, Pilar Armengol, Ignacio Blanco, Antoni E Bordoy, Francesc Catala-Moll, Pere-Joan Cardona, Maria Casadellà, Cristina Casañ, Gemma Clara, Bonaventura Clotet, Cristina Esteban, Montserrat Giménez, Mercedes Guerrero, Anna Not, Roger Paredes, Mariona Parera, Verónica Saludes, Alba Sánchez, and Elisa Martró on behalf of the Can Ruti SARS-CoV-2 Sequencing Hub. |
| EPI_ISL_1638287 | MDI Limbach Berlin GmbH; MVZ Labor Berlin | Robert Koch Institute | unknown |
| EPI_ISL_1638309 | Labor Prof. Dr. G. Enders MVZ GbR | Robert Koch Institute | unknown |
| EPI_ISL_1638315, EPI_ISL_1638326, EPI_ISL_1638343 | MDI Limbach Berlin GmbH; MVZ Labor Berlin | Robert Koch Institute | unknown |
| EPI_ISL_1638359 | MVZ Labor Dr. Limbach & Kollegen GbR | Robert Koch Institute | unknown |
| EPI_ISL_1638390, EPI_ISL_1638418 | Labor Prof. Dr. G. Enders MVZ GbR | Robert Koch Institute | unknown |
| EPI_ISL_1638423, EPI_ISL_1638447, EPI_ISL_1638454 | MDI Limbach Berlin GmbH; MVZ Labor Berlin | Robert Koch Institute | unknown |
| EPI_ISL_1640138 | Uniklinikum Carl Gustav Carus an der TU Dresden; Institut fÃ¼r Virologie | Robert Koch Institute | unknown |
| EPI_ISL_1640231 | Diagnosticum - Labor Neukirchen | Robert Koch Institute | unknown |

| | | | |
|---|---|---|---|
| EPI_ISL_1643785, EPI_ISL_1643786 | Labor Prof. Dr. G. Enders MVZ GbR | Robert Koch Institute | unknown |
| EPI_ISL_1643809 | Labor MÃ¶nchengladbach MVZ Dr. Stein + Kollegen GbR | Robert Koch Institute | unknown |
| EPI_ISL_1643861, EPI_ISL_1643867 | MDI Limbach Berlin GmbH; MVZ Labor Berlin | Robert Koch Institute | unknown |
| EPI_ISL_1653428 | Lighthouse Lab in Cambridge | Wellcome Sanger Institute for the COVID-19 Genomics UK (COG-UK) Consortium | Rob Howes, The Lighthouse Lab in Cambridge and Alex Alderton, Roberto Amato, Jeffrey Barrett, Sonia Goncalves, Ewan Harrison, David K. Jackson, Ian Johnston, Dominic Kwiatkowski, Cordelia Langford, John Sillitoe on behalf of the Wellcome Sanger Institute COVID-19 Surveillance Team |
| EPI_ISL_1653817 | Washington State Department of Health Public Health Laboratories | Washington State Department of Health Public Health Laboratories | Drew MacKellar, Philip Dykema, Denny Russell, Joenice Gonzalez, Hannah Gray, Geoff Melly, Vanessa De Los Santos, Darren Lucas, JohnAric Peterson, Avi Singh, Rebecca Cao |
| EPI_ISL_1659194 | Viollier AG | Department of Biosystems Science and Engineering, ETH Zurich | Chaoran Chen, Sarah Nadeau, Ivan Topolsky, Emmanouil Dermitzakis, Keith Harshman, Ioannis Xenarios, Henri Pegeot, Lorenzo Cerutti, Deborah Penet, Philipp Jablonski, Lara Fuhrmann, David Dreifuss, Katharina Jahn, Christiane Beckmann, Maurice Redondo, Olivier Kobel, Christoph Noppen, Sophie Seidel, Noemie Santamaria de Souza, Niko Beerenwinkel, Tanja Stadler |
| EPI_ISL_1663494 | Capital hospital, Bhubaneswar | Institute of Life Sciences - INSACOG | Sunil K. Raghav, Safal Walia, Arup Ghosh, Atimukta Jha, Amol M. Kanampalliwar, Shifu Aggarwal, Rupesh Dash, Rajeeb Swain, Punit Prasad, INSACOG Consortium, Ajay Parida |
| EPI_ISL_1669441 | Hospital Universitari Arnau de Vilanova | Hospital Universitari Vall d'Hebron - Vall d'Hebron Institut de Recerca | Cristina Andrés, Maria Piñana, Damir Garcia-Cehic, Ariadna Rando, Juliana Esperalba, Maria Gema Codina, Carla Castillo, Maria Carmen Martin, Tomàs Pumarola, Josep Quer, Andrés Antón |
| EPI_ISL_1670824 | UW Virology Lab | UW Virology Lab | Pavitra Roychoudhury, Hong Xie, Lasata Shrestha, Shah Mohamed Bakhash, Michelle Lin, Noah R. Baker, Sean Ellis, Meei-Li Huang, Keith R Jerome, Alexander Greninger |
| EPI_ISL_1671819 | CHU Lille | CHU Lille - Laboratoire de Virologie | AIT YAHYA Emilie, ALIDJINOU Enagnon Kazali, BOCKET Laurence, CREPIN Michel, DEMAY Christophe, ENGELMANN Ilka, GEFFROY Sandrine, GUIGON Aurélie, LAMBERT Valérie, LAZREK Mouna, NOBILLIAUX Florian, PREVOST Brigitte, THUILLIER Caroline, TINEZ Claire |
| EPI_ISL_1671823 | CH Armentières | CHU Lille - Laboratoire de Virologie | AIT YAHYA Emilie, ALIDJINOU Enagnon Kazali, BOCKET Laurence, CREPIN Michel, DEMAY Christophe, ENGELMANN Ilka, GEFFROY Sandrine, GUIGON Aurélie, LAMBERT Valérie, LAZREK Mouna, NOBILLIAUX Florian, PREVOST Brigitte, THUILLIER Caroline, TINEZ Claire |
| EPI_ISL_1672812, EPI_ISL_1672821, EPI_ISL_1672825, EPI_ISL_1672827 | Laboratorio Aziendale di Microbiologia e Virologia, Azienda Sanitaria dell'Alto Adige | Laboratorio Aziendale di Microbiologia e Virologia, Azienda Sanitaria dell'Alto Adige | Irene Bianconi, Bartolomeo Mobilio Rodriguez, Elisabetta Giacobazzi, Elisa Masi, Stefanie Wieser, Anne Picard, Claudia Volpato, Chiara Cantaloni, Elisabetta Pagani |
| EPI_ISL_1673424, EPI_ISL_1673587 | Istituto Zooprofilattico Sperimentale del Mezzogiorno | TIGEM | Antonio Grimaldi Patrizia Annunziata Francesco Panariello Biancamaria Pierri Claudia Tiberio Teresa Giuliano Valentina Bouche Chiara Colantuono Maria Concetta Cuomo Denise Di Concilio Lucio Di Filippo Anna Manfredi Marcello Salvi Antonio Limone Luigi Atripaldi Pellegrino Cerino Andrea Ballabio Davide Cacchiarelli |
| EPI_ISL_1680352 | UW Virology Lab | UW Virology Lab | Pavitra Roychoudhury, Hong Xie, Lasata Shrestha, Shah Mohamed Bakhash, Michelle Lin, Noah R. Baker, Sean Ellis, Meei-Li Huang, Keith R Jerome, Alexander Greninger |
| EPI_ISL_1692649, EPI_ISL_1692652 | Istituto Zooprofilattico Sperimentale del Mezzogiorno | Telethon Institute of Genetics and Medicine (TIGEM) | Antonio Grimaldi Patrizia Annunziata Francesco Panariello Biancamaria Pierri Claudia Tiberio Teresa Giuliano Valentina Bouche Chiara Colantuono Maria Concetta Cuomo Denise Di Concilio Lucio Di Filippo Anna Manfredi Marcello Salvi Antonio Limone Luigi Atripaldi Pellegrino Cerino Andrea Ballabio Davide Cacchiarelli |
| EPI_ISL_1695028, EPI_ISL_1695029 | Laboratory Corporation of America | Centers for Disease Control and Prevention Division of Viral Diseases, Pathogen Discovery | Dakota Howard, Dhwani Batra, Peter W. Cook, Kara Moser, Adrian Paskey, Jason Caravas, Benjamin Rambo-Martin, Shatavia Morrison, Christopher Gulvick, Scott Sammons, Yvette Unoarumhi, Darlene Wagner, Matthew Schmerer, Minoo Agarwal, Eyad Almasri, Debbie Boles, Ayla Burns, Nuthawin Charoensri, Oren Cohen, Susan Countryman, Mary Ann Cristobal, Bobbi Croy, Suzanne Dale, Hrushikesh Deshmukh, Amanda Douglas, Vincent Drouillon, Marcia Eisenberg, Howard Engler, Rama Ghatti, Prashant Gupta, Susan Hicks, Jake Humphrey, Lax Iyer, Manoj Jain, Mohan Kolli, Brian Krueger, Tim Kuphal, Stanley Letovsky, Michael Levandoski, Craig Lukasik, Jonathan Meltzer, Brian Norvell, Mindy Nye, Scott Parker, Christos Petropoulos, John Pruitt, Steven Ragan, Scott Ryan, Mike Sapeta, Jana Schroth, Suresh Babu Selvaraju, Goran Stevovic, Amanda Suchanek, Andrea Throop, Lyndon Tilson, Thomas Urban, Joe Voshell, Kimberly Wagner, Jonathan Williams, Mary Williamson, Qian Zeng, Tricia Zwiefelhofer, Clinton R. Paden, Duncan MacCannell |
| EPI_ISL_1695436, EPI_ISL_1695443, EPI_ISL_1695444 | LABORATOIRE DE BIOLOGIE MEDICALE | CNR Virus des Infections Respiratoires - France SUD | Antonin Bal, Gregory Destras, Gwendolyne Burfin, Hadrien Regue, Quentin Semanas, Martine Valette, Bruno Lina, Laurence Josset |
| EPI_ISL_1701870, EPI_ISL_1701871, EPI_ISL_1701875 | Laboratory Corporation of America | Centers for Disease Control and Prevention Division of Viral Diseases, Pathogen Discovery | Dakota Howard, Dhwani Batra, Peter W. Cook, Kara Moser, Adrian Paskey, Jason Caravas, Benjamin Rambo-Martin, Shatavia Morrison, Christopher Gulvick, Scott Sammons, Yvette Unoarumhi, Darlene Wagner, Matthew Schmerer, Minoo Agarwal, Eyad Almasri, Debbie Boles, Ayla Burns, Nuthawin Charoensri, Oren Cohen, Susan Countryman, Mary Ann Cristobal, Bobbi Croy, Suzanne Dale, Hrushikesh Deshmukh, Amanda Douglas, Vincent Drouillon, Marcia Eisenberg, Howard Engler, Rama Ghatti, Prashant Gupta, Susan Hicks, Jake Humphrey, Lax Iyer, Manoj Jain, Mohan Kolli, Brian Krueger, Tim Kuphal, Stanley Letovsky, Michael Levandoski, Craig Lukasik, Jonathan Meltzer, Brian Norvell, Mindy Nye, Scott Parker, Christos Petropoulos, John Pruitt, Steven Ragan, Scott Ryan, Mike Sapeta, Jana Schroth, Suresh Babu Selvaraju, Goran Stevovic, Amanda Suchanek, Andrea Throop, Lyndon Tilson, Thomas Urban, Joe Voshell, Kimberly Wagner, Jonathan Williams, Mary Williamson, Qian Zeng, Tricia Zwiefelhofer, Clinton R. Paden, Duncan MacCannell |
| EPI_ISL_1704843, EPI_ISL_1704844 | National Public Health Laboratory, National Centre for Infectious Diseases | National Public Health Laboratory, National Centre for Infectious Diseases | Tze Minn Mak, Zhenyang Zhou, Grace Jie Yin Ngan, Royce Ang, Lin Cui, Raymond Tzer Pin Lin |
| EPI_ISL_1710705 | School of Pharmacy, Shenandoah University | School of Pharmacy, Shenandoah University | Adams,S.M., Harralson,A.F., Kidd,R.S., Sawyer,G.W. |
| EPI_ISL_1715048 | Istituto Zooprofilattico Sperimentale del Mezzogiorno | TIGEM | Antonio Grimaldi Patrizia Annunziata Francesco Panariello Biancamaria Pierri Claudia Tiberio Teresa Giuliano Valentina Bouche Chiara Colantuono Maria Concetta Cuomo Denise Di Concilio Lucio Di Filippo Anna Manfredi Marcello Salvi Antonio Limone Luigi Atripaldi Pellegrino Cerino Andrea Ballabio Davide Cacchiarelli |
| EPI_ISL_1718858 | Lighthouse Lab in Milton Keynes | Wellcome Sanger Institute for the COVID-19 Genomics UK (COG-UK) Consortium | The Lighthouse Lab in Milton Keynes and Alex Alderton, Roberto Amato, Jeffrey Barrett, Sonia Goncalves, Ewan Harrison, David K. Jackson, Ian Johnston, Dominic Kwiatkowski, Cordelia Langford, John Sillitoe on behalf of the Wellcome Sanger Institute COVID-19 Surveillance Team |
| EPI_ISL_1719870 | National Public Health Laboratory, National Centre for Infectious Diseases | National Public Health Laboratory, National Centre for Infectious Diseases | Tze Minn Mak, Zhenyang Zhou, Grace Jie Yin Ngan, Royce Ang, Lin Cui, Raymond Tzer Pin Lin |
| EPI_ISL_1720971 | MVZ Dr. Eberhard & Partner Dortmund | Robert Koch Institute | unknown |
| EPI_ISL_1722672 | MVZ Labor Dr. Limbach & Kollegen GbR | Robert Koch Institute | unknown |
| EPI_ISL_1724308, EPI_ISL_1724310 | Labor Prof. Dr. G. Enders MVZ GbR | Robert Koch Institute | unknown |
| EPI_ISL_1724422, EPI_ISL_1724436 | MVZ fÃ¼r Laboratoriumsmedizin und Mikrobiologie Koblenz-Mittelrhein (Labor Koblenz) | Robert Koch Institute | unknown |
| EPI_ISL_1725248 | MDI Limbach Berlin GmbH; MVZ Labor Berlin | Robert Koch Institute | unknown |
| EPI_ISL_1725261 | MVZ fÃ¼r Laboratoriumsmedizin und Mikrobiologie Koblenz-Mittelrhein (Labor Koblenz) | Robert Koch Institute | unknown |
| EPI_ISL_1725609, EPI_ISL_1725614, EPI_ISL_1725616, EPI_ISL_1725617, EPI_ISL_1725619, EPI_ISL_1725623, EPI_ISL_1725629 | MDI Limbach Berlin GmbH; MVZ Labor Berlin | Robert Koch Institute | unknown |
| EPI_ISL_1725636, EPI_ISL_1725645 | MVZ fÃ¼r Laboratoriumsmedizin und Mikrobiologie Koblenz-Mittelrhein (Labor Koblenz) | Robert Koch Institute | unknown |

| | | | |
|---|---|---|---|
| EPI_ISL_1726900 | MVZ Labor Krone GbR | Robert Koch Institute | unknown |
| EPI_ISL_1727004, EPI_ISL_1727028, EPI_ISL_1727049 | Medizinische Laboratorien DÃ¼sseldorf | Robert Koch Institute | unknown |
| EPI_ISL_1727121 | Labor Dr. Heidrich & Kollegen MVZ GmbH Hamburg | Robert Koch Institute | unknown |
| EPI_ISL_1731352 | National Virus Reference Laboratory | National Virus Reference Laboratory | Zoe Yandle, Charlene Bennett, Gabriel Gonzalez, Michael Carr, Jonathan Dean, Cillian F De Gascun |
| EPI_ISL_1737184 | Aegis Sciences Corporation | Centers for Disease Control and Prevention Division of Viral Diseases, Pathogen Discovery | Dakota Howard, Dhwani Batra, Peter W. Cook, Kara Moser, Adrian Paskey, Jason Caravas, Benjamin Rambo-Martin, Shatavia Morrison, Christopher Gulvick, Scott Sammons, Yvette Unoarumhi, Darlene Wagner, Matthew Schmerer, Cyndi Clark, Patrick Campbell, Rob Case, Vikramsinha Ghorpade, Holly Houdeshell, Ola Kvalvaag, Dillon Nall, Ethan Sanders, Alec Vest, Shaun Westlund, Matthew Hardison, Clinton R. Paden, Duncan MacCannell |
| EPI_ISL_1742737 | Berkeley Medical Center | WVU and Marshall University Combined Genomics Core Facilities | James Denvir, Peter Stoilov, Peter Perrotta, Wesley Kimble, Ryan Percifield |
| EPI_ISL_1745239, EPI_ISL_1745277, EPI_ISL_1745284, EPI_ISL_1745291, EPI_ISL_1745299, EPI_ISL_1745303, EPI_ISL_1745306, EPI_ISL_1745324, EPI_ISL_1745328 | Azienda Sanitaria dell'Alto Adige Laboratorio Aziendale di Microbiologia e Virologia | Istituto di Genomica Applicata | Elisabetta Pagani, Irene Bianconi, Elisabetta Giacobazzi, Elisa Masi, Stefanie Wieser, Irena Jurman, Vera Vendramin, Gabriele Magris, Eleonora Paparelli, Davide Scaglione, Michele Morgante |
| EPI_ISL_1747406, EPI_ISL_1747407, EPI_ISL_1747420, EPI_ISL_1747436 | Washington State Department of Health Public Health Laboratories | Washington State Department of Health Public Health Laboratories | Drew MacKellar, Philip Dykema, Denny Russell, Joenice Gonzalez, Hannah Gray, Geoff Melly, Vanessa De Los Santos, Darren Lucas, JohnAric Peterson, Avi Singh, Rebecca Cao |
| EPI_ISL_1755433 | Biogroup Bio Lam-LCD Saint-Denis | Department of Virology, Henri Mondor University Hospital, Assistance Publique Hôpitaux de Paris, Université Paris-Est Créteil, INSERM U955 | Christophe Rodriguez, Slim Fourati, Vanessa Demontant, Guillaume Gricourt, Melissa N'Debi, Alexandre Soulier, Elisabeth Trawinski, Jean-Michel Pawlotsky |
| EPI_ISL_1755791 | Hôpital Avicenne | Department of Virology, Henri Mondor University Hospital, Assistance Publique Hôpitaux de Paris, Université Paris-Est Créteil, INSERM U955 | Christophe Rodriguez, Slim Fourati, Vanessa Demontant, Guillaume Gricourt, Melissa N'Debi, Alexandre Soulier, Elisabeth Trawinski, Jean-Michel Pawlotsky |
| EPI_ISL_1756563, EPI_ISL_1756612, EPI_ISL_1756631, EPI_ISL_1756637 | Biogroup Bio Lam-LCD Saint-Denis | Department of Virology, Henri Mondor University Hospital, Assistance Publique Hôpitaux de Paris, Université Paris-Est Créteil, INSERM U955 | Christophe Rodriguez, Slim Fourati, Vanessa Demontant, Guillaume Gricourt, Melissa N'Debi, Alexandre Soulier, Elisabeth Trawinski, Jean-Michel Pawlotsky |
| EPI_ISL_1759578, EPI_ISL_1759697 | Istituto Zooprofilattico Sperimentale del Mezzogiorno | TIGEM | Antonio Grimaldi Patrizia Annunziata Francesco Panariello Biancamaria Pierri Claudia Tiberio Teresa Giuliano Valentina Bouche Chiara Colantuono Maria Concetta Cuomo Denise Di Concilio Lucio Di Filippo Anna Manfredi Marcello Salvi Antonio Limone Luigi Atripaldi Pellegrino Cerino Andrea Ballabio Davide Cacchiarelli |
| EPI_ISL_1760146 | Ministry of Health Turkey | Ministry of Health Turkey | Fatma Bayrakdar, Yasemin Cosgun, Suleyman Yalcin, Gulay Korukluoglu |
| EPI_ISL_1760379 | Microbiologia e Virologia | Microbiologia e Virologia | L.Atripaldi, C. Tiberio, A.Perfetti |
| EPI_ISL_1761554 | P.O.CARDARELLI | P.O.CARDARELLI | Scutellà M, Felice V, Niro G |
| EPI_ISL_1785083 | SIESP CHIETI - DRIVE IN CHIETI | Istituto Zooprofilattico Sperimentale dell'Abruzzo e Molise "G. Caporale" | Lorusso A, Marcacci M, Di Domenico M, Ancora M, Curini V, Di Lollo Valeria, Mangone I, Rinaldi A, Delli Compagni E, Scialabba S, Caporale M, Di Pasquale A, Cammà C, Puglia I, Calistri P, Savini G |
| EPI_ISL_1785213, EPI_ISL_1785227 | Università degli Studi di Perugia | Istituto Zooprofilattico Sperimentale dell'Abruzzo e Molise "G. Caporale" | Mencacci A, Camilloni B,Lorusso A, Marcacci M, Di Domenico M, Ancora M, Curini V, Di Lollo Valeria, Mangone I, Rinaldi A, Delli Compagni E, Scialabba S, Caporale M, Di Pasquale A, Cammà C, Puglia I, Calistri P, Savini G |
| EPI_ISL_1785311 | National Virus Reference Laboratory | National Virus Reference Laboratory | Zoe Yandle, Charlene Bennett, Gabriel Gonzalez, Michael Carr, Jonathan Dean, Cillian F De Gascun |
| EPI_ISL_1785840, EPI_ISL_1785841 | UW Virology Lab | UW Virology Lab | Pavitra Roychoudhury, Hong Xie, Lasata Shrestha, Tien V. Nguyen, Shah Mohamed Bakhash, Michelle Lin, Noah R. Baker, Sean Ellis, Meei-Li Huang, Keith R Jerome, Alexander Greninger |
| EPI_ISL_1788425 | HOPITAL SAINT ANDRE | CNR Virus des Infections Respiratoires - France SUD | Antonin Bal, Gregory Destras, Gwendolyne Burfin, Hadrien Regue, Quentin Semanas, Martine Valette, Bruno Lina, Laurence Josset |
| EPI_ISL_1788551, EPI_ISL_1788552 | LBM CLUSES | CNR Virus des Infections Respiratoires - France SUD | Antonin Bal, Gregory Destras, Gwendolyne Burfin, Hadrien Regue, Quentin Semanas, Martine Valette, Bruno Lina, Laurence Josset |
| EPI_ISL_1789246, EPI_ISL_1789456 | UW Virology Lab | UW Virology Lab | Pavitra Roychoudhury, Hong Xie, Lasata Shrestha, Tien V. Nguyen, Shah Mohamed Bakhash, Michelle Lin, Noah R. Baker, Sean Ellis, Meei-Li Huang, Keith R Jerome, Alexander Greninger |
| EPI_ISL_1789618 | M Health Fairview | Minnesota Department of Health, Public Health Laboratory | Alexandra Lorentz, Jacob Garfin, Matt Plumb, and Xiong Wang |
| EPI_ISL_1792899 | Dutch COVID-19 response team | National Institute for Public Health and the Environment (RIVM) | Adam Meijer, Harry Vennema, Dirk Eggink, Jeroen Cremer, Sharon van den Brink, Bas van der Veer, AnneMarie van den Brandt, Lisa Wijsman, Kim Freriks, Ryanne Jaarsma, Euníce Then, Lynn Aarts, Sanne Bos, Melissa van Tuil, Robert Kohl, Linda van de Nes, Sjoerd Kuiling, James Groot, Florian Zwagemaker, Dennis Schmitz, Annelies Kroneman, Karim Hajji, Chantal Reusken, on behalf of the national COVID-19 response team |
| EPI_ISL_1793318, EPI_ISL_1793319 | Istituto Zooprofilattico Sperimentale del Mezzogiorno | TIGEM | Antonio Grimaldi Patrizia Annunziata Francesco Panariello Biancamaria Pierri Claudia Tiberio Teresa Giuliano Valentina Bouche Chiara Colantuono Maria Concetta Cuomo Denise Di Concilio Lucio Di Filippo Anna Manfredi Marcello Salvi Antonio Limone Luigi Atripaldi Pellegrino Cerino Andrea Ballabio Davide Cacchiarelli |
| EPI_ISL_1798716 | RI State Health Laboratories | Centers for Disease Control and Prevention Division of Viral Diseases, Pathogen Discovery | Mili Sheth, Sarah Nobles, Jasmine Padilla, Mark Burroughs, Shoshona Le, Katie Dillon, Peter Cook, Clinton R. Paden, Dhwani Batra, Krista Queen, Kristen Knipe, Dakota Howard, Yvette Unoarumhi, Darlene Wagner, Matthew Schmerer, Ben L. Rambo-Martin, Kristine Lacek, Sam Shepard, Alison Laufer Halpin, Dave Wentworth, Vivien Dugan, Suxiang Tong, Justin Lee |
| EPI_ISL_1804911, EPI_ISL_1804916, EPI_ISL_1804923 | Laboratorio Aziendale di Microbiologia e Virologia, Azienda Sanitaria dell'Alto Adige | Laboratorio Aziendale di Microbiologia e Virologia, Azienda Sanitaria dell'Alto Adige | Irene Bianconi, Bartolomeo Mobilio Rodriguez, Elisabetta Giacobazzi, Elisa Masi, Stefanie Wieser, Anne Picard, Claudia Volpato, Chiara Cantaloni, Elisabetta Pagani |
| EPI_ISL_1804970 | CQRC_QUALITY CONTROL CHEMICAL BIOLOGICAL RISK_AOOR Villa Sofia Cervello Palermo | CQRC_QUALITY CONTROL CHEMICAL BIOLOGICAL RISK_AOOR Villa Sofia Cervello Palermo | Di Gaudio, F., Contino, F., Brunacci, G., |
| EPI_ISL_1808671 | Swedish national genomic surveillance program of SARS-CoV-2 | The Public Health Agency of Sweden | Maximilian Riess, Maria Lind Karlberg, Alma Brolund, Swedish national genomic surveillance program of SARS-CoV-2 |
| EPI_ISL_1817303 | National Laboratory for Health, Environment and Food, OMM, Kranj | NLZOH (National Laboratory for Health, Environment and Food) / CISLD (Clinical Institute of Special Laboratory Diagnostics), University Children's Hospital, University Medical Center Ljubljana | Sandra Janezic, Aleksander Mahnic, Maja Rupnik, Tjasa Žohar retnik, Alenka Štorman, Nika Gobec, Aleksander Kocuvan, Kaja Tominc, Maša Jari, Mateja Ravnik, Monika Korošec, Marjana Petrevi / Jernej Kova, Barbara Jenko Bizjan, Tine Tesovnik, Robert Šket, Katarina Kozmos, Ana Grom, Maruša Debeljak, Marko Pokorn, Tadej Battelino |
| EPI_ISL_1818074, EPI_ISL_1818075, EPI_ISL_1818077, EPI_ISL_1818078 | Laboratory Corporation of America | Centers for Disease Control and Prevention Division of Viral Diseases, Pathogen Discovery | Dakota Howard, Dhwani Batra, Peter W. Cook, Kara Moser, Adrian Paskey, Jason Caravas, Benjamin Rambo-Martin, Shatavia Morrison, Christopher Gulvick, Scott Sammons, Yvette Unoarumhi, Darlene Wagner, Matthew Schmerer, Minoo Agarwal, Eyad Almasri, Debbie Boles, Ayla Burns, Nuthawin Charoensri, Oren Cohen, Susan Countryman, Mary Ann Cristobal, Bobbi Croy, Suzanne Dale, Hrushikesh Deshmukh, Amanda Douglas, Vincent Drouillon, Marcia Eisenberg, Howard Engler, Rama Ghatti, Prashant Gupta, Susan Hicks, Jake Humphrey, Lax Iyer, Manoj Jain, Mohan Kolli, Brian Krueger, Tim Kuphal, Stanley Letovsky, Michael Levandoski, Craig Lukasik, Jonathan Meltzer, Brian Norvell, Mindy Nye, Scott Parker, Christos Petropoulos, John Pruitt, Steven Ragan, Scott Ryan, Mike Sapeta, Jana Schroth, Suresh Babu Selvaraju, Goran Stevovic, Amanda Suchanek, Andrea Throop, Lyndon Tilson, Thomas Urban, Joe Voshell, Kimberly Wagner, Jonathan Williams, Mary Williamson, Qian Zeng, Tricia Zwiefelhofer, Clinton R. Paden, Duncan MacCannell |
| EPI_ISL_1820311, EPI_ISL_1820312, | Furst Medical Laboratory | Norwegian Institute of Public Health, Department of Virology | Kathrine Stene-Johansen, Kamilla Heddeland Instefjord, Hilde Elshaug, Garcia Llorente Ignacio, Jon Bråte, Engebretsen Serina Beate,Pedersen Benedikte |

| | | | |
|---|---|---|---|
| EPI_ISL_1820313 | | | Nevjen, Debech Nadia, Atiya R Ali,Marie Paulsen Madsen, Rasmus Riis Kopperud, Hilde Vollan, Karoline Bragstad, Olav Hungnes |
| EPI_ISL_1820314 | Medical Microbiology Unit, Department for Laboratory Medicine, Drammen Hospital, Vestre Viken Health Trust, | Norwegian Institute of Public Health, Department of Virology | Kathrine Stene-Johansen, Kamilla Heddeland Instefjord, Hilde Elshaug, Garcia Llorente Ignacio, Jon Bråte, Engebretsen Serina Beate,Pedersen Benedikte Nevjen, Debech Nadia, Atiya R Ali,Marie Paulsen Madsen, Rasmus Riis Kopperud, Hilde Vollan, Karoline Bragstad, Olav Hungnes |
| EPI_ISL_1820315 | Furst Medical Laboratory | Norwegian Institute of Public Health, Department of Virology | Kathrine Stene-Johansen, Kamilla Heddeland Instefjord, Hilde Elshaug, Garcia Llorente Ignacio, Jon Bråte, Engebretsen Serina Beate,Pedersen Benedikte Nevjen, Debech Nadia, Atiya R Ali,Marie Paulsen Madsen, Rasmus Riis Kopperud, Hilde Vollan, Karoline Bragstad, Olav Hungnes |
| EPI_ISL_1820779 | Hospital of Southern Norway - Kristiansand, Department of Medical Microbiology | Norwegian Institute of Public Health, Department of Virology | Kathrine Stene-Johansen, Kamilla Heddeland Instefjord, Hilde Elshaug, Garcia Llorente Ignacio, Jon Bråte, Engebretsen Serina Beate,Pedersen Benedikte Nevjen, Debech Nadia, Atiya R Ali,Marie Paulsen Madsen, Rasmus Riis Kopperud, Hilde Vollan, Karoline Bragstad, Olav Hungnes |
| EPI_ISL_1822672 | Ministry of Health Turkey | Ministry of Health Turkey | Fatma Bayrakdar, Yasemin Cosgun, Suleyman Yalcin, Gulay Korukluoglu |
| EPI_ISL_1824146 | Ospedale S.M. Goretti di Latina - ASL Latina presidio ospedaliero Nord | INMI Lazzaro Spallanzani IRCCS | B Bartolini, O Butera, F Messina, F De Nicola, A Massacci, A Lucci, L Di Biase, E Giombini |
| EPI_ISL_1826134, EPI_ISL_1826136 | UW Virology Lab | UW Virology Lab | Pavitra Roychoudhury, Hong Xie, Lasata Shrestha, Tien V. Nguyen, Shah Mohamed Bakhash, Michelle Lin, Noah R. Baker, Sean Ellis, Meei-Li Huang, Keith R Jerome, Alexander Greninger |
| EPI_ISL_1830347 | Lighthouse Lab in Alderley Park | Wellcome Sanger Institute for the COVID-19 Genomics UK (COG-UK) Consortium | Jacquelyn Wynn, Mairead Hyland, The Lighthouse Lab in Alderley Park and Alex Alderton, Roberto Amato, Jeffrey Barrett, Sonia Goncalves, Ewan Harrison, David K. Jackson, Ian Johnston, Dominic Kwiatkowski, Cordelia Langford, John Sillitoe on behalf of the Wellcome Sanger Institute COVID-19 Surveillance Team |
| EPI_ISL_1831596, EPI_ISL_1831620, EPI_ISL_1831643 | Laboratori Clínic Territorial de Girona | Can Ruti SARS-CoV-2 Sequencing Hub (HUGTiP/IrsiCaixa/IGTP) | Marc Noguera-Julian, Pilar Armengol, Ignacio Blanco, Antoni E Bordoy, Francesc Catala-Moll, Pere-Joan Cardona, Maria Casadellà, Cristina Casañ, Gemma Clara, Bonaventura Clotet, Cristina Esteban, Montserrat Giménez, Mercedes Guerrero, Anna Not, Roger Paredes, Mariona Parera, Verónica Saludes, Alba Sánchez, and Elisa Martró on behalf of the Can Ruti SARS-CoV-2 Sequencing Hub. |
| EPI_ISL_1831681 | Althaia. Xarxa Assistencial Universitària de Manresa | IrsiCaixa | Marc Noguera-Julian, Mariona Parera, Maria Casadellà, Pilar Armengol, Francesc Catala-Moll, Roger Paredes, Bonaventura Clotet Gloria Trujillo, Rafel Perez Vidal, Jaume Trape Pujol, Carolina Gonzalez Fernandez, Miquel Micó, Roger Paredes, Eulalia Grau, Bonaventura Clotet |
| EPI_ISL_1838837, EPI_ISL_1838838, EPI_ISL_1838848, EPI_ISL_1838857, EPI_ISL_1838858, EPI_ISL_1838859 | CSIR-Centre for Cellular and Molecular Biology | CSIR-Centre for Cellular and Molecular Biology-INSACOG | Payel Mukherjee,Lamuk Zaveri,Tulasi Nagabandi, Ara Sreenivas,Shreekant Verma, Amareshwar Vodapalli ,Blessy B John,Viswagithe S L,B Himasri,Valli Nagalakshmi Undamatla,Onkar Kulkarni,Sofia Banu,Archana Bharadwaj Siva,Sharath Chandra Thota,Karthik Bharadwaj Tallapaka,Rakesh K Mishra,Divya Tej Sowpati |
| EPI_ISL_1840705, EPI_ISL_1840706, EPI_ISL_1840707, EPI_ISL_1840725, EPI_ISL_1840726, EPI_ISL_1840774 | Azienda Sanitaria dell'Alto Adige Laboratorio Aziendale di Microbiologia e Virologia | Istituto di Genomica Applicata | Elisabetta Pagani, Irene Bianconi, Elisabetta Giacobazzi, Elisa Masi, Stefanie Wieser, Irena Jurman, Vera Vendramin, Gabriele Magris, Eleonora Paparelli, Davide Scaglione, Michele Morgante |
| EPI_ISL_1843336, EPI_ISL_1843342 | Labor Prof. Dr. G. Enders MVZ GbR | Robert Koch Institute | unknown |
| EPI_ISL_1843353 | MDI Limbach Berlin GmbH; MVZ Labor Berlin | Robert Koch Institute | unknown |
| EPI_ISL_1843356, EPI_ISL_1843359 | Labor Prof. Dr. G. Enders MVZ GbR | Robert Koch Institute | unknown |
| EPI_ISL_1847179 | MDI Limbach Berlin GmbH; MVZ Labor Berlin | Robert Koch Institute | unknown |
| EPI_ISL_1847182 | MVZ fÃ¼r Laboratoriumsmedizin und Mikrobiologie Koblenz-Mittelrhein (Labor Koblenz) | Robert Koch Institute | unknown |
| EPI_ISL_1847194, EPI_ISL_1847204 | MDI Limbach Berlin GmbH; MVZ Labor Berlin | Robert Koch Institute | unknown |
| EPI_ISL_1847217 | MVZ fÃ¼r Laboratoriumsmedizin und Mikrobiologie Koblenz-Mittelrhein (Labor Koblenz) | Robert Koch Institute | unknown |
| EPI_ISL_1847221, EPI_ISL_1847318, EPI_ISL_1847330 | MDI Limbach Berlin GmbH; MVZ Labor Berlin | Robert Koch Institute | unknown |
| EPI_ISL_1847724 | MVZ fÃ¼r Laboratoriumsmedizin und Mikrobiologie Koblenz-Mittelrhein (Labor Koblenz) | Robert Koch Institute | unknown |
| EPI_ISL_1847725, EPI_ISL_1847729, EPI_ISL_1847734 | MDI Limbach Berlin GmbH; MVZ Labor Berlin | Robert Koch Institute | unknown |
| EPI_ISL_1848754 | Labor MÃ¶nchengladbach MVZ Dr. Stein + Kollegen GbR | Robert Koch Institute | unknown |
| EPI_ISL_1848783, EPI_ISL_1848801, EPI_ISL_1848803 | MDI Limbach Berlin GmbH; MVZ Labor Berlin | Robert Koch Institute | unknown |
| EPI_ISL_1849374 | UW Virology Lab | UW Virology Lab | Pavitra Roychoudhury, Hong Xie, Lasata Shrestha, Tien V. Nguyen, Shah Mohamed Bakhash, Michelle Lin, Noah R. Baker, Sean Ellis, Meei-Li Huang, Keith R Jerome, Alexander Greninger |
| EPI_ISL_1849448 | MVZ Labor Krone GbR | Robert Koch Institute | unknown |
| EPI_ISL_1850314, EPI_ISL_1850325 | MDI Limbach Berlin GmbH; MVZ Labor Berlin | Robert Koch Institute | unknown |
| EPI_ISL_1850360 | MVZ Dr. Eberhard & Partner Dortmund | Robert Koch Institute | unknown |
| EPI_ISL_1854254 | Instituto Nacional de Saude (INSA) and Institute of Biomedicine (iBiMed), Universidade de Aveiro | Instituto Nacional de Saude (INSA) and Institute of Biomedicine (iBiMed), Universidade de Aveiro | Borges et al |
| EPI_ISL_1854858 | Clinical Molecular Microbiology Laboratory, UNC Hospitals | Jeremy Wang | Jeremy Wang, Alexander Rubinsteyn, Colleen Rice, Jason Smedberg, Shawn Hawken, Melissa Miller, Corbin Jones, Robert Hagan |
| EPI_ISL_1859259, EPI_ISL_1860240, EPI_ISL_1860959, EPI_ISL_1861130, EPI_ISL_1861219, EPI_ISL_1861375, EPI_ISL_1862921, EPI_ISL_1864252, EPI_ISL_1864267, EPI_ISL_1864300, EPI_ISL_1864685, EPI_ISL_1864956, EPI_ISL_1865121, EPI_ISL_1865528, EPI_ISL_1865859, EPI_ISL_1866443, EPI_ISL_1866463, EPI_ISL_1866643, EPI_ISL_1866657, EPI_ISL_1867049, EPI_ISL_1867586, EPI_ISL_1867794, EPI_ISL_1867948, EPI_ISL_1867985, EPI_ISL_1868453, EPI_ISL_1868805, EPI_ISL_1869041 | | | |
| see above | Department of Virus and Microbiological Special Diagnostics, Statens Serum Institut, Copenhagen, Denmark | Aalborg University | Danish Covid-19 Genome Consortium |
| EPI_ISL_1869115 | Department of Clinical Microbiology, Odense University Hospital, Odense, Denmark | Aalborg University | Danish Covid-19 Genome Consortium |
| EPI_ISL_1869166, EPI_ISL_1869173, EPI_ISL_1870601, EPI_ISL_1871094, EPI_ISL_1872516, EPI_ISL_1872655, EPI_ISL_1872832, EPI_ISL_1873667, EPI_ISL_1874383, EPI_ISL_1874957, EPI_ISL_1875518, EPI_ISL_1876635, EPI_ISL_1877251, EPI_ISL_1877698, EPI_ISL_1877699, EPI_ISL_1878067, EPI_ISL_1878539 | | | |
| see above | Department of Virus and Microbiological Special Diagnostics, Statens Serum Institut, Copenhagen, Denmark | Aalborg University | Danish Covid-19 Genome Consortium |
| EPI_ISL_1878864 | Department of Clinical Microbiology, Odense University Hospital, Odense, Denmark | Aalborg University | Danish Covid-19 Genome Consortium |
| EPI_ISL_1880159, EPI_ISL_1880486, EPI_ISL_1881473, EPI_ISL_1882091, EPI_ISL_1882513, EPI_ISL_1882669 | Department of Virus and Microbiological Special Diagnostics, Statens Serum Institut, Copenhagen, Denmark | Aalborg University | Danish Covid-19 Genome Consortium |
| EPI_ISL_1883032 | Department of Clinical Microbiology, Odense University Hospital, Odense, Denmark | Aalborg University | Danish Covid-19 Genome Consortium |
| EPI_ISL_1883194, EPI_ISL_1883530, | Department of Virus and Microbiological Special Diagnostics, | Aalborg University | Danish Covid-19 Genome Consortium |

| | | | |
|---|---|---|---|
| EPI_ISL_1883824, EPI_ISL_1884732, EPI_ISL_1885395, EPI_ISL_1885672, EPI_ISL_1885838, EPI_ISL_1885890 | Statens Serum Institut, Copenhagen, Denmark | | |
| EPI_ISL_1886140, EPI_ISL_1886168, EPI_ISL_1886196 | Labor Berlin Charite Vivantes GmbH / Institut fur Virologie | Charite Universitatsmedizin Berlin, Institut fur Virologie/Labor Berlin | Peter Menzel, Christine Stephan, Rolf Schwarzer, Victor M Corman, Barbara Muhlemann, Terry Jones, Christian Drosten |
| EPI_ISL_1886602, EPI_ISL_1887377, EPI_ISL_1887695, EPI_ISL_1887862, EPI_ISL_1888564, EPI_ISL_1888933, EPI_ISL_1888934, EPI_ISL_1888978, EPI_ISL_1889098, EPI_ISL_1889264, EPI_ISL_1889401, EPI_ISL_1889907, EPI_ISL_1889991, EPI_ISL_1890001, EPI_ISL_1890828 | | | |
| see above | Department of Virus and Microbiological Special Diagnostics, Statens Serum Institut, Copenhagen, Denmark | Aalborg University | Danish Covid-19 Genome Consortium |
| EPI_ISL_1891211, EPI_ISL_1891230, EPI_ISL_1891238, EPI_ISL_1891257, EPI_ISL_1891271, EPI_ISL_1891284 | National Virus Reference Laboratory | National Virus Reference Laboratory | Zoe Yandle, Charlene Bennett, Gabriel Gonzalez, Michael Carr, Jonathan Dean, Cillian F De Gascun |
| EPI_ISL_1891710, EPI_ISL_1892157, EPI_ISL_1892287, EPI_ISL_1892288, EPI_ISL_1892426, EPI_ISL_1892499 | Department of Virus and Microbiological Special Diagnostics, Statens Serum Institut, Copenhagen, Denmark | Aalborg University | Danish Covid-19 Genome Consortium |
| EPI_ISL_1892701 | Department of Clinical Microbiology and Center for Genomic Medicine, Rigshospitalet, Copenhagen, Denmark | Aalborg University | Danish Covid-19 Genome Consortium |
| EPI_ISL_1892959 | Laboratori Clínic Territorial de Girona | Can Ruti SARS-CoV-2 Sequencing Hub (HUGTiP/IrsiCaixa/IGTP) | Marc Noguera-Julian, Pilar Armengol, Ignacio Blanco, Antoni E Bordoy, Francesc Catala-Moll, Pere-Joan Cardona, Maria Casadellà, Cristina Casañ, Gemma Clara, Bonaventura Clotet, Cristina Esteban, Montserrat Giménez, Mercedes Guerrero, Anna Not, Roger Paredes, Mariona Parera, Verónica Saludes, Alba Sánchez, and Elisa Martró on behalf of the Can Ruti SARS-CoV-2 Sequencing Hub. |
| EPI_ISL_1892978, EPI_ISL_1893842, EPI_ISL_1894042, EPI_ISL_1894348, EPI_ISL_1894828, EPI_ISL_1894962, EPI_ISL_1894963 | Department of Virus and Microbiological Special Diagnostics, Statens Serum Institut, Copenhagen, Denmark | Aalborg University | Danish Covid-19 Genome Consortium |
| EPI_ISL_1895041 | Department of Clinical Microbiology, Odense University Hospital, Odense, Denmark | Aalborg University | Danish Covid-19 Genome Consortium |
| EPI_ISL_1895477, EPI_ISL_1895630, EPI_ISL_1895770, EPI_ISL_1896003, EPI_ISL_1896059, EPI_ISL_1896533, EPI_ISL_1896651 | Department of Virus and Microbiological Special Diagnostics, Statens Serum Institut, Copenhagen, Denmark | Aalborg University | Danish Covid-19 Genome Consortium |
| EPI_ISL_1899146 | Swedish national genomic surveillance program of SARS-CoV-2 | The Public Health Agency of Sweden | Maximilian Riess, Maria Lind Karlberg, Alma Brolund, Swedish national genomic surveillance program of SARS-CoV-2 |
| EPI_ISL_1904330 | AZDelta | AZDelta | Geert Martens; Dieter De Smet |
| EPI_ISL_1905108 | Labor Berlin Charite Vivantes GmbH / Institut fur Virologie | Charite Universitatsmedizin Berlin, Institut fur Virologie/Labor Berlin | Peter Menzel, Christine Stephan, Rolf Schwarzer, Victor M Corman, Barbara Muhlemann, Terry Jones, Christian Drosten |
| EPI_ISL_1908870 | Hospital Universitari Arnau de Vilanova | Hospital Universitari Vall d'Hebron - Vall d'Hebron Institut de Recerca | Cristina Andrés, Maria Piñana, Damir Garcia-Cehic, Ariadna Rando, Juliana Esperalba, Maria Gema Codina, Carla Castillo, Maria Carmen Martin, Tomàs Pumarola, Josep Quer, Andrés Antón |
| EPI_ISL_1913252 | Laboratorio de Virología HUCA | Laboratorio de Virología HUCA | Sandoval M, Castelló C, Gómez de Oña J, Boga JA, Rojo S, Alvarez-Arguelles ME, Abreu F, Costales I, Perez-Martínez Z, Martín-Rodríguez G, Coto E, Melón S |
| EPI_ISL_1914210 | Viollier AG | Department of Biosystems Science and Engineering, ETH Zurich | Chaoran Chen, Sarah Nadeau, Catharine Aquino, Ivan Topolsky, Philipp Jablonski, Lara Fuhrmann, David Dreifuss, Katharina Jahn, Andreia Cabral de Gouvea, Maria Domenica Moccia, Simon Gruter, Timothy Sykes, Lennart Opitz, Griffin White, Laura Neff, Doris Popovic, Andrea Patrignani, Jay Tracy, Ralph Schlapbach, Christiane Beckmann, Maurice Redondo, Olivier Kobel, Christoph Noppen, Sophie Seidel, Noemie Santamaria de Souza, Niko Beerenwinkel, Tanja Stadler |
| EPI_ISL_1914285 | ULSS 3 Venezia | Istituto Zooprofilattico Sperimentale delle Venezie | Adelaide Milani, Alessia Schivo, Annalisa Salviato, Elisa Palumbo, Erika Giorgia Quaranta, Luca Tassoni, Ambra Pastori, Edoardo Giussani, Alice Fusaro, Isabella Monne, Calogero Terregino, Antonia Ricci |
| EPI_ISL_1914286, EPI_ISL_1914287 | ULSS 2 Marca Trevigiana | Istituto Zooprofilattico Sperimentale delle Venezie | Adelaide Milani, Alessia Schivo, Annalisa Salviato, Elisa Palumbo, Erika Giorgia Quaranta, Luca Tassoni, Ambra Pastori, Edoardo Giussani, Alice Fusaro, Isabella Monne, Calogero Terregino, Antonia Ricci |
| EPI_ISL_1914646, EPI_ISL_1914650 | National Public Health Laboratory, National Centre for Infectious Diseases | National Public Health Laboratory, National Centre for Infectious Diseases | Tze Minn Mak, Zhenyang Zhou, Grace Jie Yin Ngan, Royce Ang, Lin Cui, Raymond Tzer Pin Lin |
| EPI_ISL_1918245 | Laboratoire national de sante, Microbiology, Virology | Laboratoire national de sante, Microbiology, Microbial Genomics Platform | Anke Wienecke-Baldacchino, Catherine Ragimbeau,Jessica Tapp, Fatu Djabi, Lise Pignon, Raoul Salmon, Trung Nguyen Nguyen, Tamir Abdelrahman |
| EPI_ISL_1919379 | Furst Medical Laboratory | Norwegian Institute of Public Health, Department of Virology | Kathrine Stene-Johansen, Kamilla Heddeland Instefjord, Hilde Elshaug, Garcia Llorente Ignacio, Jon Bråte, Engebretsen Serina Beate,Pedersen Benedikte Nevjen, Debech Nadia, Atiya R Ali,Marie Paulsen Madsen, Rasmus Riis Kopperud, Hilde Vollan, Karoline Bragstad, Olav Hungnes |
| EPI_ISL_1921894 | OLVZ Aalst | OLVZ Aalst | Anne Vankeerberghen |
| EPI_ISL_1923538, EPI_ISL_1923583 | Laboratorio Aziendale di Microbiologia e Virologia, Azienda Sanitaria dell'Alto Adige | Laboratorio Aziendale di Microbiologia e Virologia, Azienda Sanitaria dell'Alto Adige | Irene Bianconi, Bartolomeo Mobilio Rodriguez, Elisabetta Giacobazzi, Elisa Masi, Stefanie Wieser, Anne Picard, Claudia Volpato, Chiara Cantaloni, Elisabetta Pagani |
| EPI_ISL_1927302, EPI_ISL_1927303 | SARS-CoV-2 testing team, National Institute of Infectious Diseases | Pathogen Genomics Center, National Institute of Infectious Diseases | Tsuyoshi Sekizuka, Kentaro Itokawa, Rina Tanaka, Masanori Hashino, Nozomu Hanaoka, Masumichi Saito, Naomi Nojiri, Hazuka Y Furihata, Sana Uchikoba, Tsuguto Fujimoto, Makoto Kuroda |
| EPI_ISL_1935392 | LABORATOIRE DEPARTEMENTAL D'ANALYSES | CNR Virus des Infections Respiratoires - France SUD | Antonin Bal, Gregory Destras, Gwendolyne Burfin, Hadrien Regue, Quentin Semanas, Martine Valette, Bruno Lina, Laurence Josset |
| EPI_ISL_1935393 | LIV VILLEFRANCHE | CNR Virus des Infections Respiratoires - France SUD | Antonin Bal, Gregory Destras, Gwendolyne Burfin, Hadrien Regue, Quentin Semanas, Martine Valette, Bruno Lina, Laurence Josset |
| EPI_ISL_1936189 | MD PHL | MD PHL | Maryland Department of Health Laboratories Administration |
| EPI_ISL_1941525, EPI_ISL_1941566 | Azienda Sanitaria dell'Alto Adige Laboratorio Aziendale di Microbiologia e Virologia | Istituto di Genomica Applicata | Elisabetta Pagani, Irene Bianconi, Elisabetta Giacobazzi, Elisa Masi, Stefanie Wieser, Irena Jurman, Vera Vendramin, Gabriele Magris, Eleonora Paparelli, Davide Scaglione, Michele Morgante |
| EPI_ISL_1942236, EPI_ISL_1942238, EPI_ISL_1942239, EPI_ISL_1942240 | Laboratory Corporation of America | Centers for Disease Control and Prevention Division of Viral Diseases, Pathogen Discovery | Dakota Howard, Dhwani Batra, Peter W. Cook, Kara Moser, Adrian Paskey, Jason Caravas, Benjamin Rambo-Martin, Shatavia Morrison, Christopher Gulvick, Scott Sammons, Yvette Unoarumhi, Darlene Wagner, Matthew Schmerer, Minoo Agarwal, Eyad Almasri, Debbie Boles, Ayla Burns, Nuthawin Charoensri, Oren Cohen, Susan Countryman, Mary Ann Cristobal, Bobbi Croy, Suzanne Dale, Hrushikesh Deshmukh, Amanda Douglas, Vincent Drouillon, Marcia Eisenberg, Howard Engler, Rama Ghatti, Prashant Gupta, Susan Hicks, Jake Humphrey, Lax Iyer, Manoj Jain, Mohan Kolli, Brian Krueger, Tim Kuphal, Stanley Letovsky, Michael Levandoski, Craig Lukasik, Jonathan Meltzer, Brian Norvell, Mindy Nye, Scott Parker, Christos Petropoulos, John Pruitt, Steven Ragan, Scott Ryan, Mike Sapeta, Jana Schroth, Suresh Babu Selvaraju, Goran Stevovic, Amanda Suchanek, Andrea Throop, Lyndon Tilson, Thomas Urban, Joe Voshell, Kimberly Wagner, Jonathan Williams, Mary Williamson, Qian Zeng, Tricia Zwiefelhofer, Clinton R. Paden, Duncan MacCannell |
| EPI_ISL_1962937, EPI_ISL_1962938, EPI_ISL_1962939 | Dutch COVID-19 response team | National Institute for Public Health and the Environment (RIVM) | Adam Meijer, Harry Vennema, Dirk Eggink, Jeroen Cremer, Sharon van den Brink, Bas van der Veer, AnneMarie van den Brandt, Lisa Wijsman, Kim Freriks, Ryanne Jaarsma, Euníce Then, Lynn Aarts, Sanne Bos, Melissa van Tuil, Linda van de Nes, Sjoerd Kuiling, James Groot, Florian Zwagemaker, Dennis Schmitz, Annelies Kroneman, Karim Hajji, Chantal Reusken, on behalf of the national COVID-19 response team |

| | | | |
|---|---|---|---|
| EPI_ISL_1963287, EPI_ISL_1963288, EPI_ISL_1963289, EPI_ISL_1963290 | University Hospitals of Geneva, Laboratory of Virology | HUG, Laboratory of Virology and the Health2030 Genome Center | Samuel Cordey, Ana Rita Goncalves, Laurent Kaiser, Lorenzo Cerutti, Henri Pegeot, Melyssa Elies, Deborah Penet, Keith Harshman, Ioannis Xenarios, Emmanouil Dermitzakis |
| EPI_ISL_1970399 | Centre for Dengue Research and AICBU, Department of Immunology and Molecular Medicine | Centre for Dengue Research and AICBU, Department of Immunology and Molecular Medicine | Chandima Jeewandara, Deshni Jayathilaka, Dinuka Ariyaratne, Deshan Madhusanka, Diyanath Ranasinghe, Laksiri Gomes, Gathsaurie Neelika Malavige |
| EPI_ISL_1990051 | Laboratory Corporation of America | Centers for Disease Control and Prevention Division of Viral Diseases, Pathogen Discovery | Dakota Howard, Dhwani Batra, Peter W. Cook, Kara Moser, Adrian Paskey, Jason Caravas, Benjamin Rambo-Martin, Shatavia Morrison, Christopher Gulvick, Scott Sammons, Yvette Unoarumhi, Darlene Wagner, Matthew Schmerer, Minoo Agarwal, Eyad Almasri, Debbie Boles, Ayla Burns, Nuthawin Charoensri, Oren Cohen, Susan Countryman, Mary Ann Cristobal, Bobbi Croy, Suzanne Dale, Hrushikesh Deshmukh, Amanda Douglas, Vincent Drouillon, Marcia Eisenberg, Howard Engler, Rama Ghatti, Prashant Gupta, Susan Hicks, Jake Humphrey, Lax Iyer, Manoj Jain, Mohan Kolli, Brian Krueger, Tim Kuphal, Stanley Letovsky, Michael Levandoski, Craig Lukasik, Jonathan Meltzer, Brian Norvell, Mindy Nye, Scott Parker, Christos Petropoulos, John Pruitt, Steven Ragan, Scott Ryan, Mike Sapeta, Jana Schroth, Suresh Babu Selvaraju, Goran Stevovic, Amanda Suchanek, Andrea Throop, Lyndon Tilson, Thomas Urban, Joe Voshell, Kimberly Wagner, Jonathan Williams, Mary Williamson, Qian Zeng, Tricia Zwiefelhofer, Clinton R. Paden, Duncan MacCannell |
| EPI_ISL_1999837, EPI_ISL_1999851 | Laboratori Clínic Territorial de Girona | Can Ruti SARS-CoV-2 Sequencing Hub (HUGTiP/IrsiCaixa/IGTP) | Marc Noguera-Julian, Pilar Armengol, Ignacio Blanco, Antoni E Bordoy, Francesc Catala-Moll, Pere-Joan Cardona, Maria Casadellà, Cristina Casañ, Gemma Clara, Bonaventura Clotet, Cristina Esteban, Montserrat Giménez, Mercedes Guerrero, Anna Not, Roger Paredes, Mariona Parera, Verónica Saludes, Alba Sánchez, and Elisa Martró on behalf of the Can Ruti SARS-CoV-2 Sequencing Hub. |
| EPI_ISL_2001059 | PHV-FSS | PHV-FSS | Son Nguyen |
| EPI_ISL_2001420 | SIESP DIPARTIMENTO DI PREVENZIONE TERAMO | Istituto Zooprofilattico Sperimentale dell'Abruzzo e Molise "G. Caporale" | Lorusso A, Marcacci M, Di Domenico M, Ancora M, Curini V, Di Lollo Valeria, Mangone I, Rinaldi A, Delli Compagni E, Scialabba S, Caporale M, Di Pasquale A, Cammà C, Puglia I, Calistri P, Savini G |
| EPI_ISL_2001431 | Università degli Studi di Perugia | Istituto Zooprofilattico Sperimentale dell'Abruzzo e Molise "G. Caporale" | Mencacci A, Camilloni B, Lorusso A, Marcacci M, Di Domenico M, Ancora M, Curini V, Mangone I, Rinaldi A, Scialabba S, Di Pasquale A, Cammà C, Puglia I, Calistri P, Savini G |
| EPI_ISL_2001451, EPI_ISL_2001452 | Laboratorio Analisi Osp. Città di Castello - Azienda USL Umbria1 | Istituto Zooprofilattico Sperimentale dell'Abruzzo e Molise "G. Caporale" | Malagigi V, Tacconi P, Lorusso A, Marcacci M, Di Domenico M, Ancora M, Curini V, Mangone I, Rinaldi A, Scialabba S, Di Pasquale A, Cammà C, Puglia I, Calistri P, Savini G |
| EPI_ISL_2001459, EPI_ISL_2001460 | Azienda Ospedaliera Terni | Istituto Zooprofilattico Sperimentale dell'Abruzzo e Molise "G. Caporale" | Palumbo M, Scaccetti A, Lorusso A, Marcacci M, Di Domenico M, Ancora M, Curini V, Mangone I, Rinaldi A, Scialabba S, Di Pasquale A, Cammà C, Puglia I, Calistri P, Savini G |
| EPI_ISL_2001524 | Hospital of Southern Norway - Kristiansand, Department of Medical Microbiology | Norwegian Institute of Public Health, Department of Virology | Kathrine Stene-Johansen, Kamilla Heddeland Instefjord, Hilde Elshaug, Garcia Llorente Ignacio, Jon Bråte, Engebretsen Serina Beate,Pedersen Benedikte Nevjen, Debech Nadia, Atiya R Ali,Marie Paulsen Madsen, Rasmus Riis Kopperud, Hilde Vollan, Karoline Bragstad, Olav Hungnes |
| EPI_ISL_2001527 | Furst Medical Laboratory | Norwegian Institute of Public Health, Department of Virology | Kathrine Stene-Johansen, Kamilla Heddeland Instefjord, Hilde Elshaug, Garcia Llorente Ignacio, Jon Bråte, Engebretsen Serina Beate,Pedersen Benedikte Nevjen, Debech Nadia, Atiya R Ali,Marie Paulsen Madsen, Rasmus Riis Kopperud, Hilde Vollan, Karoline Bragstad, Olav Hungnes |
| EPI_ISL_2002145, EPI_ISL_2002161, EPI_ISL_2002166, EPI_ISL_2002191, EPI_ISL_2002192, EPI_ISL_2002250, EPI_ISL_2002281, EPI_ISL_2002345, EPI_ISL_2002353, EPI_ISL_2002383, EPI_ISL_2002386, EPI_ISL_2002694, EPI_ISL_2002885, EPI_ISL_2002887, EPI_ISL_2002892, EPI_ISL_2002921 | | | |
| see above | UW Virology Lab | UW Virology Lab | Pavitra Roychoudhury, Hong Xie, Lasata Shrestha, Tien V. Nguyen, Shah Mohamed Bakhash, Michelle Lin, Noah R. Baker, Sean Ellis, Meei-Li Huang, Keith R Jerome, Alexander Greninger |
| EPI_ISL_2003981 | Basurto University Hospital: Clinical Microbiology Laboratory | Biocruces Bizkaia | Mikel Urrutikoetxea-Gutiérrez, Mª Carmen Nieto Toboso, Estibaliz Ugalde Zarraga, Mikel Gallego Rodrigo, Ana de la Hoz, José Luis Díaz de Tuesta del Arco |
| EPI_ISL_2004140 | Instituto Nacional de Saude (INSA) and i3S - Instituto de Investigação e Inovação em Saúde | Instituto Nacional de Saude (INSA) and i3S - Instituto de Investigação e Inovação em Saúde | Borges et al |
| EPI_ISL_2004292 | Instituto Nacional de Saude (INSA) and i3S - Instituto de Investigação e Inovação em Saúde | Instituto Nacional de Saude (INSA) | Borges et al |
| EPI_ISL_2006523 | Lighthouse Lab in Milton Keynes | Wellcome Sanger Institute for the COVID-19 Genomics UK (COG-UK) Consortium | The Lighthouse Lab in Milton Keynes and Alex Alderton, Roberto Amato, Jeffrey Barrett, Sonia Goncalves, Ewan Harrison, David K. Jackson, Ian Johnston, Dominic Kwiatkowski, Cordelia Langford, John Sillitoe on behalf of the Wellcome Sanger Institute COVID-19 Surveillance Team |
| EPI_ISL_2006684 | Max von Pettenkofer Institute, Virology, National Reference Center for Retroviruses, LMU Munich | Laboratory for Functional Genome Analysis; Dept. Genomics; Gene Center of the LMU Munich | Max Muenchhoff; Stefan Krebs; Alexander Graf; Oliver Keppler; Helmut Blum |
| EPI_ISL_2007318, EPI_ISL_2007330, EPI_ISL_2007331 | Azienda Sanitaria dell'Alto Adige Laboratorio Aziendale di Microbiologia e Virologia | Istituto di Genomica Applicata | Elisabetta Pagani, Irene Bianconi, Elisabetta Giacobazzi, Elisa Masi, Stefanie Wieser, Irena Jurman, Vera Vendramin, Gabriele Magris, Eleonora Paparelli, Davide Scaglione, Michele Morgante |
| EPI_ISL_2015212 | Minnesota Department of Health, Public Health Laboratory | Minnesota Department of Health, Public Health Laboratory | Alexandra Lorentz, Jacob Garfin, Matt Plumb, and Xiong Wang |
| EPI_ISL_2015899, EPI_ISL_2015920 | SELARL MIRIALIS CLUSES | CNR Virus des Infections Respiratoires - France SUD | Antonin Bal, Gregory Destras, Gwendolyne Burfin, Hadrien Regue, Quentin Semanas, Martine Valette, Bruno Lina, Laurence Josset |
| EPI_ISL_2015999, EPI_ISL_2016006 | MIRIALIS CLUSES BECHET | CNR Virus des Infections Respiratoires - France SUD | Antonin Bal, Gregory Destras, Gwendolyne Burfin, Hadrien Regue, Quentin Semanas, Martine Valette, Bruno Lina, Laurence Josset |
| EPI_ISL_2016154 | CNR Virus des Infections Respiratoires - France SUD | CNR Virus des Infections Respiratoires - France SUD | Antonin Bal, Gregory Destras, Gwendolyne Burfin, Hadrien Regue, Quentin Semanas, Martine Valette, Bruno Lina, Laurence Josset |
| EPI_ISL_2020526 | Istituto Zooprofilattico Sperimentale del Mezzogiorno | TIGEM | Antonio Grimaldi Patrizia Annunziata Francesco Panariello Biancamaria Pierri Claudia Tiberio Teresa Giuliano Valentina Bouche Chiara Colantuono Maria Concetta Cuomo Denise Di Concilio Lucio Di Filippo Anna Manfredi Marcello Salvi Antonio Limone Luigi Atripaldi Pellegrino Cerino Andrea Ballabio Davide Cacchiarelli |
| EPI_ISL_2021918 | Lighthouse Lab in Alderley Park | Wellcome Sanger Institute for the COVID-19 Genomics UK (COG-UK) Consortium | Jacquelyn Wynn, Mairead Hyland, The Lighthouse Lab in Alderley Park and Alex Alderton, Roberto Amato, Jeffrey Barrett, Sonia Goncalves, Ewan Harrison, David K. Jackson, Ian Johnston, Dominic Kwiatkowski, Cordelia Langford, John Sillitoe on behalf of the Wellcome Sanger Institute COVID-19 Surveillance Team |
| EPI_ISL_2023710, EPI_ISL_2024170, EPI_ISL_2027877, EPI_ISL_2028587 | Department of Virus and Microbiological Special Diagnostics, Statens Serum Institut, Copenhagen, Denmark | Aalborg University | Danish Covid-19 Genome Consortium |
| EPI_ISL_2029351, EPI_ISL_2029352 | National Virus Reference Laboratory | National Virus Reference Laboratory | Zoe Yandle, Charlene Bennett, Gabriel Gonzalez, Michael Carr, Jonathan Dean, Cillian F De Gascun |
| EPI_ISL_2029366, EPI_ISL_2029441 | Istituto Zooprofilattico Sperimentale del Mezzogiorno | TIGEM | Antonio Grimaldi Patrizia Annunziata Francesco Panariello Biancamaria Pierri Claudia Tiberio Teresa Giuliano Valentina Bouche Chiara Colantuono Maria Concetta Cuomo Denise Di Concilio Lucio Di Filippo Anna Manfredi Marcello Salvi Antonio Limone Luigi Atripaldi Pellegrino Cerino Andrea Ballabio Davide Cacchiarelli |
| EPI_ISL_2031506, EPI_ISL_2031507 | UW Virology Lab | UW Virology Lab | Pavitra Roychoudhury, Hong Xie, Lasata Shrestha, Tien V. Nguyen, Shah Mohamed Bakhash, Michelle Lin, Noah R. Baker, Sean Ellis, Meei-Li Huang, Keith R Jerome, Alexander Greninger |
| EPI_ISL_2033624, EPI_ISL_2034597 | Swedish national genomic surveillance program of SARS-CoV-2 | The Public Health Agency of Sweden | Maximilian Riess, Maria Lind Karlberg, Alma Brolund, Swedish national genomic surveillance program of SARS-CoV-2 |
| EPI_ISL_2037206, EPI_ISL_2037245, EPI_ISL_2037269 | Pandemic Response Lab - NYC | Pandemic Response Lab, R&D | Henry Lee, Michael Hammerling, Melissa Hopkins, Cybill del Castillo, Shinyoung Clair Kang, William Ward, Pradeep Bugga, Sol Rey, Dylan Law, Katharine Nelson, Haiping Hao, Jon Laurent |
| EPI_ISL_2037519, EPI_ISL_2037520, EPI_ISL_2037658, EPI_ISL_2037669 | UW Virology Lab | UW Virology Lab | Pavitra Roychoudhury, Hong Xie, Lasata Shrestha, Tien V. Nguyen, Shah Mohamed Bakhash, Michelle Lin, Noah R. Baker, Sean Ellis, Meei-Li Huang, Keith R Jerome, Alexander Greninger |
| EPI_ISL_2080296, EPI_ISL_2080312, EPI_ISL_2080341, EPI_ISL_2080346, EPI_ISL_2080348 | Azienda Sanitaria dell'Alto Adige Laboratorio Aziendale di Microbiologia e Virologia | Istituto di Genomica Applicata | Elisabetta Pagani, Irene Bianconi, Elisabetta Giacobazzi, Elisa Masi, Stefanie Wieser, Irena Jurman, Vera Vendramin, Gabriele Magris, Eleonora Paparelli, Davide Scaglione, Michele Morgante |
| EPI_ISL_2082552 | UW Virology Lab | UW Virology Lab | Pavitra Roychoudhury, Hong Xie, Lasata Shrestha, Tien V. Nguyen, Shah Mohamed Bakhash, Michelle Lin, Noah R. Baker, Sean Ellis, Meei-Li Huang, |

| | | | |
|---|---|---|---|
| | | | Keith R Jerome, Alexander Greninger |
| EPI_ISL_2086174 | Microbiology Department. Complexo Hospitalario Universitario de Vigo | Microbiology Department. Complexo Hospitalario Universitario de Vigo | Alfaya N, Alonso I, Alvarez M, Cabrera JJ, Carballo R, Cores O, Cortizo S, del-Campo V, Martinez L, Mediero G, Perez S, Potel C, Regueiro B, Rey S, Vasallo FJ |
| EPI_ISL_2088160 | National Virus Reference Laboratory | National Virus Reference Laboratory | Zoe Yandle, Charlene Bennett, Gabriel Gonzalez, Michael Carr, Jonathan Dean, Cillian F De Gascun |
| EPI_ISL_2091931 | Lighthouse Lab in Alderley Park | Wellcome Sanger Institute for the COVID-19 Genomics UK (COG-UK) Consortium | Jacquelyn Wynn, Mairead Hyland, The Lighthouse Lab in Alderley Park and Alex Alderton, Roberto Amato, Jeffrey Barrett, Sonia Goncalves, Ewan Harrison, David K. Jackson, Ian Johnston, Dominic Kwiatkowski, Cordelia Langford, John Sillitoe on behalf of the Wellcome Sanger Institute COVID-19 Surveillance Team |
| EPI_ISL_2094351 | Dutch COVID-19 response team | National Institute for Public Health and the Environment (RIVM) | Adam Meijer, Harry Vennema, Dirk Eggink, Jeroen Cremer, Sharon van den Brink, Bas van der Veer, AnneMarie van den Brandt, Lisa Wijsman, Kim Freriks, Ryanne Jaarsma, Euníce Then, Lynn Aarts, Sanne Bos, Melissa van Tuil, Linda van de Nes, Sjoerd Kuiling, James Groot, Florian Zwagemaker, Dennis Schmitz, Annelies Kroneman, Karim Hajji, Chantal Reusken, on behalf of the national COVID-19 response team |
| EPI_ISL_2094436 | cerballiance-IDF | Cerba lab | Emmanuel Lecorche, Sabine Trombert-Paolantoni, Aude Lessenne, Kader Merah, Thierry Collin, Laura Verdurme, Patrice Herisson, Stéphanie Haïm-Boukobza, Bénédicte Roquebert |
| EPI_ISL_2104621, EPI_ISL_2104664 | University Hospitals of Geneva, Laboratory of Virology | HUG, Laboratory of Virology and the Health2030 Genome Center | Samuel Cordey, Ana Rita Goncalves, Laurent Kaiser, Lorenzo Cerutti, Henri Pegeot, Melyssa Elies, Deborah Penet, Keith Harshman, Ioannis Xenarios, Emmanouil Dermitzakis |
| EPI_ISL_2105968 | Respiratory Viruses Branch, Centers for Disease Control and Prevention | Respiratory Viruses Branch, Centers for Disease Control and Prevention | Howard,D., Batra,D., Cook,P.W., Moser,K., Paskey,A., Caravas,J., Rambo-Martin,B., Morrison,S., Gulvick,C., Sammons,S., Unoarumhi,Y., Wagner,D., Roychoudhury,M.S., Xie,H., Shrestha,L., Nguyen,T.V., Bakhash,S.M., Lin,M., Baker,N.R., Perez,R., Ellis,S., Breit,N., Livingston,R.J., Huang,M.-L., Jerome,K.R., Mathias,P., Greninger,A., Paden,C.R., MacCannell,D. |
| EPI_ISL_2106358, EPI_ISL_2106547, EPI_ISL_2106548 | Pandemic Response Lab - NYC | Pandemic Response Lab, R&D | Henry Lee, Michael Hammerling, Melissa Hopkins, Cybill del Castillo, Shinyoung Clair Kang, William Ward, Pradeep Bugga, Sol Rey, Dylan Law, Katharine Nelson, Haiping Hao, Jon Laurent |
| EPI_ISL_2115393 | Kath. St.-Johannes-Gesellschaft Dortmund gGmbH | Robert Koch Institute | unknown |
| EPI_ISL_2115400, EPI_ISL_2115409, EPI_ISL_2115485, EPI_ISL_2115495 | MVZ Dr. Eberhard & Partner Dortmund | Robert Koch Institute | unknown |
| EPI_ISL_2120312, EPI_ISL_2120318, EPI_ISL_2120327, EPI_ISL_2120333, EPI_ISL_2120335, EPI_ISL_2120341, EPI_ISL_2120351, EPI_ISL_2120427, EPI_ISL_2120481, EPI_ISL_2120502 | MDI Limbach Berlin GmbH; MVZ Labor Berlin | Robert Koch Institute | unknown |
| EPI_ISL_2120555 | MVZ fur Laboratoriumsmedizin und Mikrobiologie Koblenz-Mittelrhein (Labor Koblenz) | Robert Koch Institute | unknown |
| EPI_ISL_2120568 | MDI Limbach Berlin GmbH; MVZ Labor Berlin | Robert Koch Institute | unknown |
| EPI_ISL_2122436 | MVZ Dr. Eberhard & Partner Dortmund | Robert Koch Institute | unknown |
| EPI_ISL_2122454 | Laborarztpraxis Osnabruck | Robert Koch Institute | unknown |
| EPI_ISL_2122539, EPI_ISL_2122544, EPI_ISL_2122558 | MVZ Dr. Eberhard & Partner Dortmund | Robert Koch Institute | unknown |
| EPI_ISL_2122732 | MDI Limbach Berlin GmbH; MVZ Labor Berlin | Robert Koch Institute | unknown |
| EPI_ISL_2123425 | MVZ fur Laboratoriumsmedizin und Mikrobiologie Koblenz-Mittelrhein (Labor Koblenz) | Robert Koch Institute | unknown |
| EPI_ISL_2125050 | MDI Limbach Berlin GmbH; MVZ Labor Berlin | Robert Koch Institute | unknown |
| EPI_ISL_2125094, EPI_ISL_2125110 | MVZ fur Laboratoriumsmedizin und Mikrobiologie Koblenz-Mittelrhein (Labor Koblenz) | Robert Koch Institute | unknown |
| EPI_ISL_2125114 | Labor Prof. Dr. G. Enders MVZ GbR | Robert Koch Institute | unknown |
| EPI_ISL_2128961 | MDI Limbach Berlin GmbH; MVZ Labor Berlin | Robert Koch Institute | unknown |
| EPI_ISL_2128966 | MVZ fur Laboratoriumsmedizin und Mikrobiologie Koblenz-Mittelrhein (Labor Koblenz) | Robert Koch Institute | unknown |
| EPI_ISL_2128971 | MDI Limbach Berlin GmbH; MVZ Labor Berlin | Robert Koch Institute | unknown |
| EPI_ISL_2128973 | MVZ fur Laboratoriumsmedizin und Mikrobiologie Koblenz-Mittelrhein (Labor Koblenz) | Robert Koch Institute | unknown |
| EPI_ISL_2128975, EPI_ISL_2128976 | MDI Limbach Berlin GmbH; MVZ Labor Berlin | Robert Koch Institute | unknown |
| EPI_ISL_2128980, EPI_ISL_2128983 | MVZ fur Laboratoriumsmedizin und Mikrobiologie Koblenz-Mittelrhein (Labor Koblenz) | Robert Koch Institute | unknown |
| EPI_ISL_2128988 | MDI Limbach Berlin GmbH; MVZ Labor Berlin | Robert Koch Institute | unknown |
| EPI_ISL_2129577, EPI_ISL_2129603 | MVZ Dr. Eberhard & Partner Dortmund | Robert Koch Institute | unknown |
| EPI_ISL_2129605 | Medizinisches Labor Wahl LÃ¼denscheid | Robert Koch Institute | unknown |
| EPI_ISL_2129608 | MVZ Dr. Eberhard & Partner Dortmund | Robert Koch Institute | unknown |
| EPI_ISL_2131459, EPI_ISL_2131460, EPI_ISL_2131462, EPI_ISL_2131484 | Università degli Studi di Perugia | Istituto Zooprofilattico Sperimentale dell'Abruzzo e Molise "G. Caporale" | Mencacci A, Camilloni B, Lorusso A, Marcacci M, Di Domenico M, Ancora M, Curini V, Mangone I, Rinaldi A, Delli Compagni E, Scialabba S, Di Pasquale A, Cammà C, Puglia I, Calistri P, Savini G |
| EPI_ISL_2131887, EPI_ISL_2131927, EPI_ISL_2131933, EPI_ISL_2132118 | National Virus Reference Laboratory | National Virus Reference Laboratory | Zoe Yandle, Charlene Bennett, Gabriel Gonzalez, Michael Carr, Jonathan Dean, Cillian F De Gascun |
| EPI_ISL_2136915 | Laboratori Clínic Territorial de Girona | Can Ruti SARS-CoV-2 Sequencing Hub (HUGTiP/IrsiCaixa/IGTP) | Marc Noguera-Julian, Pilar Armengol, Ignacio Blanco, Antoni E Bordoy, Francesc Catala-Moll, Pere-Joan Cardona, Maria Casadellà, Cristina Casañ, Gemma Clara, Bonaventura Clotet, Cristina Esteban, Montserrat Giménez, Mercedes Guerrero, Anna Not, Roger Paredes, Mariona Parera, Verónica Saludes, Alba Sánchez, and Elisa Martró on behalf of the Can Ruti SARS-CoV-2 Sequencing Hub. |
| EPI_ISL_2137592 | Istituto Zooprofilattico Sperimentale del Mezzogiorno | TIGEM | Antonio Grimaldi Patrizia Annunziata Francesco Panariello Biancamaria Pierri Claudia Tiberio Teresa Giuliano Valentina Bouche Chiara Colantuono Maria Concetta Cuomo Denise Di Concilio Lucio Di Filippo Anna Manfredi Marcello Salvi Antonio Limone Luigi Atripaldi Pellegrino Cerino Andrea Ballabio Davide Cacchiarelli |
| EPI_ISL_2137768 | Università degli Studi di Perugia | Istituto Zooprofilattico Sperimentale dell'Abruzzo e Molise "G. Caporale" | Mencacci A, Camilloni B, Lorusso A, Marcacci M, Di Domenico M, Ancora M, Curini V, Mangone I, Rinaldi A, Delli Compagni E, Scialabba S, Di Pasquale A, Cammà C, Puglia I, Calistri P, Savini G |
| EPI_ISL_2138808 | Ostfold Hospital Trust - Kalnes, Centre for Laboratory Medicine, Section for gene technology and infection serology | Norwegian Institute of Public Health, Department of Virology | Kathrine Stene-Johansen, Kamilla Heddeland Instefjord, Hilde Elshaug, Garcia Llorente Ignacio, Jon Bråte, Engebretsen Serina Beate,Pedersen Benedikte Nevjen, Debech Nadia, Atiya R Ali,Marie Paulsen Madsen, Rasmus Riis Kopperud, Hilde Vollan, Karoline Bragstad, Olav Hungnes |
| EPI_ISL_2140548 | Laboratori Clínic Territorial de Girona | Can Ruti SARS-CoV-2 Sequencing Hub | Marc Noguera-Julian, Pilar Armengol, Ignacio Blanco, Antoni E Bordoy, Francesc Catala-Moll, Pere-Joan Cardona, Maria Casadellà, Cristina Casañ, |

| | | | |
|---|---|---|---|
| | | (HUGTiP/IrsiCaixa/IGTP) | Gemma Clara, Bonaventura Clotet, Cristina Esteban, Montserrat Giménez, Mercedes Guerrero, Anna Not, Roger Paredes, Mariona Parera, Verónica Saludes, Alba Sánchez, and Elisa Martró on behalf of the Can Ruti SARS-CoV-2 Sequencing Hub. |
| EPI_ISL_2140632 | Microbiology Department, Laboratori Clínic Metropolitana Nord. Hospital Universitari Germans Trias i Pujol | Can Ruti SARS-CoV-2 Sequencing Hub (HUGTiP/IrsiCaixa/IGTP) | Marc Noguera-Julian, Pilar Armengol, Ignacio Blanco, Antoni E Bordoy, Francesc Catala-Moll, Pere-Joan Cardona, Maria Casadellà, Cristina Casañ, Gemma Clara, Bonaventura Clotet, Cristina Esteban, Montserrat Giménez, Mercedes Guerrero, Anna Not, Roger Paredes, Mariona Parera, Verónica Saludes, Alba Sánchez, and Elisa Martró on behalf of the Can Ruti SARS-CoV-2 Sequencing Hub. |
| EPI_ISL_2141965 | BIOR | Latvian Biomedical Research and Study Centre | Janis Pjalkovskis, Nikita Zrelovs, Monta Ustinova, Ivars Silamikelis, Liga Birzniece, Kaspars Megnis, Una Krumina, Guntars Zarins, Vita Rovite, Lauma Freimane, Laila Silamikele, Laura Ansone, Davids Fridmanis, Elina Dimina, Irena Meistere, Daina Pule, Jurijs Perevoscikovs, Uga Dumpis, Janis Klovins |
| EPI_ISL_2142669 | LAM COLONNA DE CINARCA | CNR Virus des Infections Respiratoires - France SUD | Antonin Bal, Gregory Destras, Gwendolyne Burfin, Hadrien Regue, Quentin Semanas, Martine Valette, Bruno Lina, Laurence Josset |
| EPI_ISL_2143400, EPI_ISL_2143519 | CH TOURCOING | CHU Lille - Laboratoire de Virologie | AIT YAHYA Emilie, ALIDJINOU Enagnon Kazali, BOCKET Laurence, CREPIN Michel, DEMAY Christophe, ENGELMANN Ilka, GEFFROY Sandrine, GUIGON Aurélie, LAMBERT Valérie, LAZREK Mouna, NOBILLIAUX Florian, PREVOST Brigitte, THUILLIER Caroline, TINEZ Claire |
| EPI_ISL_2143688, EPI_ISL_2143703 | Istituto Zooprofilattico Sperimentale del Mezzogiorno | Telethon Institute of Genetics and Medicine (TIGEM) | Antonio Grimaldi Patrizia Annunziata Francesco Panariello Biancamaria Pierri Claudia Tiberio Teresa Giuliano Valentina Bouche Chiara Colantuono Maria Concetta Cuomo Denise Di Concilio Lucio Di Filippo Anna Manfredi Marcello Salvi Antonio Limone Luigi Atripaldi Pellegrino Cerino Andrea Ballabio Davide Cacchiarelli |
| EPI_ISL_2151335 | Basurto University Hospital Clinical Microbiology Laboratory | Biocruces Bizkaia | Mikel Urrutikoetxea-Gutierrez, Mª Carmen Nieto Toboso, Estibaliz Ugalde Zarraga, Mikel Gallego Rodrigo, Ana de la Hoz, José Luis Díaz de Tuesta del Arco |
| EPI_ISL_2153172, EPI_ISL_2153173 | Istituto Zooprofilattico Sperimentale del Mezzogiorno | TIGEM | Antonio Grimaldi Patrizia Annunziata Francesco Panariello Biancamaria Pierri Claudia Tiberio Teresa Giuliano Valentina Bouche Chiara Colantuono Maria Concetta Cuomo Denise Di Concilio Lucio Di Filippo Anna Manfredi Marcello Salvi Antonio Limone Luigi Atripaldi Pellegrino Cerino Andrea Ballabio Davide Cacchiarelli |
| EPI_ISL_2153429 | Fimlab Laboratories | Fimlab Laboratories | Minna Paloniemi, Leena Huhti, Sara Lehtinen, Bruno Luukinen, Tapio Seiskari, Mauri Keinänen |
| EPI_ISL_2156070 | Laboratorio Aziendale di Microbiologia e Virologia, Azienda Sanitaria dell'Alto Adige | Laboratorio Aziendale di Microbiologia e Virologia, Azienda Sanitaria dell'Alto Adige | Irene Bianconi, Bartolomeo Mobilio Rodriguez, Elisabetta Giacobazzi, Elisa Masi, Stefanie Wieser, Anne Picard, Claudia Volpato, Chiara Cantaloni, Elisabetta Pagani |
| EPI_ISL_2156768 | Centre de Recherches Médicales de Lambaréné (CERMEL) | Centre de Recherches Médicales de Lambaréné (CERMEL) | Gédéon Prince Manouana, Moustapha Nzamba Maloum, Sam O'neilla Oye Bingono, Georgelin Nguema Ondo, Rodrigue Bikangui, Samira Zoa Assoumou, Srinivas reddy Pallerla, Jean Bernard Lekana-Douki, Joël-Fleury Djoba Siawaya, Steffen Borrmann, Thirumalaisamy P. Velavan, Bertrand Lell and Ayola Akim Adegnika |
| EPI_ISL_2156841 | CSIR-Centre for Cellular and Molecular Biology | CSIR-Centre for Cellular and Molecular Biology-INSACOG | Tulasi Nagabandi, Lamuk Zaveri,Ara Sreenivas,Shreekant Verma, Amareshwar Vodapalli ,Blessy B John,Viswagithe S L,B Himasri,Valli Nagalakshmi Undamatla,Payel Mukherjee,Sofia Banu,Archana Bharadwaj Siva,Sharath Chandra Thota,Karthik Bharadwaj Tallapaka,Rakesh K Mishra,Divya Tej Sowpati |
| EPI_ISL_2156972 | Mapmygenome | CSIR-Centre for Cellular and Molecular Biology-INSACOG | Ara Sreenivas,Shreekant Verma,Tulasi Nagabandi, Lamuk Zaveri, Amareshwar Vodapalli ,Blessy B John,Viswagithe S L,B Himasri,Valli Nagalakshmi Undamatla,Payel Mukherjee,Sofia Banu,Archana Bharadwaj Siva,Sharath Chandra Thota,Karthik Bharadwaj Tallapaka,Rakesh K Mishra,Divya Tej Sowpati |
| EPI_ISL_2157073 | Mapmygenome | CSIR-Centre for Cellular and Molecular Biology-INSACOG | Payel Mukherjee,Lamuk Zaveri,Tulasi Nagabandi, Ara Sreenivas,Shreekant Verma, Amareshwar Vodapalli ,Blessy B John,Viswagithe S L,B Himasri,Valli Nagalakshmi Undamatla,Sofia Banu,Archana Bharadwaj Siva,Sharath Chandra Thota,Karthik Bharadwaj Tallapaka,Rakesh K Mishra,Divya Tej Sowpati |
| EPI_ISL_2157166 | Mapmygenome | CSIR-Centre for Cellular and Molecular Biology - INSACOG | Ara Sreenivas, Onkar Kulkarni, Lamuk Zaveri, Sofia Banu, Shreekant Verma, Amareshwar Vodapalli , Viswagithe S L, B Himasri, Sharath Chandra Thota, Karthik Bharadwaj Tallapaka, Rakesh K Mishra,Divya Tej Sowpati |
| EPI_ISL_2164238 | Basurto University Hospital: Clinical Microbiology Laboratory | Biocruces Bizkaia | Mikel Urrutikoetxea-Gutiérrez, Mª Carmen Nieto Toboso, Estibaliz Ugalde Zarraga, Mikel Gallego Rodrigo, Ana de la Hoz, José Luis Díaz de Tuesta del Arco |
| EPI_ISL_2164421 | Basurto University Hospital: Clinical Microbiology Laboratory | Biocruces Bizkaia | Basurto University Hospital: Clinical Microbiology Laboratory |
| EPI_ISL_2168688, EPI_ISL_2168693, EPI_ISL_2168694, EPI_ISL_2168696, EPI_ISL_2168697, EPI_ISL_2168698, EPI_ISL_2168699, EPI_ISL_2168700, EPI_ISL_2168702, EPI_ISL_2168705, EPI_ISL_2168707, EPI_ISL_2168708, EPI_ISL_2168709, EPI_ISL_2168711, EPI_ISL_2168712, EPI_ISL_2168714, EPI_ISL_2168715, EPI_ISL_2168716, EPI_ISL_2168717, EPI_ISL_2168718, EPI_ISL_2168721, EPI_ISL_2168723, EPI_ISL_2168724, EPI_ISL_2168725, EPI_ISL_2168728, EPI_ISL_2168730, EPI_ISL_2168741, EPI_ISL_2168746, EPI_ISL_2169255, EPI_ISL_2169256, EPI_ISL_2169258, EPI_ISL_2169266, EPI_ISL_2169274, EPI_ISL_2169280, EPI_ISL_2169281, EPI_ISL_2169283, EPI_ISL_2169285, EPI_ISL_2169289, EPI_ISL_2169290, EPI_ISL_2169294, EPI_ISL_2169297, EPI_ISL_2169309, EPI_ISL_2169365, EPI_ISL_2169499, EPI_ISL_2169505, EPI_ISL_2169515, EPI_ISL_2169519, EPI_ISL_2169527, EPI_ISL_2169540, EPI_ISL_2169544, EPI_ISL_2169654, EPI_ISL_2169655, EPI_ISL_2169656, EPI_ISL_2169663, EPI_ISL_2169670, EPI_ISL_2169894, EPI_ISL_2169911, EPI_ISL_2169926 | | | |
| see above | Alberta Precision Labs (APL) | Public Health Agency of Canada (PHAC) National Microbiology Laboratory | Buss, E, Croxen M, Deo A, Dieu P, Gill K, Ferrato C, Khan F, Koleva P, Li V, Lloyd C, Lynch T, Ma R, Murphy S, Pabbaraju K, Shokoples S, Tipples G, Thayer J, Whitehouse M, Wong A, Yu C, Zelyas N |
| EPI_ISL_2171315, EPI_ISL_2171556 | Istituto Zooprofilattico Sperimentale del Mezzogiorno | TIGEM | Antonio Grimaldi Patrizia Annunziata Francesco Panariello Biancamaria Pierri Claudia Tiberio Teresa Giuliano Valentina Bouche Chiara Colantuono Maria Concetta Cuomo Denise Di Concilio Lucio Di Filippo Anna Manfredi Marcello Salvi Antonio Limone Luigi Atripaldi Pellegrino Cerino Andrea Ballabio Davide Cacchiarelli |
| EPI_ISL_2171878, EPI_ISL_2171984 | Lighthouse Lab in Alderley Park | Wellcome Sanger Institute for the COVID-19 Genomics UK (COG-UK) Consortium | Jacquelyn Wynn, Mairead Hyland, The Lighthouse Lab in Alderley Park and Alex Alderton, Roberto Amato, Jeffrey Barrett, Sonia Goncalves, Ewan Harrison, David K. Jackson, Ian Johnston, Dominic Kwiatkowski, Cordelia Langford, John Sillitoe on behalf of the Wellcome Sanger Institute COVID-19 Surveillance Team |
| EPI_ISL_2178480 | Labo analyses med | National Reference Center for Viruses of Respiratory Infections, Institut Pasteur, Paris | Marion Barbet, Sylvie Behillil, Méline Bizard, Angela Brisebarre, Camille Capel, Vincent Enouf, Louise Lefrançois, Frédéric Lemoine, Christophe Malabat, Corinne Maufrais, Amaury Vaysse, Etienne Simon-Lorière, Maud Vanpeene, Sylvie Van der Werf ,Jean-FrançOis Comes |
| EPI_ISL_2178832 | Hospital General Universitario Gregorio Marañón | Hospital General Universitario Gregorio Marañón | Sergio Buenestado Serrano, Pedro Sola Campoy, Laura Pérez-Lago, Cristina Rodriguez-Grande, Marta Herranz Martin, Victor de la Cueva, Pilar Catalán, Patricia Muñoz, Darío García de Viedma |
| EPI_ISL_2179503 | SELAS BC-LAB | Department of Virology, Henri Mondor University Hospital, Assistance Publique Hôpitaux de Paris, Université Paris-Est Créteil, INSERM U955 | Christophe Rodriguez, Slim Fourati, Vanessa Demontant, Guillaume Gricourt, Melissa N'Debi, Alexandre Soulier, Elisabeth Trawinski, Jean-Michel Pawlotsky |
| EPI_ISL_2179747, EPI_ISL_2179748, EPI_ISL_2179749 | Servicio Microbiología Hospital La Paz | Servicio Microbiología Hospital La Paz | Fernando Lázaro, Rubén Cáceres, Jesús Mingorance Cruz, Elie Dahdouh |
| EPI_ISL_2188356 | Labo Analyses Med | National Reference Center for Viruses of Respiratory Infections, Institut Pasteur, Paris | Marion Barbet, Sylvie Behillil, Méline Bizard, Angela Brisebarre, Camille Capel, Vincent Enouf, Louise Lefrançois, Frédéric Lemoine, Christophe Malabat, Corinne Maufrais, Damien Mornico, Etienne Simon-Lorière, Maud Vanpeene, Sylvie Van der Werf ,Franck Ennouchi |
| EPI_ISL_2188485 | Labo Analyses Med | National Reference Center for Viruses of Respiratory Infections, Institut Pasteur, Paris | Marion Barbet, Sylvie Behillil, Méline Bizard, Angela Brisebarre, Camille Capel, Vincent Enouf, Louise Lefrançois, Frédéric Lemoine, Christophe Malabat, Corinne Maufrais, Gael Millot, Etienne Simon-Lorière, Maud Vanpeene, Sylvie Van der Werf ,Adamou Lagare |
| EPI_ISL_2193398, EPI_ISL_2193399 | University Hospitals Translational Laboratory (UHTL), University Hospitals | University Hospitals Translational Laboratory (UHTL), University Hospitals | Sadri,N., Alouani,D., Song,X. |
| EPI_ISL_2193872 | National Virus Reference Laboratory | National Virus Reference Laboratory | Zoe Yandle, Charlene Bennett, Gabriel Gonzalez, Michael Carr, Jonathan Dean, Cillian F De Gascun |
| EPI_ISL_2194359, EPI_ISL_2194447, EPI_ISL_2194467 | Istituto Zooprofilattico Sperimentale del Mezzogiorno | TIGEM | Antonio Grimaldi Patrizia Annunziata Francesco Panariello Biancamaria Pierri Claudia Tiberio Teresa Giuliano Valentina Bouche Chiara Colantuono Maria Concetta Cuomo Denise Di Concilio Lucio Di Filippo Anna Manfredi Marcello Salvi Antonio Limone Luigi Atripaldi Pellegrino Cerino Andrea Ballabio Davide Cacchiarelli |
| EPI_ISL_2195785, EPI_ISL_2195786, EPI_ISL_2195787, EPI_ISL_2195792, EPI_ISL_2195794, EPI_ISL_2195797, EPI_ISL_2195800, EPI_ISL_2195801, EPI_ISL_2195803, EPI_ISL_2195804, EPI_ISL_2195868, EPI_ISL_2195873, EPI_ISL_2195877 | | | |
| see above | Berkeley Medical Center | WVU and Marshall University Combined Genomics Core Facilities | James Denvir, Peter Stoilov, Peter Perrotta, Wesley Kimble, Ryan Percifield |
| EPI_ISL_2196064, EPI_ISL_2196067, | BMC LAB | WVU and Marshall University Combined Genomics Core | James Denvir, Peter Stoilov, Peter Perrotta, Wesley Kimble, Ryan Percifield |

| | | | |
|---|---|---|---|
| EPI_ISL_2196070 | | Facilities | |
| EPI_ISL_2196111 | West Virigina Labs | WVU and Marshall University Combined Genomics Core Facilities | James Denvir, Peter Stoilov, Peter Perrotta, Wesley Kimble, Ryan Percifield |
| EPI_ISL_2196170 | QLabs | WVU and Marshall University Combined Genomics Core Facilities | James Denvir, Peter Stoilov, Peter Perrotta, Wesley Kimble, Ryan Percifield |
| EPI_ISL_2199343 | Lighthouse Lab in Alderley Park | Wellcome Sanger Institute for the COVID-19 Genomics UK (COG-UK) Consortium | Jacquelyn Wynn, Mairead Hyland, The Lighthouse Lab in Alderley Park and Alex Alderton, Roberto Amato, Jeffrey Barrett, Sonia Goncalves, Ewan Harrison, David K. Jackson, Ian Johnston, Dominic Kwiatkowski, Cordelia Langford, John Sillitoe on behalf of the Wellcome Sanger Institute COVID-19 Surveillance Team |
| EPI_ISL_2202386, EPI_ISL_2202387 | Ministry of Health Turkey | Ministry of Health Turkey | Fatma Bayrakdar, Yasemin Cosgun, Suleyman Yalcin, Gulay Korukluoglu |
| EPI_ISL_2204356, EPI_ISL_2204389, EPI_ISL_2204392 | BMC LAB | WVU and Marshall University Combined Genomics Core Facilities | James Denvir, Peter Stoilov, Peter Perrotta, Wesley Kimble, Ryan Percifield |
| EPI_ISL_2204409, EPI_ISL_2204413, EPI_ISL_2204414, EPI_ISL_2204424, EPI_ISL_2204425, EPI_ISL_2204431, EPI_ISL_2204438, EPI_ISL_2204444, EPI_ISL_2204459, EPI_ISL_2204472, EPI_ISL_2204478, EPI_ISL_2204489, EPI_ISL_2204512, EPI_ISL_2204514 | | | |
| see above | Berkeley Medical Center | WVU and Marshall University Combined Genomics Core Facilities | James Denvir, Peter Stoilov, Peter Perrotta, Wesley Kimble, Ryan Percifield |
| EPI_ISL_2204735 | BMC LAB | WVU and Marshall University Combined Genomics Core Facilities | James Denvir, Peter Stoilov, Peter Perrotta, Wesley Kimble, Ryan Percifield |
| EPI_ISL_2204748, EPI_ISL_2204756, EPI_ISL_2204776, EPI_ISL_2204823 | Berkeley Medical Center | WVU and Marshall University Combined Genomics Core Facilities | James Denvir, Peter Stoilov, Peter Perrotta, Wesley Kimble, Ryan Percifield |
| EPI_ISL_2213595 | University Hospitals of Geneva, Laboratory of Virology | HUG, Laboratory of Virology and the Health2030 Genome Center | Samuel Cordey, Ana Rita Goncalves, Laurent Kaiser, Lorenzo Cerutti, Henri Pegeot, Melyssa Elies, Deborah Penet, Keith Harshman, Ioannis Xenarios, Emmanouil Dermitzakis |
| EPI_ISL_2225610, EPI_ISL_2225611 | Basurto University Hospital: Clinical Microbiology Laboratory | Biocruces Bizkaia | Mikel Urrutikoetxea-Gutiérrez, Mª Carmen Nieto Toboso, Estibaliz Ugalde Zarraga, Mikel Gallego Rodrigo, Ana de la Hoz, José Luis Díaz de Tuesta del Arco |
| EPI_ISL_2226117 | Furst Medical Laboratory | Norwegian Institute of Public Health, Department of Virology | Kathrine Stene-Johansen, Kamilla Heddeland Instefjord, Hilde Elshaug, Garcia Llorente Ignacio, Jon Bråte, Engebretsen Serina Beate,Pedersen Benedikte Nevjen, Debech Nadia, Atiya R Ali,Marie Paulsen Madsen, Rasmus Riis Kopperud, Hilde Vollan, Karoline Bragstad, Olav Hungnes |
| EPI_ISL_2226754, EPI_ISL_2226755, EPI_ISL_2226759 | ADMED Microbiologie | Genomics and Transcriptomics, Philip Morris International | Reto Lienhard, Marie-Lise Tritten, Emmanuel Guedj, Nicolas Sierro, Rémi Dulize, David Bornand, Mehdi Auberson, Maxime Berthouzoz, Nikolai Ivanov, Manuel Peitsch |
| EPI_ISL_2227039, EPI_ISL_2227044, EPI_ISL_2227048, EPI_ISL_2227064, EPI_ISL_2227070, EPI_ISL_2227074 | Università Federico II - Dipartimento di scienze mediche traslazionali - Napoli | TIGEM | Antonio Grimaldi Patrizia Annunziata Francesco Panariello Teresa Giuliano Michele Cennamo Valentina Bouche Chiara Colantuono Lucio Di Filippo Mariano Fiorenza Anna Manfredi Marcello Salvi Giuseppe Portella Andrea Ballabio Davide Cacchiarelli |
| EPI_ISL_2227095 | Istituto Zooprofilattico Sperimentale del Mezzogiorno | TIGEM | Antonio Grimaldi Patrizia Annunziata Francesco Panariello Biancamaria Pierri Claudia Tiberio Teresa Giuliano Valentina Bouche Chiara Colantuono Maria Concetta Cuomo Denise Di Concilio Lucio Di Filippo Anna Manfredi Marcello Salvi Antonio Limone Luigi Atripaldi Pellegrino Cerino Andrea Ballabio Davide Cacchiarelli |
| EPI_ISL_2235210 | Pandemic Response Lab - NYC | Pandemic Response Lab, R&D | Henry Lee, Michael Hammerling, Melissa Hopkins, Cybill del Castillo, Shinyoung Clair Kang, William Ward, Pradeep Bugga, Sol Rey, Dylan Law, Katharine Nelson, Haiping Hao, Jon Laurent |
| EPI_ISL_2235459 | Università degli Studi di Perugia | Istituto Zooprofilattico Sperimentale dell'Abruzzo e Molise "G. Caporale" | Mencacci A, Camilloni B, Lorusso A, Marcacci M, Di Domenico M, Ancora M, Curini V, Di Lollo Valeria, Mangone I, Rinaldi A, Delli Compagni E, Scialabba S, Caporale M, Di Pasquale A, Cammà C, Puglia I, Calistri P, Savini G |
| EPI_ISL_2236335, EPI_ISL_2236373, EPI_ISL_2237742 | Lighthouse Lab in Alderley Park | Wellcome Sanger Institute for the COVID-19 Genomics UK (COG-UK) Consortium | Jacquelyn Wynn, Mairead Hyland, The Lighthouse Lab in Alderley Park and Alex Alderton, Roberto Amato, Jeffrey Barrett, Sonia Goncalves, Ewan Harrison, David K. Jackson, Ian Johnston, Dominic Kwiatkowski, Cordelia Langford, John Sillitoe on behalf of the Wellcome Sanger Institute COVID-19 Surveillance Team |
| EPI_ISL_2239501 | Northumbria University / South Tees Hospitals NHS Foundation Trust / North Cumbria Integrated Care NHS Foundation Trust / North Tees and Hartlepool NHS Foundation Trust / Newcastle Hospitals NHS Foundation Trust | COVID-19 Genomics UK (COG-UK) Consortium | Darren L Smith,Andrew Nelson,Matthew Bashton,Greg R Young,Joshua Loh,John Allan,Mohammad A Tariq,Giles S Holt,Gary Black,Wen C Yew,Lynn Dover,Paul Baker,Steve Liggett,Sarah Essex,Jane Greenaway,Debra Padgett,Clive Graham,Garren Scott,Edward Barton,Emma Swindells,Brendan Payne,Jennifer Collins,Yusri Taha,Gary Eltringham |
| EPI_ISL_2240830, EPI_ISL_2240985, EPI_ISL_2241015, EPI_ISL_2241025, EPI_ISL_2241138, EPI_ISL_2241212, EPI_ISL_2241259, EPI_ISL_2241309 | National Virus Reference Laboratory | National Virus Reference Laboratory | Zoe Yandle, Charlene Bennett, Gabriel Gonzalez, Michael Carr, Jonathan Dean, Cillian F De Gascun |
| EPI_ISL_2241690, EPI_ISL_2241691, EPI_ISL_2241692, EPI_ISL_2241693 | Laboratory Corporation of America | Centers for Disease Control and Prevention Division of Viral Diseases, Pathogen Discovery | Dakota Howard, Dhwani Batra, Peter W. Cook, Kara Moser, Adrian Paskey, Jason Caravas, Benjamin Rambo-Martin, Shatavia Morrison, Christopher Gulvick, Scott Sammons, Yvette Unoarumhi, Darlene Wagner, Matthew Schmerer, Minoo Agarwal, Eyad Almasri, Debbie Boles, Ayla Burns, Nuthawin Charoensri, Oren Cohen, Susan Countryman, Mary Ann Cristobal, Bobbi Croy, Suzanne Dale, Hrushikesh Deshmukh, Amanda Douglas, Vincent Drouillon, Marcia Eisenberg, Howard Engler, Rama Ghatti, Prashant Gupta, Susan Hicks, Jake Humphrey, Lax Iyer, Manoj Jain, Mohan Kolli, Brian Krueger, Tim Kuphal, Stanley Letovsky, Michael Levandoski, Craig Lukasik, Jonathan Meltzer, Brian Norvell, Mindy Nye, Scott Parker, Christos Petropoulos, John Pruitt, Steven Ragan, Scott Ryan, Mike Sapeta, Jana Schroth, Suresh Babu Selvaraju, Goran Stevovic, Amanda Suchanek, Andrea Throop, Lyndon Tilson, Thomas Urban, Joe Voshell, Kimberly Wagner, Jonathan Williams, Mary Williamson, Qian Zeng, Tricia Zwiefelhofer, Clinton R. Paden, Duncan MacCannell |
| EPI_ISL_2249022 | unknown | Instituto Nacional de Saude (INSA) | Borges et al |
| EPI_ISL_2253206, EPI_ISL_2253208, EPI_ISL_2253210 | Respiratory Viruses Branch, Centers for Disease Control and Prevention | Respiratory Viruses Branch, Centers for Disease Control and Prevention | Howard,D., Batra,D., Cook,P.W., Moser,K., Paskey,A., Caravas,J., Rambo-Martin,B., Morrison,S., Gulvick,C., Sammons,S., Unoarumhi,Y., Wagner,D., Schmerer,M., Agarwal,M., Almasri,E., Boles,D., Burns,A., Charoensri,N., Cohen,O., Countryman,S., Cristobal,M.A., Croy,B., Dale,S., Deshmukh,H., Douglas,A., Drouillon,V., Eisenberg,M., Engler,H., Ghatti,R., Gupta,P., Hicks,S., Humphrey,J., Iyer,L., Jain,M., Kolli,M., Krueger,B., Kuphal,T., Letovsky,S., Levandoski,M., Lukasik,C., Meltzer,J., Norvell,B., Nye,M., Parker,S., Petropoulos,C., Pruitt,J., Ragan,S., Ryan,S., Sapeta,M., Schroth,J., Selvaraju,S.B., Stevovic,G., Suchanek,A., Throop,A., Tilson,L., Urban,T., Voshell,J., Wagner,K., Williams,J., Williamson,M., Zeng,Q., Zwiefelhofer,T., Paden,C.R., MacCannell,D. |
| EPI_ISL_2254233 | Microbiology Department, Laboratori Clínic Metropolitana Nord. Hospital Universitari Germans Trias i Pujol. | Can Ruti SARS-CoV-2 Sequencing Hub (HUGTiP/IrsiCaixa/IGTP) | Marc Noguera-Julian, Pilar Armengol, Ignacio Blanco, Antoni E Bordoy, Francesc Catala-Moll, Pere-Joan Cardona, Maria Casadellà, Cristina Casañ, Gemma Clara, Bonaventura Clotet, Cristina Esteban, Montserrat Giménez, Mercedes Guerrero, Anna Not, Roger Paredes, Mariona Parera, Verónica Saludes, Alba Sánchez, and Elisa Martró on behalf of the Can Ruti SARS-CoV-2 Sequencing Hub. |
| EPI_ISL_2260982, EPI_ISL_2260986, EPI_ISL_2260989, EPI_ISL_2260992, EPI_ISL_2260996, EPI_ISL_2260998, EPI_ISL_2261021, EPI_ISL_2261024, EPI_ISL_2261036 | Laborarztpraxis Osnabrück | Robert Koch Institute | unknown |
| EPI_ISL_2261046 | MVZ Dr. Eberhard & Partner Dortmund | Robert Koch Institute | unknown |
| EPI_ISL_2263037 | Labor Dr. Wisplinghoff - Köln | Robert Koch Institute | unknown |
| EPI_ISL_2263120, EPI_ISL_2264544 | MDI Limbach Berlin GmbH; MVZ Labor Berlin | Robert Koch Institute | unknown |

| | | | |
|---|---|---|---|
| EPI_ISL_2264550 | Labor Prof. Dr. G. Enders MVZ GbR | Robert Koch Institute | unknown |
| EPI_ISL_2264712 | MVZ für Laboratoriumsmedizin und Mikrobiologie Koblenz-Mittelrhein (Labor Koblenz) | Robert Koch Institute | unknown |
| EPI_ISL_2264731, EPI_ISL_2264733 | MDI Limbach Berlin GmbH; MVZ Labor Berlin | Robert Koch Institute | unknown |
| EPI_ISL_2265532 | Labor Prof. Dr. G. Enders MVZ GbR | Robert Koch Institute | unknown |
| EPI_ISL_2266641 | MVZ Labor Krone GbR | Robert Koch Institute | unknown |
| EPI_ISL_2275970 | Istituto Zooprofilattico Sperimentale del Mezzogiorno - Azienda Ospedaliera "Pugliese Ciaccio" di Catanzaro | TIGEM | Antonio Grimaldi Patrizia Annunziata Francesco Panariello Biancamaria Pierri Claudia Tiberio Teresa Giuliano Valentina Bouche Chiara Colantuono Maria Concetta Cuomo Denise Di Concilio Lucio Di Filippo Anna Manfredi Pasquale Minchella Marcello Salvi Antonio Limone Luigi Atripaldi Pellegrino Cerino Andrea Ballabio Davide Cacchiarelli |
| EPI_ISL_2282048, EPI_ISL_2282136 | Istituto Zooprofilattico Sperimentale del Mezzogiorno | TIGEM | Antonio Grimaldi Patrizia Annunziata Francesco Panariello Biancamaria Pierri Claudia Tiberio Teresa Giuliano Valentina Bouche Chiara Colantuono Maria Concetta Cuomo Denise Di Concilio Lucio Di Filippo Anna Manfredi Marcello Salvi Antonio Limone Luigi Atripaldi Pellegrino Cerino Andrea Ballabio Davide Cacchiarelli |
| EPI_ISL_2303932 | Dutch COVID-19 response team | National Institute for Public Health and the Environment (RIVM) | Adam Meijer, Harry Vennema, Dirk Eggink, Jeroen Cremer, Sharon van den Brink, Bas van der Veer, AnneMarie van den Brandt, Lisa Wijsman, Kim Freriks, Ryanne Jaarsma, Euníce Then, Lynn Aarts, Sanne Bos, Melissa van Tuil, Linda van de Nes, Sjoerd Kuiling, James Groot, Florian Zwagemaker, Dennis Schmitz, Annelies Kroneman, Karim Hajji, Chantal Reusken, on behalf of the national COVID-19 response team |
| EPI_ISL_2306684, EPI_ISL_2307166 | Laboratory Corporation of America | Centers for Disease Control and Prevention Division of Viral Diseases, Pathogen Discovery | Dakota Howard, Dhwani Batra, Peter W. Cook, Kara Moser, Adrian Paskey, Jason Caravas, Benjamin Rambo-Martin, Shatavia Morrison, Christopher Gulvick, Scott Sammons, Yvette Unoarumhi, Darlene Wagner, Matthew Schmerer, Minoo Agarwal, Eyad Almasri, Debbie Boles, Ayla Burns, Nuthawin Charoensri, Oren Cohen, Susan Countryman, Mary Ann Cristobal, Bobbi Croy, Suzanne Dale, Hrushikesh Deshmukh, Amanda Douglas, Vincent Drouillon, Marcia Eisenberg, Howard Engler, Rama Ghatti, Prashant Gupta, Susan Hicks, Jake Humphrey, Lax Iyer, Manoj Jain, Mohan Kolli, Brian Krueger, Tim Kuphal, Stanley Letovsky, Michael Levandoski, Craig Lukasik, Jonathan Meltzer, Brian Norvell, Mindy Nye, Scott Parker, Christos Petropoulos, John Pruitt, Steven Ragan, Scott Ryan, Mike Sapeta, Jana Schroth, Suresh Babu Selvaraju, Goran Stevovic, Amanda Suchanek, Andrea Throop, Lyndon Tilson, Thomas Urban, Joe Voshell, Kimberly Wagner, Jonathan Williams, Mary Williamson, Qian Zeng, Tricia Zwiefelhofer, Clinton R. Paden, Duncan MacCannell |
| EPI_ISL_2308562, EPI_ISL_2308563, EPI_ISL_2308571, EPI_ISL_2308572, EPI_ISL_2308573, EPI_ISL_2308593, EPI_ISL_2308595, EPI_ISL_2308596 | SIESP DIPARTIMENTO DI PREVENZIONE TERAMO | Istituto Zooprofilattico Sperimentale dell'Abruzzo e Molise "G. Caporale" | Lorusso A, Marcacci M, Di Domenico M, Ancora M, Curini V, Di Lollo Valeria, Mangone I, Rinaldi A, Delli Compagni E, Scialabba S, Caporale M, Di Pasquale A, Cammà C, Puglia I, Calistri P, Savini G |
| EPI_ISL_2308706 | Università degli Studi di Perugia | Istituto Zooprofilattico Sperimentale dell'Abruzzo e Molise "G. Caporale" | Mencacci A, Camilloni B, Biagetti M, GiammarioliM, Lorusso A, Marcacci M, Di Domenico M, Ancora M, Curini V, Mangone I, Rinaldi A, Delli Compagni E, Scialabba S, Di Pasquale A, Cammà C, Puglia I, Calistri P, Savini G |
| EPI_ISL_2313255, EPI_ISL_2313261, EPI_ISL_2313269, EPI_ISL_2313274, EPI_ISL_2313280, EPI_ISL_2313295 | Laborarztpraxis Osnabrück | Robert Koch Institute | unknown |
| EPI_ISL_2313305 | MVZ Dr. Eberhard & Partner Dortmund | Robert Koch Institute | unknown |
| EPI_ISL_2313307, EPI_ISL_2313323, EPI_ISL_2313337 | Laborarztpraxis Osnabrück | Robert Koch Institute | unknown |
| EPI_ISL_2314411, EPI_ISL_2314664 | MDI Limbach Berlin GmbH; MVZ Labor Berlin | Robert Koch Institute | unknown |
| EPI_ISL_2314778 | MVZ für Laboratoriumsmedizin und Mikrobiologie Koblenz-Mittelrhein (Labor Koblenz) | Robert Koch Institute | unknown |
| EPI_ISL_2314790 | MDI Limbach Berlin GmbH; MVZ Labor Berlin | Robert Koch Institute | unknown |
| EPI_ISL_2323015 | cerballiance-IDF | Cerba lab | Emmanuel Lecorche, Sabine Trombert-Paolantoni, Aude Lessenne, Kader Merah, Thierry Collin, Laura Verdurme, Patrice Herisson, Stéphanie Haïm-Boukobza, Bénédicte Roquebert |
| EPI_ISL_2323467 | CH ANGOULEME | CHU Poitiers | Caroline MICHAUD, Maxime PICHON, Manon PRAT, Valentin BON-BARET, Birama N'DIAYE, Magali GARCIA, Agnes BEBY-DEFAUX, Nicolas LEVEQUE |
| EPI_ISL_2331636 | Johns Hopkins Hospital Department of Pathology | Johns Hopkins Hospital Department of Pathology | C. Paul Morris, Chun Huai Luo, Adannaya Amadi, Matthew Schwartz, Nicholas Gallagher, Heba H. Mostafa |
| EPI_ISL_2332583 | Microbiology Department, University Hospital Donostia | Microbiology Department, University Hospital Donostia | Marimon JM, Montes M, Piñeiro L, Grandioso D, Martin-Peñaranda T, Gomez M, Cilla G. |
| EPI_ISL_2332616 | Azienda Sanitaria Locale di Piacenza - Presidio Ospedaliero - Laboratorio di Microbiologia | Istituto Zooprofilattico Sperimentale della Lombardia e dell'Emilia Romagna (IZSLER), Risk Analysis and Genomic Epidemiology Unit | Lo Cascio Giuliana, Schiavo Roberta, Peroni Irene, Marina Morganti, Ilaria Menozzi, Erika Scaltriti, Stefano Pongolini |
| EPI_ISL_2332986 | UW Virology Lab | UW Virology Lab | Pavitra Roychoudhury, Hong Xie, Lasata Shrestha, Tien V. Nguyen, Shah Mohamed Bakhash, Michelle Lin, Noah R. Baker, Ricardo Perez, Sean Ellis, Nathan Breit, Robert J. Livingston, Meei-Li Huang, Keith R Jerome, Patrick Mathias, Alexander Greninger |
| EPI_ISL_2333501 | Norwegian Institute of Public Health, Department of Virology | Norwegian Institute of Public Health, Department of Virology | 'Kathrine Stene-Johansen, Kamilla Heddeland Instefjord, Hilde Elshaug, Garcia Llorente Ignacio, Jon Bråte, Engebretsen Serina Beate,Pedersen Benedikte Nevjen, Debech Nadia, Line Victoria Moen, Atiya R Ali,Marie Paulsen Madsen, Rasmus Riis Kopperud, Hilde Vollan, Karoline Bragstad, Olav Hungnes |
| EPI_ISL_2340278 | Instituto Nacional de Saude (INSA) | Instituto Nacional de Saude (INSA) | Borges et al |
| EPI_ISL_2340797 | UW Virology Lab | UW Virology Lab | Pavitra Roychoudhury, Hong Xie, Lasata Shrestha, Tien V. Nguyen, Shah Mohamed Bakhash, Michelle Lin, Noah R. Baker, Sean Ellis, Meei-Li Huang, Keith R Jerome, Alexander Greninger |
| EPI_ISL_2346425, EPI_ISL_2346427, EPI_ISL_2346428, EPI_ISL_2346429, EPI_ISL_2346433, EPI_ISL_2346435 | MRC/UVRI & LSHTM Uganda Research Unit, Central Public Health Laboratories | MRC/UVRI & LSHTM Uganda Research Unit, Central Public Health Laboratories | Matthew Cotten, Dan Lule Bugembe, My V.T. Phan, Pontiano Kaleebu,Isaac Sseeewanyana, Patrick Semanda, Susan Nabadda |
| EPI_ISL_2348189 | Lighthouse Lab in Alderley Park | Wellcome Sanger Institute for the COVID-19 Genomics UK (COG-UK) Consortium | Jacquelyn Wynn, Mairead Hyland, The Lighthouse Lab in Alderley Park and Alex Alderton, Roberto Amato, Jeffrey Barrett, Sonia Goncalves, Ewan Harrison, David K. Jackson, Ian Johnston, Dominic Kwiatkowski, Cordelia Langford, John Sillitoe on behalf of the Wellcome Sanger Institute COVID-19 Surveillance Team |
| EPI_ISL_2348662 | Genome Center | Genome Center | Ovinu Kibria Islam, M. Tanvir Islam, A. S. M. Rubayet Ul Alam, Md. Shazid Hasan, Shovon Lal Sarkar, Ali Ahsan Setu, Tanay Chakrabarty,Prosanto Kumar Das, M. Shaminur Rahman, Hassan M. Al-Emran, Iqbal Kabir Jahid, M. Anwar Hossain |
| EPI_ISL_2361529 | CHU LILLE | CHU Lille - Laboratoire de Virologie | AIT YAHYA Emilie, ALIDJINOU Enagnon Kazali, BOCKET Laurence, CREPIN Michel, DEMAY Christophe, ENGELMANN Ilka, GEFFROY Sandrine, GUIGON Aurélie, LAMBERT Valérie, LAZREK Mouna, NOBILLIAUX Florian, PREVOST Brigitte, THUILLIER Caroline, TINEZ Claire ,TCHANTCHOU NJOSSE YANICK |
| EPI_ISL_2363193 | National Virus Reference Laboratory | National Virus Reference Laboratory | Zoe Yandle, Charlene Bennett, Gabriel Gonzalez, Michael Carr, Jonathan Dean, Cillian F De Gascun |
| EPI_ISL_2363374 | National Virus Reference Laboratory | National Virus Reference Laboratory | Fiona Crispie, Calum Walsh, Matthew McCabe, Zoe Yandle, Charlene Bennet, Gabriel Gonzalez, Michael Carr, Jonathan Dean, Paul Cotter, Cillian F De Gascun |
| EPI_ISL_2373194 | AP SSO | CSIR-Centre for Cellular and Molecular Biology-INSACOG | Blessy B John,Viswagithe S L,Tulasi Nagabandi, Lamuk Zaveri,Ara Sreenivas,Shreekant Verma, Amareshwar Vodapalli ,B Himasri,Valli Nagalakshmi Undamatla,Payel Mukherjee,Sofia Banu,Archana Bharadwaj Siva,Sharath Chandra Thota,Karthik Bharadwaj Tallapaka,Rakesh K Mishra,Divya Tej |

| | | | |
|---|---|---|---|
| | | | Sowpati |
| EPI_ISL_2373258 | CSIR-Centre for Cellular and Molecular Biology | CSIR-Centre for Cellular and Molecular Biology-INSACOG | Lamuk Zaveri,Tulasi Nagabandi, Ara Sreenivas,Shreekant Verma, Amareshwar Vodapalli ,Blessy B John,Viswagithe S L,B Himasri,Valli Nagalakshmi Undamatla,Payel Mukherjee,Sofia Banu,Archana Bharadwaj Siva,Sharath Chandra Thota,Karthik Bharadwaj Tallapaka,Rakesh K Mishra,Divya Tej Sowpati |
| EPI_ISL_2373312 | Mapmygenome | CSIR-Centre for Cellular and Molecular Biology-INSACOG | Ara Sreenivas,Shreekant Verma,Tulasi Nagabandi, Lamuk Zaveri, Amareshwar Vodapalli ,Blessy B John,Viswagithe S L,B Himasri,Valli Nagalakshmi Undamatla,Payel Mukherjee,Sofia Banu,Archana Bharadwaj Siva,Sharath Chandra Thota,Karthik Bharadwaj Tallapaka,Rakesh K Mishra,Divya Tej Sowpati |
| EPI_ISL_2373375 | Mapmygenome | CSIR-Centre for Cellular and Molecular Biology-INSACOG | B Himasri,Valli Nagalakshmi Undamatla,Lamuk Zaveri,Tulasi Nagabandi, Ara Sreenivas,Shreekant Verma, Amareshwar Vodapalli ,Blessy B John,Viswagithe S L,Payel Mukherjee,Sofia Banu,Archana Bharadwaj Siva,Sharath Chandra Thota,Karthik Bharadwaj Tallapaka,Rakesh K Mishra,Divya Tej Sowpati |
| EPI_ISL_2373848 | BIOMNIS LYON | CNR Virus des Infections Respiratoires - France SUD | Antonin Bal, Gregory Destras, Gwendolyne Burfin, Hadrien Regue, Quentin Semanas, Martine Valette, Bruno Lina, Laurence Josset |
| EPI_ISL_2373986, EPI_ISL_2373987, EPI_ISL_2373988, EPI_ISL_2373989, EPI_ISL_2373991, EPI_ISL_2373992, EPI_ISL_2373993, EPI_ISL_2373995 | Laboratoire Virologie Saint Louis APHP | Laboratoire Virologie Saint Louis APHP | Maud Salmona, Marie Laure Chaix, Severine Mercier Delarue, Marie Laure Néré, Linda Feghoul, Jérome Le Goff, Constance Delauguerre, Sophia Achaibou |
| EPI_ISL_2376398 | ULSS 2 Marca Trevigiana | Istituto Zooprofilattico Sperimentale delle Venezie | Adelaide Milani, Alessia Schivo, Annalisa Salviato, Elisa Palumbo, Erika Giorgia Quaranta, Luca Tassoni, Ambra Pastori, Edoardo Giussani, Alice Fusaro, Isabella Monne, Calogero Terregino, Antonia Ricci |
| EPI_ISL_2379487 | Dr S Raju, Director of Public Heath and Preventive Medicine | inStem NCBS - INSACOG | Uma Ramakrishnan Dasaradhi Palakodeti Aswin SaiNarain |
| EPI_ISL_2385971, EPI_ISL_2385974 | PHV-FSS | PHV-FSS | Son Nguyen |
| EPI_ISL_2386845 | Labor Becker & Kollegen (Standort München) | Robert Koch Institute | unknown |
| EPI_ISL_2387394 | MVZ Dr. Eberhard & Partner Dortmund | Robert Koch Institute | unknown |
| EPI_ISL_2388219, EPI_ISL_2388220, EPI_ISL_2388222, EPI_ISL_2388246, EPI_ISL_2388250, EPI_ISL_2388315 | Laborarztpraxis Osnabrück | Robert Koch Institute | unknown |
| EPI_ISL_2396153 | Lighthouse Lab in Alderley Park | Wellcome Sanger Institute for the COVID-19 Genomics UK (COG-UK) Consortium | Jacquelyn Wynn, Mairead Hyland, The Lighthouse Lab in Alderley Park and Alex Alderton, Roberto Amato, Jeffrey Barrett, Sonia Goncalves, Ewan Harrison, David K. Jackson, Ian Johnston, Dominic Kwiatkowski, Cordelia Langford, John Sillitoe on behalf of the Wellcome Sanger Institute COVID-19 Surveillance Team |
| EPI_ISL_2398498 | Laboratory Corporation of America | Centers for Disease Control and Prevention Division of Viral Diseases, Pathogen Discovery | Dakota Howard, Dhwani Batra, Peter W. Cook, Kara Moser, Adrian Paskey, Jason Caravas, Benjamin Rambo-Martin, Shatavia Morrison, Christopher Gulvick, Scott Sammons, Yvette Unoarumhi, Darlene Wagner, Matthew Schmerer, Minoo Agarwal, Eyad Almasri, Debbie Boles, Ayla Burns, Nuthawin Charoensri, Oren Cohen, Susan Countryman, Mary Ann Cristobal, Bobbi Croy, Suzanne Dale, Hrushikesh Deshmukh, Amanda Douglas, Vincent Drouillon, Marcia Eisenberg, Howard Engler, Rama Ghatti, Prashant Gupta, Susan Hicks, Jake Humphrey, Lax Iyer, Lisa Pfefferle, Manoj Jain, Matthew Robinson, Mohan Kolli, Brian Krueger, Tim Kuphal, Stanley Letovsky, Michael Levandoski, Craig Lukasik, Jonathan Meltzer, Brian Norvell, Mindy Nye, Scott Parker, Christos Petropoulos, John Pruitt, Steven Ragan, Scott Ryan, Mike Sapeta, Jana Schroth, Suresh Babu Selvaraju, Goran Stevovic, Amanda Suchanek, Andrea Throop, Lyndon Tilson, Thomas Urban, Joe Voshell, Kimberly Wagner, Jonathan Williams, Mary Williamson, Qian Zeng, Tricia Zwiefelhofer, Clinton R. Paden, Duncan MacCannell |
| EPI_ISL_2399786, EPI_ISL_2399836 | National Virus Reference Laboratory | National Virus Reference Laboratory | Zoe Yandle, Charlene Bennett, Gabriel Gonzalez, Michael Carr, Jonathan Dean, Cillian F De Gascun |
| EPI_ISL_2400460 | Microbiologia e Virologia | Istituto Zooprofilattico Sperimentale delle Venezie | Adelaide Milani, Alessia Schivo, Annalisa Salviato, Elisa Palumbo, Erika Giorgia Quaranta, Luca Tassoni, Ambra Pastori, Edoardo Giussani, Alice Fusaro, Isabella Monne, Calogero Terregino, Antonia Ricci |
| EPI_ISL_2400504 | cerballiance-NOR | Cerba lab | Emmanuel Lecorche, Sabine Trombert-Paolantoni, Gwénolé Prigent, Antoine Prigent, Francois Xavier Le Guillou, Alexandre Leriche, Laura Verdurme, Sylvain Metge, Stéphanie Haïm-Boukobza, Bénédicte Roquebert |
| EPI_ISL_2400558, EPI_ISL_2400636 | Laboratoires d'analyses medicales - Ketterthill | Laboratoire national de sante, Microbiology, Microbial Genomics Platform | Anke Wienecke-Baldacchino, Catherine Ragimbeau,Jessica Tapp, Fatu Djabi, Lise Pignon, Raoul Salmon, Serge Vedy, Caroline Scheiber, Tamir Abdelrahman |
| EPI_ISL_2401539, EPI_ISL_2401597 | BioneXt Lab | Laboratoire national de sante, Microbiology, Microbial Genomics Platform | Anke Wienecke-Baldacchino, Catherine Ragimbeau,Jessica Tapp, Fatu Djabi, Lise Pignon, Raoul Salmon, Thibault Ferrandon, Tamir Abdelrahman |
| EPI_ISL_2403649 | Ministry of Health Turkey | Ministry of Health Turkey | Fatma Bayrakdar, Yasemin Cosgun, Suleyman Yalcin, Gulay Korukluoglu |
| EPI_ISL_2403794 | Dianalabs SA | Genesupport | Tanguy ARAUD |
| EPI_ISL_2406496, EPI_ISL_2406497 | Laboratoire Virologie Saint Louis APHP | Laboratoire Virologie Saint Louis APHP | Maud Salmona, Marie Laure Chaix, Severine Mercier Delarue, Marie Laure Néré, Linda Feghoul, Jérome Le Goff, Constance Delauguerre, Sophia Achaibou |
| EPI_ISL_2420349 | cerballiance-IDF | Cerba lab | Emmanuel Lecorche, Sabine Trombert-Paolantoni, Aude Lessenne, Kader Merah, Thierry Collin, Laura Verdurme, Patrice Herisson, Stéphanie Haïm-Boukobza, Bénédicte Roquebert |
| EPI_ISL_2425247, EPI_ISL_2425251, EPI_ISL_2425252, EPI_ISL_2425330, EPI_ISL_2425331 | KU Leuven, Rega Institute, Clinical and Epidemiological Virology | KU Leuven, Rega Institute, Clinical and Epidemiological Virology | Tony Wawina-Bokalanga, Bert Vanmechelen, Joan Marti-Carerras, Piet Maes |
| EPI_ISL_2425566 | cerballiance-PACA | Cerba lab | Emmanuel Lecorche, Sabine Trombert-Paolantoni, Laurence Prots, Jean-Christophe Roig, Edouard Delaunay, Isabelle Butin, Laura Verdurme, Bruno Roure, Stéphanie Haïm-Boukobza, Bénédicte Roquebert |
| EPI_ISL_2425571 | cerballiance-IDF | Cerba lab | Emmanuel Lecorche, Sabine Trombert-Paolantoni, Aude Lessenne, Kader Merah, Thierry Collin, Laura Verdurme, Patrice Herisson, Stéphanie Haïm-Boukobza, Bénédicte Roquebert |
| EPI_ISL_2425598 | cerballiance-PACA | Cerba lab | Emmanuel Lecorche, Sabine Trombert-Paolantoni, Laurence Prots, Jean-Christophe Roig, Edouard Delaunay, Isabelle Butin, Laura Verdurme, Bruno Roure, Stéphanie Haïm-Boukobza, Bénédicte Roquebert |
| EPI_ISL_2425607 | cerballiance-IDF | Cerba lab | Emmanuel Lecorche, Sabine Trombert-Paolantoni, Aude Lessenne, Kader Merah, Thierry Collin, Laura Verdurme, Patrice Herisson, Stéphanie Haïm-Boukobza, Bénédicte Roquebert |
| EPI_ISL_2429686 | Department of Bacteria, Parasites and Fungi, Statens Serum Institut, Copenhagen, Denmark | Statens Serum Institut Bioinformatics and Microbial Genomics | Danish Covid-19 Genome Consortium |
| EPI_ISL_2445698, EPI_ISL_2445790, EPI_ISL_2445844, EPI_ISL_2445864, EPI_ISL_2445865 | unknown | Instituto Nacional de Saude (INSA) | Borges et al |
| EPI_ISL_2446047, EPI_ISL_2446082, EPI_ISL_2446084 | Pandemic Response Lab - NYC | Pandemic Response Lab, R&D | Henry Lee, Michael Hammerling, Melissa Hopkins, Cybill del Castillo, Shinyoung Clair Kang, William Ward, Pradeep Bugga, Sol Rey, Dylan Law, Katharine Nelson, Haiping Hao, Jon Laurent |
| EPI_ISL_2448424 | ULSS 3 Venezia | Istituto Zooprofilattico Sperimentale delle Venezie | Adelaide Milani, Alessia Schivo, Annalisa Salviato, Elisa Palumbo, Erika Giorgia Quaranta, Luca Tassoni, Ambra Pastori, Edoardo Giussani, Alice Fusaro, Isabella Monne, Calogero Terregino, Antonia Ricci |
| EPI_ISL_2448436, EPI_ISL_2448439, EPI_ISL_2448440, EPI_ISL_2448444 | ULSS 2 Marca Trevigiana | Istituto Zooprofilattico Sperimentale delle Venezie | Adelaide Milani, Alessia Schivo, Annalisa Salviato, Elisa Palumbo, Erika Giorgia Quaranta, Luca Tassoni, Ambra Pastori, Edoardo Giussani, Alice Fusaro, Isabella Monne, Calogero Terregino, Antonia Ricci |

| | | | |
|---|---|---|---|
| EPI_ISL_2448447, EPI_ISL_2448448, EPI_ISL_2448451, EPI_ISL_2448452, EPI_ISL_2448455, EPI_ISL_2448461, EPI_ISL_2448462, EPI_ISL_2448463, EPI_ISL_2448464 | ULSS 6 Euganea | Istituto Zooprofilattico Sperimentale delle Venezie | Adelaide Milani, Alessia Schivo, Annalisa Salviato, Elisa Palumbo, Erika Giorgia Quaranta, Luca Tassoni, Ambra Pastori, Edoardo Giussani, Alice Fusaro, Isabella Monne, Calogero Terregino, Antonia Ricci |
| EPI_ISL_2448467, EPI_ISL_2448468 | ULSS 5 Polesana | Istituto Zooprofilattico Sperimentale delle Venezie | Adelaide Milani, Alessia Schivo, Annalisa Salviato, Elisa Palumbo, Erika Giorgia Quaranta, Luca Tassoni, Ambra Pastori, Edoardo Giussani, Alice Fusaro, Isabella Monne, Calogero Terregino, Antonia Ricci |
| EPI_ISL_2449937 | UniversitŠtsSpital ZŸrich | Institute of Medical Virology | Daniel Ehrsam, Isabel StŸrmer, Catharine Aquino, Joel Wirz, Weihong Qi, Hubert Rehrauer, Verena Kufner, Gabriela Ziltener, Maryam Zaheri, Stefan Schmutz, Annette AudigŽ, Maria GrŸnberg, Kevin Steiner, Jon Huder, Cyril Shah, Riccarda Capaul, Guido Bloemberg, JŸrg Bšni, Michael Huber, Alexandra Trkola |
| EPI_ISL_2450791 | National Public Health Laboratory, Ministry of Health, Ministry of Health, Republic of South Sudan | South Sudan Ministry of Health, WHO South Sudan, MRC/UVRI & LSHTM Uganda Research Unit | Joseph Francis Wamala, Abe G. Abias, Dennis Kenyi Lodiongo, Sudhir Bunga, James Ayei, Lul Lojok Deng, Richard Lino Loro Lako, John Rumunu, Juma John HM, Dan Lule Bugembe, My V.T. Phan, Pontiano Kaleebu, Matthew Cotten |
| EPI_ISL_2455642, EPI_ISL_2455710, EPI_ISL_2455728 | Università Federico II - Dipartimento di scienze mediche traslazionali - Napoli | TIGEM | Antonio Grimaldi Patrizia Annunziata Francesco Panariello Teresa Giuliano Michele Cennamo Valentina Bouche Chiara Colantuono Lucio Di Filippo Mariano Fiorenza Anna Manfredi Marcello Salvi Giuseppe Portella Andrea Ballabio Davide Cacchiarelli |
| EPI_ISL_2462425 | South Eastern Area Laboratory Services (SEALS) | NSW Health Pathology - Institute of Clinical Pathology and Medical Research; Westmead Hospital; University of Sydney | CIDM-PH et al. |
| EPI_ISL_2467865 | Usansolo-Galdakao University Hospital | Cruces University Hospital | Ana Gual-de-Torrella, Izaskun Alejo-Cancho, Mikel Gallego, Ana Belén de la Hoz |
| EPI_ISL_2469753 | Med. Labor Prof. Schenk Dr. Ansorge & Kollegen | Robert Koch Institute | unknown |
| EPI_ISL_2471612 | Eurofins LifeCodexx GmbH | Robert Koch Institute | unknown |
| EPI_ISL_2471762 | Medizinisch-Diagnostisches Labor Kempten allgäulab | Robert Koch Institute | unknown |
| EPI_ISL_2471936 | MDI Limbach Berlin GmbH; MVZ Labor Berlin | Robert Koch Institute | unknown |
| EPI_ISL_2476255 | Dutch COVID-19 response team | National Institute for Public Health and the Environment (RIVM) | Adam Meijer, Harry Vennema, Dirk Eggink, Jeroen Cremer, Sharon van den Brink, Bas van der Veer, AnneMarie van den Brandt, Lisa Wijsman, Kim Freriks, Ryanne Jaarsma, Euníce Then, Lynn Aarts, Sanne Bos, Melissa van Tuil, Linda van de Nes, Florian Zwagemaker, Dennis Schmitz, Annelies Kroneman, Karim Hajji, Chantal Reusken, on behalf of the national COVID-19 response team |
| EPI_ISL_2477718, EPI_ISL_2477730, EPI_ISL_2478300, EPI_ISL_2478308, EPI_ISL_2478332, EPI_ISL_2478338, EPI_ISL_2479079, EPI_ISL_2479082, EPI_ISL_2479089, EPI_ISL_2479140, EPI_ISL_2479141, EPI_ISL_2479147, EPI_ISL_2479165, EPI_ISL_2479227, EPI_ISL_2479229 | | | |
| see above | Edmonton Provincial Lab | Public Health Agency of Canada (PHAC) National Microbiology Laboratory | Buss, E, Croxen M, Deo A, Dieu P, Gill K, Ferrato C, Khan F, Koleva P, Li V, Lloyd C, Lynch T, Ma R, Murphy S, Pabbaraju K, Shokoples S, Tipples G, Thayer J, Whitehouse M, Wong A, Yu C, Zelyas N |
| EPI_ISL_2483575, EPI_ISL_2483576 | SIESP DIPARTIMENTO DI PREVENZIONE TERAMO | Istituto Zooprofilattico Sperimentale dell'Abruzzo e Molise "G. Caporale" | Lorusso A, Marcacci M, Di Domenico M, Ancora M, Curini V, Di Lollo Valeria, Mangone I, Rinaldi A, Delli Compagni E, Scialabba S, Caporale M, Di Pasquale A, Cammà C, Puglia I, Calistri P, Savini G |
| EPI_ISL_2489270 | Aegis Sciences Corporation | Centers for Disease Control and Prevention Division of Viral Diseases, Pathogen Discovery | Dakota Howard, Dhwani Batra, Peter W. Cook, Kara Moser, Adrian Paskey, Jason Caravas, Benjamin Rambo-Martin, Shatavia Morrison, Christopher Gulvick, Scott Sammons, Yvette Unoarumhi, Darlene Wagner, Matthew Schmerer, Cyndi Clark, Patrick Campbell, Rob Case, Vikramsinha Ghorpade, Holly Houdeshell, Ola Kvalvaag, Dillon Nall, Ethan Sanders, Alec Vest, Shaun Westlund, Matthew Hardison, Clinton R. Paden, Duncan MacCannell |
| EPI_ISL_2490026, EPI_ISL_2490036 | Seattle Flu Study | Seattle Flu Study | Deborah A. Nickerson, Chris D. Frazar, Jover Lee, Benjamin Pelle, Erica Ryke, Matthew Richardson, Amanda Adler, Elisabeth Brandstetter, Peter D. Han, Kairsten Fay, Misja Ilcisin, Kirsten Lacombe, Thomas R. Sibley, Melissa Truong, Caitlin R. Wolf, Karen Cowgill, Stephanie Schrag, Jeff Duchin, Michael Boeckh, Janet A. Englund, Michael Famulare, Barry R. Lutz, Mark J. Rieder, Lea M. Starita, Matthew Thompson, Helen Y. Chu, Trevor Bedford, Jay Shendure |
| EPI_ISL_2495344 | HOSPITAL GENERAL UNIVERSITARIO DE GUADALAJARA | Instituto de Salud Carlos III | Iglesias-Caballero, M. Sandonís,V. Vázquez-Morón, S. Camarero, S. Pozo, F. Casas, I. Jiménez, P. Zaballos, A. Monzón, S. Varona, S. Cuesta, I.GONZALEZ PRAETORIUS, ALEJANDRO |
| EPI_ISL_2500436 | CT Department of Public Health | CT Department of Public Health | Claire_Pearson,Tu_N_Nguyen |
| EPI_ISL_2533640, EPI_ISL_2533669 | National Virus Reference Laboratory | National Virus Reference Laboratory | Zoe Yandle, Charlene Bennett, Gabriel Gonzalez, Michael Carr, Jonathan Dean, Cillian F De Gascun |
| EPI_ISL_2536216 | Laboratorio HUB -Azienda Ospedaliero Universitaria - AOU - Cagliari | Laboratorio SPOKE Biologia Molecolare -Azienda Ospedaliero Universitaria - AOU - Cagliari | Germano Orrù, Sara Fais, Valentina Medda, Alessandra Scano, Miriam Loddo, Riccardo Cappai, Ferdinando Coghe |
| EPI_ISL_2536480 | Incyte Diagnostics Spokane | Seattle Flu Study | Deborah A. Nickerson, Chris D. Frazar, Jover Lee, Benjamin Pelle, Erica Ryke, Matthew Richardson, Amanda Adler, Elisabeth Brandstetter, Peter D. Han, Kairsten Fay, Misja Ilcisin, Kirsten Lacombe, Thomas R. Sibley, Melissa Truong, Caitlin R. Wolf, Romesh Gautom, Geoff Melly, Brian Hiatt, Philip Dykema, Scott Lindquist, Michael Boeckh, Janet A. Englund, Michael Famulare, Barry R. Lutz, Mark J. Rieder, Lea M. Starita, Matthew Thompson, Helen Y. Chu, Jay Shendure, Trevor Bedford |
| EPI_ISL_2546745 | ICMR-National Institute of Virology - INSACOG | NIV Influenza | Dr. Varsha Potdar and NIC Team |
| EPI_ISL_2548609 | INSACOG-WB | National Institute of Biomedical Genomics - INSACOG | Arindam Maitra, Bhaswati Bandyopadhyay, Nidhan Kumar Biswas, Tamal Ghosh, Sreedhar Chinnaswamy, Ajay Chakraborti, Saumitra Das |
| EPI_ISL_2549566, EPI_ISL_2549567, EPI_ISL_2549568, EPI_ISL_2549569, EPI_ISL_2549570, EPI_ISL_2549571, EPI_ISL_2549572, EPI_ISL_2549573, EPI_ISL_2549574, EPI_ISL_2549575 | Virology, Istituto Zooprofilattico Sperimentale dell'Abruzzo e del Molise 'G. Caporale' | Virology, Istituto Zooprofilattico Sperimentale dell'Abruzzo e del Molise 'G. Caporale' | Delli Compagni,E., Mangone,I., Scialabba,S., Caporale,M., Lorusso,A. |
| EPI_ISL_2557459 | MOH - Jaber Al-Ahmad Hospital (Innovation Research Laboratory) | MOH - Jaber Al-Ahmad Hospital (Innovation Research Laboratory) | Salman Al-Sabah , Mohammad Alghounaim |
| EPI_ISL_2558084 | LBM ALPHABIO, Marseille | LBM ALPHABIO, Marseille | Vincent GARCIA |
| EPI_ISL_2561989 | cerballiance-IDF | Cerba lab | Emmanuel Lecorche, Sabine Trombert-Paolantoni, Laura Verdurme, Stéphanie Haïm-Boukobza, Bénédicte Roquebert |
| EPI_ISL_2562007 | cerballiance-cote d'azur | Cerba lab | Emmanuel Lecorche, Sabine Trombert-Paolantoni, Laura Verdurme, Stéphanie Haïm-Boukobza, Bénédicte Roquebert |
| EPI_ISL_2597134, EPI_ISL_2597152, EPI_ISL_2597160, EPI_ISL_2597189 | MEPHI, Aix Marseille University | MEPHI, Aix Marseille University | Anthony LEVASSEUR |
| EPI_ISL_2604147 | Azienda Ospedaliera Terni | Istituto Zooprofilattico Sperimentale dell'Abruzzo e Molise "G. Caporale" | Palumbo M, Scaccetti A, Lorusso A, Marcacci M, Di Domenico M, Ancora M, Curini V, Mangone I, Rinaldi A, Delli Compagni E, Scialabba S, Di Pasquale A, Cammà C, Puglia I, Calistri P, Savini G |
| EPI_ISL_2615960 | Berkeley Medical Center | WVU and Marshall University Combined Genomics Core Facilities | James Denvir, Peter Stoilov, Peter Perrotta, Wesley Kimble, Ryan Percifield |
| EPI_ISL_2615961 | QLabs | WVU and Marshall University Combined Genomics Core Facilities | James Denvir, Peter Stoilov, Peter Perrotta, Wesley Kimble, Ryan Percifield |
| EPI_ISL_2615962 | WV Labs | WVU and Marshall University Combined Genomics Core Facilities | James Denvir, Peter Stoilov, Peter Perrotta, Wesley Kimble, Ryan Percifield |
| EPI_ISL_2615963, EPI_ISL_2615964, EPI_ISL_2615965 | Berkeley Medical Center | WVU and Marshall University Combined Genomics Core Facilities | James Denvir, Peter Stoilov, Peter Perrotta, Wesley Kimble, Ryan Percifield |

| | | | |
|---|---|---|---|
| EPI_ISL_2615966 | WV Labs | WVU and Marshall University Combined Genomics Core Facilities | James Denvir, Peter Stoilov, Peter Perrotta, Wesley Kimble, Ryan Percifield |
| EPI_ISL_2615967 | J.W. Ruby Memorial Hospital | WVU and Marshall University Combined Genomics Core Facilities | James Denvir, Peter Stoilov, Peter Perrotta, Wesley Kimble, Ryan Percifield |
| EPI_ISL_2615968, EPI_ISL_2615969, EPI_ISL_2615972 | Berkeley Medical Center | WVU and Marshall University Combined Genomics Core Facilities | James Denvir, Peter Stoilov, Peter Perrotta, Wesley Kimble, Ryan Percifield |
| EPI_ISL_2617525 | Lab Genomik Solidaritas Indonesia | National Institute of Health Research and Development | Vivi Setiawaty, Hana Apsari Pawestri, Hartanti Dian Ikawati, Subangkit, Kartika Dewi Puspa, Arie Ardiansyah Nugraha, Triyani Soekarso, Krisna Pangesti,Meutia Kumaheri, Nelly Puspandari |
| EPI_ISL_2620734 | Functional Genomics and Proteomics Laboratory (FGPL), Department of Genetic Engineering and Biotechnology, University of Chittagong | Virology Laboratory, International Centre for Diarrhoeal Disease Research, Bangladesh (ICDDR,B) | Dr. Mohammad Al-Forkan, Dr. Sujat Paul, Dr. Laila Khaleda, Mojnu Miah, Rashedul Hasan, Md. Jibran Alam, Amit Datta, Rahee Hasan Chowdhury, Shuva Das, Inzamamul Ismail Shawon, Md. Abdur Rahman, Miftah Mushfique, Amlan Bhattacharjee, Dr. Mustafizur Rahman, Dr. Mohammad Enayet Hossain |
| EPI_ISL_2620903, EPI_ISL_2620905 | Fondazione IRCCS Ca' Granda Ospedale Maggiore Policlinico | Fondazione IRCCS Ca' Granda Ospedale Maggiore Policlinico | Ferruccio Ceriotti, Sara Uceda Renteria |
| EPI_ISL_2622164, EPI_ISL_2622165, EPI_ISL_2622166, EPI_ISL_2622167, EPI_ISL_2622169, EPI_ISL_2622171, EPI_ISL_2622172, EPI_ISL_2622174, EPI_ISL_2622175 | Azienda Ospedaliera Pugliese Ciaccio di Catanzaro SOC Microbiologia e Virologia | Azienda Ospedaliera Pugliese Ciaccio di Catanzaro SOC Microbiologia e Virologia | Rossana Tallerico Federica Pasceri Marco De Fazio Ilenia Talotta Giuseppina Panduri Pasquale Minchella |
| EPI_ISL_2628710 | UNILABS | Instituto Nacional de Saude (INSA) | Borges et al |
| EPI_ISL_2631766, EPI_ISL_2631775, EPI_ISL_2631779 | MVZ für Laboratoriumsmedizin und Mikrobiologie Koblenz-Mittelrhein (Labor Koblenz) | Robert Koch Institute | unknown |
| EPI_ISL_2631829, EPI_ISL_2631835 | Labor Dr. Wisplinghoff - Köln | Robert Koch Institute | unknown |
| EPI_ISL_2632026 | Medizinisches Labor Wahl Lüdenscheid | Robert Koch Institute | unknown |
| EPI_ISL_2632037, EPI_ISL_2632085 | Laborarztpraxis Osnabrück | Robert Koch Institute | unknown |
| EPI_ISL_2632102, EPI_ISL_2633436 | Medizinisches Labor Wahl Lüdenscheid | Robert Koch Institute | unknown |
| EPI_ISL_2633440, EPI_ISL_2633443 | Laborarztpraxis Osnabrück | Robert Koch Institute | unknown |
| EPI_ISL_2634508 | Eurofins LifeCodexx GmbH | Robert Koch Institute | unknown |
| EPI_ISL_2635787 | Med. Labor Prof. Schenk Dr. Ansorge & Kollegen | Robert Koch Institute | unknown |
| EPI_ISL_2635841 | MVZ für Laboratoriumsmedizin und Mikrobiologie Koblenz-Mittelrhein (Labor Koblenz) | Robert Koch Institute | unknown |
| EPI_ISL_2636094 | Medizinisch-Diagnostisches Labor Kempten allgäulab | Robert Koch Institute | unknown |
| EPI_ISL_2636153 | Corona-Testzentrum ifp Institut für Produktqualität GmbH | Robert Koch Institute | unknown |
| EPI_ISL_2636491 | MVZ für Laboratoriumsmedizin und Mikrobiologie Koblenz-Mittelrhein (Labor Koblenz) | Robert Koch Institute | unknown |
| EPI_ISL_2636898 | MVZ Dr. Eberhard & Partner Dortmund | Robert Koch Institute | unknown |
| EPI_ISL_2636900 | Laborarztpraxis Osnabrück | Robert Koch Institute | unknown |
| EPI_ISL_2646019, EPI_ISL_2646083 | UNILABS | Instituto Nacional de Saude (INSA) | Borges et al |
| EPI_ISL_2649124, EPI_ISL_2649497 | SARS-CoV-2 Sequencing Castilla y Leon-Spain Consortium | SARS-CoV-2 Sequencing Castilla y Leon-Spain Consortium | Antonio Orduña-Domingo, Marta Hernandez, David Abad, Marta Dominguez-Gil, Silvia Rojo, Laura Sánchez de Prada, Sonsoles Garcinuño Pérez, Carmen Aldea-Mansilla, Mª Fe Brezmes-Valdivieso, Gregoria Megías Lobón, María Antonia García Castro, Carmen Gimeno Crespo, Noelia Arenal Andrés, Carlos Fuster Foz, M. Isabel Fernandez-Natal, Jose María Eiros Bouza |
| EPI_ISL_2650188, EPI_ISL_2650252, EPI_ISL_2650289 | Microbiology Department, Laboratori Clínic Metropolitana Nord. Hospital Universitari Germans Trias i Pujol | Can Ruti SARS-CoV-2 Sequencing Hub (HUGTiP/IrsiCaixa/IGTP) | Marc Noguera-Julian, Pilar Armengol, Ignacio Blanco, Antoni E Bordoy, Francesc Catala-Moll, Pere-Joan Cardona, Maria Casadellà, Cristina Casañ, Gemma Clara, Bonaventura Clotet, Cristina Esteban, Montserrat Giménez, Mercedes Guerrero, Anna Not, Roger Paredes, Mariona Parera, Verónica Saludes, Alba Sánchez, and Elisa Martró on behalf of the Can Ruti SARS-CoV-2 Sequencing Hub |
| EPI_ISL_2652057 | Hospital Universitari Joan XXIII | Hospital Universitari Joan XXIII | Gemma Recio, Cristina Gutiérrez, Clara Benavent, Carla Martín, Margarida Terrón, Ester Picó, Natalia Bastón |
| EPI_ISL_2654226, EPI_ISL_2654944, EPI_ISL_2654990, EPI_ISL_2655020, EPI_ISL_2655731, EPI_ISL_2655915, EPI_ISL_2656238, EPI_ISL_2656241, EPI_ISL_2656310 | BCCDC Public Health Laboratory | BCCDC Public Health Laboratory | Prystajecky Natalie, Linda Hoang, Dan Fornika, John Tyson, Shannon Russell, Kim Macdonald, Kimia Kamelian, Ana Pacagnella, Corrinne Ng, Loretta Janz, Robert Azana, Mel Krajden |
| EPI_ISL_2661118 | Mapmygenome | CSIR-Centre for Cellular and Molecular Biology-INSACOG | Tulasi Nagabandi,Lamuk Zaveri,Payel Mukherjee,Onkar Kulkarni, Sofia Banu, Ara Sreenivas,Shreekant Verma, Amareshwar Vodapalli ,B Himasri,Valli Nagalakshmi Undamatla, Priya Nurkurthy,Sumedha Avadhanula, Vidhyadhari Methuku, Archana Bharadwaj Siva, Karthik Bharadwaj Tallapaka,Rakesh K Mishra,Divya Tej Sowpati |
| EPI_ISL_2661132, EPI_ISL_2661141 | Mapmygenome | CSIR-Centre for Cellular and Molecular Biology-INSACOG | Ara Sreenivas,Shreekant Verma, Amareshwar Vodapalli ,B Himasri,Valli Nagalakshmi Undamatla,Payel Mukherjee, Lamuk Zaveri, Onkar Kulkarni,Tulasi Nagabandi, Sofia Banu, ,Priya Nurkurthy, Vidhyadhari Methuku, Sumedha Avadhanula, Archana Bharadwaj Siva, Karthik Bharadwaj Tallapaka,Rakesh K Mishra,Divya Tej Sowpati |
| EPI_ISL_2672210, EPI_ISL_2672216 | INSP, Bamako | Malaria Research and Training Center-Parasito | Antoine Dara, Ibrehima Guindo, Amadou Daou, Demba Koita, Abdoulaye Djimde |
| EPI_ISL_2678695 | MVZ Labor Dr. Limbach & Kollegen GbR | Robert Koch Institute | unknown |
| EPI_ISL_2681581 | Microbiology Department, Laboratori Clínic Metropolitana Nord. Hospital Universitari Germans Trias i Pujol | Can Ruti SARS-CoV-2 Sequencing Hub (HUGTiP/IrsiCaixa/IGTP) | Marc Noguera-Julian, Pilar Armengol, Ignacio Blanco, Antoni E Bordoy, Francesc Catala-Moll, Pere-Joan Cardona, Maria Casadellà, Cristina Casañ, Gemma Clara, Bonaventura Clotet, Cristina Esteban, Montserrat Giménez, Mercedes Guerrero, Anna Not, Roger Paredes, Mariona Parera, Verónica Saludes, Alba Sánchez, and Elisa Martró on behalf of the Can Ruti SARS-CoV-2 Sequencing Hub |
| EPI_ISL_2690456, EPI_ISL_2690457, EPI_ISL_2690459 | MRC/UVRI & LSHTM Uganda Research Unit, Central Public Health Laboratories, Rakai Health Sciences Program | MRC/UVRI & LSHTM Uganda Research Unit, Central Public Health Laboratories | Matthew Cotten, Dan Lule Bugembe, My V.T. Phan, Pontiano Kaleebu,Isaac Sseeewanyana, Patrick Semanda, Susan Nabadda, Hellen Nansumba |
| EPI_ISL_2690465, EPI_ISL_2690466, EPI_ISL_2690467, EPI_ISL_2690468, EPI_ISL_2690469, EPI_ISL_2690470, EPI_ISL_2690471, EPI_ISL_2690472, EPI_ISL_2690473, EPI_ISL_2690474, EPI_ISL_2690475, EPI_ISL_2690476, EPI_ISL_2690477, EPI_ISL_2690478 | | | |
| see above | MRC/UVRI & LSHTM Uganda Research Unit, Central Public Health Laboratories, Rakai Health Sciences Program | MRC/UVRI & LSHTM Uganda Research Unit, Rakai Health Sciences Program | Matthew Cotten, Dan Lule Bugembe, My V.T. Phan, Pontiano Kaleebu, Charles Ssuuna, Ronald Moses Galiwango, Steven J Reynolds |
| EPI_ISL_2691703 | Analytica | Institute of Medical Virology | Verena Kufner, Gabriela Ziltener, Maryam Zaheri, Stefan Schmutz, Annette Audigé, Maria Grünberg, Kevin Steiner, Jon Huder, Cyril Shah, Riccarda Capaul, Guido Bloemberg, Jürg Böni, Michael Huber, Alexandra Trkola |
| EPI_ISL_2694831 | Labor Berlin Charite Vivantes GmbH / Institut fur Virologie | Charite Universitatsmedizin Berlin, Institut fur Virologie/Labor Berlin | Peter Menzel, Christine Stephan, Rolf Schwarzer, Victor M Corman, Barbara Muhlemann, Terry Jones, Christian Drosten |
| EPI_ISL_2695344 | CHEDV | Instituto Nacional de Saude (INSA) | Borges et al |

| | | | |
|---|---|---|---|
| EPI_ISL_2718650, EPI_ISL_2718858, EPI_ISL_2719916, EPI_ISL_2720267, EPI_ISL_2720309, EPI_ISL_2720322 | BCCDC Public Health Laboratory | BCCDC Public Health Laboratory | Prystajecky Natalie, Linda Hoang, Dan Fornika, John Tyson, Shannon Russell, Kim Macdonald, Kimia Kamelian, Ana Pacagnella, Corrinne Ng, Loretta Janz, Robert Azana, Mel Krajden |
| EPI_ISL_2720402 | A.S.L. VCO | Fondazione del Piemonte per l'Oncologia IRCCS | Antonino Sottile, Silvia Brossa, Paola Marino, Giorgio Giardina |
| EPI_ISL_2720404 | A.S.L. VERCELLI | Fondazione del Piemonte per l'Oncologia IRCCS | Antonino Sottile, Silvia Brossa, Paola Marino, Giorgio Giardina |
| EPI_ISL_2720511 | A.S.L. TO4 | Fondazione del Piemonte per l'Oncologia IRCCS | Antonino Sottile, Silvia Brossa, Paola Marino, Giorgio Giardina |
| EPI_ISL_2720584, EPI_ISL_2720585, EPI_ISL_2720807, EPI_ISL_2721136 | BCCDC Public Health Laboratory | BCCDC Public Health Laboratory | Prystajecky Natalie, Linda Hoang, Dan Fornika, John Tyson, Shannon Russell, Kim Macdonald, Kimia Kamelian, Ana Pacagnella, Corrinne Ng, Loretta Janz, Robert Azana, Mel Krajden |
| EPI_ISL_2721308 | A.S.L. TO4 | Fondazione del Piemonte per l'Oncologia IRCCS | Antonino Sottile, Silvia Brossa, Paola Marino, Giorgio Giardina |
| EPI_ISL_2721422, EPI_ISL_2721426, EPI_ISL_2721453, EPI_ISL_2721840, EPI_ISL_2721869 | BCCDC Public Health Laboratory | BCCDC Public Health Laboratory | Prystajecky Natalie, Linda Hoang, Dan Fornika, John Tyson, Shannon Russell, Kim Macdonald, Kimia Kamelian, Ana Pacagnella, Corrinne Ng, Loretta Janz, Robert Azana, Mel Krajden |
| EPI_ISL_2725219, EPI_ISL_2725247, EPI_ISL_2725296 | Lab. Microbiologia e Virologia Cotugno A.O. dei Colli - Istituto Zooprofilattico Sperimentale del Mezzogiorno | Telethon Institute of Genetics and Medicine (TIGEM) | Antonio Grimaldi Patrizia Annunziata Francesco Panariello Biancamaria Pierri Claudia Tiberio Teresa Giuliano Valentina Bouche Chiara Colantuono Maria Concetta Cuomo Denise Di Concilio Lucio Di Filippo Anna Manfredi Marcello Salvi Antonio Limone Luigi Atripaldi Pellegrino Cerino Andrea Ballabio Davide Cacchiarelli |
| EPI_ISL_2726488 | KU Leuven, Rega Institute, Clinical and Epidemiological Virology | KU Leuven, Rega Institute, Clinical and Epidemiological Virology | Tony Wawina-Bokalanga, Bert Vanmechelen, Joan Marti-Carerras, Piet Maes |
| EPI_ISL_2727349 | ZARV/NHLS, Department Medical Virology, University of Pretoria | KRISP, KZn Research Innovation and Sequencing Platform | Amy Strydom, Adriano Mendes, Micheala Davids, Sim Mayaphi and Marietjie Venter, Giandhari J, Pillay S, Naidoo Y, Ramphal U, Lessells R, Emmanuel SJ, Tegally H, Wilkinson E, de Oliveira T |
| EPI_ISL_2747989, EPI_ISL_2748655 | Ohio Department of Health Laboratory | Ohio Department of Health Laboratory | Holmes, Jennifer; Eric Brandt, Keoni Omura, Glen McGillivary, Caitlin McDonnell, Jade Mowery, Stephanie Mccracken, Tyler Payne, Kirtana Ramadugu, Erica Leasure, Brent Lee, Kelsey Florek, Heather Blankenship, Allison Black, Quanta Brown, and Tammy Bannerman |
| EPI_ISL_2753948 | NYU Langone Health | Departments of Pathology and Medicine, New York University School of Medicine | Adriana Heguy, Dacia Dimartino, Emily Guzman, Christian Marier, Peter Meyn, Sitharam Ramaswami, Gael Westby, Paul Zappile, Yutong Zhang, Paolo Cotzia, Guiqing Wang |
| EPI_ISL_2758356 | QLabs | WVU and Marshall University Combined Genomics Core Facilities | James Denvir, Peter Stoilov, Peter Perrotta, Wesley Kimble, Ryan Percifield |
| EPI_ISL_2762285 | MVZ Labor Dr. Fenner und Kollegen (Standort Hamburg) | Robert Koch Institute | unknown |
| EPI_ISL_2771492 | Ohio Department of Health Laboratory | Ohio Department of Health Laboratory | Holmes, Jennifer; Eric Brandt, Keoni Omura, Glen McGillivary, Caitlin McDonnell, Jade Mowery, Stephanie Mccracken, Tyler Payne, Kirtana Ramadugu, Erica Leasure, Brent Lee, Kelsey Florek, Heather Blankenship, Allison Black, Quanta Brown, and Tammy Bannerman |

### supp. Table S8: Iota

We gratefully acknowledge the following Authors from the Originating laboratories responsible for obtaining the specimens, as well as the Submitting laboratories where the genome data were generated and shared via GISAID, on which this research is based.

All Submitters of data may be contacted directly via www.gisaid.org

Authors are sorted alphabetically.

| Accession ID | Originating Laboratory | Submitting Laboratory | Authors |
|---|---|---|---|
| EPI_ISL_2349561, EPI_ISL_2349577, EPI_ISL_2349580, EPI_ISL_2349597, EPI_ISL_2349608, EPI_ISL_2349652, EPI_ISL_2349653, EPI_ISL_2349682 | Pandemic Response Lab - NYC | Pandemic Response Lab, R&D | Henry Lee, Michael Hammerling, Melissa Hopkins, Cybill del Castillo, Shinyoung Clair Kang, William Ward, Pradeep Bugga, Sol Rey, Dylan Law, Katharine Nelson, Haiping Hao, Jon Laurent |
| EPI_ISL_2362005 | HOSPITAL UNIVERSITARIO SON ESPASES | HOSPITAL UNIVERSITARIO SON ESPASES | Carla López-Causapé, Pablo Fraile-Ribot, Antonio Oliver, SeqCovid |
| EPI_ISL_2370368, EPI_ISL_2370374, EPI_ISL_2370384, EPI_ISL_2370566, EPI_ISL_2370606, EPI_ISL_2370608, EPI_ISL_2370630, EPI_ISL_2370706 | Aegis Sciences Corporation | Centers for Disease Control and Prevention Division of Viral Diseases, Pathogen Discovery | Dakota Howard, Dhwani Batra, Peter W. Cook, Kara Moser, Adrian Paskey, Jason Caravas, Benjamin Rambo-Martin, Shatavia Morrison, Christopher Gulvick, Scott Sammons, Yvette Unoarumhi, Darlene Wagner, Matthew Schmerer, Cyndi Clark, Patrick Campbell, Rob Case, Vikramsinha Ghorpade, Holly Houdeshell, Ola Kvalvaag, Dillon Nall, Ethan Sanders, Alec Vest, Shaun Westlund, Matthew Hardison, Clinton R. Paden, Duncan MacCannell |
| EPI_ISL_2371541, EPI_ISL_2371551, EPI_ISL_2371565 | Fulgent Genetics | Centers for Disease Control and Prevention Division of Viral Diseases, Pathogen Discovery | Dakota Howard, Dhwani Batra, Peter W. Cook, Kara Moser, Adrian Paskey, Jason Caravas, Benjamin Rambo-Martin, Shatavia Morrison, Christopher Gulvick, Scott Sammons, Yvette Unoarumhi, Darlene Wagner, Matthew Schmerer, Harry Gao, Mickey Li, John Gao, Joseph Fierro, Benafsh Sapra, Becky Tsai, Yan Meng, Doreen Ng, James Xie, Clinton R. Paden, Duncan MacCannell |
| EPI_ISL_2380811, EPI_ISL_2380814 | Clinical Molecular Microbiology Laboratory, UNC Hospitals | Jeremy Wang | Jeremy Wang, Alexander Rubinsteyn, Colleen Rice, Jason Smedberg, Shawn Hawken, Melissa Miller, Corbin Jones, Robert Hagan |
| EPI_ISL_2398063, EPI_ISL_2399345, EPI_ISL_2399360, EPI_ISL_2399361, EPI_ISL_2399362, EPI_ISL_2399369, EPI_ISL_2399370, EPI_ISL_2399371, EPI_ISL_2399382, EPI_ISL_2399393, EPI_ISL_2399396, EPI_ISL_2399409 | | | |
| see above | Laboratory Corporation of America | Centers for Disease Control and Prevention Division of Viral Diseases, Pathogen Discovery | Dakota Howard, Dhwani Batra, Peter W. Cook, Kara Moser, Adrian Paskey, Jason Caravas, Benjamin Rambo-Martin, Shatavia Morrison, Christopher Gulvick, Scott Sammons, Yvette Unoarumhi, Darlene Wagner, Matthew Schmerer, Minoo Agarwal, Eyad Almasri, Debbie Boles, Ayla Burns, Nuthawin Charoensri, Oren Cohen, Susan Countryman, Mary Ann Cristobal, Bobbi Croy, Suzanne Dale, Hrushikesh Deshmukh, Amanda Douglas, Vincent Drouillon, Marcia Eisenberg, Howard Engler, Rama Ghatti, Prashant Gupta, Susan Hicks, Jake Humphrey, Lax Iyer, Lisa Pfefferle, Manoj Jain, Matthew Robinson, Mohan Kolli, Brian Krueger, Tim Kuphal, Stanley Letovsky, Michael Levandoski, Craig Lukasik, Jonathan Meltzer, Brian Norvell, Mindy Nye, Scott Parker, Christos Petropoulos, John Pruitt, Steven Ragan, Scott Ryan, Mike Sapeta, Jana Schroth, Suresh Babu Selvaraju, Goran Stevovic, Amanda Suchanek, Andrea Throop, Lyndon Tilson, Thomas Urban, Joe Voshell, Kimberly Wagner, Jonathan Williams, Mary Williamson, Qian Zeng, Tricia Zwiefelhofer, Clinton R. Paden, Duncan MacCannell |
| EPI_ISL_2400241 | Wichita State University - Molecular Diagnostics Lab | Kansas Health and Environmental Lab | Mike Grose, Katherine Wiggins, Jonathan Barnell, Ben Olsen, and Phil Adam |
| EPI_ISL_2405148, EPI_ISL_2405149 | INTERLAB | Omics Sciences Laboratory | Derly Andrade Molina, Gabriel Morey León, Darlyn Amaya, Rubén Armas González, Juan Carlos Fernández Cadena |
| EPI_ISL_2405151 | Hospital Luis Vernaza | Omics Sciences Laboratory | Derly Andrade Molina, Gabriel Morey León, Darlyn Amaya, Rubén Armas González, Juan Carlos Fernández Cadena |
| EPI_ISL_2405153 | Omics Sciences Laboratory | Omics Sciences Laboratory | Derly Andrade Molina, Gabriel Morey León, Darlyn Amaya, Rubén Armas González, Juan Carlos Fernández Cadena |
| EPI_ISL_2405156, EPI_ISL_2405164, EPI_ISL_2405166 | Hospital Luis Vernaza | Omics Sciences Laboratory | Derly Andrade Molina, Gabriel Morey León, Darlyn Amaya, Rubén Armas González, Juan Carlos Fernández Cadena |
| EPI_ISL_2405548, EPI_ISL_2405595, EPI_ISL_2405629 | Dutch COVID-19 response team | National Institute for Public Health and the Environment (RIVM) | Adam Meijer, Harry Vennema, Dirk Eggink, Jeroen Cremer, Sharon van den Brink, Bas van der Veer, AnneMarie van den Brandt, Lisa Wijsman, Kim Freriks, Ryanne Jaarsma, Euníce Then, Lynn Aarts, Sanne Bos, Melissa van Tuil, Linda van de Nes, Sjoerd Kuiling, James Groot, Florian Zwagemaker, Dennis Schmitz, Annelies Kroneman, Karim Hajji, Chantal Reusken, on behalf of the national COVID-19 response team |
| EPI_ISL_2420987 | Montana Public Health Laboratory | Montana Public Health Laboratory | Joy Ritter, Michelle Mozer, Carrie Biskupiak, Deborah Gibson, Michael Dills |
| EPI_ISL_2422013, EPI_ISL_2422025, EPI_ISL_2422044, EPI_ISL_2422109, EPI_ISL_2422118 | Aegis Sciences Corporation | Centers for Disease Control and Prevention Division of Viral Diseases, Pathogen Discovery | Dakota Howard, Dhwani Batra, Peter W. Cook, Kara Moser, Adrian Paskey, Jason Caravas, Benjamin Rambo-Martin, Shatavia Morrison, Christopher Gulvick, Scott Sammons, Yvette Unoarumhi, Darlene Wagner, Matthew Schmerer, Cyndi Clark, Patrick Campbell, Rob Case, Vikramsinha Ghorpade, Holly Houdeshell, Ola Kvalvaag, Dillon Nall, Ethan Sanders, Alec Vest, Shaun Westlund, Matthew Hardison, Clinton R. Paden, Duncan MacCannell |
| EPI_ISL_2424257, EPI_ISL_2424261 | Oregon State Public Health Laboratory | Oregon State Public Health Laboratory | Rafia Razzaque, Eugene Yeboah, Vanda Makris, Laura Tsaknaridis, John Fontana and Shane Sevey |
| EPI_ISL_2425398, EPI_ISL_2425399 | Minnesota Department of Health, Public Health Laboratory | Minnesota Department of Health, Public Health Laboratory | Alexandra Lorentz, Jacob Garfin, Matt Plumb, and Xiong Wang |
| EPI_ISL_2427480 | NYU Langone Health | Departments of Pathology and Medicine, New York University School of Medicine | Adriana Heguy, Dacia Dimartino, Emily Guzman, Christian Marier, Peter Meyn, Sitharam Ramaswami, Gael Westby, Paul Zappile, Yutong Zhang, Paolo Cotzia, Guiqing Wang |
| EPI_ISL_2432609, EPI_ISL_2432626, EPI_ISL_2432648 | UW Virology Lab | UW Virology Lab | Pavitra Roychoudhury, Hong Xie, Lasata Shrestha, Tien V. Nguyen, Shah Mohamed Bakhash, Michelle Lin, Noah R. Baker, Ricardo Perez, Sean Ellis, Nathan Breit, Robert J. Livingston, Meei-Li Huang, Keith R Jerome, Patrick Mathias, Alexander Greninger |
| EPI_ISL_2438698, EPI_ISL_2438717, EPI_ISL_2438725 | HOSPITAL UNIVERSITARIO SON ESPASES | HOSPITAL UNIVERSITARIO SON ESPASES | Servicio de Microbiología, Hospital Universitario Son Espases |
| EPI_ISL_2439546, EPI_ISL_2439703, EPI_ISL_2440635, EPI_ISL_2440726, EPI_ISL_2440792, EPI_ISL_2440810, EPI_ISL_2440818, EPI_ISL_2440826, EPI_ISL_2440852, EPI_ISL_2440872, EPI_ISL_2440887, EPI_ISL_2440890 | | | |
| see above | Quest Diagnostics Incorporated | Centers for Disease Control and Prevention Division of Viral Diseases, Pathogen Discovery | Dakota Howard, Dhwani Batra, Peter W. Cook, Kara Moser, Adrian Paskey, Jason Caravas, Benjamin Rambo-Martin, Shatavia Morrison, Christopher Gulvick, Scott Sammons, Yvette Unoarumhi, Darlene Wagner, Matthew Schmerer, S. H. Rosenthal, A. Gerasimova, R. M. Kagan, B. Anderson, M. Hua, Y. Liu, L.E. Bernstein, K.E. Livingston, A. Perez, I. A. Shlyakhter, R. V. Rolando, R. Owen, P. Tanpaiboon, F. Lacbawan, Clinton R. Paden, Duncan MacCannell |
| EPI_ISL_2441920 | Fulgent Genetics | Centers for Disease Control and Prevention Division of Viral Diseases, Pathogen Discovery | Dakota Howard, Dhwani Batra, Peter W. Cook, Kara Moser, Adrian Paskey, Jason Caravas, Benjamin Rambo-Martin, Shatavia Morrison, Christopher Gulvick, Scott Sammons, Yvette Unoarumhi, Darlene Wagner, Matthew Schmerer, Harry Gao, Mickey Li, John Gao, Joseph Fierro, Benafsh Sapra, Becky Tsai, Yan Meng, Doreen Ng, James Xie, Clinton R. Paden, Duncan MacCannell |
| EPI_ISL_2443110, EPI_ISL_2443125 | NJDOH, Public Health and Environmental Laboratories | NJ_PHEL | Lindsey Bodnar, Shiv K. Verma, Ryan Pachucki, Chelsea San Filippo, Dana Woell, Allison Roder, Byeong Jeong |
| EPI_ISL_2443167 | IN State Department of Health Laboratory Services | IN State Department of Health Laboratory Services | Cassandra Campion, Jamie Yeadon, Brian Pope, Lixia Liu, Kyle Brownlee, Melissa Hindenlang, Mark Glazier |
| EPI_ISL_2443851, EPI_ISL_2443852 | Minnesota Department of Health, Public Health Laboratory | Minnesota Department of Health, Public Health Laboratory | Alexandra Lorentz, Jacob Garfin, Matt Plumb, and Xiong Wang |
| EPI_ISL_2444439, EPI_ISL_2444447 | Salud Digna | Instituto Nacional de Medicina Genomica | Hidalgo-Miranda A, Cedro-Tanda A, Mendoza-Vargas A, Reyes-Grajeda JP, Abraham Campos-Romero, Moreno-Camacho José Luis, Rodriguez-Gallegos Jorge, Luna-Ruiz Marco, Gonzalez-Barrera D, Rangel-DeLeon D, Munguia-Garza P, Ramirez-Vega O, Escobar-Arrazola, M, Herrera-Montalvo LA. |
| EPI_ISL_2446040, EPI_ISL_2446042, EPI_ISL_2446048, EPI_ISL_2446054, EPI_ISL_2446056, EPI_ISL_2446067, EPI_ISL_2446074, EPI_ISL_2446079, EPI_ISL_2446096 | Pandemic Response Lab - NYC | Pandemic Response Lab, R&D | Henry Lee, Michael Hammerling, Melissa Hopkins, Cybill del Castillo, Shinyoung Clair Kang, William Ward, Pradeep Bugga, Sol Rey, Dylan Law, Katharine Nelson, Haiping Hao, Jon Laurent |
| EPI_ISL_2448099 | Microbiology Department, Laboratori Clínic Metropolitana | Can Ruti SARS-CoV-2 Sequencing Hub | |

| | | | |
|---|---|---|---|
| Nord. Hospital Universitari Germans Trias i Pujol | (HUGTiP/IrsiCaixa/IGTP) | | Marc Noguera-Julian, Pilar Armengol, Ignacio Blanco, Antoni E Bordoy, Francesc Catala-Moll, Pere-Joan Cardona, Maria Casadell, Cristina Casa, Gemma Clara, Bonaventura Clotet, Cristina Esteban, Montserrat Gimnez, Mercedes Guerrero, Anna Not, Roger Paredes, Mariona Parera, Vernica Saludes, Alba S nchez, and Elisa Martron behalf of the Can Ruti SARS-CoV-2 Sequencing Hub. |
| EPI_ISL_2451268, EPI_ISL_2451269, EPI_ISL_2451277, EPI_ISL_2451317, EPI_ISL_2451325, EPI_ISL_2451326, EPI_ISL_2451332, EPI_ISL_2451335, EPI_ISL_2451340, EPI_ISL_2451344, EPI_ISL_2451355, EPI_ISL_2451360, EPI_ISL_2451367, EPI_ISL_2451372, EPI_ISL_2451387, EPI_ISL_2451392, EPI_ISL_2451394, EPI_ISL_2451399, EPI_ISL_2451401 | | | |
| see above | Quest Diagnostics Incorporated | Centers for Disease Control and Prevention Division of Viral Diseases, Pathogen Discovery | Dakota Howard, Dhwani Batra, Peter W. Cook, Kara Moser, Adrian Paskey, Jason Caravas, Benjamin Rambo-Martin, Shatavia Morrison, Christopher Gulvick, Scott Sammons, Yvette Unoarumhi, Darlene Wagner, Matthew Schmerer, S. H. Rosenthal, A. Gerasimova, R. M. Kagan, B. Anderson, M. Hua, Y. Liu, L.E. Bernstein, K.E. Livingston, A. Perez, I. A. Shlyakhter, R. V. Rolando, R. Owen, P. Tanpaiboon, F. Lacbawan, Clinton R. Paden, Duncan MacCannell |
| EPI_ISL_2455301 | Johns Hopkins Hospital Department of Pathology | Johns Hopkins Hospital Department of Pathology | C. Paul Morris, Chun Huai Luo, Adannaya Amadi, Matthew Schwartz, Nicholas Gallagher, Heba H. Mostafa |
| EPI_ISL_2455346, EPI_ISL_2455351, EPI_ISL_2455355, EPI_ISL_2455361 | Minnesota Department of Health, Public Health Laboratory | Minnesota Department of Health, Public Health Laboratory | Alexandra Lorentz, Jacob Garfin, Matt Plumb, and Xiong Wang |
| EPI_ISL_2455380 | HealthPartners Central Lab | Minnesota Department of Health, Public Health Laboratory | Alexandra Lorentz, Jacob Garfin, Matt Plumb, and Xiong Wang |
| EPI_ISL_2458208 | NYU Langone Health | Departments of Pathology and Medicine, New York University School of Medicine | Adriana Heguy, Dacia Dimartino, Emily Guzman, Christian Marier, Peter Meyn, Sitharam Ramaswami, Gael Westby, Paul Zappile, Yutong Zhang, Paolo Cotzia, Guiqing Wang |
| EPI_ISL_2463036, EPI_ISL_2463039, EPI_ISL_2463044, EPI_ISL_2463063, EPI_ISL_2463065, EPI_ISL_2463090, EPI_ISL_2463122, EPI_ISL_2463130, EPI_ISL_2463136, EPI_ISL_2463145, EPI_ISL_2463165, EPI_ISL_2463173, EPI_ISL_2463195, EPI_ISL_2463197 | | | |
| see above | Quest Diagnostics Incorporated | Centers for Disease Control and Prevention Division of Viral Diseases, Pathogen Discovery | Dakota Howard, Dhwani Batra, Peter W. Cook, Kara Moser, Adrian Paskey, Jason Caravas, Benjamin Rambo-Martin, Shatavia Morrison, Christopher Gulvick, Scott Sammons, Yvette Unoarumhi, Darlene Wagner, Matthew Schmerer, S. H. Rosenthal, A. Gerasimova, R. M. Kagan, B. Anderson, M. Hua, Y. Liu, L.E. Bernstein, K.E. Livingston, A. Perez, I. A. Shlyakhter, R. V. Rolando, R. Owen, P. Tanpaiboon, F. Lacbawan, Clinton R. Paden, Duncan MacCannell |
| EPI_ISL_2464863 | Biogroup Bio Lam-LCD Saint-Denis | Department of Virology, Henri Mondor University Hospital, Assistance Publique Hôpitaux de Paris, Université Paris-Est Créteil, INSERM U955 | Christophe Rodriguez, Slim Fourati, Vanessa Demontant, Guillaume Gricourt, Melissa N'Debi, Alexandre Soulier, Elisabeth Trawinski, Jean-Michel Pawlotsky |
| EPI_ISL_2466670 | Stony Brook Medicine | New York Genome Center | Michael Zody, Andre Corvelo, Dayna M. Oschwald, Samantha Fennessey, Tom Maniatis , Sarah A. Halbohn, Alaa B. Salaheldin, Sarah A. Malik, Josue M. Flores Acuna, Dimitri Gnatenko, Silvia G. Spitzer, Margaret M. McGovern, Eric D. Spitzer, Richard R. Kew, Kenneth R. Shroyer, Jiang Chen, Susan V. Donelan |
| EPI_ISL_2466695 | Sherman Abrams Laboratory | New York City Public Health Laboratory | Jade Wang, et al. |
| EPI_ISL_2466697 | Department of Homeless Services | New York City Public Health Laboratory | Jade Wang, et al. |
| EPI_ISL_2466700 | OCME Office Of Chief Medical Examiner | New York City Public Health Laboratory | Jade Wang, et al. |
| EPI_ISL_2470118 | LabKom - Labor an der Salzbrücke MVZ GmbH | Robert Koch Institute | unknown |
| EPI_ISL_2473668, EPI_ISL_2473669, EPI_ISL_2473673, EPI_ISL_2473679 | Oregon State Public Health Laboratory | Oregon State Public Health Laboratory | Rafia Razzaque, Eugene Yeboah, Vanda Makris, Laura Tsaknaridis, John Fontana and Shane Sevey |
| EPI_ISL_2476337, EPI_ISL_2476341 | Dutch COVID-19 response team | National Institute for Public Health and the Environment (RIVM) | Adam Meijer, Harry Vennema, Dirk Eggink, Jeroen Cremer, Sharon van den Brink, Bas van der Veer, AnneMarie van den Brandt, Lisa Wijsman, Kim Freriks, Ryanne Jaarsma, Euníce Then, Lynn Aarts, Sanne Bos, Melissa van Tuil, Linda van de Nes, Florian Zwagemaker, Dennis Schmitz, Annelies Kroneman, Karim Hajji, Chantal Reusken, on behalf of the national COVID-19 response team |
| EPI_ISL_2480024, EPI_ISL_2480094 | Fulgent Genetics | Centers for Disease Control and Prevention Division of Viral Diseases, Pathogen Discovery | Dakota Howard, Dhwani Batra, Peter W. Cook, Kara Moser, Adrian Paskey, Jason Caravas, Benjamin Rambo-Martin, Shatavia Morrison, Christopher Gulvick, Scott Sammons, Yvette Unoarumhi, Darlene Wagner, Matthew Schmerer, Harry Gao, Mickey Li, John Gao, Joseph Fierro, Benafsh Sapra, Becky Tsai, Yan Meng, Doreen Ng, James Xie, Clinton R. Paden, Duncan MacCannell |
| EPI_ISL_2480205, EPI_ISL_2480206, EPI_ISL_2480219, EPI_ISL_2480228, EPI_ISL_2480231, EPI_ISL_2480249, EPI_ISL_2480257, EPI_ISL_2480291, EPI_ISL_2480296, EPI_ISL_2480320, EPI_ISL_2480327, EPI_ISL_2480328, EPI_ISL_2480340, EPI_ISL_2480386, EPI_ISL_2480388, EPI_ISL_2480412, EPI_ISL_2480463, EPI_ISL_2480464, EPI_ISL_2480466, EPI_ISL_2480480, EPI_ISL_2480482, EPI_ISL_2480496, EPI_ISL_2480497, EPI_ISL_2480498, EPI_ISL_2480503, EPI_ISL_2480508, EPI_ISL_2480514, EPI_ISL_2480518, EPI_ISL_2480523, EPI_ISL_2480529, EPI_ISL_2480788, EPI_ISL_2480795, EPI_ISL_2480802, EPI_ISL_2480813, EPI_ISL_2480820, EPI_ISL_2480833, EPI_ISL_2480838, EPI_ISL_2480843, EPI_ISL_2480850, EPI_ISL_2480879, EPI_ISL_2480885, EPI_ISL_2480894, EPI_ISL_2480897, EPI_ISL_2480916, EPI_ISL_2480925, EPI_ISL_2480937, EPI_ISL_2480946, EPI_ISL_2480947, EPI_ISL_2480948, EPI_ISL_2480950, EPI_ISL_2480958, EPI_ISL_2480966, EPI_ISL_2480971, EPI_ISL_2481122, EPI_ISL_2481139, EPI_ISL_2481150, EPI_ISL_2481160, EPI_ISL_2481176, EPI_ISL_2481198, EPI_ISL_2481210, EPI_ISL_2481226, EPI_ISL_2481227, EPI_ISL_2481228, EPI_ISL_2481239, EPI_ISL_2481241, EPI_ISL_2481242, EPI_ISL_2481246, EPI_ISL_2481271, EPI_ISL_2481275, EPI_ISL_2481277, EPI_ISL_2481284, EPI_ISL_2481304, EPI_ISL_2481305, EPI_ISL_2481306, EPI_ISL_2481315, EPI_ISL_2481316, EPI_ISL_2481317, EPI_ISL_2481392, EPI_ISL_2481407, EPI_ISL_2481436, EPI_ISL_2481439, EPI_ISL_2481440, EPI_ISL_2481444, EPI_ISL_2481452, EPI_ISL_2481453, EPI_ISL_2481457, EPI_ISL_2481482, EPI_ISL_2481483, EPI_ISL_2481584, EPI_ISL_2481585, EPI_ISL_2481586, EPI_ISL_2481629, EPI_ISL_2481665, EPI_ISL_2481675, EPI_ISL_2481735, EPI_ISL_2481747, EPI_ISL_2481773, EPI_ISL_2481791, EPI_ISL_2481818, EPI_ISL_2481824, EPI_ISL_2481863, EPI_ISL_2481873, EPI_ISL_2481923, EPI_ISL_2481924, EPI_ISL_2481925, EPI_ISL_2481978, EPI_ISL_2481983, EPI_ISL_2481995, EPI_ISL_2482005, EPI_ISL_2482019, EPI_ISL_2482028, EPI_ISL_2482071, EPI_ISL_2482091, EPI_ISL_2482095, EPI_ISL_2482098, EPI_ISL_2482132, EPI_ISL_2482187, EPI_ISL_2482207, EPI_ISL_2482258, EPI_ISL_2482265, EPI_ISL_2482268 | | | |
| see above | Laboratory Corporation of | Centers for Disease | Dakota Howard, Dhwani Batra, Peter W. Cook, Kara Moser, Adrian Paskey, Jason |

| | | | |
|---|---|---|---|
| | America | Control and Prevention Division of Viral Diseases, Pathogen Discovery | Caravas, Benjamin Rambo-Martin, Shatavia Morrison, Christopher Gulvick, Scott Sammons, Yvette Unoarumhi, Darlene Wagner, Matthew Schmerer, Minoo Agarwal, Eyad Almasri, Debbie Boles, Ayla Burns, Nuthawin Charoensri, Oren Cohen, Susan Countryman, Mary Ann Cristobal, Bobbi Croy, Suzanne Dale, Hrushikesh Deshmukh, Amanda Douglas, Vincent Drouillon, Marcia Eisenberg, Howard Engler, Rama Ghatti, Prashant Gupta, Susan Hicks, Jake Humphrey, Lax Iyer, Lisa Pfefferle, Manoj Jain, Matthew Robinson, Mohan Kolli, Brian Krueger, Tim Kuphal, Stanley Letovsky, Michael Levandoski, Craig Lukasik, Jonathan Meltzer, Brian Norvell, Mindy Nye, Scott Parker, Christos Petropoulos, John Pruitt, Steven Ragan, Scott Ryan, Mike Sapeta, Jana Schroth, Suresh Babu Selvaraju, Goran Stevovic, Amanda Suchanek, Andrea Throop, Lyndon Tilson, Thomas Urban, Joe Voshell, Kimberly Wagner, Jonathan Williams, Mary Williamson, Qian Zeng, Tricia Zwiefelhofer, Clinton R. Paden, Duncan MacCannell |
| EPI_ISL_2484115 | Arizona State University | Arizona State University | Peter T. Skidmore, LaRinda A. Holland, Matthew F. Smith, Rabia Maqsood, Nicholas J. Mellor, Joy M. Blain, Valerie Harris, Joshua LaBaer, Vel Murugan, Efrem S. Lim |
| EPI_ISL_2489070, EPI_ISL_2489071, EPI_ISL_2489111, EPI_ISL_2489140, EPI_ISL_2489179, EPI_ISL_2489180, EPI_ISL_2489188, EPI_ISL_2489254, EPI_ISL_2489256, EPI_ISL_2489259, EPI_ISL_2489264, EPI_ISL_2489276, EPI_ISL_2489287, EPI_ISL_2489296, EPI_ISL_2489299, EPI_ISL_2489304, EPI_ISL_2489310, EPI_ISL_2489311, EPI_ISL_2489326, EPI_ISL_2489333, EPI_ISL_2489336, EPI_ISL_2489339, EPI_ISL_2489359, EPI_ISL_2489388, EPI_ISL_2489399, EPI_ISL_2489400, EPI_ISL_2489408, EPI_ISL_2489466, EPI_ISL_2489467, EPI_ISL_2489468, EPI_ISL_2489469, EPI_ISL_2489484, EPI_ISL_2489487, EPI_ISL_2489556, EPI_ISL_2489563, EPI_ISL_2489569, EPI_ISL_2489571, EPI_ISL_2489586, EPI_ISL_2489595, EPI_ISL_2489596, EPI_ISL_2489600, EPI_ISL_2489606, EPI_ISL_2489641, EPI_ISL_2489644, EPI_ISL_2489660, EPI_ISL_2489661, EPI_ISL_2489663, EPI_ISL_2489669, EPI_ISL_2489738, EPI_ISL_2489744, EPI_ISL_2489745, EPI_ISL_2489747, EPI_ISL_2489772, EPI_ISL_2489775, EPI_ISL_2489778, EPI_ISL_2489780, EPI_ISL_2489794, EPI_ISL_2489796, EPI_ISL_2489799 | | | |
| see above | Aegis Sciences Corporation | Centers for Disease Control and Prevention Division of Viral Diseases, Pathogen Discovery | Dakota Howard, Dhwani Batra, Peter W. Cook, Kara Moser, Adrian Paskey, Jason Caravas, Benjamin Rambo-Martin, Shatavia Morrison, Christopher Gulvick, Scott Sammons, Yvette Unoarumhi, Darlene Wagner, Matthew Schmerer, Cyndi Clark, Patrick Campbell, Rob Case, Vikramsinha Ghorpade, Holly Houdeshell, Ola Kvalvaag, Dillon Nall, Ethan Sanders, Alec Vest, Shaun Westlund, Matthew Hardison, Clinton R. Paden, Duncan MacCannell |
| EPI_ISL_2492341 | Institute of Microbiology, Universidad San Francisco de Quito | Institute of Microbiology, Universidad San Francisco de Quito | Belén Prado-Vivar, Sully Márquez, Juan José Guadalupe, Monica Becerra-Wong, Fernanda Zurita, Bernardo Gutiérrez, Luis Morales, Verónica Barragán, Patricio Rojas-Silva, Gabriel Trueba, Michelle Grunauer, Paúl Cárdenas |
| EPI_ISL_2492353 | Institute of Microbiology, Universidad San Francisco de Quito | Institute of Microbiology, Universidad San Francisco de Quito | Fernanda Zurita, Sully Márquez, Juan José Guadalupe, Belén Prado-Vivar, Monica Becerra-Wong, Bernardo Gutiérrez, Lofrein Vera, Verónica Barragán, Patricio Rojas-Silva, Gabriel Trueba, Michelle Grunauer, Paúl Cárdenas |
| EPI_ISL_2495707, EPI_ISL_2495708, EPI_ISL_2495710, EPI_ISL_2495711, EPI_ISL_2495712 | MB-Cadham Provincial laboratory | National Microbiology Laboratory (NML) | Anna Majer, Shari Tyson, Grace Seo, Philip Mabon, Elsie Grudeski, Rhiannon Huzarewich, Russell Mandes, Anneliese Landgraff, Jennifer Tanner, Natalie Knox, Morag Graham, Gary Van Domselaar, Paul Van Caeseele, Jared Bullard, David Alexander, Kerry Dust, Nathalie Bastien, Yan Li, Timothy Booth, Darian Hole, Madison Chapel, Kirsten Biggar, CanCOGeN's metadata curation team, Public Health Agency of Canada CanCOGeN team |
| EPI_ISL_2495921 | Nacionaline visuomenes sveikatos prieziuros laboratorija | Vilnius University Hospital Santaros Klinikos, Center of Laboratory Medicine | Ligita Raugaite, Monika Katenaite, Rimvydas Norvilas, Dovile Ezerskyte, Ingrida Olendraite, Gytis Dudas, Daniel Naumovas |
| EPI_ISL_2496092 | North Dakota Department of Health, Public Health Laboratory | North Dakota Department of Health, Public Health Laboratory | Lisa Wingerter |
| EPI_ISL_2497192, EPI_ISL_2497224, EPI_ISL_2497270, EPI_ISL_2497317, EPI_ISL_2497417, EPI_ISL_2497427 | UW Virology Lab | UW Virology Lab | Pavitra Roychoudhury, Hong Xie, Lasata Shrestha, Tien V. Nguyen, Shah Mohamed Bakhash, Michelle Lin, Noah R. Baker, Ricardo Perez, Sean Ellis, Nathan Breit, Robert J. Livingston, Meei-Li Huang, Keith R Jerome, Patrick Mathias, Alexander Greninger |
| EPI_ISL_2497892 | University of Mississippi Medical Center, Department of Pathology | University of Mississippi Medical Center, Molecular and Genomics Core Facility | Ashley C. Johnson, Ithiel J. Frame, Krishna K. Ayyalasomayajula, Michael R. Garrett, D. Ashley Robinson |
| EPI_ISL_2497926, EPI_ISL_2497927 | UW Virology Lab | UW Virology Lab | Pavitra Roychoudhury, Hong Xie, Lasata Shrestha, Tien V. Nguyen, Shah Mohamed Bakhash, Michelle Lin, Noah R. Baker, Ricardo Perez, Sean Ellis, Nathan Breit, Robert J. Livingston, Meei-Li Huang, Keith R Jerome, Patrick Mathias, Alexander Greninger |
| EPI_ISL_2499856 | DC Public Health Lab/ Dept. of Forensic Sciences | DC Public Health Lab/ Dept. of Forensic Sciences | Janis Doss, Scott Nguyen, Elizabeth Zelaya, Sarah Scott, Connie Maza, Monica Mann, Brittany Hamilton, David Payne, Jocelyn Hauser |
| EPI_ISL_2500459 | Hackensack Medical Center | New York Genome Center | Michael Zody, Andre Corvelo, Dayna M. Oschwald, Samantha Fennessey, Tom Maniatis, Liang Chen, Jose Mediavilla, Marcus Cunningham, Kaelea Composto, Kar Chow, David Perlin, Barry Kreiswirth |
| EPI_ISL_2501004, EPI_ISL_2501013 | Gundersen Clinical Microbiology Laboratory | Kabara Cancer Research Institute | Craig S. Richmond, Paraic A. Kenny |
| EPI_ISL_2501377, EPI_ISL_2501378 | Microbiology Department. Complexo Hospitalario Universitario de Vigo | Microbiology Department. Complexo Hospitalario Universitario de Vigo | Alfaya N, Alonso I, Alvarez M, Cabrera JJ, Carballo R, Cores O, Cortizo S, del-Campo V, Martinez L, Mediero G, Perez S, Potel C, Regueiro B, Rey S, Vasallo FJ |
| EPI_ISL_2501498, EPI_ISL_2501508, EPI_ISL_2501517, EPI_ISL_2501518, EPI_ISL_2501519, EPI_ISL_2501520, EPI_ISL_2501521, EPI_ISL_2501522 | UW Virology Lab | UW Virology Lab | Pavitra Roychoudhury, Hong Xie, Lasata Shrestha, Tien V. Nguyen, Shah Mohamed Bakhash, Michelle Lin, Noah R. Baker, Ricardo Perez, Sean Ellis, Nathan Breit, Robert J. Livingston, Meei-Li Huang, Keith R Jerome, Patrick Mathias, Alexander Greninger |

| | | | |
|---|---|---|---|
| EPI_ISL_2501539 | CT Department of Public Health | CT Department of Public Health | Claire_Pearson,Tu_N_Nguyen |
| EPI_ISL_2501696, EPI_ISL_2501706, EPI_ISL_2501727 | Yale Clinical Virology Lab | Grubaugh Lab - Yale School of Public Health | Joseph Fauver, Mallery Breban, Isabel Ott, Tara Alpert, Mary Petrone, Anderson Brito, Chantal Vogels, Annie Watkins, Chaney Kalinich, Jessica Rothman, Marie L. Landry, Nathan Grubaugh |
| EPI_ISL_2503994, EPI_ISL_2503996 | Alaska State Virology Laboratory | Alaska State Virology Laboratory | Stephanie DeRonde, Elva House, Jacob Zidek, Lisa Smith, Ph.D., Jack Chen, Ph.D. |
| EPI_ISL_2508533 | Microbiology Division, SC DHEC | Microbiology Division, SC DHEC | Freeman,J., Flores,H. |
| EPI_ISL_2525599, EPI_ISL_2525754 | Fulgent Genetics | Centers for Disease Control and Prevention Division of Viral Diseases, Pathogen Discovery | Dakota Howard, Dhwani Batra, Peter W. Cook, Kara Moser, Adrian Paskey, Jason Caravas, Benjamin Rambo-Martin, Shatavia Morrison, Christopher Gulvick, Scott Sammons, Yvette Unoarumhi, Darlene Wagner, Matthew Schmerer, Harry Gao, Mickey Li, John Gao, Joseph Fierro, Benafsh Sapra, Becky Tsai, Yan Meng, Doreen Ng, James Xie, Clinton R. Paden, Duncan MacCannell |
| EPI_ISL_2526772, EPI_ISL_2526783, EPI_ISL_2526811, EPI_ISL_2526816, EPI_ISL_2526827, EPI_ISL_2526844, EPI_ISL_2526867, EPI_ISL_2526875, EPI_ISL_2526976, EPI_ISL_2526994, EPI_ISL_2527028, EPI_ISL_2527034 | | | |
| see above | Quest Diagnostics Incorporated | Centers for Disease Control and Prevention Division of Viral Diseases, Pathogen Discovery | Dakota Howard, Dhwani Batra, Peter W. Cook, Kara Moser, Adrian Paskey, Jason Caravas, Benjamin Rambo-Martin, Shatavia Morrison, Christopher Gulvick, Scott Sammons, Yvette Unoarumhi, Darlene Wagner, Matthew Schmerer, S. H. Rosenthal, A. Gerasimova, R. M. Kagan, B. Anderson, M. Hua, Y. Liu, L.E. Bernstein, K.E. Livingston, A. Perez, I. A. Shlyakhter, R. V. Rolando, R. Owen, P. Tanpaiboon, F. Lacbawan, Clinton R. Paden, Duncan MacCannell |
| EPI_ISL_2527594, EPI_ISL_2527601, EPI_ISL_2527714, EPI_ISL_2527745, EPI_ISL_2527831 | Fulgent Genetics | Centers for Disease Control and Prevention Division of Viral Diseases, Pathogen Discovery | Dakota Howard, Dhwani Batra, Peter W. Cook, Kara Moser, Adrian Paskey, Jason Caravas, Benjamin Rambo-Martin, Shatavia Morrison, Christopher Gulvick, Scott Sammons, Yvette Unoarumhi, Darlene Wagner, Matthew Schmerer, Harry Gao, Mickey Li, John Gao, Joseph Fierro, Benafsh Sapra, Becky Tsai, Yan Meng, Doreen Ng, James Xie, Clinton R. Paden, Duncan MacCannell |
| EPI_ISL_2527904, EPI_ISL_2527929, EPI_ISL_2528008, EPI_ISL_2528010, EPI_ISL_2528058, EPI_ISL_2528123, EPI_ISL_2528173, EPI_ISL_2528199, EPI_ISL_2528257, EPI_ISL_2528314, EPI_ISL_2528354, EPI_ISL_2528430, EPI_ISL_2528464, EPI_ISL_2528472, EPI_ISL_2528479, EPI_ISL_2528486, EPI_ISL_2528510, EPI_ISL_2528513, EPI_ISL_2528528, EPI_ISL_2528530, EPI_ISL_2528659, EPI_ISL_2528668, EPI_ISL_2528670, EPI_ISL_2528686, EPI_ISL_2528714, EPI_ISL_2528800, EPI_ISL_2528898, EPI_ISL_2528900, EPI_ISL_2528916, EPI_ISL_2528927, EPI_ISL_2528929, EPI_ISL_2528931, EPI_ISL_2528951 | | | |
| see above | Aegis Sciences Corporation | Centers for Disease Control and Prevention Division of Viral Diseases, Pathogen Discovery | Dakota Howard, Dhwani Batra, Peter W. Cook, Kara Moser, Adrian Paskey, Jason Caravas, Benjamin Rambo-Martin, Shatavia Morrison, Christopher Gulvick, Scott Sammons, Yvette Unoarumhi, Darlene Wagner, Matthew Schmerer, Cyndi Clark, Patrick Campbell, Rob Case, Vikramsinha Ghorpade, Holly Houdeshell, Ola Kvalvaag, Dillon Nall, Ethan Sanders, Alec Vest, Shaun Westlund, Matthew Hardison, Clinton R. Paden, Duncan MacCannell |
| EPI_ISL_2529102, EPI_ISL_2529104, EPI_ISL_2529152, EPI_ISL_2529190, EPI_ISL_2529219, EPI_ISL_2529244, EPI_ISL_2529256, EPI_ISL_2529276, EPI_ISL_2529293, EPI_ISL_2529310, EPI_ISL_2529335 | | | |
| see above | Quest Diagnostics Incorporated | Centers for Disease Control and Prevention Division of Viral Diseases, Pathogen Discovery | Dakota Howard, Dhwani Batra, Peter W. Cook, Kara Moser, Adrian Paskey, Jason Caravas, Benjamin Rambo-Martin, Shatavia Morrison, Christopher Gulvick, Scott Sammons, Yvette Unoarumhi, Darlene Wagner, Matthew Schmerer, S. H. Rosenthal, A. Gerasimova, R. M. Kagan, B. Anderson, M. Hua, Y. Liu, L.E. Bernstein, K.E. Livingston, A. Perez, I. A. Shlyakhter, R. V. Rolando, R. Owen, P. Tanpaiboon, F. Lacbawan, Clinton R. Paden, Duncan MacCannell |
| EPI_ISL_2534389, EPI_ISL_2534397, EPI_ISL_2534400 | HOSPITAL UNIVERSITARIO SON ESPASES | HOSPITAL UNIVERSITARIO SON ESPASES | Servicio de Microbiología, Hospital Universitario Son Espases |
| EPI_ISL_2534404 | HOSPITAL CAN MISSES | HOSPITAL UNIVERSITARIO SON ESPASES | Servicio de Microbiología, Hospital Universitario Son Espases |
| EPI_ISL_2534405, EPI_ISL_2534407, EPI_ISL_2534415, EPI_ISL_2534417, EPI_ISL_2534425 | HOSPITAL UNIVERSITARIO SON ESPASES | HOSPITAL UNIVERSITARIO SON ESPASES | Servicio de Microbiología, Hospital Universitario Son Espases |
| EPI_ISL_2536950 | Centracare Laboratory Services | Minnesota Department of Health, Public Health Laboratory | Alexandra Lorentz, Jacob Garfin, Matt Plumb, and Xiong Wang |
| EPI_ISL_2537154, EPI_ISL_2537170, EPI_ISL_2537175, EPI_ISL_2537224, EPI_ISL_2537245, EPI_ISL_2537267 | Gravity Diagnostics, LLC | Gravity Diagnostics, LLC | Gravity Diagnostics |
| EPI_ISL_2537367, EPI_ISL_2537371 | Montana Public Health Laboratory | Montana Public Health Laboratory | Joy Ritter, Michelle Mozer, Carrie Biskupiak, Deborah Gibson, Michael Dills |
| EPI_ISL_2544621 | IN State Department of Health Laboratory Services | IN State Department of Health Laboratory Services | Cassandra Campion, Jamie Yeadon, Brian Pope, Lixia Liu, Kyle Brownlee, Melissa Hindenlang, Mark Glazier |
| EPI_ISL_2544915, EPI_ISL_2545005, EPI_ISL_2545015, EPI_ISL_2545016, EPI_ISL_2545017, EPI_ISL_2545021, EPI_ISL_2545029, EPI_ISL_2545055, EPI_ISL_2545125, EPI_ISL_2545134, EPI_ISL_2545149 | | | |
| see above | UW Virology Lab | UW Virology Lab | Pavitra Roychoudhury, Hong Xie, Lasata Shrestha, Shah Mohamed Bakhash, Tien V. Nguyen, Noah R. Baker, Sean Ellis, Meei-Li Huang, Keith R Jerome, Alexander Greninger |
| EPI_ISL_2547288, EPI_ISL_2547289 | UHTL, University Hospitals | UHTL, University Hospitals | Sadri,N., Alouani,D., Song,X. |
| EPI_ISL_2549330, EPI_ISL_2549360, EPI_ISL_2549364, EPI_ISL_2549373, EPI_ISL_2549377, EPI_ISL_2549402, EPI_ISL_2549403, EPI_ISL_2549410, EPI_ISL_2549420, EPI_ISL_2549427, EPI_ISL_2549429, EPI_ISL_2549461, EPI_ISL_2549463, EPI_ISL_2549467, EPI_ISL_2549468, EPI_ISL_2549470, EPI_ISL_2549486, EPI_ISL_2549487, EPI_ISL_2549489, EPI_ISL_2549491, EPI_ISL_2549500, EPI_ISL_2549513, EPI_ISL_2549514 | | | |
| see above | UW Virology Lab | UW Virology Lab | Pavitra Roychoudhury, Hong Xie, Lasata Shrestha, Tien V. Nguyen, Shah Mohamed |

| | | | |
|---|---|---|---|
| | | | Bakhash, Michelle Lin, Noah R. Baker, Ricardo Perez, Sean Ellis, Nathan Breit, Robert J. Livingston, Meei-Li Huang, Keith R Jerome, Patrick Mathias, Alexander Greninger |
| EPI_ISL_2550374, EPI_ISL_2550427 | Colorado Department of Public Health and Environment | Colorado Department of Public Health and Environment | Laura Bankers, Molly C. Hetherington-Rauth, Diana Ir, Alexandria Rossheim, Shannon R. Matzinger, Sarah Elizabeth Totten, Emily A. Travanty |
| EPI_ISL_2550763 | Maryland Public Health Laboratory (MD PHL) | Maryland Public Health Laboratory (MD PHL) | Maryland Department of Health Laboratories Administration |
| EPI_ISL_2557051 | Washington State Department of Health Public Health Laboratories | Washington State Department of Health Public Health Laboratories | Drew MacKellar, Philip Dykema, Denny Russell, Joenice Gonzalez, Hannah Gray, Geoff Melly, Vanessa De Los Santos, Darren Lucas, JohnAric Peterson, Avi Singh, Rebecca Cao |
| EPI_ISL_2557124, EPI_ISL_2557128, EPI_ISL_2557146, EPI_ISL_2557147, EPI_ISL_2557151 | NJDOH, Public Health and Environmental Laboratories | NJ_PHEL | Lindsey Bodnar, Shiv K. Verma, Ryan Pachucki, Chelsea San Filippo, Dana Woell, Allison Roder, Byeong Jeong |
| EPI_ISL_2557433, EPI_ISL_2557439, EPI_ISL_2557526, EPI_ISL_2557540, EPI_ISL_2557561, EPI_ISL_2557591, EPI_ISL_2557595 | Infinity Biologix | Centers for Disease Control and Prevention Division of Viral Diseases, Pathogen Discovery | Dakota Howard, Dhwani Batra, Peter W. Cook, Kara Moser, Adrian Paskey, Jason Caravas, Benjamin Rambo-Martin, Shatavia Morrison, Christopher Gulvick, Scott Sammons, Yvette Unoarumhi, Darlene Wagner, Matthew Schmerer, Christian Bixby, Yihe Wang, Jonathan Schultz, Chirayu Goswami, Russ Hager, Robin Grimwood, Clinton R. Paden, Duncan MacCannell |
| EPI_ISL_2557929, EPI_ISL_2557934 | Oregon State Public Health Laboratory | Oregon State Public Health Laboratory | Rafia Razzaque, Eugene Yeboah, Vanda Makris, Laura Tsaknaridis, John Fontana and Shane Sevey |
| EPI_ISL_2560433 | Wyoming Public Health Laboratory | Wyoming Public Health Laboratory | Jim Mildenberger, Taylor Fearing, Channing Weber, Ashley Norberg, Chayse Rowley, Marley Goetz, Brian Dominguez, Elliot Thomasson, Sam Britz, Cari Sloma, Robert Petit, and Rob Christensen |
| EPI_ISL_2562121, EPI_ISL_2562122 | The Jackson Laboratory | The Jackson Laboratory | Long J,Bergeron D, Renzette N, Adams M, Omerza G, Kelly K, Li L |
| EPI_ISL_2565775 | TXDSHS | TXDSHS | Rashmi Tuladhar, Bonnie Oh, Jenny Zhang, Maliha Rahman, Mayela Pedrueza, Anita Pokharel, Karen Bobier, Lorraine Rodriguez, Myong Koag, Chun Wang, Rachel Lee, Grace Kubin |
| EPI_ISL_2565868 | Minnesota Department of Health, Public Health Laboratory | Minnesota Department of Health, Public Health Laboratory | Alexandra Lorentz, Jacob Garfin, Matt Plumb, and Xiong Wang |
| EPI_ISL_2565915, EPI_ISL_2565957, EPI_ISL_2565964 | North Dakota Department of Health, Public Health Laboratory | North Dakota Department of Health, Public Health Laboratory | Lisa Wingerter |
| EPI_ISL_2566090 | Wyoming Public Health Laboratory | Wyoming Public Health Laboratory | Jim Mildenberger, Taylor Fearing, Channing Weber, Ashley Norberg, Chayse Rowley, Marley Goetz, Brian Dominguez, Elliot Thomasson, Sam Britz, Cari Sloma, Robert Petit, and Rob Christensen |
| EPI_ISL_2566122 | Nevada State Public Health Laboratory | Nevada State Public Health Laboratory | Andrew Gorzalski, Mark Pandori |
| EPI_ISL_2597647, EPI_ISL_2597694, EPI_ISL_2597695, EPI_ISL_2597704 | Hospital of the University of Pennsylvania Molecular Pathology Lab | Bushman Lab - University of Pennsylvania | John K. Everett, Kyle Rodino, Shantan Reddy, Aoife M. Roche, Young Hwang, Scott Sherrill-Mix, Samantha A. Whiteside, Jevon Graham-Wooten, Layla A. Khatib, Ayannah S. Fitzgerald, Arupa Ganguly, Mike Feldman, Brendan Kelly, Ronald G. Collman and Frederic Bushan |
| EPI_ISL_2597928 | Nebraska Public Health Laboratory | NPHL COVID-19 Response Team | NPHL COVID-19 Response Team |
| EPI_ISL_2598016 | Hackensack Medical Center | New York Genome Center | Michael Zody, Andre Corvelo, Dayna M. Oschwald, Samantha Fennessey, Tom Maniatis, Liang Chen, Jose Mediavilla, Marcus Cunningham, Kaelea Composto, Kar Chow, David Perlin, Barry Kreiswirth |
| EPI_ISL_2598076, EPI_ISL_2598078 | MD PHL | MD PHL | Maryland Department of Health Laboratories Administration |
| EPI_ISL_2598468, EPI_ISL_2598482 | Maryland Genomics, Institute for Genome Sciences, University of Maryland School of Medicine | Maryland Genomics, Institute for Genome Sciences, University of Maryland School of Medicine | Tallon, Luke J; Sadzewicz, Lisa D; Humphrys, Mike; Ott, Sandra; Roussey, Holly; Santana-Cruz, Ivette; Vavikolanu, Kranthi; Lim, Jonathan;Hazen, Tracy;Fraser, Claire M; Ravel, Jacques |
| EPI_ISL_2598735 | Fulgent Genetics | Centers for Disease Control and Prevention Division of Viral Diseases, Pathogen Discovery | Dakota Howard, Dhwani Batra, Peter W. Cook, Kara Moser, Adrian Paskey, Jason Caravas, Benjamin Rambo-Martin, Shatavia Morrison, Christopher Gulvick, Scott Sammons, Yvette Unoarumhi, Darlene Wagner, Matthew Schmerer, Harry Gao, Mickey Li, John Gao, Joseph Fierro, Benafsh Sapra, Becky Tsai, Yan Meng, Doreen Ng, James Xie, Clinton R. Paden, Duncan MacCannell |
| EPI_ISL_2599642, EPI_ISL_2599673, EPI_ISL_2599683, EPI_ISL_2599695, EPI_ISL_2599700, EPI_ISL_2599747, EPI_ISL_2599749, EPI_ISL_2599750, EPI_ISL_2599782, EPI_ISL_2599835, EPI_ISL_2599848, EPI_ISL_2599883, EPI_ISL_2599916 | | | |
| see above | Quest Diagnostics Incorporated | Centers for Disease Control and Prevention Division of Viral Diseases, Pathogen Discovery | Dakota Howard, Dhwani Batra, Peter W. Cook, Kara Moser, Adrian Paskey, Jason Caravas, Benjamin Rambo-Martin, Shatavia Morrison, Christopher Gulvick, Scott Sammons, Yvette Unoarumhi, Darlene Wagner, Matthew Schmerer, S. H. Rosenthal, A. Gerasimova, R. M. Kagan, B. Anderson, M. Hua, Y. Liu, L.E. Bernstein, K.E. Livingston, A. Perez, I. A. Shlyakhter, R. V. Rolando, R. Owen, P. Tanpaiboon, F. Lacbawan, Clinton R. Paden, Duncan MacCannell |
| EPI_ISL_2601037 | Instituto de Medicina Tropical & Salud Global, Universidad Iberoamericana (UNIBE) | Instituto de Medicina Tropical & Salud Global, Universidad Iberoamericana (UNIBE) | Victor Virgilio Calderon, Alejandro Vallejo Degaudenzi & Robert Paulino-Ramirez |
| EPI_ISL_2603211, EPI_ISL_2603216 | Virginia Division of Consolidated Laboratory Services | Virginia Division of Consolidated Laboratory Services | Virginia DCLS |

| | | | |
|---|---|---|---|
| EPI_ISL_2603317 | Rhode Island State Health Laboratory | Rhode Island State Health Laboratory | Ewa King, Richard C. Huard, Kristin Carpenter-Azevedo |
| EPI_ISL_2603351, EPI_ISL_2603354 | Virginia Division of Consolidated Laboratory Services | Virginia Division of Consolidated Laboratory Services | Virginia DCLS |
| EPI_ISL_2603805 | Salud Digna | Instituto Nacional de Medicina Genomica | Hidalgo-Miranda A, Cedro-Tanda A, Mendoza-Vargas A, Reyes-Grajeda JP, Abraham Campos-Romero, Moreno-Camacho José Luis, Rodriguez-Gallegos Jorge, Luna-Ruiz Marco, Gonzalez-Barrera D, Rangel-DeLeon D, Munguia-Garza P, Ramirez-Vega O, Escobar-Arrazola, M, Herrera-Montalvo LA. |
| EPI_ISL_2604126, EPI_ISL_2604128, EPI_ISL_2604168 | Arizona State University | Arizona State University | Peter T. Skidmore, LaRinda A. Holland, Matthew F. Smith, Rabia Maqsood, Nicholas J. Mellor, Joy M. Blain, Valerie Harris, Joshua LaBaer, Vel Murugan, Efrem S. Lim |
| EPI_ISL_2604199, EPI_ISL_2604200, EPI_ISL_2604201, EPI_ISL_2604203, EPI_ISL_2604206 | Oregon State Public Health Laboratory | Oregon State Public Health Laboratory | Rafia Razzaque, Eugene Yeboah, Vanda Makris, Laura Tsaknaridis, John Fontana and Shane Sevey |
| EPI_ISL_2604227 | Montana Public Health Laboratory | Montana Public Health Laboratory | Joy Ritter, Michelle Mozer, Carrie Biskupiak, Deborah Gibson, Michael Dills |
| EPI_ISL_2604868 | Omics Sciences Laboratory | Omics Sciences Laboratory | Derly Andrade Molina, Gabriel Morey León, Darlyn Amaya, Rubén Armas González, Juan Carlos Fernández Cadena |
| EPI_ISL_2604873, EPI_ISL_2604874, EPI_ISL_2604879, EPI_ISL_2604880 | INTERLAB | Omics Sciences Laboratory | Derly Andrade Molina, Gabriel Morey León, Darlyn Amaya, Rubén Armas González, Juan Carlos Fernández Cadena |
| EPI_ISL_2604881 | Omics Sciences Laboratory | Omics Sciences Laboratory | Derly Andrade Molina, Gabriel Morey León, Darlyn Amaya, Rubén Armas González, Juan Carlos Fernández Cadena |
| EPI_ISL_2609623, EPI_ISL_2610127, EPI_ISL_2610178, EPI_ISL_2610186, EPI_ISL_2610683, EPI_ISL_2610692, EPI_ISL_2610712, EPI_ISL_2610724 | Dutch COVID-19 response team | National Institute for Public Health and the Environment (RIVM) | Adam Meijer, Harry Vennema, Dirk Eggink, Jeroen Cremer, Sharon van den Brink, Bas van der Veer, AnneMarie van den Brandt, Lisa Wijsman, Kim Freriks, Ryanne Jaarsma, Euníce Then, Lynn Aarts, Sanne Bos, Melissa van Tuil, Linda van de Nes, Florian Zwagemaker, Dennis Schmitz, Annelies Kroneman, Karim Hajji, Chantal Reusken, on behalf of the national COVID-19 response team |
| EPI_ISL_2611126, EPI_ISL_2611127, EPI_ISL_2611128, EPI_ISL_2611156, EPI_ISL_2611157, EPI_ISL_2611158, EPI_ISL_2611159, EPI_ISL_2611171, EPI_ISL_2611173, EPI_ISL_2611180, EPI_ISL_2611182, EPI_ISL_2611215, EPI_ISL_2611217, EPI_ISL_2611235, EPI_ISL_2611295, EPI_ISL_2611297, EPI_ISL_2611300, EPI_ISL_2611302, EPI_ISL_2611306, EPI_ISL_2611308, EPI_ISL_2611318, EPI_ISL_2611319, EPI_ISL_2611358, EPI_ISL_2611360, EPI_ISL_2611394, EPI_ISL_2611432, EPI_ISL_2611438, EPI_ISL_2611439, EPI_ISL_2611440, EPI_ISL_2611445, EPI_ISL_2611454, EPI_ISL_2611456, EPI_ISL_2611457, EPI_ISL_2611460, EPI_ISL_2611467, EPI_ISL_2611468, EPI_ISL_2611491, EPI_ISL_2611492, EPI_ISL_2611523, EPI_ISL_2611530, EPI_ISL_2611531, EPI_ISL_2611537, EPI_ISL_2611541, EPI_ISL_2611542, EPI_ISL_2611543, EPI_ISL_2611545, EPI_ISL_2611547, EPI_ISL_2611566, EPI_ISL_2611588, EPI_ISL_2611591, EPI_ISL_2611599, EPI_ISL_2611618, EPI_ISL_2611621, EPI_ISL_2611636, EPI_ISL_2611650, EPI_ISL_2611675, EPI_ISL_2611686, EPI_ISL_2611695, EPI_ISL_2611696, EPI_ISL_2611698, EPI_ISL_2611716, EPI_ISL_2611720, EPI_ISL_2611728, EPI_ISL_2611757, EPI_ISL_2611774, EPI_ISL_2611775, EPI_ISL_2611783, EPI_ISL_2611800, EPI_ISL_2611806, EPI_ISL_2611814, EPI_ISL_2611815, EPI_ISL_2611820, EPI_ISL_2611821 | | | |
| see above | Laboratory Corporation of America | Centers for Disease Control and Prevention Division of Viral Diseases, Pathogen Discovery | Dakota Howard, Dhwani Batra, Peter W. Cook, Kara Moser, Adrian Paskey, Jason Caravas, Benjamin Rambo-Martin, Shatavia Morrison, Christopher Gulvick, Scott Sammons, Yvette Unoarumhi, Darlene Wagner, Matthew Schmerer, Minoo Agarwal, Eyad Almasri, Debbie Boles, Ayla Burns, Nuthawin Charoensri, Oren Cohen, Susan Countryman, Mary Ann Cristobal, Bobbi Croy, Suzanne Dale, Hrushikesh Deshmukh, Amanda Douglas, Vincent Drouillon, Marcia Eisenberg, Howard Engler, Rama Ghatti, Prashant Gupta, Susan Hicks, Jake Humphrey, Lax Iyer, Lisa Pfefferle, Manoj Jain, Matthew Robinson, Mohan Kolli, Brian Krueger, Tim Kuphal, Stanley Letovsky, Michael Levandoski, Craig Lukasik, Jonathan Meltzer, Brian Norvell, Mindy Nye, Scott Parker, Christos Petropoulos, John Pruitt, Steven Ragan, Scott Ryan, Mike Sapeta, Jana Schroth, Suresh Babu Selvaraju, Goran Stevovic, Amanda Suchanek, Andrea Throop, Lyndon Tilson, Thomas Urban, Joe Voshell, Kimberly Wagner, Jonathan Williams, Mary Williamson, Qian Zeng, Tricia Zwiefelhofer, Clinton R. Paden, Duncan MacCannell |
| EPI_ISL_2611834, EPI_ISL_2611854, EPI_ISL_2611856 | Quest Diagnostics Incorporated | Centers for Disease Control and Prevention Division of Viral Diseases, Pathogen Discovery | Dakota Howard, Dhwani Batra, Peter W. Cook, Kara Moser, Adrian Paskey, Jason Caravas, Benjamin Rambo-Martin, Shatavia Morrison, Christopher Gulvick, Scott Sammons, Yvette Unoarumhi, Darlene Wagner, Matthew Schmerer, S. H. Rosenthal, A. Gerasimova, R. M. Kagan, B. Anderson, M. Hua, Y. Liu, L.E. Bernstein, K.E. Livingston, A. Perez, I. A. Shlyakhter, R. V. Rolando, R. Owen, P. Tanpaiboon, F. Lacbawan, Clinton R. Paden, Duncan MacCannell |
| EPI_ISL_2612459 | Montana Public Health Laboratory | Montana Public Health Laboratory | Joy Ritter, Michelle Mozer, Carrie Biskupiak, Deborah Gibson, Michael Dills |
| EPI_ISL_2612532 | Centracare Laboratory Services | Minnesota Department of Health, Public Health Laboratory | Alexandra Lorentz, Jacob Garfin, Matt Plumb, and Xiong Wang |
| EPI_ISL_2612538, EPI_ISL_2612547 | Oregon State Public Health Laboratory | Oregon State Public Health Laboratory | Rafia Razzaque, Eugene Yeboah, Vanda Makris, Laura Tsaknaridis, John Fontana and Shane Sevey |
| EPI_ISL_2614898 | Nacionaline visuomenes sveikatos prieziuros laboratorija | National Public Health Surveillance Laboratory | Lukas Zemaitis, Migle Gabrielaite, Jelena Razmuk, Svajune Muralyte, Ana Steponkiene, Lukas Vasionis, Danas Baksa |
| EPI_ISL_2615045 | Nacionalinis maisto ir veterinarijos rizikos vertinimo institutas | National Public Health Surveillance Laboratory | Lukas Zemaitis, Migle Gabrielaite, Jelena Razmuk, Svajune Muralyte, Ana Steponkiene, Lukas Vasionis, Danas Baksa |
| EPI_ISL_2615726 | QLabs | WVU and Marshall University Combined Genomics Core Facilities | James Denvir, Peter Stoilov, Peter Perrotta, Wesley Kimble, Ryan Percifield |
| EPI_ISL_2615984 | J.W. Ruby Memorial Hospital | WVU and Marshall University Combined | James Denvir, Peter Stoilov, Peter Perrotta, Wesley Kimble, Ryan Percifield |

| | | | |
|---|---|---|---|
| | | Genomics Core Facilities | |
| EPI_ISL_2615985, EPI_ISL_2615986 | Summersville Regional Medical Center | WVU and Marshall University Combined Genomics Core Facilities | James Denvir, Peter Stoilov, Peter Perrotta, Wesley Kimble, Ryan Percifield |
| EPI_ISL_2615987 | QLabs | WVU and Marshall University Combined Genomics Core Facilities | James Denvir, Peter Stoilov, Peter Perrotta, Wesley Kimble, Ryan Percifield |
| EPI_ISL_2615992 | Mon Health Medical Center | WVU and Marshall University Combined Genomics Core Facilities | James Denvir, Peter Stoilov, Peter Perrotta, Wesley Kimble, Ryan Percifield |
| EPI_ISL_2616041 | Berkeley Medical Center | WVU and Marshall University Combined Genomics Core Facilities | James Denvir, Peter Stoilov, Peter Perrotta, Wesley Kimble, Ryan Percifield |
| EPI_ISL_2616042 | QLabs | WVU and Marshall University Combined Genomics Core Facilities | James Denvir, Peter Stoilov, Peter Perrotta, Wesley Kimble, Ryan Percifield |
| EPI_ISL_2616057, EPI_ISL_2616058 | Berkeley Medical Center | WVU and Marshall University Combined Genomics Core Facilities | James Denvir, Peter Stoilov, Peter Perrotta, Wesley Kimble, Ryan Percifield |
| EPI_ISL_2616062 | Mon Health Medical Center | WVU and Marshall University Combined Genomics Core Facilities | James Denvir, Peter Stoilov, Peter Perrotta, Wesley Kimble, Ryan Percifield |
| EPI_ISL_2616063, EPI_ISL_2616064, EPI_ISL_2616065, EPI_ISL_2616066, EPI_ISL_2616067 | QLabs | WVU and Marshall University Combined Genomics Core Facilities | James Denvir, Peter Stoilov, Peter Perrotta, Wesley Kimble, Ryan Percifield |
| EPI_ISL_2616074, EPI_ISL_2616075, EPI_ISL_2616076 | J.W. Ruby Memorial Hospital | WVU and Marshall University Combined Genomics Core Facilities | James Denvir, Peter Stoilov, Peter Perrotta, Wesley Kimble, Ryan Percifield |
| EPI_ISL_2616094 | Berkeley Medical Center | WVU and Marshall University Combined Genomics Core Facilities | James Denvir, Peter Stoilov, Peter Perrotta, Wesley Kimble, Ryan Percifield |
| EPI_ISL_2616095 | J.W. Ruby Memorial Hospital | WVU and Marshall University Combined Genomics Core Facilities | James Denvir, Peter Stoilov, Peter Perrotta, Wesley Kimble, Ryan Percifield |
| EPI_ISL_2616098 | QLabs | WVU and Marshall University Combined Genomics Core Facilities | James Denvir, Peter Stoilov, Peter Perrotta, Wesley Kimble, Ryan Percifield |
| EPI_ISL_2616103 | Berkeley Medical Center | WVU and Marshall University Combined Genomics Core Facilities | James Denvir, Peter Stoilov, Peter Perrotta, Wesley Kimble, Ryan Percifield |
| EPI_ISL_2616104 | Hampshire Memorial Hospital | WVU and Marshall University Combined Genomics Core Facilities | James Denvir, Peter Stoilov, Peter Perrotta, Wesley Kimble, Ryan Percifield |
| EPI_ISL_2616105, EPI_ISL_2616106 | QLabs | WVU and Marshall University Combined Genomics Core Facilities | James Denvir, Peter Stoilov, Peter Perrotta, Wesley Kimble, Ryan Percifield |
| EPI_ISL_2616107 | Berkeley Medical Center | WVU and Marshall University Combined Genomics Core Facilities | James Denvir, Peter Stoilov, Peter Perrotta, Wesley Kimble, Ryan Percifield |
| EPI_ISL_2617018, EPI_ISL_2617019 | Salud Digna | Instituto Nacional de Medicina Genomica | Hidalgo-Miranda A, Cedro-Tanda A, Mendoza-Vargas A, Reyes-Grajeda JP, Abraham Campos-Romero, Moreno-Camacho José Luis, Rodriguez-Gallegos Jorge, Luna-Ruiz Marco, Gonzalez-Barrera D, Rangel-DeLeon D, Munguia-Garza P, Ramirez-Vega O, Escobar-Arrazola, M, Herrera-Montalvo LA. |
| EPI_ISL_2617629, EPI_ISL_2617630, EPI_ISL_2617980, EPI_ISL_2617983, EPI_ISL_2618054, EPI_ISL_2618073, EPI_ISL_2618080 | UW Virology Lab | UW Virology Lab | Pavitra Roychoudhury, Hong Xie, Lasata Shrestha, Tien V. Nguyen, Shah Mohamed Bakhash, Michelle Lin, Noah R. Baker, Ricardo Perez, Sean Ellis, Nathan Breit, Robert J. Livingston, Meei-Li Huang, Keith R Jerome, Patrick Mathias, Alexander Greninger |
| EPI_ISL_2626098 | HOSPITAL UNIVERSITARIO SON ESPASES | HOSPITAL UNIVERSITARIO SON ESPASES | Servicio de Microbiología, Hospital Universitario Son Espases |
| EPI_ISL_2626128 | HOSPITAL MANACOR | HOSPITAL UNIVERSITARIO SON ESPASES | Servicio de Microbiología, Hospital Universitario Son Espases |
| EPI_ISL_2626131 | HOSPITAL UNIVERSITARIO SON ESPASES | HOSPITAL UNIVERSITARIO SON ESPASES | Servicio de Microbiología, Hospital Universitario Son Espases |
| EPI_ISL_2626147, EPI_ISL_2626155 | HOSPITAL MANACOR | HOSPITAL UNIVERSITARIO SON ESPASES | Servicio de Microbiología, Hospital Universitario Son Espases |
| EPI_ISL_2626165 | HOSPITAL CAN MISSES | HOSPITAL UNIVERSITARIO SON ESPASES | Servicio de Microbiología, Hospital Universitario Son Espases |
| EPI_ISL_2626168 | HOSPITAL MANACOR | HOSPITAL UNIVERSITARIO SON ESPASES | Servicio de Microbiología, Hospital Universitario Son Espases |
| EPI_ISL_2626704, | Alaska State Virology | Alaska State Virology | Stephanie DeRonde, Elva House, Jacob Zidek, Lisa Smith, Ph.D., Jack Chen, Ph.D. |

| | | | |
|---|---|---|---|
| EPI_ISL_2626742, EPI_ISL_2626747 | Laboratory | Laboratory | |
| EPI_ISL_2626784, EPI_ISL_2626796 | Baylor Scott & White-Temple | Baylor Scott & White-Temple | Ari Rao, Linden Morales, Kimberly Walker, Marcus Volz, Shelby Johnson |
| EPI_ISL_2626932, EPI_ISL_2626937, EPI_ISL_2626939, EPI_ISL_2626940, EPI_ISL_2626953, EPI_ISL_2626958, EPI_ISL_2626971 | TEMPUS LABS INC | Wadsworth Center, New York State Department of Health | Kirsten St. George, Daryl M. Lamson, Alexis Russell, Matthew Shudt, Melissa A Leisner, Jonathan Plitnick, Catharine Prussing, Navjot Singh, John Kelly, Erasmus Schneider, Erica Lasek-Nesselquist |
| EPI_ISL_2626987 | URMC LABS | Wadsworth Center, New York State Department of Health | Kirsten St. George, Daryl M. Lamson, Alexis Russell, Matthew Shudt, Melissa A Leisner, Jonathan Plitnick, Catharine Prussing, Navjot Singh, John Kelly, Erasmus Schneider, Erica Lasek-Nesselquist |
| EPI_ISL_2627000, EPI_ISL_2627002 | ALBANY MEDICAL CENTER | Wadsworth Center, New York State Department of Health | Kirsten St. George, Daryl M. Lamson, Alexis Russell, Matthew Shudt, Melissa A Leisner, Jonathan Plitnick, Catharine Prussing, Navjot Singh, John Kelly, Erasmus Schneider, Erica Lasek-Nesselquist |
| EPI_ISL_2627144, EPI_ISL_2627145 | MONTEFIORE MEDICAL CENTER LABORATORIES | Wadsworth Center, New York State Department of Health | Kirsten St. George, Daryl M. Lamson, Alexis Russell, Matthew Shudt, Melissa A Leisner, Jonathan Plitnick, Catharine Prussing, Navjot Singh, John Kelly, Erasmus Schneider, Erica Lasek-Nesselquist |
| EPI_ISL_2627770 | NCSLPH | NCSLPH | Chase K, Miller MC, Greene S, Glover W |
| EPI_ISL_2629191, EPI_ISL_2629195, EPI_ISL_2629206, EPI_ISL_2629209, EPI_ISL_2629216 | Microbiology Division, South Carolina Department of Health and Environmental Control | Microbiology Division, South Carolina Department of Health and Environmental Control | Freeman,J.S., Flores,H.V. |
| EPI_ISL_2631368, EPI_ISL_2631373, EPI_ISL_2631377 | Pandemic Response Lab - NYC | Pandemic Response Lab, R&D | Henry Lee, Michael Hammerling, Melissa Hopkins, Cybill del Castillo, Shinyoung Clair Kang, William Ward, Pradeep Bugga, Sol Rey, Dylan Law, Katharine Nelson, Haiping Hao, Jon Laurent |
| EPI_ISL_2631590, EPI_ISL_2631596 | SYNLAB MVZ Trier | Robert Koch Institute | unknown |
| EPI_ISL_2633584 | CENTOGENE Frankfurt Laboratory: Niederlassung Industriepark Höchst | Robert Koch Institute | unknown |
| EPI_ISL_2634588 | Eurofins LifeCodexx GmbH | Robert Koch Institute | unknown |
| EPI_ISL_2634936 | Pandemic Response Lab - NYC | Pandemic Response Lab, R&D | Henry Lee, Michael Hammerling, Melissa Hopkins, Cybill del Castillo, Shinyoung Clair Kang, William Ward, Pradeep Bugga, Sol Rey, Dylan Law, Katharine Nelson, Haiping Hao, Jon Laurent |
| EPI_ISL_2645310, EPI_ISL_2645313, EPI_ISL_2645315 | SC Dept of Health and Env. Control-Bureau of Laboratories | Centers for Disease Control and Prevention Division of Viral Diseases, Pathogen Discovery | Mili Sheth, Sarah Nobles, Jasmine Padilla, Mark Burroughs, Shoshona Le, Katie Dillon, Peter Cook, Clinton R. Paden, Dhwani Batra, Krista Queen, Kristen Knipe, Dakota Howard, Yvette Unoarumhi, Darlene Wagner, Matthew Schmerer, Ben L. Rambo-Martin, Kristine Lacek, Sam Shepard, Alison Laufer Halpin, Dave Wentworth, Vivien Dugan, Suxiang Tong, Justin Lee |
| EPI_ISL_2645351 | RI State Health Laboratories | Centers for Disease Control and Prevention Division of Viral Diseases, Pathogen Discovery | Mili Sheth, Sarah Nobles, Jasmine Padilla, Mark Burroughs, Shoshona Le, Katie Dillon, Peter Cook, Clinton R. Paden, Dhwani Batra, Krista Queen, Kristen Knipe, Dakota Howard, Yvette Unoarumhi, Darlene Wagner, Matthew Schmerer, Ben L. Rambo-Martin, Kristine Lacek, Sam Shepard, Alison Laufer Halpin, Dave Wentworth, Vivien Dugan, Suxiang Tong, Justin Lee |
| EPI_ISL_2645373 | NH Dept. of Health and Human Services Public Health Labs | Centers for Disease Control and Prevention Division of Viral Diseases, Pathogen Discovery | Mili Sheth, Sarah Nobles, Jasmine Padilla, Mark Burroughs, Shoshona Le, Katie Dillon, Peter Cook, Clinton R. Paden, Dhwani Batra, Krista Queen, Kristen Knipe, Dakota Howard, Yvette Unoarumhi, Darlene Wagner, Matthew Schmerer, Ben L. Rambo-Martin, Kristine Lacek, Sam Shepard, Alison Laufer Halpin, Dave Wentworth, Vivien Dugan, Suxiang Tong, Justin Lee |
| EPI_ISL_2647397, EPI_ISL_2647462, EPI_ISL_2647490, EPI_ISL_2647497, EPI_ISL_2647514, EPI_ISL_2647531, EPI_ISL_2647591, EPI_ISL_2647597, EPI_ISL_2647606, EPI_ISL_2647632, EPI_ISL_2647640, EPI_ISL_2647660, EPI_ISL_2647677, EPI_ISL_2647697, EPI_ISL_2647703, EPI_ISL_2647719, EPI_ISL_2647808, EPI_ISL_2647817, EPI_ISL_2647829, EPI_ISL_2647892, EPI_ISL_2647893, EPI_ISL_2647895, EPI_ISL_2647908, EPI_ISL_2647955, EPI_ISL_2648005 | | | |
| see above | Mako Medical | Centers for Disease Control and Prevention Division of Viral Diseases, Pathogen Discovery | Dakota Howard, Dhwani Batra, Peter W. Cook, Kara Moser, Adrian Paskey, Jason Caravas, Benjamin Rambo-Martin, Shatavia Morrison, Christopher Gulvick, Scott Sammons, Yvette Unoarumhi, Darlene Wagner, Matthew Schmerer,Matthew Tugwell, Lauren Moon, Clinton R. Paden, Duncan MacCannell |
| EPI_ISL_2649850, EPI_ISL_2649875, EPI_ISL_2649883, EPI_ISL_2649884 | NJDOH, Public Health and Environmental Laboratories | NJ_PHEL | Lindsey Bodnar, Shiv K. Verma, Ryan Pachucki, Chelsea San Filippo, Jacquelyn Deverell, Dana Woell, Allison Roder, Byeong Jeong |
| EPI_ISL_2650248 | Microbiologia CATLAB | Can Ruti SARS-CoV-2 Sequencing Hub (HUGTiP/IrsiCaixa/IGTP) | Marc Noguera-Julian, Pilar Armengol, Ignacio Blanco, Antoni E Bordoy, Francesc Catala-Moll, Pere-Joan Cardona, Maria Casadellà, Cristina Casañ, Gemma Clara, Bonaventura Clotet, Cristina Esteban, Montserrat Giménez, Mercedes Guerrero, Anna Not, Roger Paredes, Mariona Parera, Verónica Saludes, Alba Sánchez, and Elisa Martró on behalf of the Can Ruti SARS-CoV-2 Sequencing Hub |
| EPI_ISL_2650521 | University of Oregon COVID-19 MAP Laboratory | University of Oregon Genomics and Cell Characterization Core Facility (GC3F) | Douglas Turnbull, Ariana White, Jeff Bishop, Jason Carriere, Jason Sydes, Peter Batzel |
| EPI_ISL_2651044 | Washington State Department of Health Public Health Laboratories | Washington State Department of Health Public Health Laboratories | Drew MacKellar, Philip Dykema, Denny Russell, Joenice Gonzalez, Hannah Gray, Geoff Melly, Vanessa De Los Santos, Darren Lucas, JohnAric Peterson, Avi Singh, Rebecca Cao |
| EPI_ISL_2652041 | New Mexico Department | New Mexico Department | Ellie Johnson, D'eldra Malone, Jennifer Benoit, Ratheesh Rajan, Linda Salazar, |

| | | | |
|---|---|---|---|
| | of Health Scientific Laboratory | of Health Scientific Laboratory | Keila Gutierrez, Anastacia Griego-Fisher |
| EPI_ISL_2652270, EPI_ISL_2652296, EPI_ISL_2652320, EPI_ISL_2652329, EPI_ISL_2652330, EPI_ISL_2652355, EPI_ISL_2652370, EPI_ISL_2652385, EPI_ISL_2652418, EPI_ISL_2652420, EPI_ISL_2652426, EPI_ISL_2652430, EPI_ISL_2652471, EPI_ISL_2652475 | | | |
| see above | Quest Diagnostics Incorporated | Centers for Disease Control and Prevention Division of Viral Diseases, Pathogen Discovery | Dakota Howard, Dhwani Batra, Peter W. Cook, Kara Moser, Adrian Paskey, Jason Caravas, Benjamin Rambo-Martin, Shatavia Morrison, Christopher Gulvick, Scott Sammons, Yvette Unoarumhi, Darlene Wagner, Matthew Schmerer, S. H. Rosenthal, A. Gerasimova, R. M. Kagan, B. Anderson, M. Hua, Y. Liu, L.E. Bernstein, K.E. Livingston, A. Perez, I. A. Shlyakhter, R. V. Rolando, R. Owen, P. Tanpaiboon, F. Lacbawan, Clinton R. Paden, Duncan MacCannell |
| EPI_ISL_2652716, EPI_ISL_2652959, EPI_ISL_2652975, EPI_ISL_2653043, EPI_ISL_2653053, EPI_ISL_2653066 | Infinity Biologix | Centers for Disease Control and Prevention Division of Viral Diseases, Pathogen Discovery | Dakota Howard, Dhwani Batra, Peter W. Cook, Kara Moser, Adrian Paskey, Jason Caravas, Benjamin Rambo-Martin, Shatavia Morrison, Christopher Gulvick, Scott Sammons, Yvette Unoarumhi, Darlene Wagner, Matthew Schmerer, Christian Bixby, Yihe Wang, Jonathan Schultz, Chirayu Goswami, Russ Hager, Robin Grimwood, Clinton R. Paden, Duncan MacCannell |
| EPI_ISL_2657665, EPI_ISL_2657666 | NCSLPH | NCSLPH | Chase K, Miller MC, Greene S, Glover W |
| EPI_ISL_2658561, EPI_ISL_2658563 | Minnesota Department of Health, Public Health Laboratory | Minnesota Department of Health, Public Health Laboratory | Alexandra Lorentz, Jacob Garfin, Matt Plumb, and Xiong Wang |
| EPI_ISL_2660408 | Infectious Disease Program, Broad Institute of Harvard and MIT | Infectious Disease Program, Broad Institute of Harvard and MIT | Lemieux,J.E., Siddle,K.J., Shaw,B., Adams,G., Pierce,V., Turbett,S., Anahtar,M., Branda,J., Slater,D., Harris,J., Lin,A.E., Gladden-Young,A., Lagerborg,K., Rudy,M., DeRuff,K., Carter,A., Normandin,E., Bauer,M., Reilly,S., Tomkins-Tinch,C., Loreth,C., Chaluvadi,S., Neumann,A., Cusick,C., Chapman,S.B., Gnirke,A., Flowers,K., Cerrato,F., Birren,B.W., Gallagher,G., Smole,S., Park,D.J., MacInnis,B.L., Ryan,E., LaRocque,R., Rosenberg,E., Sabeti,P.C. |
| EPI_ISL_2661536, EPI_ISL_2661537 | UHTL, University Hospitals | UHTL, University Hospitals | Sadri,N., Alouani,D., Song,X. |
| EPI_ISL_2662751, EPI_ISL_2662767, EPI_ISL_2662770, EPI_ISL_2662778, EPI_ISL_2662814 | DPHL | Delaware Public Health Lab | Rebecca Savage |
| EPI_ISL_2663233 | Mayo Clinic & Mayo Clinic Laboratories | Minnesota Department of Health, Public Health Laboratory | Alexandra Lorentz, Jacob Garfin, Matt Plumb, and Xiong Wang |
| EPI_ISL_2663500, EPI_ISL_2663503 | Virginia Division of Consolidated Laboratory Services | Virginia Division of Consolidated Laboratory Services | Virginia Division of Consolidated Laboratory Services |
| EPI_ISL_2663516, EPI_ISL_2663517, EPI_ISL_2663534, EPI_ISL_2663537, EPI_ISL_2663540, EPI_ISL_2663541, EPI_ISL_2663542, EPI_ISL_2663543, EPI_ISL_2663550 | NYU Langone Health | Departments of Pathology and Medicine, New York University School of Medicine | Adriana Heguy, Dacia Dimartino, Emily Guzman, Christian Marier, Peter Meyn, Sitharam Ramaswami, Gael Westby, Paul Zappile, Yutong Zhang, Paolo Cotzia, Guiqing Wang |
| EPI_ISL_2665592 | Department of Bacteria, Parasites and Fungi, Statens Serum Institut, Copenhagen, Denmark | Statens Serum Institut Bioinformatics and Microbial Genomics | Danish Covid-19 Genome Consortium |
| EPI_ISL_2671584 | Centro de Investigacion Biomedica de Occidente (CIBO) | Unidad de Genomica Avanzada | Consorcio Mexicano de Vigilancia Genomica (CoViGen-Mex). Authors (in alphabetical order): Julio Elias Alvarado-Yaah, Carlos F. Arias, Santiago avila-Rios, Eduardo Becerril-Vargas, Victor Hugo Borja-Aburto, Celia Boukadida, Cristobal Chaidez-Quiroz, Juan Bautista Chale-Dzul, Ricardo Ciria Merce, Andreu Comas-Garcia, Celida Duque Molina, Julissa Enciso-Ibarra, Jose Antonio Enciso-Moreno, Gloria Elena Espinosa-Ayala, Fernando Fontove-Herrera, Daniel Fregoso-Rueda, Victor Eduardo Garcia-Arias, Alejandra Garcia-Gasca, Bruno Gomez-Gil, Jean Pierre Gonzalez, Irvin Gonzalez-Lopez, Concepcion Grajales-Muñiz, Ricardo Grande, Rosa Maria Gutierrez Rios, Jesus Hernandez, Alejandra Hernandez-Teran, Alfredo Herrera-Estrella, Carla Ivon Herrera-Najera, Pavel Isa, Veronica Mata-Haro, Daniel Lira Morales, Susana Lopez, Antonio Loza Roman, Brenda Irasema Maldonado-Meza, Bernardo Martinez-Miguel, Jose Arturo Martinez-Orozco, Celida Martinez- Rodriguez, Margarita Matias-Florentino, Fidencio Mejia-Nepomuceno, Maria Guadalupe de Jesus Mireles-Rivera, Gloria Maria Molina-Salinas, Hector Montoya-Fuentes, , Mario Mujica-Sanchez, Jose Esteban Muñoz-Medina, Jose de Jesus Nuñez-Contreras, Alicia Ocaña-Mondragon, Luis Alberto Ochoa-Carrera, Hector Esteban Paz-Juarez, Marissa Perez-Garcia, Francisco Pulido, Helen Haydee Fernanda Ramirez-Plascencia, Jorge Salas-Hernandez, Angel Gustavo Salas-Lais, Alejandro Sanchez-Flores, Clara Esperanza Santacruz-Tinoco, Maria Guadalupe Santiago-Mauricio, Selene Zarate, Nelly Selem-Mojica, Blanca Taboada, Gloria Vazquez, Joel Armando Vazquez-Perez. |
| EPI_ISL_2673234, EPI_ISL_2673252, EPI_ISL_2673262, EPI_ISL_2673274 | Dutch COVID-19 response team | National Institute for Public Health and the Environment (RIVM) | Adam Meijer, Harry Vennema, Dirk Eggink, Jeroen Cremer, Sharon van den Brink, Bas van der Veer, AnneMarie van den Brandt, Lisa Wijsman, Kim Freriks, Ryanne Jaarsma, Euníce Then, Lynn Aarts, Sanne Bos, Melissa van Tuil, Linda van de Nes, Florian Zwagemaker, Dennis Schmitz, Annelies Kroneman, Karim Hajji, Chantal Reusken, on behalf of the national COVID-19 response team |
| EPI_ISL_2678987, EPI_ISL_2678989, EPI_ISL_2679303 | Utah Public Health Laboratory | Utah Public Health Laboratory | Erin L. Young, Kelly F. Oakeson, Tara Gallagher |

| | | | |
|---|---|---|---|
| EPI_ISL_2680476 | Humboldt County Public Health Laboratory | Humboldt County Public Health Laboratory | Jeremy Corrigan |
| EPI_ISL_2681110 | Centro de Investigación Biomédica del Noreste (CIBIN) | Instituto de Biotecnología de la UNAM | Consorcio Mexicano de Vigilancia Genómica (CoViGen-Mex). Authors (in alphabetical order): Julio Elias Alvarado-Yaah, Carlos F. Arias, Santiago Ávila-Ríos, Eduardo Becerril-Vargas, Víctor Hugo Borja-Aburto, Celia Boukadida, Cristóbal Cháidez-Quiróz, Juan Bautista Chale-Dzul, Ricardo Ciria Merce, Andreu Comas-García, Célida Duque Molina, Julissa Enciso-Ibarra, José Antonio Enciso-Moreno, Gloria Elena Espinosa-Ayala, Fernando Fontove-Herrera, Daniel Fregoso-Rueda, Victor Eduardo García-Arias, Alejandra García-Gasca, Bruno Gómez-Gil, Jean Pierre González, Irvin González-López, Concepción Grajales-Muñiz, Ricardo Grande, Rosa María Gutiérrez Rios, Jesús Hernández, Alejandra Hernández-Terán, Alfredo Herrera-Estrella, Carla Ivón Herrera-Najera, Pavel Isa, Verónica Mata-Haro, Daniel Lira Morales, Susana Lopez, Antonio Loza Román, Brenda Irasema Maldonado-Meza, Bernardo Martínez-Miguel, José Arturo Martínez-Orozco, Célida Martínez- Rodríguez, Margarita Matías-Florentino, Fidencio Mejía-Nepomuceno, María Guadalupe de Jesús Mireles-Rivera, Gloria María Molina-Salinas, Hector Montoya-Fuentes, , Mario Mújica-Sánchez, José Esteban Muñoz-Medina, José de Jesús Nuñez-Contreras, Alicia Ocaña-Mondragón, Luis Alberto Ochoa-Carrera, Hector Esteban Paz-Juárez, Marissa Perez-Garcia, Francisco Pulido, Helen Haydee Fernanda Ramírez-Plascencia, Jorge Salas-Hernández, Angel Gustavo Salas-Lais, Alejandro Sánchez-Flores, Clara Esperanza Santacruz-Tinoco, María Guadalupe Santiago-Mauricio, Selene Zárate, Nelly Sélem-Mojica, Blanca Taboada, Gloria Vazquez, Joel Armando Vázquez-Pérez. |
| EPI_ISL_2681221 | Unidad de Investigación Biomédica de Zacatecas (UIBZ) | Instituto de Biotecnología de la UNAM | Consorcio Mexicano de Vigilancia Genómica (CoViGen-Mex). Authors (in alphabetical order): Julio Elias Alvarado-Yaah, Carlos F. Arias, Santiago Ávila-Ríos, Eduardo Becerril-Vargas, Víctor Hugo Borja-Aburto, Celia Boukadida, Cristóbal Cháidez-Quiróz, Juan Bautista Chale-Dzul, Ricardo Ciria Merce, Andreu Comas-García, Célida Duque Molina, Julissa Enciso-Ibarra, José Antonio Enciso-Moreno, Gloria Elena Espinosa-Ayala, Fernando Fontove-Herrera, Daniel Fregoso-Rueda, Victor Eduardo García-Arias, Alejandra García-Gasca, Bruno Gómez-Gil, Jean Pierre González, Irvin González-López, Concepción Grajales-Muñiz, Ricardo Grande, Rosa María Gutiérrez Rios, Jesús Hernández, Alejandra Hernández-Terán, Alfredo Herrera-Estrella, Carla Ivón Herrera-Najera, Pavel Isa, Verónica Mata-Haro, Daniel Lira Morales, Susana Lopez, Antonio Loza Román, Brenda Irasema Maldonado-Meza, Bernardo Martínez-Miguel, José Arturo Martínez-Orozco, Célida Martínez- Rodríguez, Margarita Matías-Florentino, Fidencio Mejía-Nepomuceno, María Guadalupe de Jesús Mireles-Rivera, Gloria María Molina-Salinas, Hector Montoya-Fuentes, , Mario Mújica-Sánchez, José Esteban Muñoz-Medina, José de Jesús Nuñez-Contreras, Alicia Ocaña-Mondragón, Luis Alberto Ochoa-Carrera, Hector Esteban Paz-Juárez, Marissa Perez-Garcia, Francisco Pulido, Helen Haydee Fernanda Ramírez-Plascencia, Jorge Salas-Hernández, Angel Gustavo Salas-Lais, Alejandro Sánchez-Flores, Clara Esperanza Santacruz-Tinoco, María Guadalupe Santiago-Mauricio, Selene Zárate, Nelly Sélem-Mojica, Blanca Taboada, Gloria Vazquez, Joel Armando Vázquez-Pérez. |
| EPI_ISL_2686412, EPI_ISL_2686414, EPI_ISL_2686455, EPI_ISL_2686506, EPI_ISL_2686603, EPI_ISL_2686634, EPI_ISL_2686677, EPI_ISL_2686700, EPI_ISL_2686701, EPI_ISL_2686703, EPI_ISL_2686748 | | | |
| see above | Fulgent Genetics | Centers for Disease Control and Prevention Division of Viral Diseases, Pathogen Discovery | Dakota Howard, Dhwani Batra, Peter W. Cook, Kara Moser, Adrian Paskey, Jason Caravas, Benjamin Rambo-Martin, Shatavia Morrison, Christopher Gulvick, Scott Sammons, Yvette Unoarumhi, Darlene Wagner, Matthew Schmerer, Harry Gao, Mickey Li, John Gao, Joseph Fierro, Benafsh Sapra, Becky Tsai, Yan Meng, Doreen Ng, James Xie, Clinton R. Paden, Duncan MacCannell |
| EPI_ISL_2686842, EPI_ISL_2686858, EPI_ISL_2686876 | Infinity Biologix | Centers for Disease Control and Prevention Division of Viral Diseases, Pathogen Discovery | Dakota Howard, Dhwani Batra, Peter W. Cook, Kara Moser, Adrian Paskey, Jason Caravas, Benjamin Rambo-Martin, Shatavia Morrison, Christopher Gulvick, Scott Sammons, Yvette Unoarumhi, Darlene Wagner, Matthew Schmerer, Christian Bixby, Yihe Wang, Jonathan Schultz, Chirayu Goswami, Russ Hager, Robin Grimwood, Clinton R. Paden, Duncan MacCannell |
| EPI_ISL_2686915, EPI_ISL_2686931, EPI_ISL_2686945, EPI_ISL_2686951, EPI_ISL_2686957, EPI_ISL_2687007, EPI_ISL_2687008, EPI_ISL_2687016, EPI_ISL_2687017, EPI_ISL_2687064, EPI_ISL_2687071, EPI_ISL_2687072, EPI_ISL_2687079, EPI_ISL_2687082, EPI_ISL_2687116, EPI_ISL_2687122, EPI_ISL_2687140, EPI_ISL_2687173, EPI_ISL_2687216, EPI_ISL_2687271, EPI_ISL_2687384, EPI_ISL_2687430, EPI_ISL_2687441, EPI_ISL_2687466, EPI_ISL_2687484, EPI_ISL_2687519, EPI_ISL_2687552 | | | |
| see above | Aegis Sciences Corporation | Centers for Disease Control and Prevention Division of Viral Diseases, Pathogen Discovery | Dakota Howard, Dhwani Batra, Peter W. Cook, Kara Moser, Adrian Paskey, Jason Caravas, Benjamin Rambo-Martin, Shatavia Morrison, Christopher Gulvick, Scott Sammons, Yvette Unoarumhi, Darlene Wagner, Matthew Schmerer, Cyndi Clark, Patrick Campbell, Rob Case, Vikramsinha Ghorpade, Holly Houdeshell, Ola Kvalvaag, Dillon Nall, Ethan Sanders, Alec Vest, Shaun Westlund, Matthew Hardison, Clinton R. Paden, Duncan MacCannell |
| EPI_ISL_2688317 | Puerto Rico Department of Health | Centers for Disease Control and Prevention, Dengue Branch | Gilberto A. Santiago, Glenda Gonzalez, Betzabel Flores, Keyla Charriez, Gabriela Paz-Bailey, Jorge L. Munoz-Jordan |
| EPI_ISL_2688999 | DC Public Health Lab/ Dept. of Forensic Sciences | DC Public Health Lab/ Dept. of Forensic Sciences | Janis Doss, Scott Nguyen, Elizabeth Zelaya, Sarah Scott, Connie Maza, Monica Mann, Brittany Hamilton, David Payne, Jocelyn Hauser |
| EPI_ISL_2689031, EPI_ISL_2689032, EPI_ISL_2689037 | Mayo Clinic & Mayo Clinic Laboratories | Minnesota Department of Health, Public Health Laboratory | Alexandra Lorentz, Jacob Garfin, Matt Plumb, and Xiong Wang |
| EPI_ISL_2689057, EPI_ISL_2689058, EPI_ISL_2689102 | MD PHL | MD PHL | Maryland Department of Health Laboratories Administration |
| EPI_ISL_2689601 | GA Department of Public Health Laboratory | Genomics and Discovery, Respiratory Viruses Branch, Division of Viral Diseases, | Jing Zhang, Anna Kelleher, Ying Tao, Yan Li, Brian Lynch, Krista Queen, Anna Uehara, Peter Cook, Han Jia Justin Ng, Rachel Marine, Clinton R. Paden, Dhwani Batra, Haibin Wang, Tara Coalter, Jasmine Padilla, Morgan Davis, Mili Sheth, Sarah Nobles, Mark Burroughs, Justin Lee, Adam Retchless, Suxiang Tong |

| | | | |
|---|---|---|---|
| | | Centers for Disease Control and Prevention | |
| EPI_ISL_2689603 | OK Public Health Laboratory, Oklahoma State DOH | Genomics and Discovery, Respiratory Viruses Branch, Division of Viral Diseases, Centers for Disease Control and Prevention | Jing Zhang, Anna Kelleher, Ying Tao, Yan Li, Brian Lynch, Krista Queen, Anna Uehara, Peter Cook, Han Jia Justin Ng, Rachel Marine, Clinton R. Paden, Dhwani Batra, Haibin Wang, Tara Coalter, Jasmine Padilla, Morgan Davis, Mili Sheth, Sarah Nobles, Mark Burroughs, Justin Lee, Adam Retchless, Suxiang Tong |
| EPI_ISL_2689848, EPI_ISL_2689856, EPI_ISL_2689863 | Virginia Division of Consolidated Laboratory Services | Virginia Division of Consolidated Laboratory Services | Virginia Division of Consolidated Laboratory Services |
| EPI_ISL_2689910, EPI_ISL_2689911 | RI State Health Laboratories | Centers for Disease Control and Prevention Division of Viral Diseases, Pathogen Discovery | Mili Sheth, Sarah Nobles, Jasmine Padilla, Mark Burroughs, Shoshona Le, Katie Dillon, Peter Cook, Clinton R. Paden, Dhwani Batra, Krista Queen, Kristen Knipe, Dakota Howard, Yvette Unoarumhi, Darlene Wagner, Matthew Schmerer, Ben L. Rambo-Martin, Kristine Lacek, Sam Shepard, Alison Laufer Halpin, Dave Wentworth, Vivien Dugan, Suxiang Tong, Justin Lee |
| EPI_ISL_2689961 | LA Office of Public Health Laboratories | Centers for Disease Control and Prevention Division of Viral Diseases, Pathogen Discovery | Mili Sheth, Sarah Nobles, Jasmine Padilla, Mark Burroughs, Shoshona Le, Katie Dillon, Peter Cook, Clinton R. Paden, Dhwani Batra, Krista Queen, Kristen Knipe, Dakota Howard, Yvette Unoarumhi, Darlene Wagner, Matthew Schmerer, Ben L. Rambo-Martin, Kristine Lacek, Sam Shepard, Alison Laufer Halpin, Dave Wentworth, Vivien Dugan, Suxiang Tong, Justin Lee |
| EPI_ISL_2689980 | OR State PHL-Virology/Immunology Section | Centers for Disease Control and Prevention Division of Viral Diseases, Pathogen Discovery | Mili Sheth, Sarah Nobles, Jasmine Padilla, Mark Burroughs, Shoshona Le, Katie Dillon, Peter Cook, Clinton R. Paden, Dhwani Batra, Krista Queen, Kristen Knipe, Dakota Howard, Yvette Unoarumhi, Darlene Wagner, Matthew Schmerer, Ben L. Rambo-Martin, Kristine Lacek, Sam Shepard, Alison Laufer Halpin, Dave Wentworth, Vivien Dugan, Suxiang Tong, Justin Lee |
| EPI_ISL_2690005 | IN State Department of Health Laboratory Services | Centers for Disease Control and Prevention Division of Viral Diseases, Pathogen Discovery | Mili Sheth, Sarah Nobles, Jasmine Padilla, Mark Burroughs, Shoshona Le, Katie Dillon, Peter Cook, Clinton R. Paden, Dhwani Batra, Krista Queen, Kristen Knipe, Dakota Howard, Yvette Unoarumhi, Darlene Wagner, Matthew Schmerer, Ben L. Rambo-Martin, Kristine Lacek, Sam Shepard, Alison Laufer Halpin, Dave Wentworth, Vivien Dugan, Suxiang Tong, Justin Lee |
| EPI_ISL_2690023 | UT-Unified State Labs: Public Health Utah DOH | Centers for Disease Control and Prevention Division of Viral Diseases, Pathogen Discovery | Mili Sheth, Sarah Nobles, Jasmine Padilla, Mark Burroughs, Shoshona Le, Katie Dillon, Peter Cook, Clinton R. Paden, Dhwani Batra, Krista Queen, Kristen Knipe, Dakota Howard, Yvette Unoarumhi, Darlene Wagner, Matthew Schmerer, Ben L. Rambo-Martin, Kristine Lacek, Sam Shepard, Alison Laufer Halpin, Dave Wentworth, Vivien Dugan, Suxiang Tong, Justin Lee |
| EPI_ISL_2690074 | ID Bureau of Laboratories | Centers for Disease Control and Prevention Division of Viral Diseases, Pathogen Discovery | Mili Sheth, Sarah Nobles, Jasmine Padilla, Mark Burroughs, Shoshona Le, Katie Dillon, Peter Cook, Clinton R. Paden, Dhwani Batra, Krista Queen, Kristen Knipe, Dakota Howard, Yvette Unoarumhi, Darlene Wagner, Matthew Schmerer, Ben L. Rambo-Martin, Kristine Lacek, Sam Shepard, Alison Laufer Halpin, Dave Wentworth, Vivien Dugan, Suxiang Tong, Justin Lee |
| EPI_ISL_2690113 | WA State Department of Health | Centers for Disease Control and Prevention Division of Viral Diseases, Pathogen Discovery | Mili Sheth, Sarah Nobles, Jasmine Padilla, Mark Burroughs, Shoshona Le, Katie Dillon, Peter Cook, Clinton R. Paden, Dhwani Batra, Krista Queen, Kristen Knipe, Dakota Howard, Yvette Unoarumhi, Darlene Wagner, Matthew Schmerer, Ben L. Rambo-Martin, Kristine Lacek, Sam Shepard, Alison Laufer Halpin, Dave Wentworth, Vivien Dugan, Suxiang Tong, Justin Lee |
| EPI_ISL_2690116, EPI_ISL_2690119 | PR Public Health Lab | Centers for Disease Control and Prevention Division of Viral Diseases, Pathogen Discovery | Mili Sheth, Sarah Nobles, Jasmine Padilla, Mark Burroughs, Shoshona Le, Katie Dillon, Peter Cook, Clinton R. Paden, Dhwani Batra, Krista Queen, Kristen Knipe, Dakota Howard, Yvette Unoarumhi, Darlene Wagner, Matthew Schmerer, Ben L. Rambo-Martin, Kristine Lacek, Sam Shepard, Alison Laufer Halpin, Dave Wentworth, Vivien Dugan, Suxiang Tong, Justin Lee |
| EPI_ISL_2690121, EPI_ISL_2690123 | MT Public Health Laboratory | Centers for Disease Control and Prevention Division of Viral Diseases, Pathogen Discovery | Mili Sheth, Sarah Nobles, Jasmine Padilla, Mark Burroughs, Shoshona Le, Katie Dillon, Peter Cook, Clinton R. Paden, Dhwani Batra, Krista Queen, Kristen Knipe, Dakota Howard, Yvette Unoarumhi, Darlene Wagner, Matthew Schmerer, Ben L. Rambo-Martin, Kristine Lacek, Sam Shepard, Alison Laufer Halpin, Dave Wentworth, Vivien Dugan, Suxiang Tong, Justin Lee |
| EPI_ISL_2690357, EPI_ISL_2690387 | Arizona State University | Arizona State University | Peter T. Skidmore, LaRinda A. Holland, Matthew F. Smith, Rabia Maqsood, Nicholas J. Mellor, Joy M. Blain, Valerie Harris, Joshua LaBaer, Vel Murugan, Efrem S. Lim |
| EPI_ISL_2691299 | Santa Clara Valley Medical Center | Santa Clara County Public Health Laboratory | Santa Clara County Public Health Department |
| EPI_ISL_2691950 | Rush University Medical Center | RIPHL at Rush University Medical Center | Stefan Green, Kevin Kunstman, Max Kolton, Marieta Hyde, Laura Furtado, Felix Araujo Perez, Mary Hayden, Joyce Houlihan, Diane Springer |
| EPI_ISL_2692020, EPI_ISL_2692021 | University of Chicago Medicine | RIPHL at Rush University Medical Center | Stefan Green, Kevin Kunstman, Max Kolton, Marieta Hyde, Laura Furtado, Felix Araujo Perez, Lauren Megger |
| EPI_ISL_2693029, EPI_ISL_2693030 | Oregon State Public Health Laboratory | Oregon State Public Health Laboratory | Rafia Razzaque, Eugene Yeboah, Vanda Makris, Laura Tsaknaridis, John Fontana and Shane Sevey |
| EPI_ISL_2693137, EPI_ISL_2693141, EPI_ISL_2693142, EPI_ISL_2693266, EPI_ISL_2693288, EPI_ISL_2693292, EPI_ISL_2693311 | UW Virology Lab | UW Virology Lab | Pavitra Roychoudhury, Hong Xie, Lasata Shrestha, Tien V. Nguyen, Shah Mohamed Bakhash, Michelle Lin, Noah R. Baker, Ricardo Perez, Sean Ellis, Nathan Breit, Robert J. Livingston, Meei-Li Huang, Keith R Jerome, Patrick Mathias, Alexander Greninger |
| EPI_ISL_2693678, EPI_ISL_2693682, EPI_ISL_2693693, EPI_ISL_2693711, EPI_ISL_2693714, EPI_ISL_2693721, EPI_ISL_2693722, EPI_ISL_2693723, EPI_ISL_2693730, EPI_ISL_2693734, EPI_ISL_2693735, EPI_ISL_2693745, EPI_ISL_2693773, EPI_ISL_2693774, EPI_ISL_2693788, EPI_ISL_2693789, EPI_ISL_2693791, EPI_ISL_2693799, EPI_ISL_2693808, EPI_ISL_2693818 | | | |
| see above | UW Virology Lab | UW Virology Lab | Pavitra Roychoudhury, Hong Xie, Lasata Shrestha, Shah Mohamed Bakhash, Tien V. Nguyen, Noah R. Baker, Sean Ellis, Meei-Li Huang, Keith R Jerome, Alexander Greninger |

| | | | |
|---|---|---|---|
| EPI_ISL_2699440, EPI_ISL_2699466 | Public Health Laboratory, Minnesota Department of Health | University of Minnesota Genomics Center | Daryl M. Gohl, John Garbe, Jaquelyn Kuriger-Laber, Corbin Dirkx, and Sean Wang |
| EPI_ISL_2709537 | Aegis Sciences Corporation | Centers for Disease Control and Prevention Division of Viral Diseases, Pathogen Discovery | Dakota Howard, Dhwani Batra, Peter W. Cook, Kara Moser, Adrian Paskey, Jason Caravas, Benjamin Rambo-Martin, Shatavia Morrison, Christopher Gulvick, Scott Sammons, Yvette Unoarumhi, Darlene Wagner, Matthew Schmerer, Cyndi Clark, Patrick Campbell, Rob Case, Vikramsinha Ghorpade, Holly Houdeshell, Ola Kvalvaag, Dillon Nall, Ethan Sanders, Alec Vest, Shaun Westlund, Matthew Hardison, Clinton R. Paden, Duncan MacCannell |
| EPI_ISL_2709687 | LESP Baja California | Instituto de Diagnostico y Referencia Epidemiologicos (INDRE) | Claudia Wong-Arambula, Abril Rodriguez-Maldonado, Vanessa Rivero-Arredondo, Ariadna Medina-Benitez, Joaquin Quiroz-Mercado, Sergio Rangel-Guerrero, Natividad Cruz-Ortiz, Tatiana Nunez-Garcia, Gisela Barrera-Badillo, Lucia Hernandez-Rivas, Irma Lopez-Martinez, Ernesto Ramirez-Gonzalez. |
| EPI_ISL_2710201 | HOSPITAL UNIVERSITARIO SON ESPASES | HOSPITAL UNIVERSITARIO SON ESPASES | Servicio de Microbiología, Hospital Universitario Son Espases |
| EPI_ISL_2712543, EPI_ISL_2712549, EPI_ISL_2712553 | Sharp HealthCare Laboratory | Andersen lab at Scripps Research | SEARCH Alliance San Diego with Aaron Harding, Jacquelyn Berumen, Cathy Woerle, Liam McGinnis, Art Mendoza, Omid Bakhtar |
| EPI_ISL_2713062 | Oregon State Public Health Laboratory | Oregon State Public Health Laboratory | Rafia Razzaque, Eugene Yeboah, Vanda Makris, Laura Tsaknaridis, John Fontana and Shane Sevey |
| EPI_ISL_2713292 | Infectious Disease Program, Broad Institute of Harvard and MIT | Infectious Disease Program, Broad Institute of Harvard and MIT | Lemieux,J.E., Siddle,K.J., Shaw,B., Adams,G., Pierce,V., Turbett,S., Anahtar,M., Branda,J., Slater,D., Harris,J., Lin,A.E., Gladden-Young,A., Lagerborg,K., Rudy,M., DeRuff,K., Carter,A., Normandin,E., Bauer,M., Reilly,S., Tomkins-Tinch,C., Loreth,C., Chaluvadi,S., Neumann,A., Cusick,C., Chapman,S.B., Gnirke,A., Flowers,K., Cerrato,F., Birren,B.W., Gallagher,G., Smole,S., Park,D.J., MacInnis,B.L., Ryan,E., LaRocque,R., Rosenberg,E., Sabeti,P.C. |
| EPI_ISL_2715247 | North Dakota Department of Health, Public Health Laboratory | North Dakota Department of Health, Public Health Laboratory | Lisa Wingerter |
| EPI_ISL_2716212, EPI_ISL_2716213, EPI_ISL_2716214 | The Jackson Laboratory | The Jackson Laboratory | Long J, Renzette N, Adams M, Omerza G, Kelly K, Li L |
| EPI_ISL_2716222, EPI_ISL_2716229, EPI_ISL_2716240, EPI_ISL_2716241, EPI_ISL_2716253, EPI_ISL_2716254 | Yale Clinical Virology Lab | Grubaugh Lab - Yale School of Public Health | Joseph Fauver, Mallery Breban, Isabel Ott, Tara Alpert, Mary Petrone, Anderson Brito, Chantal Vogels, Annie Watkins, Chaney Kalinich, Jessica Rothman, Marie L. Landry, Nathan Grubaugh |
| EPI_ISL_2716916 | Southern Nevada Public Health Laboratory | Southern Nevada Public Health Laboratory | Michael Picker |
| EPI_ISL_2725058 | IN State Department of Health Laboratory Services | IN State Department of Health Laboratory Services | Cassandra Campion, Jamie Yeadon, Brian Pope, Lixia Liu, Kyle Brownlee, Melissa Hindenlang, Mark Glazier |
| EPI_ISL_2725591 | Rhode Island State Health Laboratory | Rhode Island State Health Laboratory | Ewa King, Richard C. Huard, Kristin Carpenter-Azevedo |
| EPI_ISL_2727104, EPI_ISL_2727113 | ALBANY MEDICAL CENTER | Wadsworth Center, New York State Department of Health | Kirsten St. George, Daryl M. Lamson, Alexis Russell, Matthew Shudt, Melissa A Leisner, Jonathan Plitnick, Catharine Prussing, Navjot Singh, John Kelly, Erasmus Schneider, Erica Lasek-Nesselquist |
| EPI_ISL_2727473 | Oregon State Public Health Laboratory | Oregon State Public Health Laboratory | Rafia Razzaque, Eugene Yeboah, Vanda Makris, Laura Tsaknaridis, John Fontana and Shane Sevey |
| EPI_ISL_2727572, EPI_ISL_2727579, EPI_ISL_2727588 | NJDOH, Public Health and Environmental Laboratories | NJ_PHEL | Lindsey Bodnar, Shiv K. Verma, Jacquelyn Deverell, Ryan Pachucki, Dana Woell, Allison Roder, Byeong Jeong |
| EPI_ISL_2727726, EPI_ISL_2727728, EPI_ISL_2727737, EPI_ISL_2727743, EPI_ISL_2727784, EPI_ISL_2727802, EPI_ISL_2727805 | Wisconsin State Laboratory of Hygiene Communicable Disease Division | Wisconsin State Laboratory of Hygiene Communicable Disease Division | Abigail C. Shockey, Alicia J. Mooney, Erika M. Hanson, Tonya Danz, Richard Griesser, Sara Wagner, Kelsey R. Florek |
| EPI_ISL_2727871 | Centracare Laboratory Services | Minnesota Department of Health, Public Health Laboratory | Alexandra Lorentz, Jacob Garfin, Matt Plumb, and Xiong Wang |
| EPI_ISL_2728144 | Wyoming Public Health Laboratory | Wyoming Public Health Laboratory | Jim Mildenberger, Taylor Fearing, Channing Weber, Ashley Norberg, Chayse Rowley, Marley Goetz, Brian Dominguez, Elliot Thomasson, Sam Britz, Cari Sloma, Robert Petit, and Rob Christensen |
| EPI_ISL_2735900 | Seattle Flu Study | Seattle Flu Study | Deborah A. Nickerson, Chris D. Frazar, Jover Lee, Benjamin Pelle, Erica Ryke, Matthew Richardson, Amanda Adler, Elisabeth Brandstetter, Peter D. Han, Kairsten Fay, Misja Ilcisin, Kirsten Lacombe, Thomas R. Sibley, Melissa Truong, Caitlin R. Wolf, Karen Cowgill, Stephanie Schrag, Jeff Duchin, Michael Boeckh, Janet A. Englund, Michael Famulare, Barry R. Lutz, Mark J. Rieder, Lea M. Starita, Matthew Thompson, Helen Y. Chu, Trevor Bedford, Jay Shendure |
| EPI_ISL_2735987 | Incyte Diagnostics Spokane | Seattle Flu Study | Deborah A. Nickerson, Chris D. Frazar, Jover Lee, Benjamin Pelle, Erica Ryke, Matthew Richardson, Amanda Adler, Elisabeth Brandstetter, Peter D. Han, Kairsten Fay, Misja Ilcisin, Kirsten Lacombe, Thomas R. Sibley, Melissa Truong, Caitlin R. Wolf, Romesh Gautom, Geoff Melly, Brian Hiatt, Philip Dykema, Scott Lindquist, Michael Boeckh, Janet A. Englund, Michael Famulare, Barry R. Lutz, Mark J. Rieder, Lea M. Starita, Matthew Thompson, Helen Y. Chu, Jay Shendure, Trevor Bedford |
| EPI_ISL_2736230 | Atlas Genomics | Seattle Flu Study | Deborah A. Nickerson, Chris D. Frazar, Jover Lee, Benjamin Pelle, Erica Ryke, Matthew Richardson, Amanda Adler, Elisabeth Brandstetter, Peter D. Han, Kairsten |

| | | | |
|---|---|---|---|
| | | | Fay, Misja Ilcisin, Kirsten Lacombe, Thomas R. Sibley, Melissa Truong, Caitlin R. Wolf, Romesh Gautom, Geoff Melly, Brian Hiatt, Philip Dykema, Scott Lindquist, Michael Boeckh, Janet A. Englund, Michael Famulare, Barry R. Lutz, Mark J. Rieder, Lea M. Starita, Matthew Thompson, Helen Y. Chu, Jay Shendure, Trevor Bedford |
| EPI_ISL_2736837 | LESP Tabasco | Instituto de Diagnostico y Referencia Epidemiologicos (INDRE) | Claudia Wong-Arambula, Abril Rodriguez-Maldonado, Vanessa Rivero-Arredondo, Ariadna Medina-Benitez, Joaquin Quiroz-Mercado, Sergio Rangel-Guerrero, Natividad Cruz-Ortiz, Tatiana Nunez-Garcia, Gisela Barrera-Badillo, Lucia Hernandez-Rivas, Irma Lopez-Martinez, Ernesto Ramirez-Gonzalez. |
| EPI_ISL_2753764 | Missouri State Public Health Laboratory | Missouri State Public Health Laboratory | Matthew Sinn, Joshua Barry, Ashley New |
| EPI_ISL_2754008 | Kaiser Regional | Santa Clara County Public Health Laboratory | Santa Clara County Public Health Department |
| EPI_ISL_2754921 | GA Department of Public Health Laboratory | Genomics and Discovery, Respiratory Viruses Branch, Division of Viral Diseases, Centers for Disease Control and Prevention | Anna Kelleher, Ying Tao, Yan Li, Jing Zhang, Brian Lynch, Krista Queen, Anna Uehara, Peter Cook, Han Jia Justin Ng, Rachel Marine, Clinton R. Paden, Dhwani Batra, Haibin Wang, Tara Coalter, Jasmine Padilla, Morgan Davis, Mili Sheth, Sarah Nobles, Mark Burroughs, Justin Lee, Adam Retchless, Suxiang Tong |
| EPI_ISL_2757590, EPI_ISL_2757591, EPI_ISL_2757601, EPI_ISL_2757603, EPI_ISL_2757605, EPI_ISL_2757606, EPI_ISL_2757607, EPI_ISL_2757610, EPI_ISL_2757615, EPI_ISL_2757617, EPI_ISL_2757620, EPI_ISL_2757625, EPI_ISL_2757627, EPI_ISL_2757631, EPI_ISL_2757632, EPI_ISL_2757634, EPI_ISL_2757637, EPI_ISL_2757639, EPI_ISL_2757643, EPI_ISL_2757646, EPI_ISL_2757648 | | | |
| see above | INSPI-CRN DE INFLUENZA Y OTROS VIRUS RESPIRATORIOS | NIC-INSPI | Alfredo Bruno , Maritza Olmedo, Michelle Páez, Jimmy Garcés, Johanna Laines, Lizbeth Patiño, Manuel Gonzalez, Domenica de Mora. |
| EPI_ISL_2758278 | Berkeley Medical Center | WVU and Marshall University Combined Genomics Core Facilities | James Denvir, Peter Stoilov, Peter Perrotta, Wesley Kimble, Ryan Percifield |
| EPI_ISL_2758354, EPI_ISL_2758355, EPI_ISL_2758357, EPI_ISL_2758381, EPI_ISL_2758384, EPI_ISL_2758387, EPI_ISL_2758389, EPI_ISL_2758393 | QLabs | WVU and Marshall University Combined Genomics Core Facilities | James Denvir, Peter Stoilov, Peter Perrotta, Wesley Kimble, Ryan Percifield |
| EPI_ISL_2758617 | Microbiology Division, South Carolina Department of Health and Environmental Control | Microbiology Division, South Carolina Department of Health and Environmental Control | Freeman,J.S., Flores,H.V. |
| EPI_ISL_2758887, EPI_ISL_2758908 | The Ohio State University Applied Microbiology Services Laboratory | The Ohio State University Applied Microbiology Services Laboratory | Seth A. Faith PhD |
| EPI_ISL_2769232 | TGen North | TGen North | "Jolene Bowers, Heather Centner, Brett Van Tassel, Chris French, Hayley Yaglom, Darrin Lemmer, Dave Engelthaler" |
| EPI_ISL_2774272 | Baylor Scott & White-Temple | Baylor Scott & White-Temple | Ari Rao, Linden Morales, Kimberly Walker, Marcus Volz, Shelby Johnson |
| EPI_ISL_2774596, EPI_ISL_2774608, EPI_ISL_2774613 | Santa Clara Valley Medical Center | Santa Clara County Public Health Laboratory | Santa Clara County Public Health Department |
